# Cerebrospinal Fluid and Plasma Metabolites with Parkinson’s Disease: A Mendelian Randomization Study

**DOI:** 10.1101/2024.07.19.24310687

**Authors:** Jia-Li Wang, Ran Zheng, Yi Fang, Jin Cao, Bao-Rong Zhang

**Affiliations:** Department of Neurology, The Second Affiliated Hospital, College of Medicine, Zhejiang University, Hangzhou, Zhejiang 310009, China

**Keywords:** Parkinson’s disease, Metabolomics, Mendelian Randomization, Cerebrospinal fluid metabolites, Plasma metabolites

## Abstract

**Background and Objective:** Previous studies have identified associations between metabolites and Parkinson’s disease (PD), but the causal relationships remain unclear. This study aims to identify causal relationships between specific cerebrospinal fluid (CSF) and plasma metabolites and the PD risk using Mendelian Randomization (MR).

**Methods:** We utilized data on 338 CSF metabolites from the Wisconsin Alzheimer’s Disease Research Center and the Wisconsin Registry for Alzheimer’s Prevention, and 1,400 plasma metabolites from the Canadian Longitudinal Study on Aging. PD outcome data were obtained from a GWAS meta-analysis by the International Parkinson’s Disease Genomics Consortium. MR analysis was conducted using the TwoSampleMR package in R.

**Results:** MR analysis identified 49 plasma metabolites with suggestive causal relationships with PD risk, including 21 positively associated metabolites, 23 negatively associated metabolites, and 5 unknown compounds. In the CSF, six metabolites showed suggestive causal relationships with PD, including positively associated dimethylglycine, gluconate, oxalate (ethanedioate), and the unknown metabolite X-12015, while (1-enyl-palmitoyl)-2-arachidonoyl-GPC (P-16:0/20:4) and the unknown metabolite X-23587 were negatively associated. Among the plasma metabolites, those with a positive association with PD risk include hydroxy-3-carboxy-4-methyl-5-propyl-2-furanpropanoic acid (hydroxy-CMPF), carnitine C14, 1-linoleoyl-GPG (18:2), glucose to maltose ratio, and cis-3,4-methyleneheptanoate. Conversely, metabolites with a negative association with PD risk include tryptophan, succinate to acetoacetate ratio, N,N,N-trimethyl-alanylproline betaine (TMAP), glucuronide of piperine metabolite C17H21NO3, and linoleoylcholine.

**Conclusion:** Our study underscores the correlation between CSF and plasma metabolites and PD risk, highlighting specific metabolites as potential biomarkers for diagnosis and therapeutic targets.

## Introduction

Parkinson’s disease (PD) is a progressive neurodegenerative disorder primarily characterized by the loss of dopaminergic neurons in the substantia nigra, leading to symptoms such as tremors, bradykinesia, and rigidity^1,2^. Affecting millions globally, the prevalence of PD increases with age^3^. Despite extensive research, the exact etiology of PD remains unclear, involving a complex interplay of genetic, environmental, and lifestyle factors^4,5^.

Metabolomics, the comprehensive study of small molecule metabolites within biological systems, has emerged as a crucial tool in understanding PD^6^. This approach allows for the identification of metabolic changes that occur in response to disease processes, offering potential biomarkers for early diagnosis and targets for therapeutic intervention. Recent studies have underscored significant metabolic disruptions in both cerebrospinal fluid (CSF) and plasma of PD patients, pointing to systemic metabolic dysfunction associated with the disease^7^. Analyses of plasma metabolomics have revealed alterations in various metabolic pathways, including those related to amino acids, lipids, and energy metabolism^8,9^. Similarly, CSF metabolomics studies have identified changes in neurotransmitter metabolites, oxidative stress markers, and other pathways specific to the central nervous system^10–12^. These findings indicate that metabolic biomarkers in both CSF and plasma could serve as valuable indicators for PD^13^.

Mendelian Randomization (MR) is a robust analytical method that uses genetic variants as instrumental variables (IVs) to infer causal relationships between exposures (e.g., metabolite levels) and outcomes (e.g., PD). This method addresses confounding factors and reverse causation issues common in observational studies, providing more reliable evidence for causality. Applying MR to metabolomic data in PD can identify causal metabolic pathways and potential biomarkers, advancing our understanding of PD pathogenesis and aiding in the development of targeted therapeutic strategies and more effective clinical interventions^14^.

## Patients and Methods

### Exposure data

In this study, we utilized plasma metabolite data from 8,299 participants aged 45-85 from the Canadian Longitudinal Study on Aging. The data included summary statistics for 1,091 blood metabolites and 309 metabolite ratios obtained via genome-wide association studies (GWAS)^15^. Additionally, the GWAS summary data for CSF metabolites were sourced from a study by Panyard et al., which included 338 metabolites in the CSF of 291 participants^16^. These data were derived from participants enrolled in the Wisconsin Alzheimer’s Disease Research Center, with a mean age of 64.7 years, and the Wisconsin Registry for Alzheimer’s Prevention, with a mean age of 62.0 years.

### Outcome data

The main results for PD were summarized in a recent GWAS by Nalls et al^17^. We obtained genetic information from a recent meta-analysis of GWAS conducted by the International Parkinson’s Disease Genomics Consortium and 23andMe, which included a total of 16 cohorts. The study population consisted of 33,674 individuals, including 15,056 PD patients and 18,618 control subjects. The GWAS analysis in this study took into account a range of factors including sex, various biomarkers, educational level, smoking status, and brain volume. Since the 23andMe data was not included in the publicly accessible datasets, we simply omitted this cohort from our analysis.

### Instrument selection

To ensure the effectiveness of instrumental genetic variables, we selected qualified single-nucleotide polymorphisms (SNPs) through a rigorous process. First, considering that genome-wide significance might be too stringent for CSF and plasma metabolites, we used a relatively relaxed threshold (P < 5.00E-5) to extract IVs from plasma metabolites^18,19^. Second, to ensure the independence of the IVs, we applied clumping to the SNPs based on European samples from the 1000 Genomes Project Linkage Disequilibrium reference panels, using a clumping threshold of R2 < 0.001 and a window size of 10 kb. This step aimed to minimize the potential impact of linkage disequilibrium on the stochastic allocation of alleles. Third, we harmonized the data to eliminate unclear SNPs with non-concordant alleles, ensuring consistent estimates of effect by aligning with the same alleles. The maximum threshold for minor allele frequency alignment for palindromic SNPs was set at 0.3. Fourth, we calculated the F-statistics for each SNP and excluded those with F < 10 to avoid weak instrument bias^20^. Finally, we conducted MR analysis on metabolites with more than two SNPs.

### Mendelian randomization assumptions

MR was used to evaluate the causal relationship between exposure (plasma and CSF metabolites) and disease outcome (PD). MR analysis has three core assumptions: (1) SNPs are closely related to exposure, (2) SNPs are not associated with confounding factors, and (3) SNPs have no other (pleiotropic) way of affecting the results except through exposure^21^.

### Statistical Analysis

We conducted MR analyses using the TwoSampleMR package^22^ in R version 4.4.0. For the primary analysis, the inverse variance-weighted (IVW) method was employed to evaluate MR estimates, supplemented by four additional MR methods: MR-Egger, weighted median, simple mode, and weighted mode. The MR results were expressed as odds ratios (ORs) with corresponding 95% confidence intervals (CIs). The IVW method is considered robust for estimating causal effects, provided that genetic variations comply with the three instrumental variable assumptions and are not influenced by pleiotropy.

### Sensitivity Analysis

Several sensitivity analyses were conducted to validate the robustness of the MR estimates. These included the MR-Egger intercept^22^, Cochran’s Q test, leave-one-out test^23^, and funnel plots. Cochran’s Q test was utilized to detect potential heterogeneity, with a P < 0.05 indicating its presence. The MR-Egger intercept method assessed horizontal pleiotropy. Leave-one-out sampling identified influential IV outliers, and funnel plots visualized SNP heterogeneity. Forest and scatter plots were created to display the study results. Additionally, a reverse MR analysis was performed on the positive results from the primary analysis to ensure that reverse causality did not affect the findings, thereby ensuring the reliability of the MR study.

### Metabolic Pathway Analysis

The metabolic pathway analysis was carried out using MetaboAnalyst 6.0, an online tool available at https://www.metaboanalyst.ca/. To maintain the accuracy and robustness of the results, only metabolites with a statistical significance of P < 0.05 were considered. This analysis leveraged two key databases: the Small Molecule Pathway Database (SMPDB) and the Kyoto Encyclopedia of Genes and Genomes (KEGG). A significance threshold of 0.05 was established for the pathway analysis. Functional enrichment and pathway analysis modules were applied to pinpoint relevant metabolite groups or pathways associated with the biological processes of interest.

## Results

We conducted an MR study on CSF and plasma metabolites in a European population (Figure 2). From a total of 338 CSF metabolites and 1400 plasma metabolites, we selected between 3 to 404 independent SNPs as IVs. Each of these IVs had an F value greater than 10, ensuring no weak IV bias. Detailed summary information for these SNPs is provided in Tables S1 and S2.

**Figure 1:**
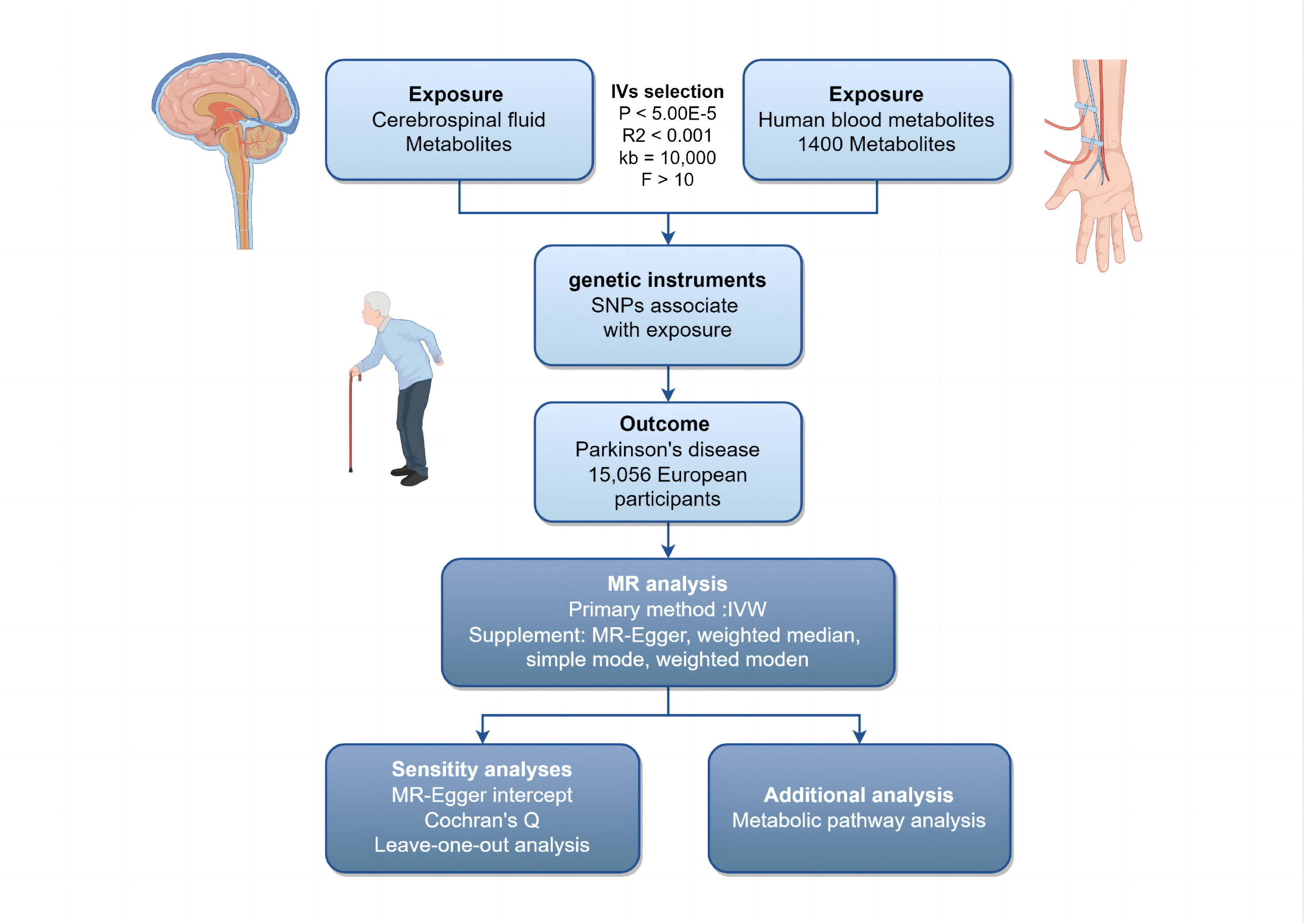
Flowchart of Mendelian Randomization Study on Metabolites and Parkinson’s Disease. IVs: Instrumental Variables; SNPs: Single Nucleotide Polymorphisms; MR: Mendelian Randomization; IVW: Inverse Variance Weighted.

**Figure 2:**
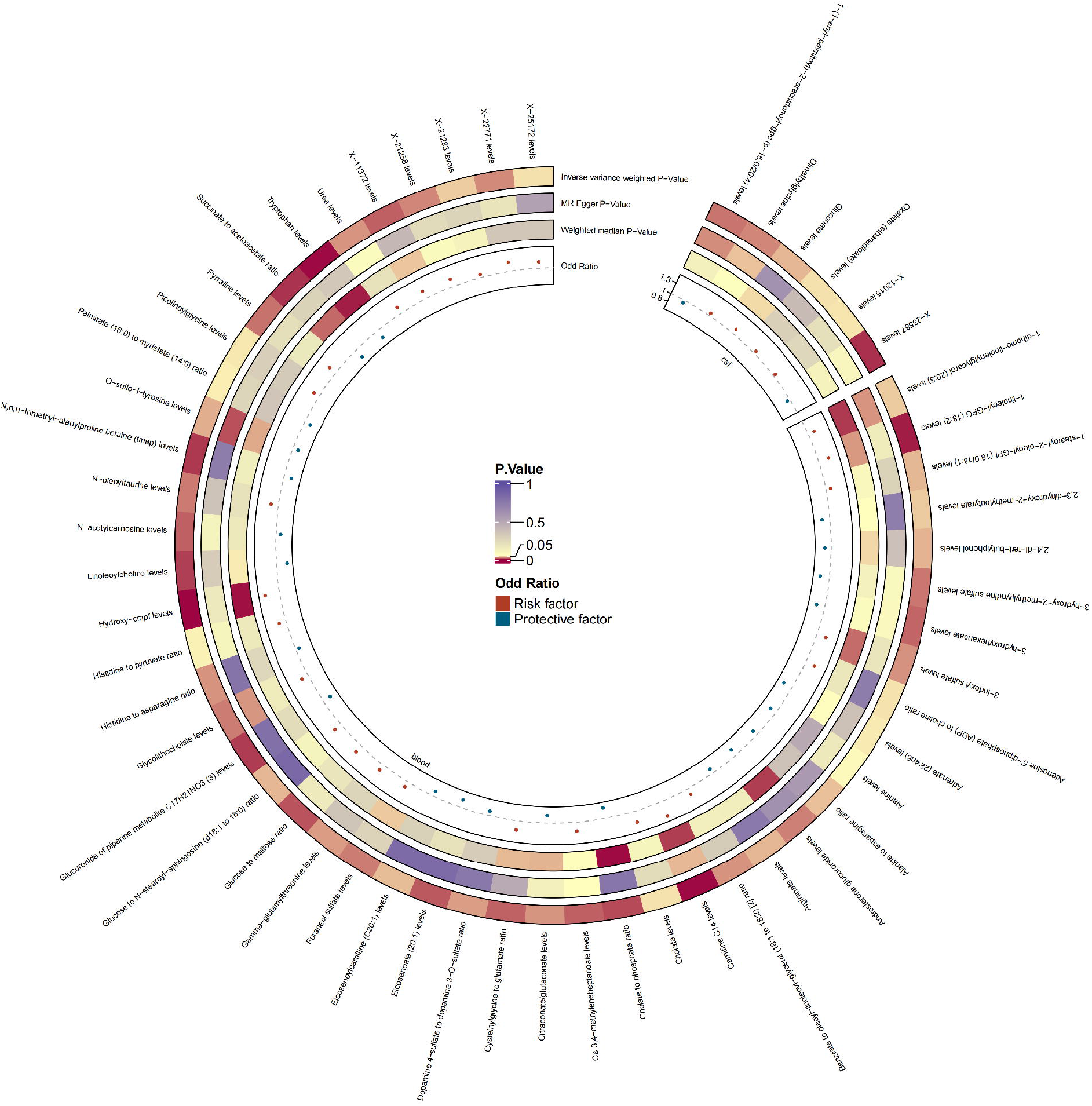
Circular Heatmap of Combined Cerebrospinal Fluid and Plasma Metabolites Associated with Parkinson’s Disease in Mendelian Randomization Analysis. The circular heatmap displays the odds ratios and p-values of cerebrospinal fluid and plasma metabolites associated with Parkinson’s Disease. Red indicates risk factors and blue indicates protective factors.

In CSF metabolites, a total of six metabolites were identified to have suggestive causal relationships with the PD risk, including two unknown metabolites (Figure 3). Among them, levels of dimethylglycine (OR [95% CI] = 1.06 [1.01-1.12], P = 0.023), gluconate (OR [95% CI] = 1.12 [1.01-1.25], P = 0.034), oxalate (ethanedioate) (OR [95% CI] = 1.07 [1.00-1.13], P = 0.043), and the unknown metabolite X-12015 (OR [95% CI] = 1.06 [1.00-1.11], P = 0.044) were positively associated with the PD risk. Conversely, levels of (1-enyl-palmitoyl)-2-arachidonoyl-gpc (p-16:0/20:4) (OR [95% CI] = 0.93 [0.88-0.99], P = 0.019) and the unknown metabolite X-23587 (OR [95% CI] = 0.93 [0.88-0.98], P = 0.005) were negatively associated with the PD risk (Figure 3).

**Figure 3:**
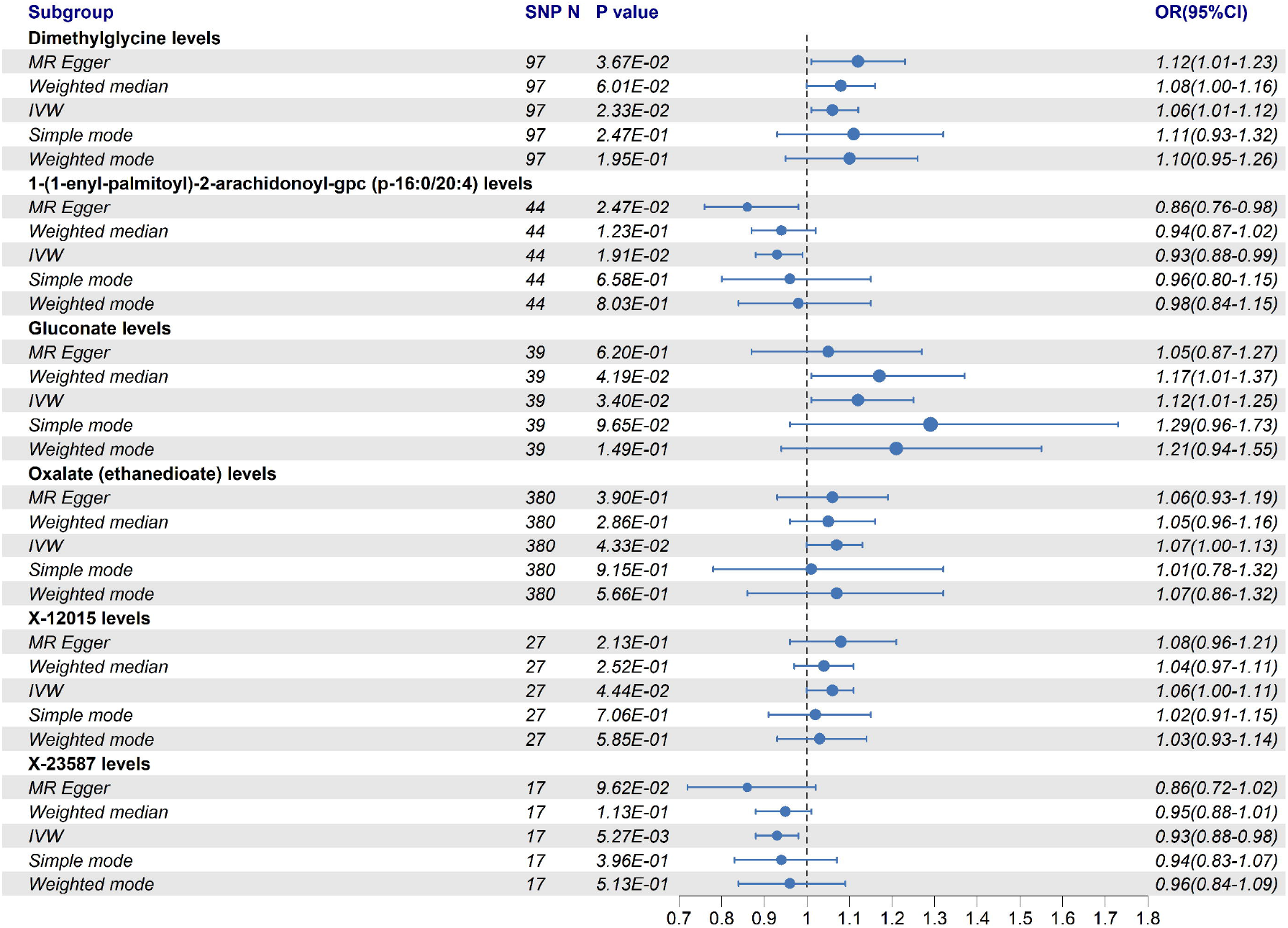
Mendelian Randomization Results of Cerebrospinal Fluid Metabolites and Parkinson’s Disease. This figure summarizes the causal relationships between cerebrospinal fluid metabolites and Parkinson’s Disease, determined by Mendelian randomization analysis. The size of each point represents the odds ratio, and horizontal lines indicate 95% confidence intervals. OR: Odds Ratio; CI: Confidence Interval; IVW: Inverse Variance Weighted.

In the MR analysis of plasma metabolites, a total of 49 metabolites were identified to have suggestive causal relationships with the PD risk (Figure 4), including five unknown metabolites. Among them, 21 metabolites were positively associated with the PD risk. The top five metabolites with the smallest p-values are hydroxy-3-carboxy-4-methyl-5-propyl-2-furanpropanoic acid (hydroxy-CMPF) (OR [95% CI] = 1.36 [1.15-1.60], P = 0.0003), carnitine C14 (OR [95% CI] = 1.38 [1.15-1.66], P = 0.0005), 1-linoleoyl-GPG (18:2) (OR [95% CI] = 1.26 [1.09-1.47], P = 0.002), glucose to maltose ratio (OR [95% CI] = 1.19 [1.04-1.36], P = 0.012), and cis-3,4-methyleneheptanoate (OR [95% CI] = 1.21 [1.04-1.42], P = 0.015). Conversely, 23 metabolites were negatively associated with the PD risk. The top five protective metabolites with the smallest p-values are tryptophan (OR [95% CI] = 0.79 [0.69-0.90], P = 0.0006), succinate to acetoacetate ratio (OR [95% CI] = 0.79 [0.66-0.93], P = 0.0055), N,N,N-trimethyl-alanylproline betaine (TMAP) (OR [95% CI] = 0.82 [0.71-0.95], P = 0.0067), glucuronide of piperine metabolite C17H21NO3 (3) (OR [95% CI] = 0.80 [0.68-0.94], P = 0.0073), and linoleoylcholine (OR [95% CI] = 0.73 [0.58-0.92], P = 0.0078).

**Figure 4:**
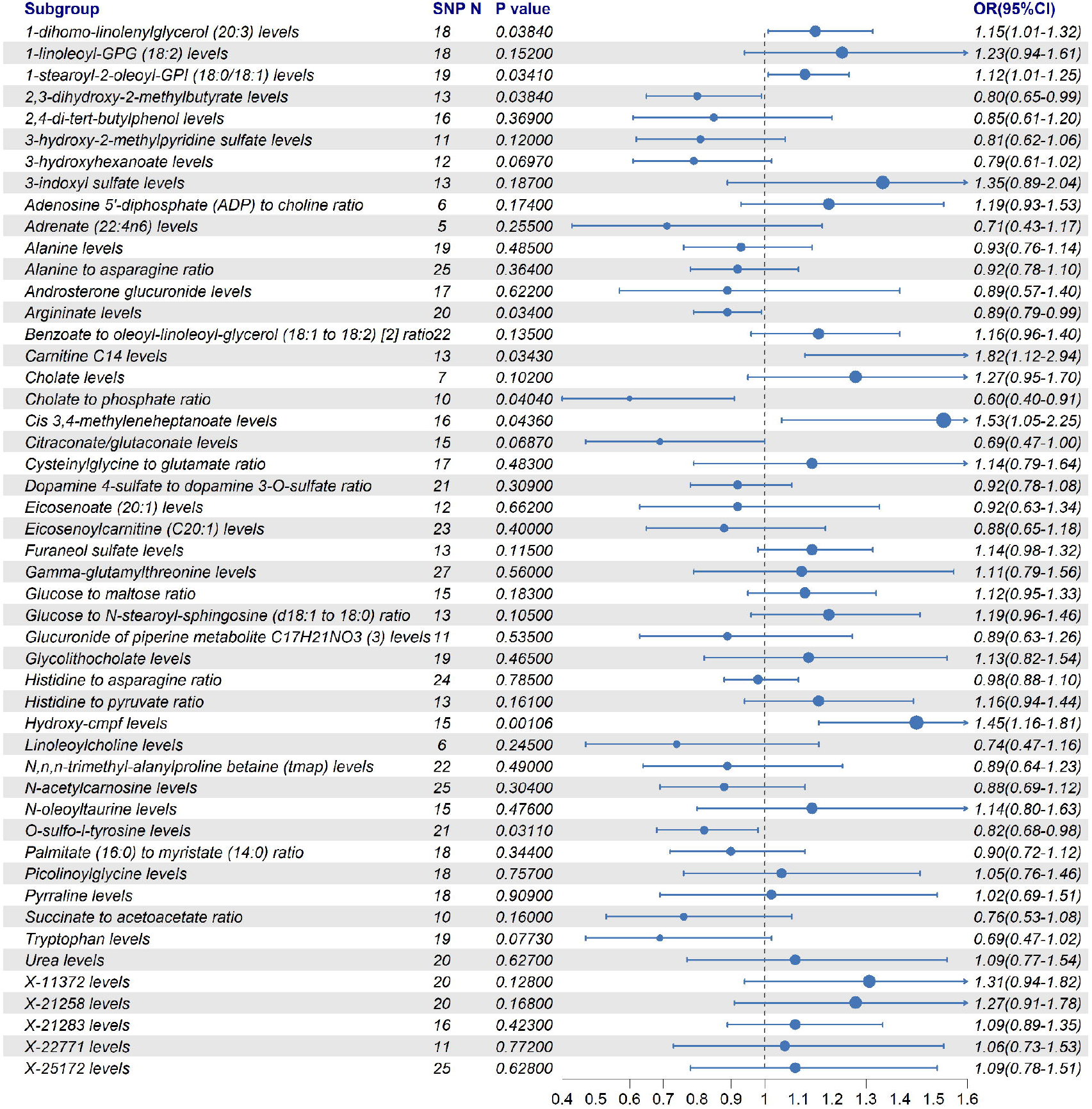
Mendelian Randomization Results of Plasma Metabolites and Parkinson’s Disease. This figure summarizes the causal relationships between plasma metabolites and Parkinson’s Disease, determined by Mendelian randomization analysis using the inverse variance weighted method. The size of each point represents the odds ratio, and horizontal lines indicate 95% confidence intervals. OR: Odds Ratio; CI: Confidence Interval.

To explore the possibility of reverse causality, we performed a reverse MR analysis to determine whether an increased PD risk could influence the levels of certain metabolites (Figure 5). In the CSF metabolites, PD risk was associated with a decrease in gluconate levels (OR [95% CI] = 0.95 [0.91-0.99], p = 0.0276). In the plasma metabolites, PD risk was associated with decreases in 1-stearoyl-2-oleoyl-GPI (18:0/18:1) levels (OR [95% CI] = 0.94 [0.89-0.99], p = 0.0123), argininate levels (OR [95% CI] = 0.94 [0.89-0.99], p = 0.0153), and 2,3-dihydroxy-2-methylbutyrate levels (OR [95% CI] = 0.95 [0.90-1.00], p = 0.0331). These findings suggest that an increased PD risk may lead to alterations in specific CSF and plasma metabolites, underscoring the impact of reverse causality on our study.

**Figure 5:**
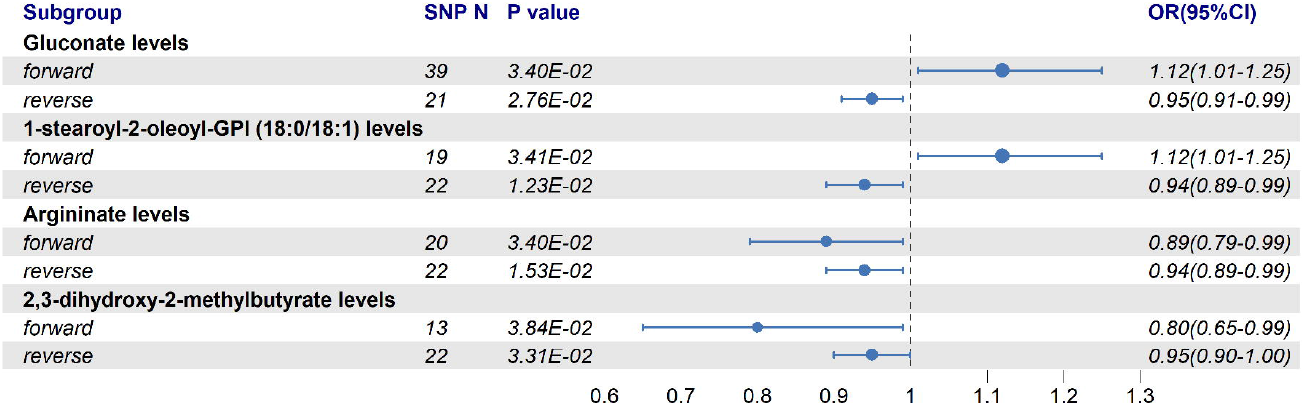
Reverse Mendelian Randomization Results of Metabolites and Parkinson’s Disease. This figure shows the reverse Mendelian randomization results of metabolites and Parkinson’s Disease, determined using the inverse variance weighted method from the positive findings of the primary analysis. Forward indicates the causal effect of metabolites on Parkinson’s Disease, and reverse indicates the causal effect of Parkinson’s Disease on metabolite levels. OR: Odds Ratio; CI: Confidence Interval.

The sensitivity analyses confirmed the robustness of the MR estimates. Despite the IVW methods being susceptible to weak IV bias, the MR-Egger, weighted median, simple mode, and weighted mode methods showed directions consistent with the MR estimates. Heterogeneity was evaluated using Cochran’s Q statistic in MR-Egger regression, and results with notable heterogeneity were excluded (Tables S3 and S4). The MR-Egger intercept was used to detect pleiotropy, and any results indicating pleiotropy were excluded (Tables S5 and S6). The leave-one-out sensitivity analysis (Figures S1-S2) and forest plots (Figures S3-S4) did not reveal any anomalous SNPs, confirming the stability of the MR estimates. Scatter plots (Figures S5-S6) and funnel plots (Figures S7-S8) further supported these findings, showing directions consistent with the IVW approach and symmetrical distribution of IVs.

### Metabolic Pathway Analysis

In this study, we utilized MetaboAnalyst 6.0 to conduct a comprehensive metabolic pathway analysis of CSF and plasma metabolites in PD patients (Table S7 and Figures S9). Our results identified 15 significant metabolic pathways, including the urea cycle (4/23, P = 0.0006; D-alanine, pyruvic acid, urea, ADP), ammonia recycling (4/25, P = 0.0009; pyruvic acid, L-asparagine, L-histidine, ADP), alanine metabolism (3/14, P = 0.0018; D-alanine, pyruvic acid, ADP), glucose-alanine cycle (2/9, P = 0.0112), glycine and serine metabolism (4/50, P = 0.0119), trehalose degradation (2/11, P = 0.0167), bile acid biosynthesis (4/59, P = 0.0211), fatty acid biosynthesis (3/33, P = 0.0216), phosphatidylethanolamine biosynthesis (2/13, P = 0.0232), riboflavin metabolism (2/14, P = 0.0267), transfer of acetyl groups into mitochondria (2/18, P = 0.0430), betaine metabolism (2/18, P = 0.0430), phosphatidylcholine biosynthesis (2/18, P = 0.0430), glutathione metabolism (2/19, P = 0.0475), and glutamate metabolism (3/45, P = 0.0487). These findings highlight the involvement of multiple metabolic pathways in PD pathogenesis, providing valuable insights into the underlying metabolic disruptions associated with this condition.

## Discussion

The findings from our MR study provide robust evidence for the metabolic underpinnings of PD. By leveraging genetic variants as instrumental variables, we inferred causal relationships between specific CSF and plasma metabolites and PD risk. Our MR analysis identified 49 metabolites with suggestive causal relationships with PD risk: 21 positively associated, 23 negatively associated, and 5 unknown compounds. In the context of CSF metabolites, we identified six with suggestive causal relationships to PD: dimethylglycine, gluconate, oxalate (ethanedioate), and the unknown metabolite X-12015 were positively associated with PD risk, whereas (1-enyl-palmitoyl)-2-arachidonoyl-GPC (P-16:0/20:4) and the unknown metabolite X-23587 were negatively associated. Notably, in the plasma metabolites, we identified hydroxy-CMPF, carnitine C14, 1-linoleoyl-GPG (18:2), glucose to maltose ratio, and cis-3,4-methyleneheptanoate as having the strongest positive associations with PD risk, while metabolites such as tryptophan, the succinate to acetoacetate ratio, N,N,N-trimethyl-alanylproline betaine (TMAP), the glucuronide of piperine metabolite C17H21NO3, and linoleoylcholine were found to be negatively associated with PD risk.

These findings highlight the multifaceted metabolic alterations in PD. Our study found that tryptophan is negatively associated with PD risk. This amino acid is not only crucial for protein synthesis but also serves as a precursor to serotonin, a neurotransmitter that regulates numerous functions in the central nervous system. In PD patients, serotonin system dysfunction is associated with non-motor symptoms such as depression and sleep disturbances^24^. Additionally, tryptophan metabolites possess antioxidative properties, reducing oxidative stress by decreasing the production of free radicals and enhancing the activity of antioxidant enzymes, thereby protecting neurons^25^. The kynurenine pathway of tryptophan metabolism also generates anti-inflammatory metabolites, which is important given the role of inflammation in PD pathology^26^.

Our study found that a lower succinate to acetoacetate ratio is associated with reduced PD risk. Previous research has shown that this ratio reflects the metabolic state of cells, particularly mitochondrial function, which is often compromised in PD. Mitochondrial dysfunction leads to disrupted energy metabolism, a hallmark of PD^27^. An increase in succinate may indicate a block in the tricarboxylic acid cycle, contributing to oxidative stress by generating reactive oxygen species. Conversely, an increase in acetoacetate suggests enhanced ketone body metabolism, which can serve as an alternative energy source when mitochondrial function is impaired, thereby reducing oxidative stress by decreasing reactive oxygen species production^28^.

Our comprehensive metabolic pathway analysis revealed that these metabolites are enriched in 15 metabolic pathways in the plasma of PD patients. The main affected pathways include the urea cycle and alanine metabolism. Research indicates that the urea cycle is significantly disrupted in PD patients, with elevated urea levels observed in various brain regions. Elevated urea levels in the brain can lead to conditions similar to uremic encephalopathy, characterized by severe cognitive impairments and neurological deficits^29^. The widespread increase in brain urea levels indicates potential metabolic disturbances across PD and underscores the critical role of effective ammonia detoxification in preventing neurotoxicity and subsequent neuronal damage in PD^30^. Furthermore, studies have shown that elevated blood urea nitrogen and proteinuria are associated with increased severity and progression of PD. For example, higher blood urea nitrogen and creatinine levels are linked to longer disease duration and worse motor symptoms^31^. In summary, these findings highlight the significant relationship between elevated urea levels and the progression and severity of Parkinson’s disease.

Additionally, alterations in alanine metabolism were identified as significant in PD patients. Elevated plasma alanine levels and increased alanine to asparagine ratios, which show a negative association with PD risk, suggest a protective role of alanine in PD. Alanine plays a crucial role in glucose metabolism and energy production, and its increased levels might support better neuronal function and energy homeostasis, potentially offering neuroprotection. Studies have shown that specific amino acid profiles, including elevated alanine levels, could serve as biochemical markers of PD progression^32^. Moreover, beta-alanine has been associated with the expression of Wnt pathway genes, further supporting its potential neuroprotective role in PD^33^.

This study represents the initial application of MR to investigate CSF and plasma metabolites in PD, effectively addressing confounding factors and issues of reverse causation commonly encountered in observational studies. This approach provides robust insights into metabolic dysregulation associated with Parkinson’s disease. However, certain limitations should be acknowledged. Firstly, our study population was primarily of European descent, which may limit the generalizability of the findings to other ethnic groups. Future research should aim to include more diverse populations to validate these results across different genetic backgrounds. Secondly, our analysis was restricted to metabolites for which genetic data were available, potentially overlooking other relevant metabolites and pathways involved in PD. Expanding the metabolomic profiling to encompass a broader range of metabolites could provide a more comprehensive understanding of the disease. Additionally, due to the limited number of SNPs, we employed a relatively relaxed significance threshold (P < 5.00E-5)^19^, which might have introduced some false positives. Furthermore, we did not perform multiple testing corrections, which is consistent with practices in similar studies^18^, but this could affect the robustness of our findings. Future studies should incorporate more stringent criteria and correction methods to enhance the reliability of the results.

In conclusion, our MR study identified 6 CSF metabolites and 49 plasma metabolites with suggestive causal relationships with PD. Among these, 1 CSF metabolite and 23 plasma metabolites were identified as protective, while 3 CSF metabolites and 21 plasma metabolites were classified as risk factors. Additionally, 2 CSF metabolites and 5 plasma metabolites were categorized as unknown compounds. Our metabolic pathway analysis revealed enrichment in 15 pathways, notably highlighting the urea cycle and alanine metabolism. These findings underscore the potential of these metabolites as biomarkers for diagnosis and therapeutic targets. Further research is needed to validate these findings and explore their clinical applications, thereby enhancing our understanding of PD pathogenesis and improving patient outcomes.

## Data Availability

The underlying code for this study is available at https://github.com/598047655/MR_metabolomics.

## Data Availability Statement

All data generated or analyzed during this study are included in this published article and its Supplementary Materials.

## Code Availability

The underlying code for this study is available at https://github.com/598047655/MR_metabolomics.

## Acknowledgments

We express our gratitude to the participants of the UK Biobank for their contribution to this research, which utilized the UK Biobank Resource. We also extend our sincere gratitude to Blauwendraat and colleagues for providing the essential data that greatly contributed to our research.

## Author Contributions

B.Z. contributed to the conception and design of the study. J.W., R.Z., Y.F., and J.C. contributed to the acquisition and analysis of data. J.W. contributed to drafting the text and preparing the figures. All authors read and approved the final manuscript.

## Standard Protocol Approvals, Registrations, and Patient Consents

Ethical review and approval were waived for this study because no individual patients were directly involved in the overall process of our study. Our study was based only on publicly available GWAS data.

## Declaration of Generative AI and AI-Assisted Technologies in the Writing Process

During the preparation of this work, the authors used ChatGPT-4 to refine and polish the language. After using this tool, the authors reviewed and edited the content as needed and take full responsibility for the content of the publication.

## Supplementary File legends

Supplementary Figure 1: Leave-One-Out Sensitivity Analysis Results for Cerebrospinal Fluid Metabolites in MR Study of Parkinson’s Disease

Supplementary Figure 2: Leave-One-Out Sensitivity Analysis Results for Plasma Metabolites in MR Study of Parkinson’s Disease

Supplementary Figure 3: Forest Plots of Cerebrospinal Fluid Metabolites in MR Study of Parkinson’s Disease

Supplementary Figure 4: Forest Plots of Plasma Metabolites in MR Study of Parkinson’s Disease

Supplementary Figure 5: Scatter Plots of Cerebrospinal Fluid Metabolites in MR Study of Parkinson’s Disease

Supplementary Figure 6: Scatter Plots of Plasma Metabolites in MR Study of Parkinson’s Disease

Supplementary Figure 7: Funnel Plots of Cerebrospinal Fluid Metabolites in MR Study of Parkinson’s Disease

Supplementary Figure 8: Funnel Plots of Plasma Metabolites in MR Study of Parkinson’s Disease

Supplementary Figure 9: Metabolic Pathway Analysis of Metabolites Associated with Parkinson’s Disease. This bubble plot shows the pathway impact score on the x-axis and the -log10 (p-value) on the y-axis. Larger bubbles indicate higher impact, and the color gradient from yellow to red represents increasing significance, with redder bubbles indicating more statistically significant pathways.

Supplementary Table 1: Summary of SNPs for Cerebrospinal Fluid Metabolites in MR Study of Parkinson’s Disease

Supplementary Table 2: Summary of SNPs for Plasma Metabolites in MR Study of Parkinson’s Disease

Supplementary Table 3: Heterogeneity Analysis Using Cochran’s Q statistic for Cerebrospinal Fluid Metabolites in MR Study of Parkinson’s Disease

Supplementary Table 4: Heterogeneity Analysis Using Cochran’s Q statistic for Plasma Metabolites in MR Study of Parkinson’s Disease

Supplementary Table 5: Pleiotropy Analysis Using MR-Egger Intercept for Cerebrospinal Fluid Metabolites in MR Study of Parkinson’s Disease

Supplementary Table 6: Pleiotropy Analysis Using MR-Egger Intercept for Plasma Metabolites in MR Study of Parkinson’s Disease

Supplementary Table 7: Metabolic Pathway Analysis of Metabolites Associated with Parkinson’s Disease

