## Supplementary figures and images for "Cerebrospinal Fluid and Plasma Metabolites with Parkinson’s Disease: A Mendelian Randomization Study"

### Supplemental Figure 1

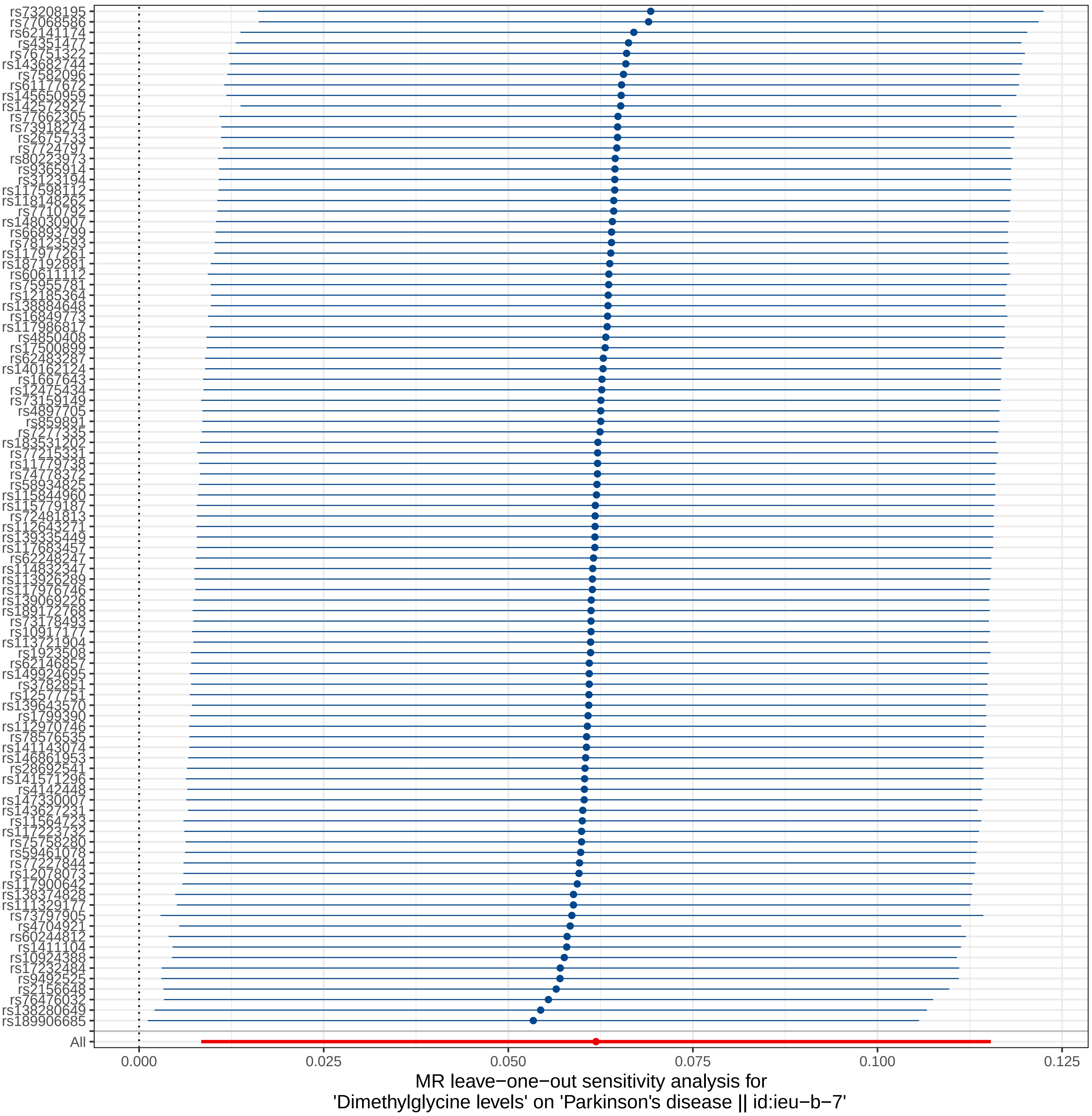

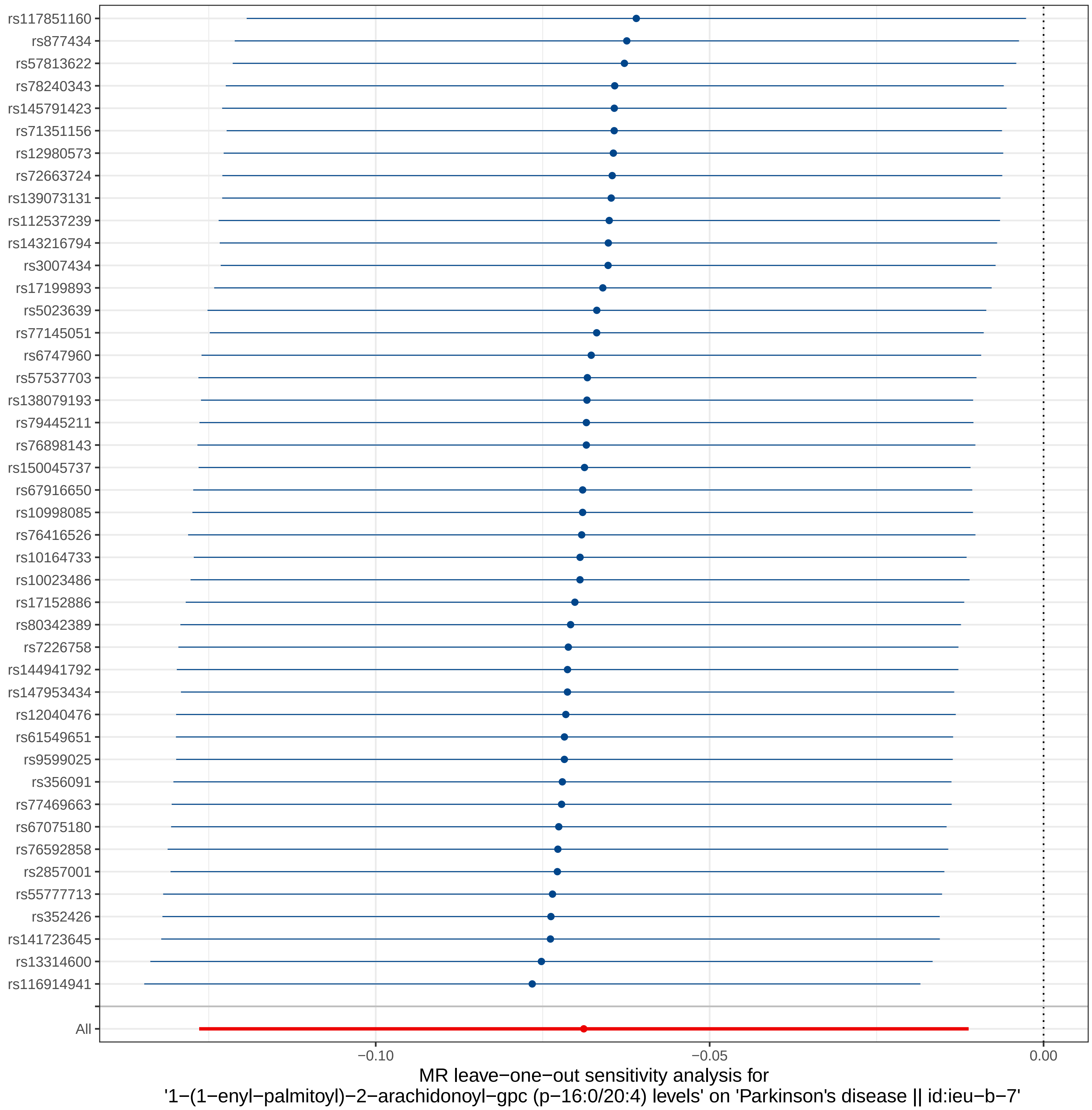

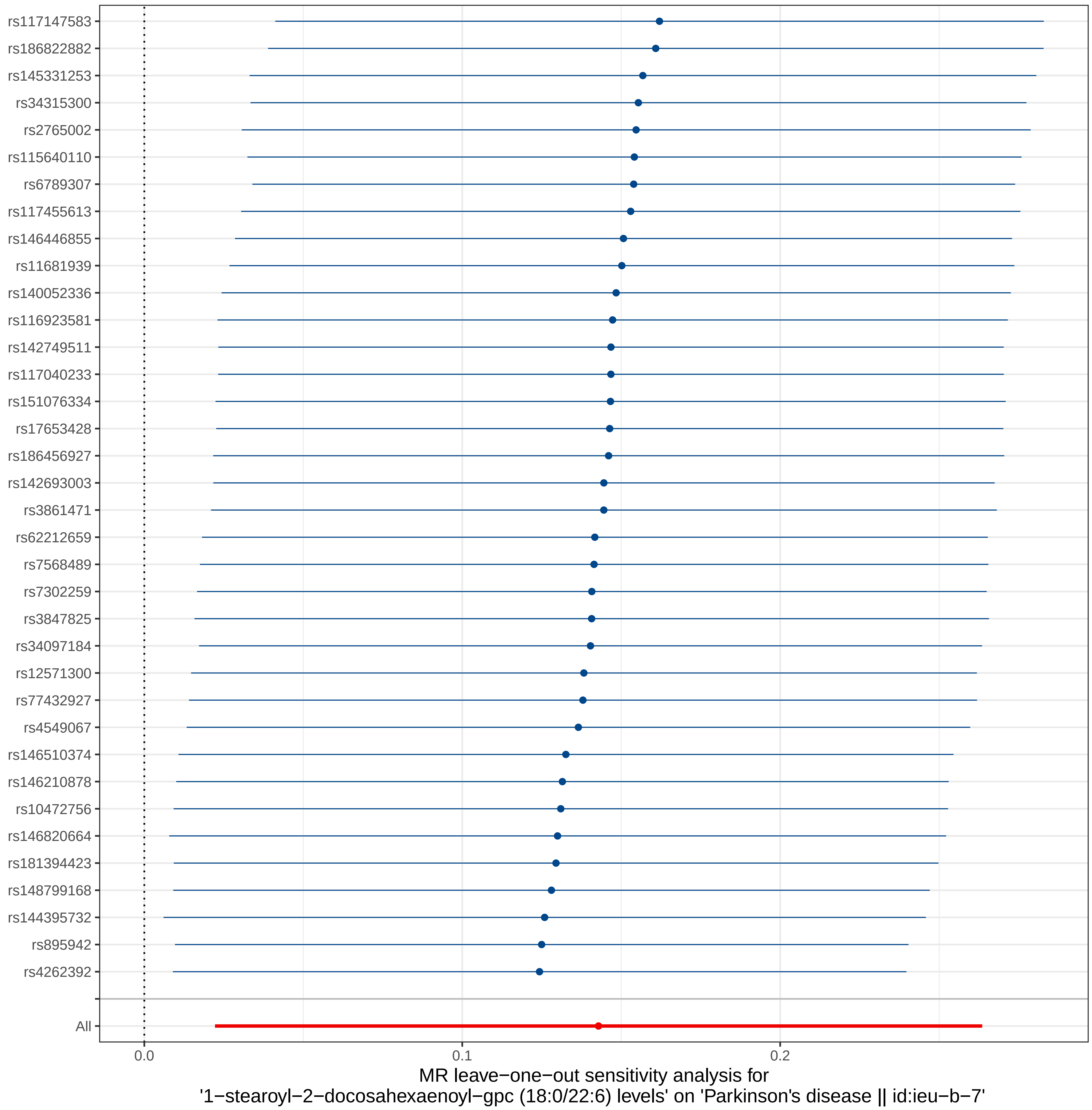

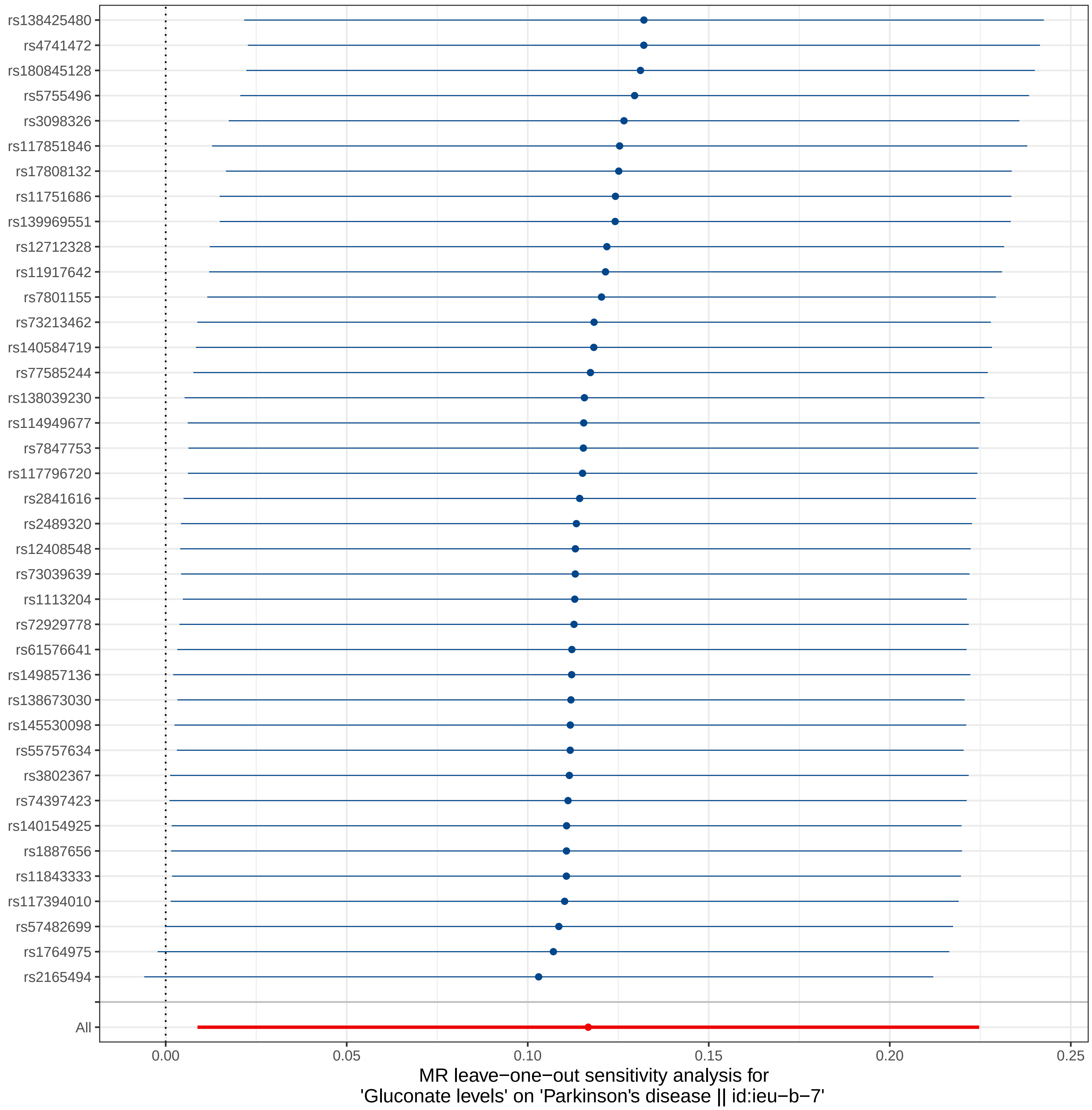

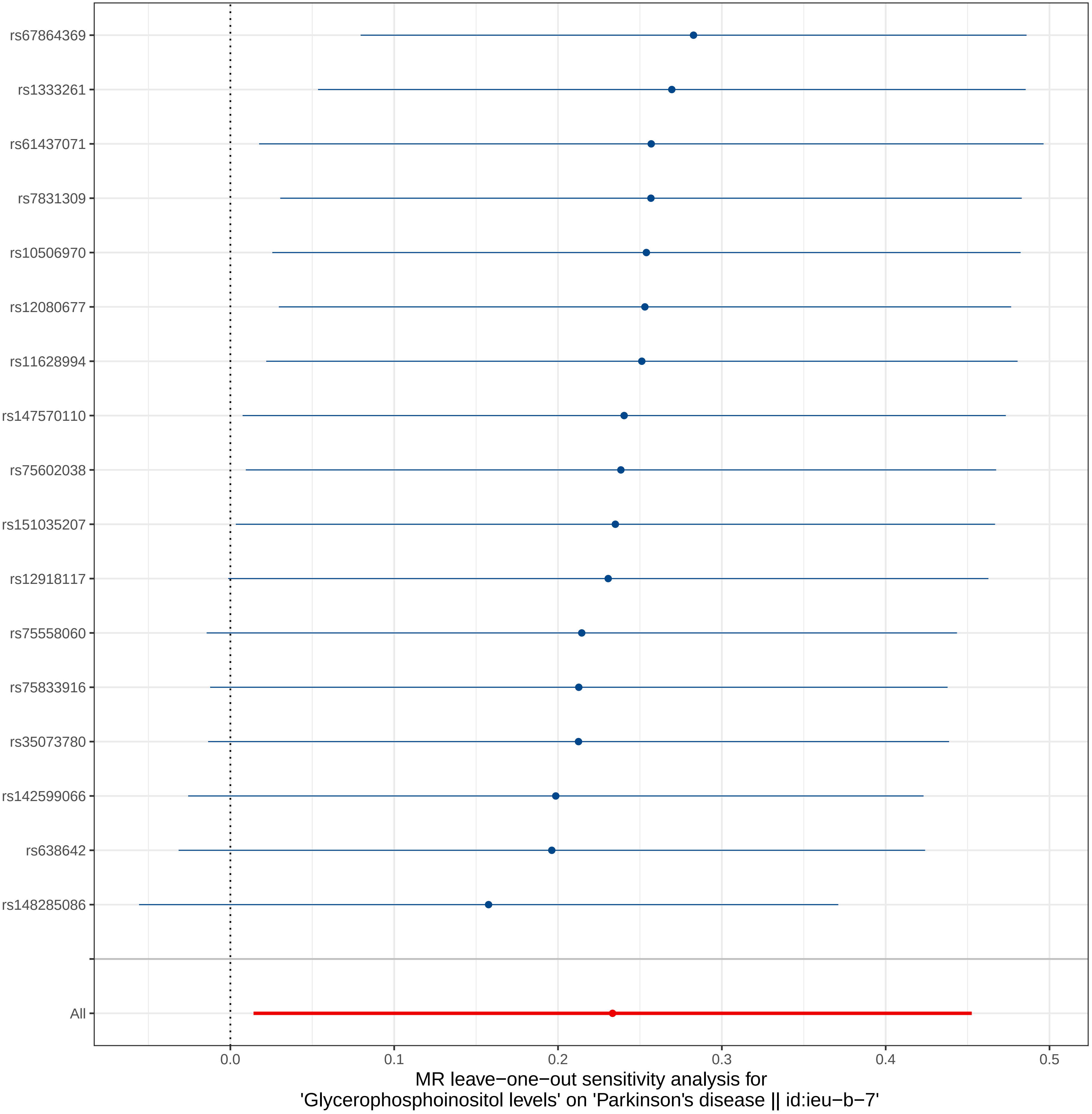

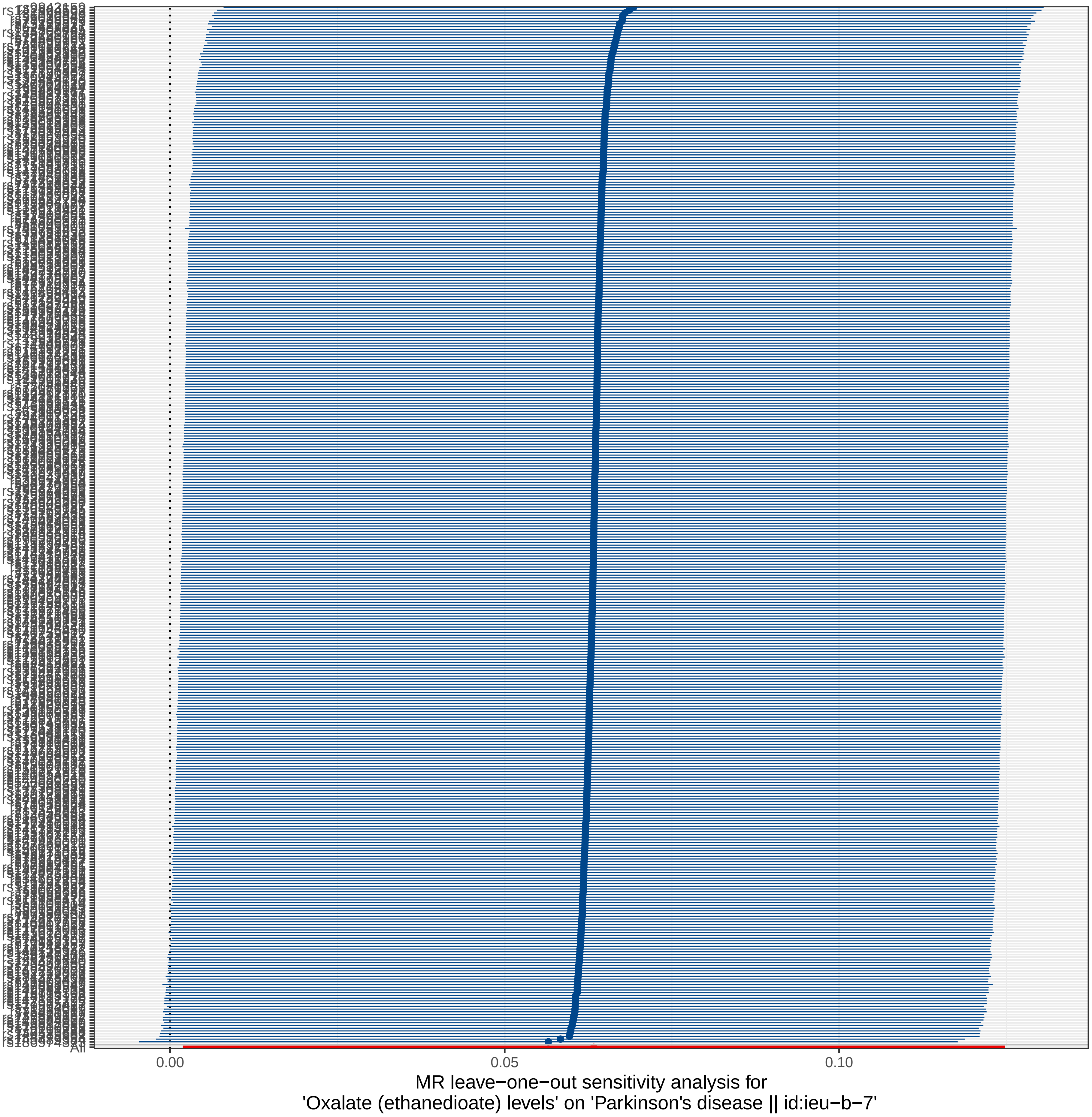

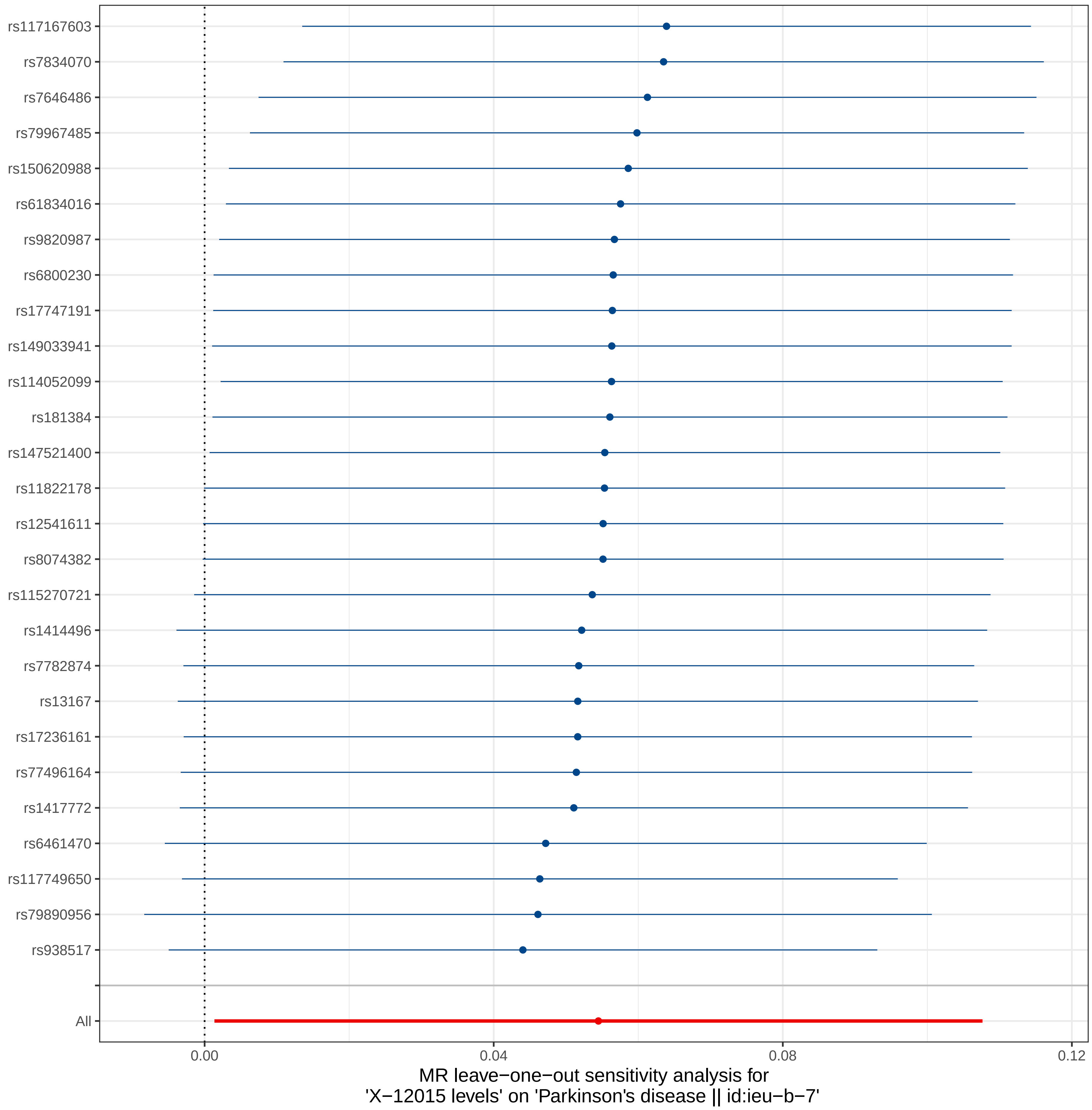

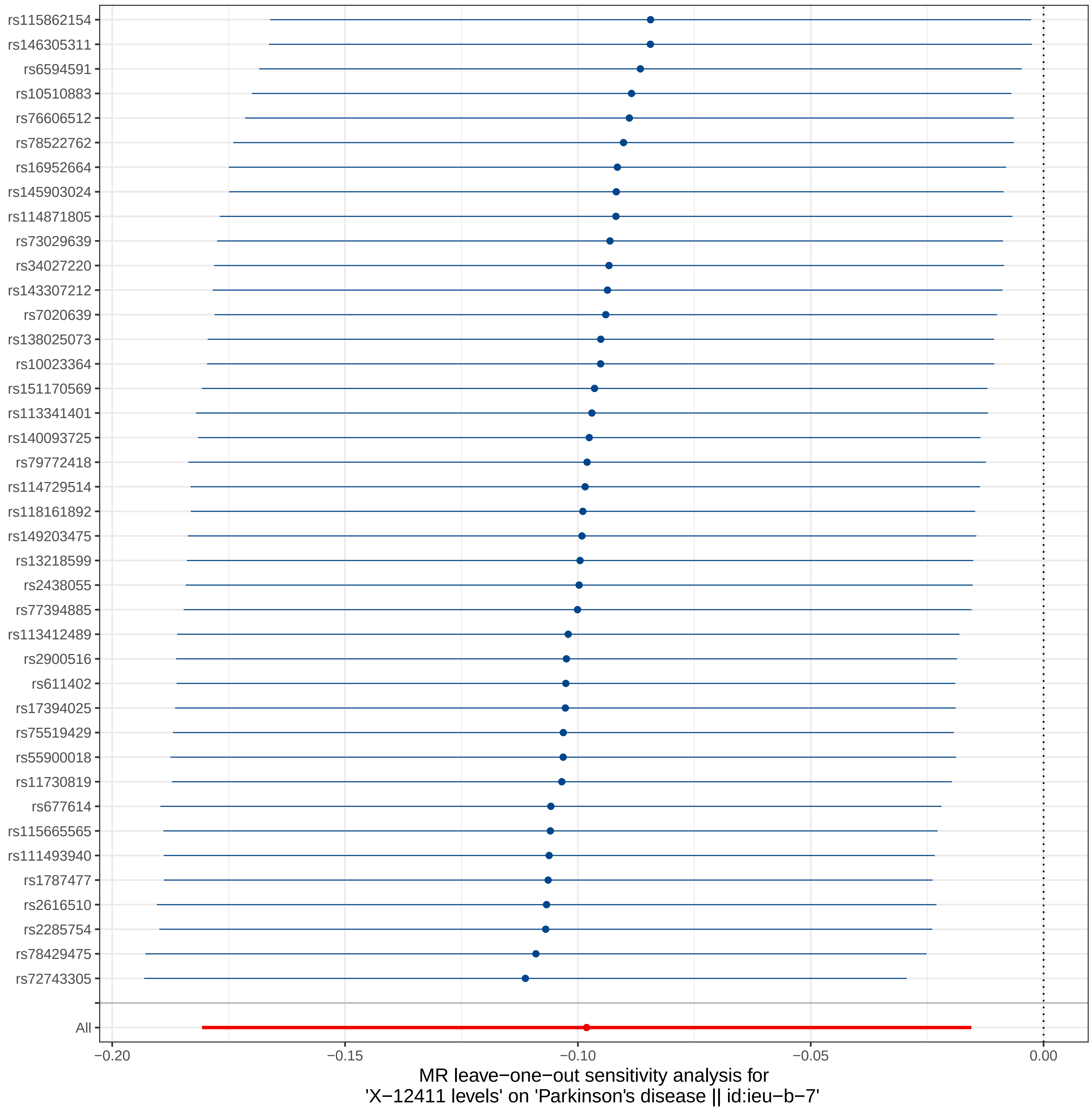

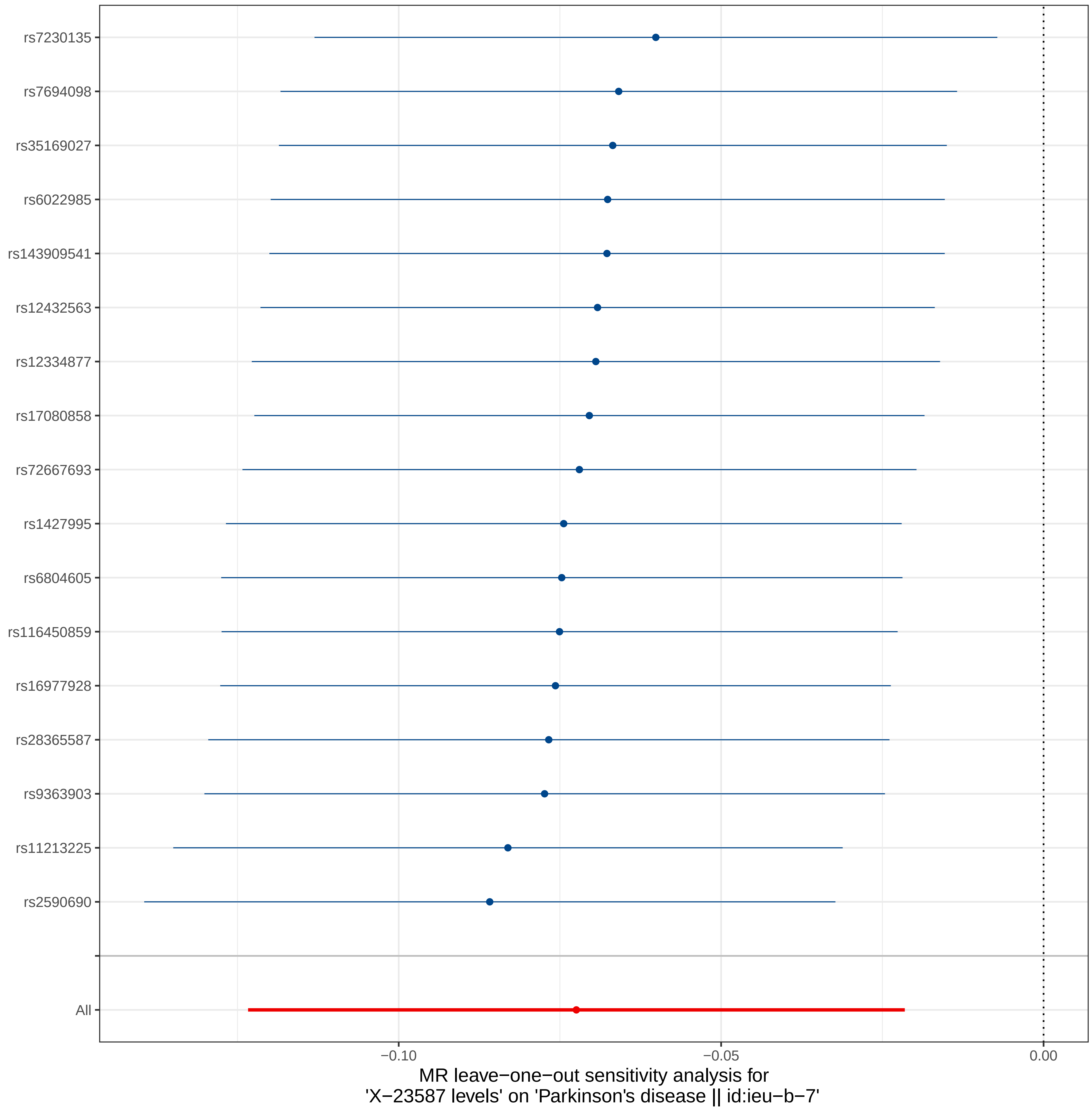

### Supplemental Figure 3

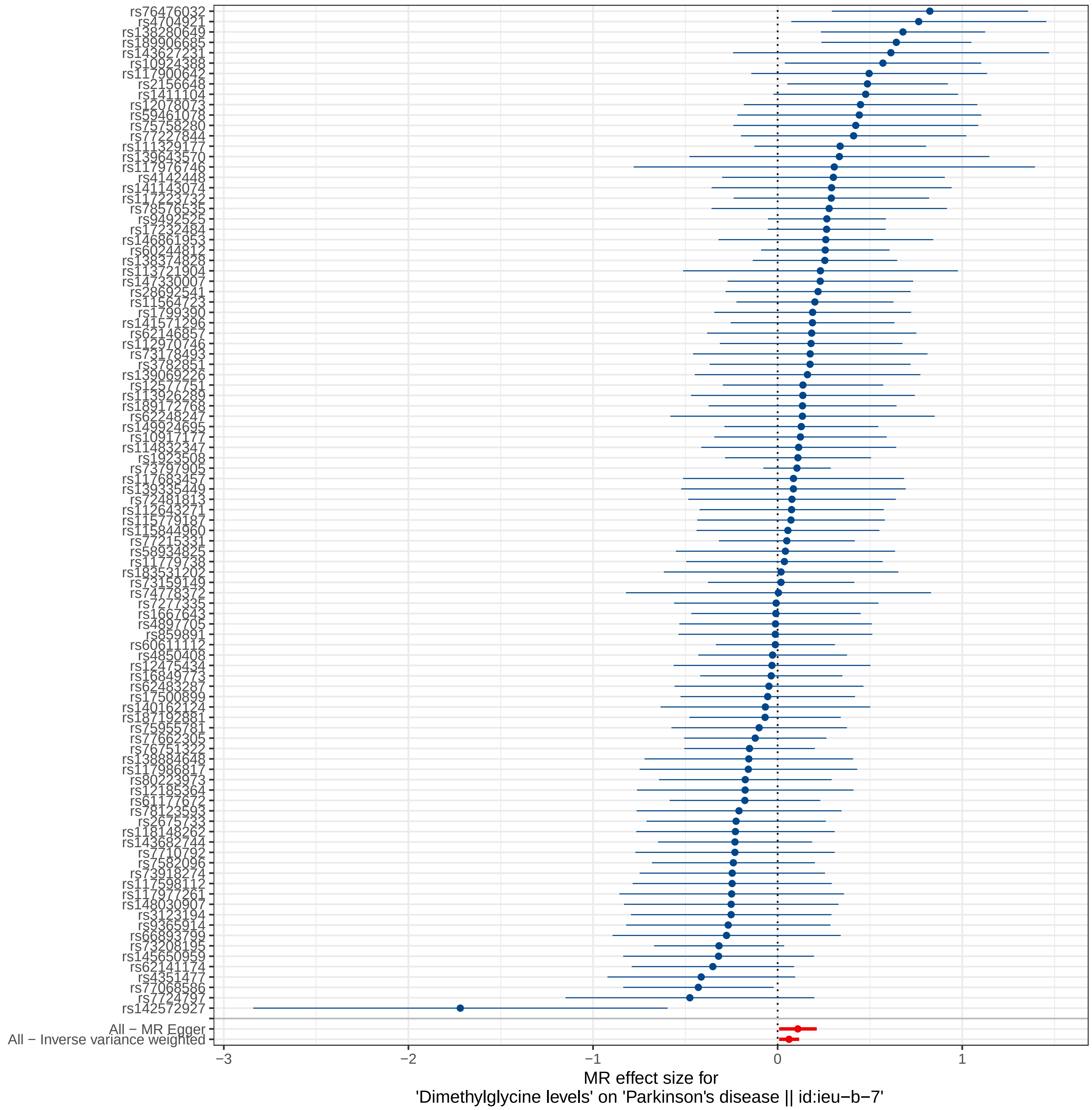

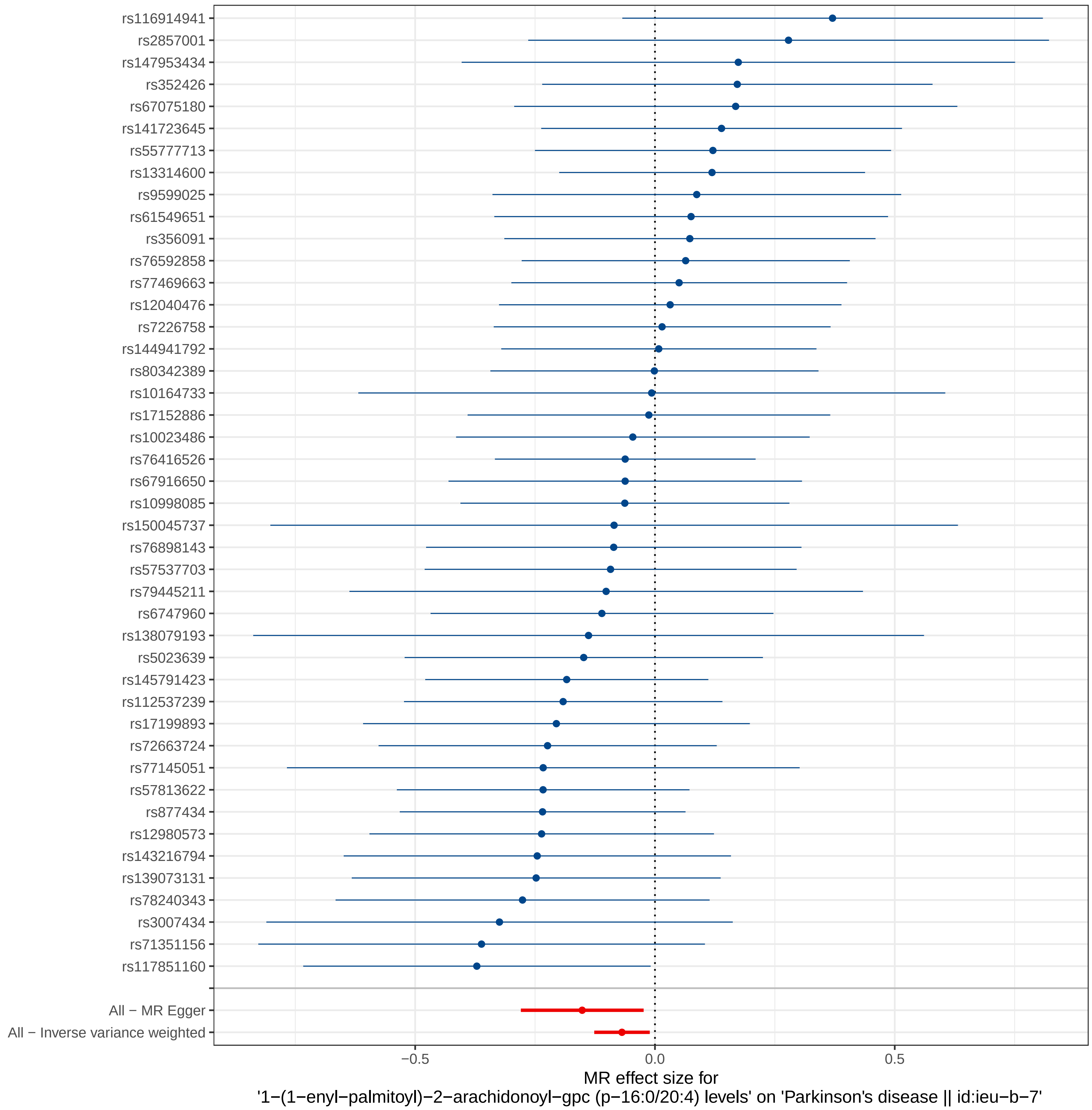

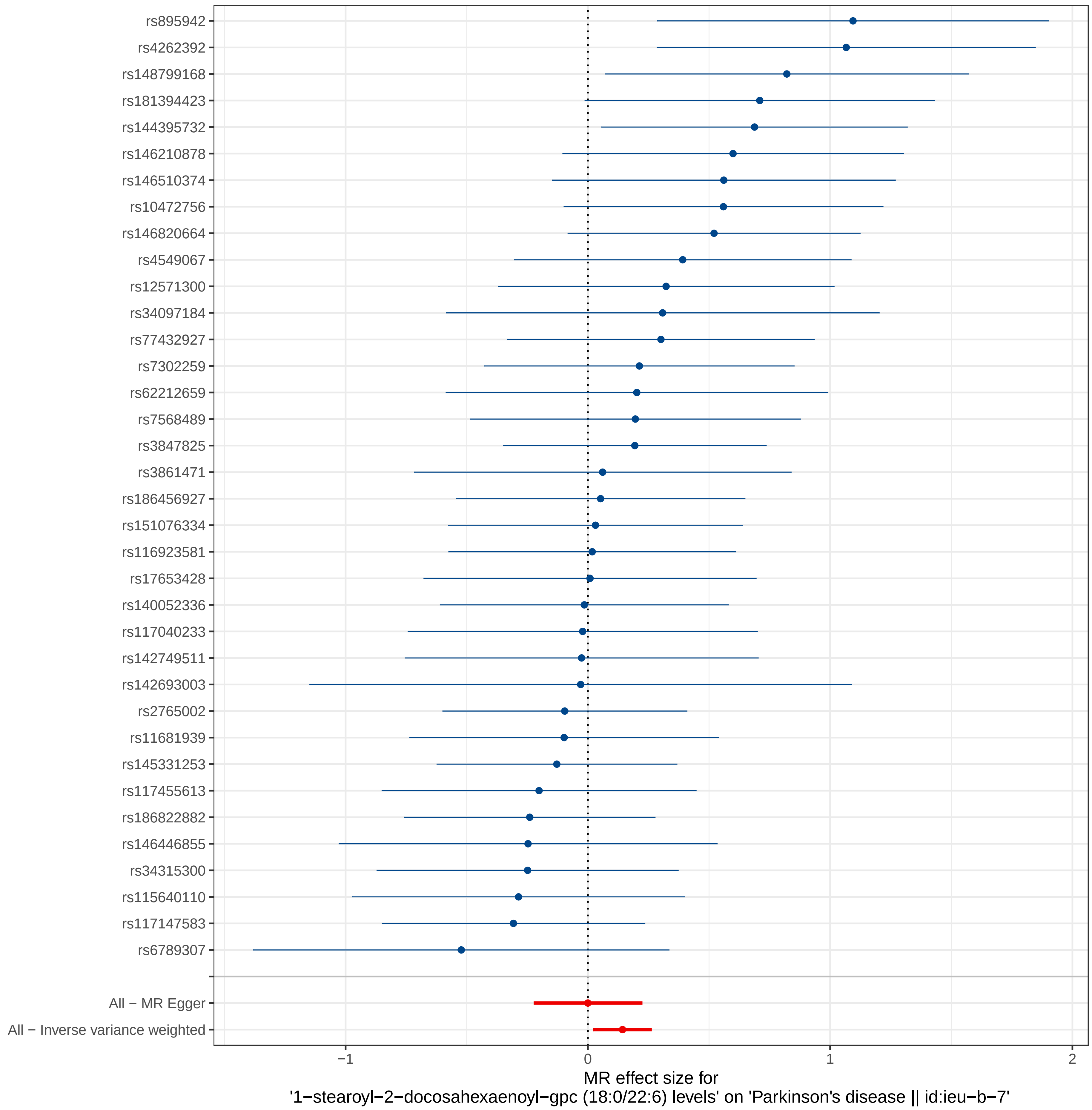

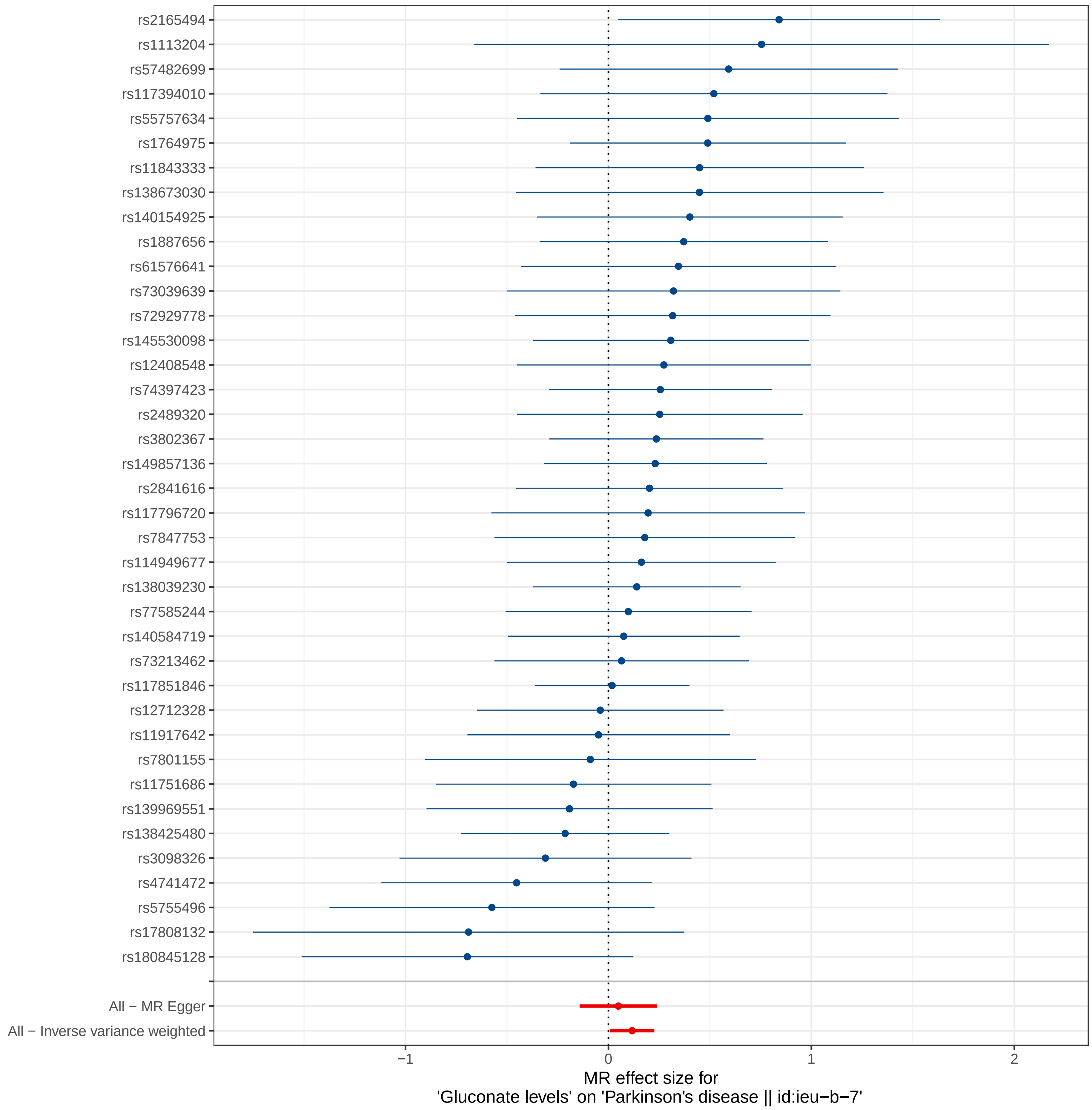

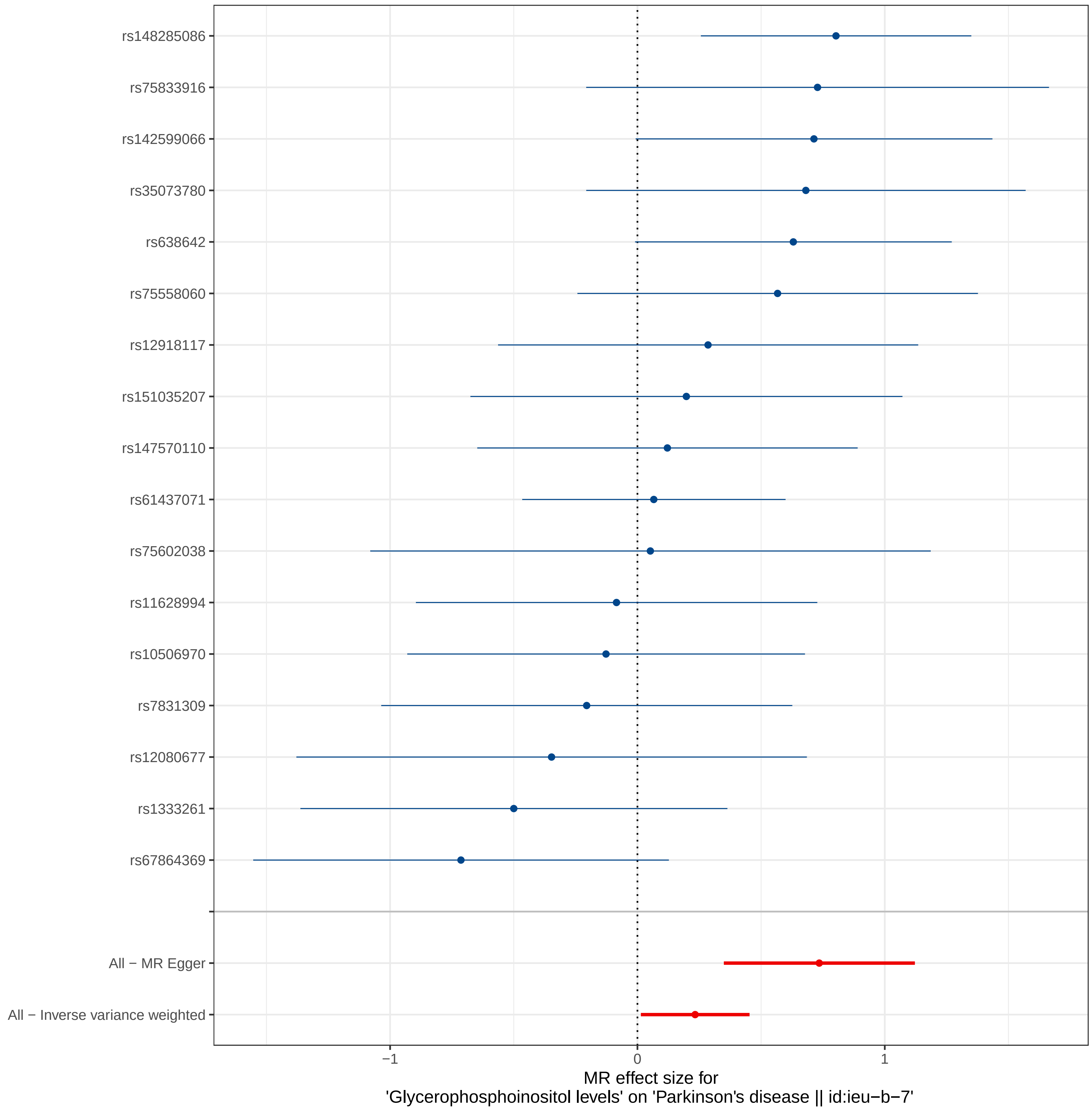

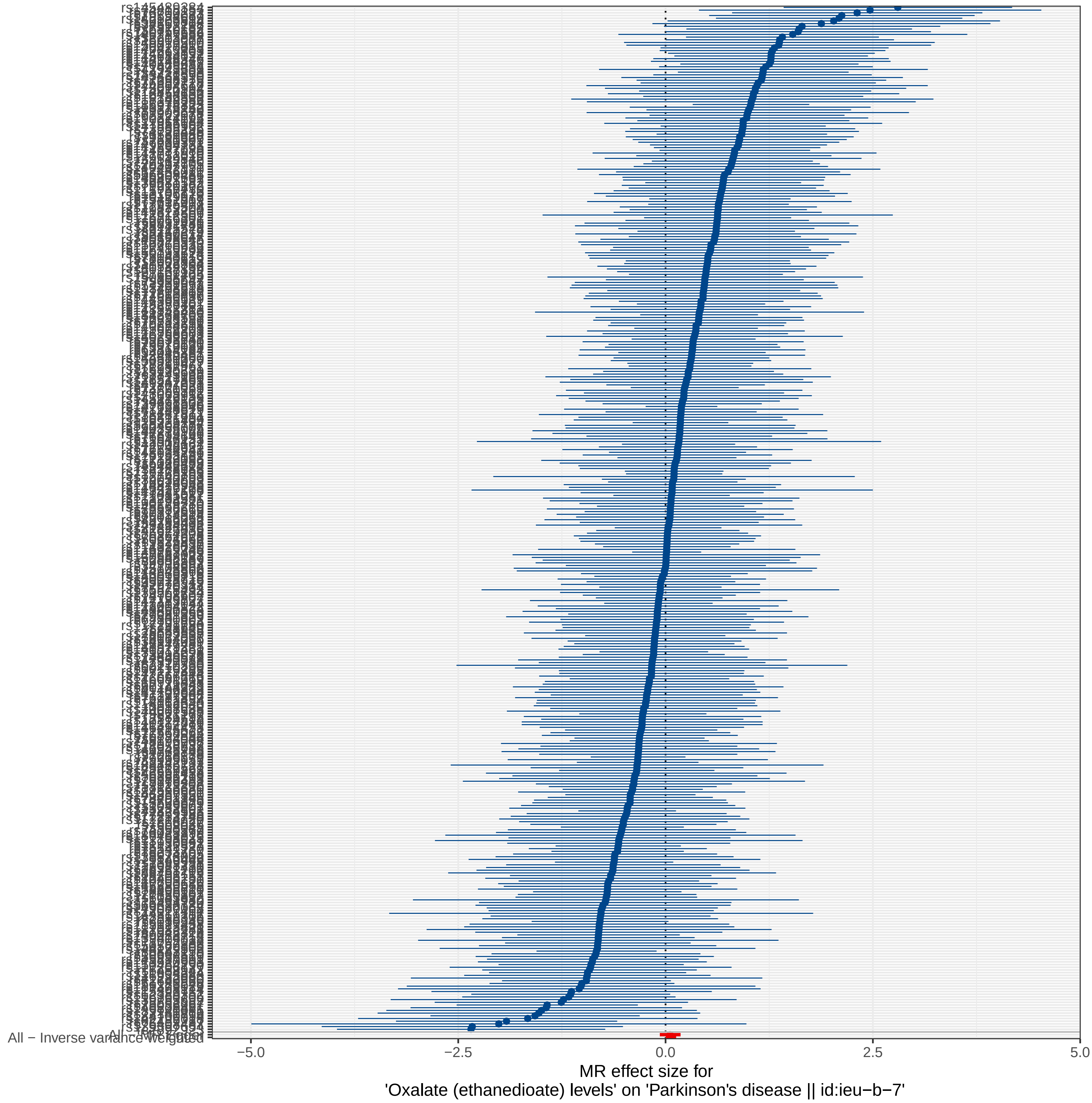

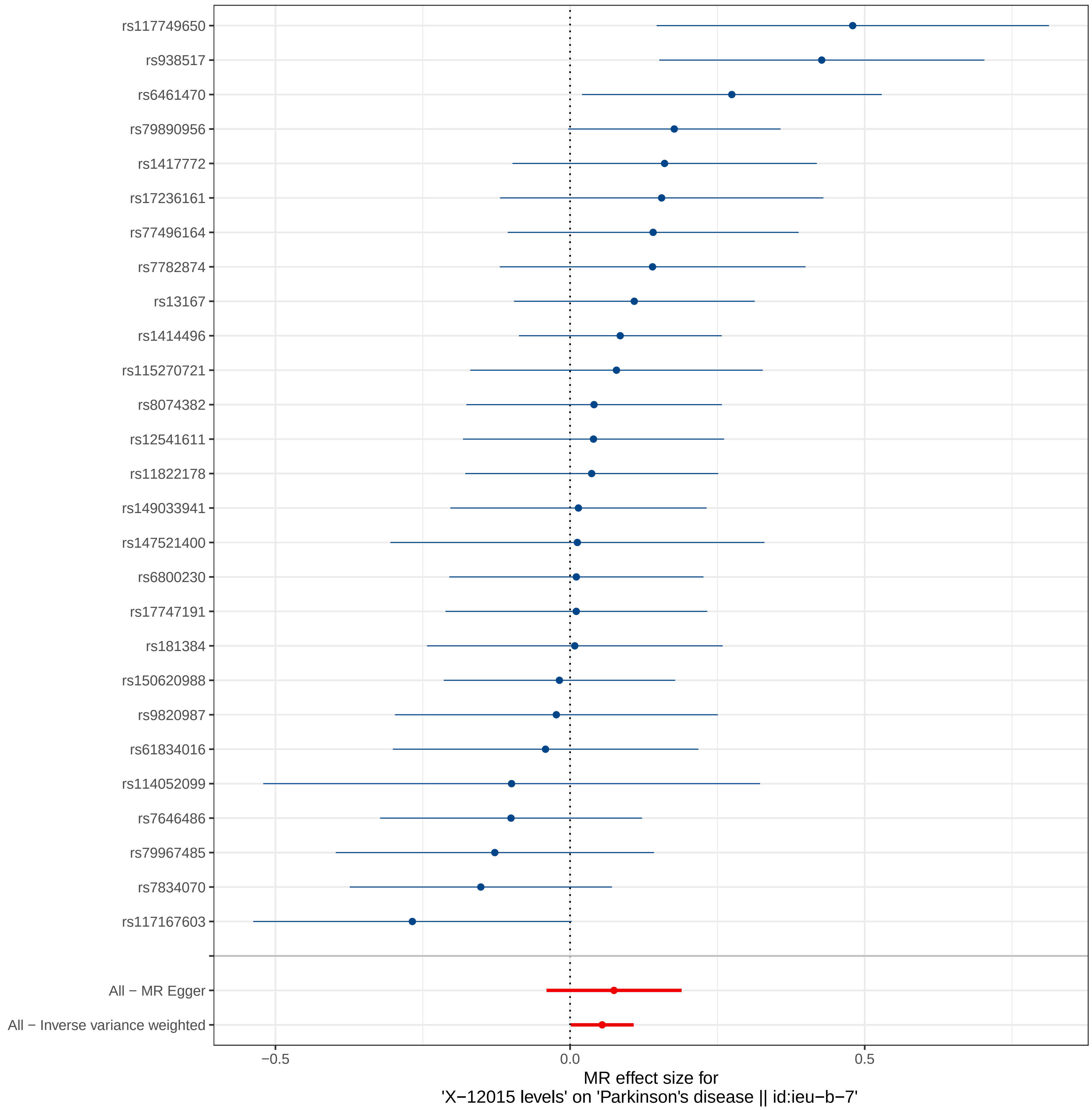

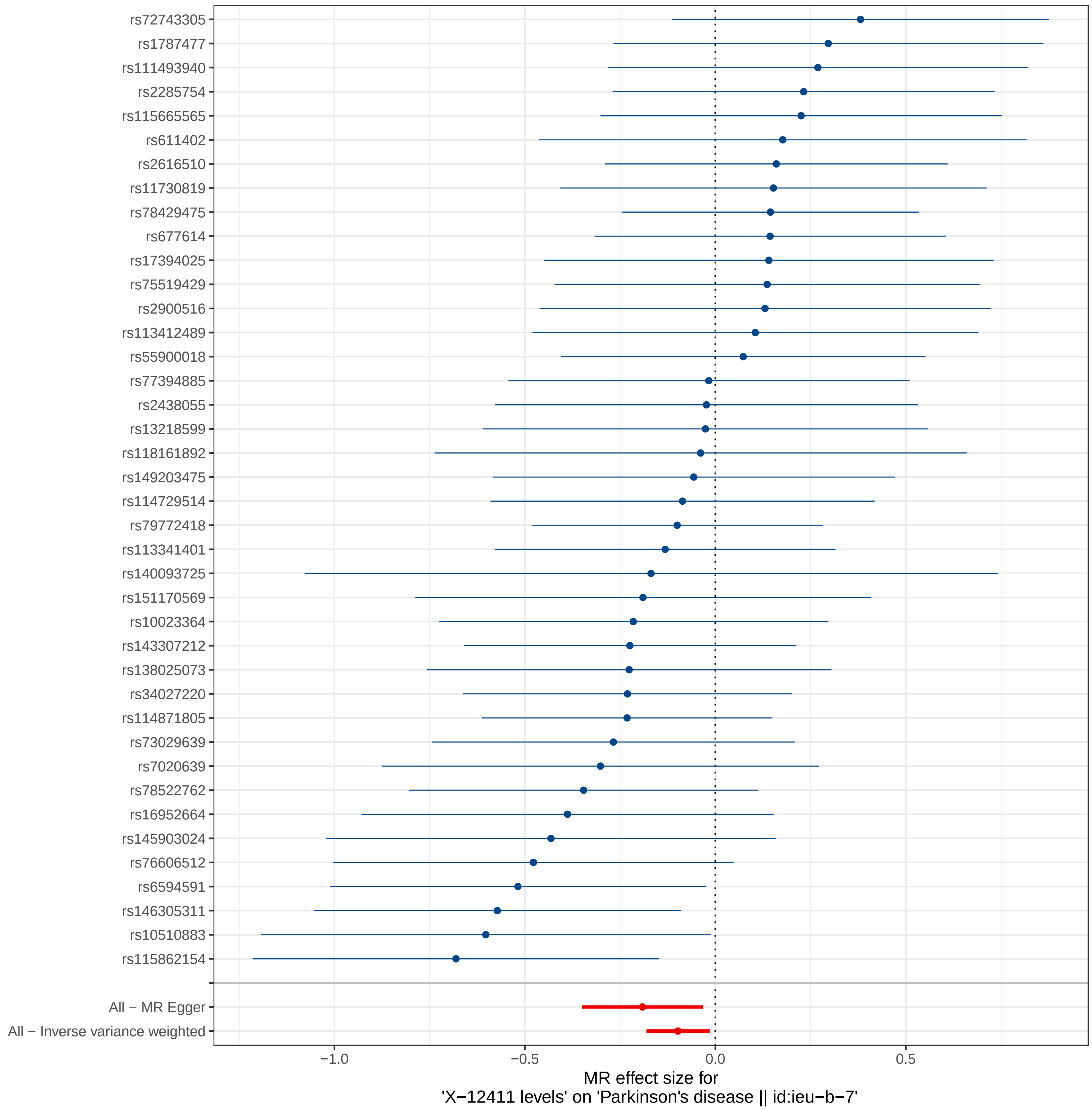

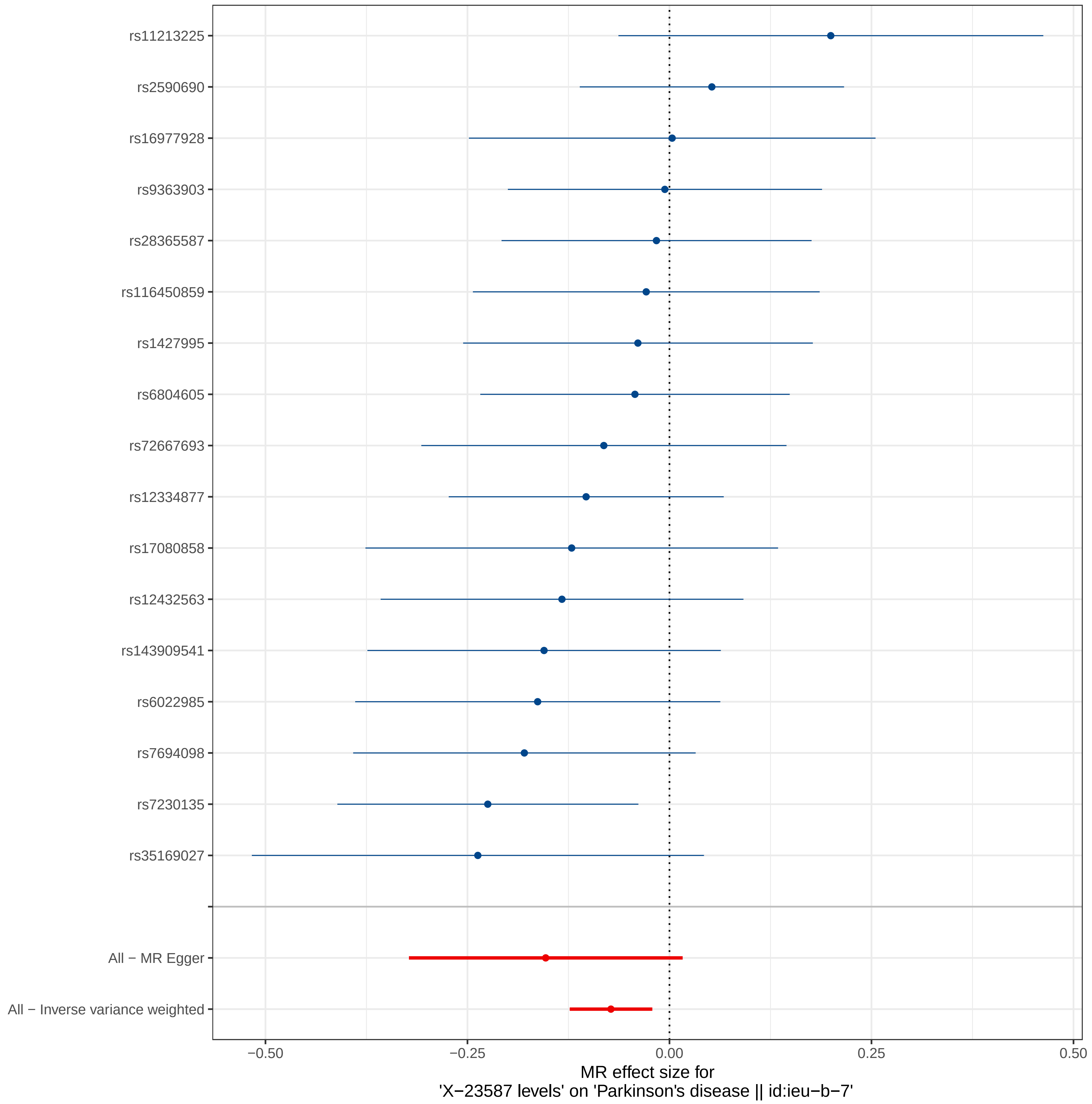

### Supplemental Figure 5

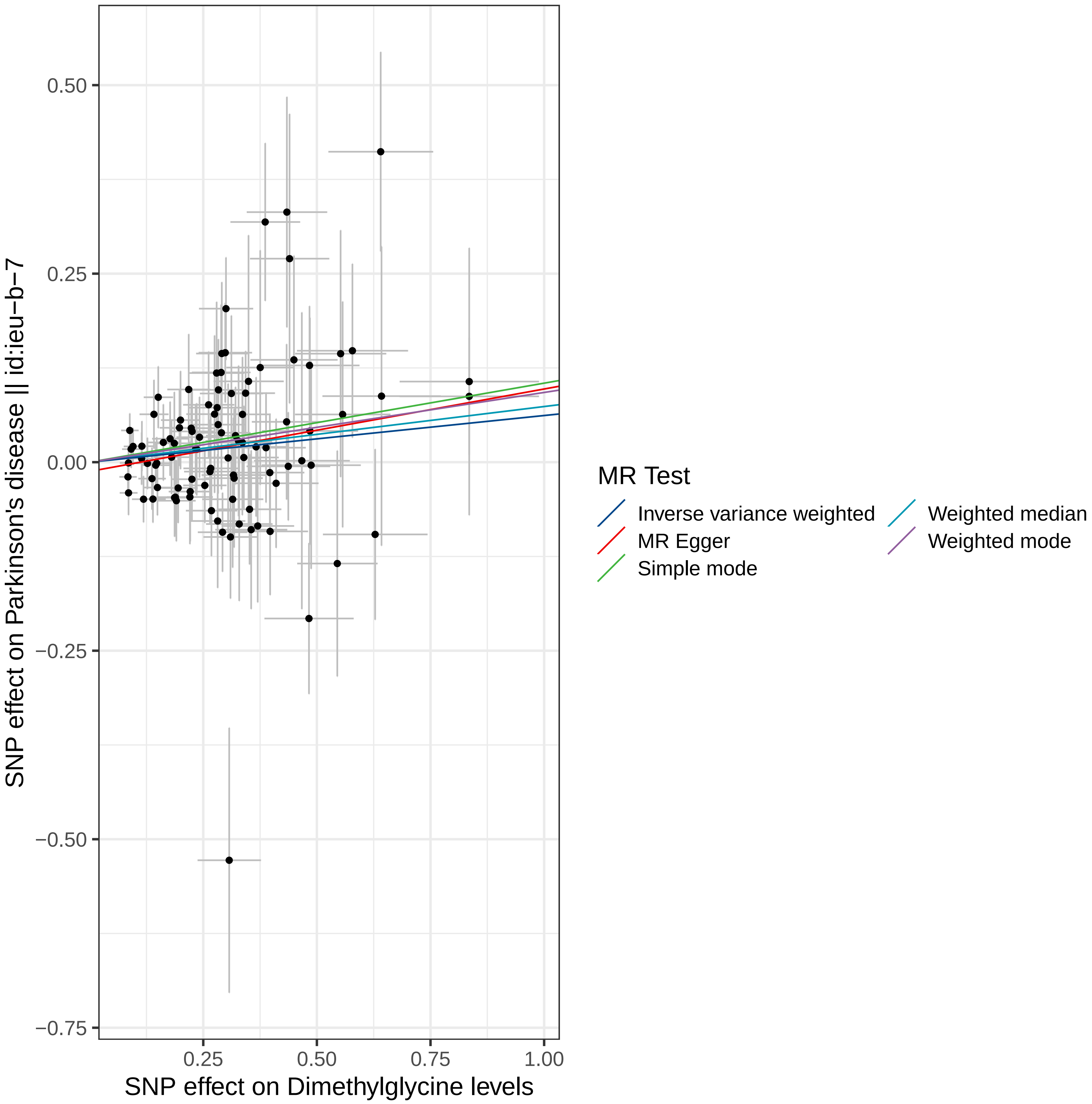

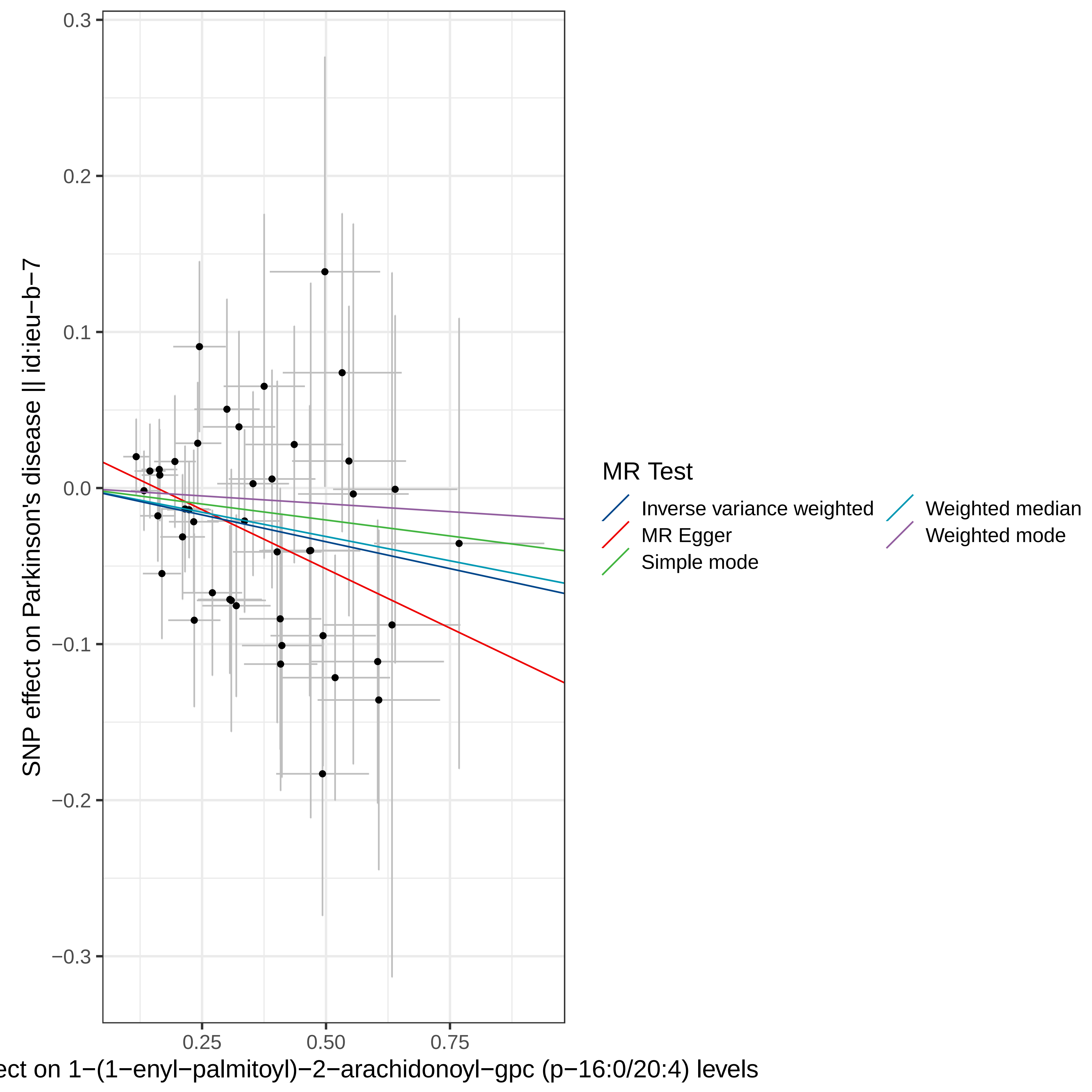

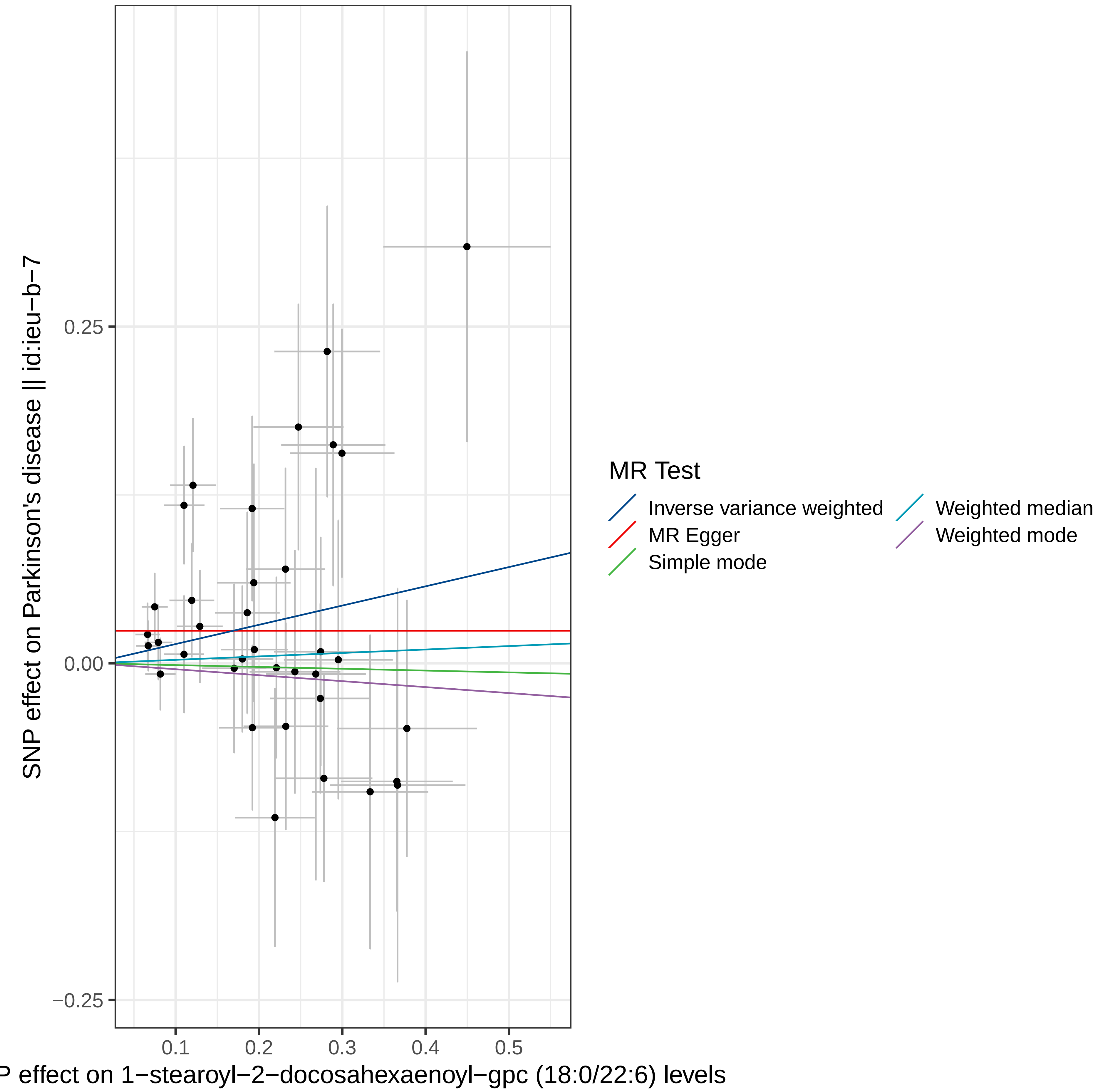

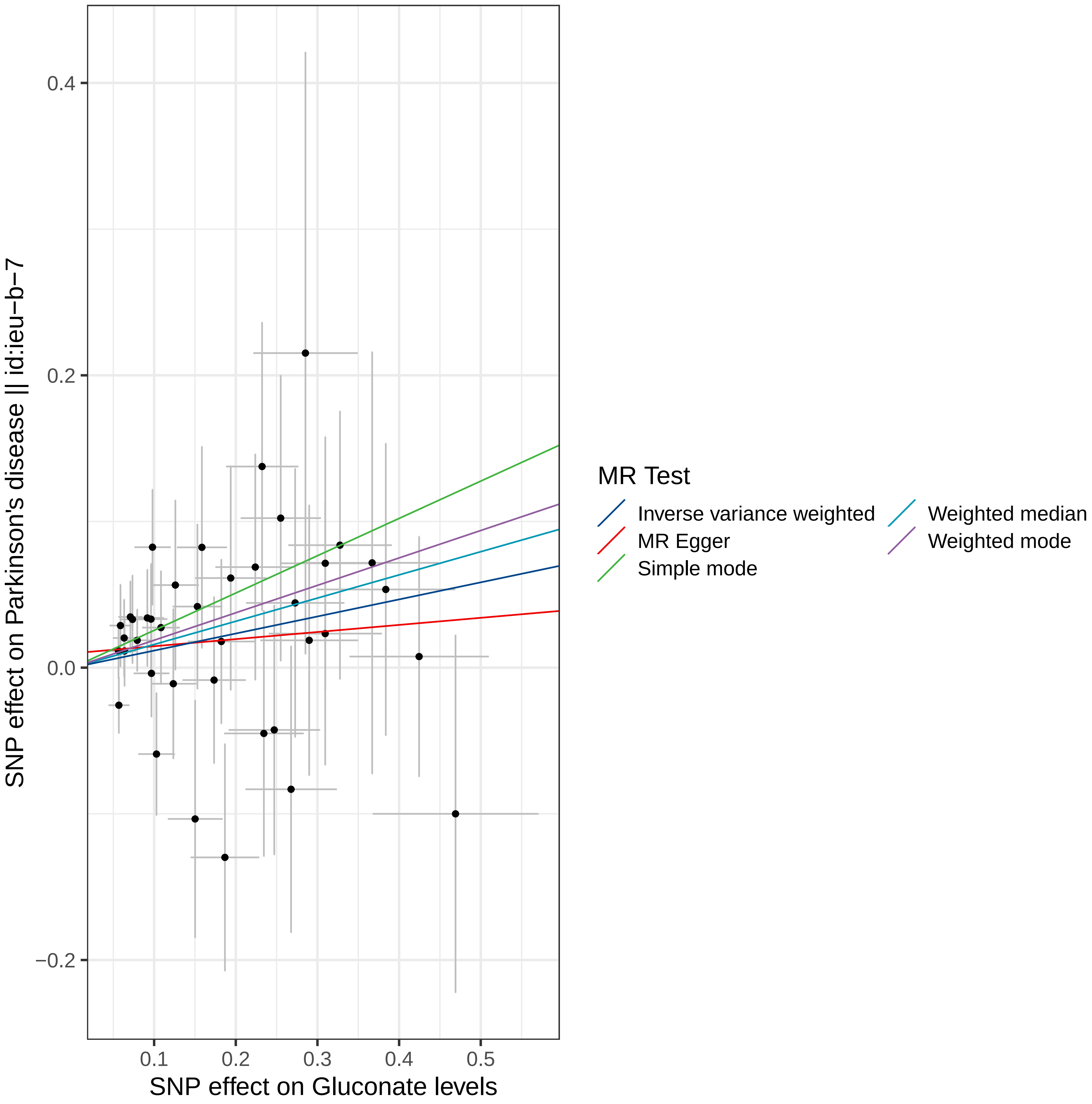

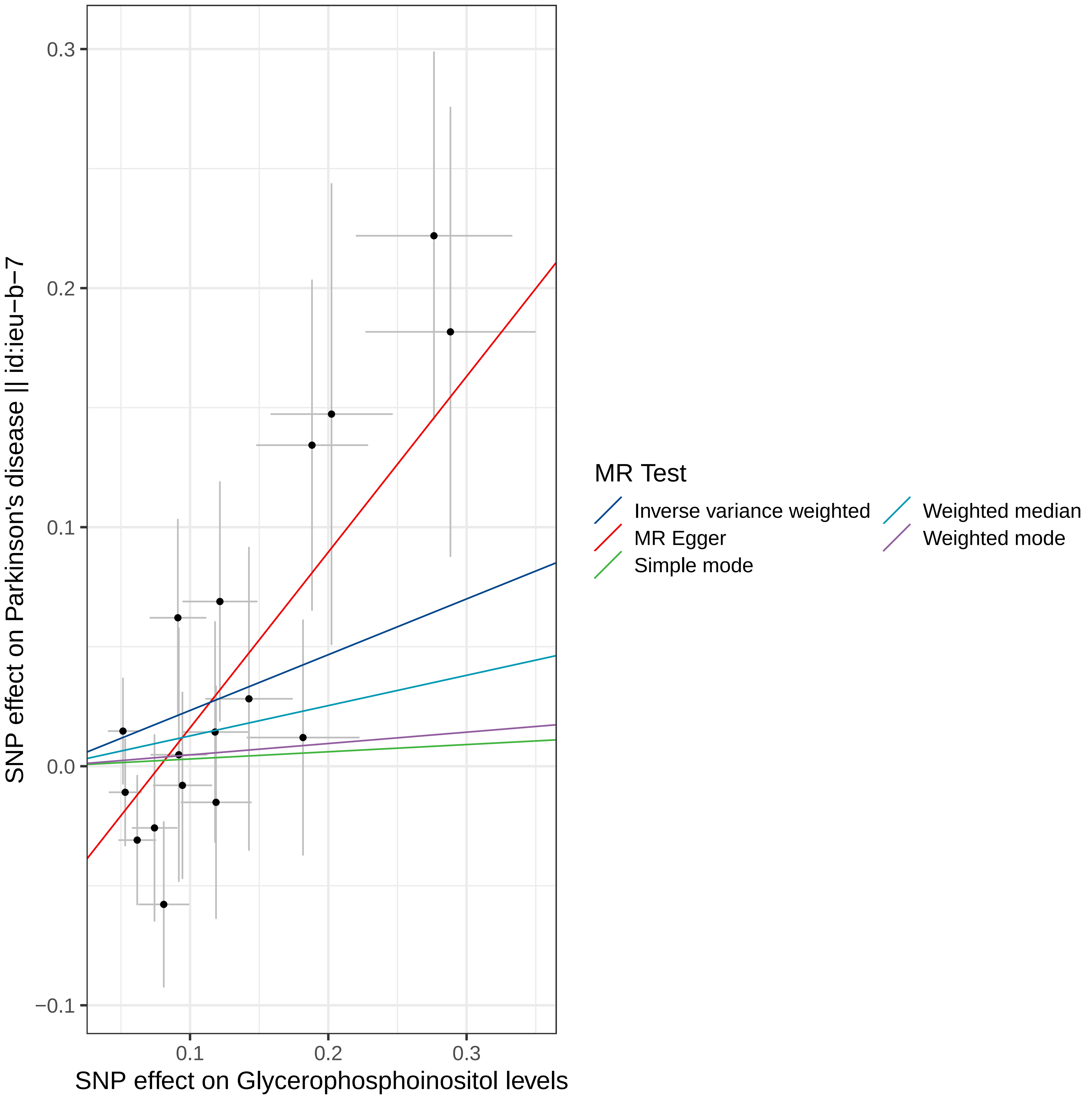

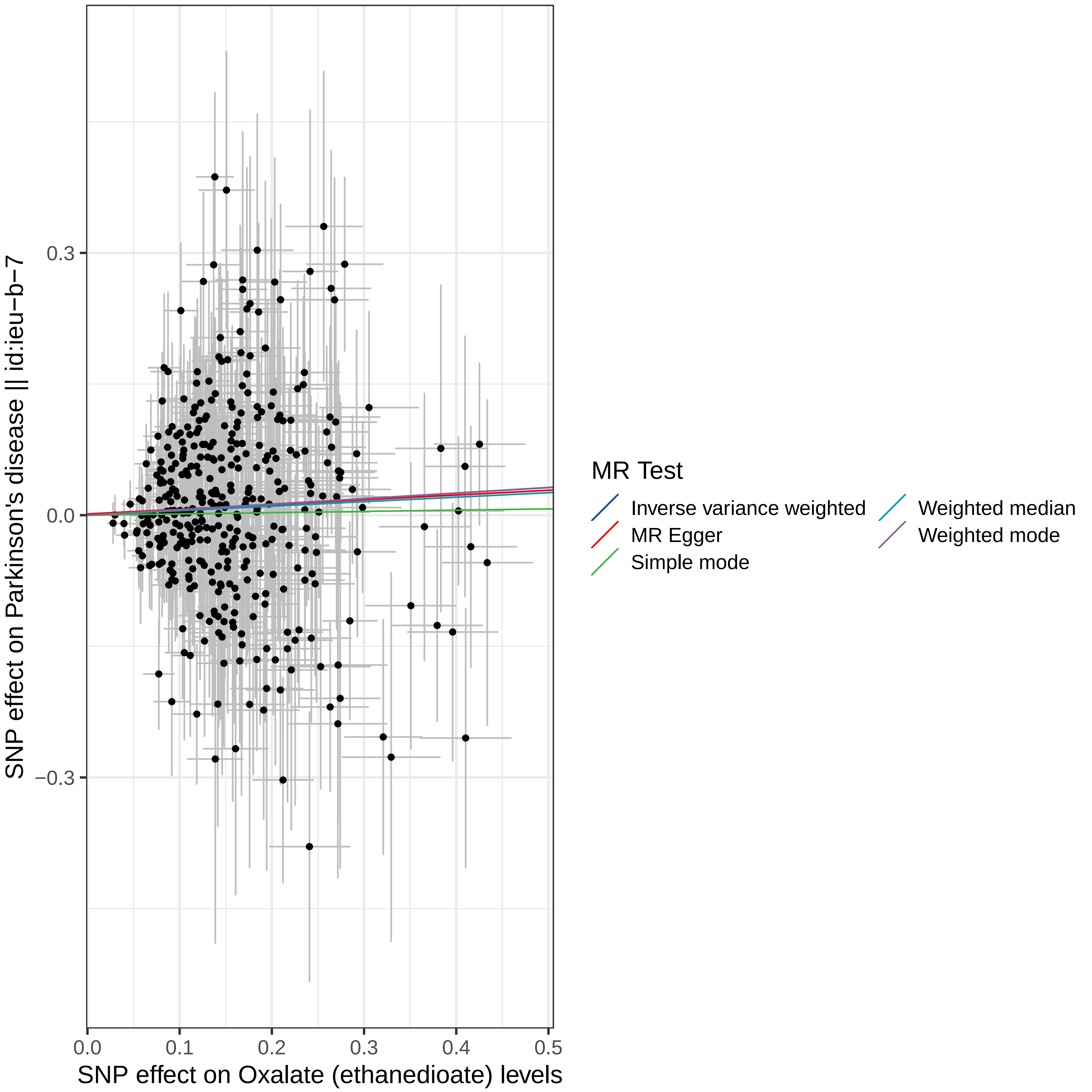

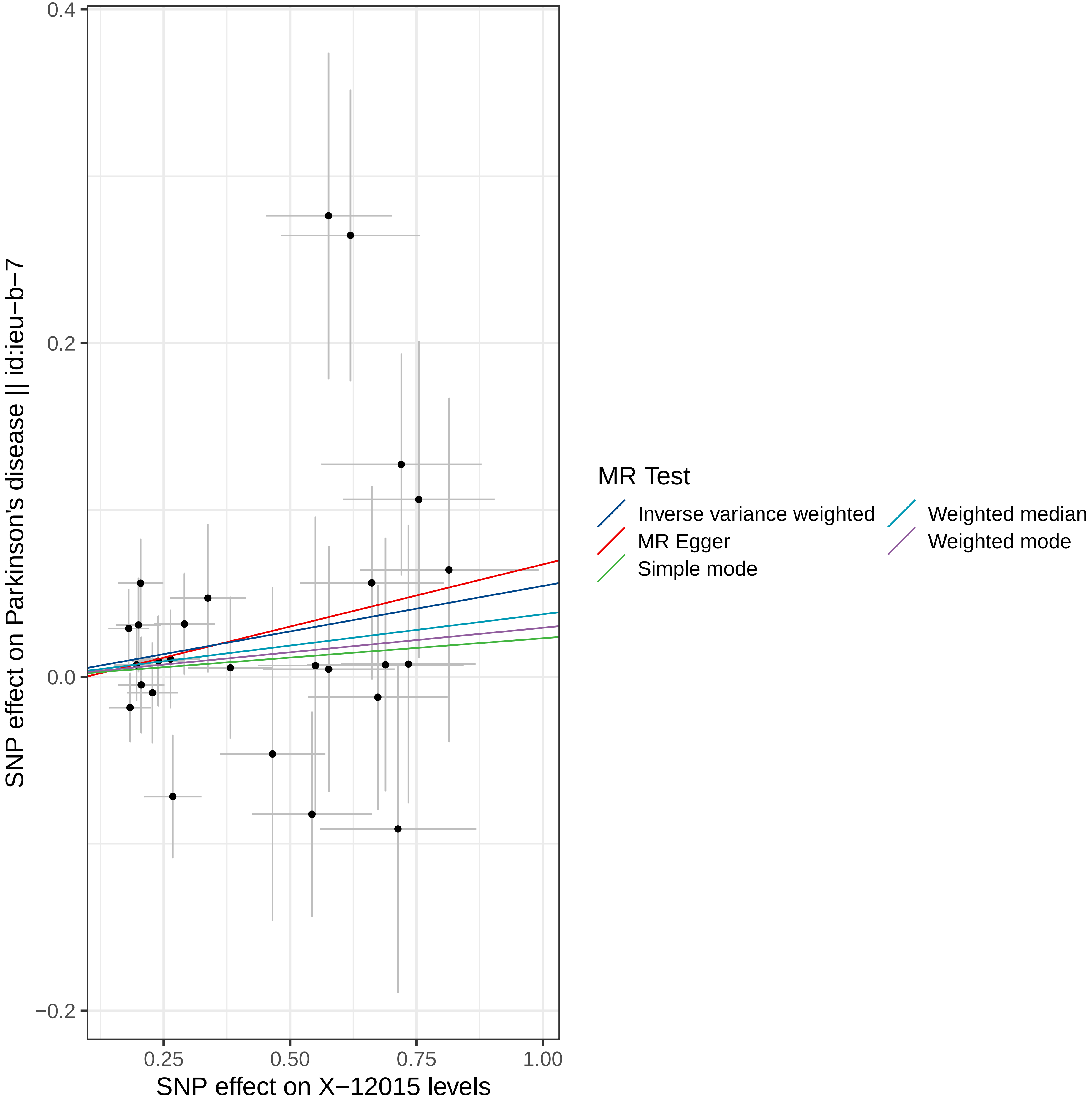

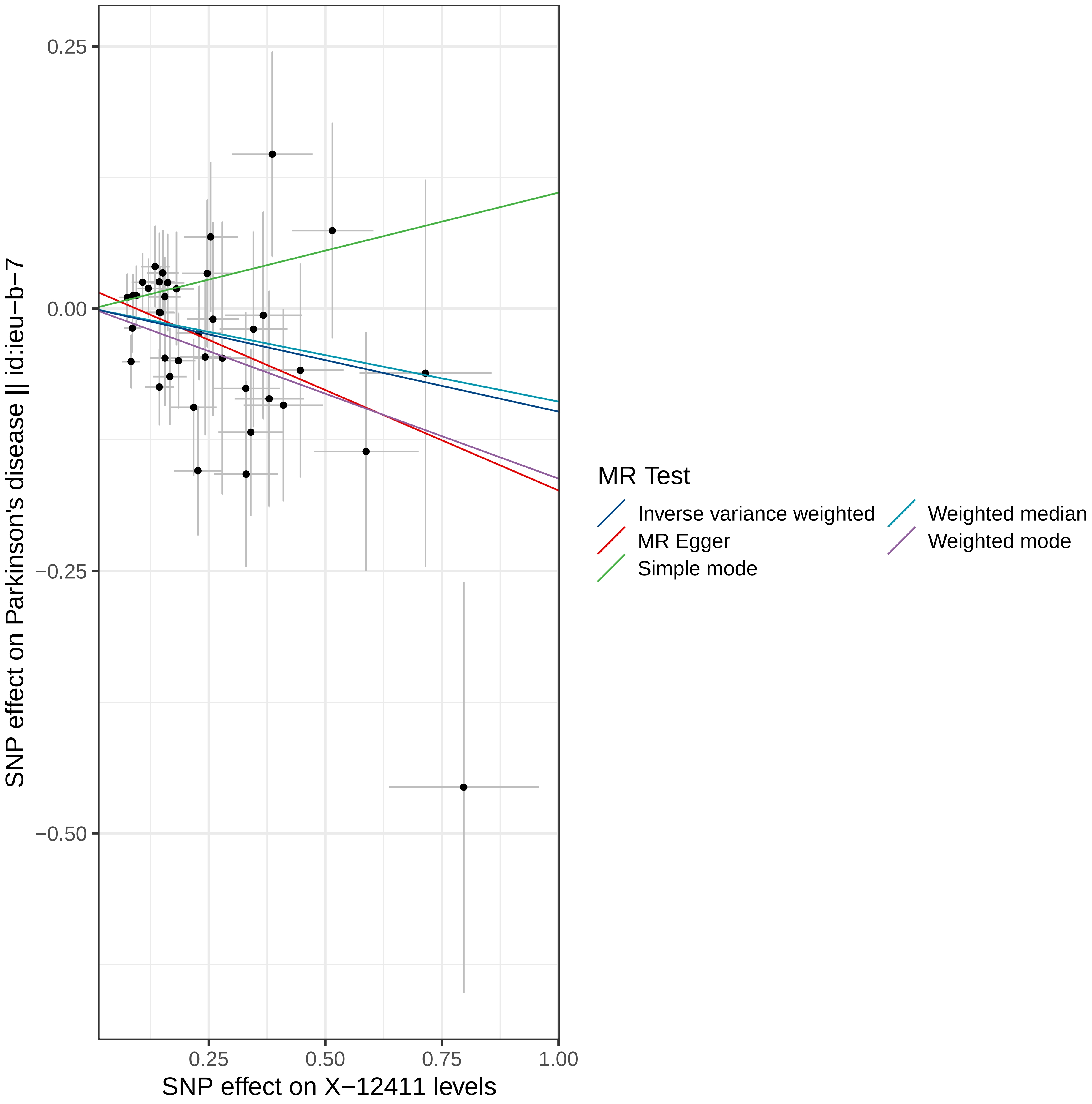

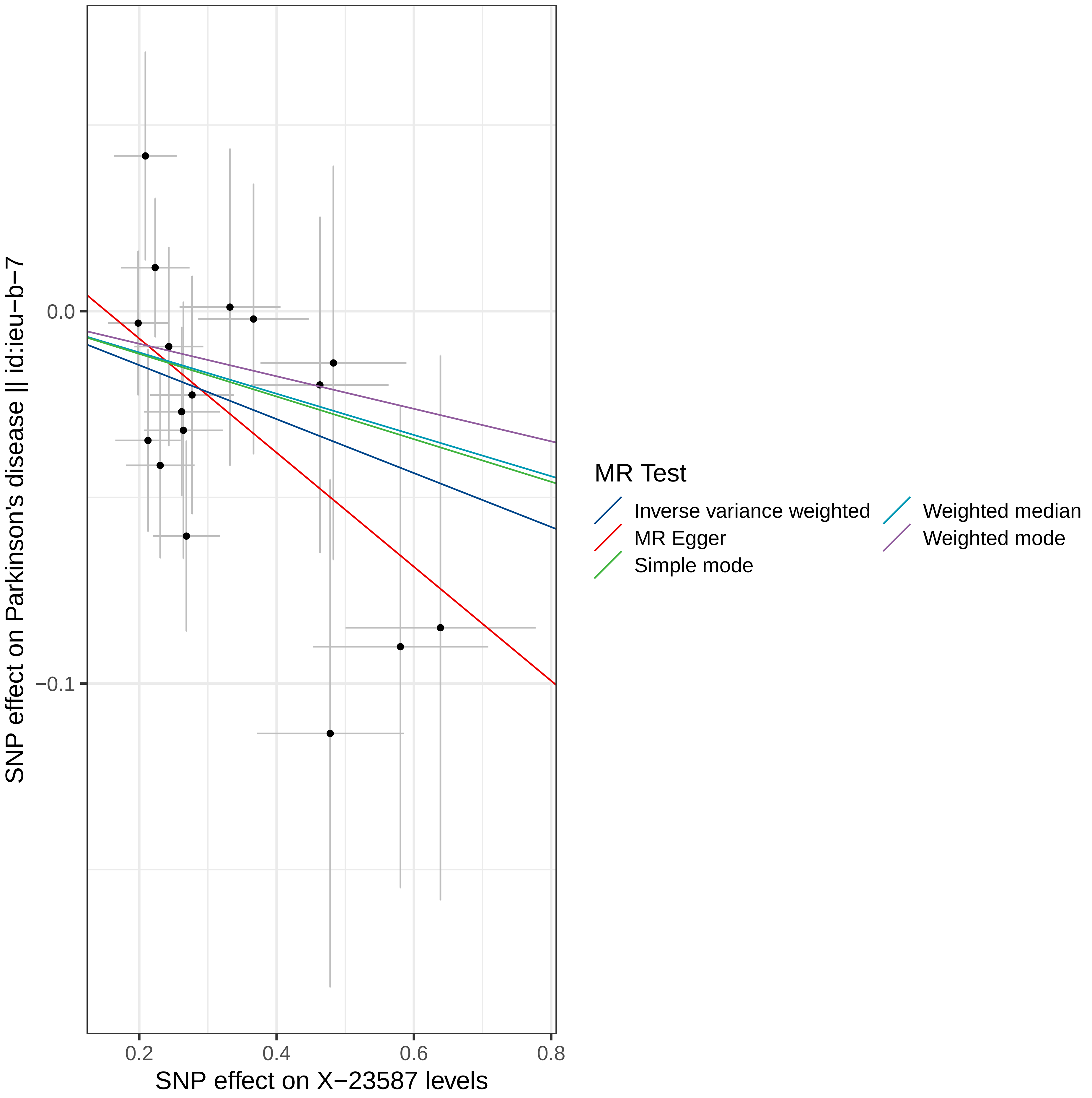

### Supplemental Figure 7

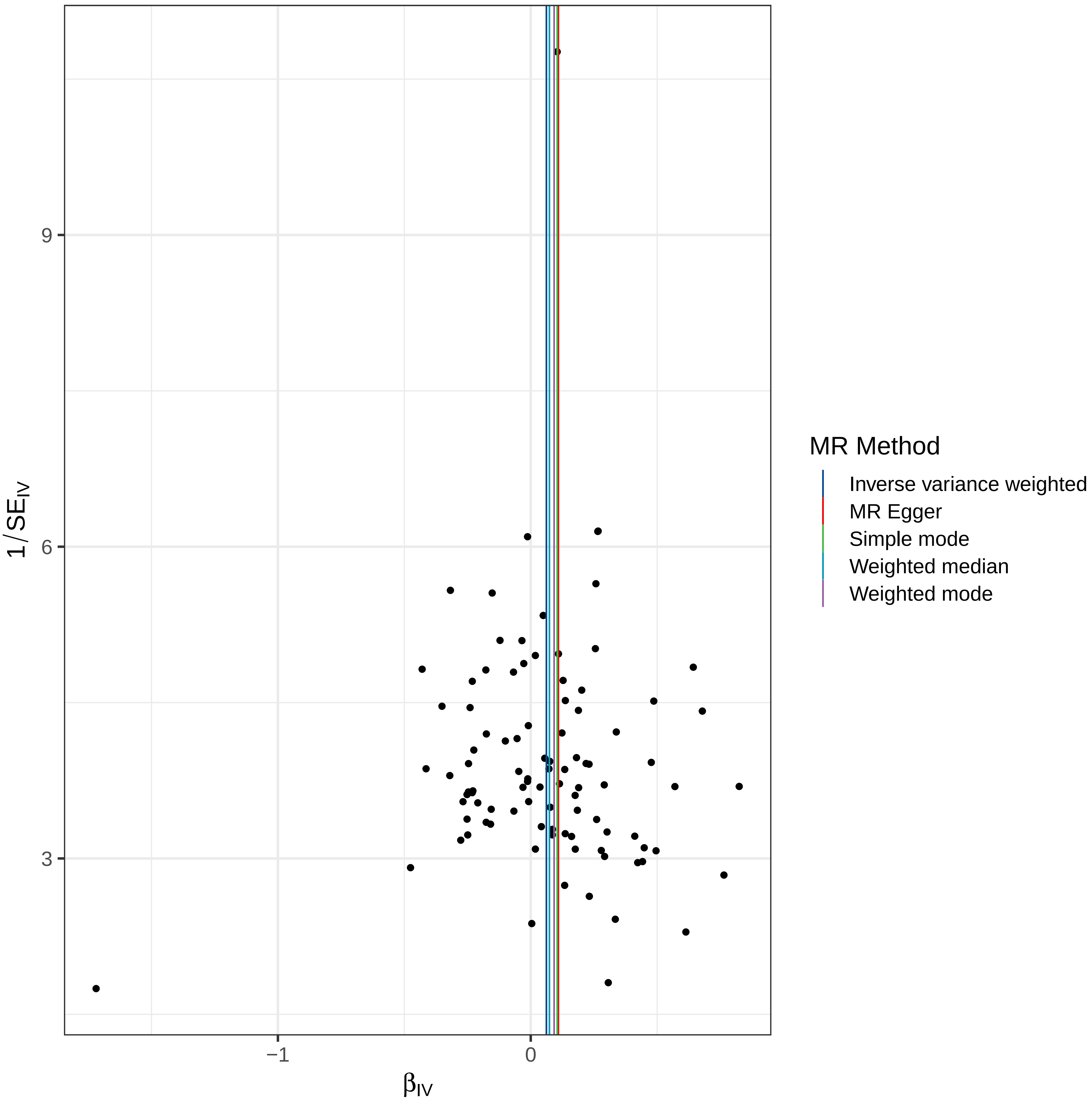

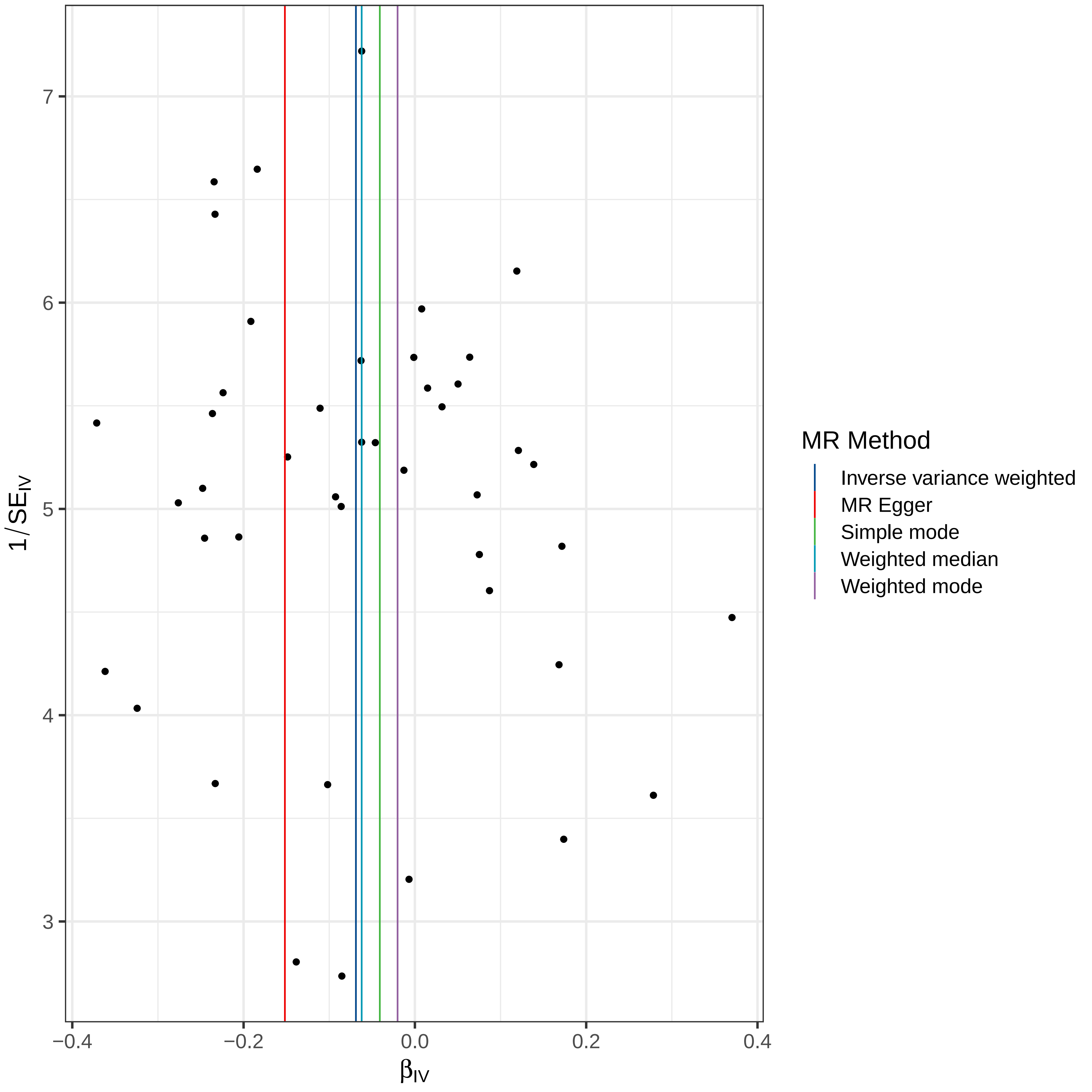

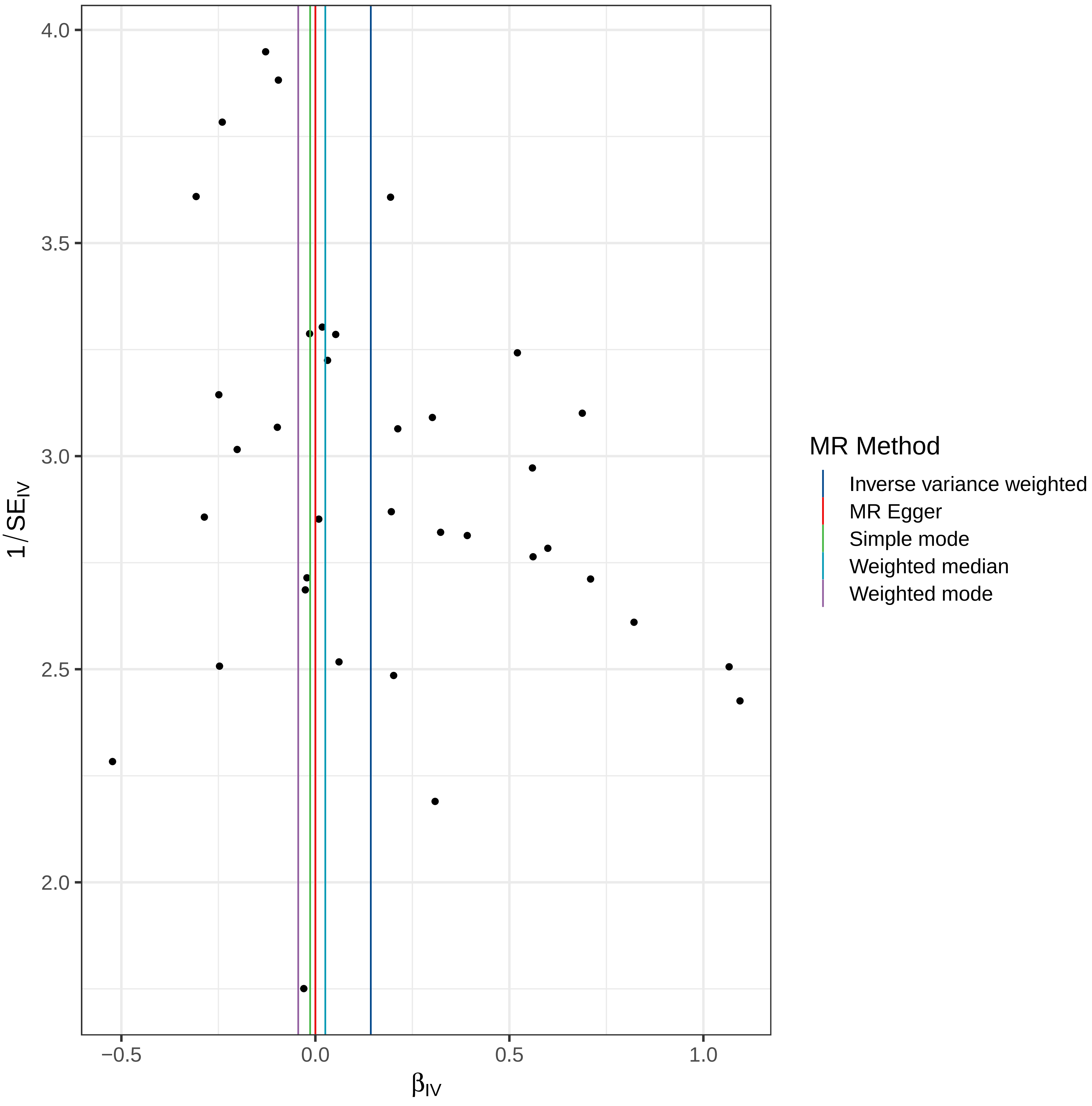

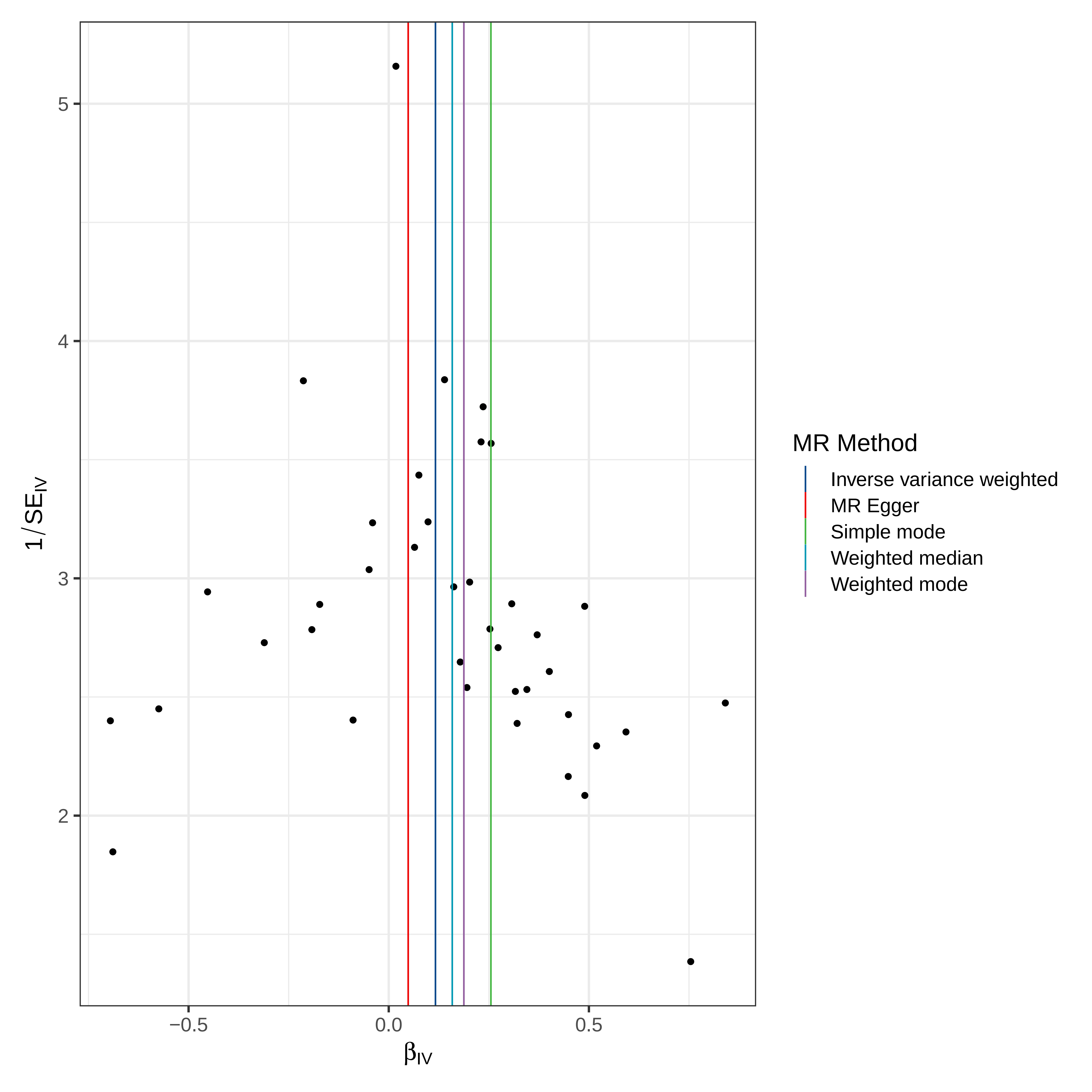

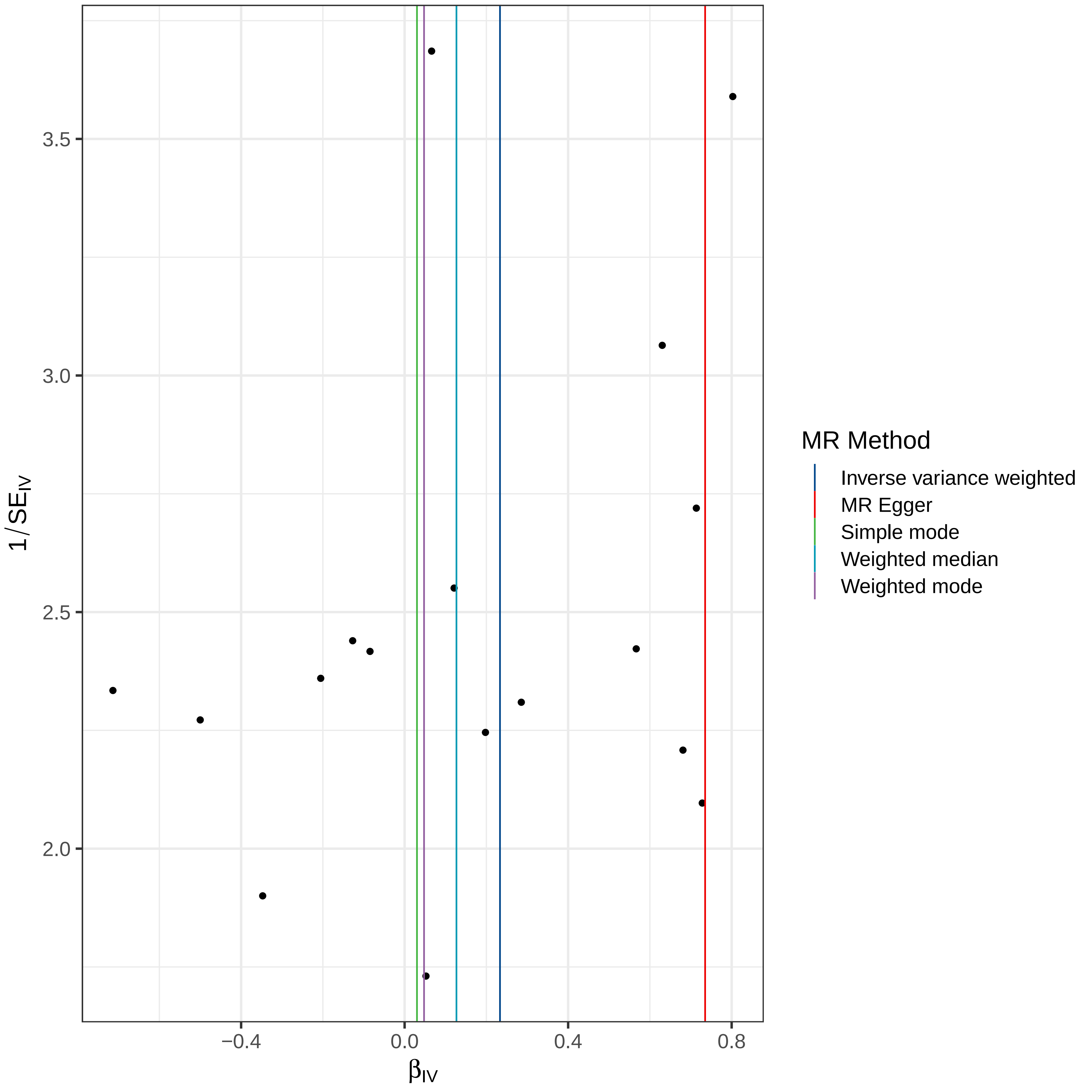

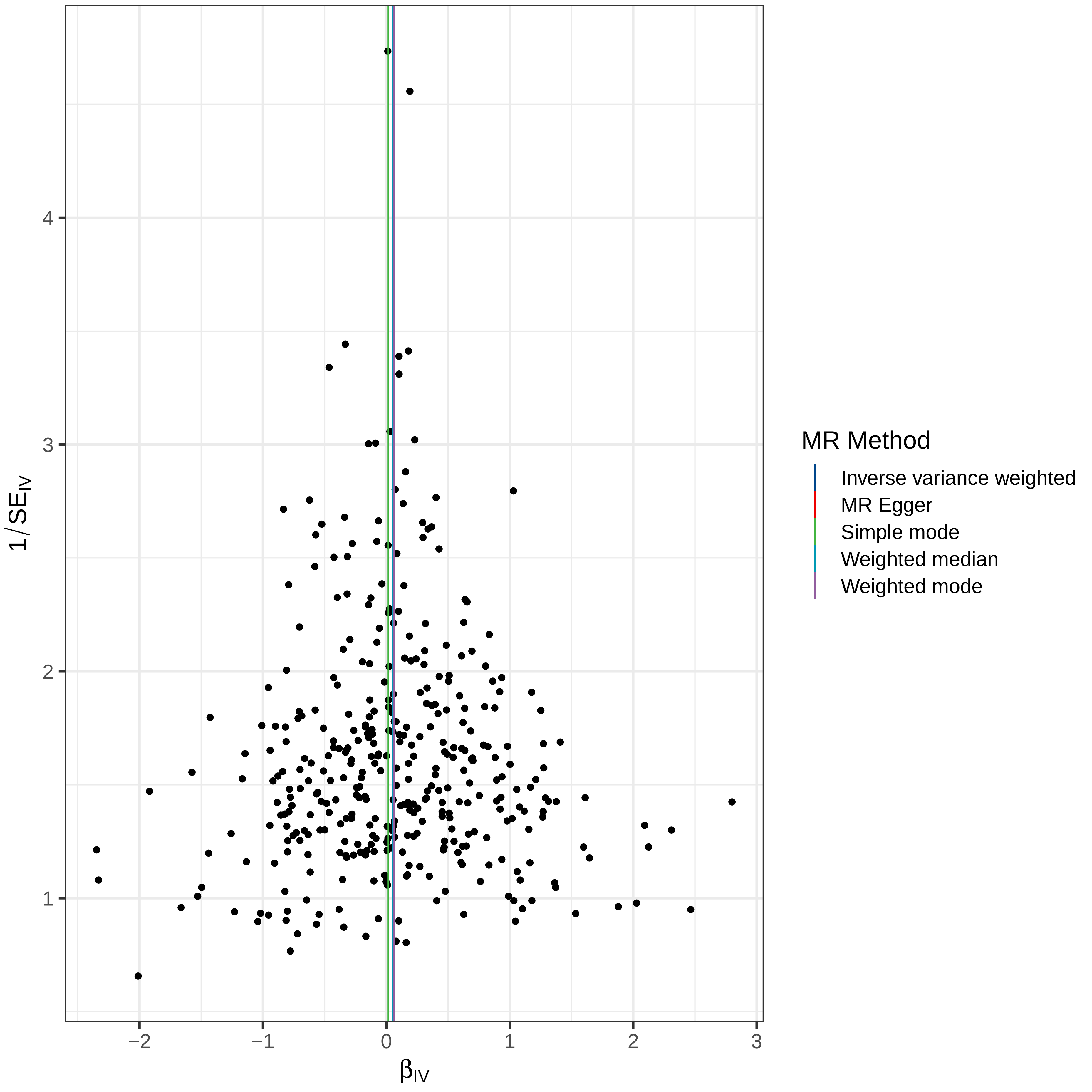

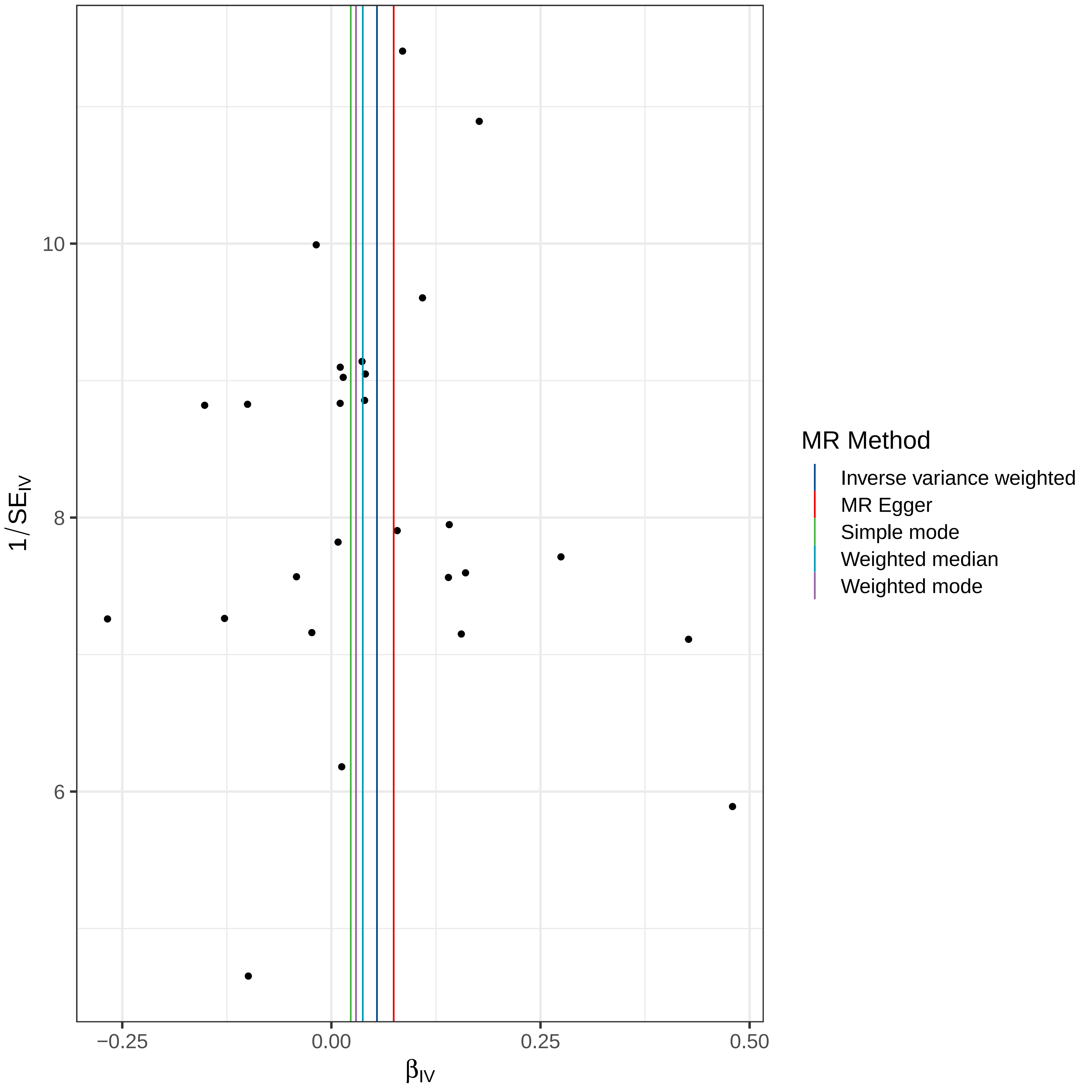

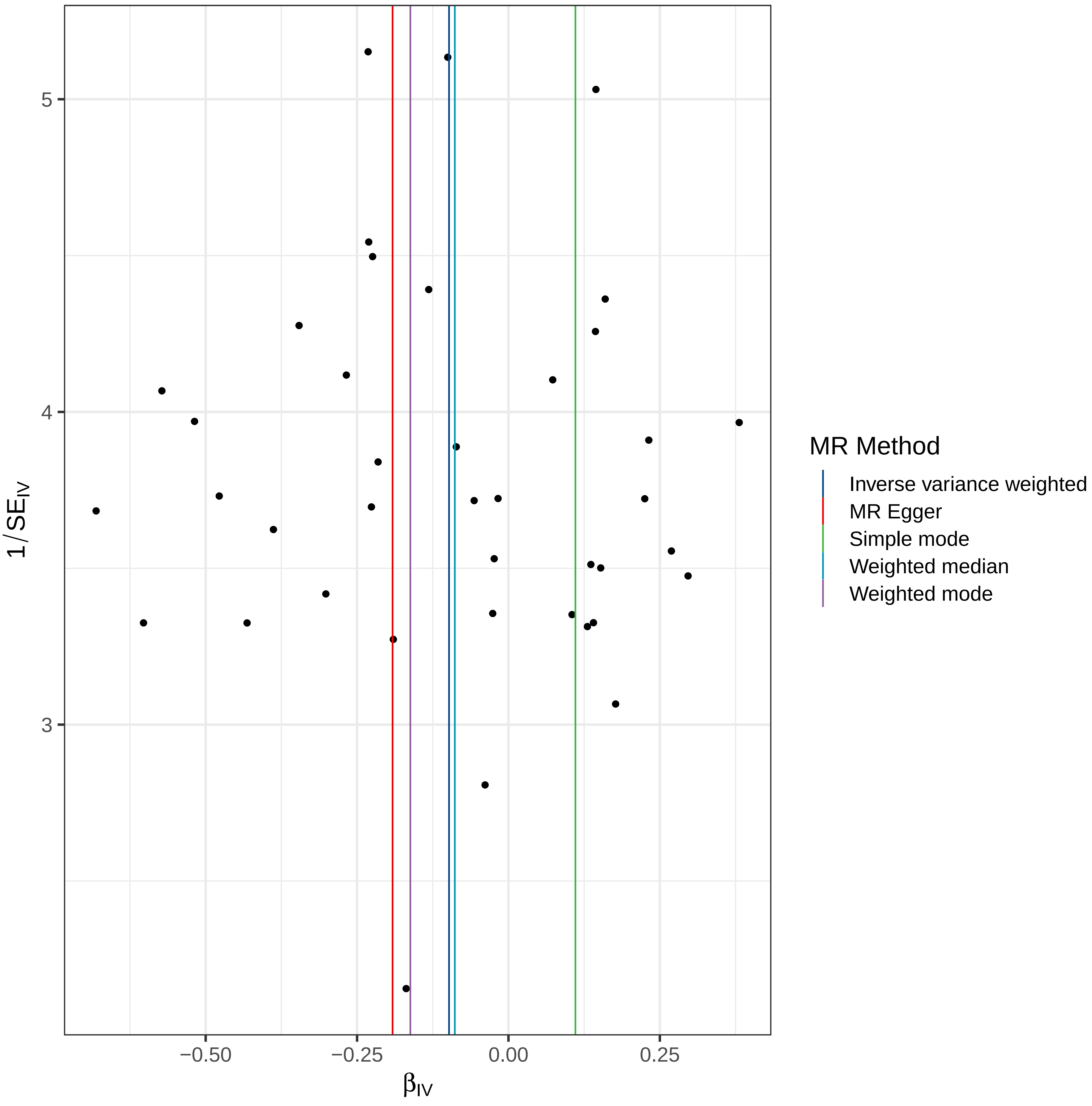

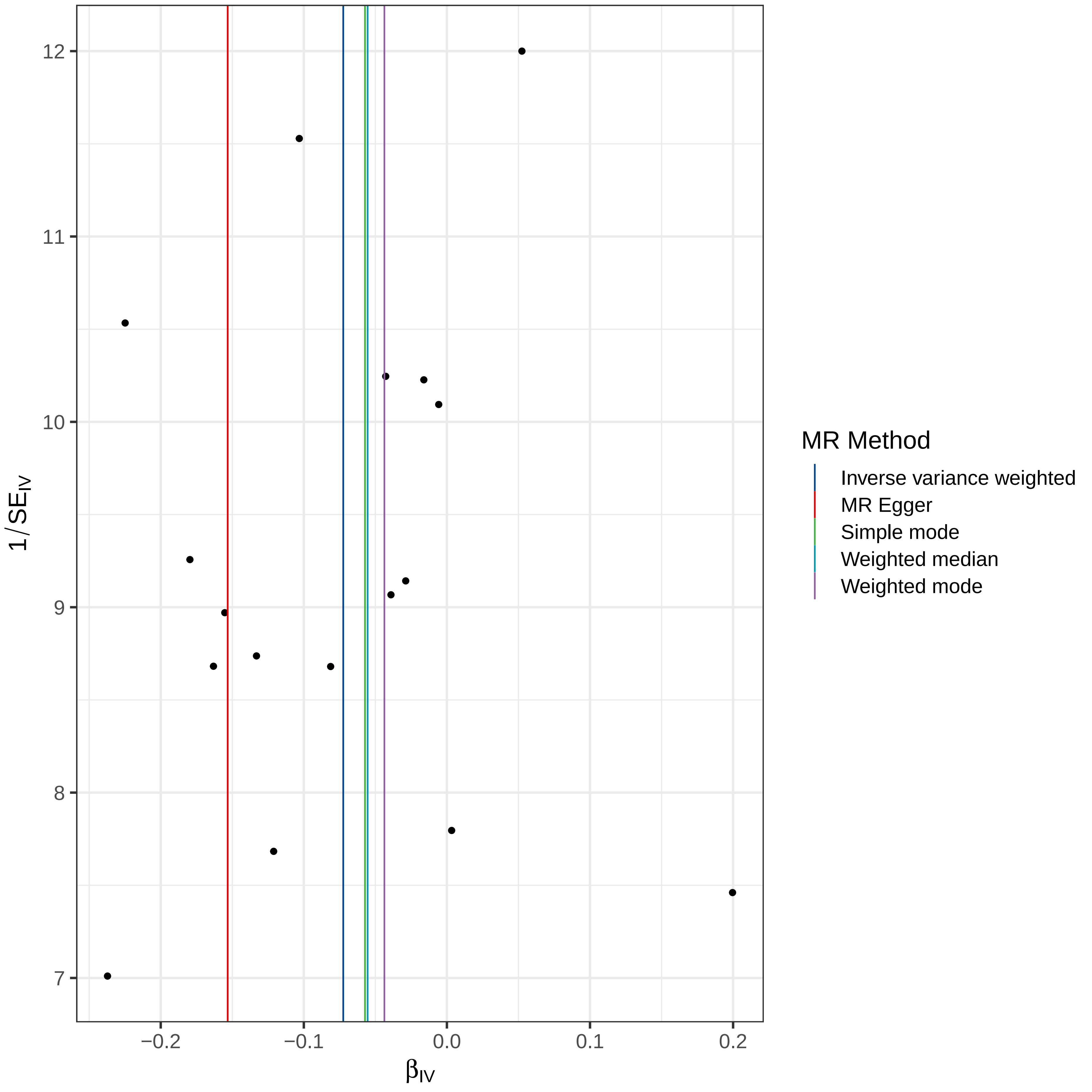

### Supplemental Figure 9

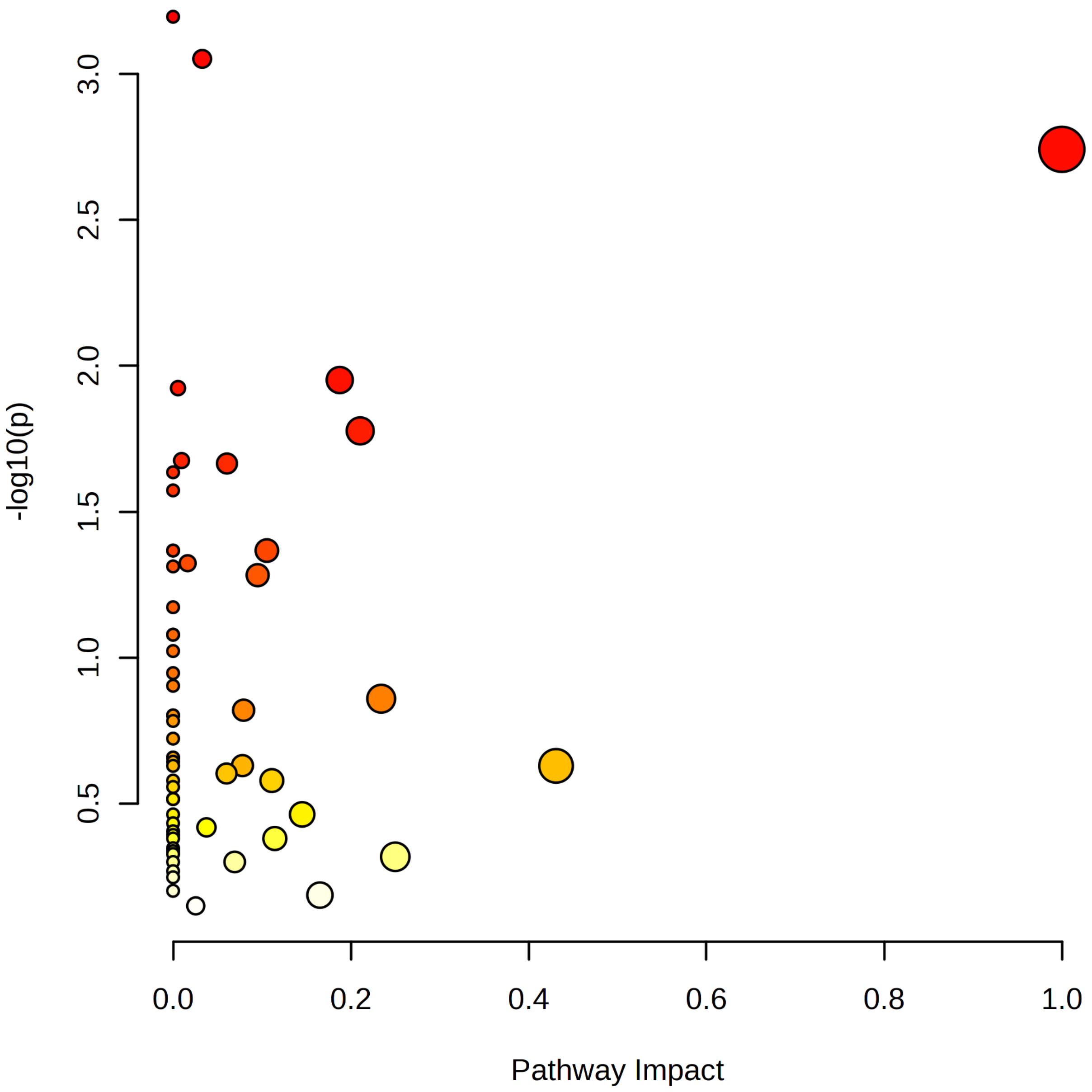
