## Supplemental Figure 2 for "Cerebrospinal Fluid and Plasma Metabolites with Parkinson’s Disease: A Mendelian Randomization Study"

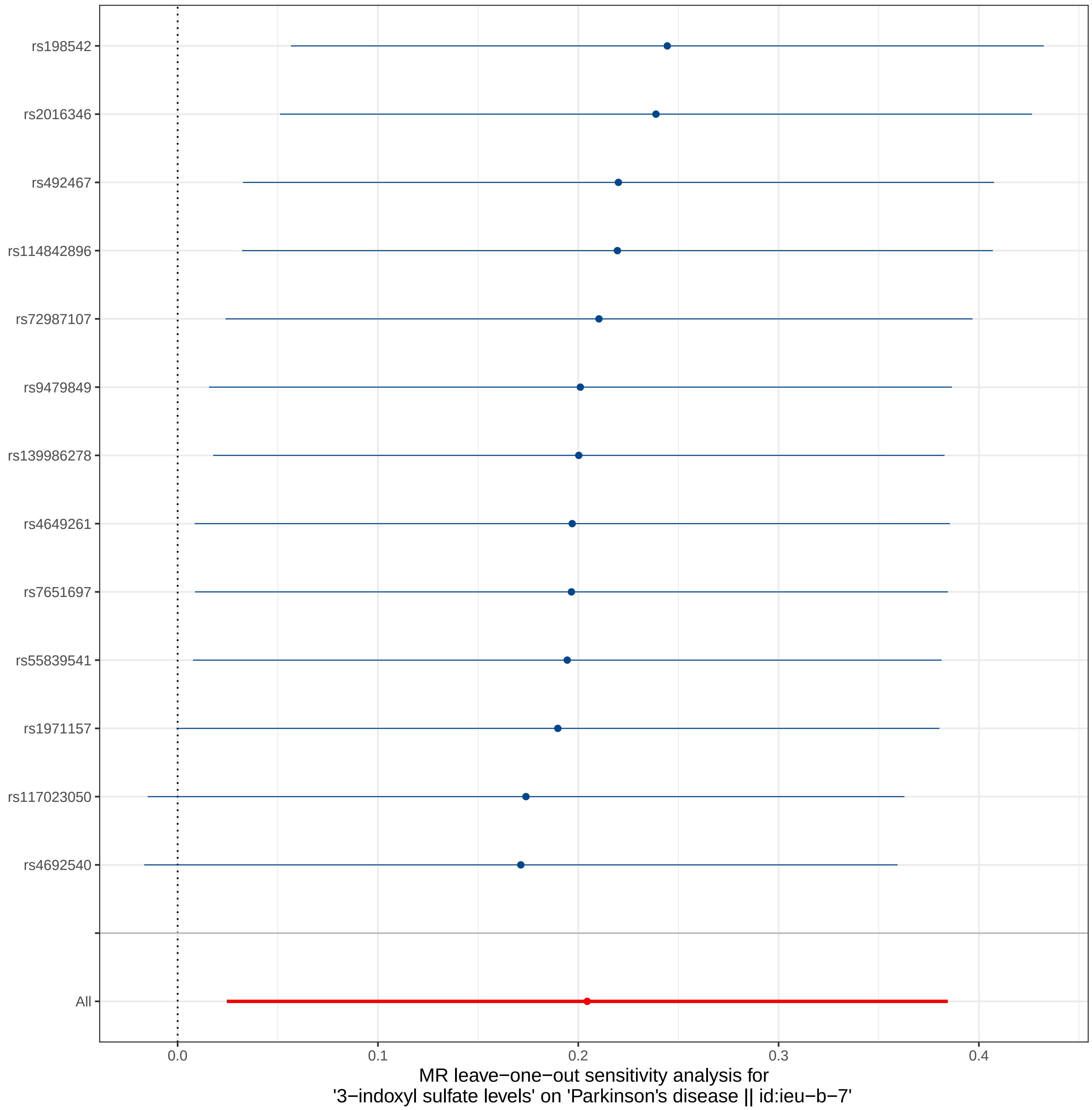
rs198542
rs2016346
rs492467
rs114842896
rs72987107
rs9479849
rs139986278
rs4649261
rs7651697
rs55839541
rs1971157
rs117023050
rs4692540
All
0.0
0.1
0.2
0.3
0.4
MR leave−one−out sensitivity analysis for
'3−indoxyl sulfate levels' on 'Parkinson's disease || id:ieu−b−7'

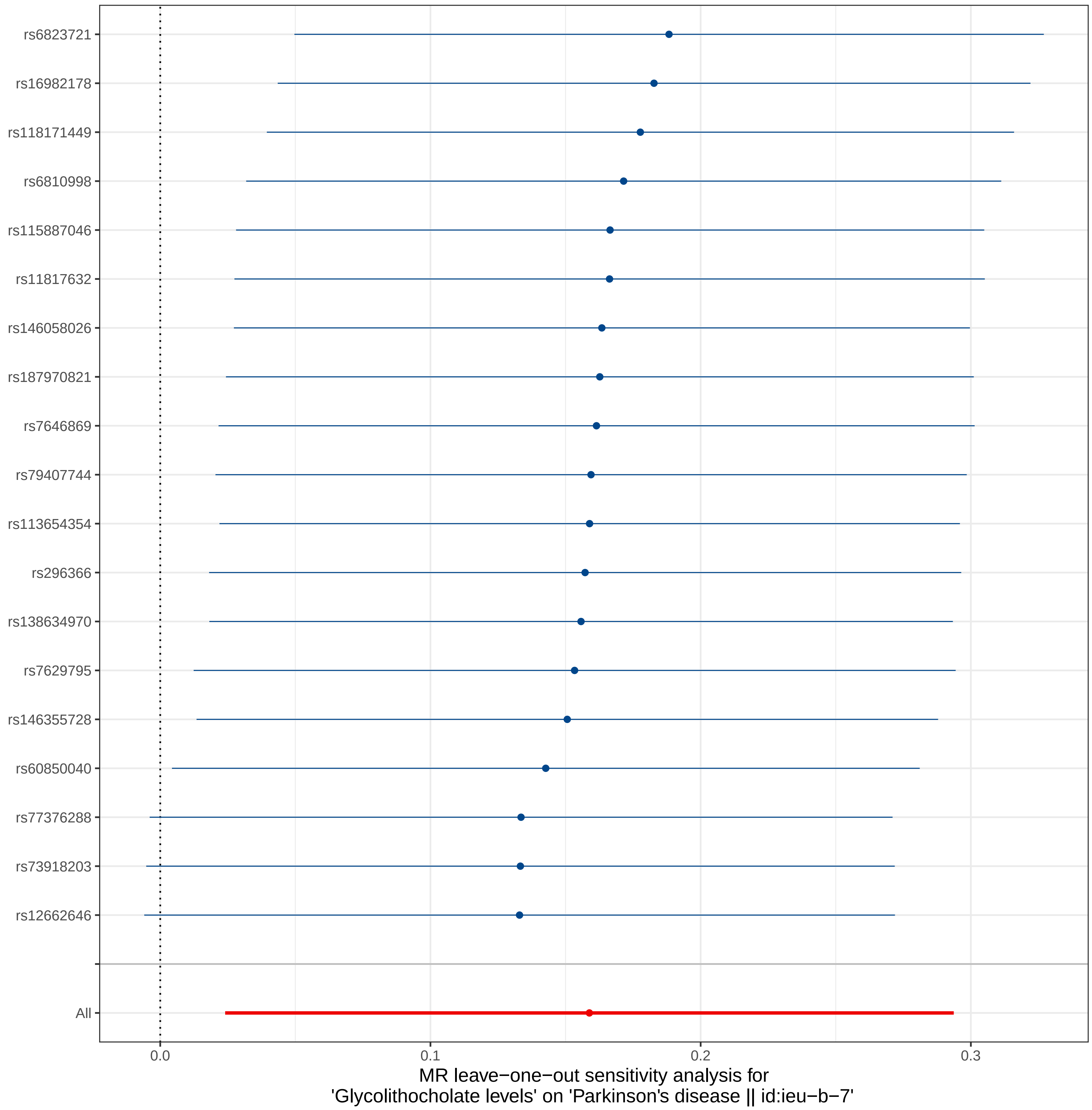

rs6823721
rs16982178
rs118171449
rs6810998
rs115887046
rs11817632
rs146058026
rs187970821
rs7646869
rs79407744
rs113654354
rs296366
rs138634970
rs7629795
rs146355728
rs60850040
rs77376288
rs73918203
rs12662646
All
0.0
0.1
0.2
0.3
MR leave−one−out sensitivity analysis for
'Glycolithocholate levels' on 'Parkinson's disease || id:ieu−b−7'

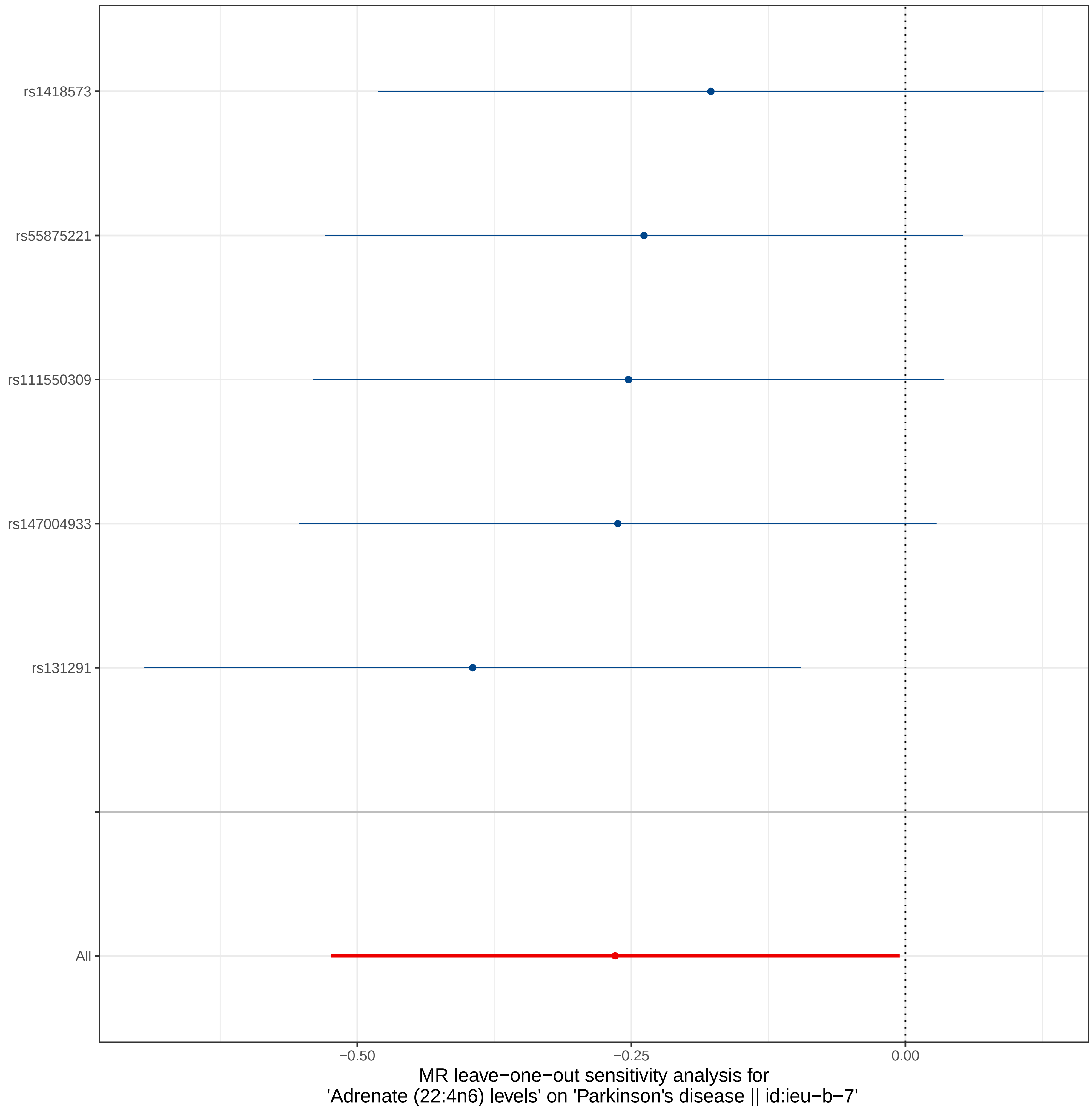

rs1418573
rs55875221
rs111550309
rs147004933
rs131291
All
−0.50
−0.25
0.00
MR leave−one−out sensitivity analysis for
'Adrenate (22:4n6) levels' on 'Parkinson's disease || id:ieu−b−7'

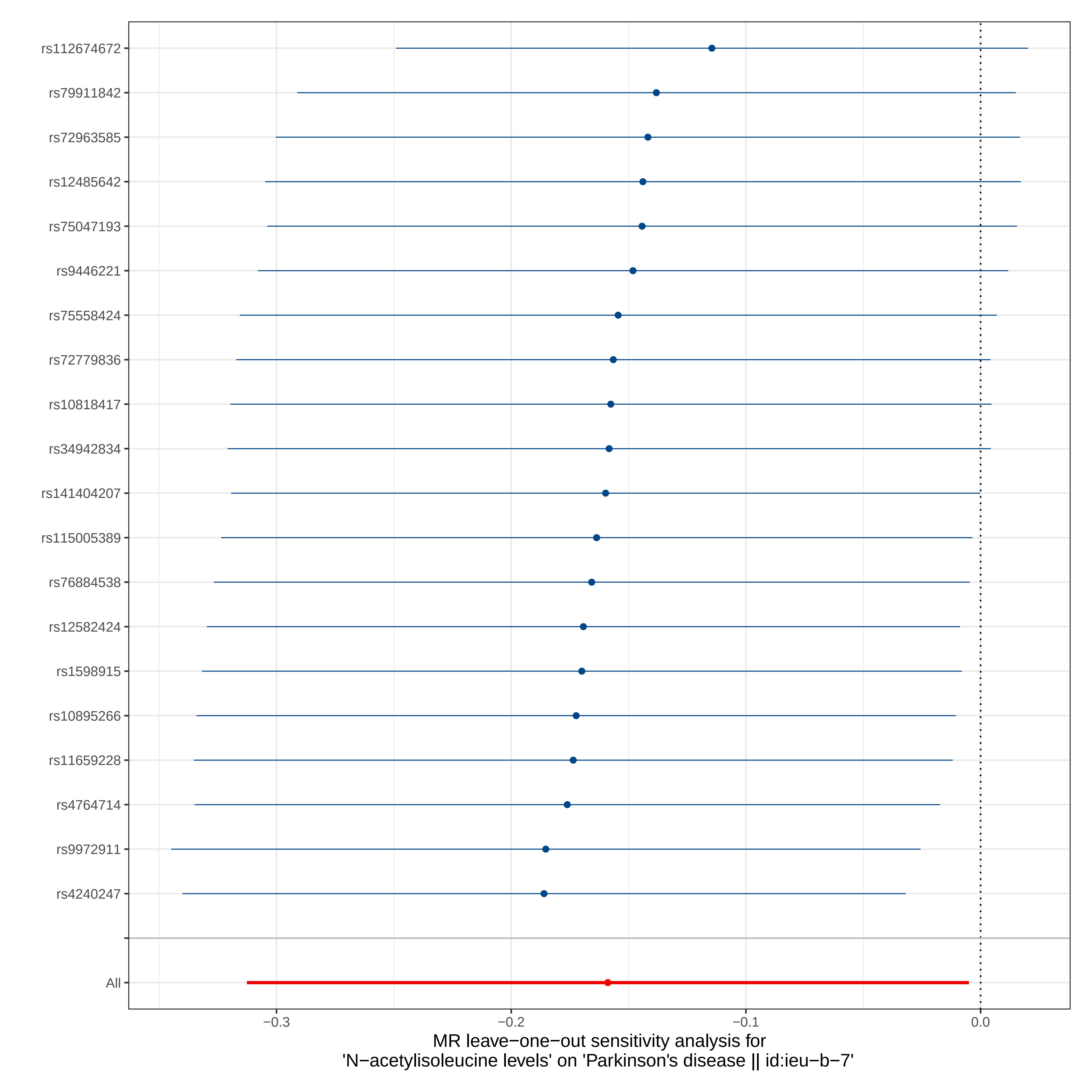

rs112674672
rs79911842
rs72963585
rs12485642
rs75047193
rs9446221
rs75558424
rs72779836
rs10818417
rs34942834
rs141404207
rs115005389
rs76884538
rs12582424
rs1598915
rs10895266
rs11659228
rs4764714
rs9972911
rs4240247
All
−0.3
−0.2
−0.1
0.0
MR leave−one−out sensitivity analysis for
'N−acetylisoleucine levels' on 'Parkinson's disease || id:ieu−b−7'

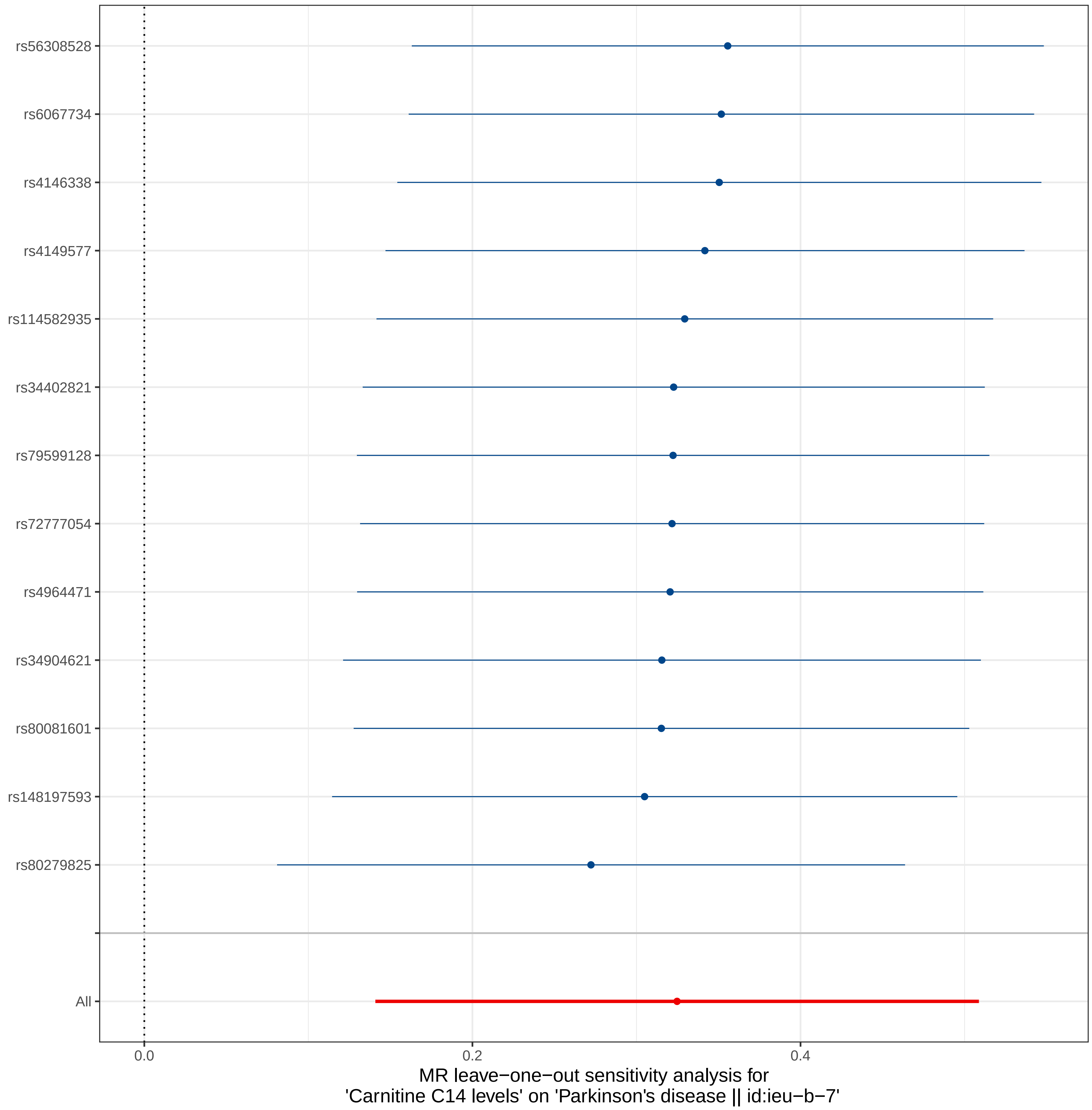
rs56308528
rs6067734
rs4146338
rs4149577
rs114582935
rs34402821
rs79599128
rs72777054
rs4964471
rs34904621
rs80081601
rs148197593
rs80279825
All
0.0
0.2
0.4
MR leave−one−out sensitivity analysis for
'Carnitine C14 levels' on 'Parkinson's disease || id:ieu−b−7'

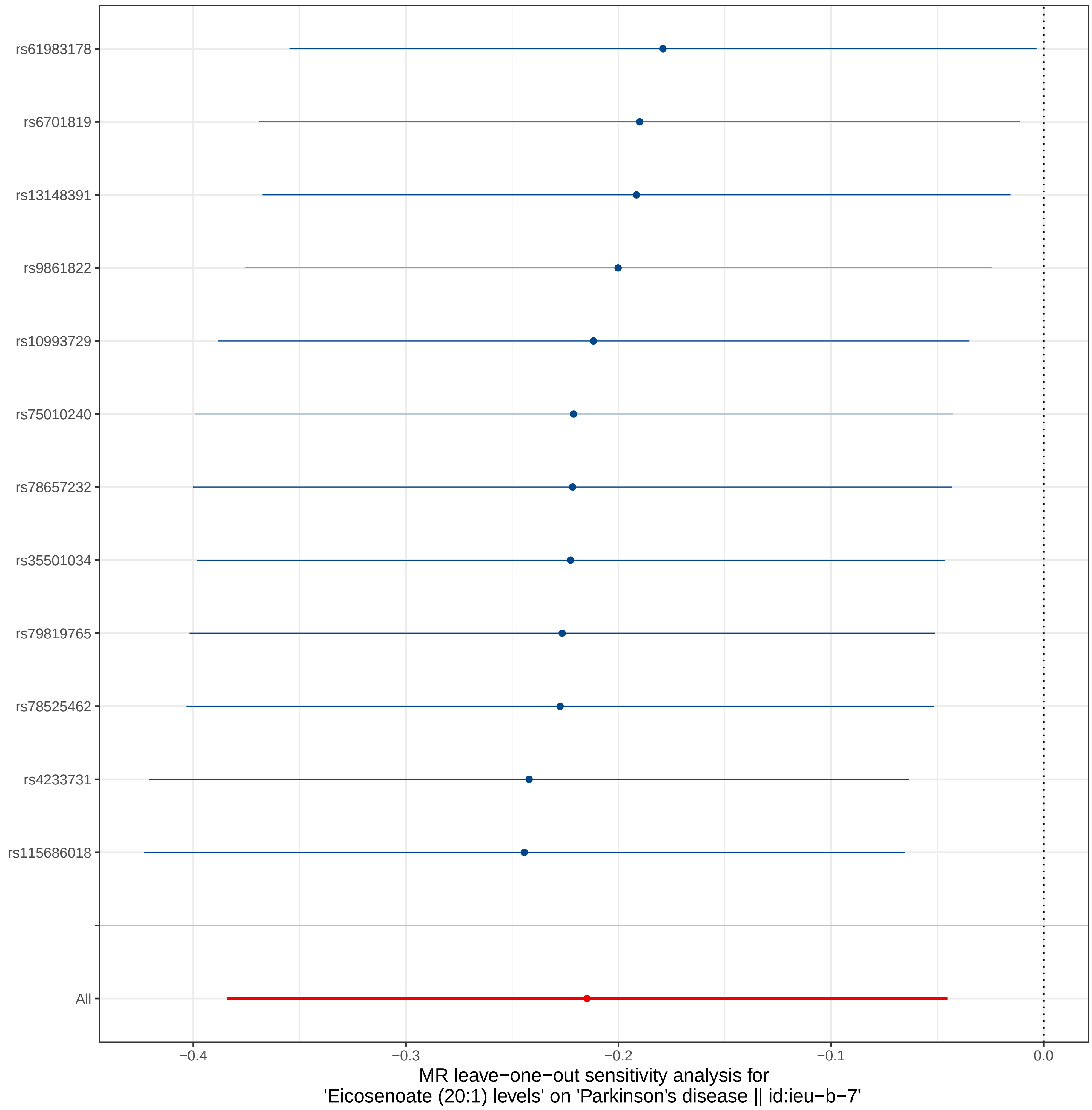

rs61983178
rs6701819
rs13148391
rs9861822
rs10993729
rs75010240
rs78657232
rs35501034
rs79819765
rs78525462
rs4233731
rs115686018
All
−0.4
−0.3
−0.2
−0.1
0.0
MR leave−one−out sensitivity analysis for
'Eicosenoate (20:1) levels' on 'Parkinson's disease || id:ieu−b−7'

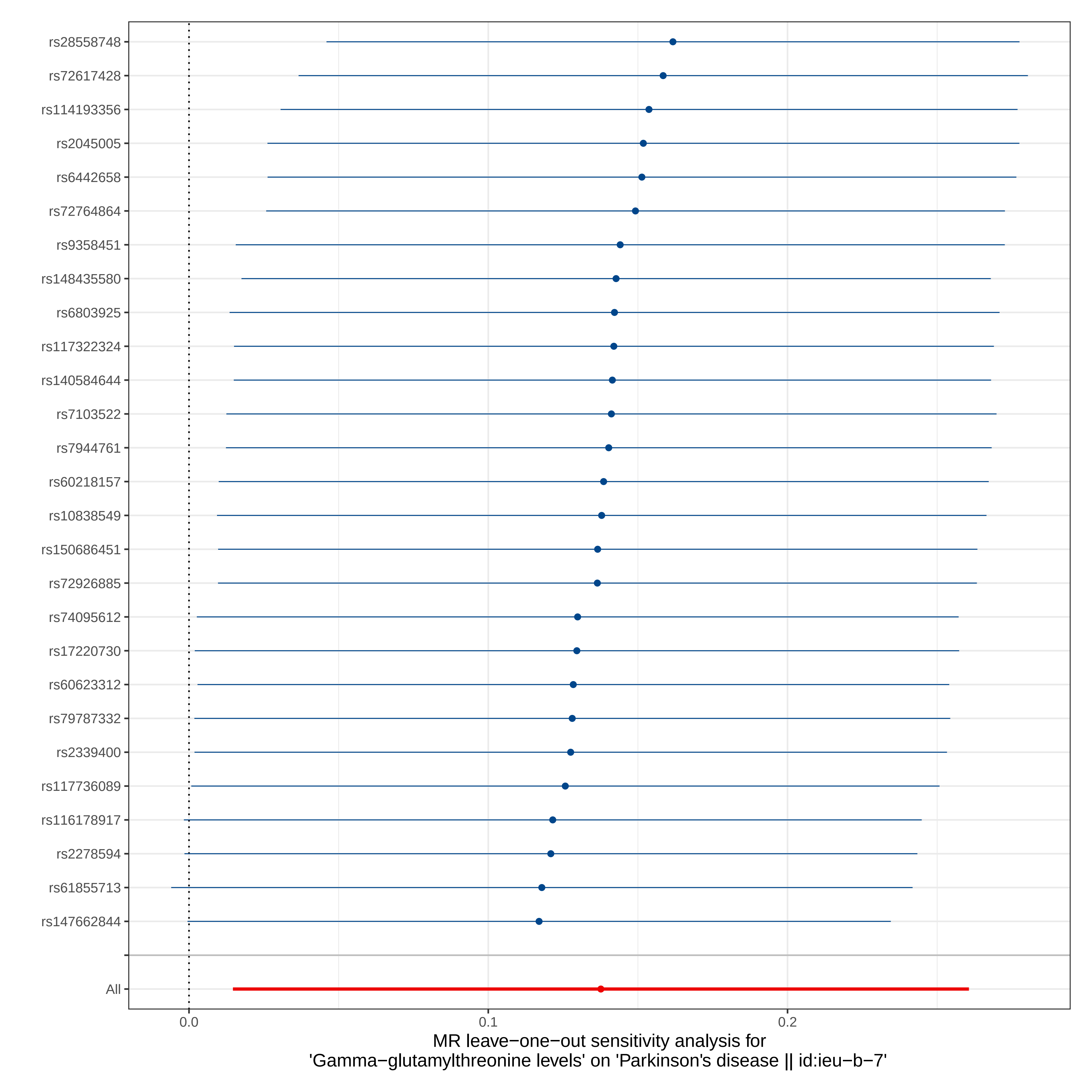
rs28558748
rs72617428
rs114193356
rs2045005
rs6442658
rs72764864
rs9358451
rs148435580
rs6803925
rs117322324
rs140584644
rs7103522
rs7944761
rs60218157
rs10838549
rs150686451
rs72926885
rs74095612
rs17220730
rs60623312
rs79787332
rs2339400
rs117736089
rs116178917
rs2278594
rs61855713
rs147662844
All
0.0
0.1
0.2
MR leave−one−out sensitivity analysis for
'Gamma−glutamylthreonine levels' on 'Parkinson's disease || id:ieu−b−7'

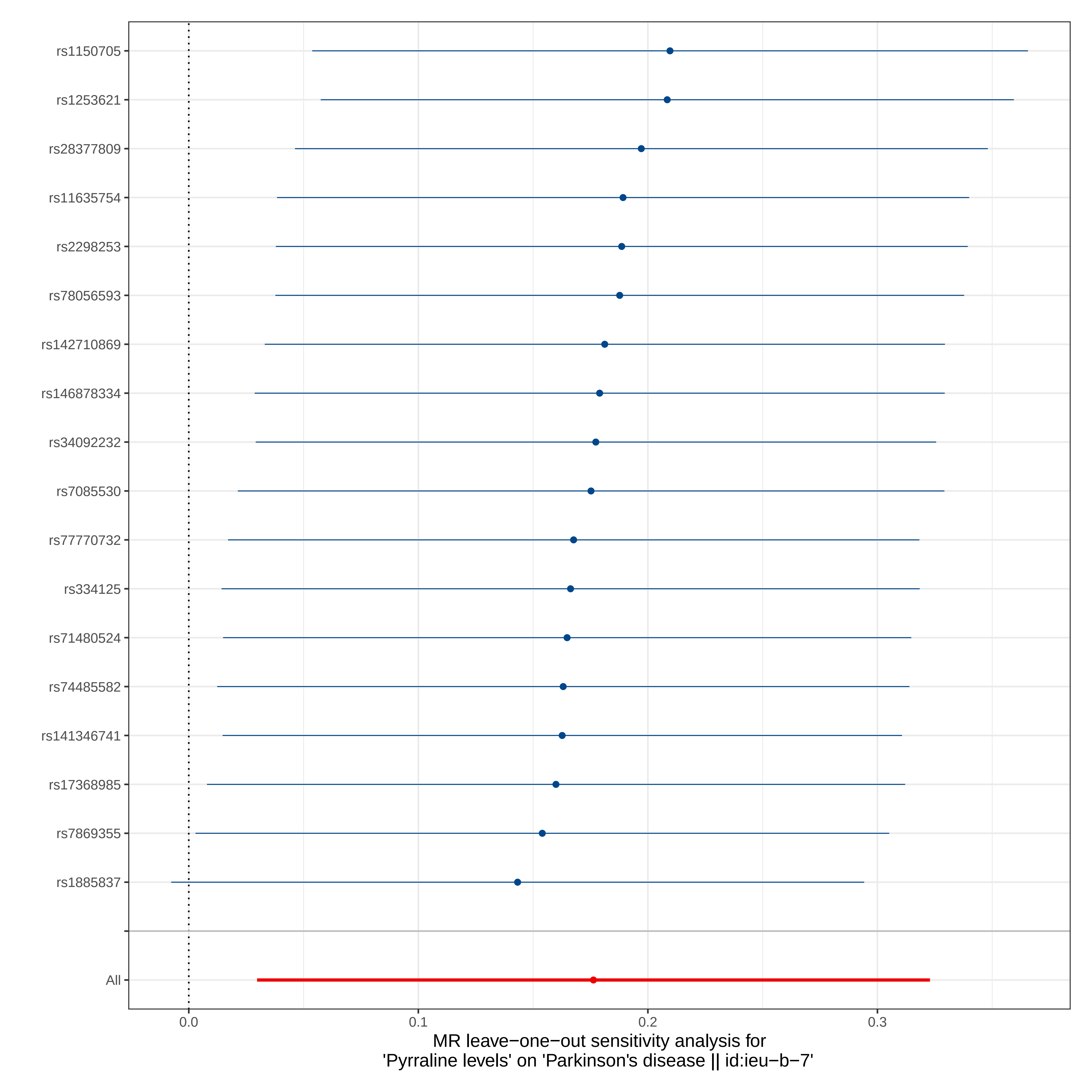

rs1150705
rs1253621
rs28377809
rs11635754
rs2298253
rs78056593
rs142710869
rs146878334
rs34092232
rs7085530
rs77770732
rs334125
rs71480524
rs74485582
rs141346741
rs17368985
rs7869355
rs1885837
All
0.0
0.1
0.2
0.3
MR leave−one−out sensitivity analysis for
'Pyrraline levels' on 'Parkinson's disease || id:ieu−b−7'

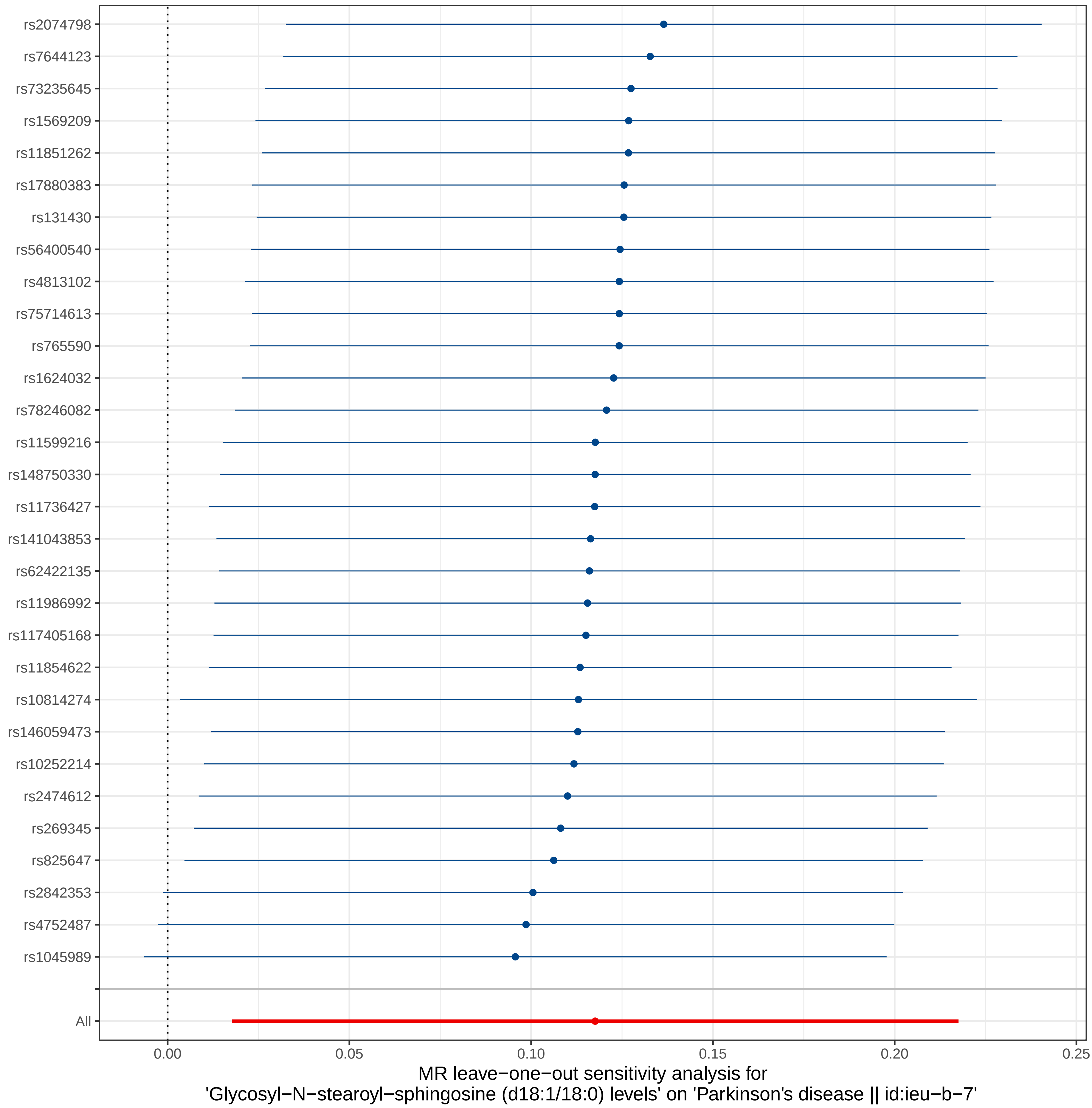
rs2074798
rs7644123
rs73235645
rs1569209
rs11851262
rs17880383
rs131430
rs56400540
rs4813102
rs75714613
rs765590
rs1624032
rs78246082
rs11599216
rs148750330
rs11736427
rs141043853
rs62422135
rs11986992
rs117405168
rs11854622
rs10814274
rs146059473
rs10252214
rs2474612
rs269345
rs825647
rs2842353
rs4752487
rs1045989
All
0.00
0.05
0.10
0.15
0.20
0.25
MR leave−one−out sensitivity analysis for
'Glycosyl−N−stearoyl−sphingosine (d18:1/18:0) levels' on 'Parkinson's disease || id:ieu−b−7'

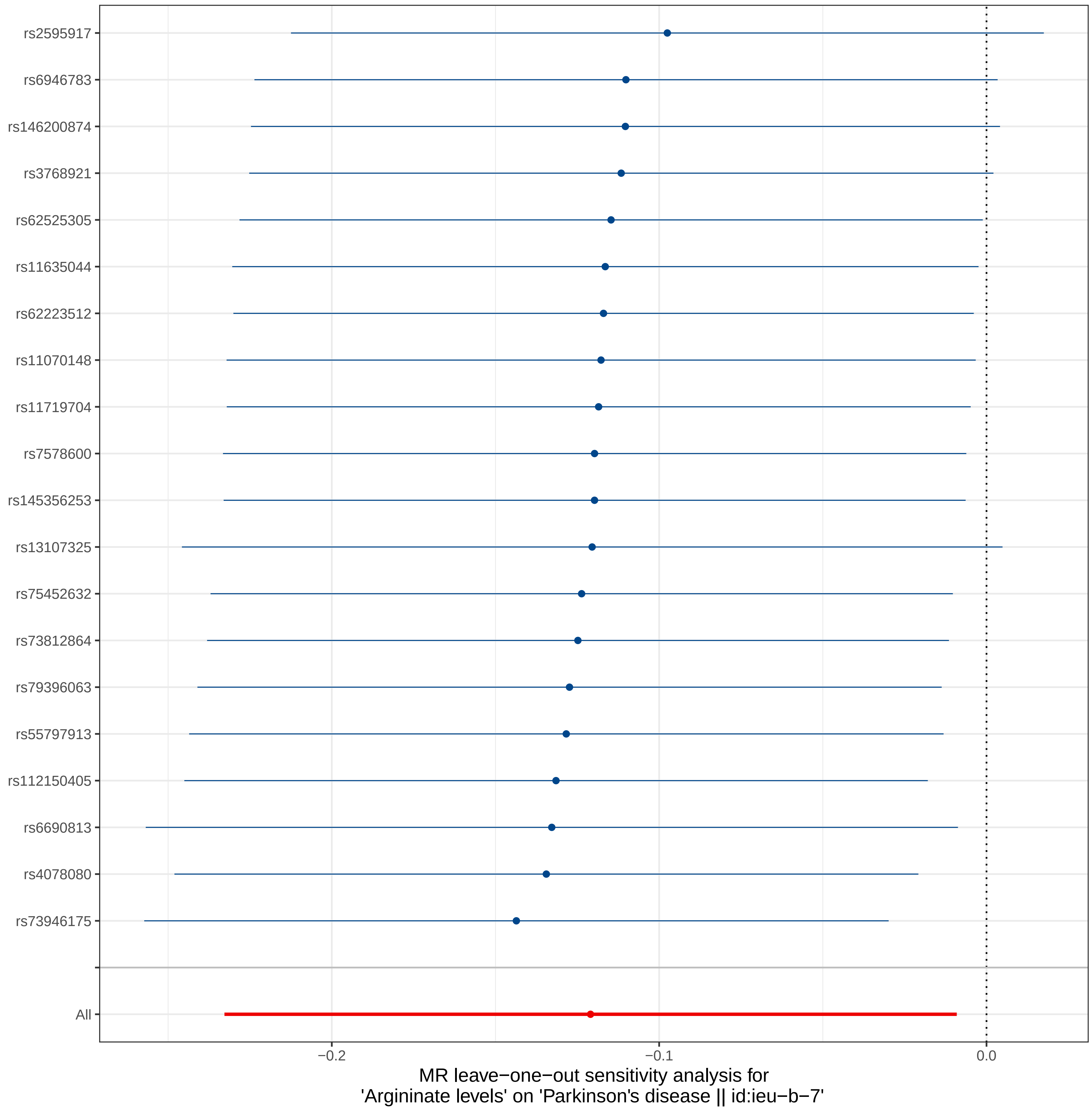
rs2595917
rs6946783
rs146200874
rs3768921
rs62525305
rs11635044
rs62223512
rs11070148
rs11719704
rs7578600
rs145356253
rs13107325
rs75452632
rs73812864
rs79396063
rs55797913
rs112150405
rs6690813
rs4078080
rs73946175
All
−0.2
−0.1
0.0
MR leave−one−out sensitivity analysis for
'Arginate levels' on 'Parkinson's disease || id:ieu−b−7'

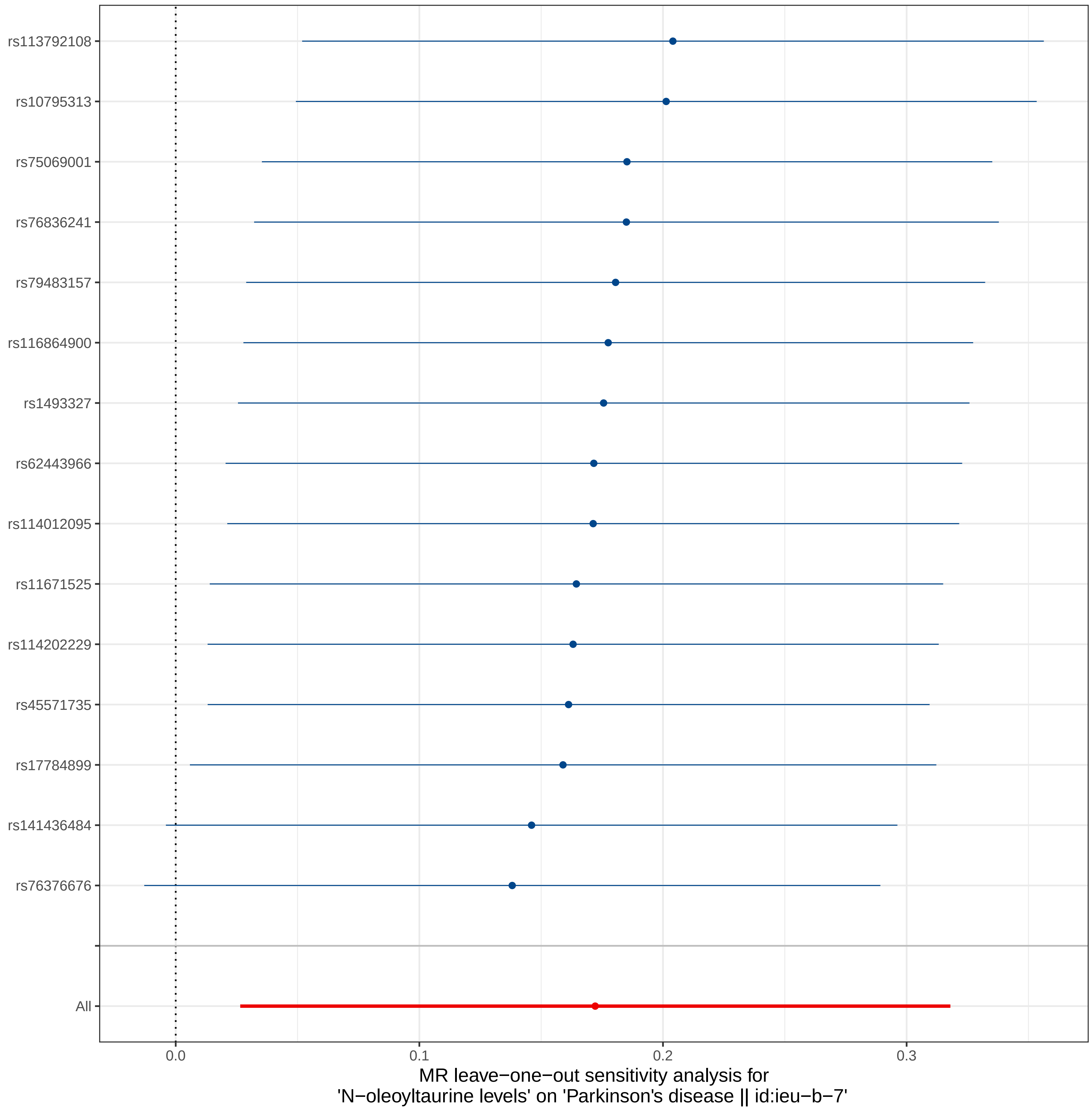

rs113792108
rs10795313
rs75069001
rs76836241
rs79483157
rs116864900
rs1493327
rs62443966
rs114012095
rs11671525
rs114202229
rs45571735
rs17784899
rs141436484
rs76376676
All
0.0
0.1
0.2
0.3
MR leave−one−out sensitivity analysis for
'N−oleoyltaurine levels' on 'Parkinson's disease || id:ieu−b−7'

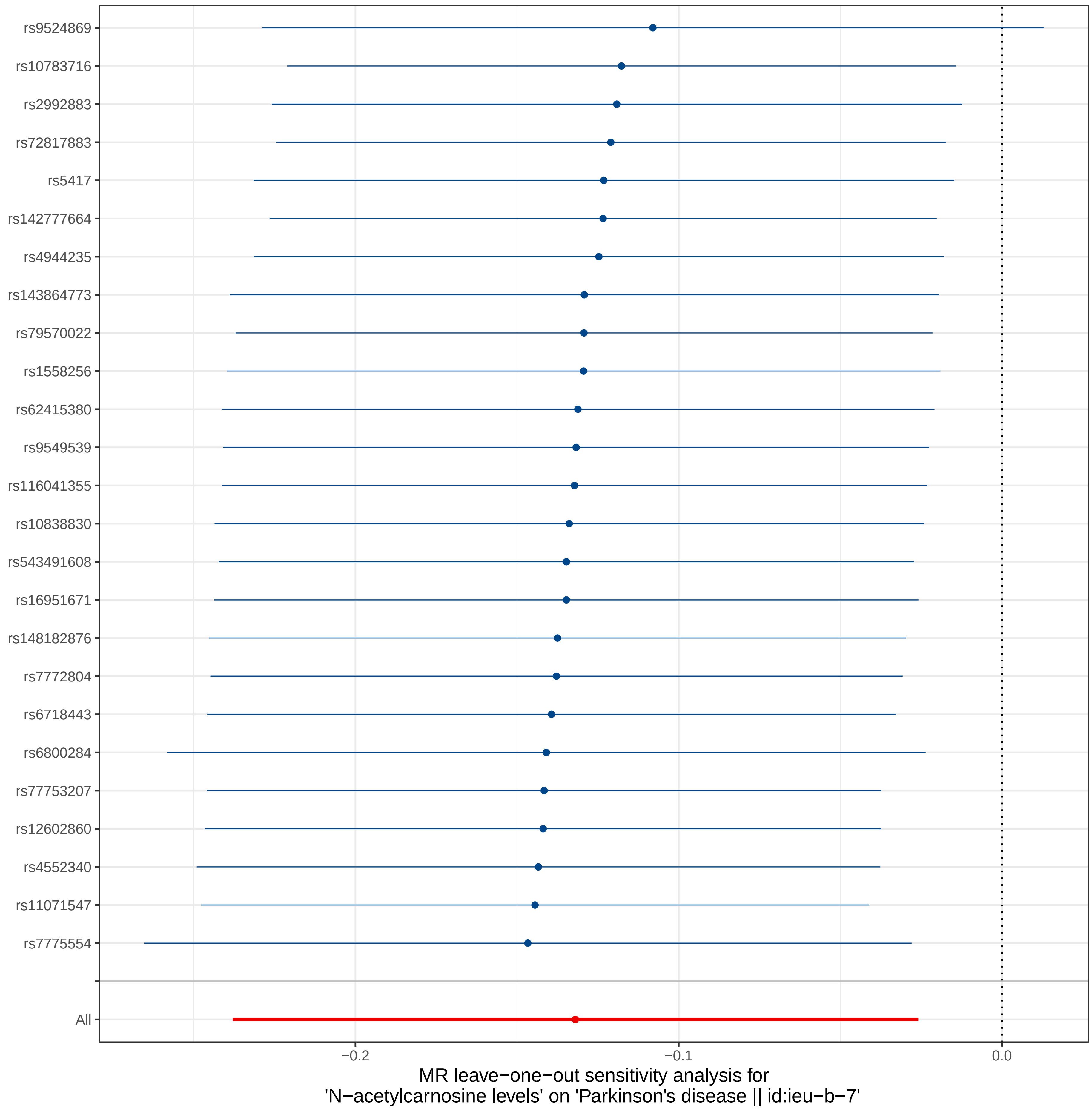
rs9524869
rs10783716
rs2992883
rs72817883
rs5417
rs142777664
rs4944235
rs143864773
rs79570022
rs1558256
rs62415380
rs9549539
rs116041355
rs10838830
rs543491608
rs16951671
rs148182876
rs7772804
rs6718443
rs6800284
rs77753207
rs12602860
rs4552340
rs11071547
rs7775554
All
−0.2
−0.1
0.0
MR leave−one−out sensitivity analysis for
'N−acetylcarnosine levels' on 'Parkinson's disease || id:ieu−b−7'

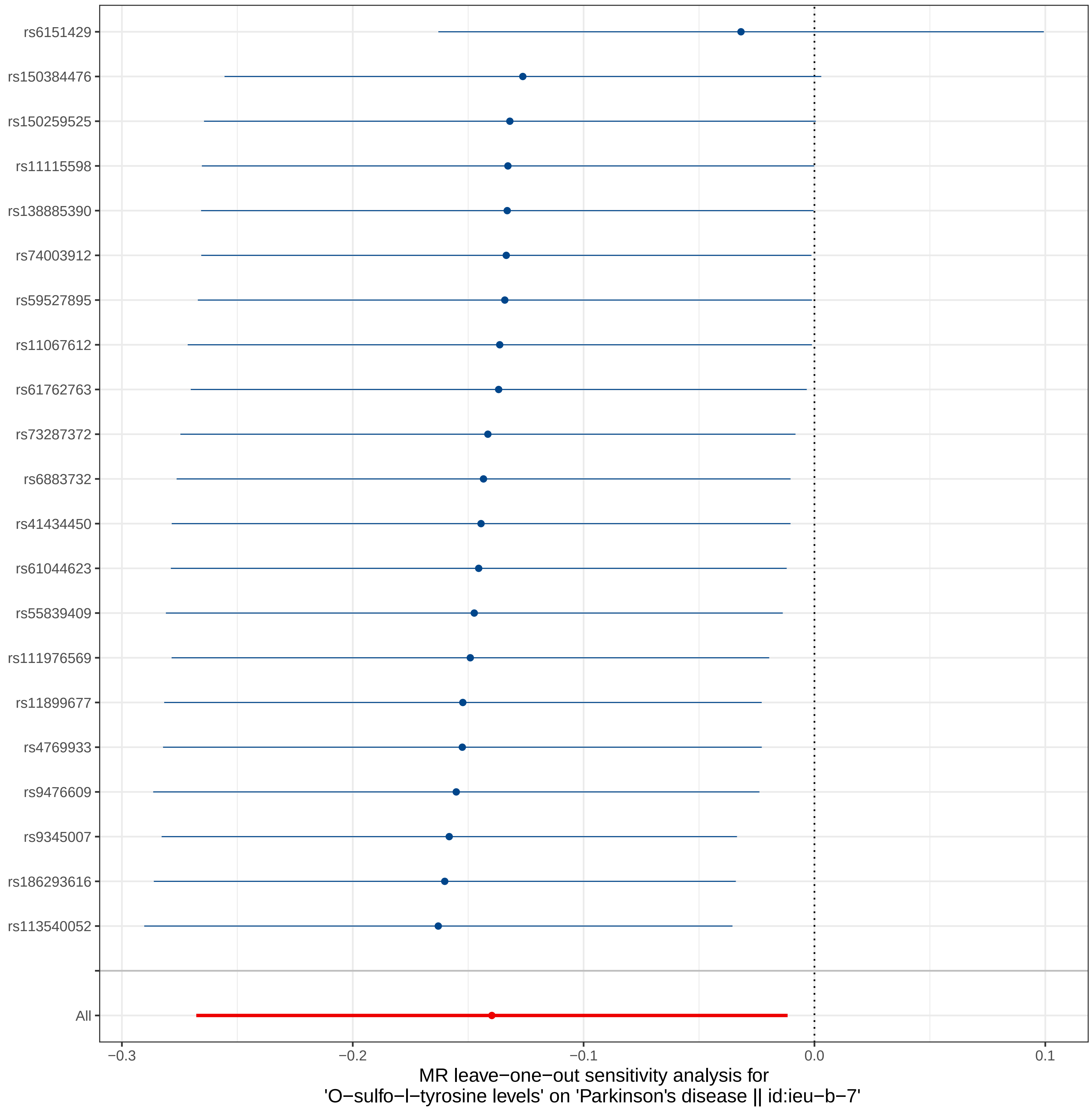

rs6151429
rs150384476
rs150259525
rs11115598
rs138885390
rs74003912
rs59527895
rs11067612
rs61762763
rs73287372
rs6883732
rs41434450
rs61044623
rs55839409
rs111976569
rs11899677
rs4769933
rs9476609
rs9345007
rs186293616
rs113540052
All
−0.3
−0.2
−0.1
0.0
0.1
MR leave−one−out sensitivity analysis for
'O−sulfo−l−tyrosine levels' on 'Parkinson's disease || id:ieu−b−7'

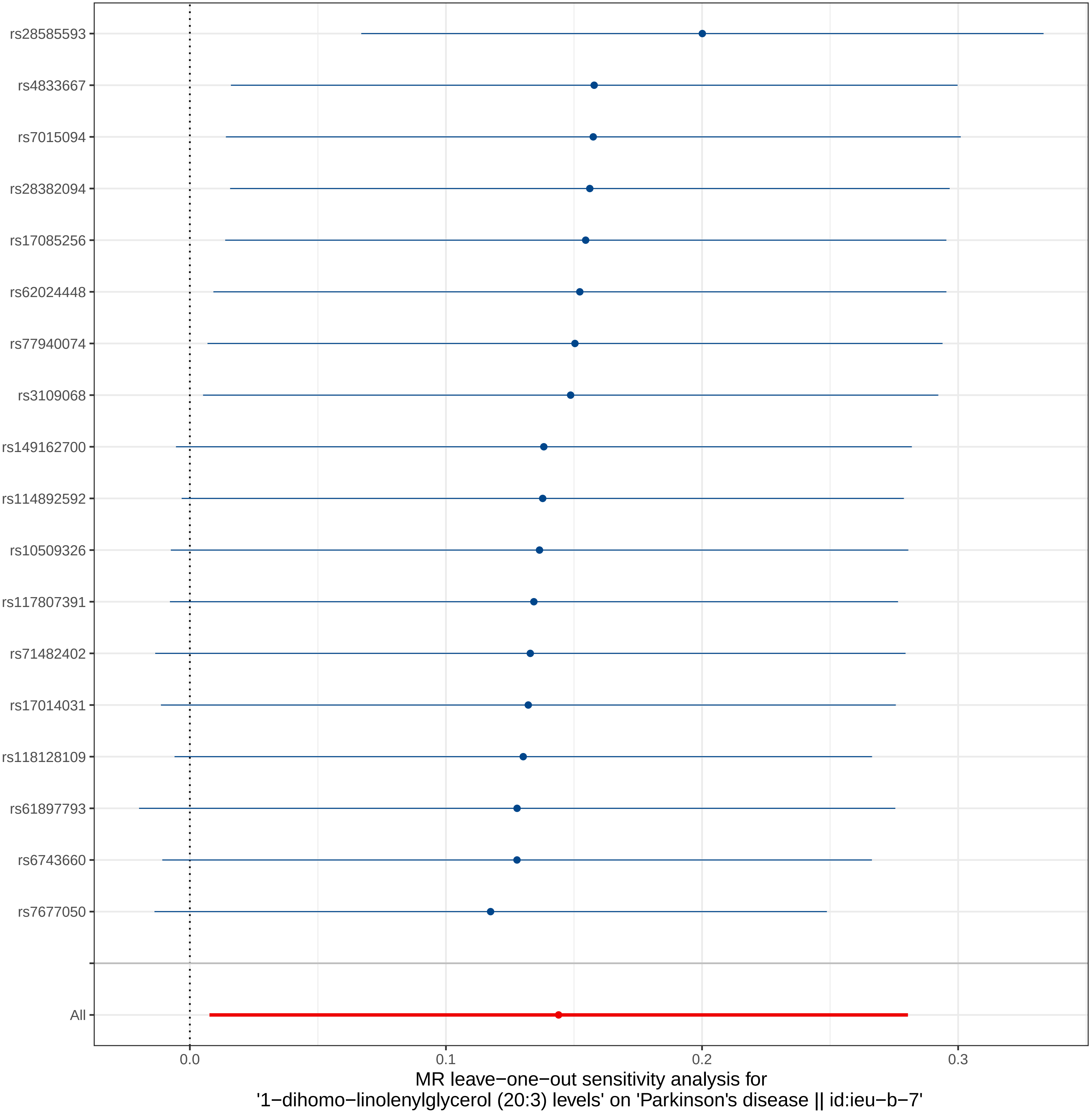
rs28585593
rs4833667
rs7015094
rs28382094
rs17085256
rs62024448
rs77940074
rs3109068
rs149162700
rs114892592
rs10509326
rs117807391
rs71482402
rs17014031
rs118128109
rs61897793
rs6743660
rs7677050
All
0.0
0.1
0.2
0.3
MR leave−one−out sensitivity analysis for
'1−dihomo−linolenylglycerol (20:3) levels' on 'Parkinson's disease || id:ieu−b−7'

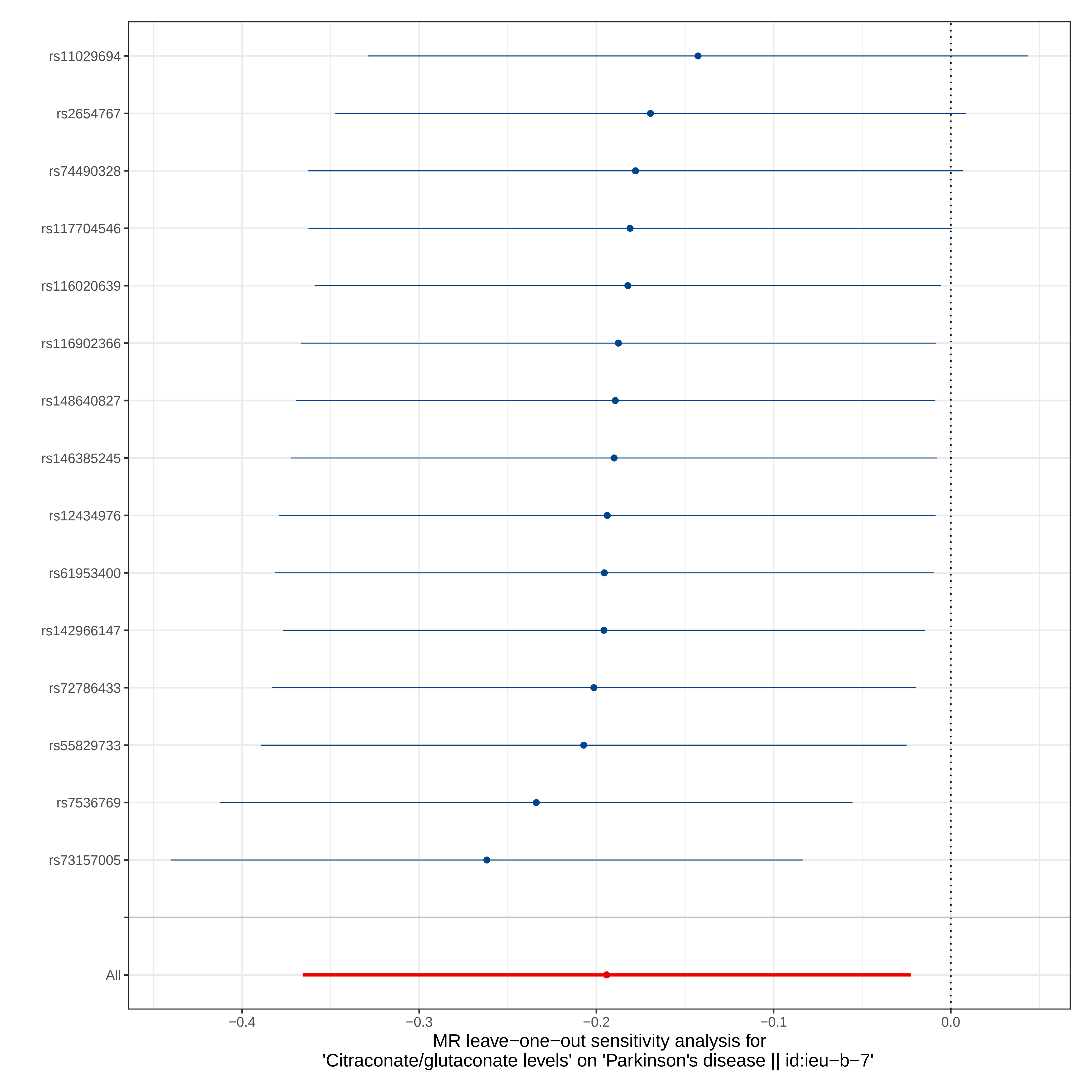
rs11029694
rs2654767
rs74490328
rs117704546
rs116020639
rs116902366
rs148640827
rs146385245
rs12434976
rs61953400
rs142966147
rs72786433
rs55829733
rs7536769
rs73157005
All
−0.4
−0.3
−0.2
−0.1
0.0
MR leave−one−out sensitivity analysis for
'Citraconate/glutaconate levels' on 'Parkinson's disease || id:ieu−b−7'

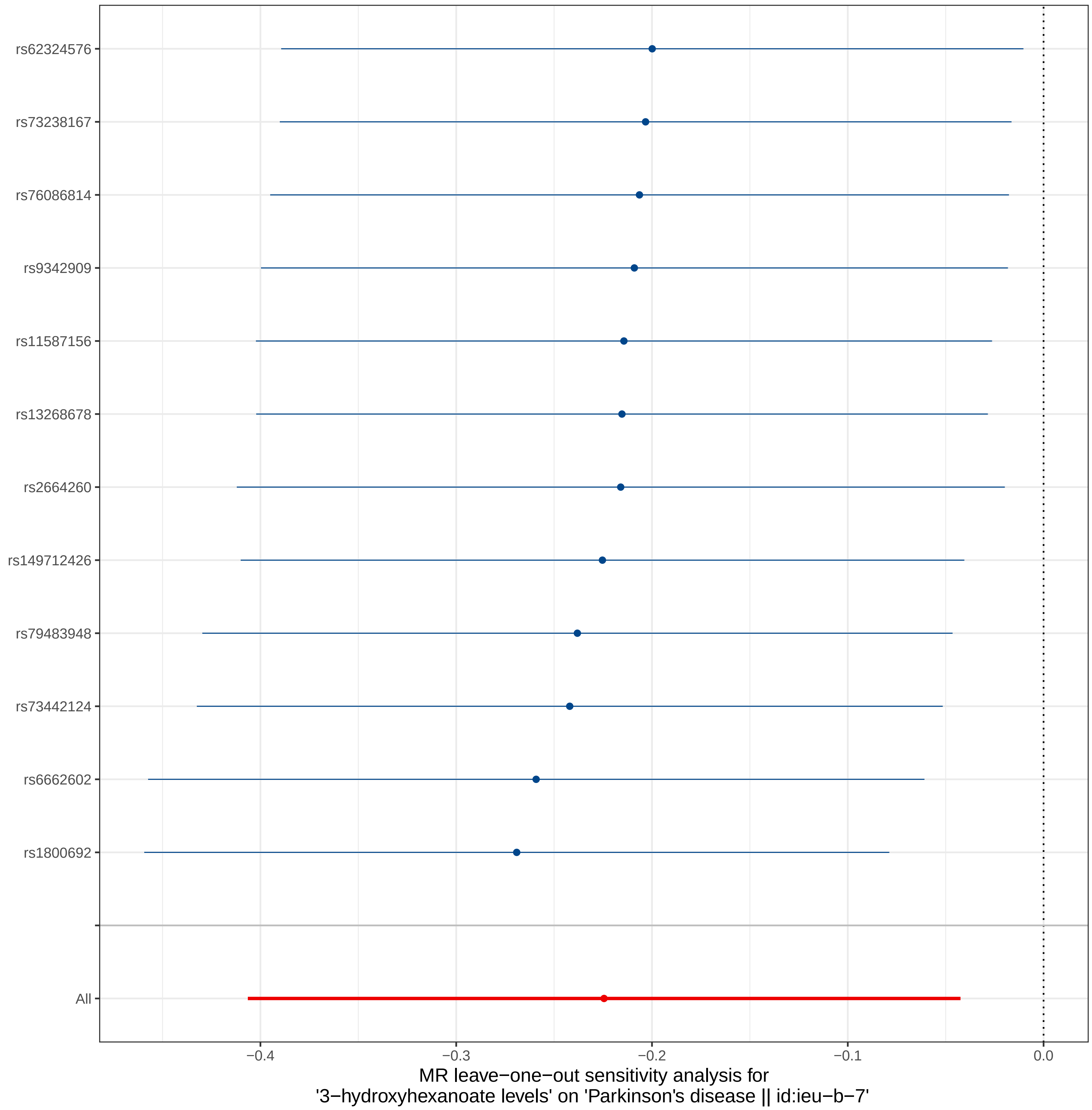

rs62324576
rs73238167
rs76086814
rs9342909
rs11587156
rs13268678
rs2664260
rs149712426
rs79483948
rs73442124
rs6662602
rs1800692
All
−0.4
−0.3
−0.2
−0.1
0.0
MR leave−one−out sensitivity analysis for
'3−hydroxyhexanoate levels' on 'Parkinson's disease || id:ieu−b−7'

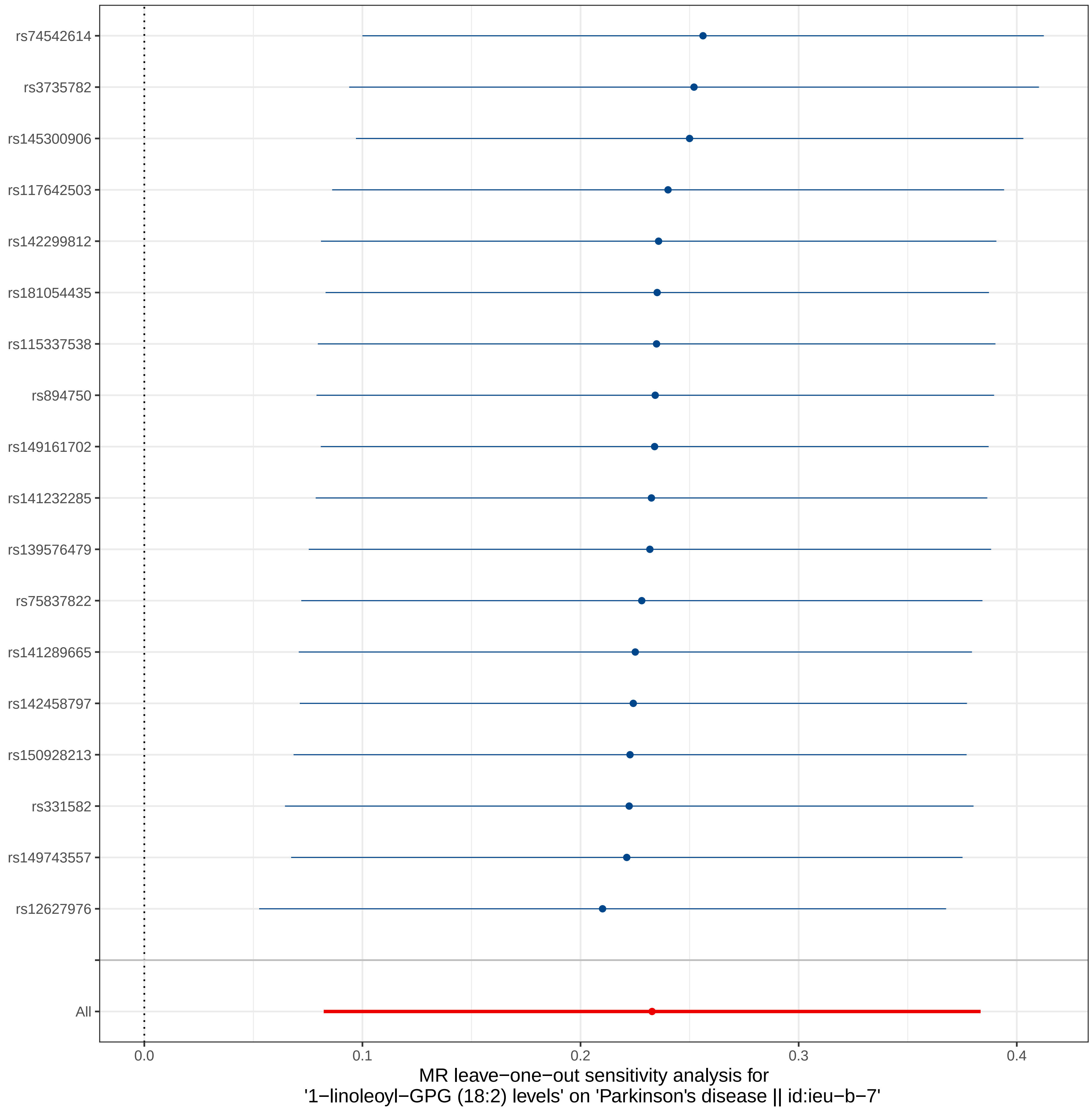
rs74542614
rs3735782
rs145300906
rs117642503
rs142299812
rs181054435
rs115337538
rs894750
rs149161702
rs141232285
rs139576479
rs75837822
rs141289665
rs142458797
rs150928213
rs331582
rs149743557
rs12627976
All
0.0
0.1
0.2
0.3
0.4
MR leave−one−out sensitivity analysis for
'1−linoleoyl−GPG (18:2) levels' on 'Parkinson's disease || id:ieu−b−7'

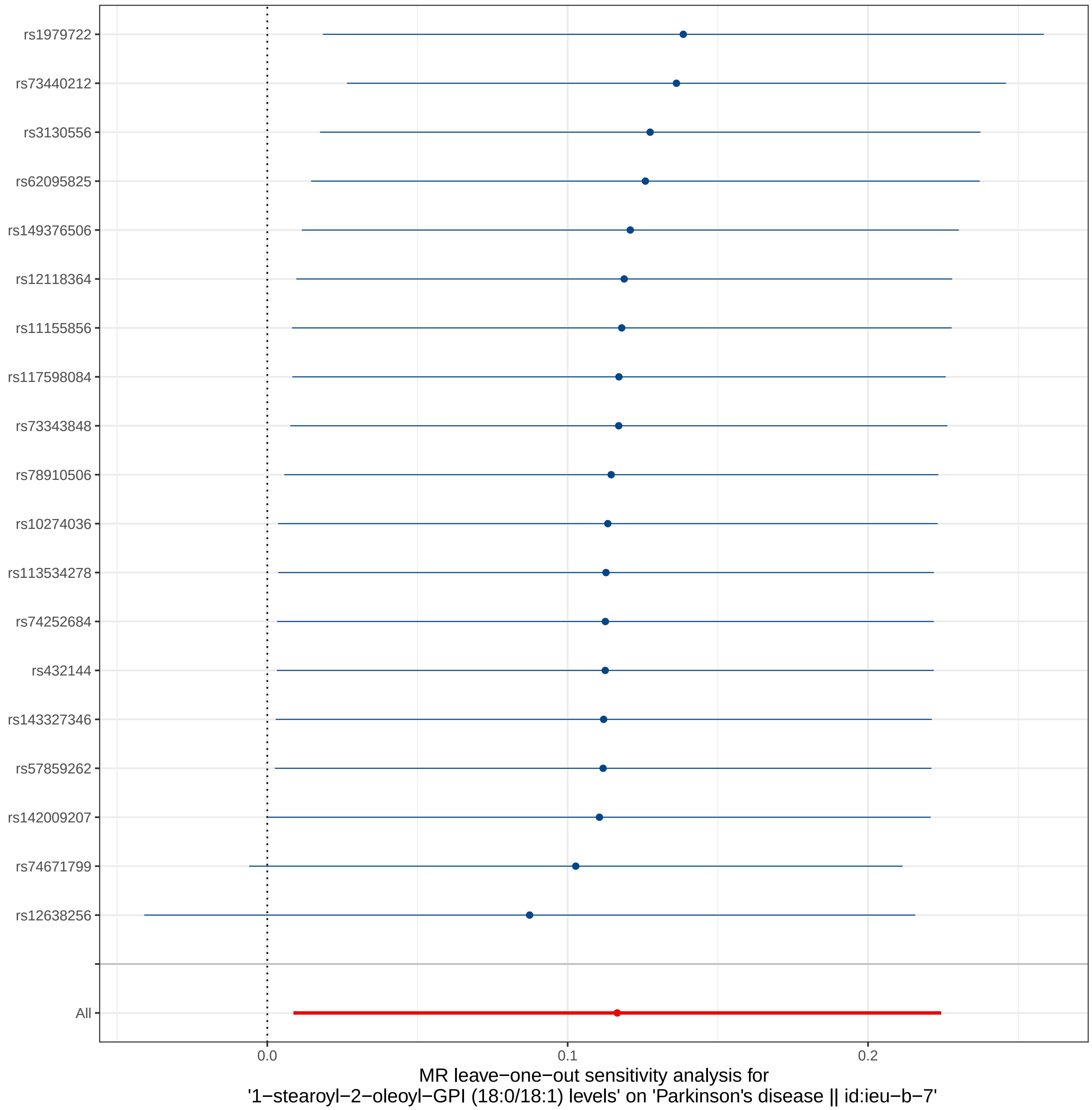
rs1979722
rs73440212
rs3130556
rs62095825
rs149376506
rs12118364
rs11155856
rs117598084
rs73343848
rs78910506
rs10274036
rs113534278
rs74252684
rs432144
rs143327346
rs57859262
rs142009207
rs74671799
rs12638256
All
0.0
0.1
0.2
MR leave−one−out sensitivity analysis for
'1−stearoyl−2−oleoyl−GPI (18:0/18:1) levels' on 'Parkinson's disease || id:ieu−b−7'

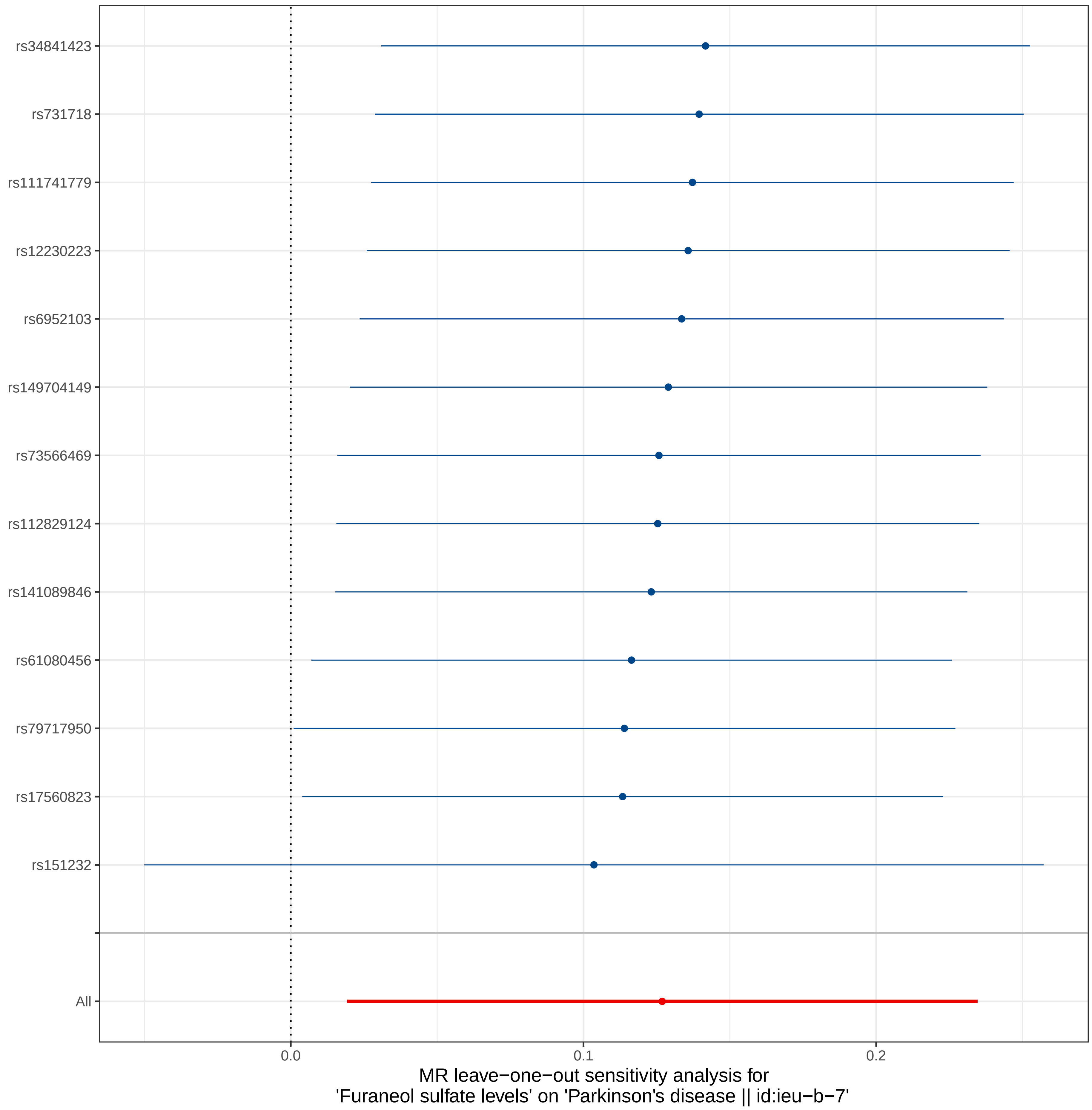
rs34841423
rs731718
rs111741779
rs12230223
rs6952103
rs149704149
rs73566469
rs112829124
rs141089846
rs61080456
rs79717950
rs17560823
rs151232
All
0.0
0.1
0.2
MR leave−one−out sensitivity analysis for
'Furaneol sulfate levels' on 'Parkinson's disease || id:ieu−b−7'

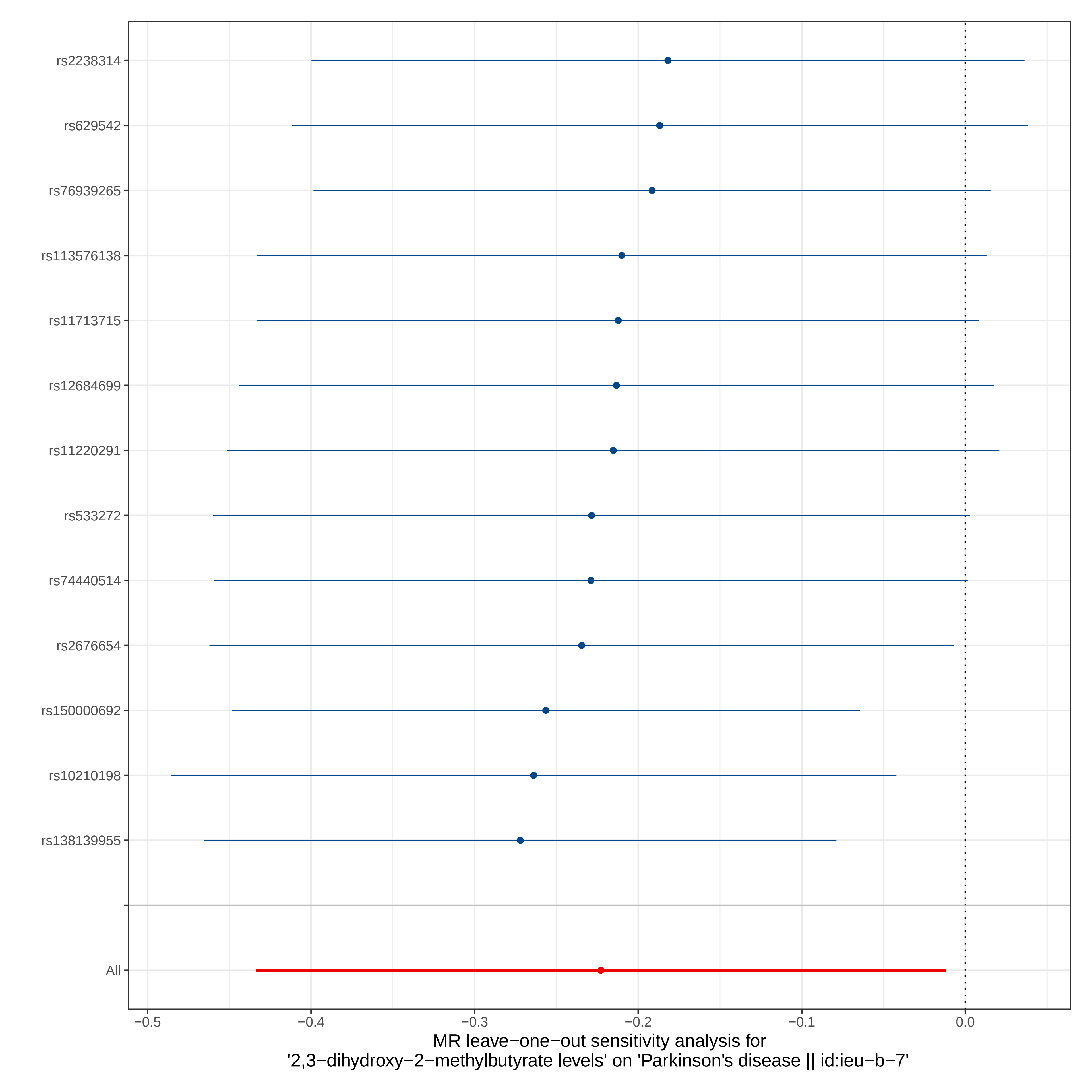
rs2238314
rs629542
rs76939265
rs113576138
rs11713715
rs12684699
rs11220291
rs533272
rs74440514
rs2676654
rs150000692
rs10210198
rs138139955
All
−0.5
−0.4
−0.3
−0.2
−0.1
0.0
MR leave−one−out sensitivity analysis for
'2,3−dihydroxy−2−methylbutyrate levels' on 'Parkinson's disease || id:ieu−b−7'

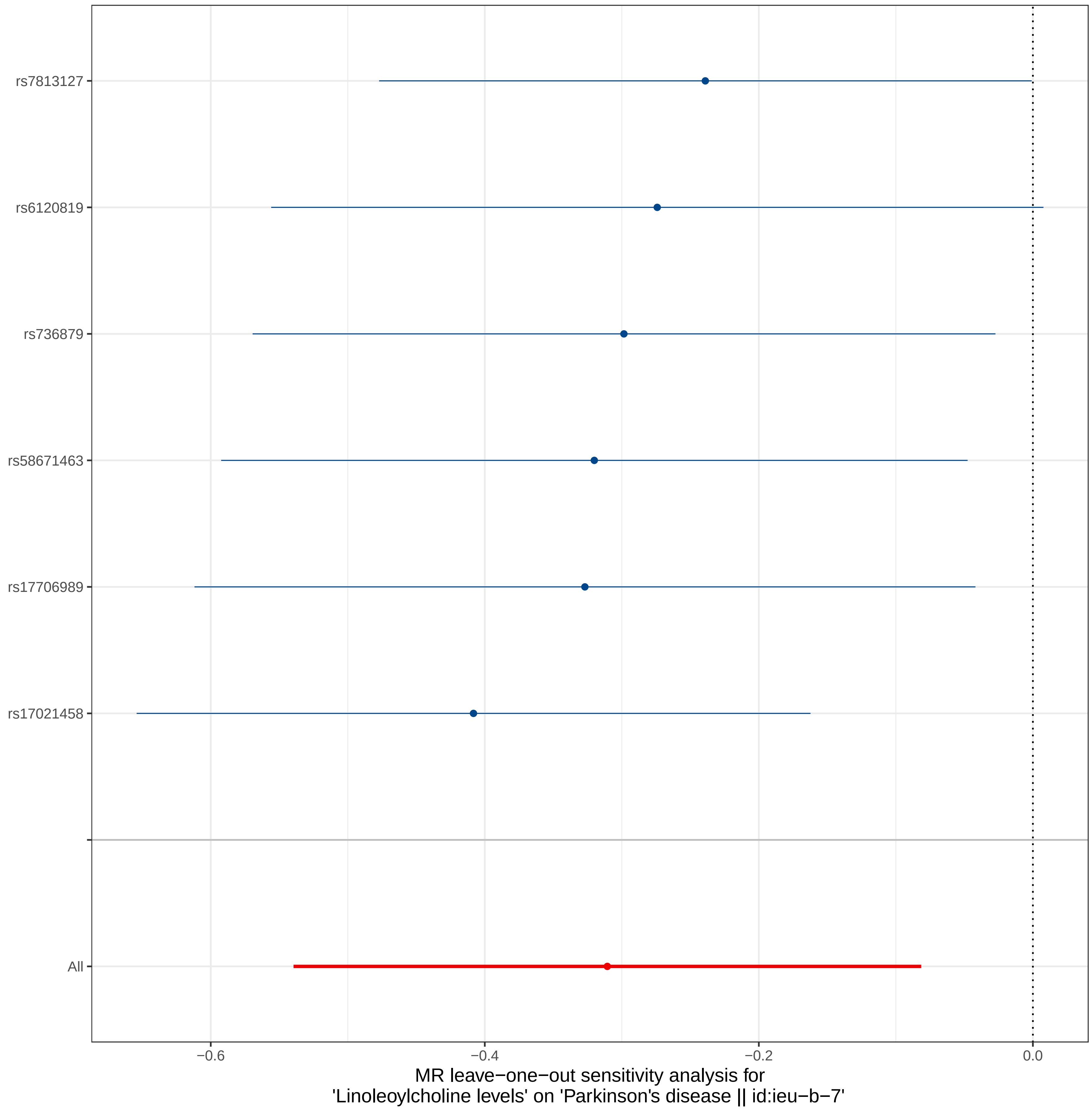
rs7813127
rs6120819
rs736879
rs58671463
rs17706989
rs17021458
All
−0.6
−0.4
−0.2
0.0
MR leave−one−out sensitivity analysis for
'Linoleoylcholine levels' on 'Parkinson's disease || id:ieu−b−7'

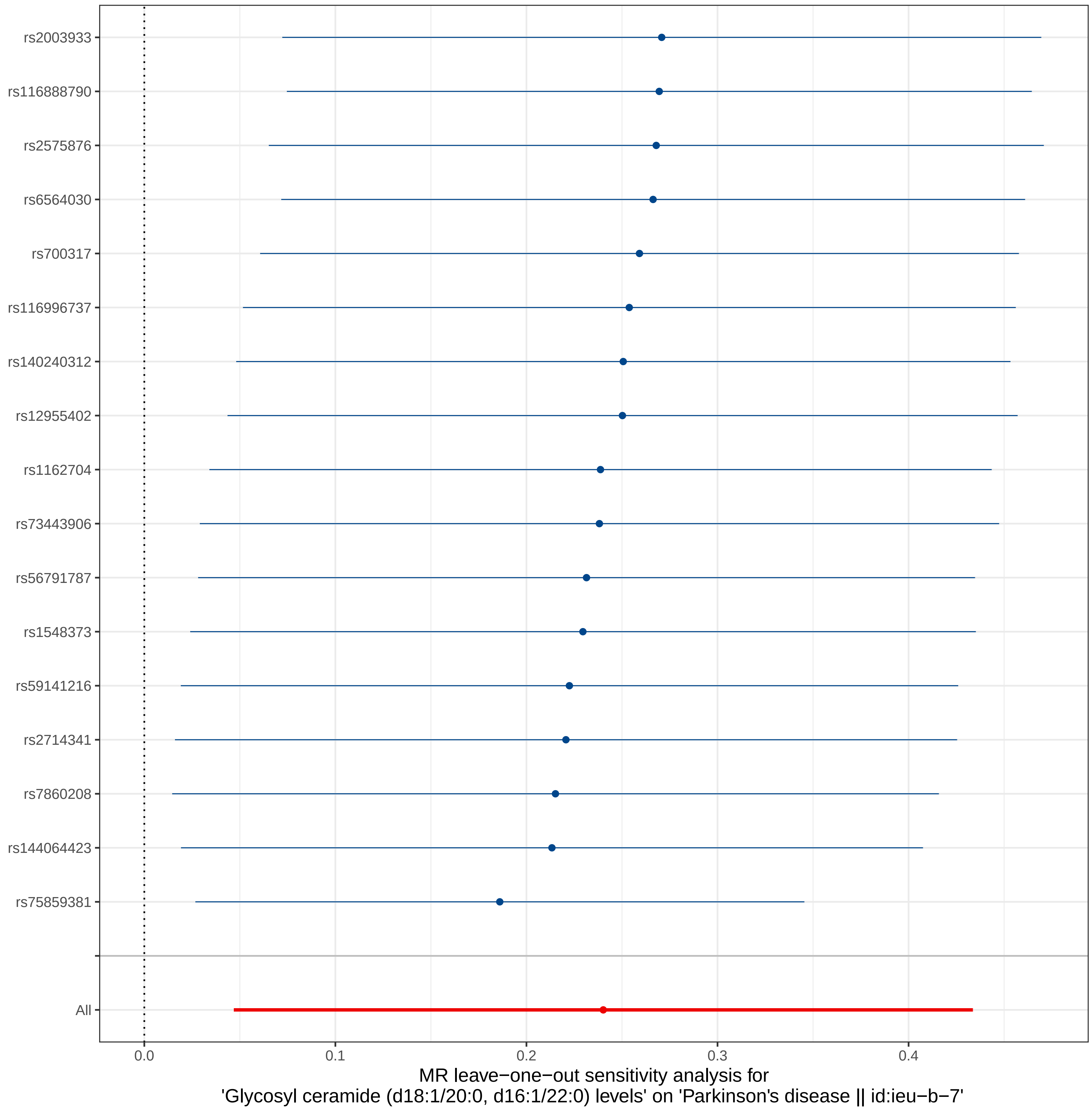

rs2003933
rs116888790
rs2575876
rs6564030
rs700317
rs116996737
rs140240312
rs12955402
rs1162704
rs73443906
rs56791787
rs1548373
rs59141216
rs2714341
rs7860208
rs144064423
rs75859381
All
0.0
0.1
0.2
0.3
0.4
MR leave−one−out sensitivity analysis for
'Glycosyl ceramide (d18:1/20:0, d16:1/22:0) levels' on 'Parkinson's disease || id:ieu−b−7'

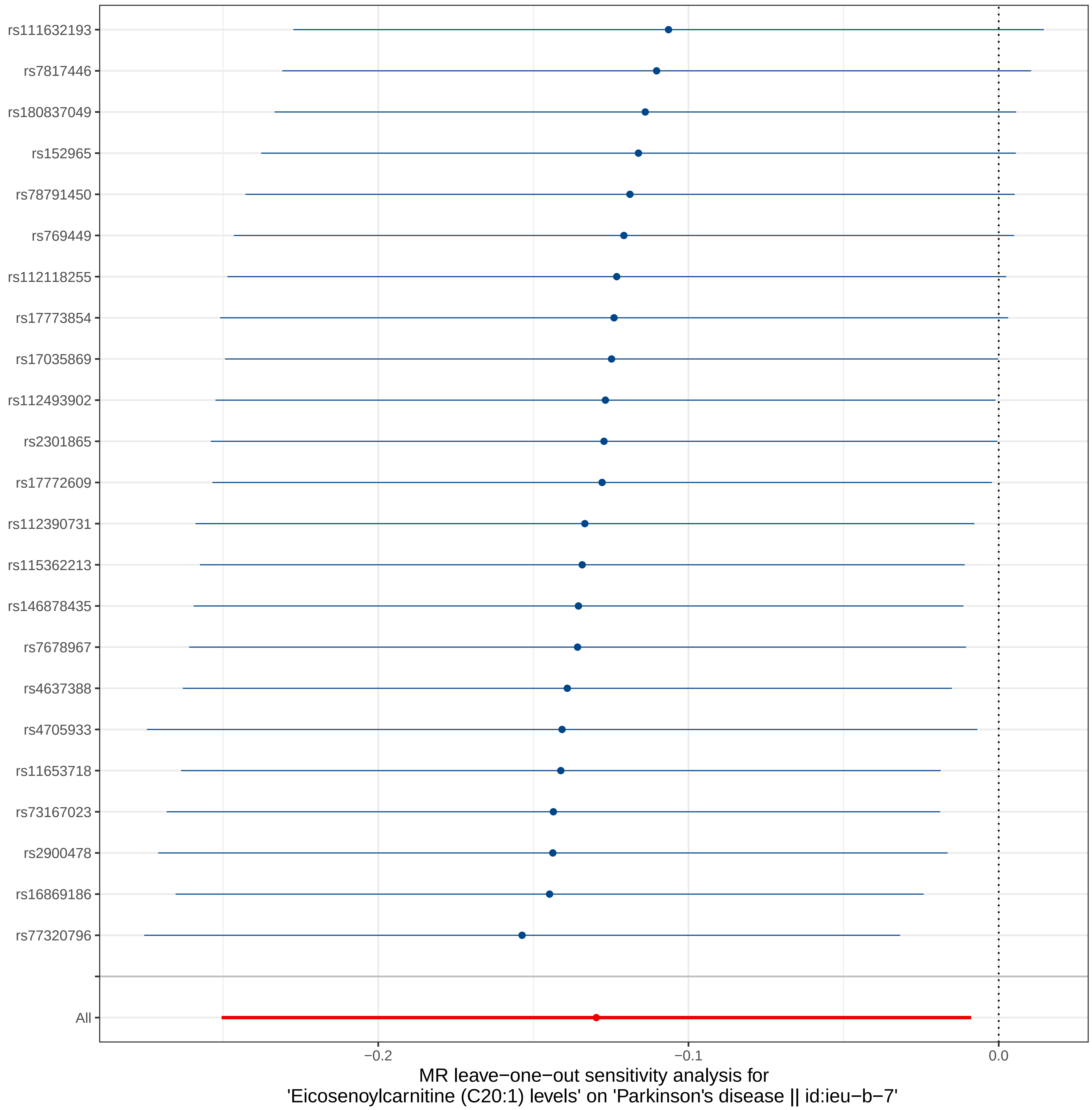

rs111632193
rs7817446
rs180837049
rs152965
rs78791450
rs769449
rs112118255
rs17773854
rs17035869
rs112493902
rs2301865
rs17772609
rs112390731
rs115362213
rs146878435
rs7678967
rs4637388
rs4705933
rs11653718
rs73167023
rs2900478
rs16869186
rs77320796
All
−0.2
−0.1
0.0
MR leave−one−out sensitivity analysis for
'Eicosenoylcarnitine (C20:1) levels' on 'Parkinson's disease || id:ieu−b−7'

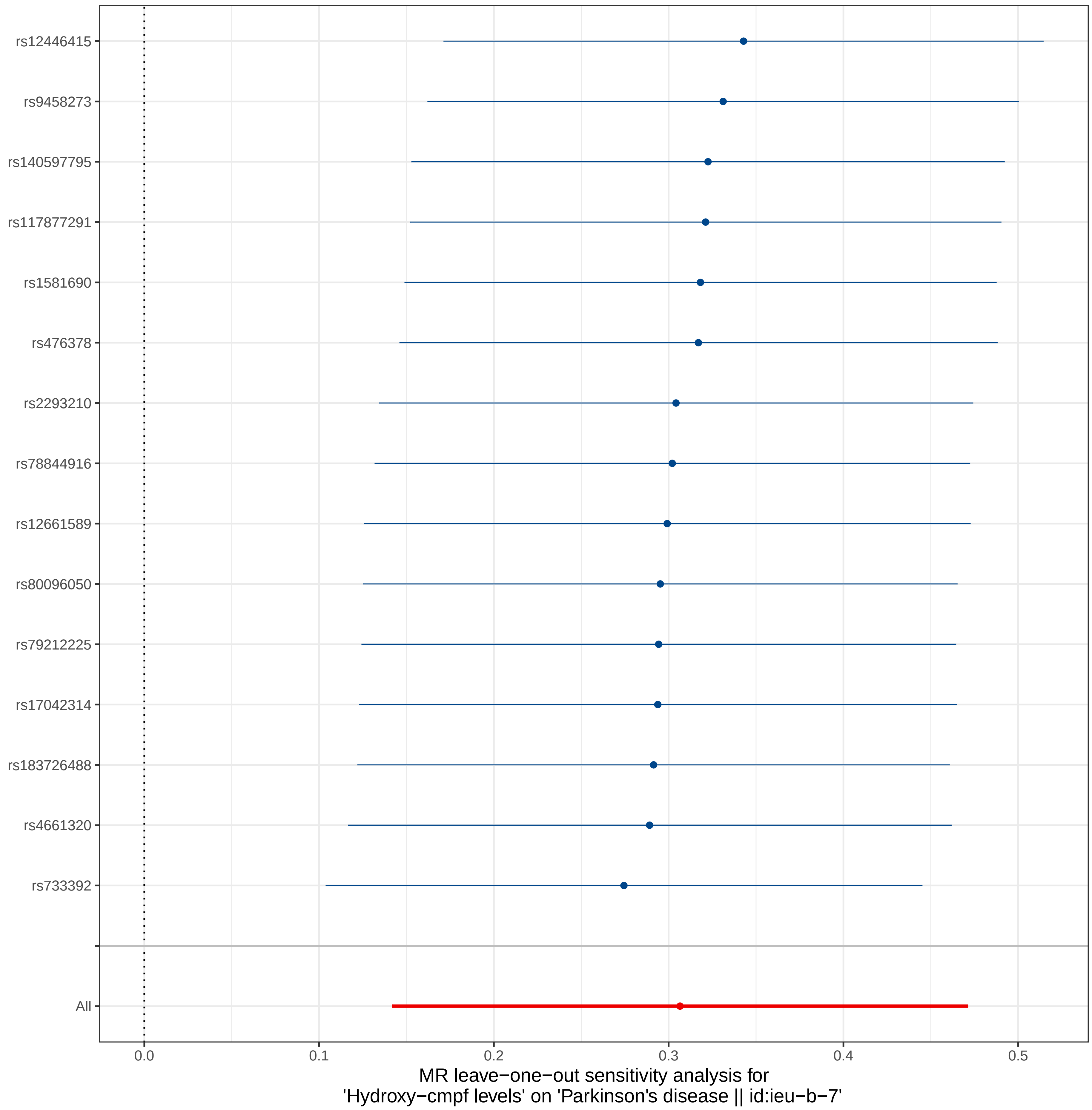
rs12446415
rs9458273
rs140597795
rs117877291
rs1581690
rs476378
rs2293210
rs78844916
rs12661589
rs80096050
rs79212225
rs17042314
rs183726488
rs4661320
rs733392
All
0.0
0.1
0.2
0.3
0.4
0.5
MR leave−one−out sensitivity analysis for
'Hydroxy−cmpf levels' on 'Parkinson's disease || id:ieu−b−7'

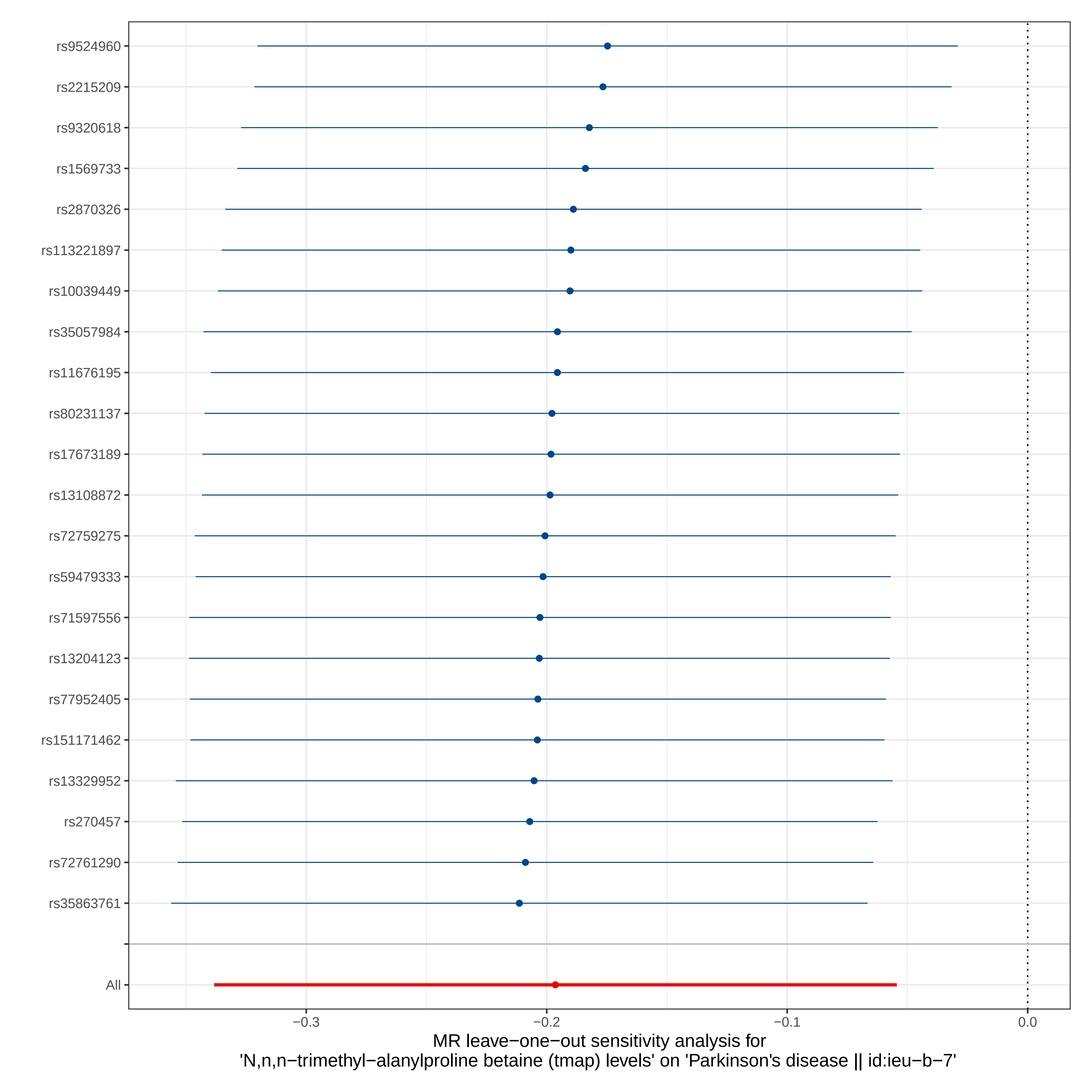
rs9524960
rs2215209
rs9320618
rs1569733
rs2870326
rs113221897
rs10039449
rs35057984
rs11676195
rs80231137
rs17673189
rs13108872
rs72759275
rs59479333
rs71597556
rs13204123
rs77952405
rs151171462
rs13329952
rs270457
rs72761290
rs35863761
All
−0.3
−0.2
−0.1
0.0
MR leave−one−out sensitivity analysis for
'N,n,n−trimethyl−alanylproline betaine (tmap) levels' on 'Parkinson's disease || id:ieu−b−7'

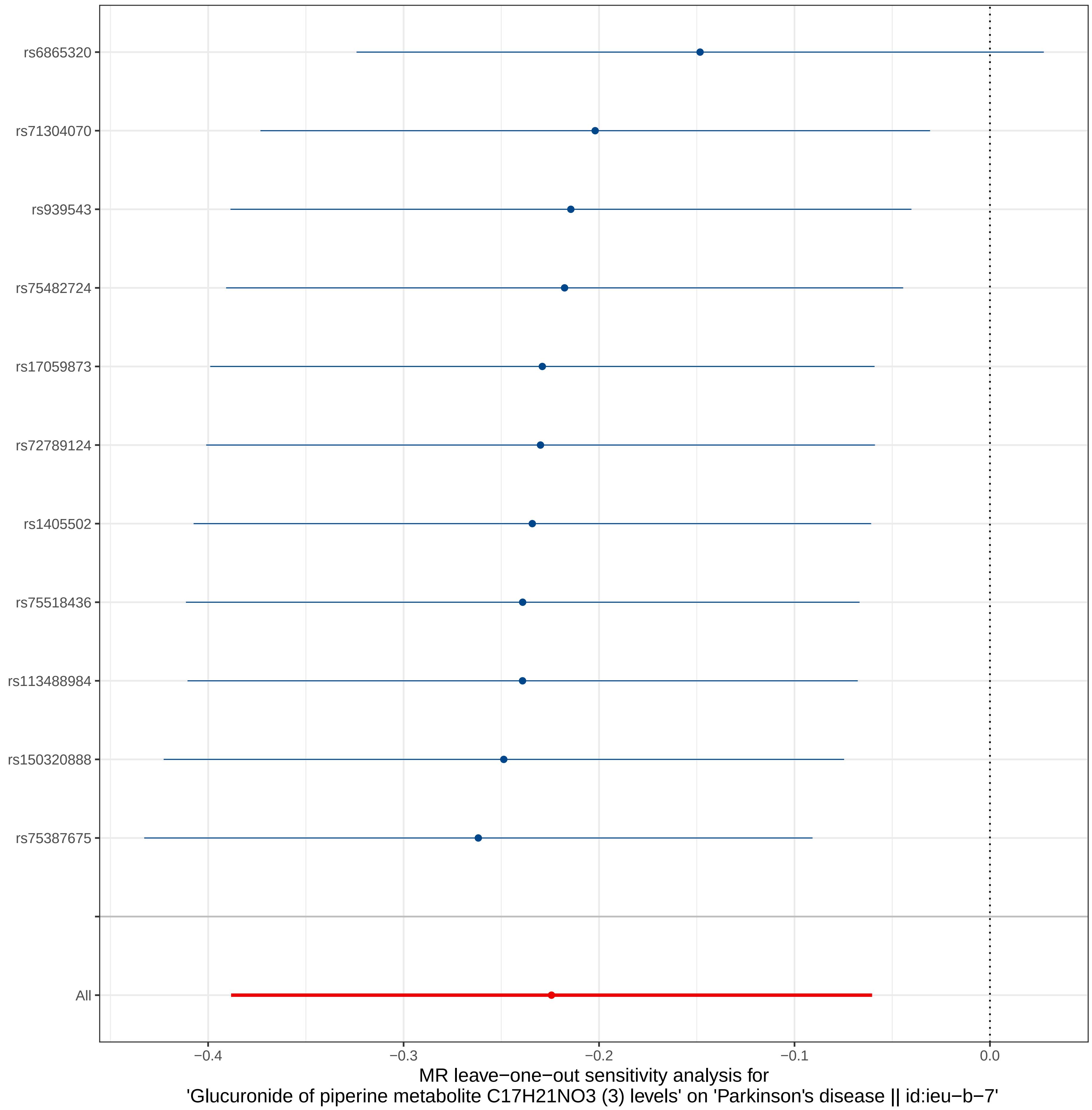

rs6865320
rs71304070
rs939543
rs75482724
rs17059873
rs72789124
rs1405502
rs75518436
rs113488984
rs150320888
rs75387675
All
−0.4
−0.3
−0.2
−0.1
0.0
MR leave−one−out sensitivity analysis for
'Glucuronide of piperine metabolite C17H21NO3 (3) levels' on 'Parkinson's disease || id:ieu−b−7'

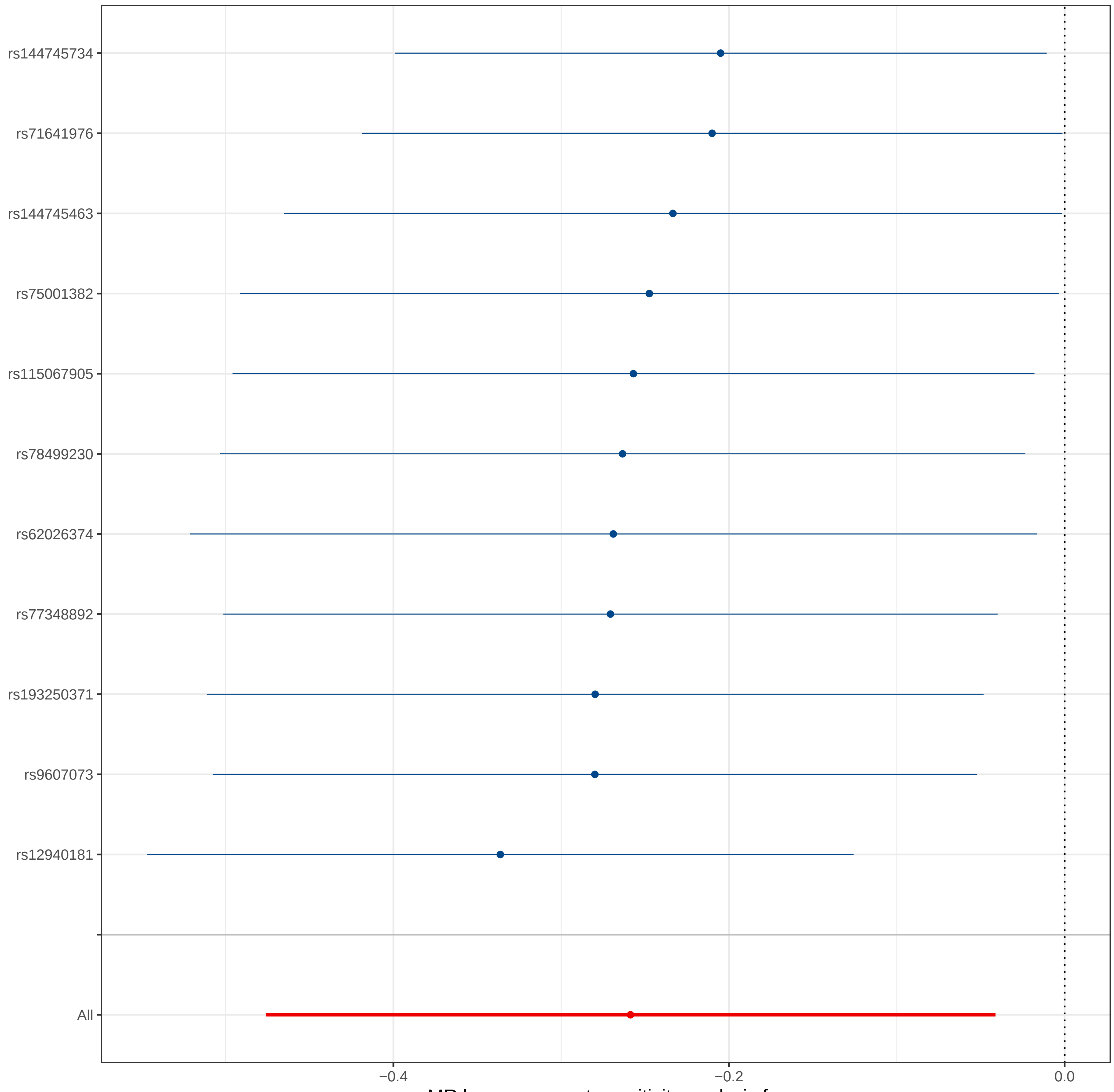

MR leave−one−out sensitivity analysis for
'3−hydroxy−2−methylpyridine sulfate levels' on 'Parkinson's disease || id:ieu−b−7'

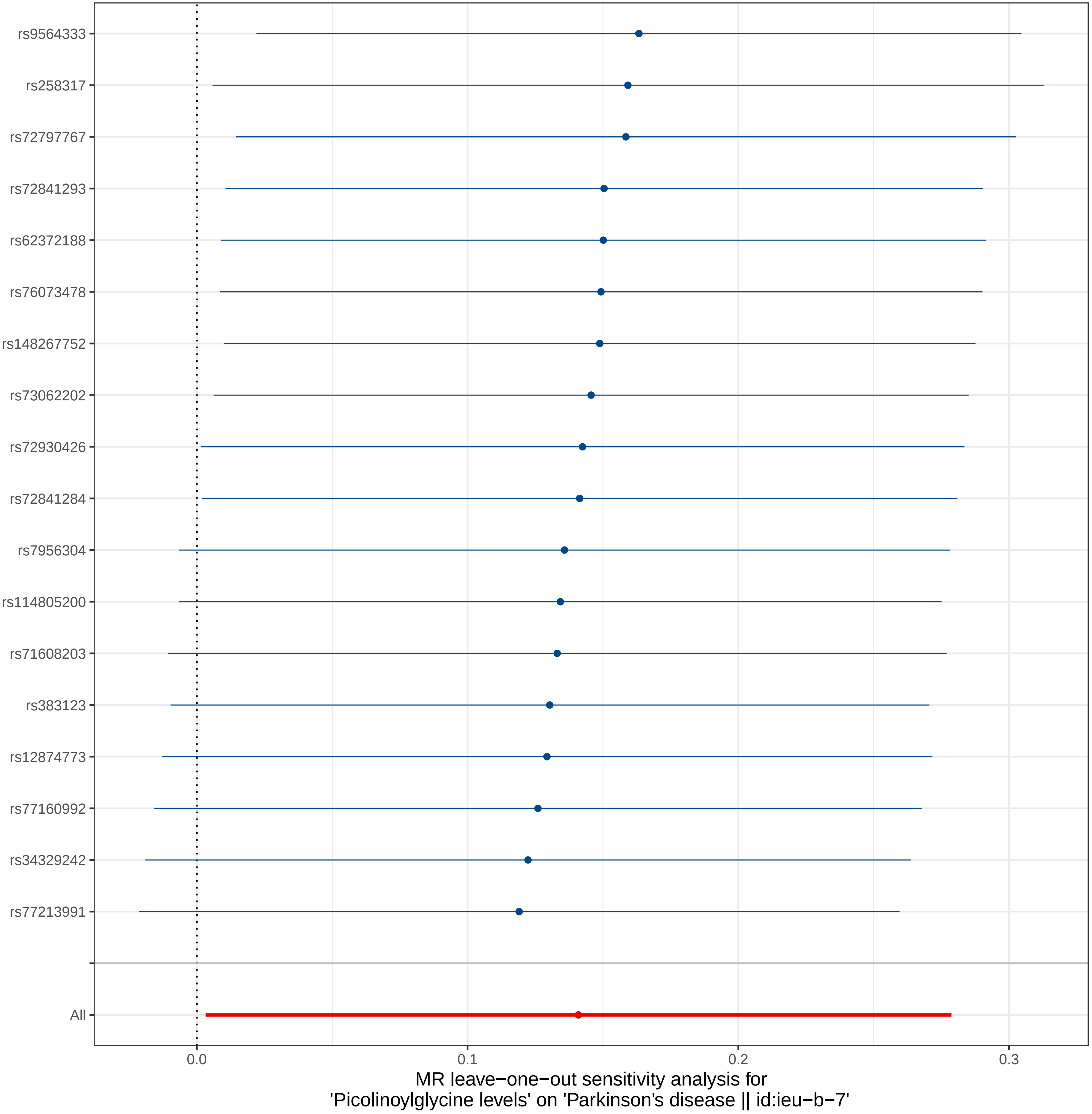

rs9564333
rs258317
rs72797767
rs72841293
rs62372188
rs76073478
rs148267752
rs73062202
rs72930426
rs72841284
rs7956304
rs114805200
rs71608203
rs383123
rs12874773
rs77160992
rs34329242
rs77213991
All
0.0
0.1
0.2
0.3
MR leave−one−out sensitivity analysis for
'Picolinoylglycine levels' on 'Parkinson's disease || id:ieu−b−7'

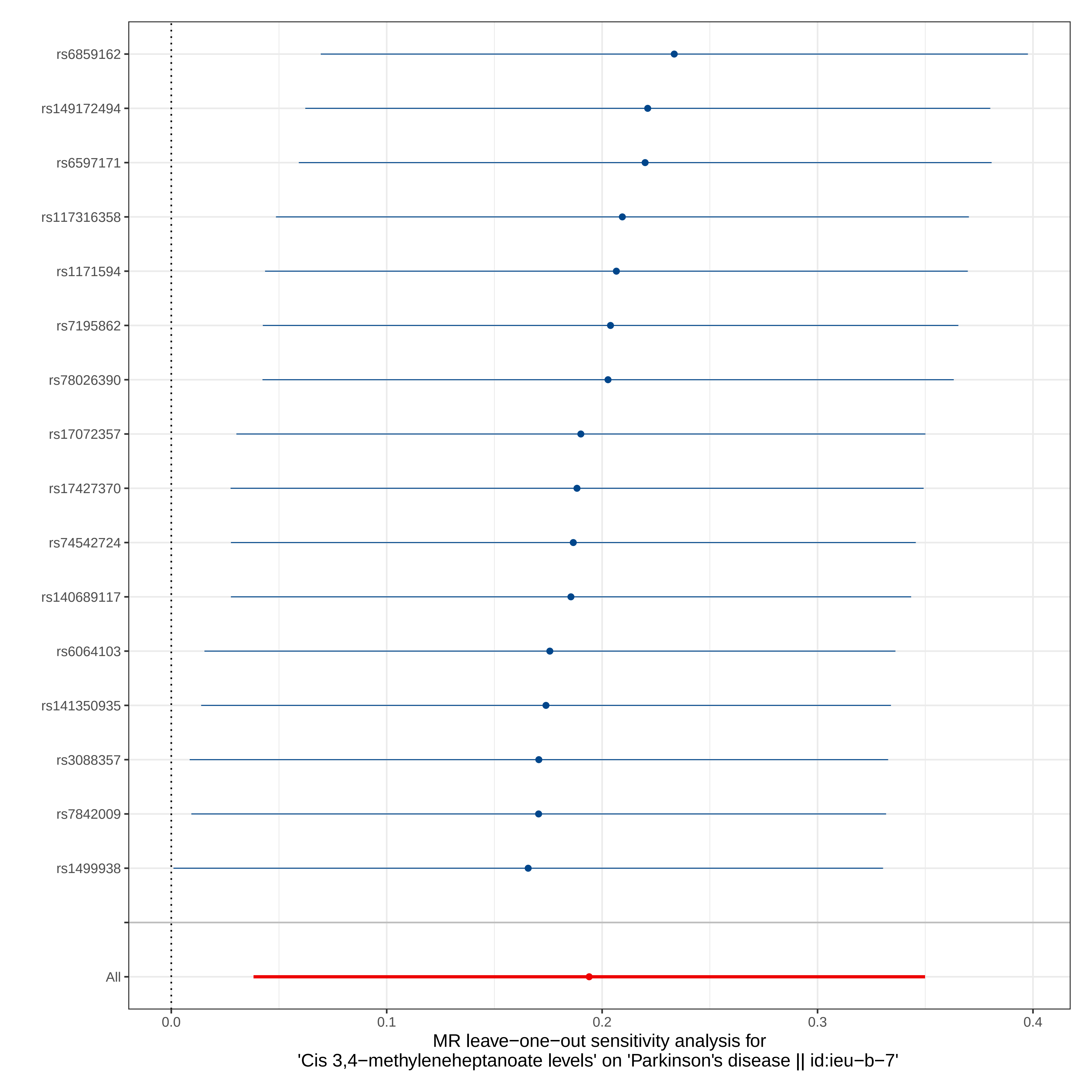
rs6859162
rs149172494
rs6597171
rs117316358
rs1171594
rs7195862
rs78026390
rs17072357
rs17427370
rs74542724
rs140689117
rs6064103
rs141350935
rs3088357
rs7842009
rs1499938
All
0.0
0.1
0.2
0.3
0.4
MR leave−one−out sensitivity analysis for
'Cis 3,4−methyleneheptanoate levels' on 'Parkinson's disease || id:ieu−b−7'

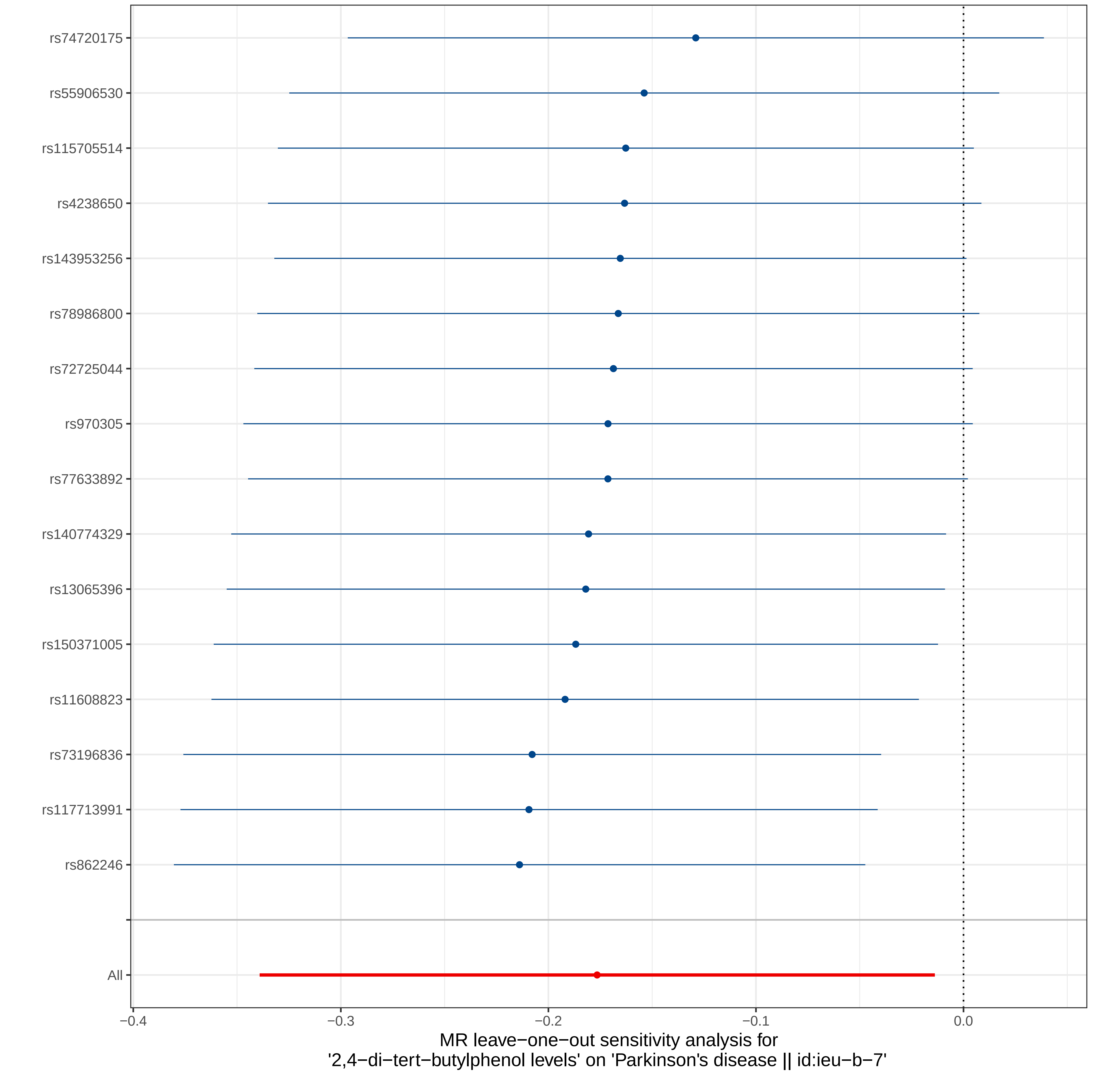

rs74720175
rs55906530
rs115705514
rs4238650
rs143953256
rs78986800
rs72725044
rs970305
rs77633892
rs140774329
rs13065396
rs150371005
rs11608823
rs73196836
rs117713991
rs862246
All
−0.4
−0.3
−0.2
−0.1
0.0
MR leave−one−out sensitivity analysis for
'2,4−di−tert−butylphenol levels' on 'Parkinson's disease || id:ieu−b−7'

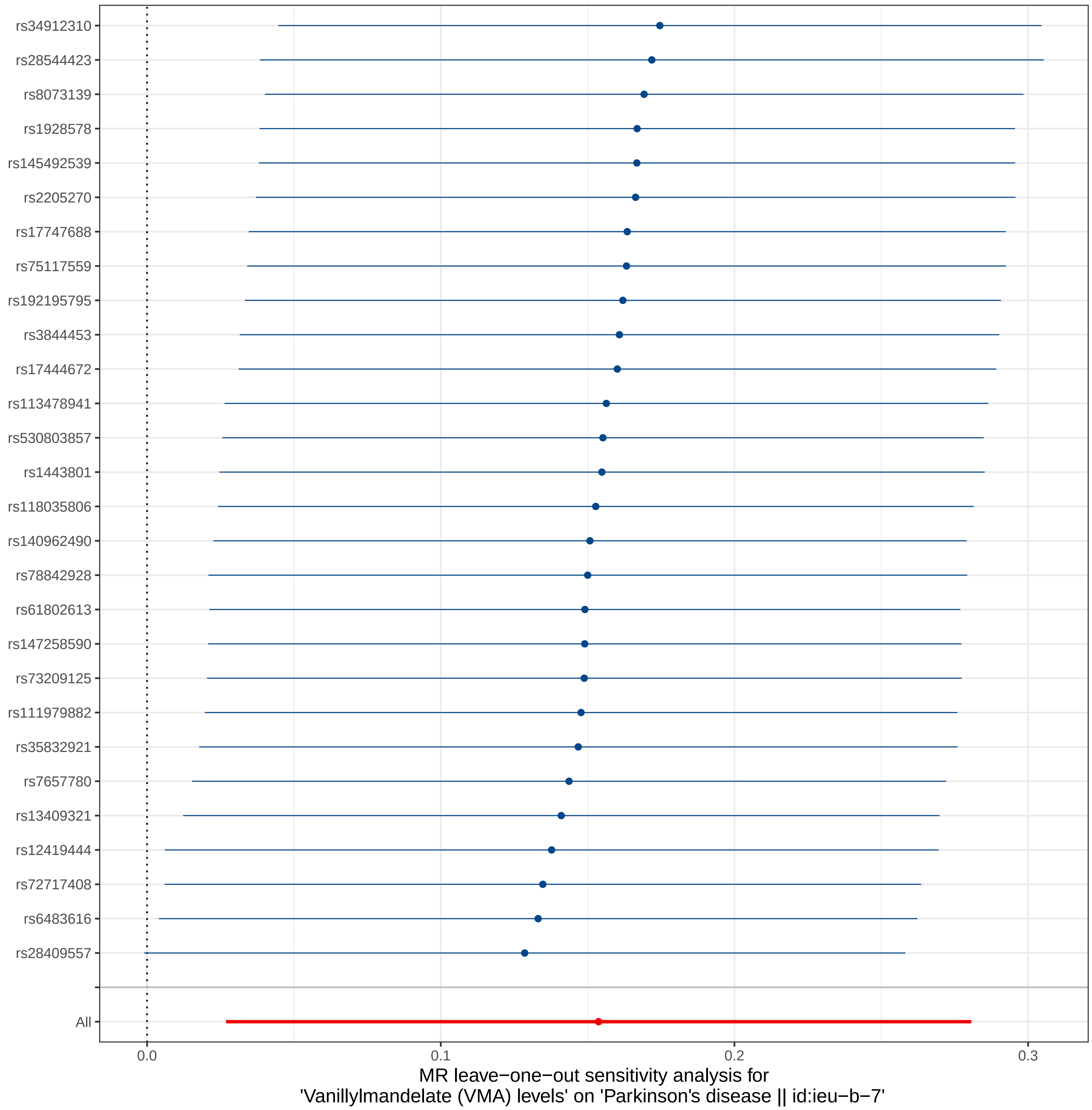
rs34912310
rs28544423
rs8073139
rs1928578
rs145492539
rs2205270
rs17747688
rs75117559
rs192195795
rs3844453
rs17444672
rs113478941
rs530803857
rs1443801
rs118035806
rs140962490
rs78842928
rs61802613
rs147258590
rs73209125
rs111979882
rs35832921
rs7657780
rs13409321
rs12419444
rs72717408
rs6483616
rs28409557
All
0.0
0.1
0.2
0.3
MR leave−one−out sensitivity analysis for
'Vanillylmandelate (VMA) levels' on 'Parkinson's disease || id:ieu−b−7'

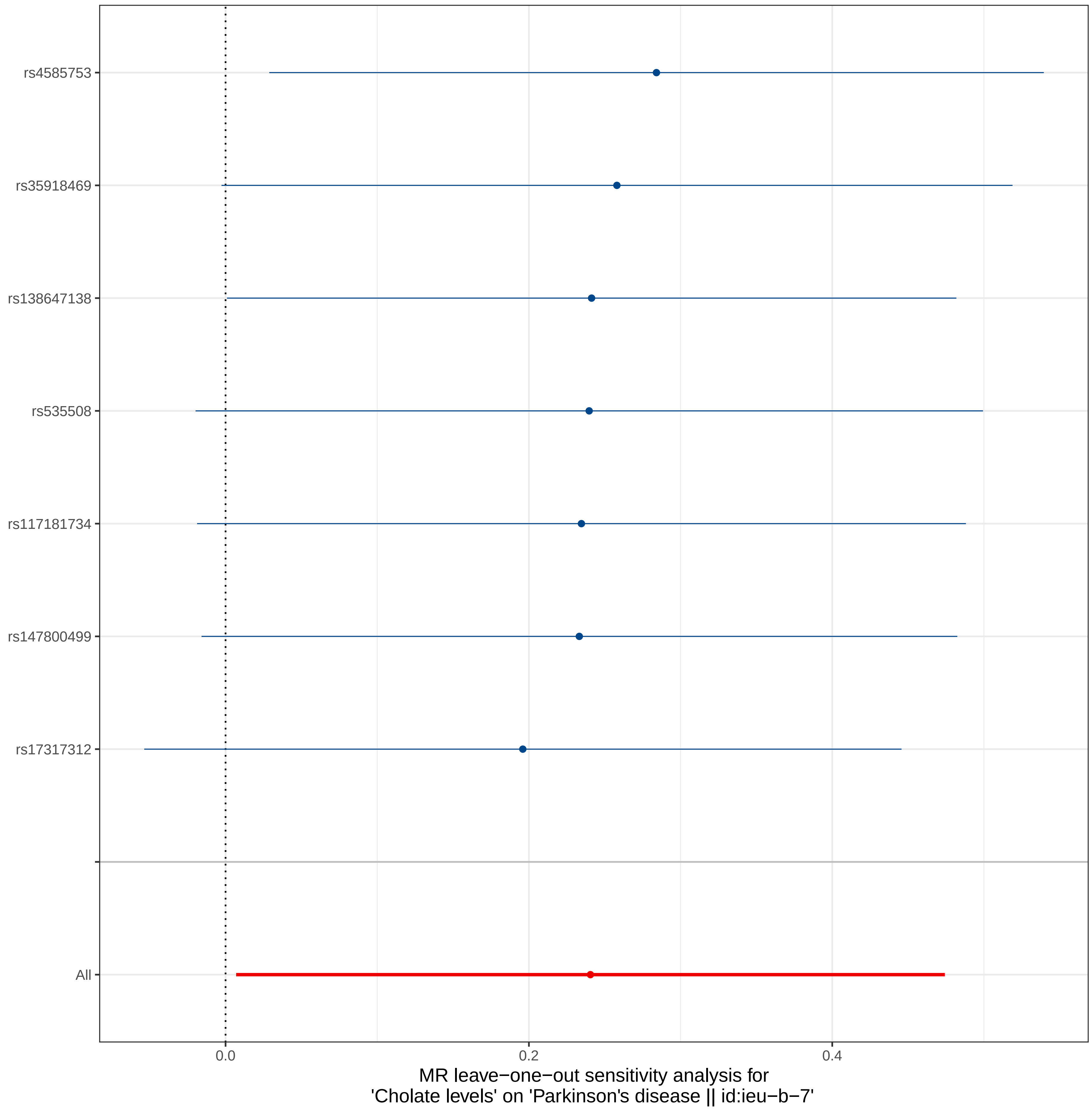

rs4585753
rs35918469
rs138647138
rs535508
rs117181734
rs147800499
rs17317312
All
0.0
0.2
0.4
MR leave−one−out sensitivity analysis for
'Cholate levels' on 'Parkinson's disease || id:ieu−b−7'

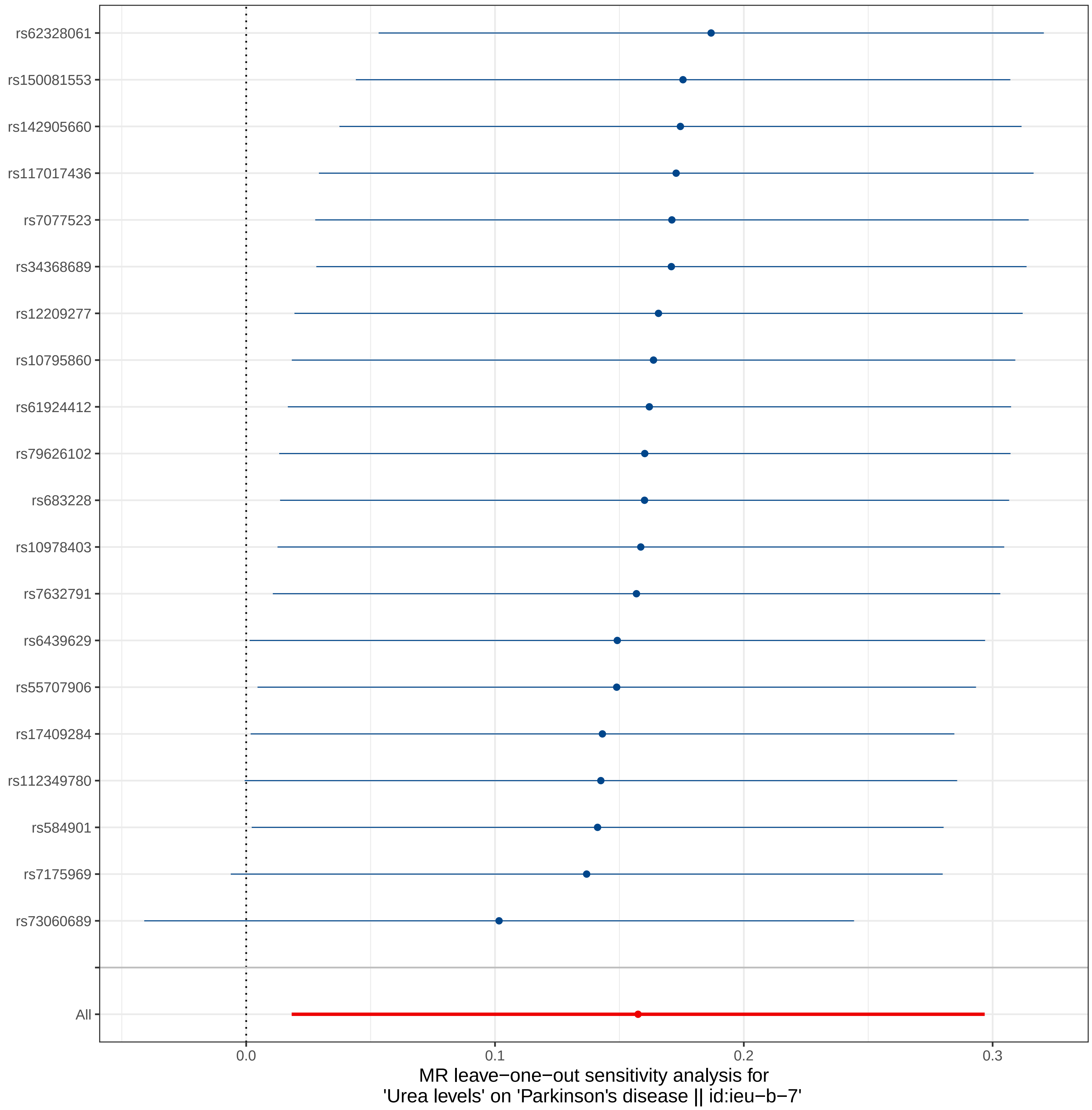
rs62328061
rs150081553
rs142905660
rs117017436
rs7077523
rs34368689
rs12209277
rs10795860
rs61924412
rs79626102
rs683228
rs10978403
rs7632791
rs6439629
rs55707906
rs17409284
rs112349780
rs584901
rs7175969
rs73060689
All
0.0
0.1
0.2
0.3
MR leave−one−out sensitivity analysis for
'Urea levels' on 'Parkinson's disease || id:ieu−b−7'

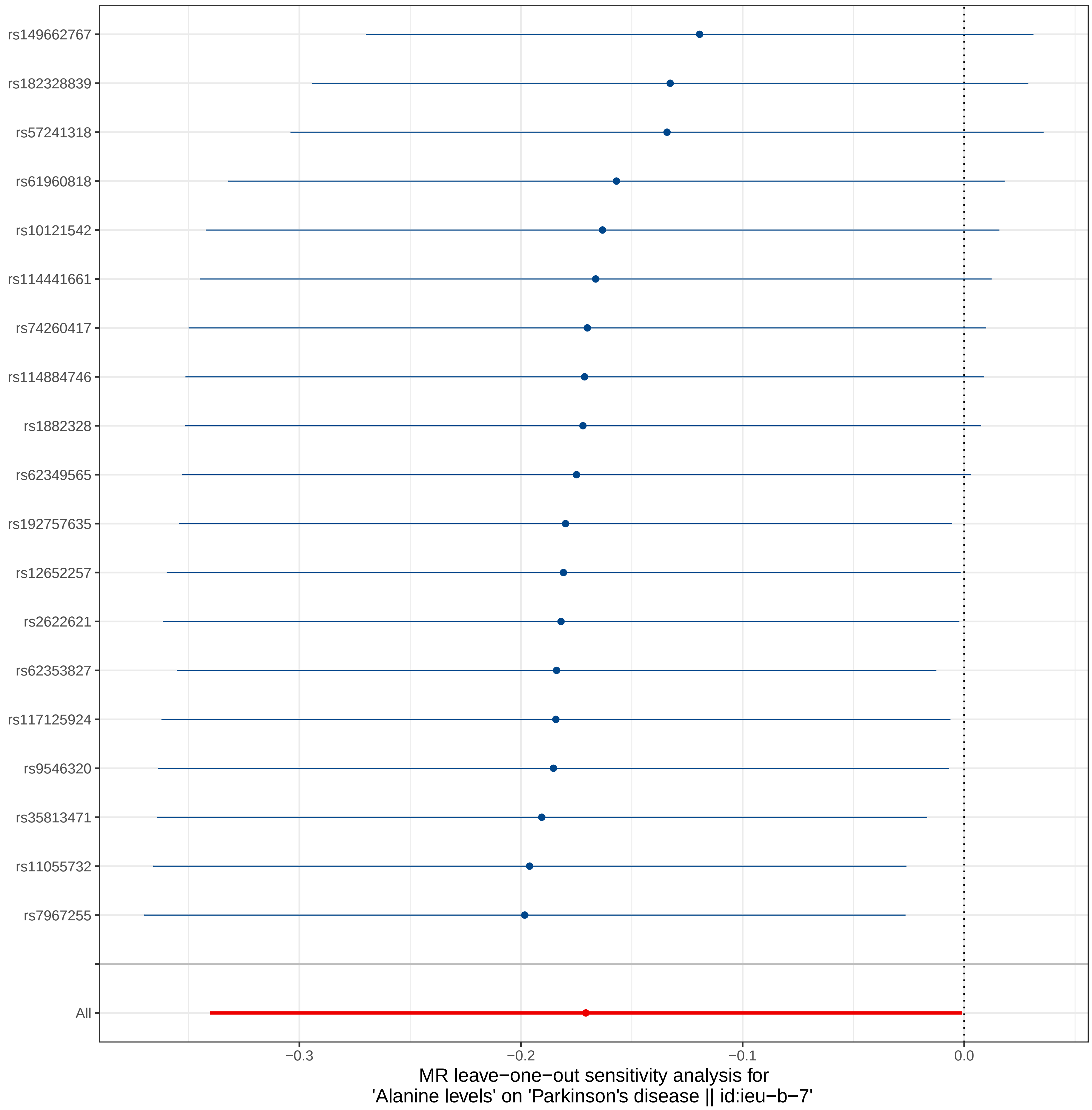
rs149662767
rs182328839
rs57241318
rs61960818
rs10121542
rs114441661
rs74260417
rs114884746
rs1882328
rs62349565
rs192757635
rs12652257
rs2622621
rs62353827
rs117125924
rs9546320
rs35813471
rs11055732
rs7967255
All
−0.3
−0.2
−0.1
0.0
MR leave−one−out sensitivity analysis for
'Alanine levels' on 'Parkinson's disease || id:ieu−b−7'

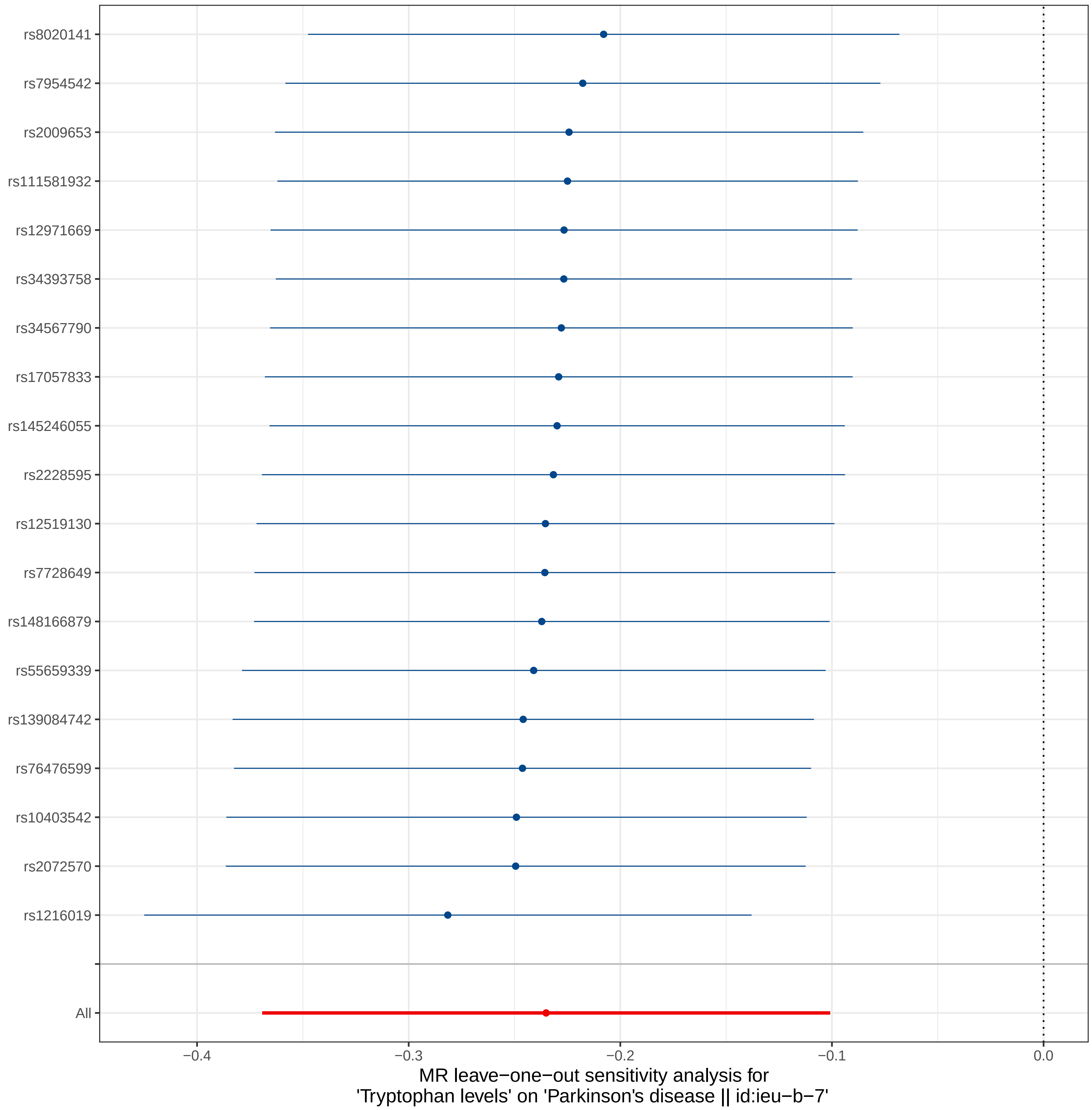

rs8020141
rs7954542
rs2009653
rs111581932
rs12971669
rs34393758
rs34567790
rs17057833
rs145246055
rs2228595
rs12519130
rs7728649
rs148166879
rs55659339
rs139084742
rs76476599
rs10403542
rs2072570
rs1216019
All
−0.4
−0.3
−0.2
−0.1
0.0
MR leave−one−out sensitivity analysis for
'Tryptophan levels' on 'Parkinson's disease || id:ieu−b−7'

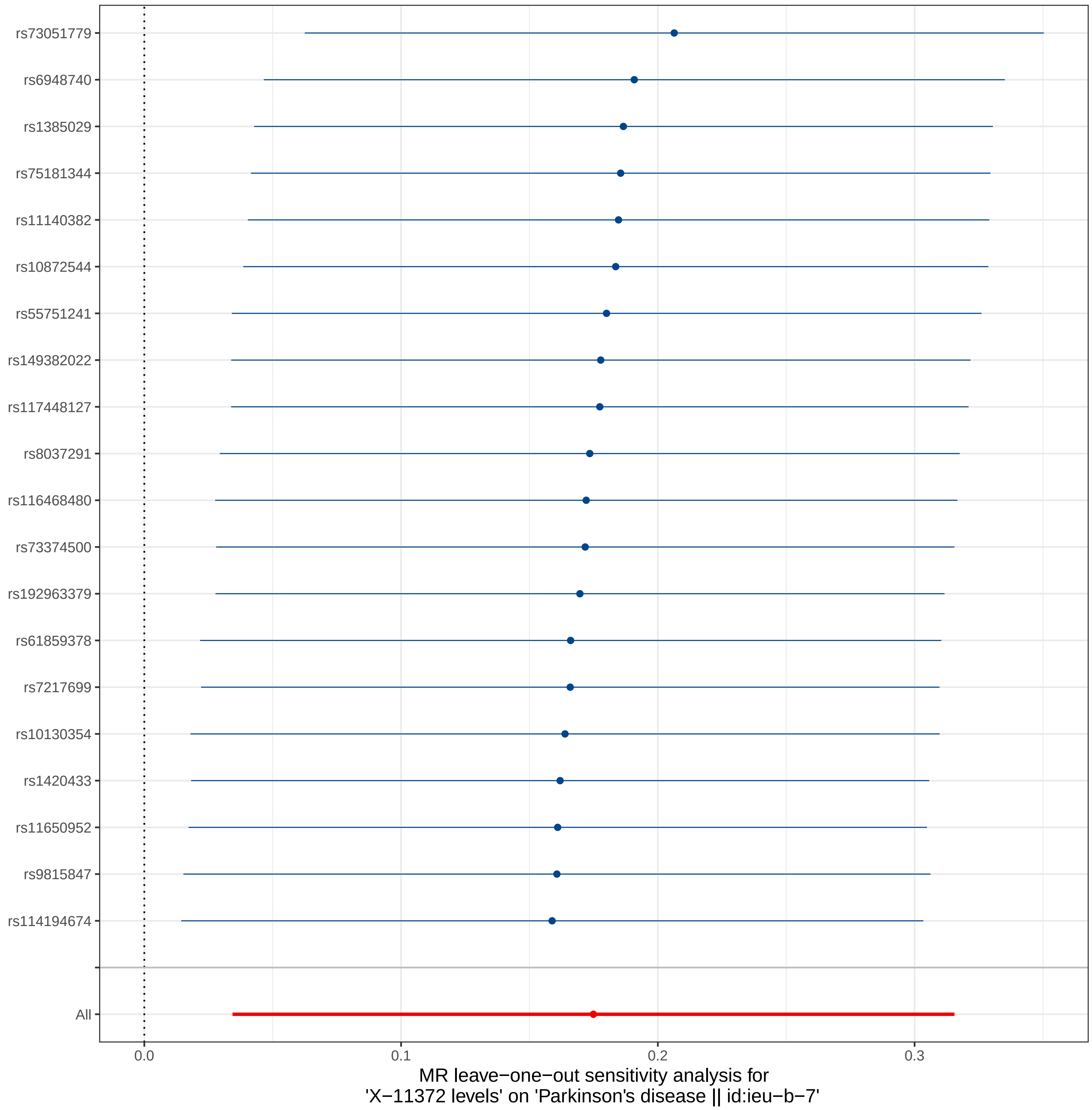

rs73051779
rs6948740
rs1385029
rs75181344
rs11140382
rs10872544
rs55751241
rs149382022
rs117448127
rs8037291
rs116468480
rs73374500
rs192963379
rs61859378
rs7217699
rs10130354
rs1420433
rs11650952
rs9815847
rs114194674
All
0.0
0.1
0.2
0.3
MR leave−one−out sensitivity analysis for
'X−11372 levels' on 'Parkinson's disease || id:ieu−b−7'

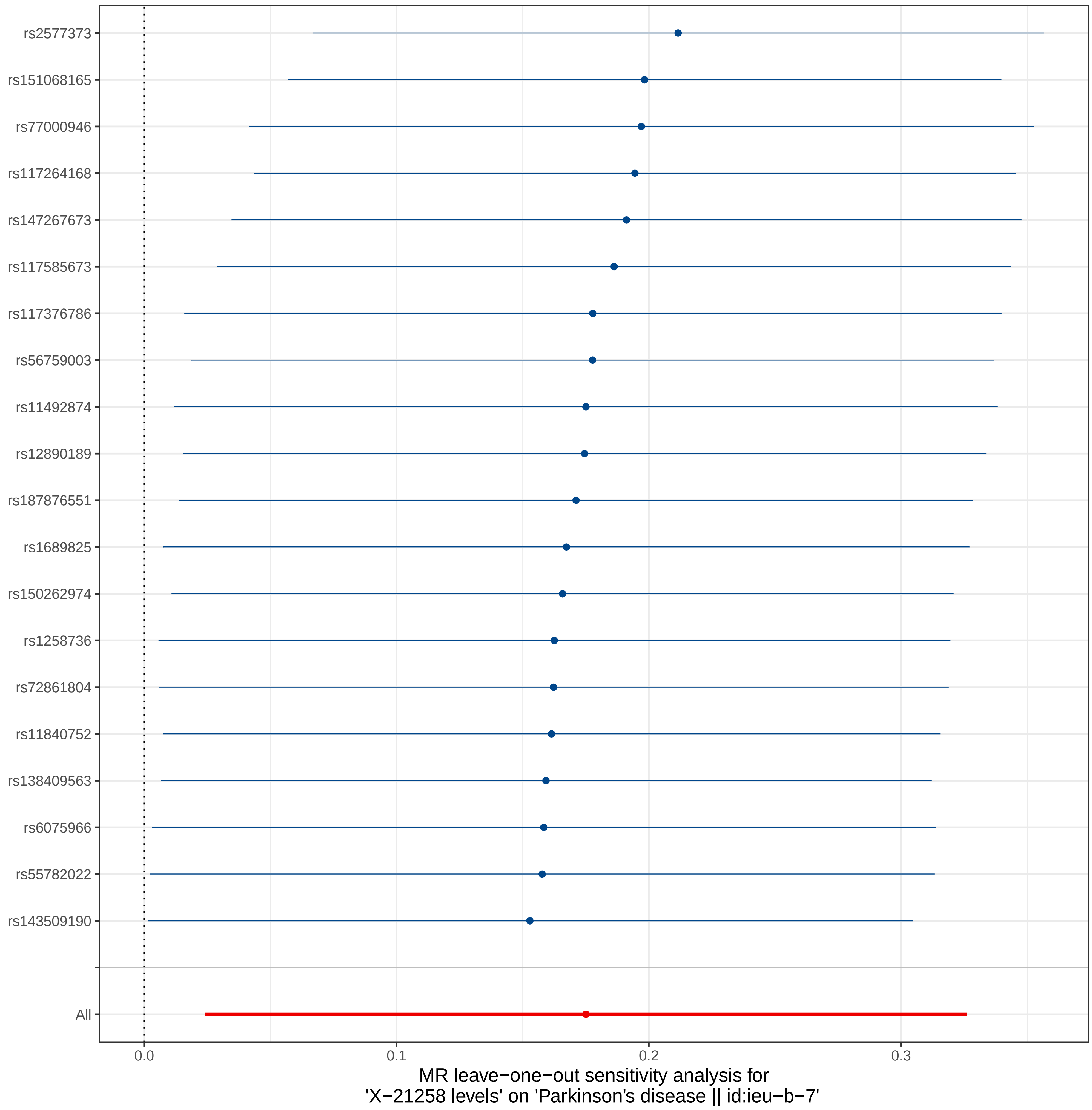
rs2577373
rs151068165
rs77000946
rs117264168
rs147267673
rs117585673
rs117376786
rs56759003
rs11492874
rs12890189
rs187876551
rs1689825
rs150262974
rs1258736
rs72861804
rs11840752
rs138409563
rs6075966
rs55782022
rs143509190
All
0.0
0.1
0.2
0.3
MR leave−one−out sensitivity analysis for
'X−21258 levels' on 'Parkinson's disease || id:ieu−b−7'

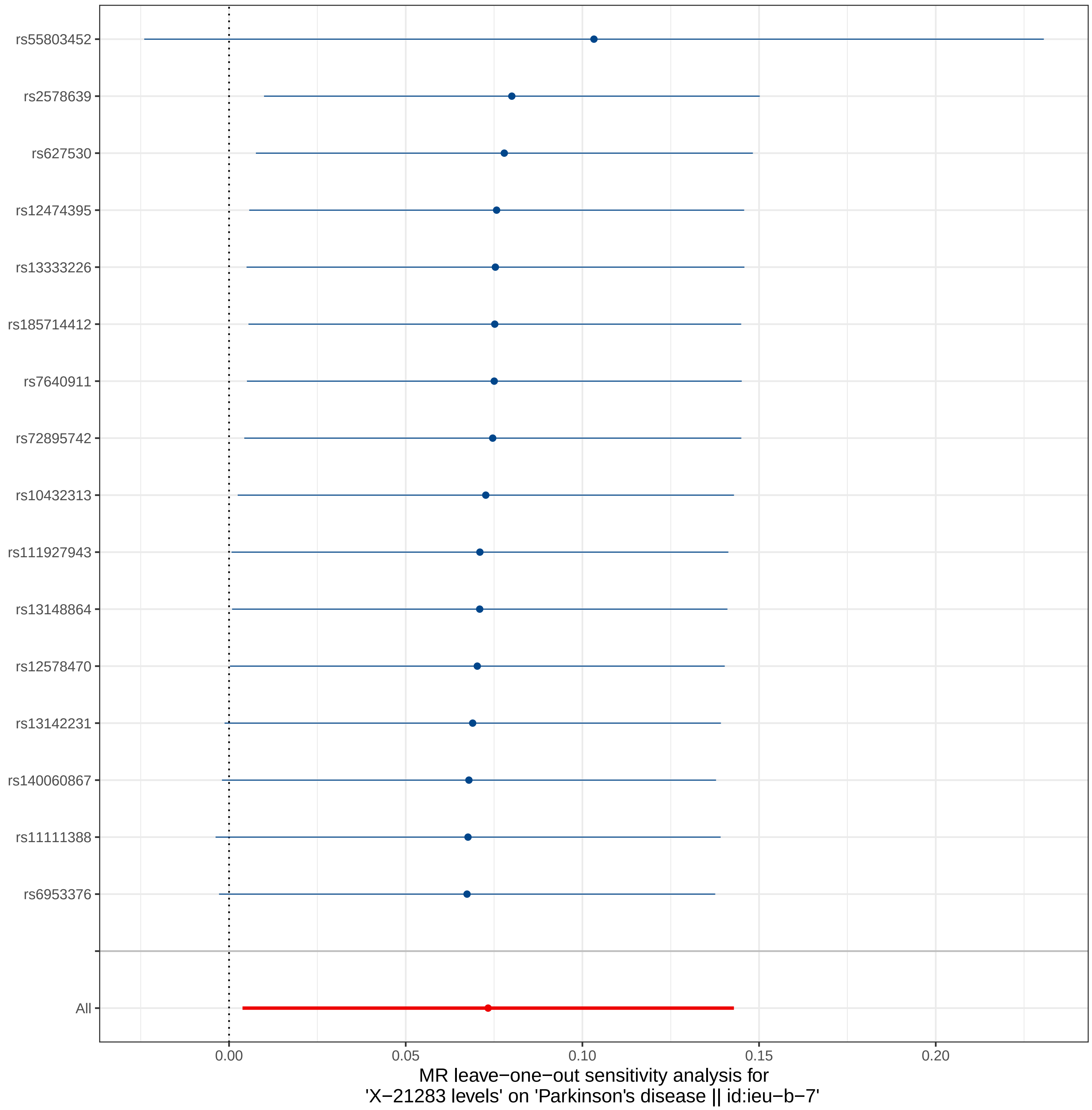

rs55803452
rs2578639
rs627530
rs12474395
rs13333226
rs185714412
rs7640911
rs72895742
rs10432313
rs111927943
rs13148864
rs12578470
rs13142231
rs140060867
rs11111388
rs6953376
All
0.00
0.05
0.10
0.15
0.20
MR leave−one−out sensitivity analysis for
'X−21283 levels' on 'Parkinson's disease || id:ieu−b−7'

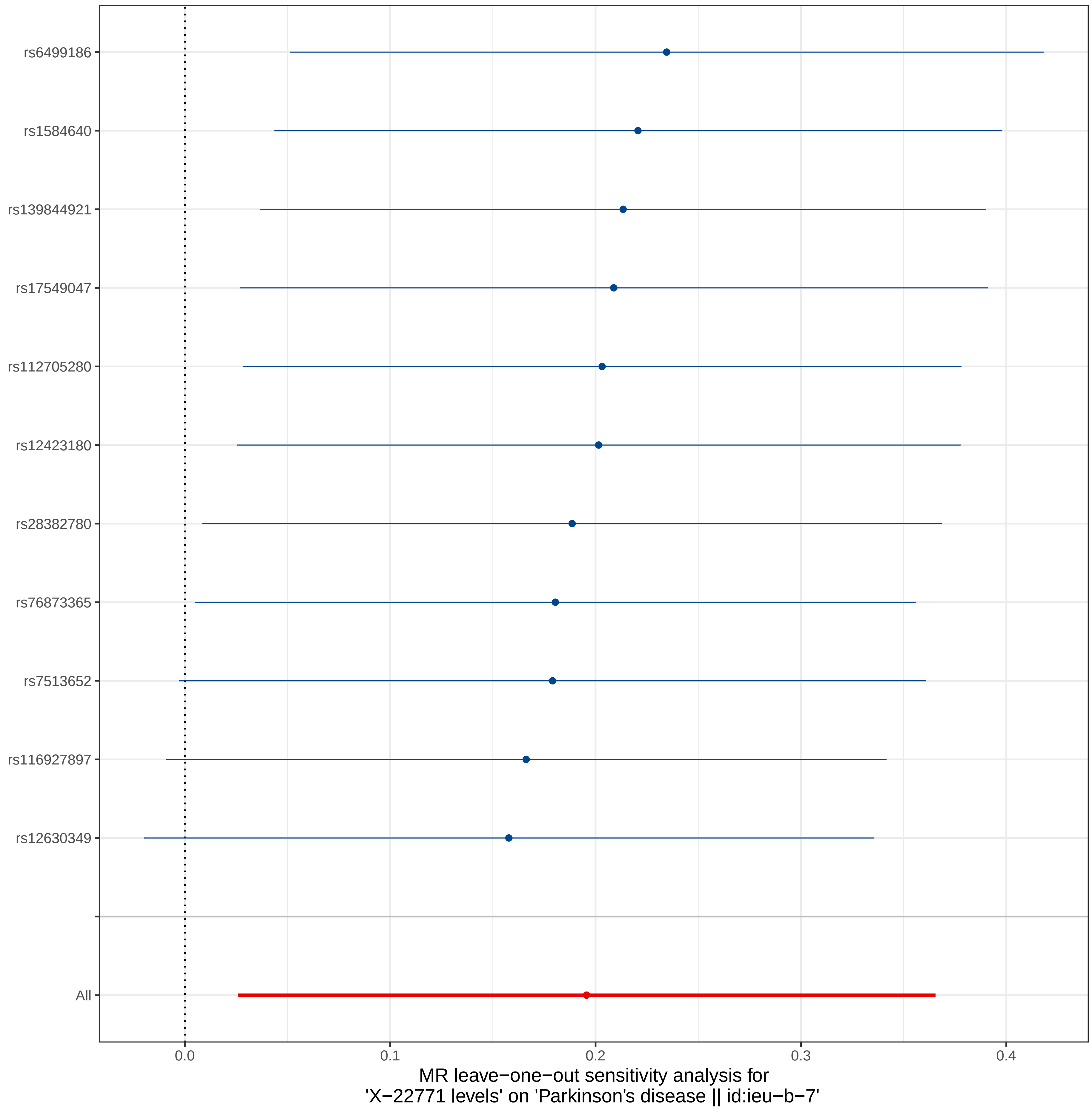
rs6499186
rs1584640
rs139844921
rs17549047
rs112705280
rs12423180
rs28382780
rs76873365
rs7513652
rs116927897
rs12630349
All
0.0
0.1
0.2
0.3
0.4
MR leave−one−out sensitivity analysis for
'X−22771 levels' on 'Parkinson's disease || id:ieu−b−7'

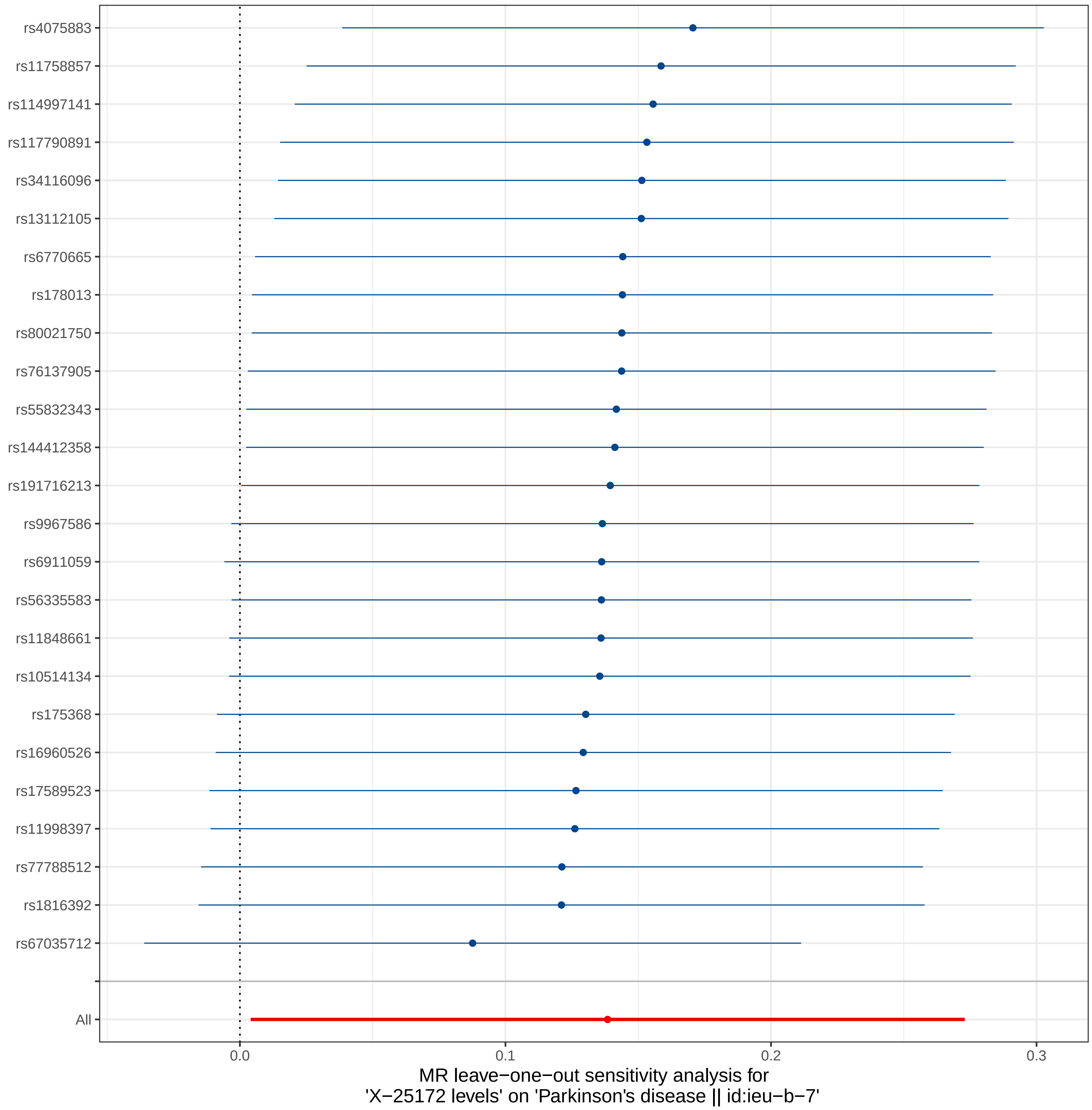

rs4075883
rs11758857
rs114997141
rs117790891
rs34116096
rs13112105
rs6770665
rs178013
rs80021750
rs76137905
rs55832343
rs144412358
rs191716213
rs9967586
rs6911059
rs56335583
rs11848661
rs10514134
rs175368
rs16960526
rs17589523
rs11998397
rs77788512
rs1816392
rs67035712
All
0.0
0.1
0.2
0.3
MR leave−one−out sensitivity analysis for
'X−25172 levels' on 'Parkinson's disease || id:ieu−b−7'

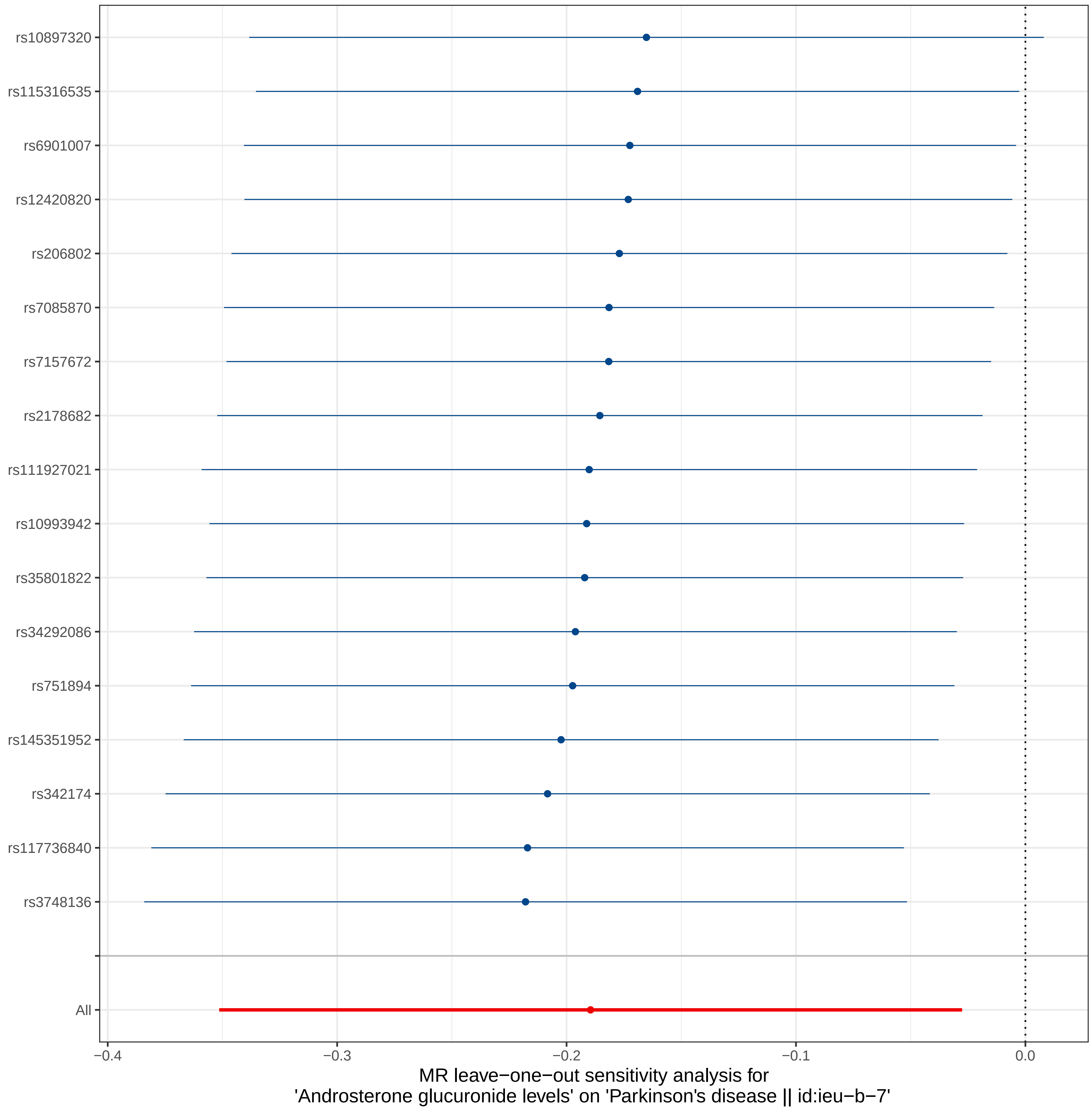

rs10897320
rs115316535
rs6901007
rs12420820
rs206802
rs7085870
rs7157672
rs2178682
rs111927021
rs10993942
rs35801822
rs34292086
rs751894
rs145351952
rs342174
rs117736840
rs3748136
All
−0.4
−0.3
−0.2
−0.1
0.0
MR leave−one−out sensitivity analysis for
'Androsterone glucuronide levels' on 'Parkinson's disease || id:ieu−b−7'

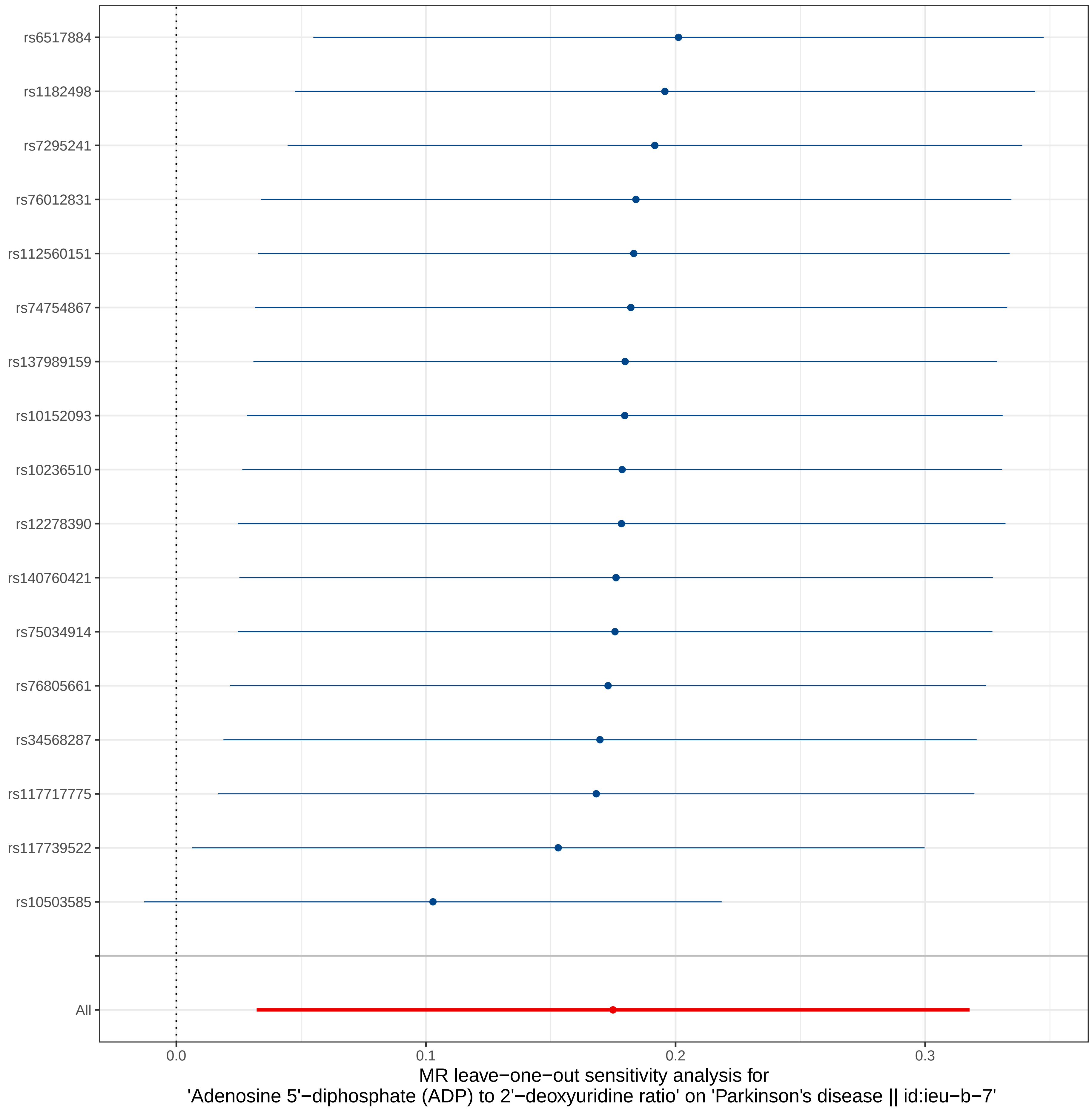
rs6517884
rs1182498
rs7295241
rs76012831
rs112560151
rs74754867
rs137989159
rs10152093
rs10236510
rs12278390
rs140760421
rs75034914
rs76805661
rs34568287
rs117717775
rs117739522
rs10503585
All
0.0
0.1
0.2
0.3
MR leave−one−out sensitivity analysis for
'Adenosine 5'−diphosphate (ADP) to 2'−deoxyuridine ratio' on 'Parkinson's disease || id:ieu−b−7'

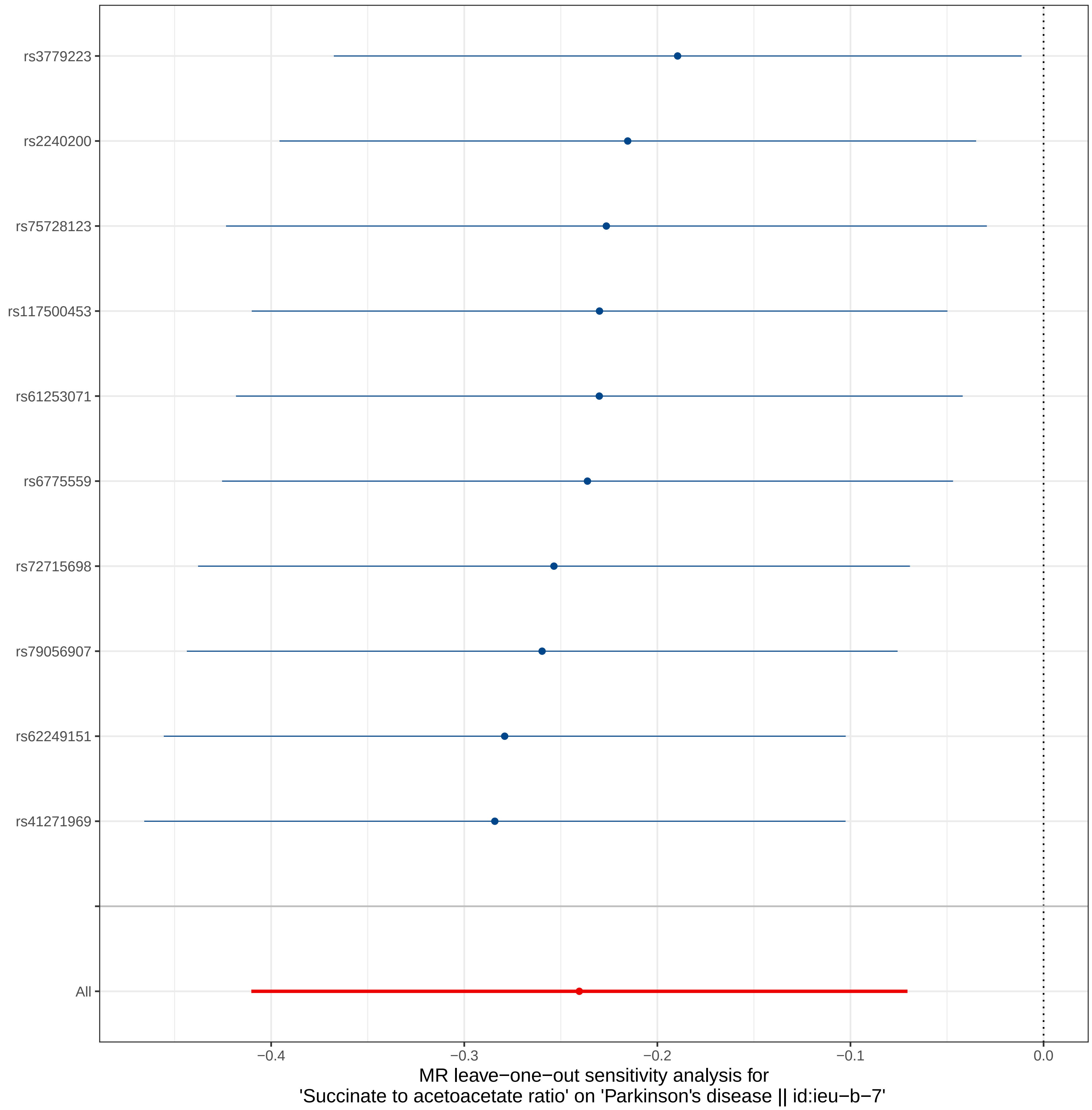

rs3779223
rs2240200
rs75728123
rs117500453
rs61253071
rs6775559
rs72715698
rs79056907
rs62249151
rs41271969
All
−0.4
−0.3
−0.2
−0.1
0.0
MR leave−one−out sensitivity analysis for
'Succinate to acetoacetate ratio' on 'Parkinson's disease || id:ieu−b−7'

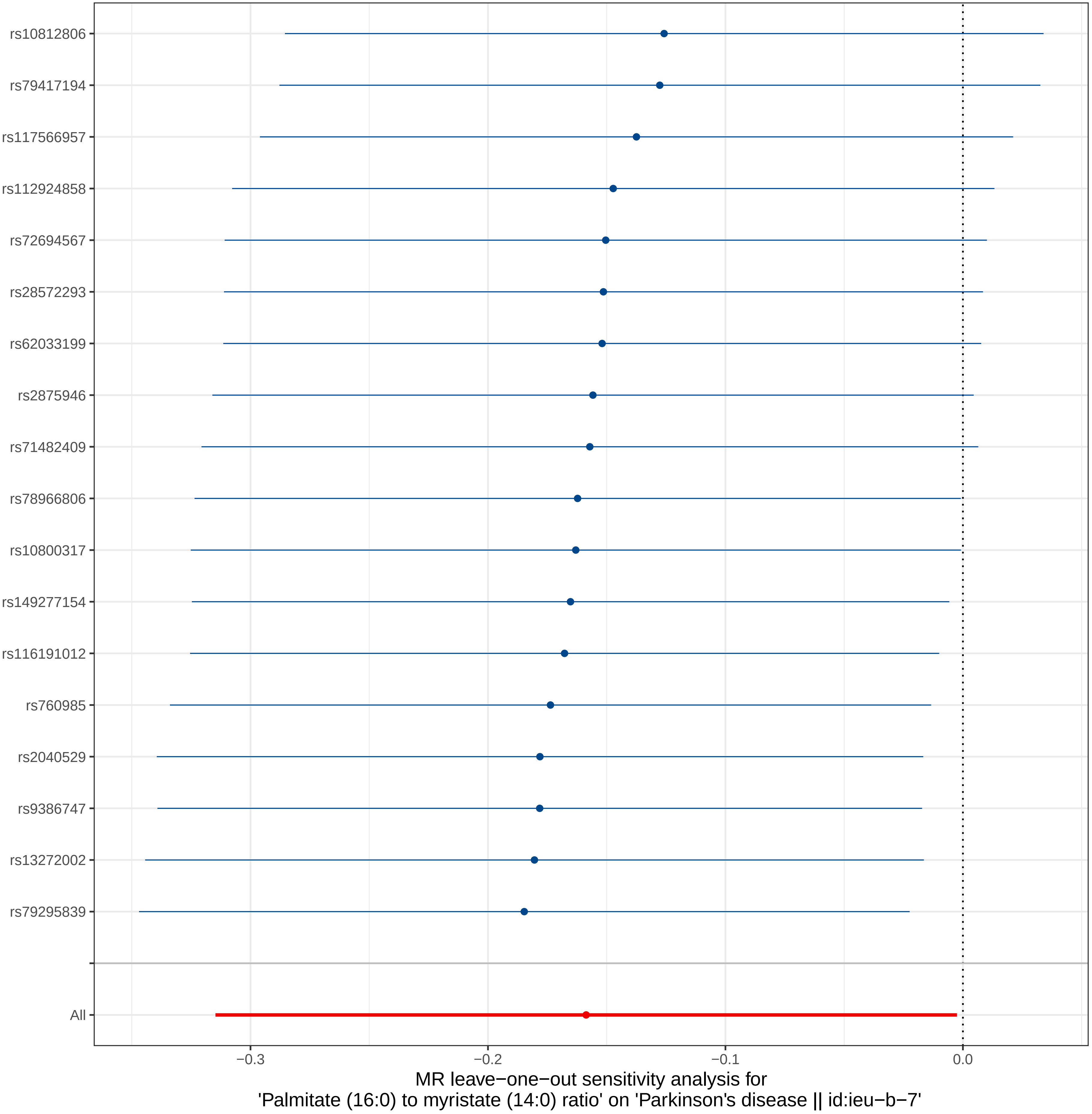
rs10812806
rs79417194
rs117566957
rs112924858
rs72694567
rs28572293
rs62033199
rs2875946
rs71482409
rs78966806
rs10800317
rs149277154
rs116191012
rs760985
rs2040529
rs9386747
rs13272002
rs79295839
All
−0.3
−0.2
−0.1
0.0
MR leave−one−out sensitivity analysis for
'Palmitate (16:0) to myristate (14:0) ratio' on 'Parkinson's disease || id:ieu−b−7'

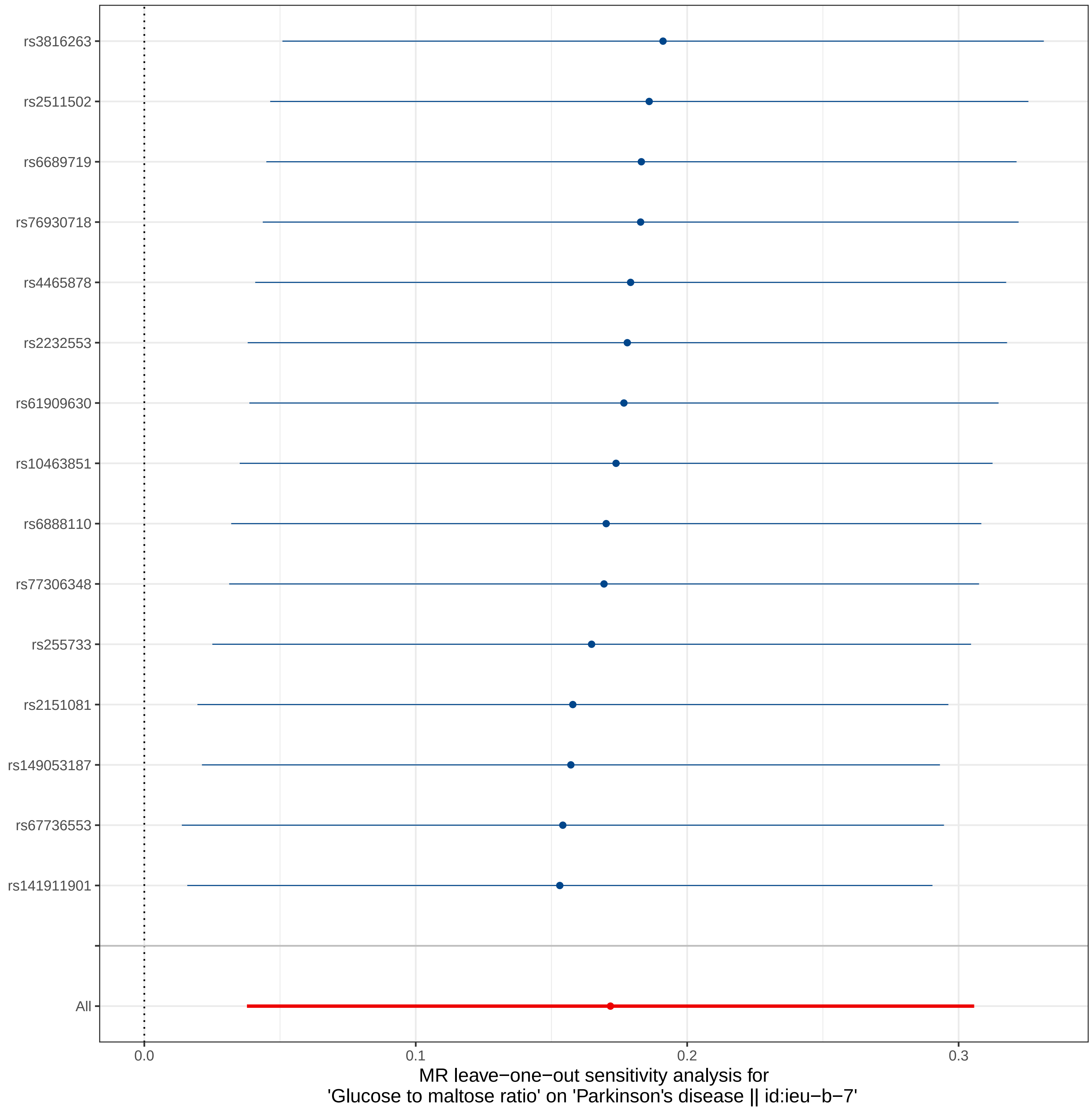
rs3816263
rs2511502
rs6689719
rs76930718
rs4465878
rs2232553
rs61909630
rs10463851
rs6888110
rs77306348
rs255733
rs2151081
rs149053187
rs67736553
rs141911901
All
0.0
0.1
0.2
0.3
MR leave−one−out sensitivity analysis for
'Glucose to maltose ratio' on 'Parkinson's disease || id:ieu−b−7'

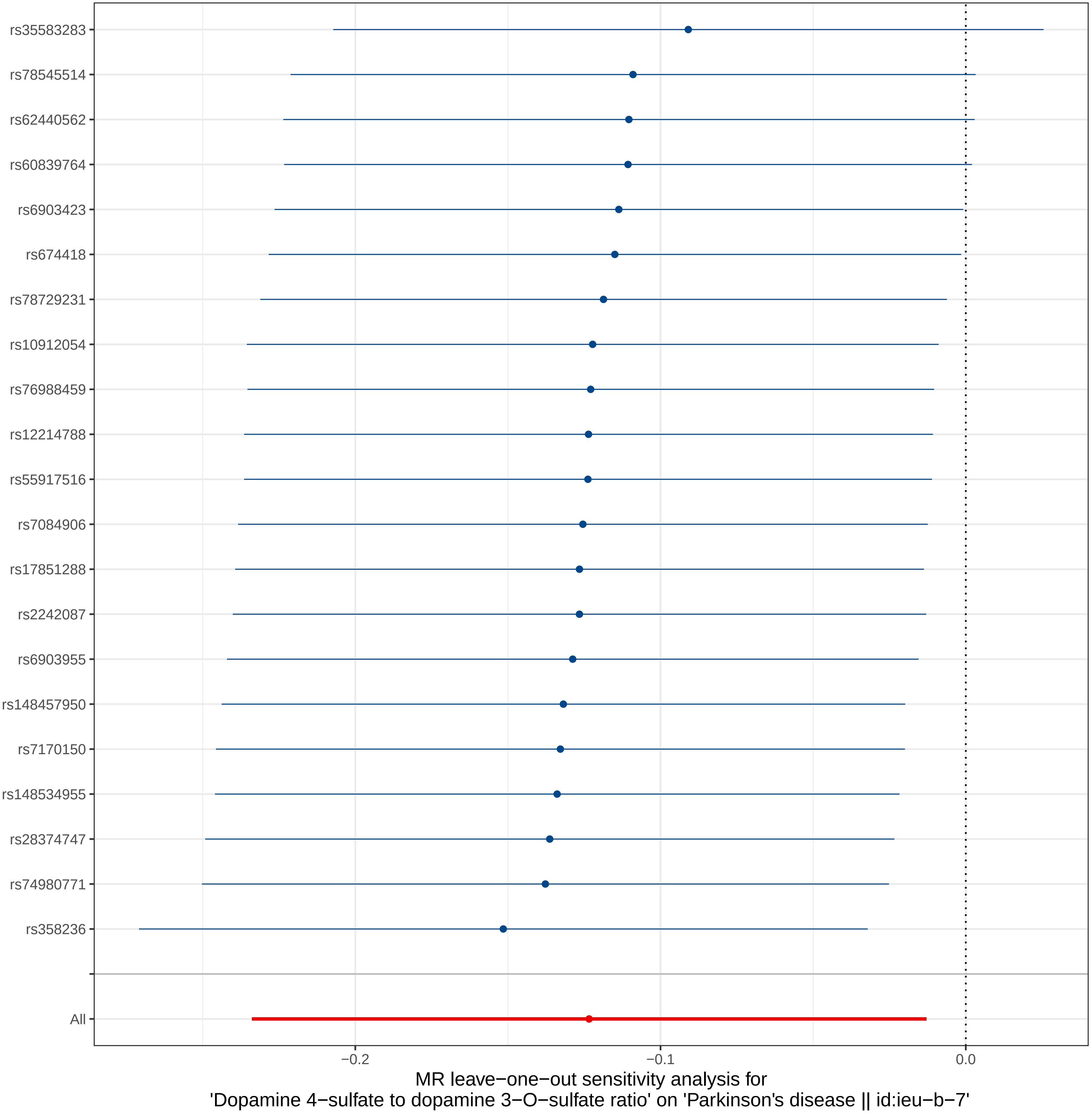
rs35583283
rs78545514
rs62440562
rs60839764
rs6903423
rs674418
rs78729231
rs10912054
rs76988459
rs12214788
rs55917516
rs7084906
rs17851288
rs2242087
rs6903955
rs148457950
rs7170150
rs148534955
rs28374747
rs74980771
rs358236
All
−0.2
−0.1
0.0
MR leave−one−out sensitivity analysis for
'Dopamine 4−sulfate to dopamine 3−O−sulfate ratio' on 'Parkinson's disease || id:ieu−b−7'

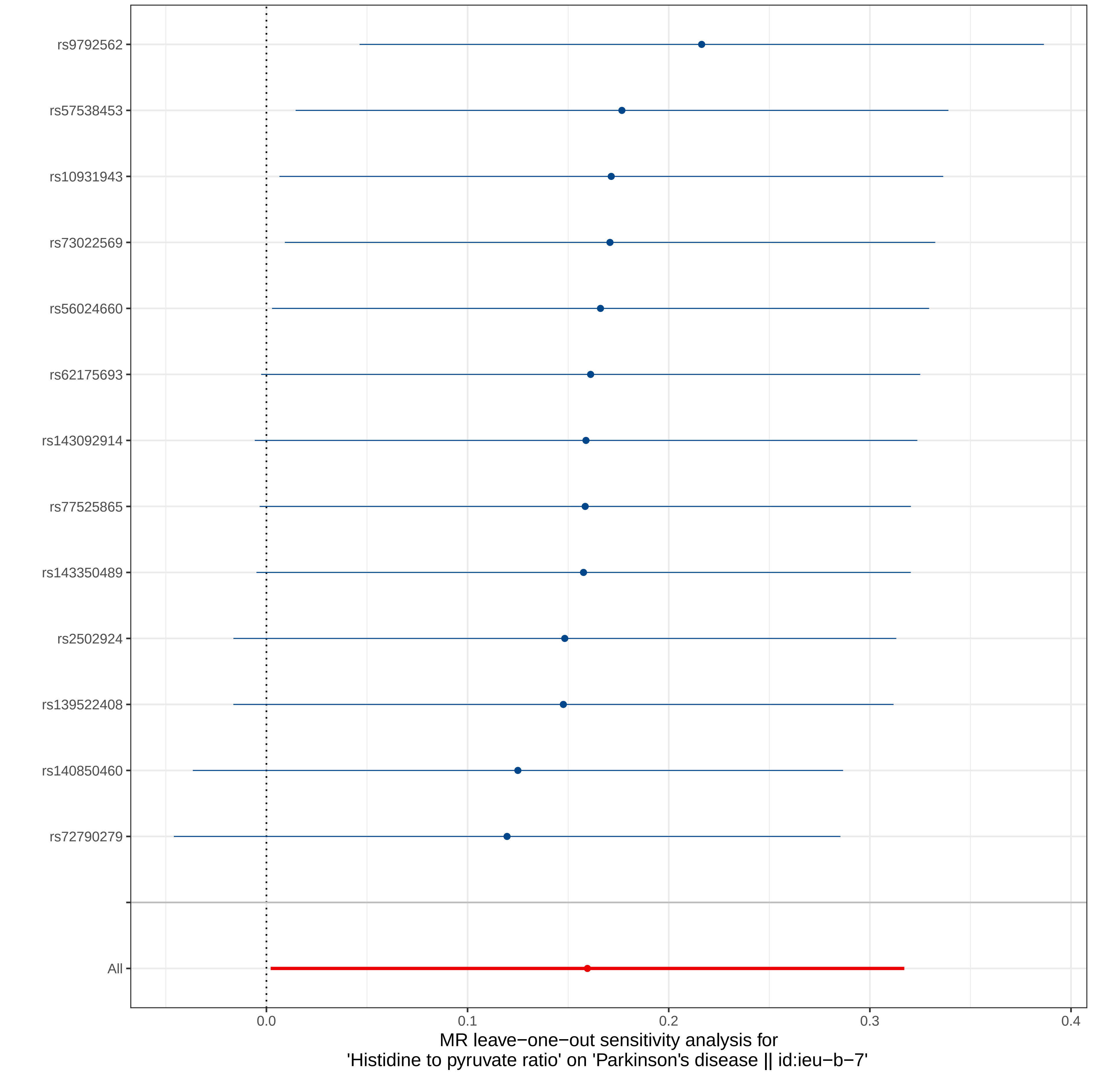

rs9792562
rs57538453
rs10931943
rs73022569
rs56024660
rs62175693
rs143092914
rs77525865
rs143350489
rs2502924
rs139522408
rs140850460
rs72790279
All
0.0
0.1
0.2
0.3
0.4
MR leave−one−out sensitivity analysis for
'Histidine to pyruvate ratio' on 'Parkinson's disease || id:ieu−b−7'

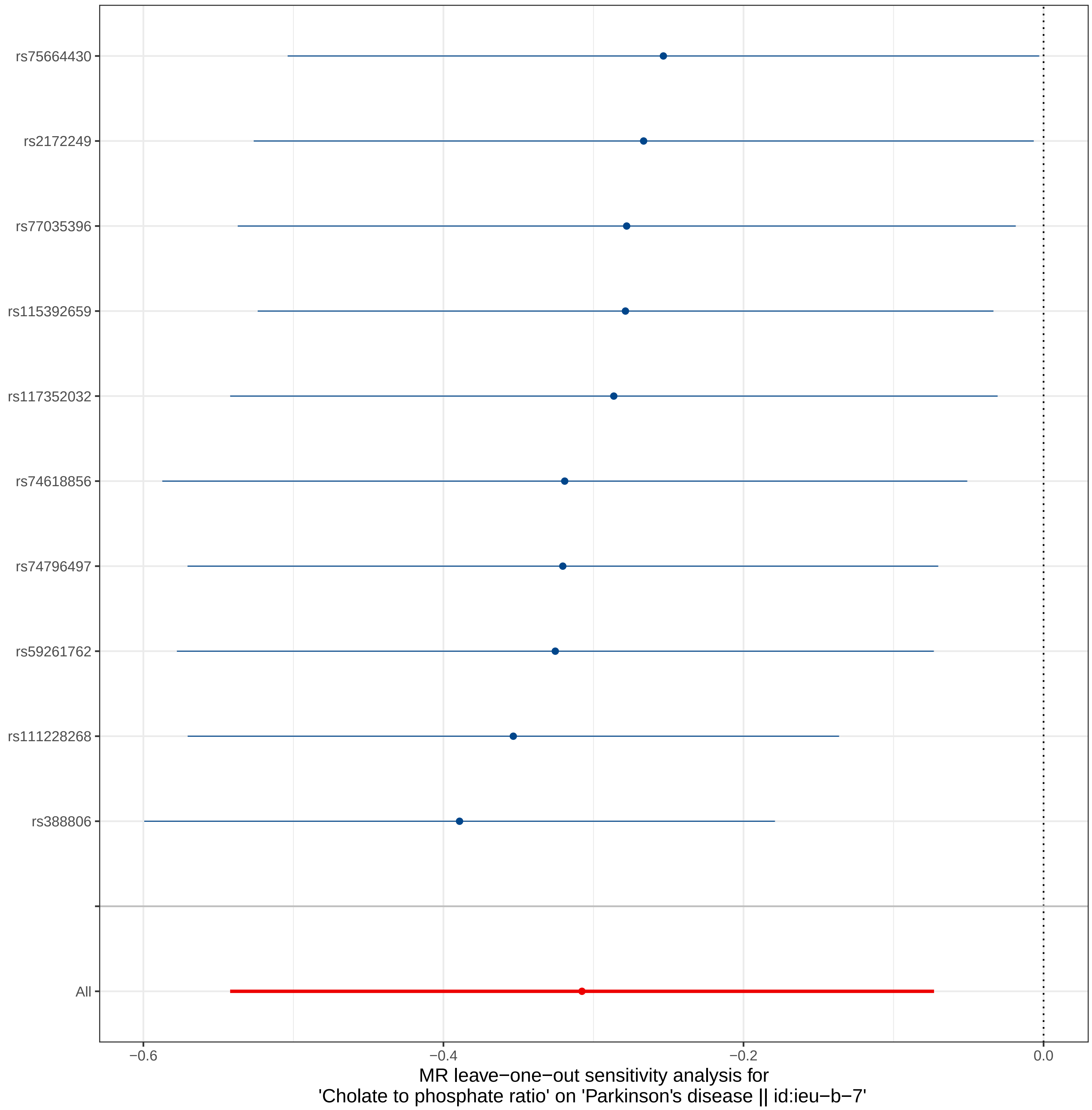
rs75664430
rs2172249
rs77035396
rs115392659
rs117352032
rs74618856
rs74796497
rs59261762
rs111228268
rs388806
All
−0.6
−0.4
−0.2
0.0
MR leave−one−out sensitivity analysis for
'Cholate to phosphate ratio' on 'Parkinson's disease || id:ieu−b−7'

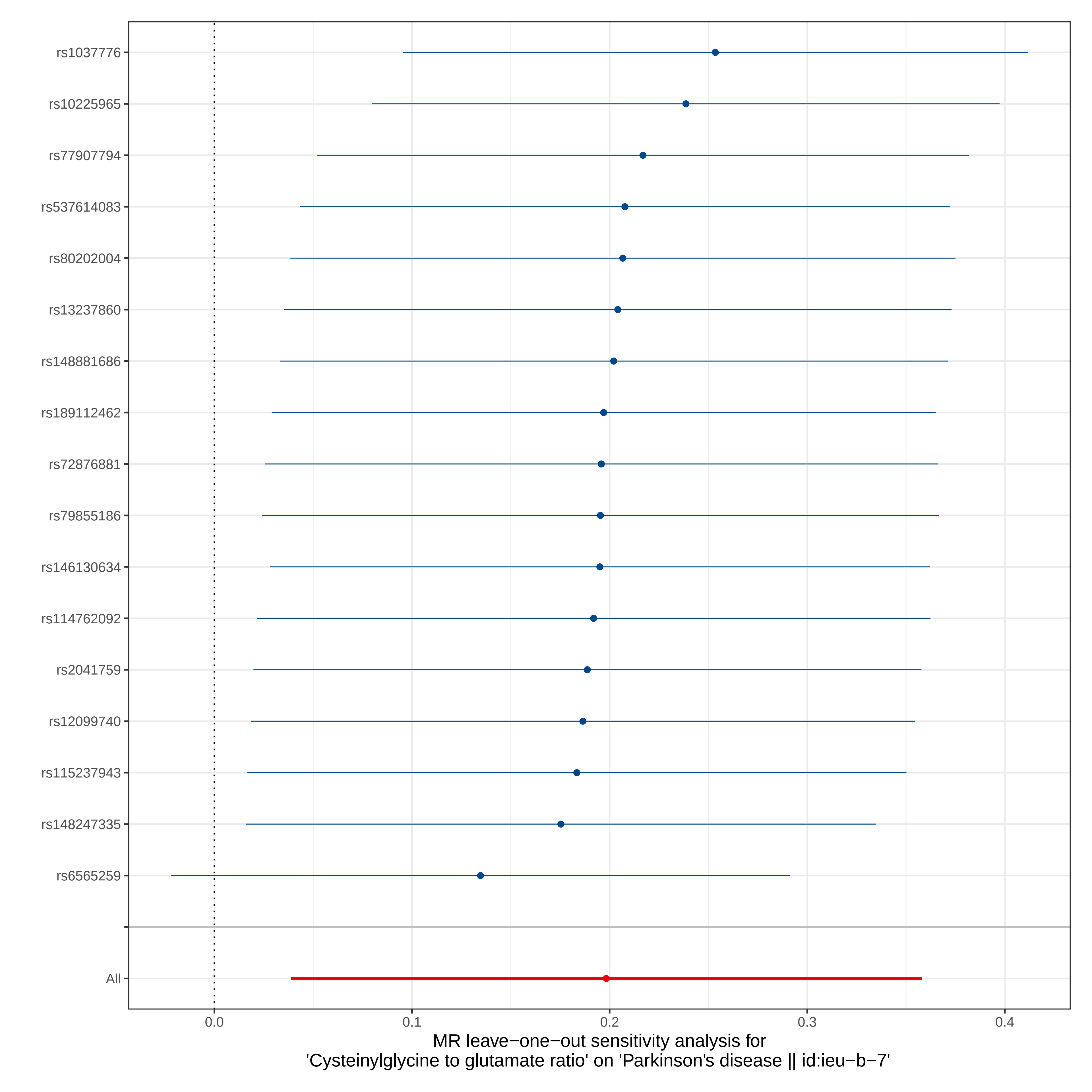

rs1037776
rs10225965
rs77907794
rs537614083
rs80202004
rs13237860
rs148881686
rs189112462
rs72876881
rs79855186
rs146130634
rs114762092
rs2041759
rs12099740
rs115237943
rs148247335
rs6565259
All
0.0
0.1
0.2
0.3
0.4
MR leave−one−out sensitivity analysis for
'Cysteinylglycine to glutamate ratio' on 'Parkinson's disease || id:ieu−b−7'

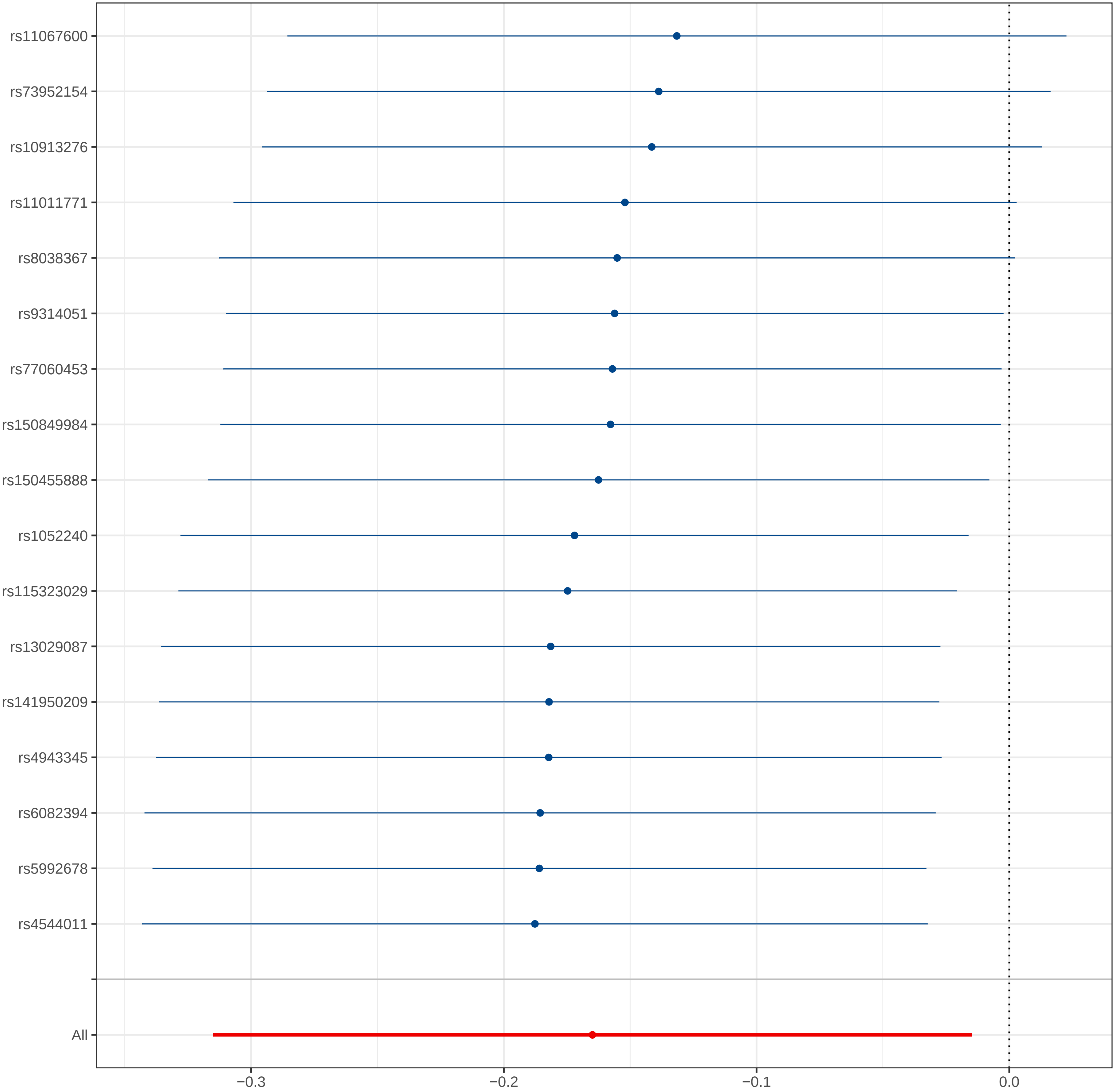

MR leave−one−out sensitivity analysis for
'Glycerol to mannitol to sorbitol ratio' on 'Parkinson's disease || id:ieu−b−7'

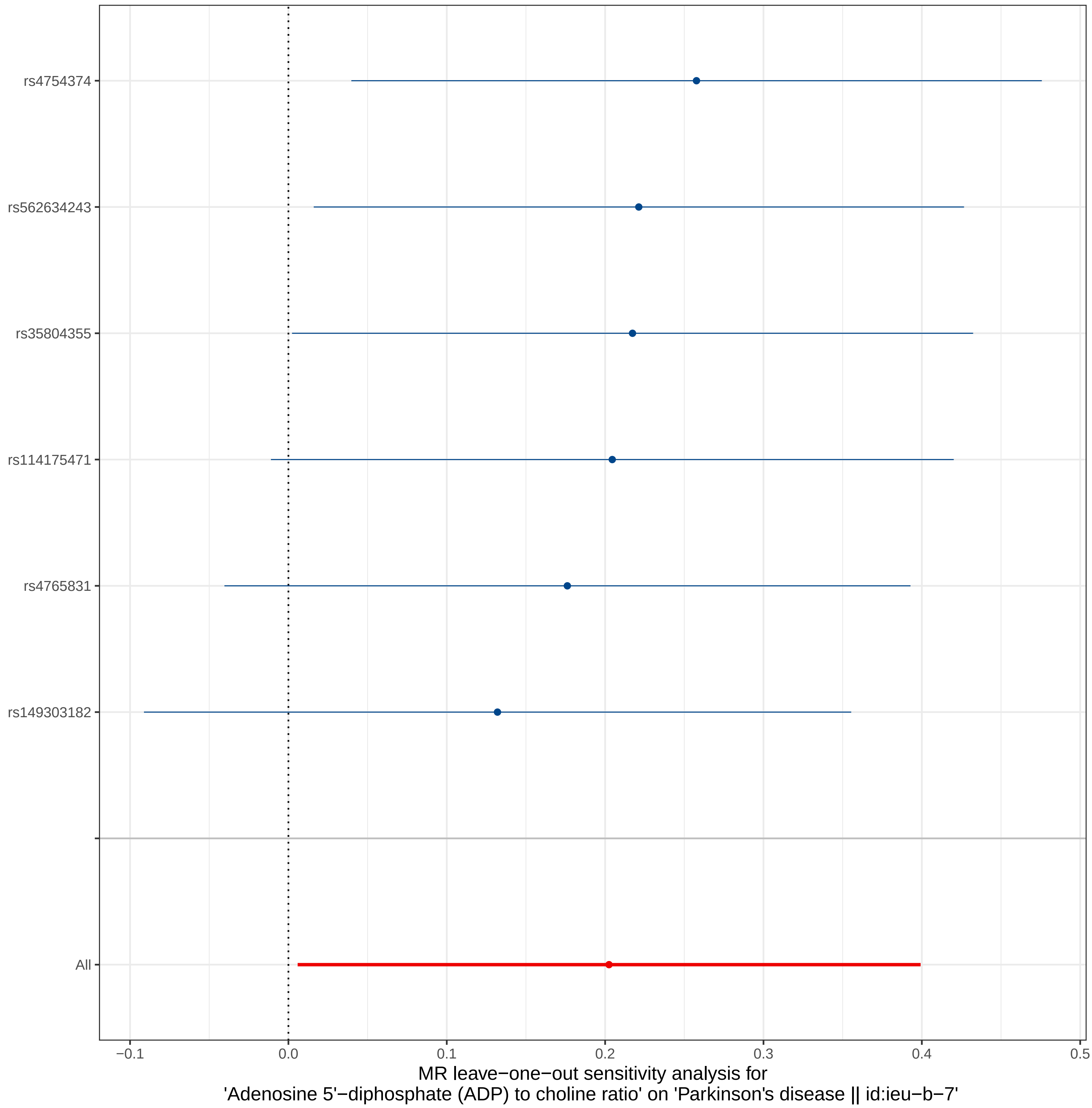
rs4754374
rs562634243
rs35804355
rs114175471
rs4765831
rs149303182
All
−0.1
0.0
0.1
0.2
0.3
0.4
0.5
MR leave−one−out sensitivity analysis for
'Adenosine 5'−diphosphate (ADP) to choline ratio' on 'Parkinson's disease || id:ieu−b−7'

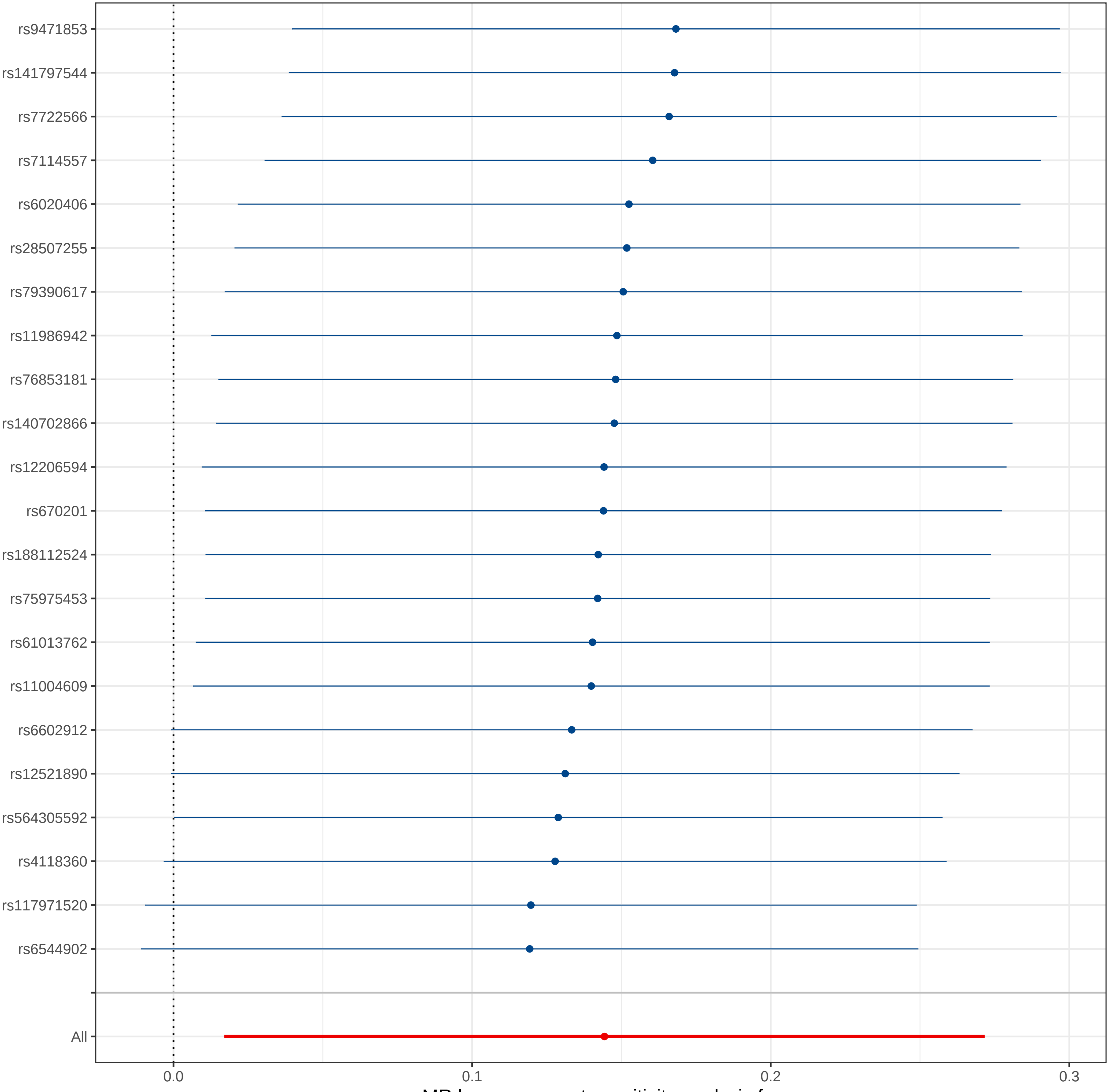

MR leave−one−out sensitivity analysis for
'Benzoate to oleoyl−linoleoyl−glycerol (18:1 to 18:2) [2] ratio' on 'Parkinson's disease || id:ieu−b−7'

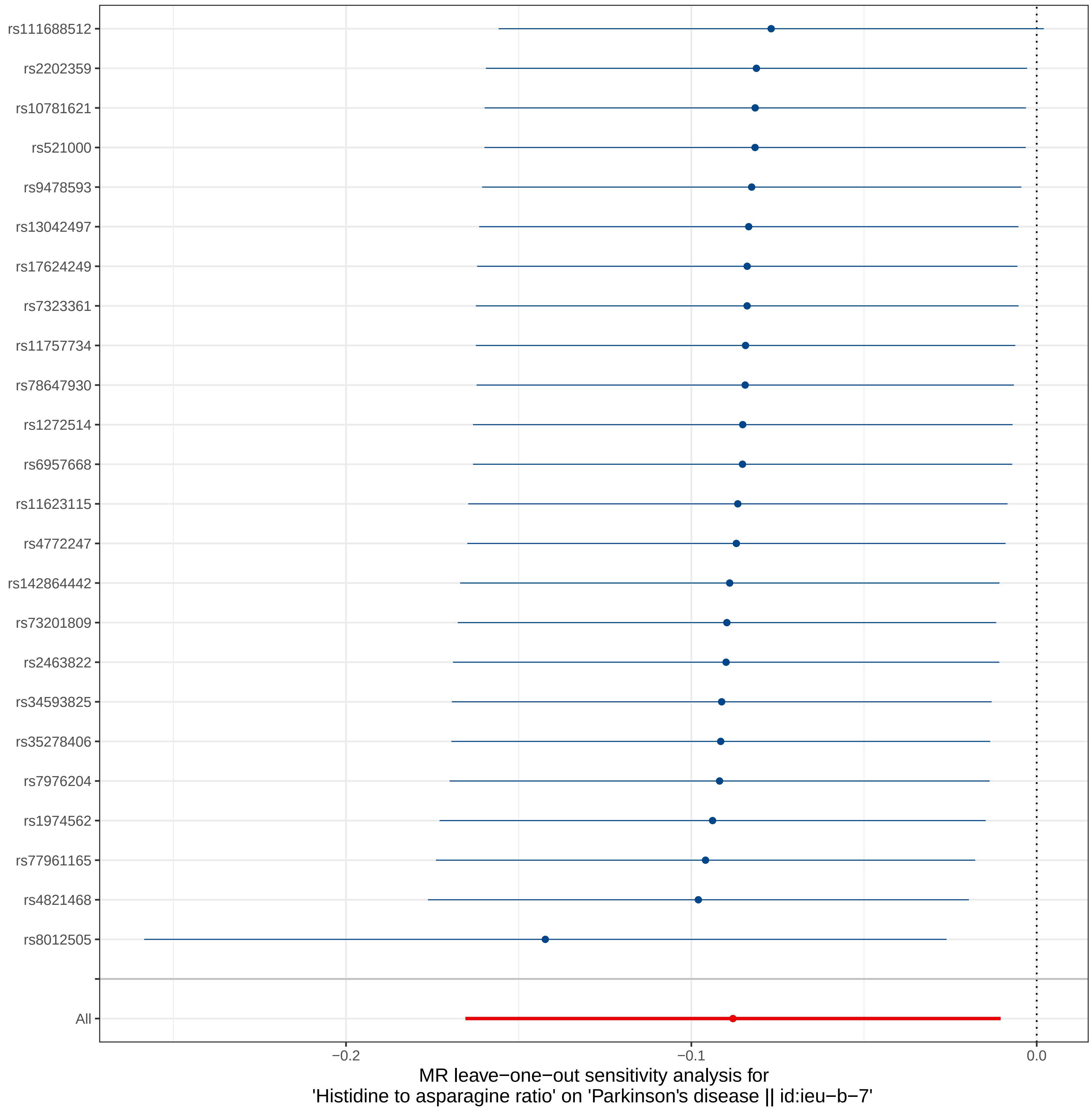
rs111688512
rs2202359
rs10781621
rs521000
rs9478593
rs13042497
rs17624249
rs7323361
rs11757734
rs78647930
rs1272514
rs6957668
rs11623115
rs4772247
rs142864442
rs73201809
rs2463822
rs34593825
rs35278406
rs7976204
rs1974562
rs77961165
rs4821468
rs8012505
All
−0.2
−0.1
0.0
MR leave−one−out sensitivity analysis for
'Histidine to asparagine ratio' on 'Parkinson's disease || id:ieu−b−7'

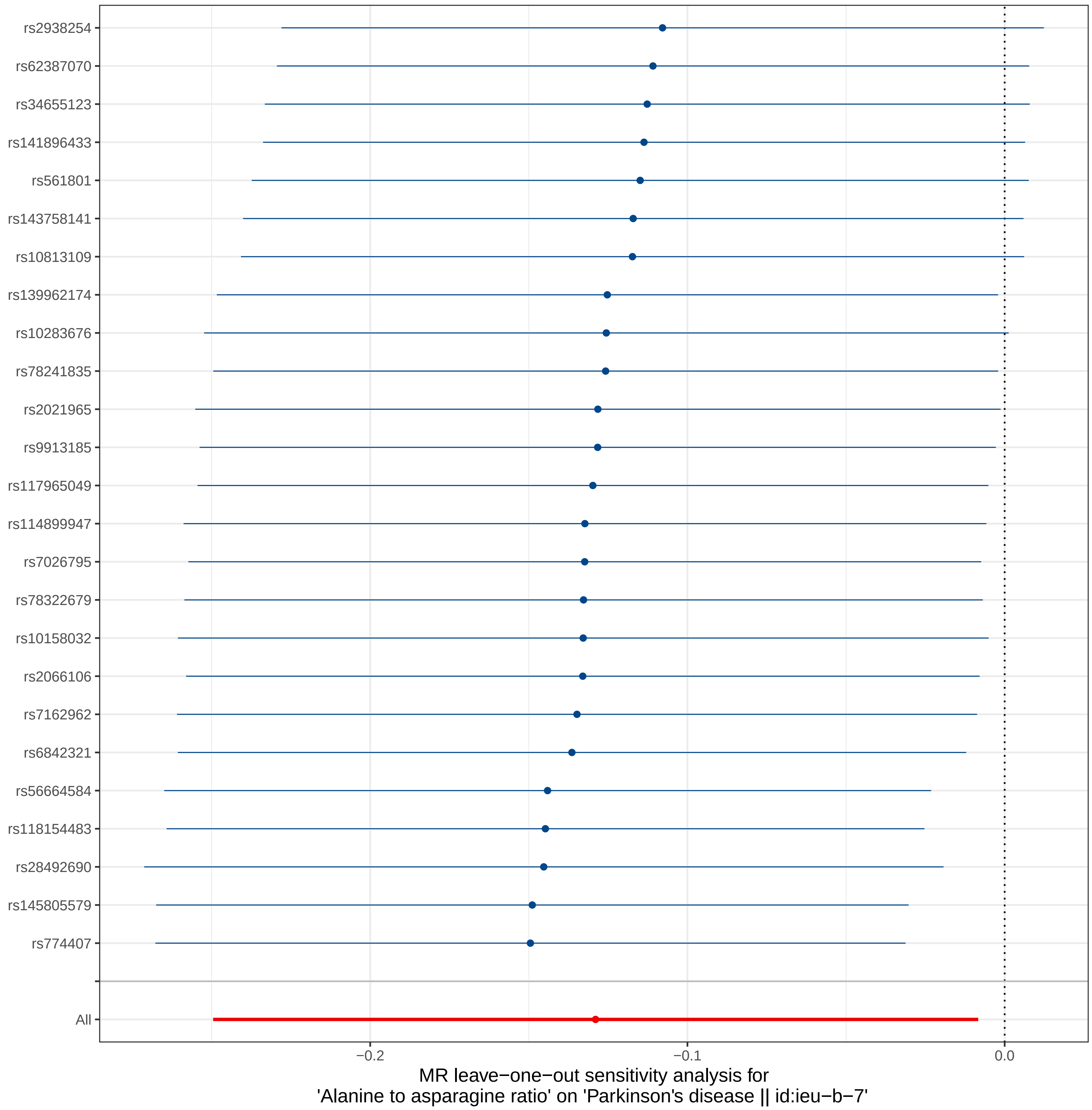

rs2938254
rs62387070
rs34655123
rs141896433
rs561801
rs143758141
rs10813109
rs139962174
rs10283676
rs78241835
rs2021965
rs9913185
rs117965049
rs114899947
rs7026795
rs78322679
rs10158032
rs2066106
rs7162962
rs6842321
rs56664584
rs118154483
rs28492690
rs145805579
rs774407
All
−0.2
−0.1
0.0
MR leave−one−out sensitivity analysis for
'Alanine to asparagine ratio' on 'Parkinson's disease || id:ieu−b−7'

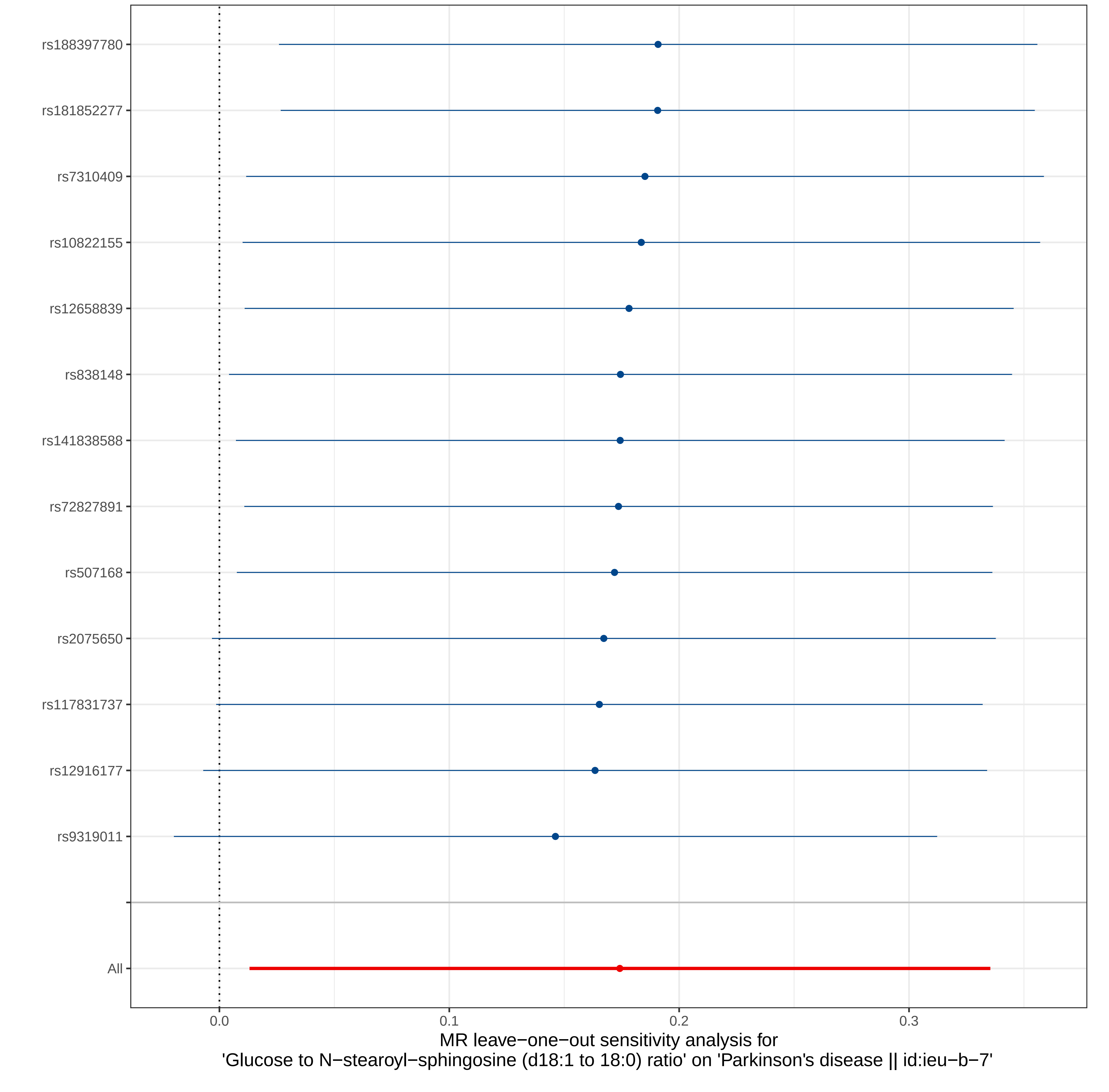

rs188397780
rs181852277
rs7310409
rs10822155
rs12658839
rs838148
rs141838588
rs72827891
rs507168
rs2075650
rs117831737
rs12916177
rs9319011
All
0.0
0.1
0.2
0.3
MR leave−one−out sensitivity analysis for
'Glucose to N−stearoyl−sphingosine (d18:1 to 18:0) ratio' on 'Parkinson's disease || id:ieu−b−7'
