## Supplemental Figure 4 for "Cerebrospinal Fluid and Plasma Metabolites with Parkinson’s Disease: A Mendelian Randomization Study"

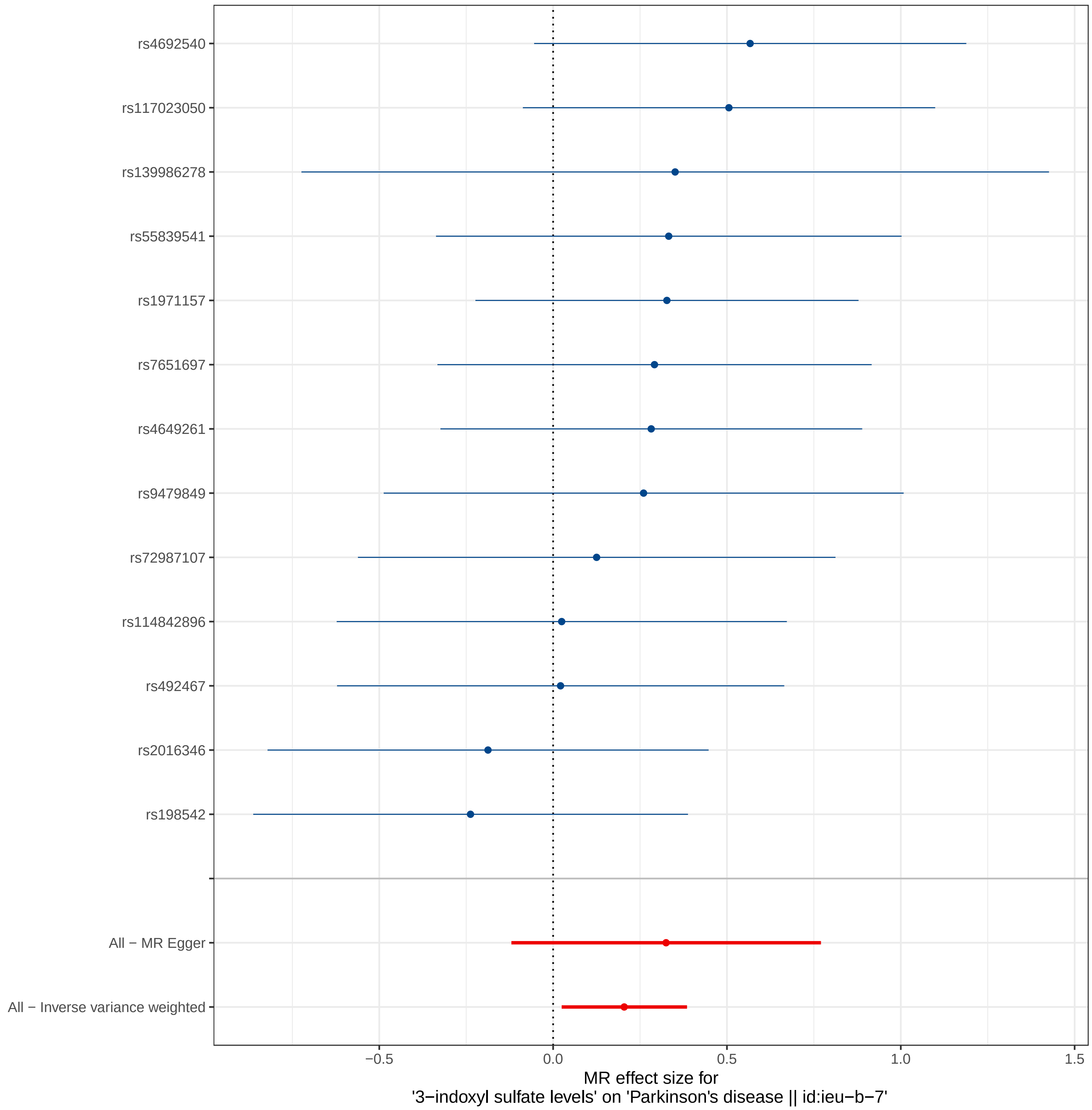
rs4692540
rs117023050
rs139986278
rs55839541
rs1971157
rs7651697
rs4649261
rs9479849
rs72987107
rs114842896
rs492467
rs2016346
rs198542
All − MR Egger
All − Inverse variance weighted
−0.5
0.0
0.5
1.0
1.5
MR effect size for
'3−indoxyl sulfate levels' on 'Parkinson's disease || id:ieu−b−7'

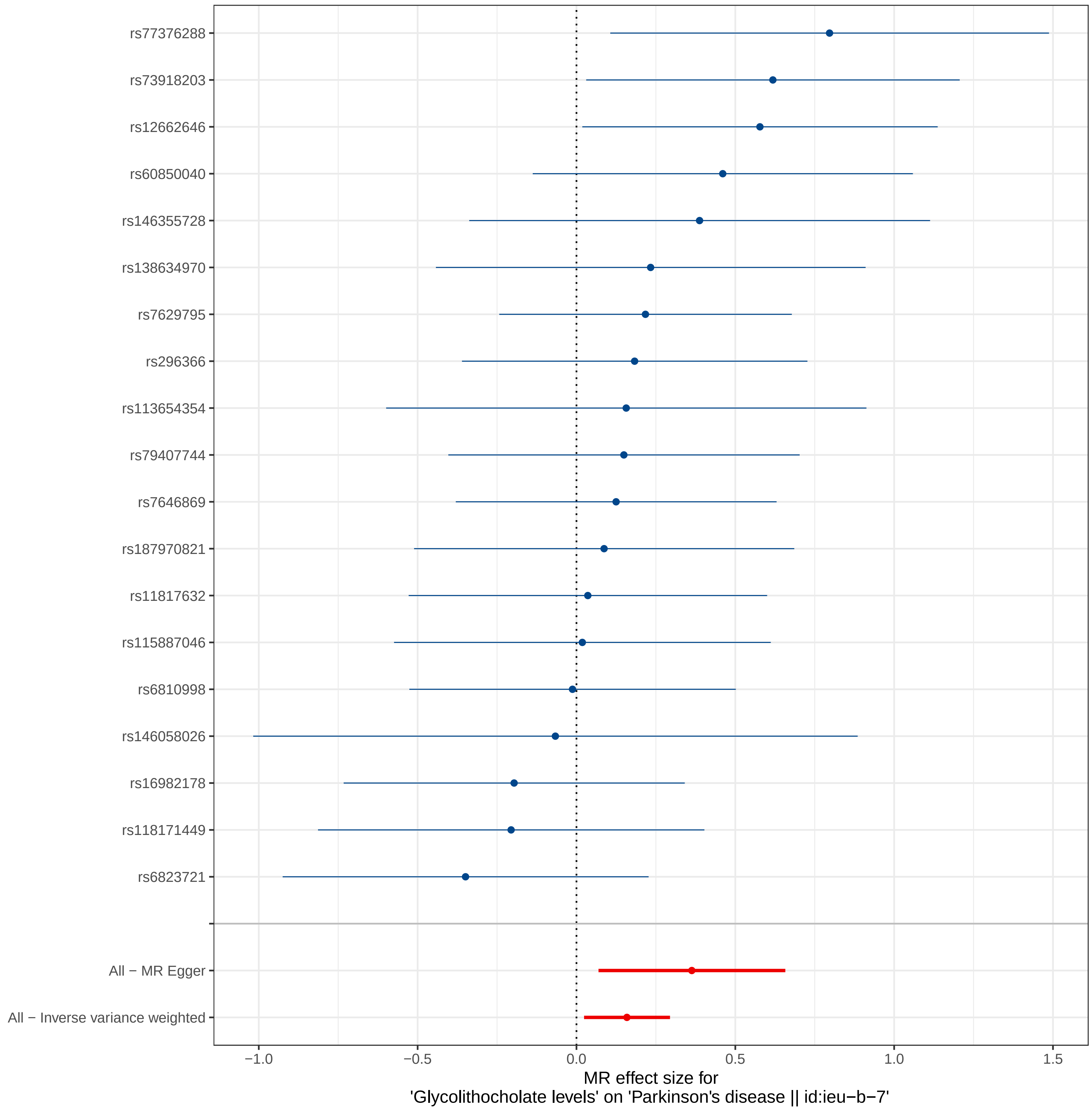
rs77376288
rs73918203
rs12662646
rs60850040
rs146355728
rs138634970
rs7629795
rs296366
rs113654354
rs79407744
rs7646869
rs187970821
rs11817632
rs115887046
rs6810998
rs146058026
rs16982178
rs118171449
rs6823721
All − MR Egger
All − Inverse variance weighted
−1.0
−0.5
0.0
0.5
1.0
1.5
MR effect size for
'Glycolithocholate levels' on 'Parkinson's disease || id:ieu−b−7'

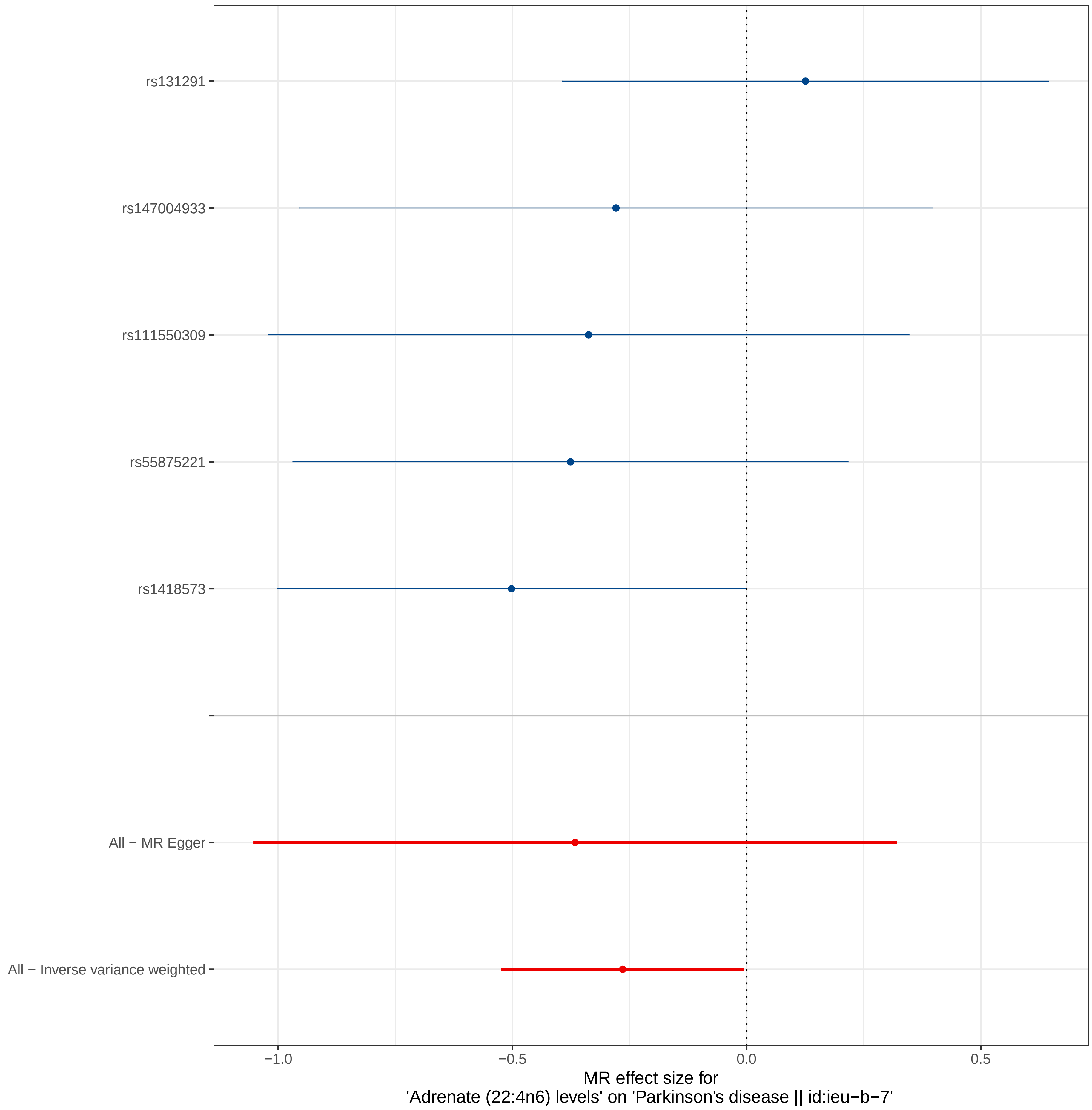

rs131291
rs147004933
rs111550309
rs55875221
rs1418573
All − MR Egger
All − Inverse variance weighted
−1.0
−0.5
0.0
0.5
MR effect size for
'Adrenate (22:4n6) levels' on 'Parkinson's disease || id:ieu−b−7'

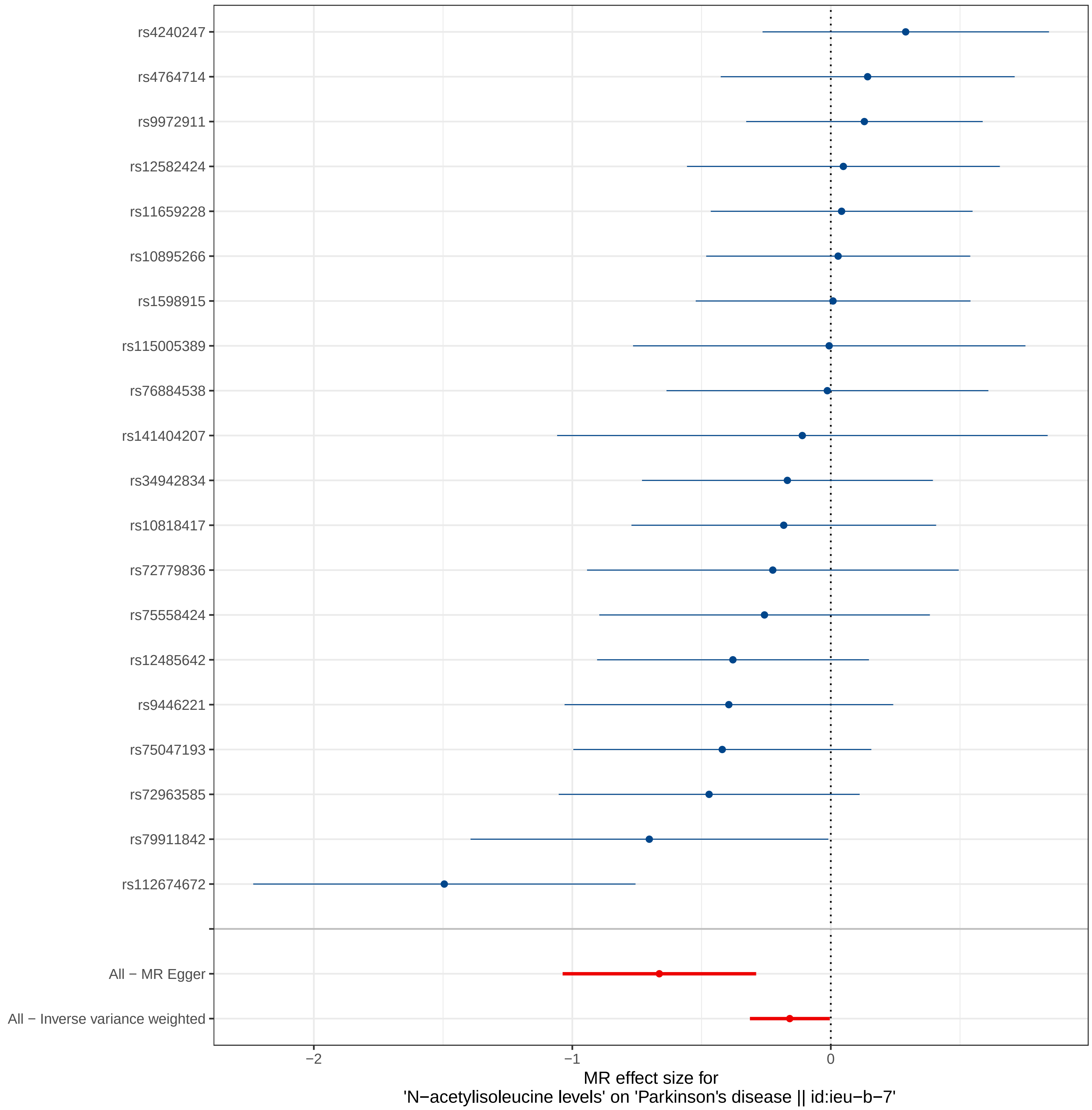

rs4240247
rs4764714
rs9972911
rs12582424
rs11659228
rs10895266
rs1598915
rs115005389
rs76884538
rs141404207
rs34942834
rs10818417
rs72779836
rs75558424
rs12485642
rs9446221
rs75047193
rs72963585
rs79911842
rs112674672
All − MR Egger
All − Inverse variance weighted
−2
−1
0
MR effect size for
'N−acetylisoleucine levels' on 'Parkinson's disease || id:ieu−b−7'

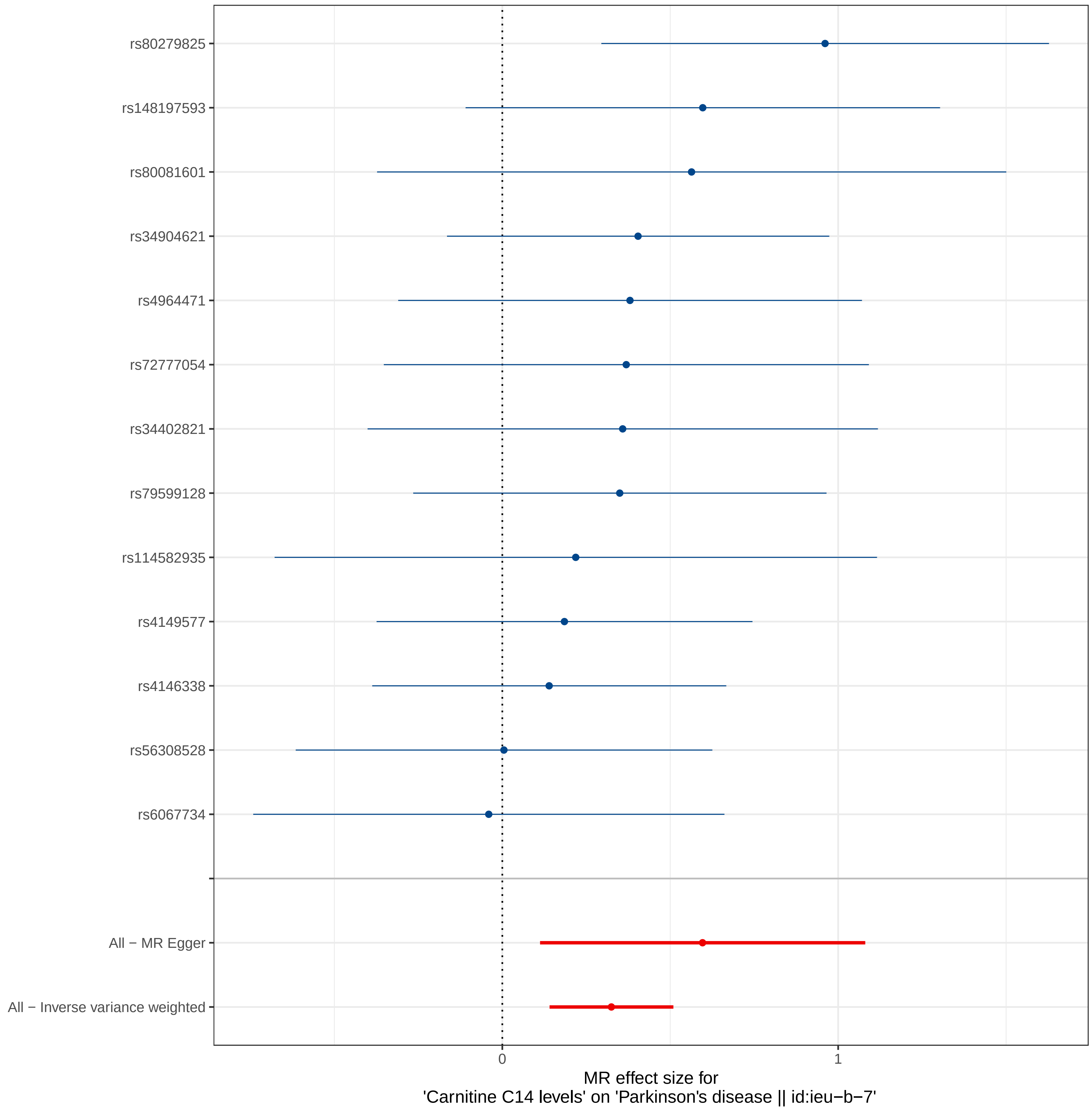

rs80279825
rs148197593
rs80081601
rs34904621
rs4964471
rs72777054
rs34402821
rs79599128
rs114582935
rs4149577
rs4146338
rs56308528
rs6067734
All − MR Egger
All − Inverse variance weighted
0
1
MR effect size for
'Carnitine C14 levels' on 'Parkinson's disease || id:ieu−b−7'

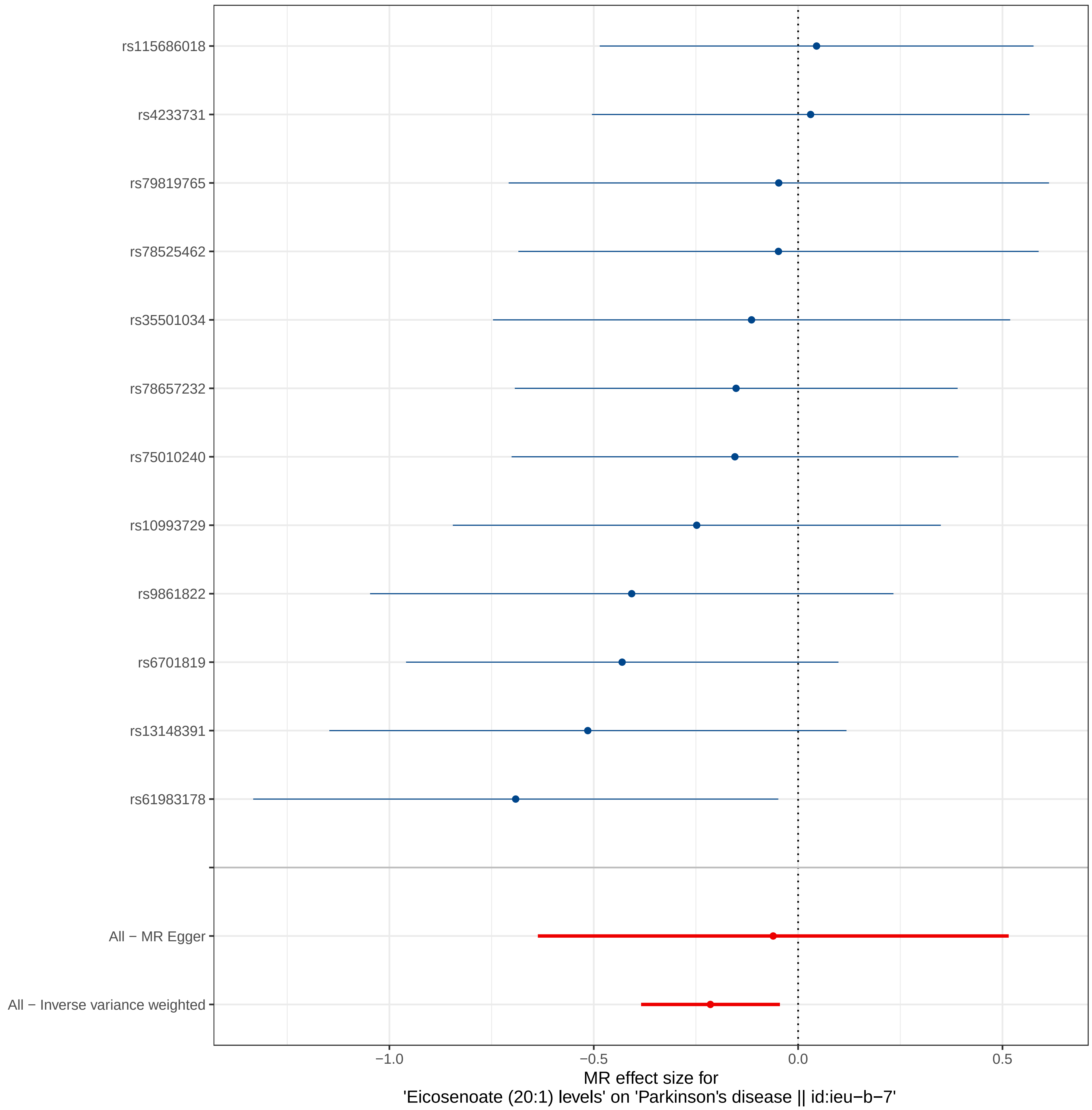
rs115686018
rs4233731
rs79819765
rs78525462
rs35501034
rs78657232
rs75010240
rs10993729
rs9861822
rs6701819
rs13148391
rs61983178
All − MR Egger
All − Inverse variance weighted
−1.0
−0.5
0.0
0.5
MR effect size for
'Eicosenoate (20:1) levels' on 'Parkinson's disease || id:ieu−b−7'

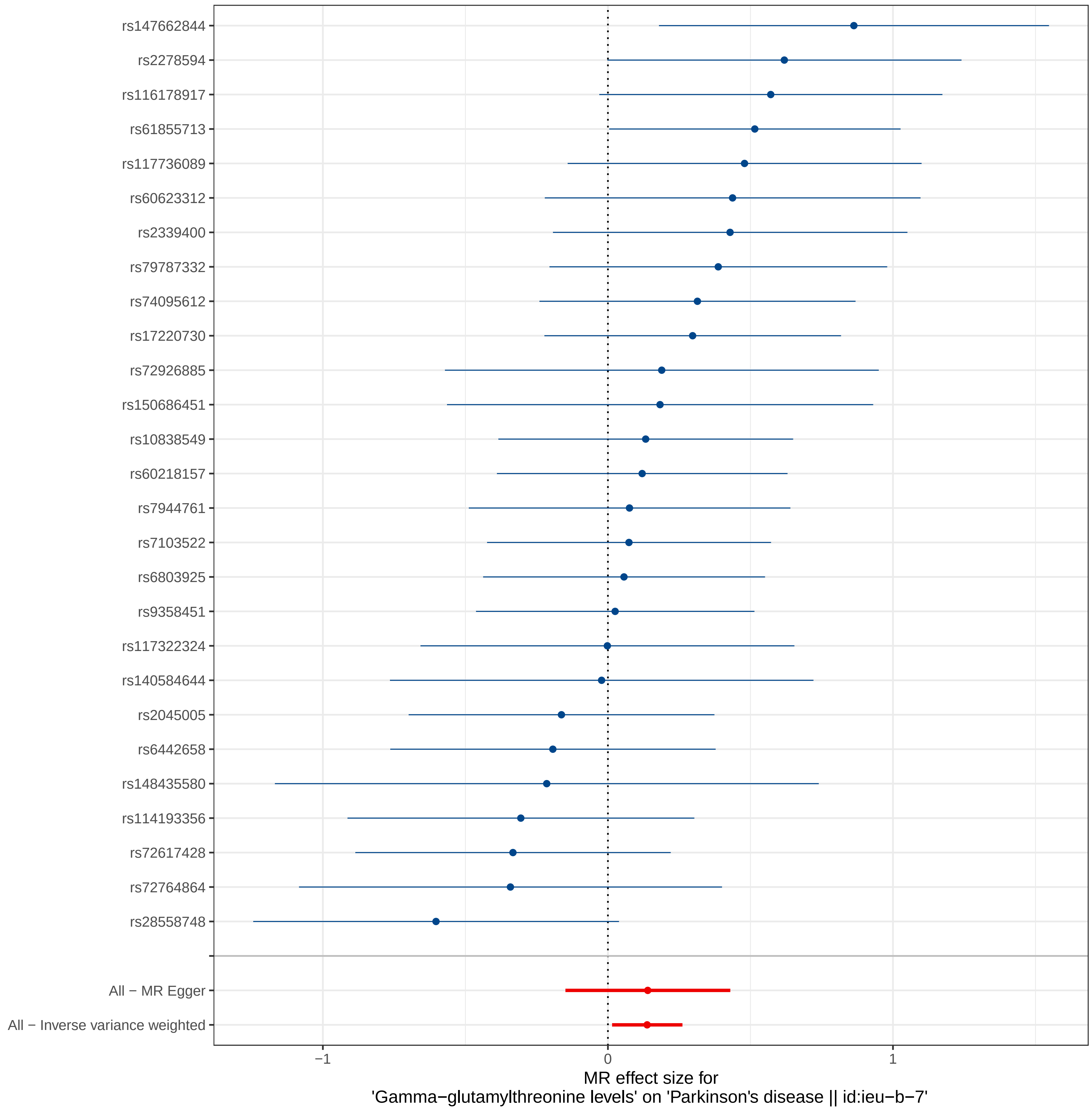
rs147662844
rs2278594
rs116178917
rs61855713
rs117736089
rs60623312
rs2339400
rs79787332
rs74095612
rs17220730
rs72926885
rs150686451
rs10838549
rs60218157
rs7944761
rs7103522
rs6803925
rs9358451
rs117322324
rs140584644
rs2045005
rs6442658
rs148435580
rs114193356
rs72617428
rs72764864
rs28558748
All − MR Egger
All − Inverse variance weighted
−1
0
1
MR effect size for
'Gamma−glutamylthreonine levels' on 'Parkinson's disease || id:ieu−b−7'

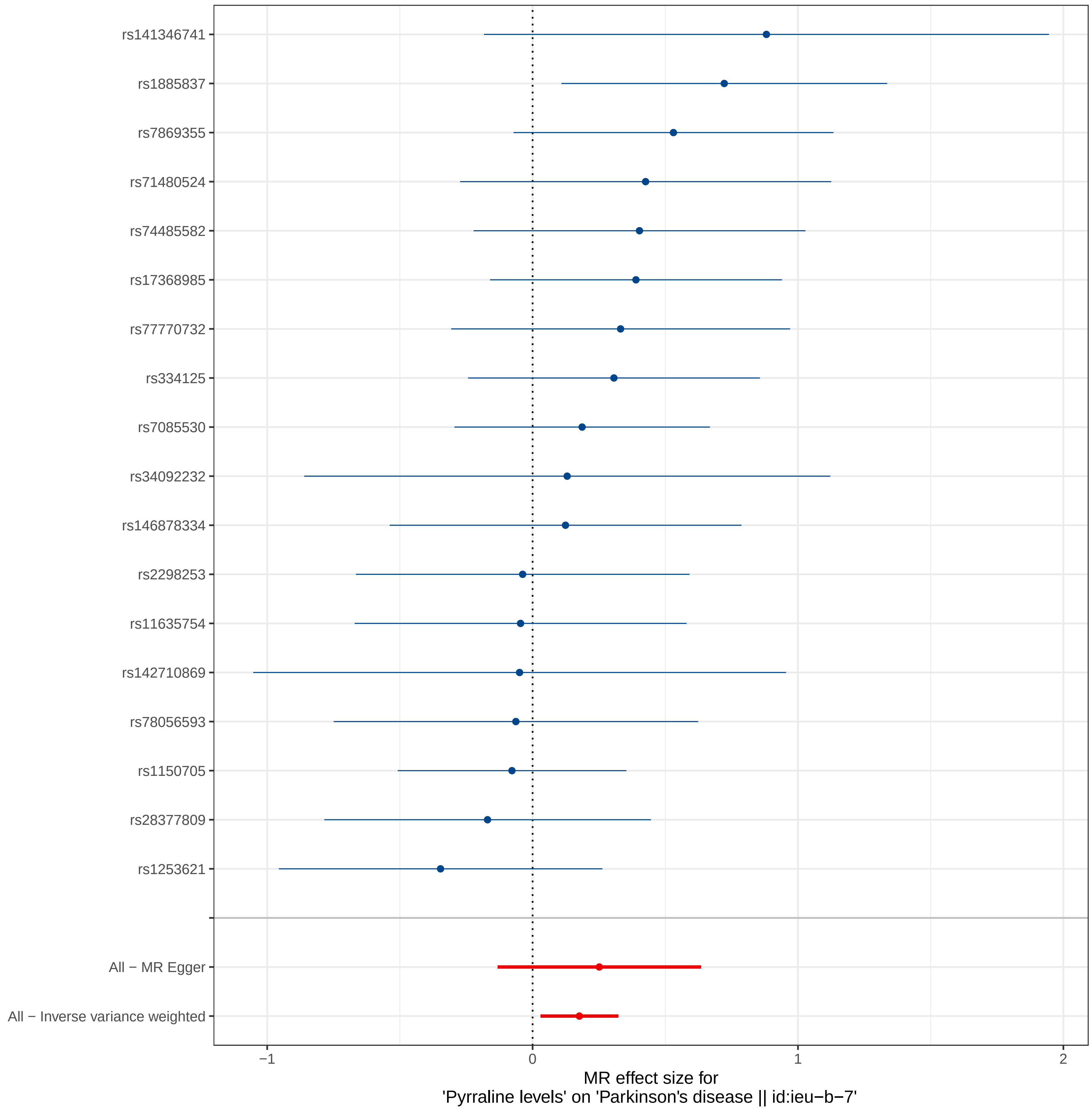

rs141346741
rs1885837
rs7869355
rs71480524
rs74485582
rs17368985
rs77770732
rs334125
rs7085530
rs34092232
rs146878334
rs2298253
rs11635754
rs142710869
rs78056593
rs1150705
rs28377809
rs1253621
All − MR Egger
All − Inverse variance weighted
−1
0
1
2
MR effect size for
'Pyrraline levels' on 'Parkinson's disease || id:ieu−b−7'

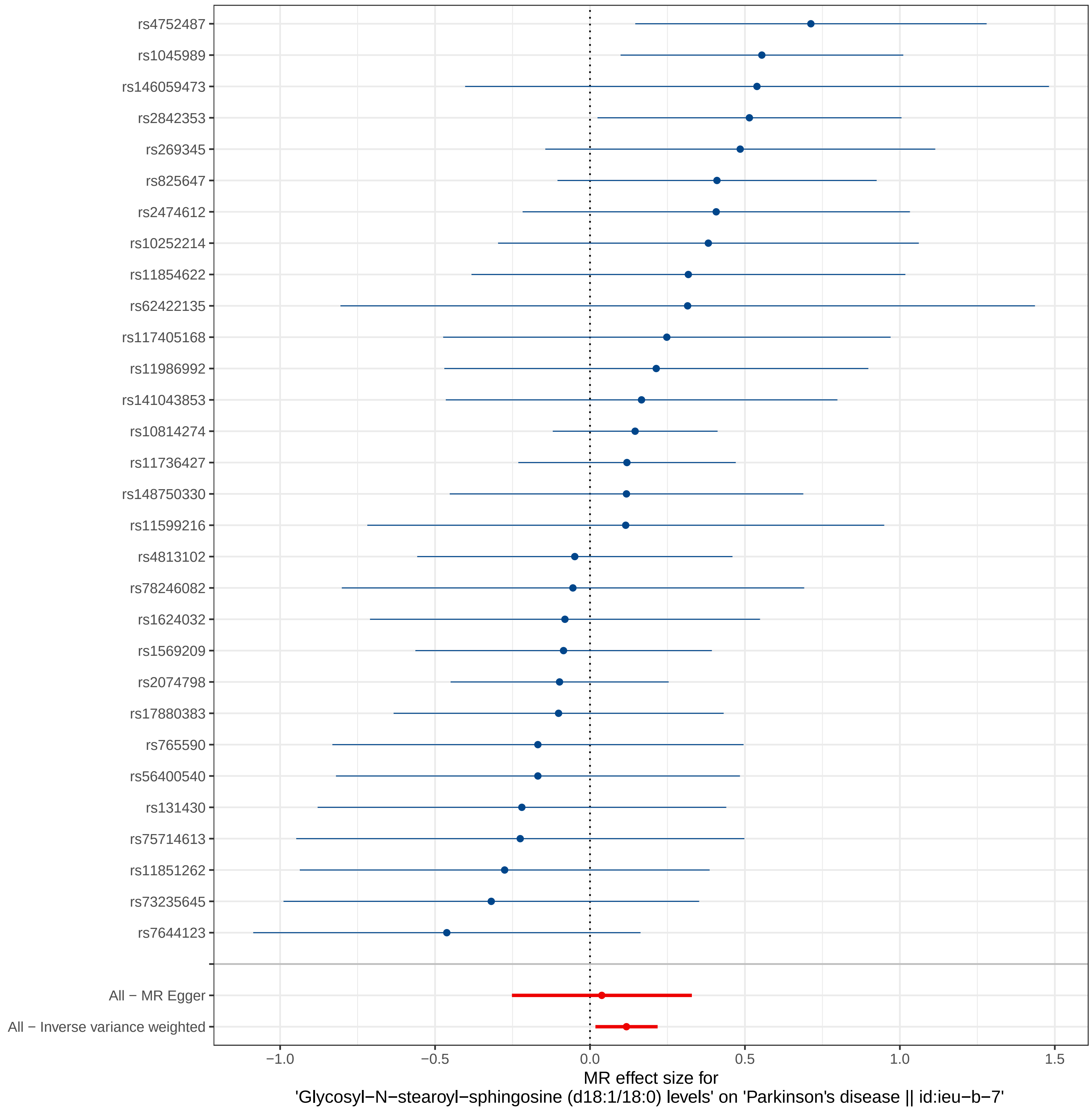
rs4752487
rs1045989
rs146059473
rs2842353
rs269345
rs825647
rs2474612
rs10252214
rs11854622
rs62422135
rs117405168
rs11986992
rs141043853
rs10814274
rs11736427
rs148750330
rs11599216
rs4813102
rs78246082
rs1624032
rs1569209
rs2074798
rs17880383
rs765590
rs56400540
rs131430
rs75714613
rs11851262
rs73235645
rs7644123
All − MR Egger
All − Inverse variance weighted
−1.0
−0.5
0.0
0.5
1.0
1.5
MR effect size for
'Glycosyl−N−stearoyl−sphingosine (d18:1/18:0) levels' on 'Parkinson's disease || id:ieu−b−7'

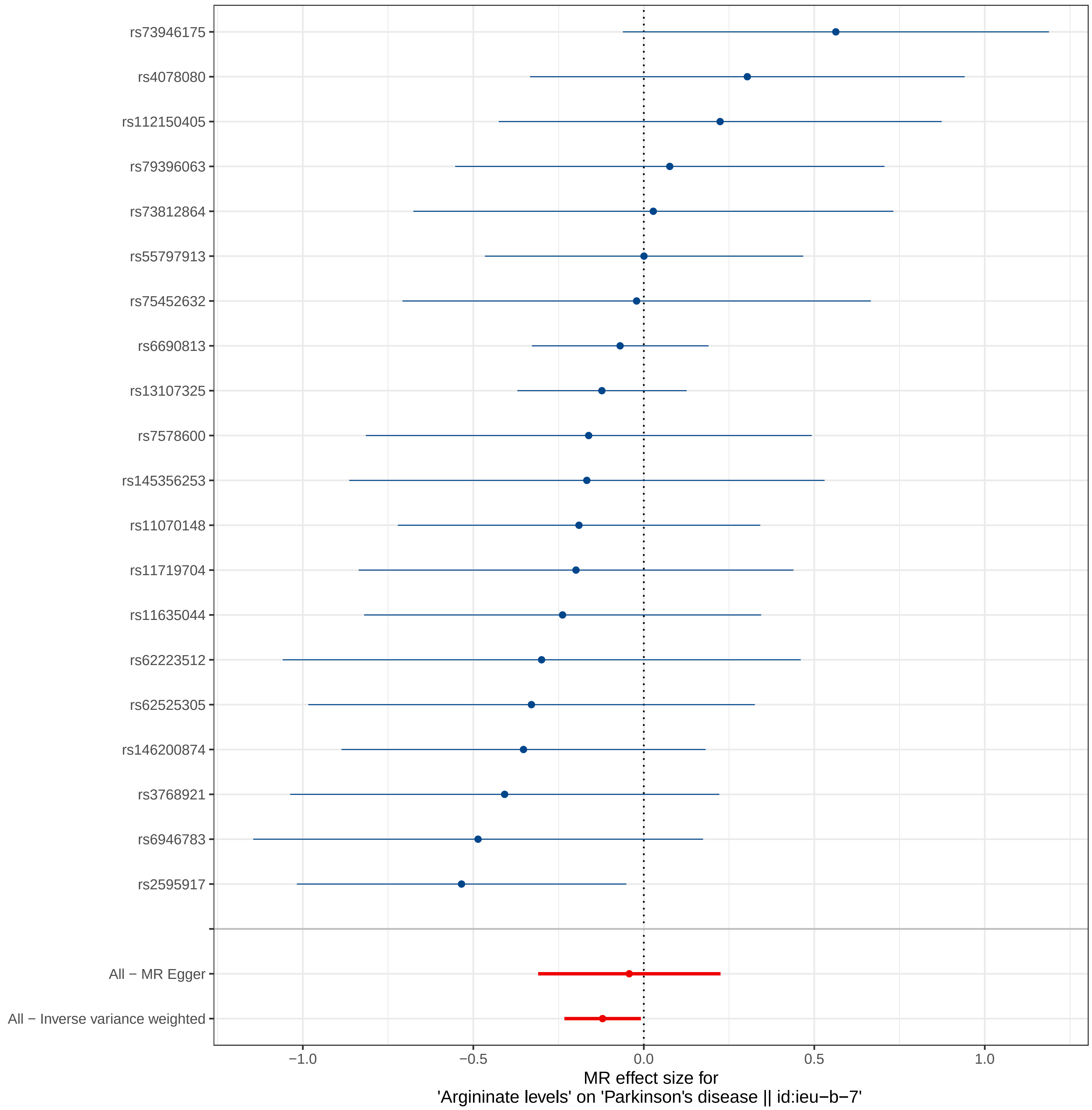

rs73946175
rs4078080
rs112150405
rs79396063
rs73812864
rs55797913
rs75452632
rs6690813
rs13107325
rs7578600
rs145356253
rs11070148
rs11719704
rs11635044
rs62223512
rs62525305
rs146200874
rs3768921
rs6946783
rs2595917
All − MR Egger
All − Inverse variance weighted
−1.0
−0.5
0.0
0.5
1.0
MR effect size for
'Argininate levels' on 'Parkinson's disease || id:ieu−b−7'

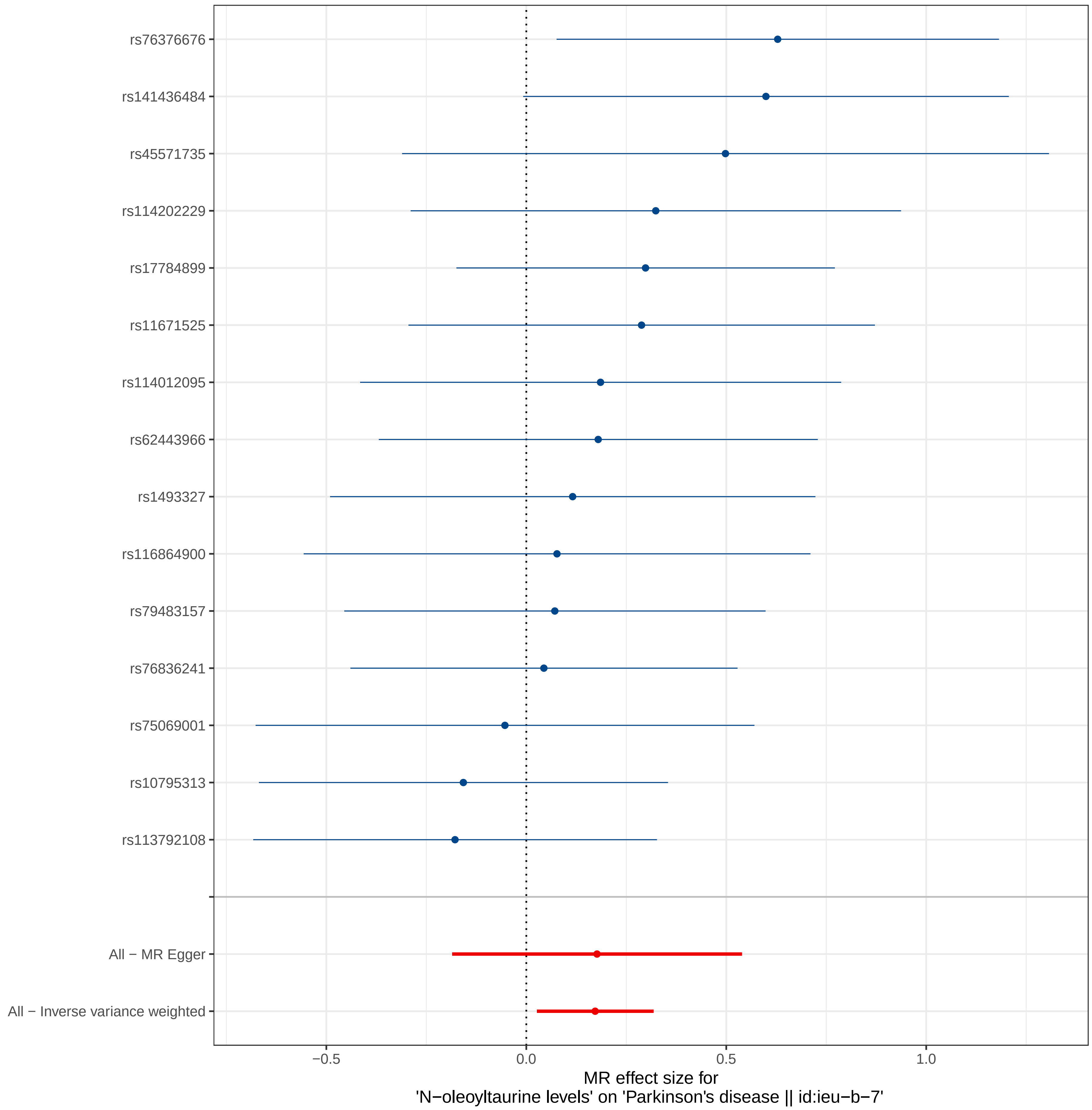

rs76376676
rs141436484
rs45571735
rs114202229
rs17784899
rs11671525
rs114012095
rs62443966
rs1493327
rs116864900
rs79483157
rs76836241
rs75069001
rs10795313
rs113792108
All − MR Egger
All − Inverse variance weighted
−0.5
0.0
0.5
1.0
MR effect size for
'N−oleoyltaurine levels' on 'Parkinson's disease || id:ieu−b−7'

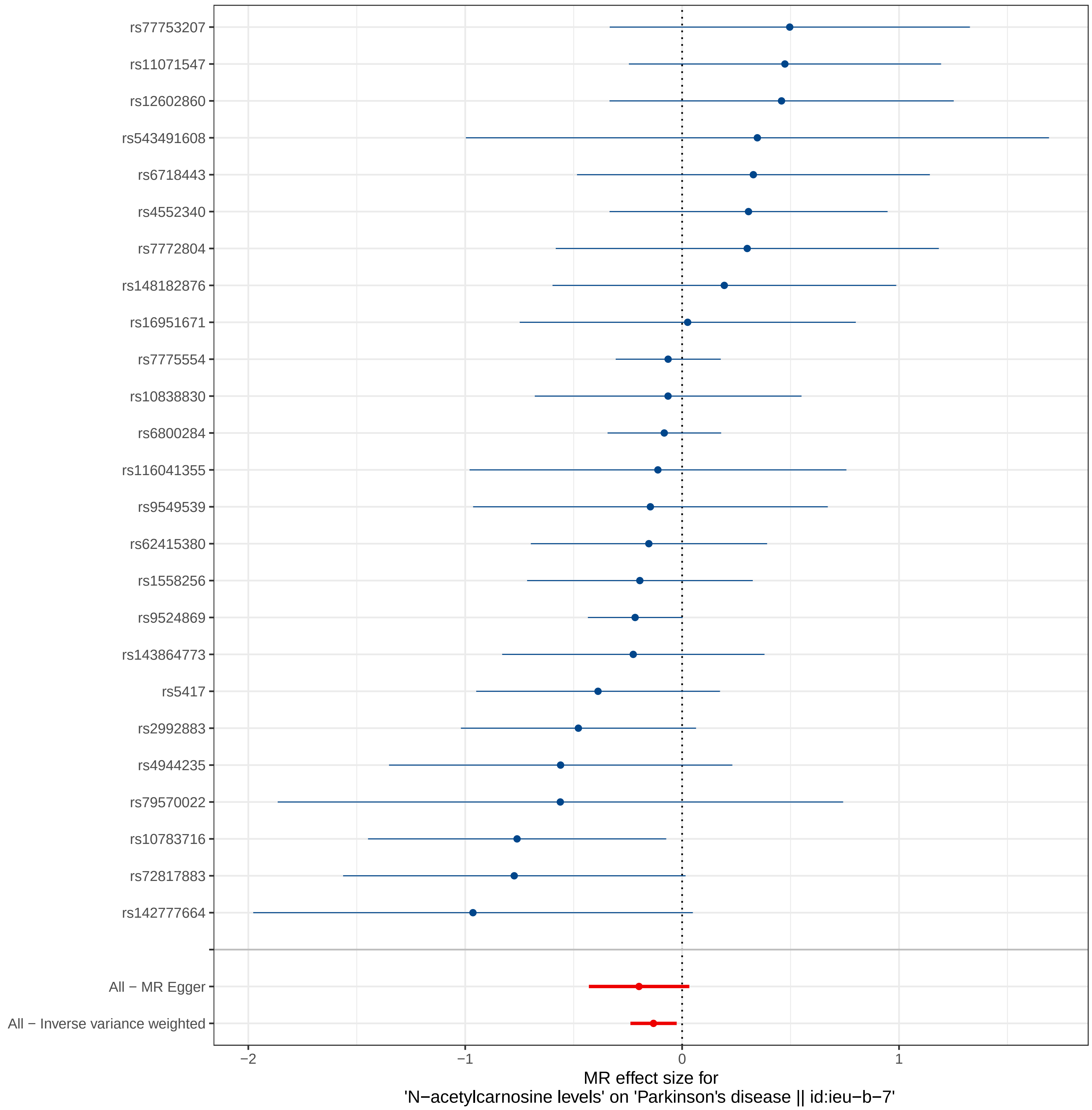
rs77753207
rs11071547
rs12602860
rs543491608
rs6718443
rs4552340
rs7772804
rs148182876
rs16951671
rs7775554
rs10838830
rs6800284
rs116041355
rs9549539
rs62415380
rs1558256
rs9524869
rs143864773
rs5417
rs2992883
rs4944235
rs79570022
rs10783716
rs72817883
rs142777664
All − MR Egger
All − Inverse variance weighted
−2
−1
0
1
MR effect size for
'N−acetylcarnosine levels' on 'Parkinson's disease || id:ieu−b−7'

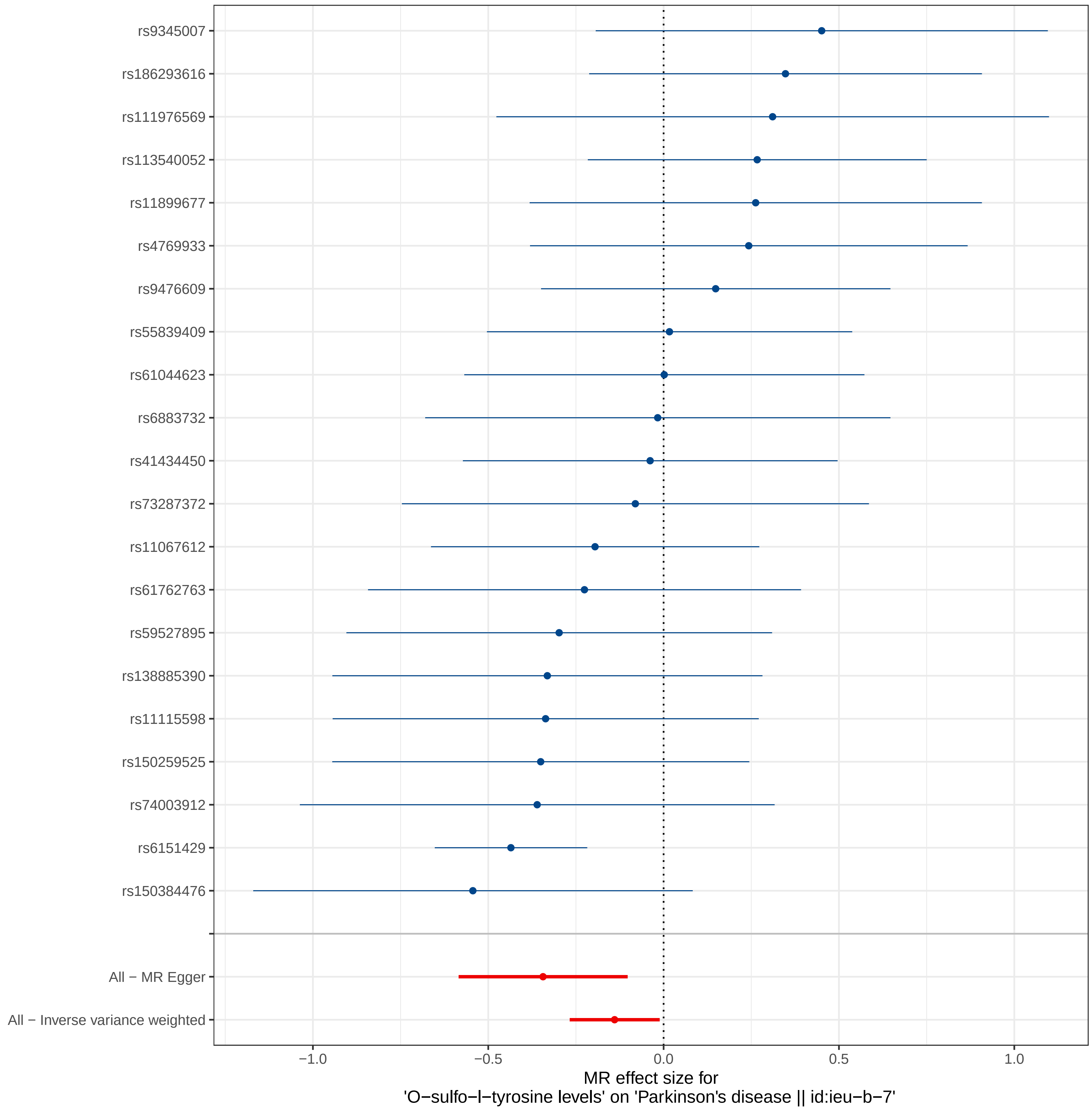
rs9345007
rs186293616
rs111976569
rs113540052
rs11899677
rs4769933
rs9476609
rs55839409
rs61044623
rs6883732
rs41434450
rs73287372
rs11067612
rs61762763
rs59527895
rs138885390
rs11115598
rs150259525
rs74003912
rs6151429
rs150384476
All − MR Egger
All − Inverse variance weighted
−1.0
−0.5
0.0
0.5
1.0
MR effect size for
'O−sulfo−l−tyrosine levels' on 'Parkinson's disease || id:ieu−b−7'

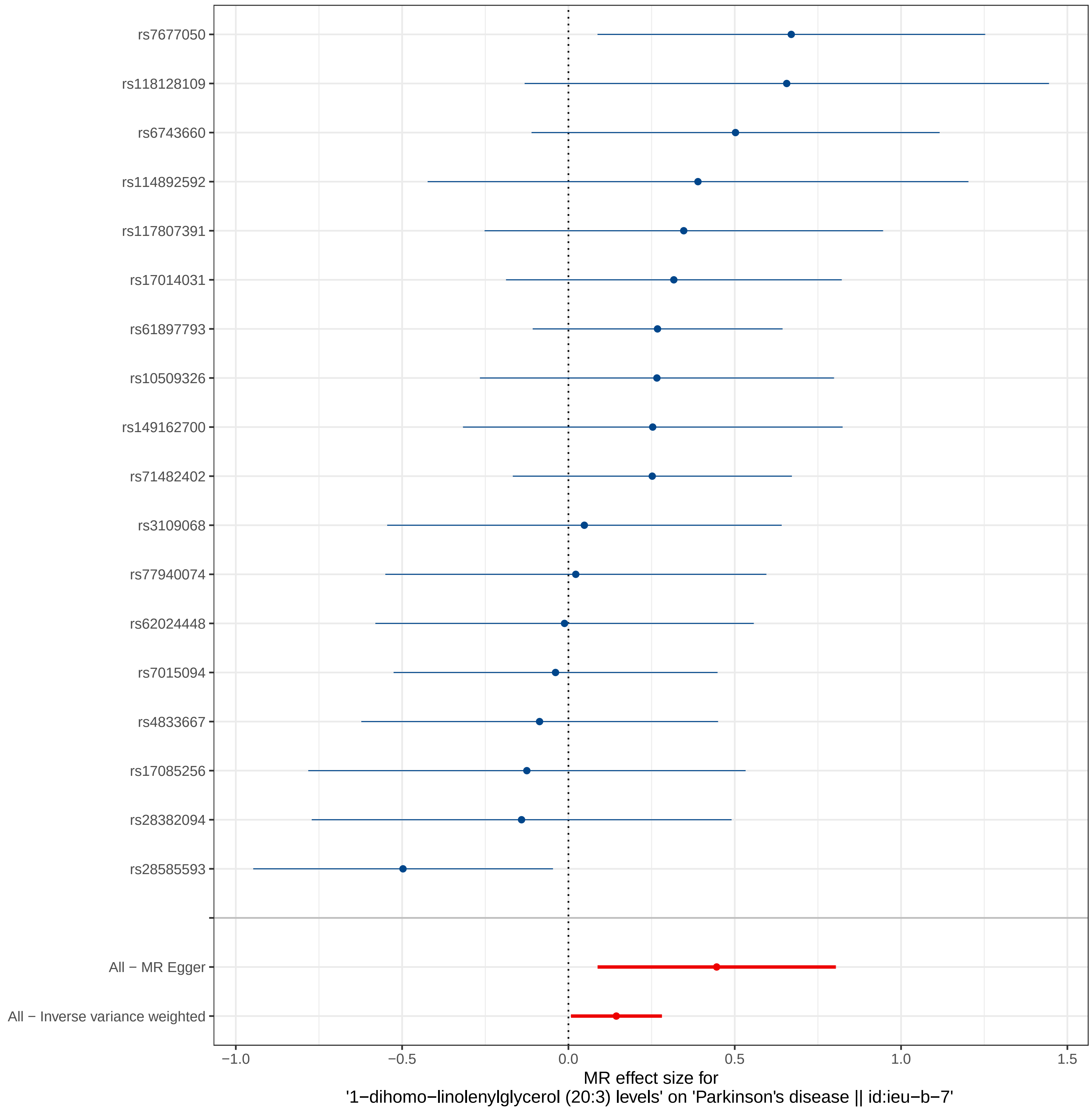
rs7677050
rs118128109
rs6743660
rs114892592
rs117807391
rs17014031
rs61897793
rs10509326
rs149162700
rs71482402
rs3109068
rs77940074
rs62024448
rs7015094
rs4833667
rs17085256
rs28382094
rs28585593
All − MR Egger
All − Inverse variance weighted
−1.0
−0.5
0.0
0.5
1.0
1.5
MR effect size for
'1−dihomo−linolenylglycerol (20:3) levels' on 'Parkinson's disease || id:ieu−b−7'

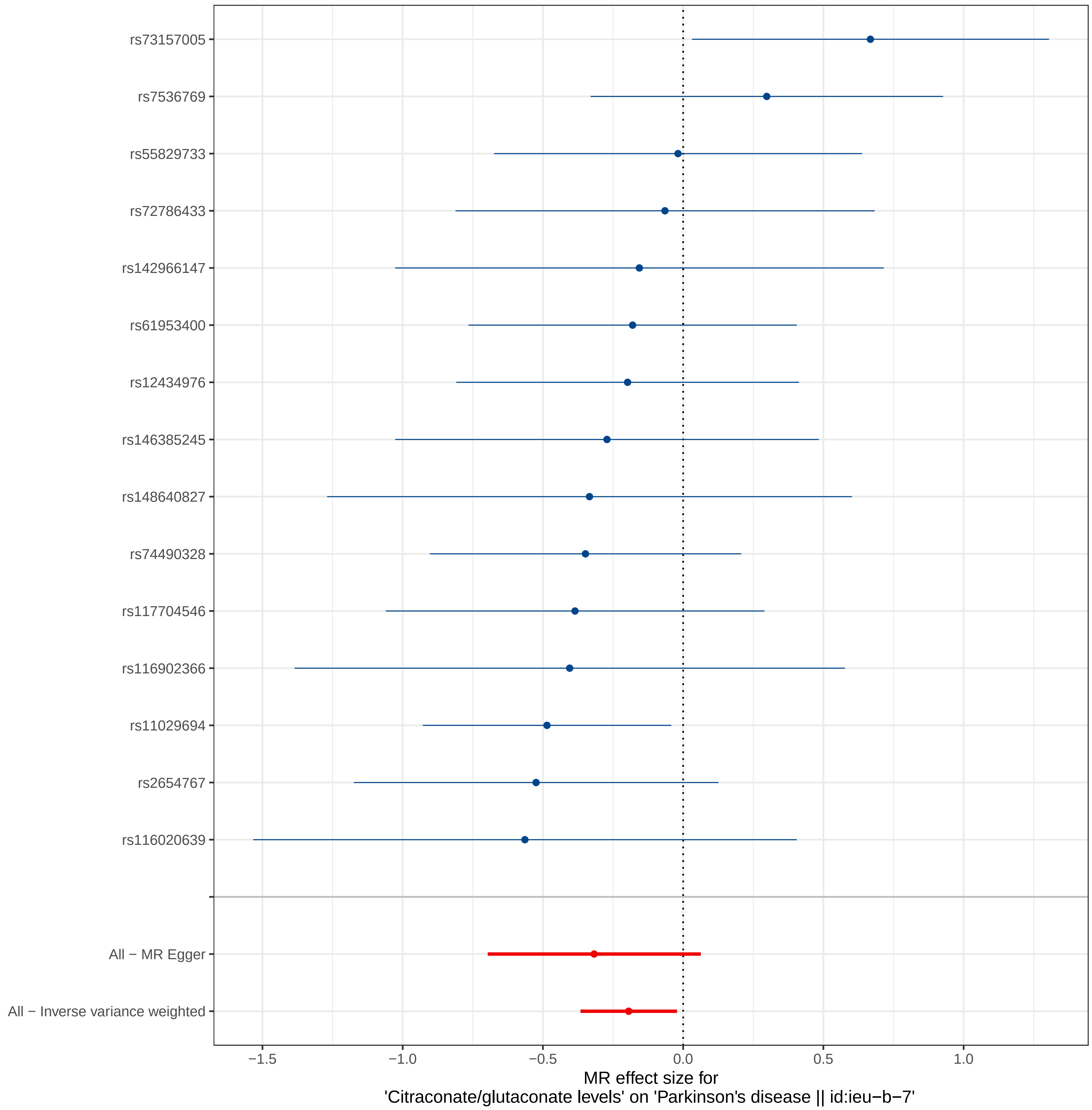

rs73157005
rs7536769
rs55829733
rs72786433
rs142966147
rs61953400
rs12434976
rs146385245
rs148640827
rs74490328
rs117704546
rs116902366
rs11029694
rs2654767
rs116020639
All − MR Egger
All − Inverse variance weighted
−1.5
−1.0
−0.5
0.0
0.5
1.0
MR effect size for
'Citraconate/glutaconate levels' on 'Parkinson's disease || id:ieu−b−7'

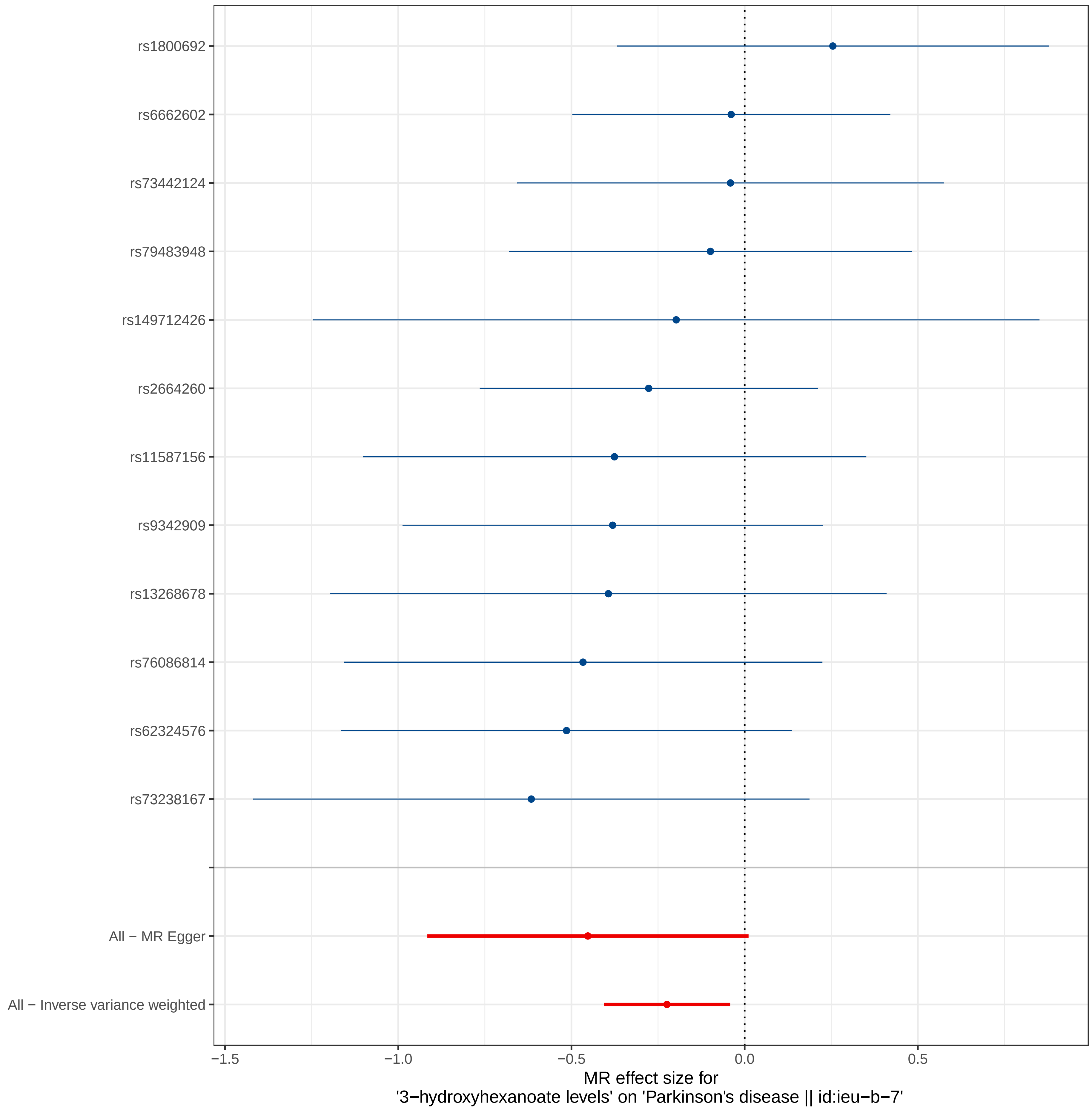
rs1800692
rs6662602
rs73442124
rs79483948
rs149712426
rs2664260
rs11587156
rs9342909
rs13268678
rs76086814
rs62324576
rs73238167
All − MR Egger
All − Inverse variance weighted
−1.5
−1.0
−0.5
0.0
0.5
MR effect size for
'3−hydroxyhexanoate levels' on 'Parkinson's disease || id:ieu−b−7'

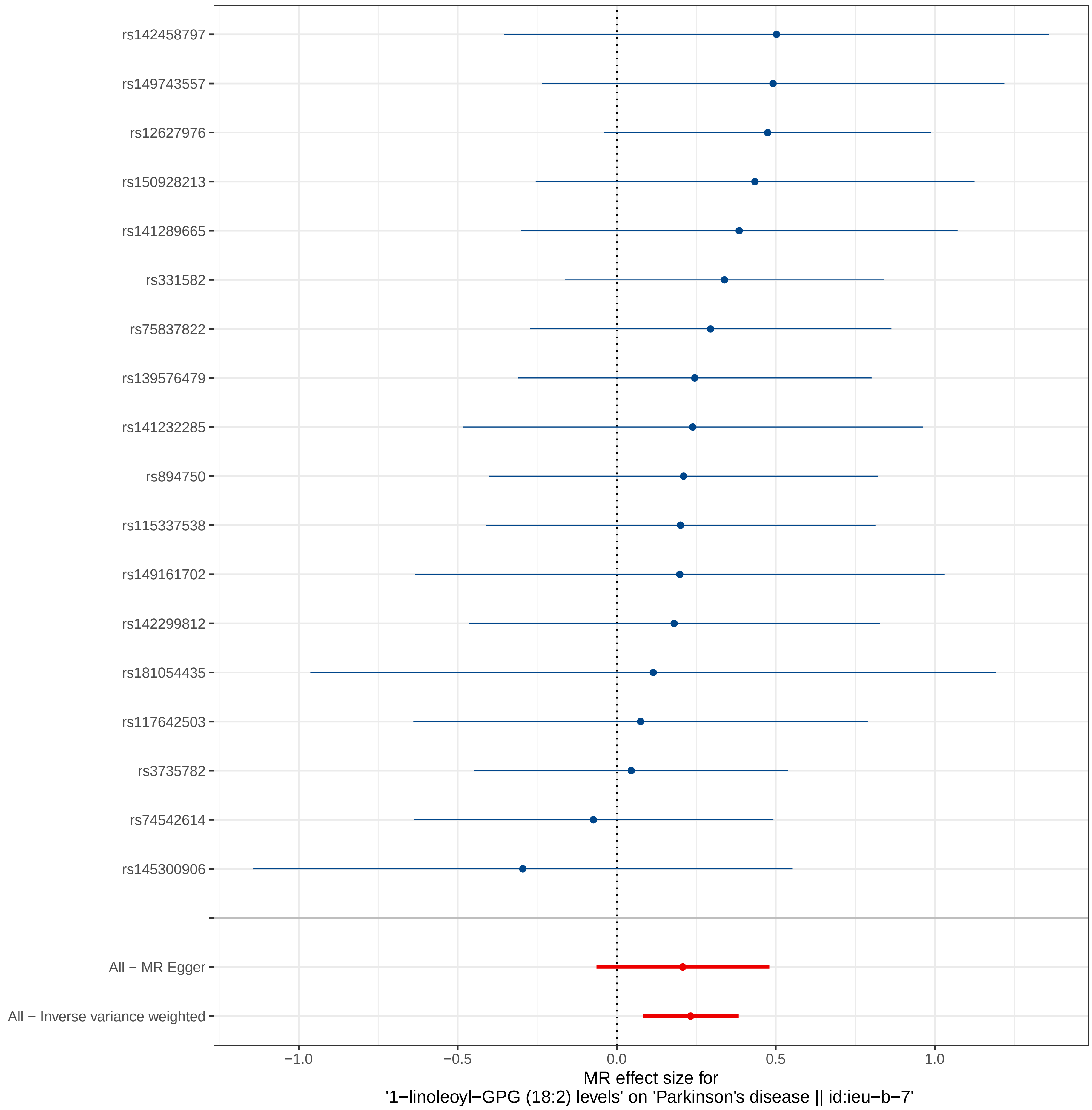
rs142458797
rs149743557
rs12627976
rs150928213
rs141289665
rs331582
rs75837822
rs139576479
rs141232285
rs894750
rs115337538
rs149161702
rs142299812
rs181054435
rs117642503
rs3735782
rs74542614
rs145300906
All − MR Egger
All − Inverse variance weighted
−1.0
−0.5
0.0
0.5
1.0
MR effect size for
'1−linoleoyl−GPG (18:2) levels' on 'Parkinson's disease || id:ieu−b−7'

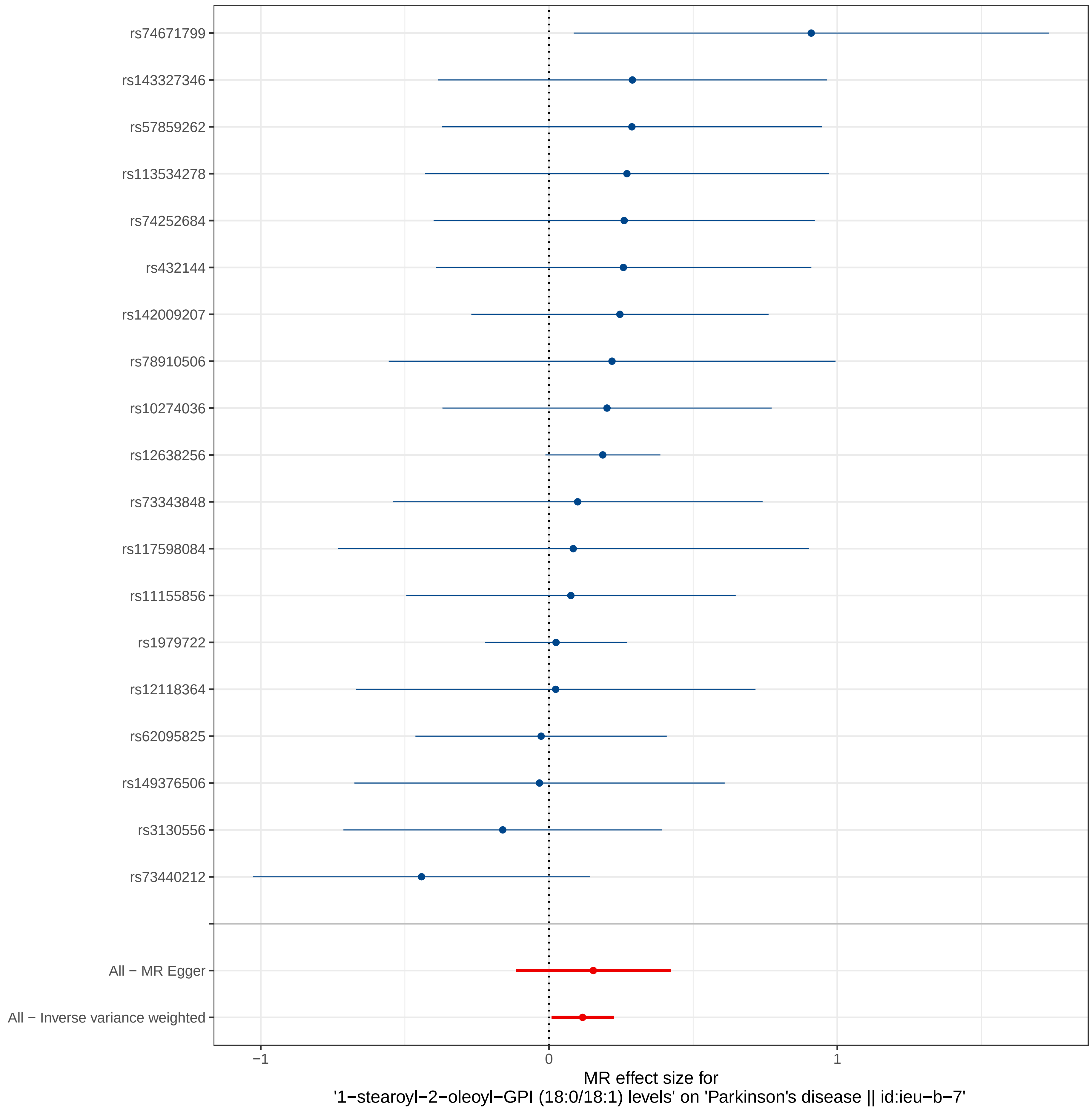
rs74671799
rs143327346
rs57859262
rs113534278
rs74252684
rs432144
rs142009207
rs78910506
rs10274036
rs12638256
rs73343848
rs117598084
rs11155856
rs1979722
rs12118364
rs62095825
rs149376506
rs3130556
rs73440212
All − MR Egger
All − Inverse variance weighted
−1
0
1
MR effect size for
'1−stearoyl−2−oleoyl−GPI (18:0/18:1) levels' on 'Parkinson's disease || id:ieu−b−7'

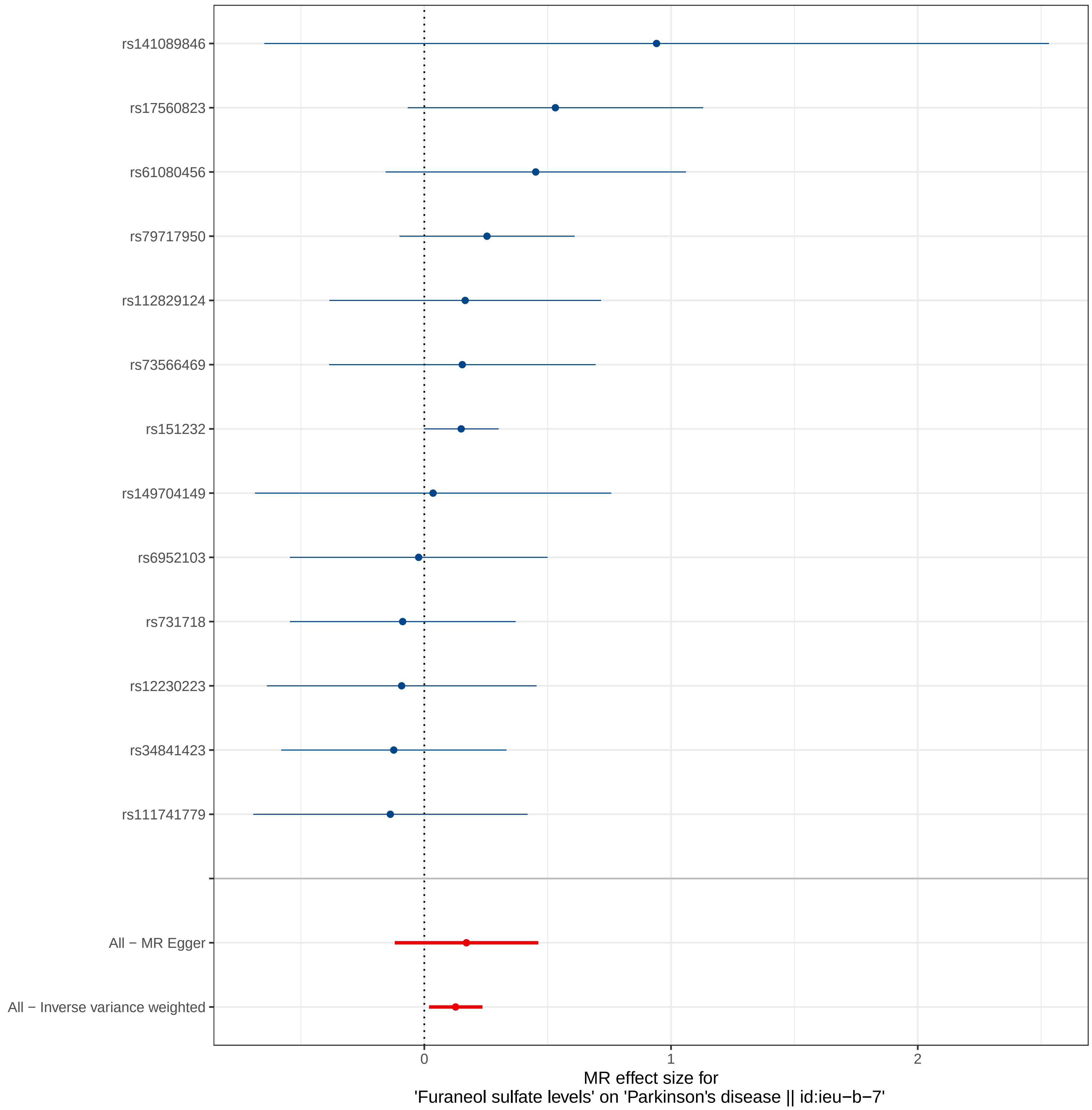
rs141089846
rs17560823
rs61080456
rs79717950
rs112829124
rs73566469
rs151232
rs149704149
rs6952103
rs731718
rs12230223
rs34841423
rs111741779
All − MR Egger
All − Inverse variance weighted
0
1
2
MR effect size for
'Furaneol sulfate levels' on 'Parkinson's disease || id:ieu−b−7'

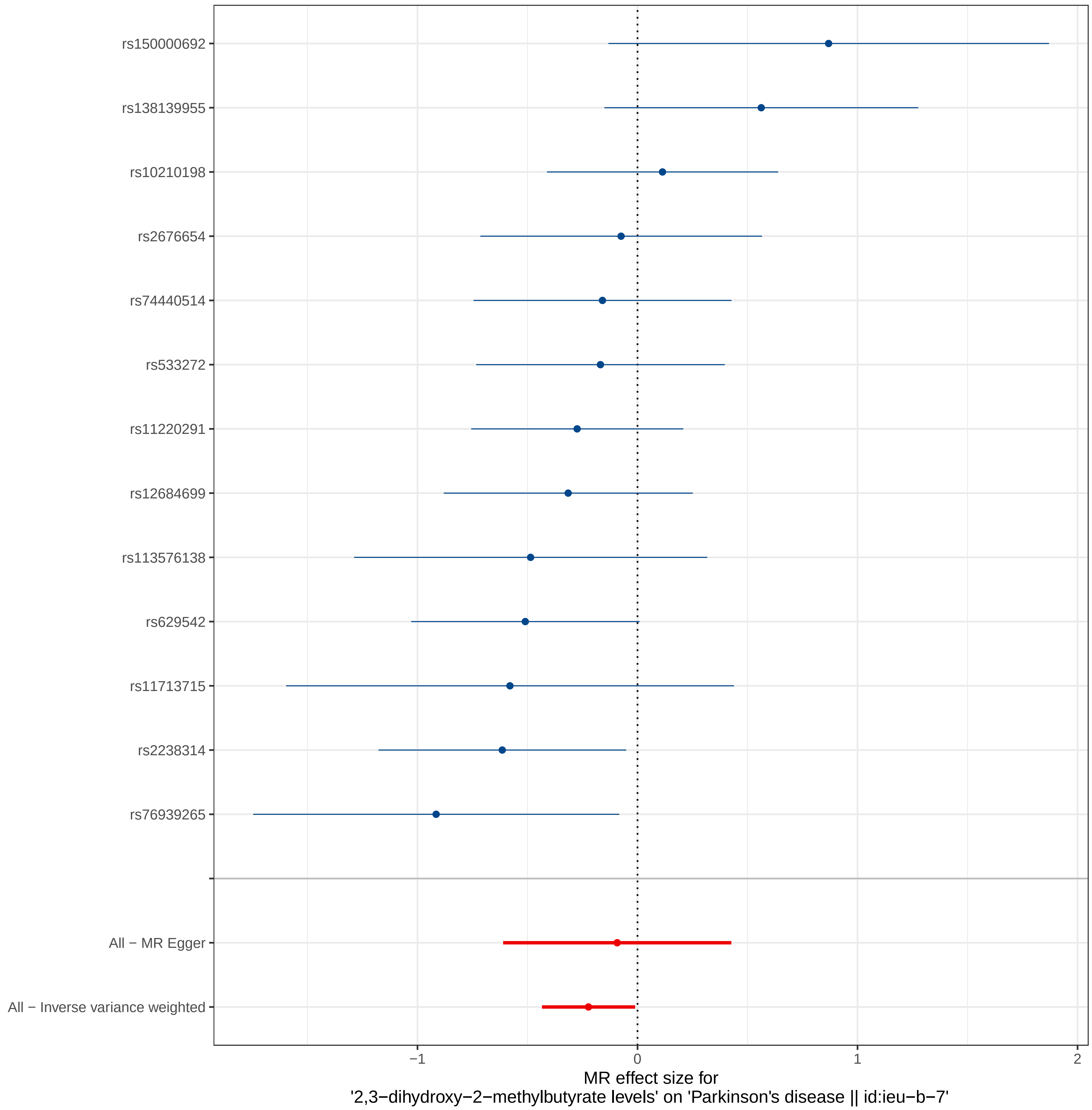
rs150000692
rs138139955
rs10210198
rs2676654
rs74440514
rs533272
rs11220291
rs12684699
rs113576138
rs629542
rs11713715
rs2238314
rs76939265
All − MR Egger
All − Inverse variance weighted
−1
0
1
2
MR effect size for
'2,3−dihydroxy−2−methylbutyrate levels' on 'Parkinson's disease || id:ieu−b−7'

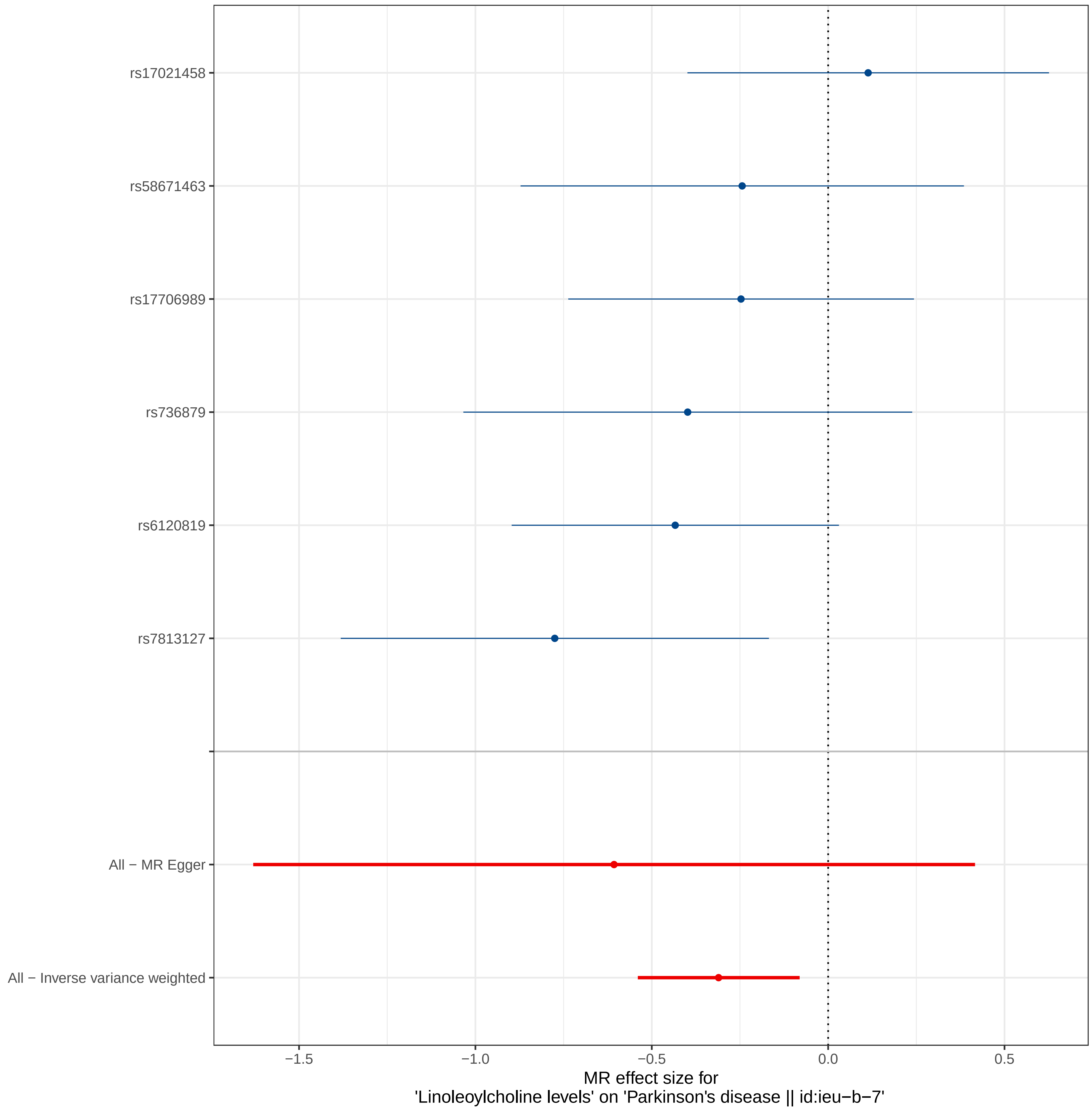
rs17021458
rs58671463
rs17706989
rs736879
rs6120819
rs7813127
All − MR Egger
All − Inverse variance weighted
−1.5
−1.0
−0.5
0.0
0.5
MR effect size for
'Linoleoylcholine levels' on 'Parkinson's disease || id:ieu−b−7'

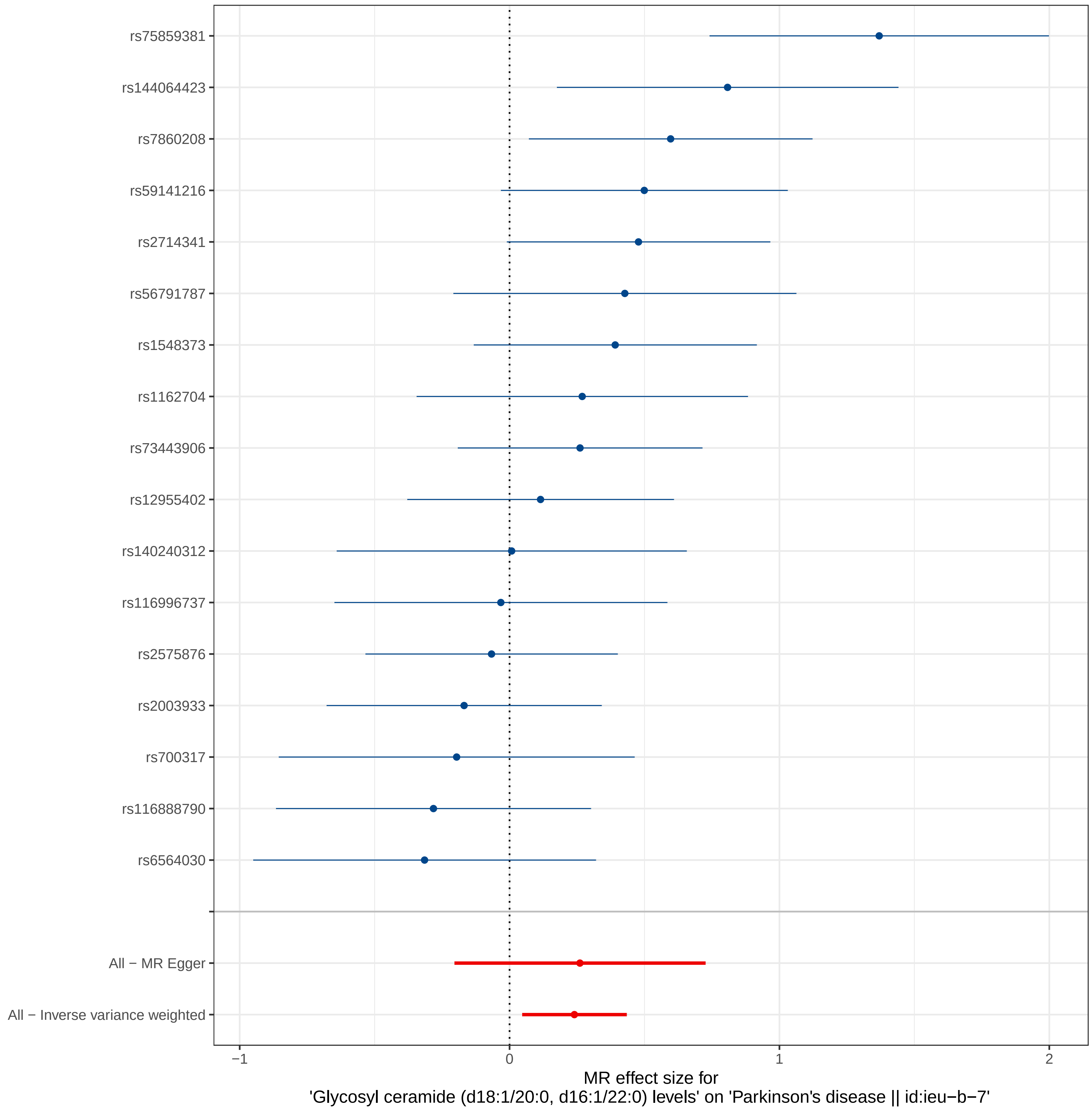
rs75859381
rs144064423
rs7860208
rs59141216
rs2714341
rs56791787
rs1548373
rs1162704
rs73443906
rs12955402
rs140240312
rs116996737
rs2575876
rs2003933
rs700317
rs116888790
rs6564030
All − MR Egger
All − Inverse variance weighted
−1
0
1
2
MR effect size for
'Glycosyl ceramide (d18:1/20:0, d16:1/22:0) levels' on 'Parkinson's disease || id:ieu−b−7'

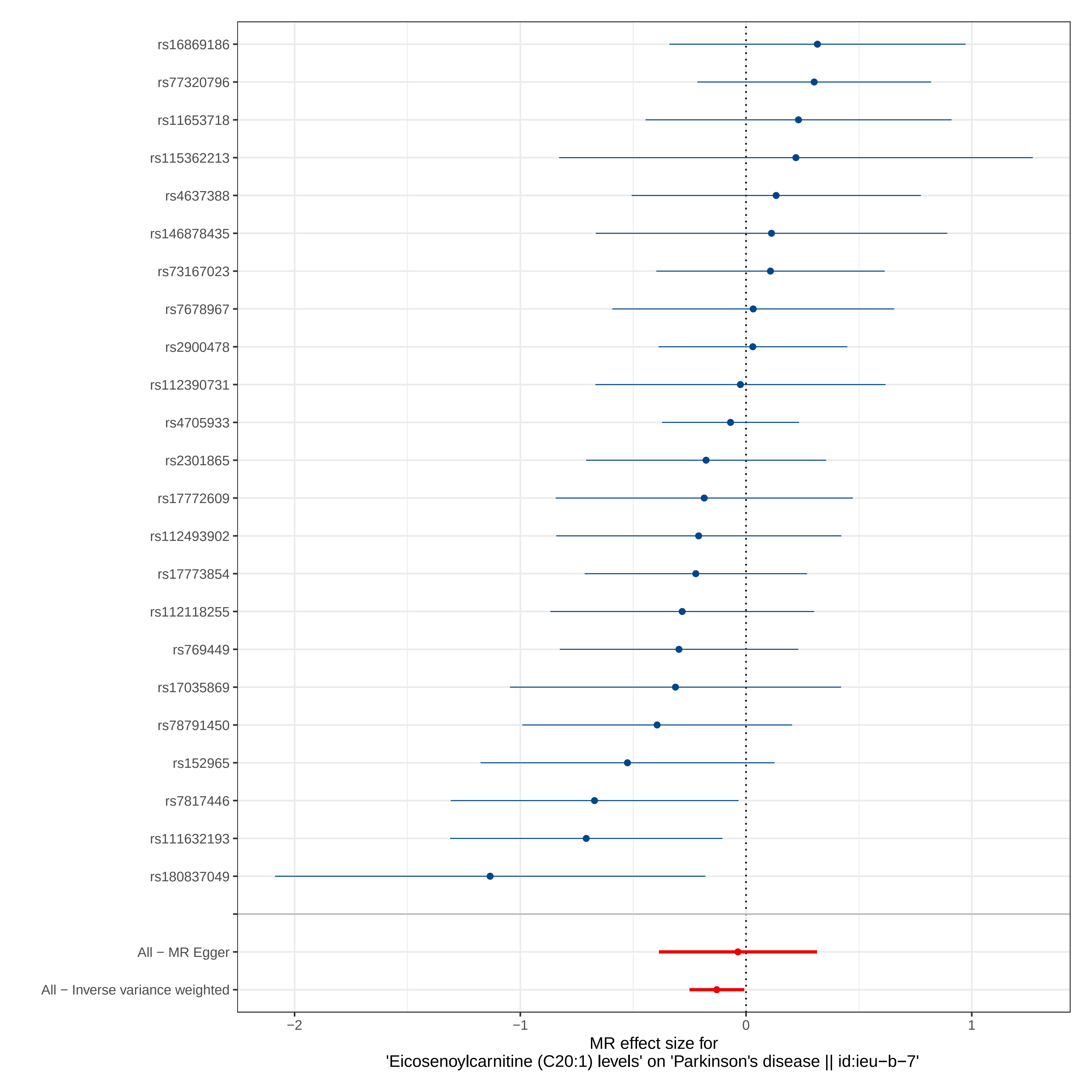
rs16869186
rs77320796
rs11653718
rs115362213
rs4637388
rs146878435
rs73167023
rs7678967
rs2900478
rs112390731
rs4705933
rs2301865
rs17772609
rs112493902
rs17773854
rs112118255
rs769449
rs17035869
rs78791450
rs152965
rs7817446
rs111632193
rs180837049
All − MR Egger
All − Inverse variance weighted
−2
−1
0
1
MR effect size for
'Eicosenoylcarnitine (C20:1) levels' on 'Parkinson's disease || id:ieu−b−7'

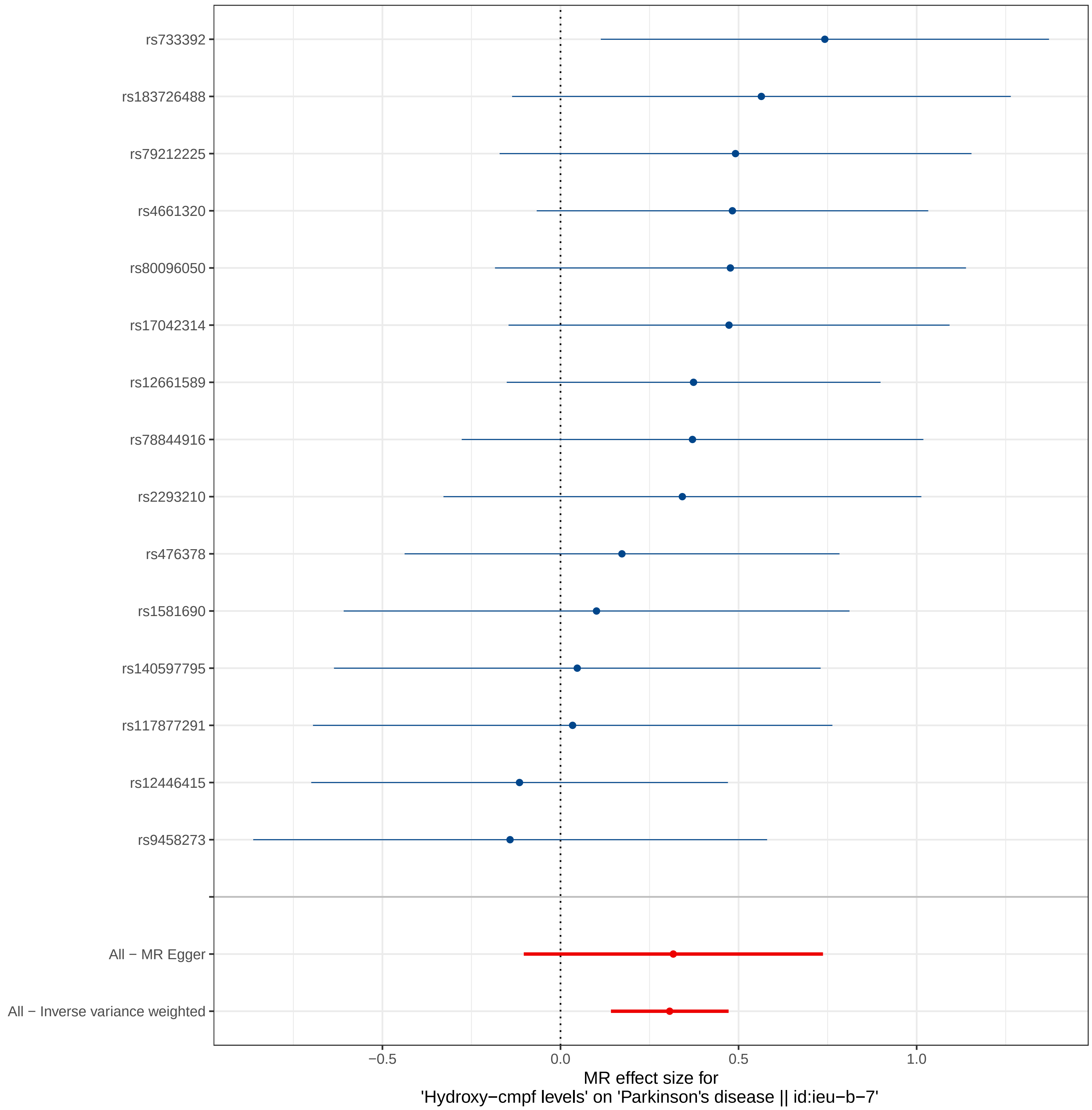

rs733392
rs183726488
rs79212225
rs4661320
rs80096050
rs17042314
rs12661589
rs78844916
rs2293210
rs476378
rs1581690
rs140597795
rs117877291
rs12446415
rs9458273
All − MR Egger
All − Inverse variance weighted
−0.5
0.0
0.5
1.0
MR effect size for
'Hydroxy−cmpf levels' on 'Parkinson's disease || id:ieu−b−7'

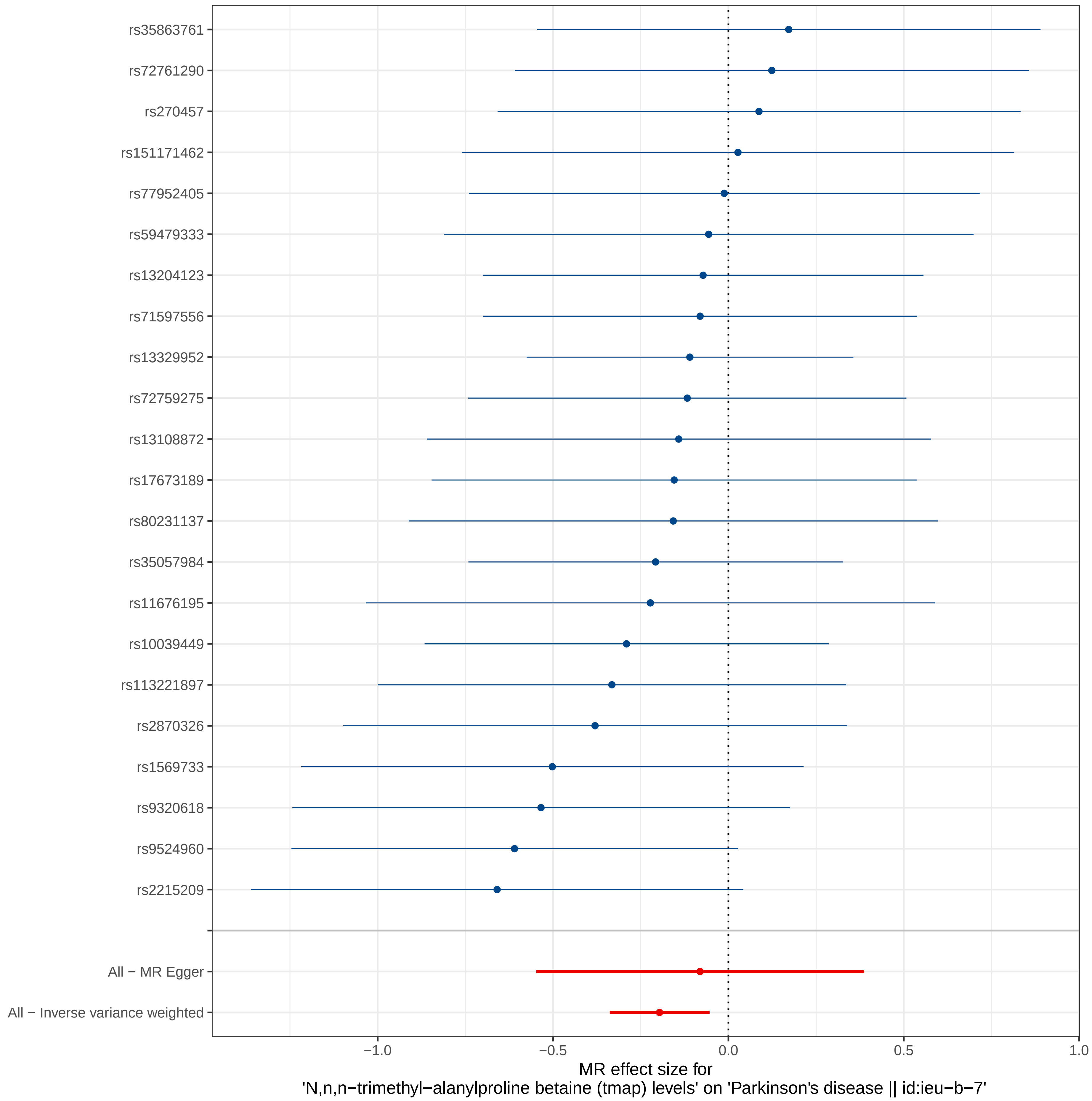
rs35863761
rs72761290
rs270457
rs151171462
rs77952405
rs59479333
rs13204123
rs71597556
rs13329952
rs72759275
rs13108872
rs17673189
rs80231137
rs35057984
rs11676195
rs10039449
rs113221897
rs2870326
rs1569733
rs9320618
rs9524960
rs2215209
All − MR Egger
All − Inverse variance weighted
−1.0
−0.5
0.0
0.5
1.0
MR effect size for
'N,n,n−trimethyl−alanylproline betaine (tmap) levels' on 'Parkinson's disease || id:ieu−b−7'

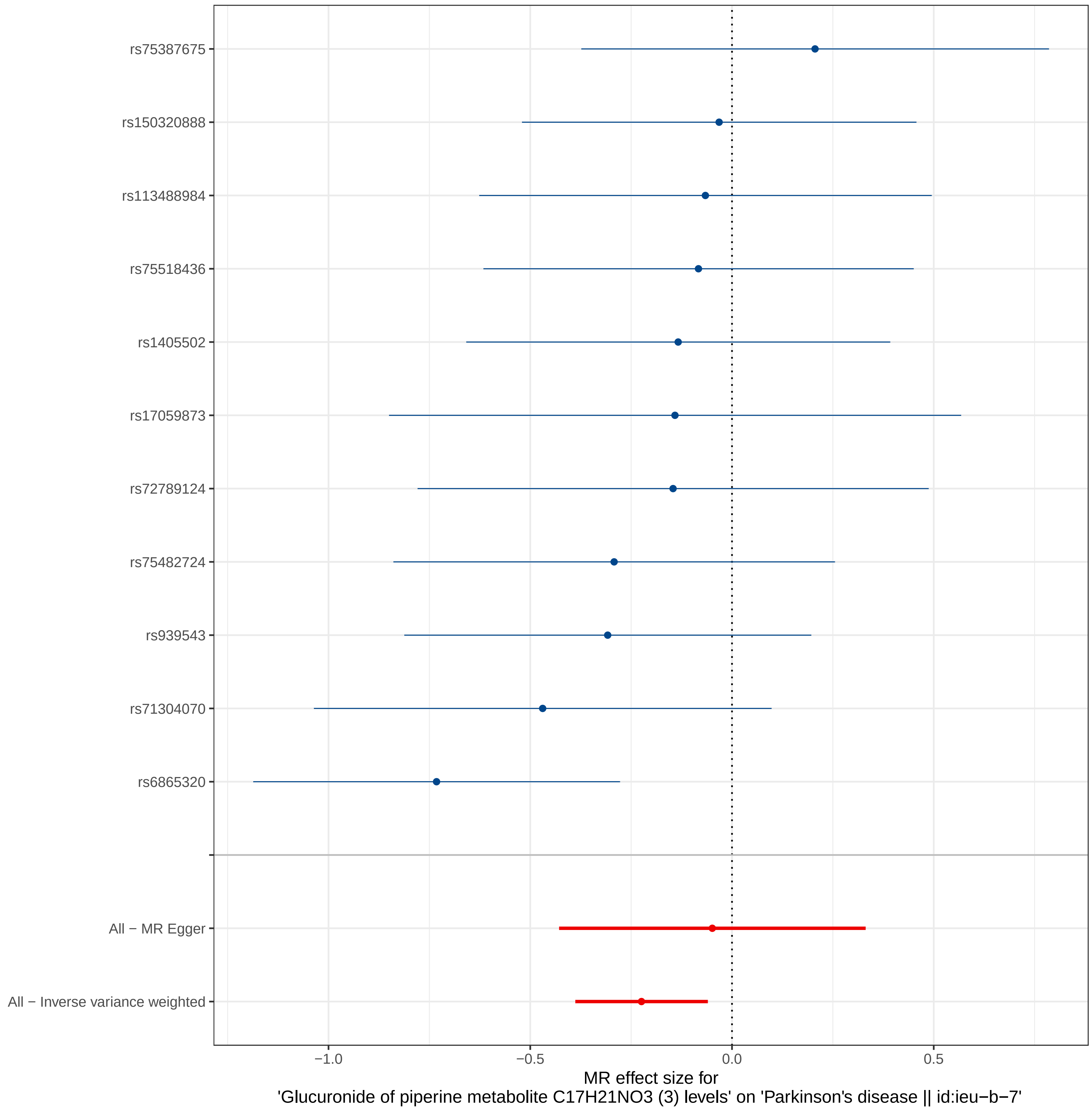
rs75387675
rs150320888
rs113488984
rs75518436
rs1405502
rs17059873
rs72789124
rs75482724
rs939543
rs71304070
rs6865320
All − MR Egger
All − Inverse variance weighted
−1.0
−0.5
0.0
0.5
MR effect size for
'Glucuronide of piperine metabolite C17H21NO3 (3) levels' on 'Parkinson's disease || id:ieu−b−7'

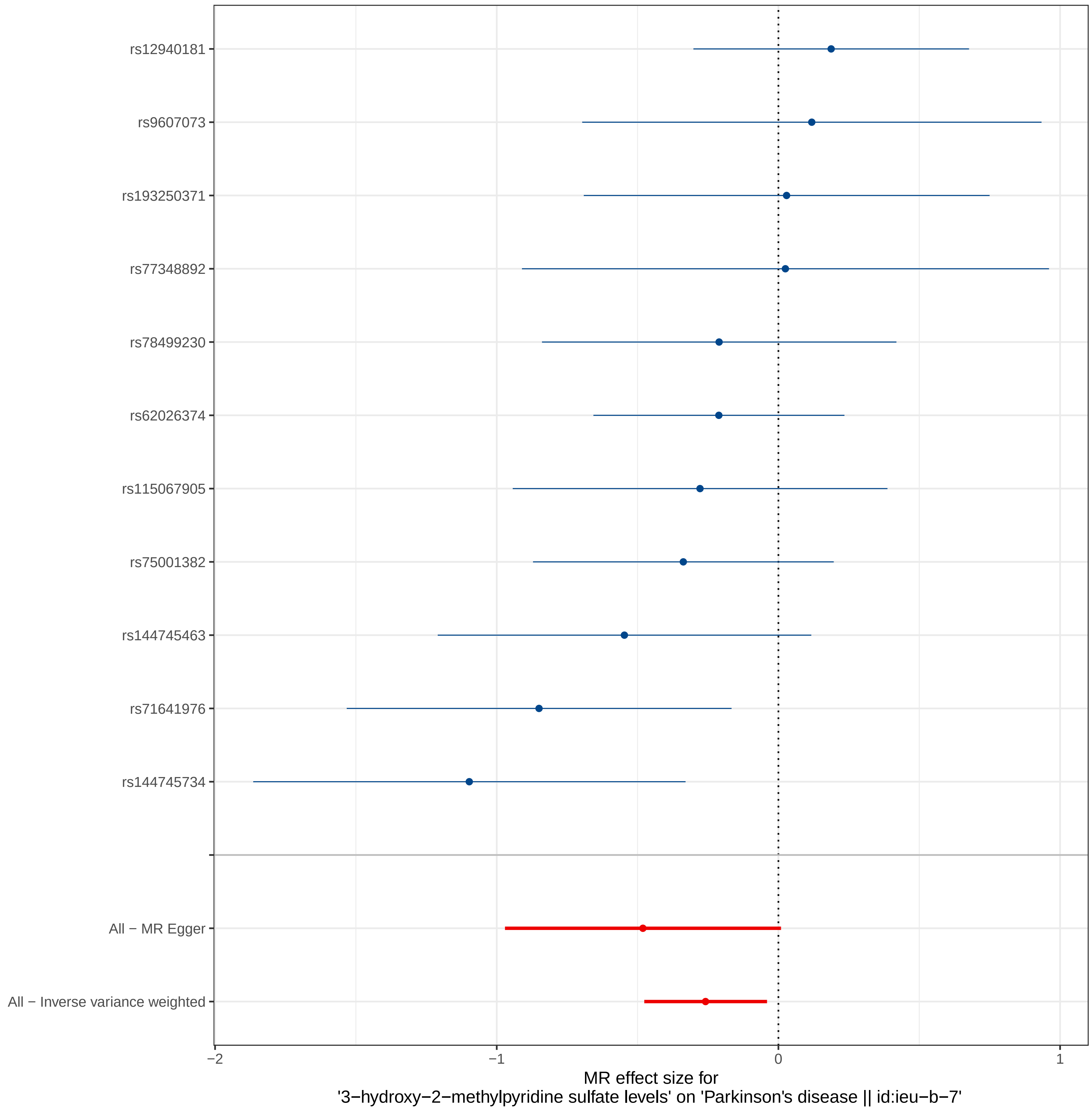
rs12940181
rs9607073
rs193250371
rs77348892
rs78499230
rs62026374
rs115067905
rs75001382
rs144745463
rs71641976
rs144745734
All − MR Egger
All − Inverse variance weighted
−2
−1
0
1
MR effect size for
'3−hydroxy−2−methylpyridine sulfate levels' on 'Parkinson's disease || id:ieu−b−7'

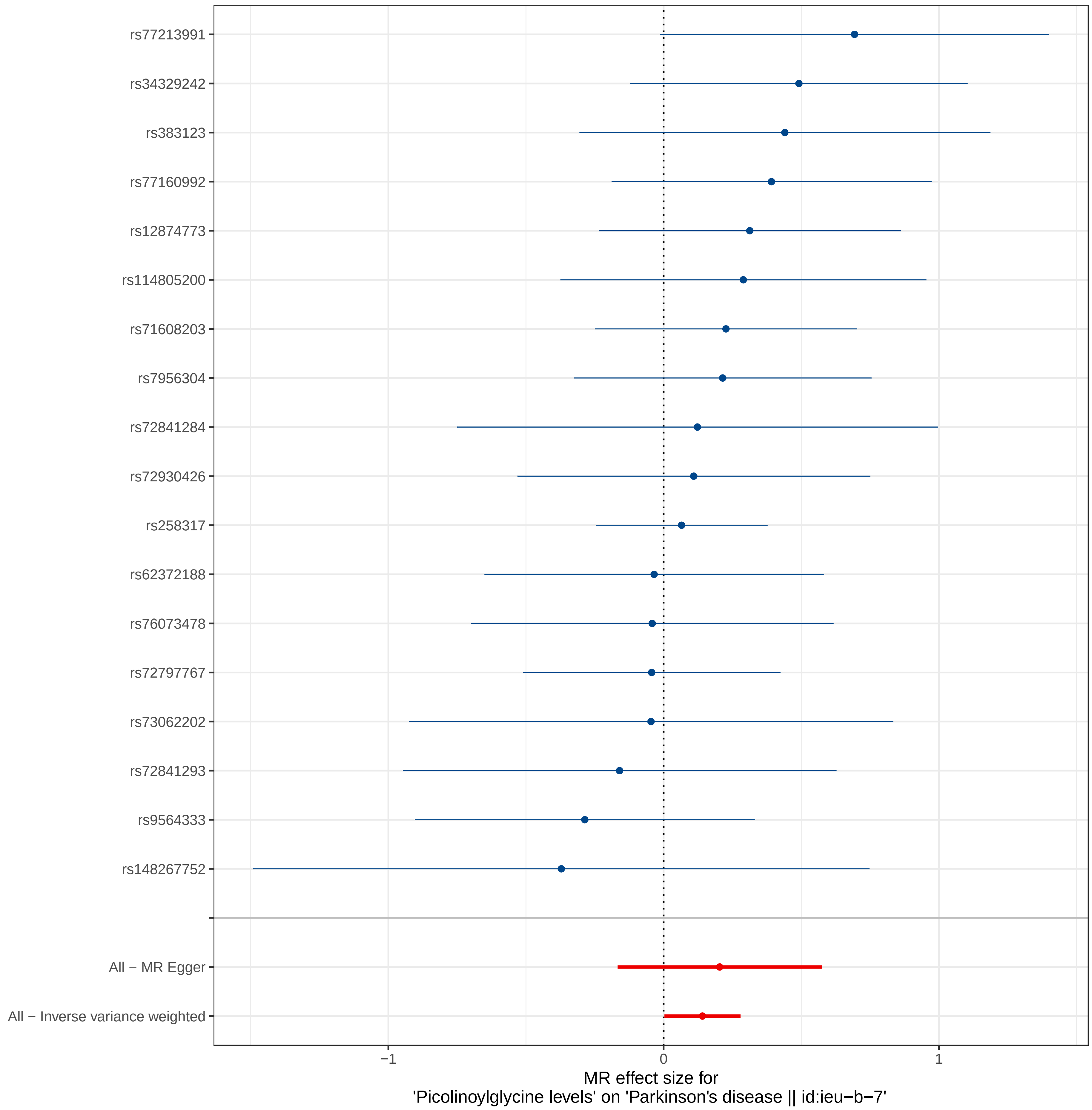

rs77213991
rs34329242
rs383123
rs77160992
rs12874773
rs114805200
rs71608203
rs7956304
rs72841284
rs72930426
rs258317
rs62372188
rs76073478
rs72797767
rs73062202
rs72841293
rs9564333
rs148267752
All − MR Egger
All − Inverse variance weighted
−1
0
1
MR effect size for
'Picolinoylglycine levels' on 'Parkinson's disease || id:ieu−b−7'

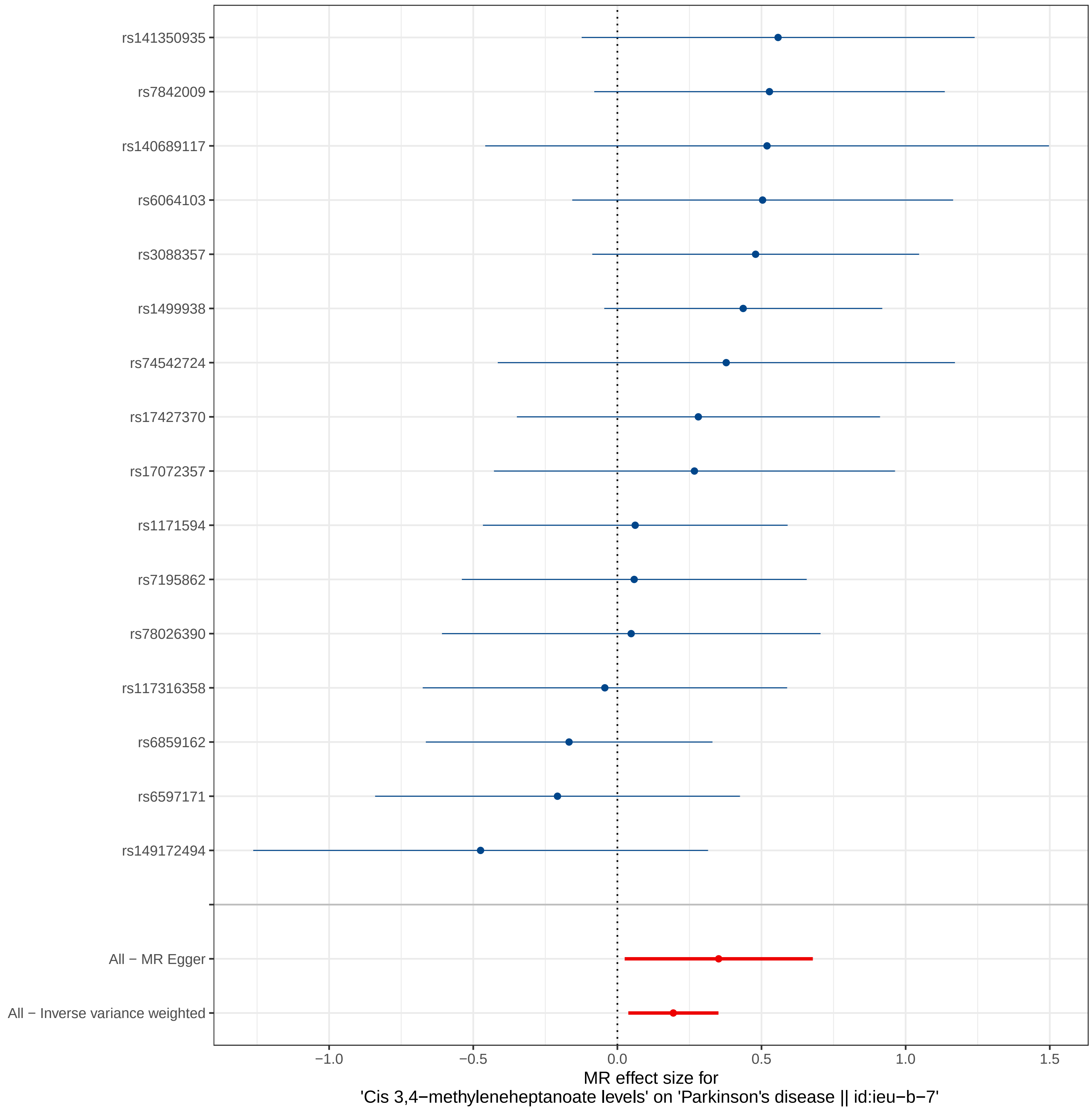
rs141350935
rs7842009
rs140689117
rs6064103
rs3088357
rs1499938
rs74542724
rs17427370
rs17072357
rs1171594
rs7195862
rs78026390
rs117316358
rs6859162
rs6597171
rs149172494
All − MR Egger
All − Inverse variance weighted
−1.0
−0.5
0.0
0.5
1.0
1.5
MR effect size for
'Cis 3,4−methyleneheptanoate levels' on 'Parkinson's disease || id:ieu−b−7'

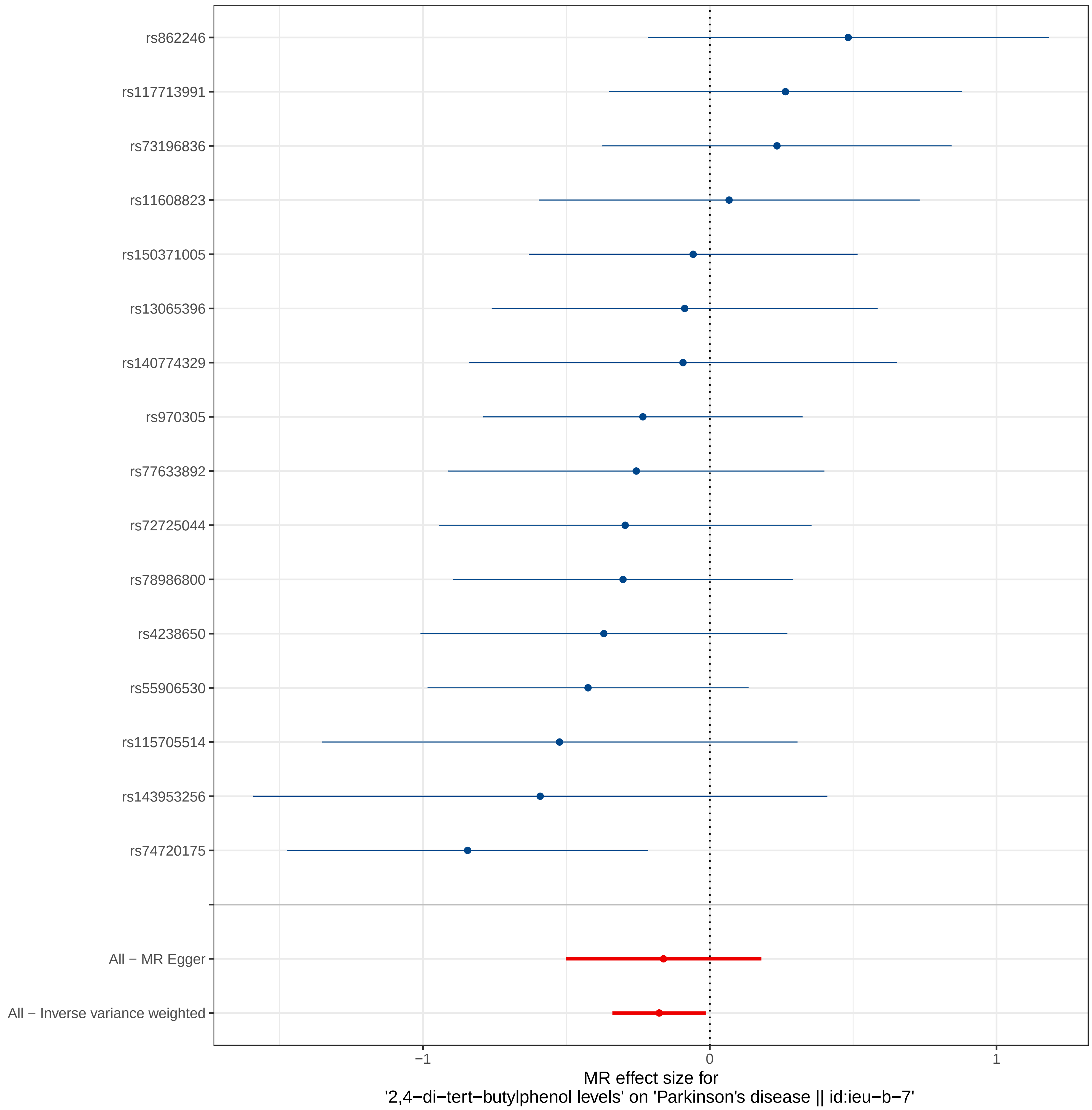

rs862246
rs117713991
rs73196836
rs11608823
rs150371005
rs13065396
rs140774329
rs970305
rs77633892
rs72725044
rs78986800
rs4238650
rs55906530
rs115705514
rs143953256
rs74720175
All − MR Egger
All − Inverse variance weighted
−1
0
1
MR effect size for
'2,4−di−tert−butylphenol levels' on 'Parkinson's disease || id:ieu−b−7'

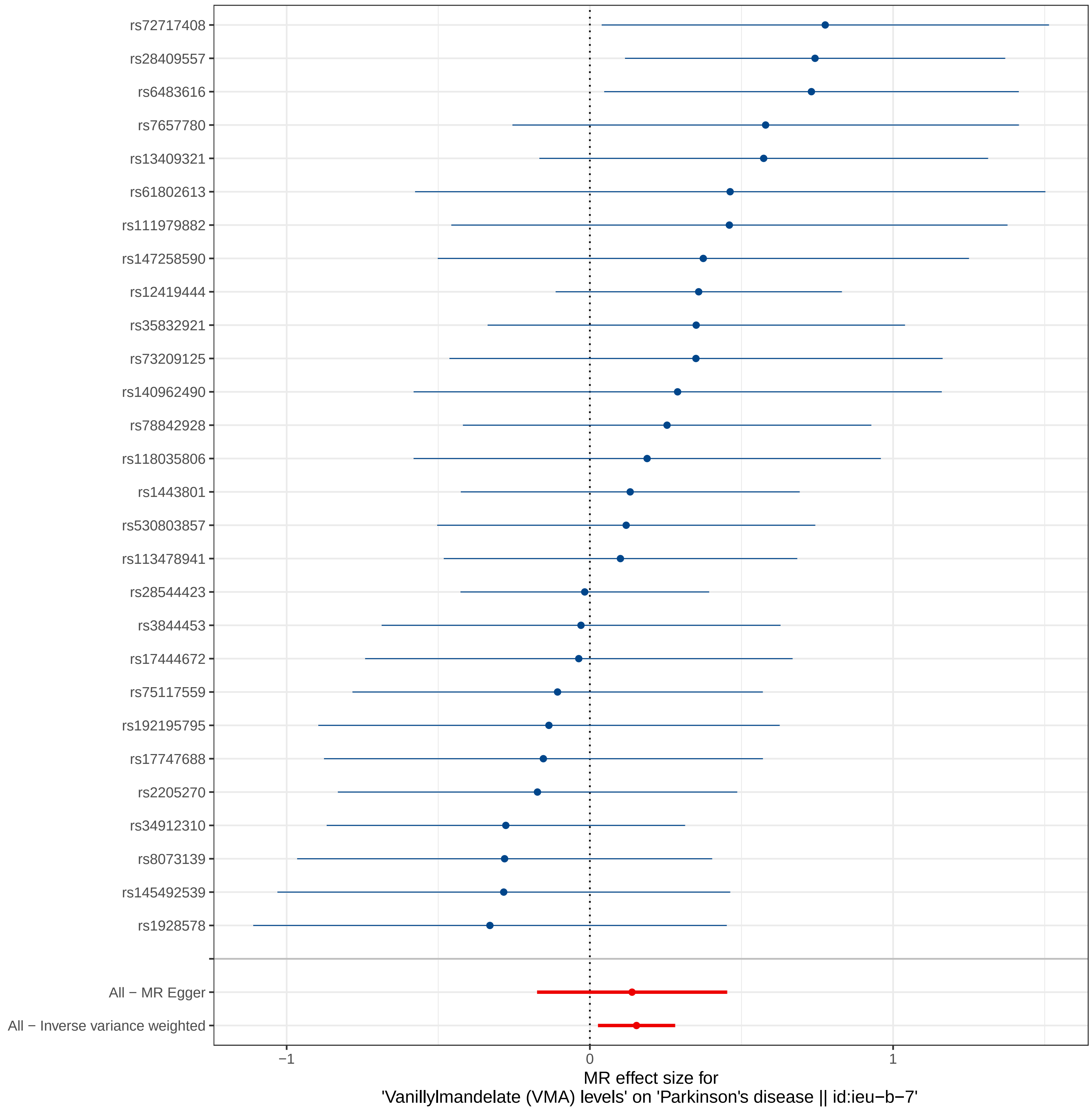
rs72717408
rs28409557
rs6483616
rs7657780
rs13409321
rs61802613
rs111979882
rs147258590
rs12419444
rs35832921
rs73209125
rs140962490
rs78842928
rs118035806
rs1443801
rs530803857
rs113478941
rs28544423
rs3844453
rs17444672
rs75117559
rs192195795
rs17747688
rs2205270
rs34912310
rs8073139
rs145492539
rs1928578
All − MR Egger
All − Inverse variance weighted
−1
0
1
MR effect size for
'Vanillylmandelate (VMA) levels' on 'Parkinson's disease || id:ieu-b-7'

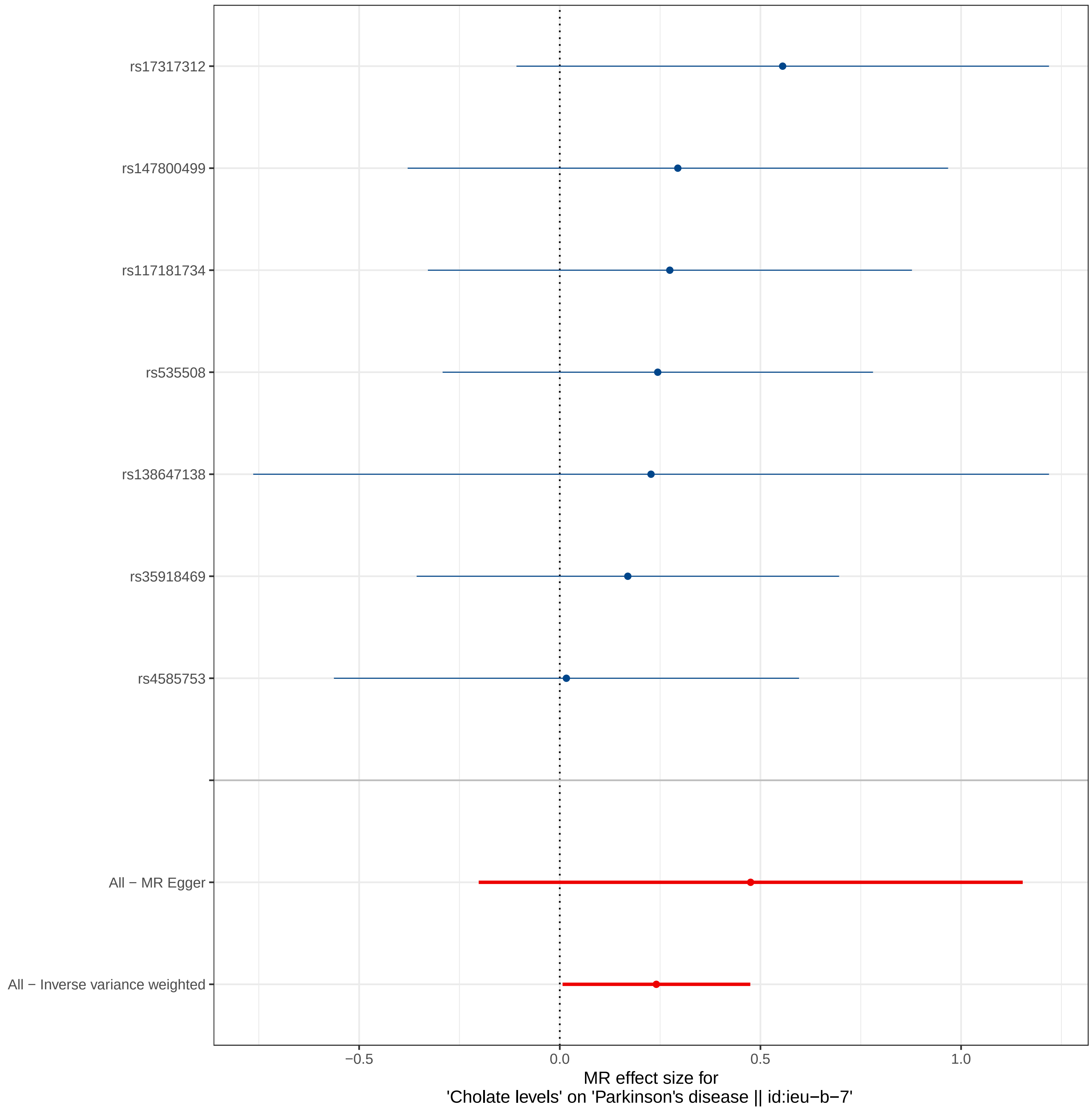
rs17317312
rs147800499
rs117181734
rs535508
rs138647138
rs35918469
rs4585753
All − MR Egger
All − Inverse variance weighted
−0.5
0.0
0.5
1.0
MR effect size for
'Cholate levels' on 'Parkinson's disease || id:ieu−b−7'

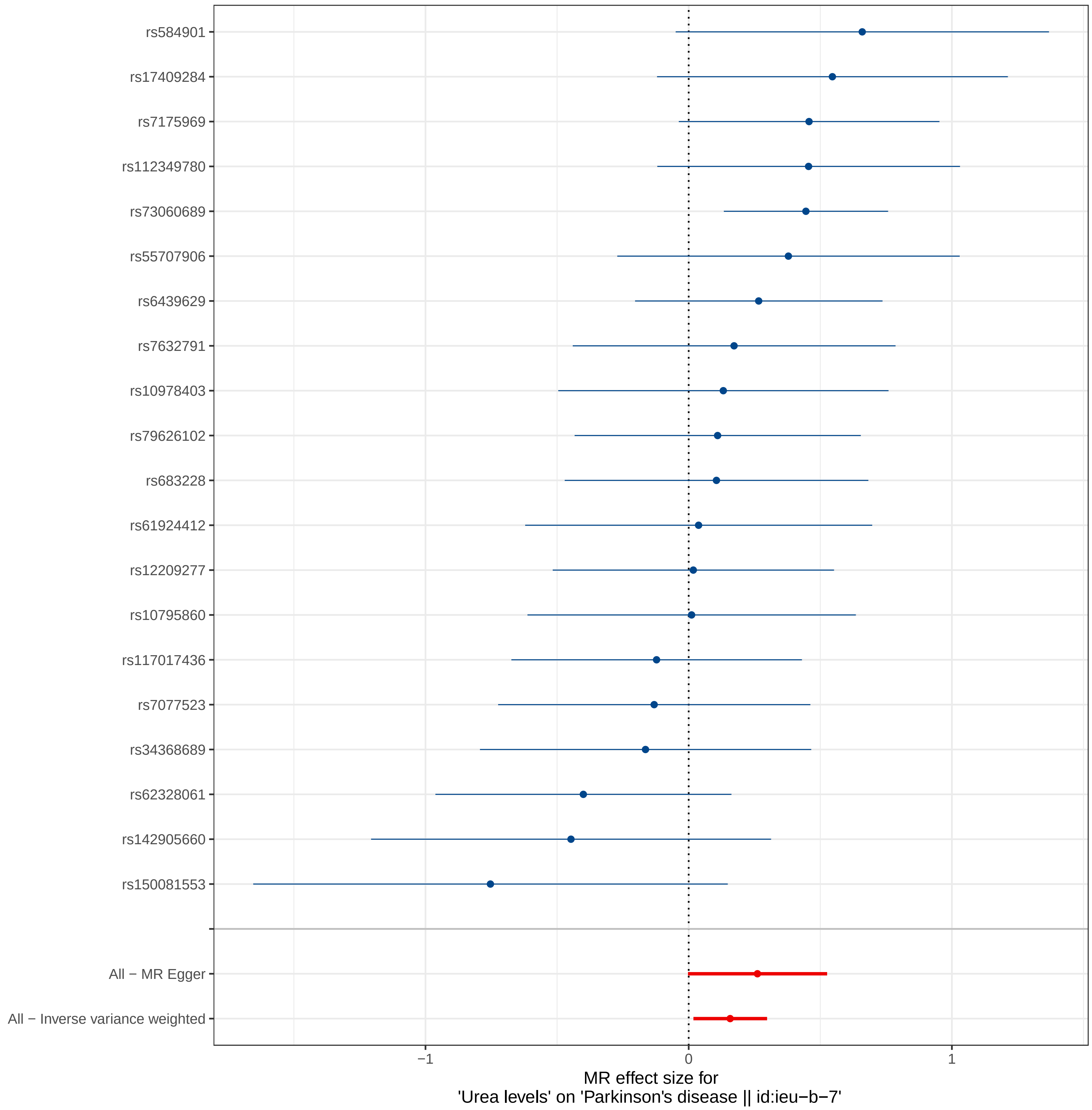
rs584901
rs17409284
rs7175969
rs112349780
rs73060689
rs55707906
rs6439629
rs7632791
rs10978403
rs79626102
rs683228
rs61924412
rs12209277
rs10795860
rs117017436
rs7077523
rs34368689
rs62328061
rs142905660
rs150081553
All − MR Egger
All − Inverse variance weighted
−1
0
1
MR effect size for
'Urea levels' on 'Parkinson's disease || id:ieu−b−7'

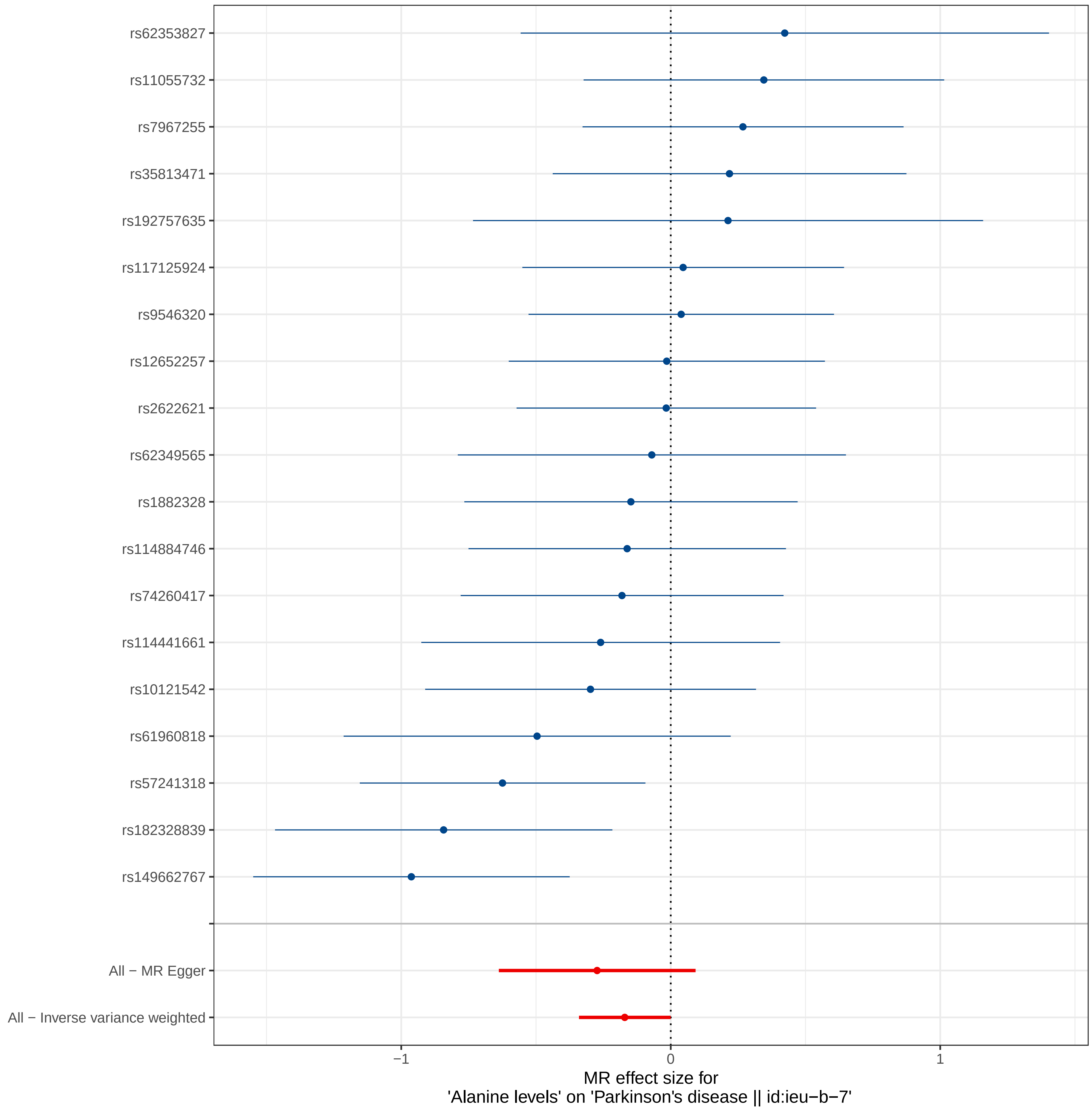

rs62353827
rs11055732
rs7967255
rs35813471
rs192757635
rs117125924
rs9546320
rs12652257
rs2622621
rs62349565
rs1882328
rs114884746
rs74260417
rs114441661
rs10121542
rs61960818
rs57241318
rs182328839
rs149662767
All − MR Egger
All − Inverse variance weighted
−1
0
1
MR effect size for
'Alanine levels' on 'Parkinson's disease || id:ieu−b−7'

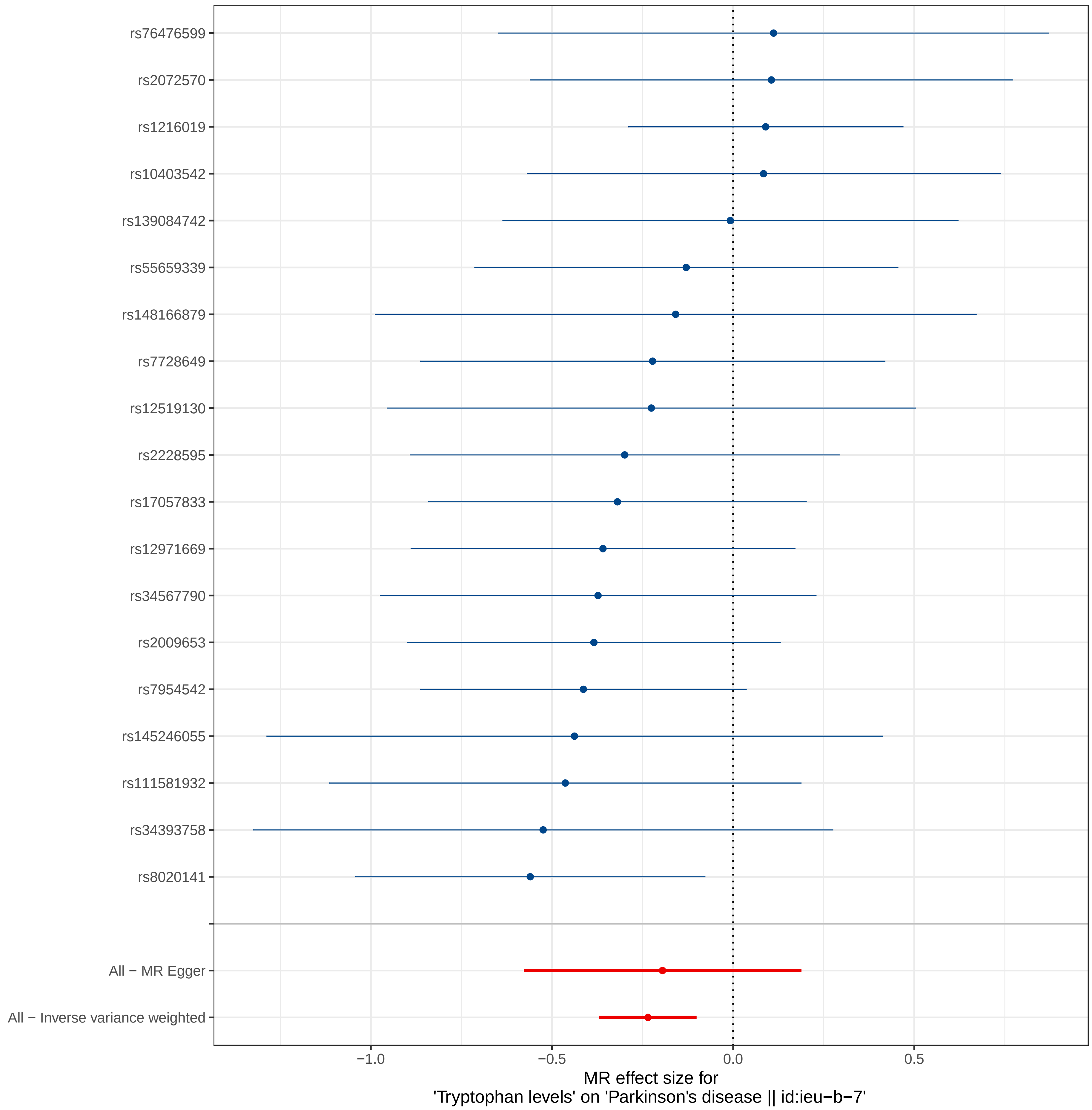
rs76476599
rs2072570
rs1216019
rs10403542
rs139084742
rs55659339
rs148166879
rs7728649
rs12519130
rs2228595
rs17057833
rs12971669
rs34567790
rs2009653
rs7954542
rs145246055
rs111581932
rs34393758
rs8020141
All − MR Egger
All − Inverse variance weighted
−1.0
−0.5
0.0
0.5
MR effect size for
'Tryptophan levels' on 'Parkinson's disease || id:ieu−b−7'

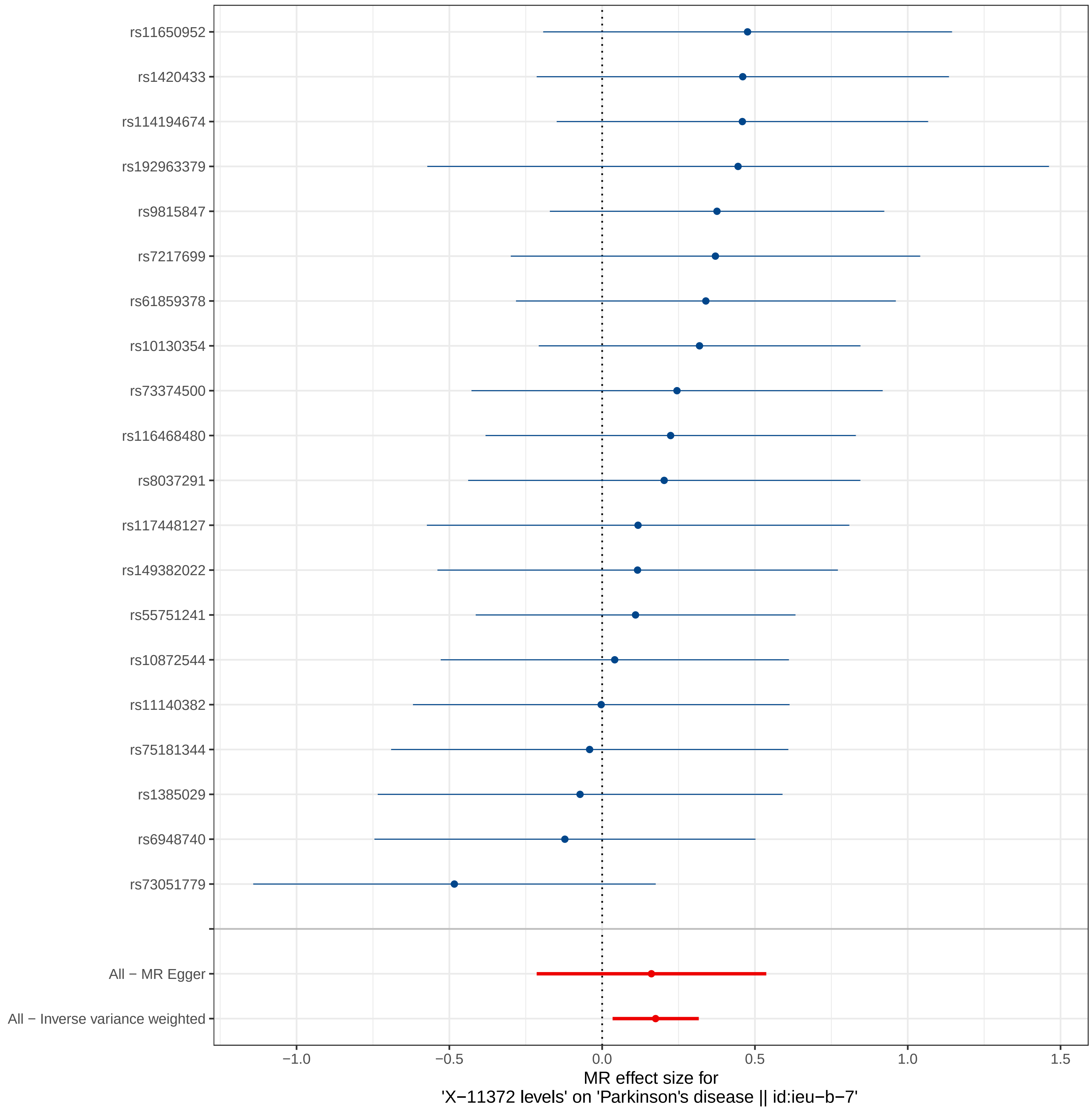
rs11650952
rs1420433
rs114194674
rs192963379
rs9815847
rs7217699
rs61859378
rs10130354
rs73374500
rs116468480
rs8037291
rs117448127
rs149382022
rs55751241
rs10872544
rs11140382
rs75181344
rs1385029
rs6948740
rs73051779
All − MR Egger
All − Inverse variance weighted
−1.0
−0.5
0.0
0.5
1.0
1.5
MR effect size for
'X−11372 levels' on 'Parkinson's disease || id:ieu−b−7'

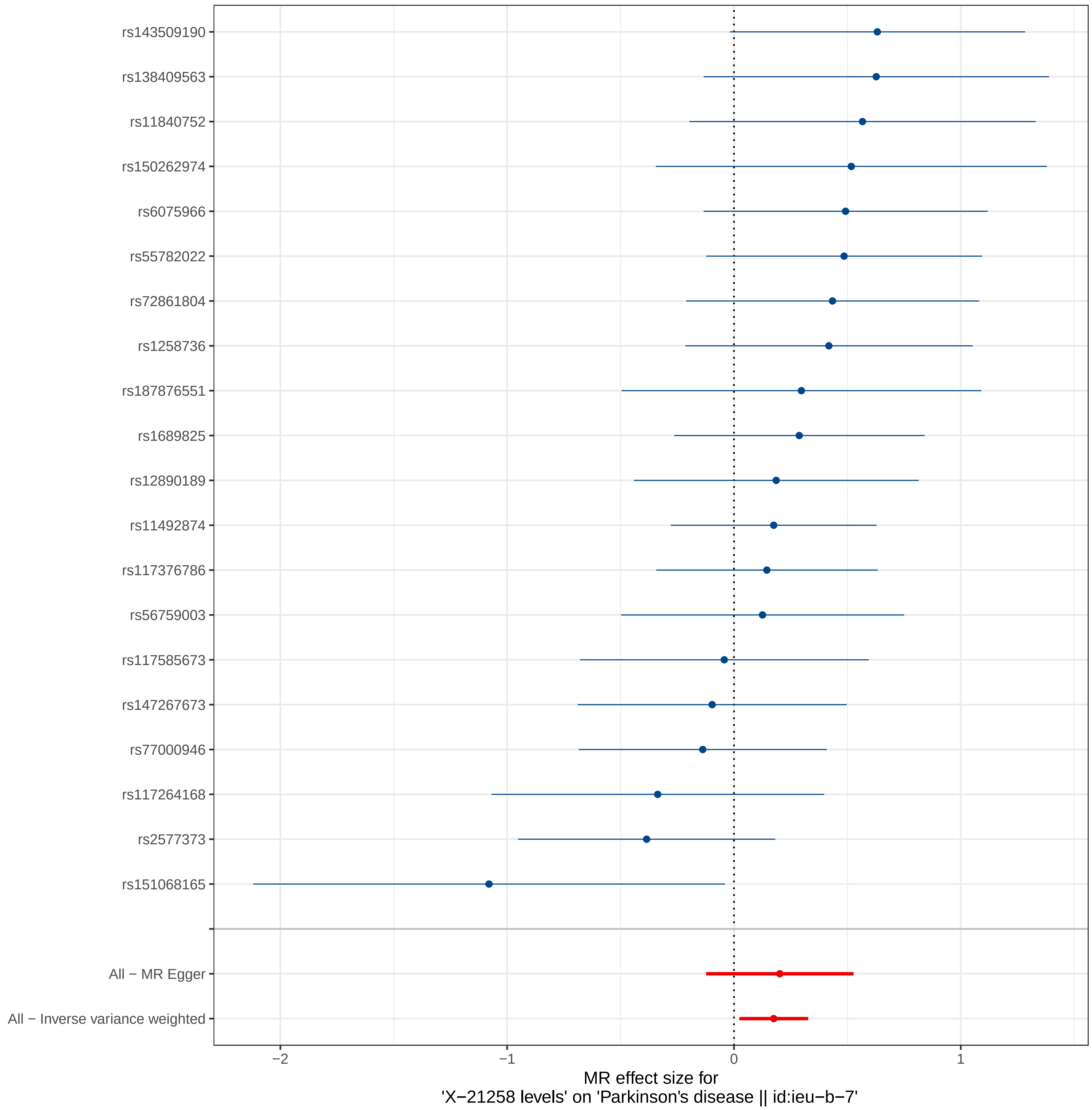
rs143509190
rs138409563
rs11840752
rs150262974
rs6075966
rs55782022
rs72861804
rs1258736
rs187876551
rs1689825
rs12890189
rs11492874
rs117376786
rs56759003
rs117585673
rs147267673
rs77000946
rs117264168
rs2577373
rs151068165
All − MR Egger
All − Inverse variance weighted
−2
−1
0
1
MR effect size for
'X−21258 levels' on 'Parkinson's disease || id:ieu−b−7'

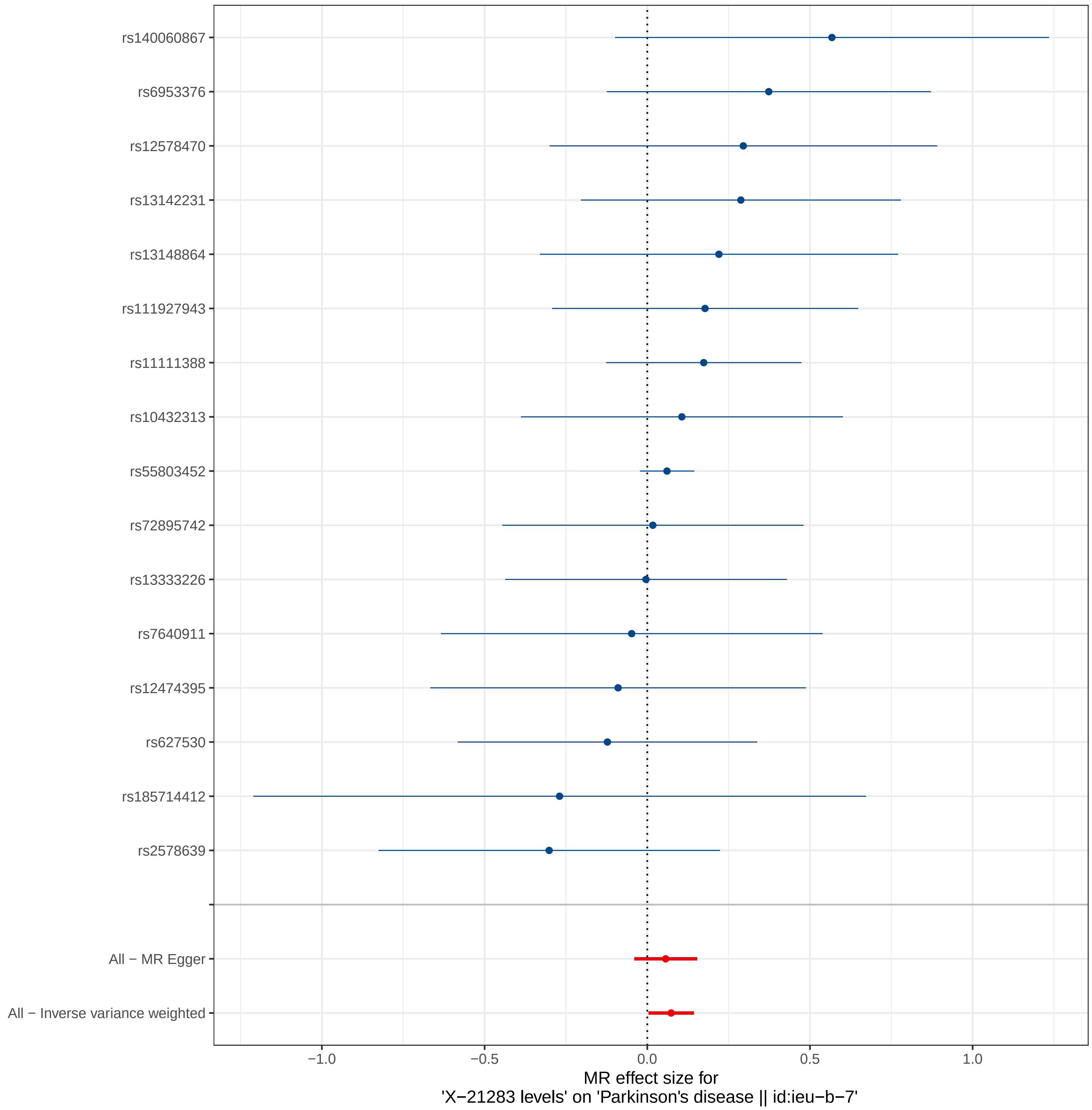
rs140060867
rs6953376
rs12578470
rs13142231
rs13148864
rs111927943
rs11111388
rs10432313
rs55803452
rs72895742
rs13333226
rs7640911
rs12474395
rs627530
rs185714412
rs2578639
All − MR Egger
All − Inverse variance weighted
−1.0
−0.5
0.0
0.5
1.0
MR effect size for
'X−21283 levels' on 'Parkinson's disease || id:ieu−b−7'

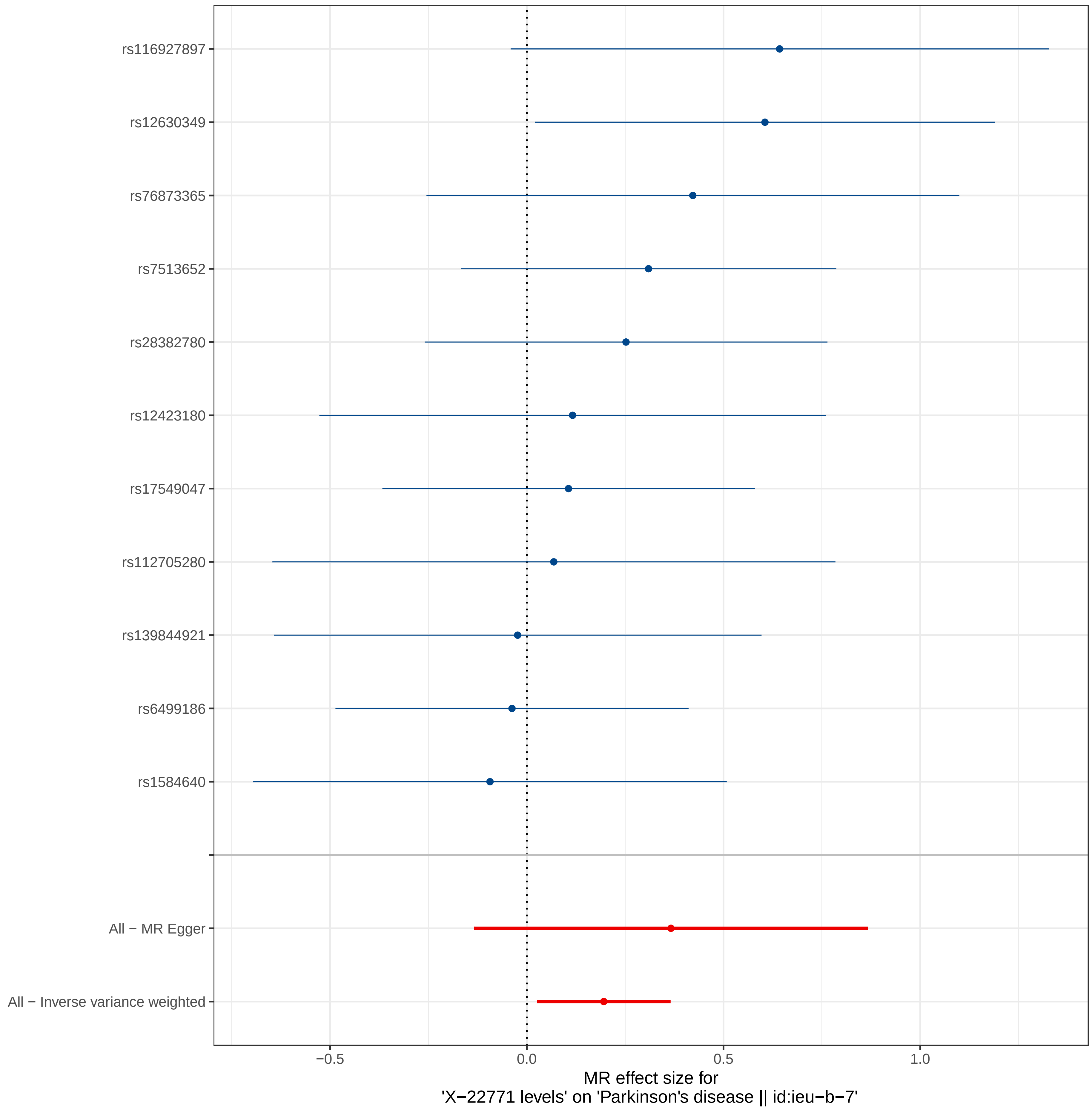
rs116927897
rs12630349
rs76873365
rs7513652
rs28382780
rs12423180
rs17549047
rs112705280
rs139844921
rs6499186
rs1584640
All − MR Egger
All − Inverse variance weighted
−0.5
0.0
0.5
1.0
MR effect size for
'X−22771 levels' on 'Parkinson's disease || id:ieu−b−7'

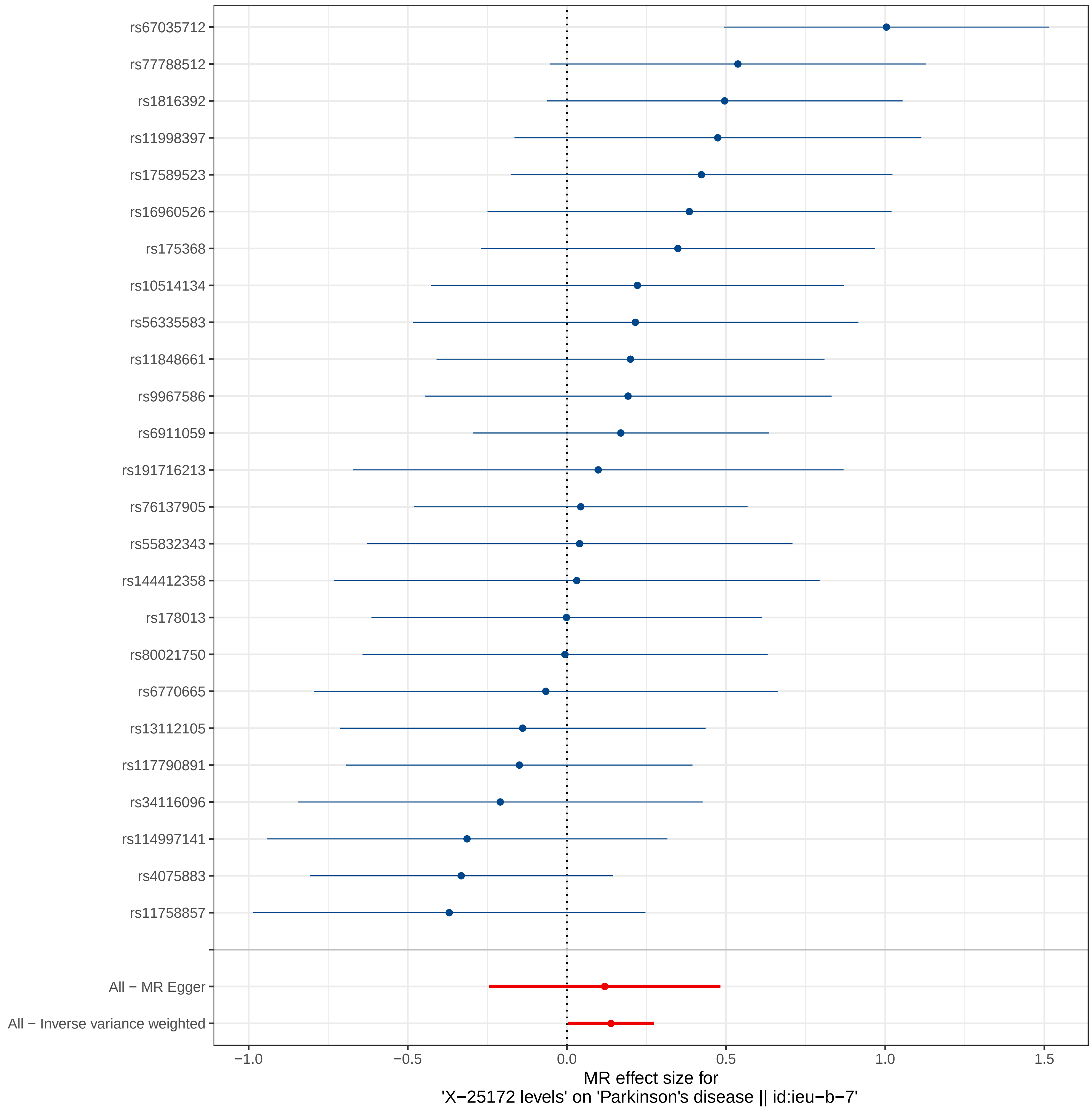
rs67035712
rs77788512
rs1816392
rs11998397
rs17589523
rs16960526
rs175368
rs10514134
rs56335583
rs11848661
rs9967586
rs6911059
rs191716213
rs76137905
rs55832343
rs144412358
rs178013
rs80021750
rs6770665
rs13112105
rs117790891
rs34116096
rs114997141
rs4075883
rs11758857
All − MR Egger
All − Inverse variance weighted
−1.0
−0.5
0.0
0.5
1.0
1.5
MR effect size for
'X−25172 levels' on 'Parkinson's disease || id:ieu−b−7'

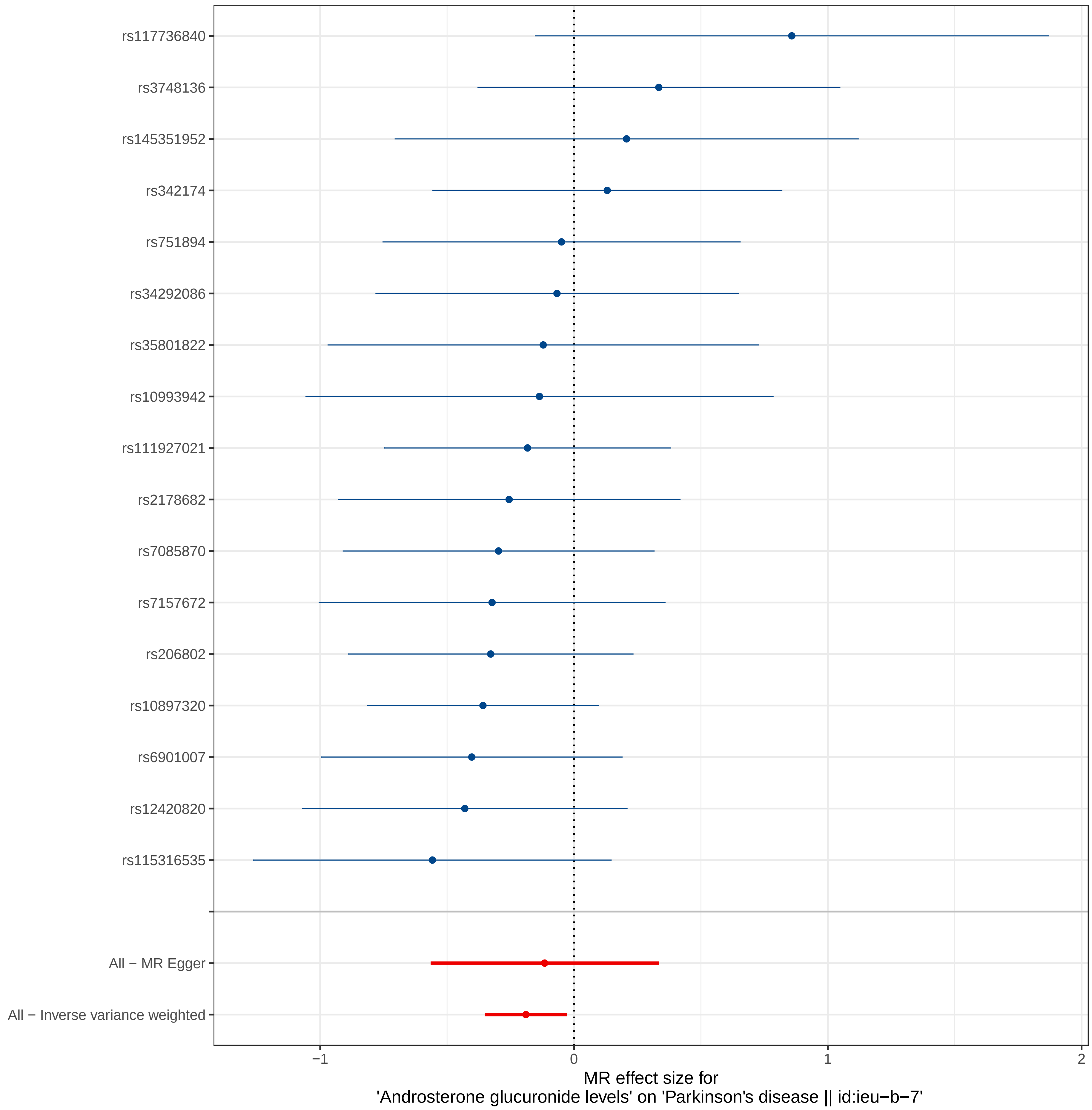

rs117736840
rs3748136
rs145351952
rs342174
rs751894
rs34292086
rs35801822
rs10993942
rs111927021
rs2178682
rs7085870
rs7157672
rs206802
rs10897320
rs6901007
rs12420820
rs115316535
All − MR Egger
All − Inverse variance weighted
−1
0
1
2
MR effect size for
'Androsterone glucuronide levels' on 'Parkinson's disease || id:ieu−b−7'

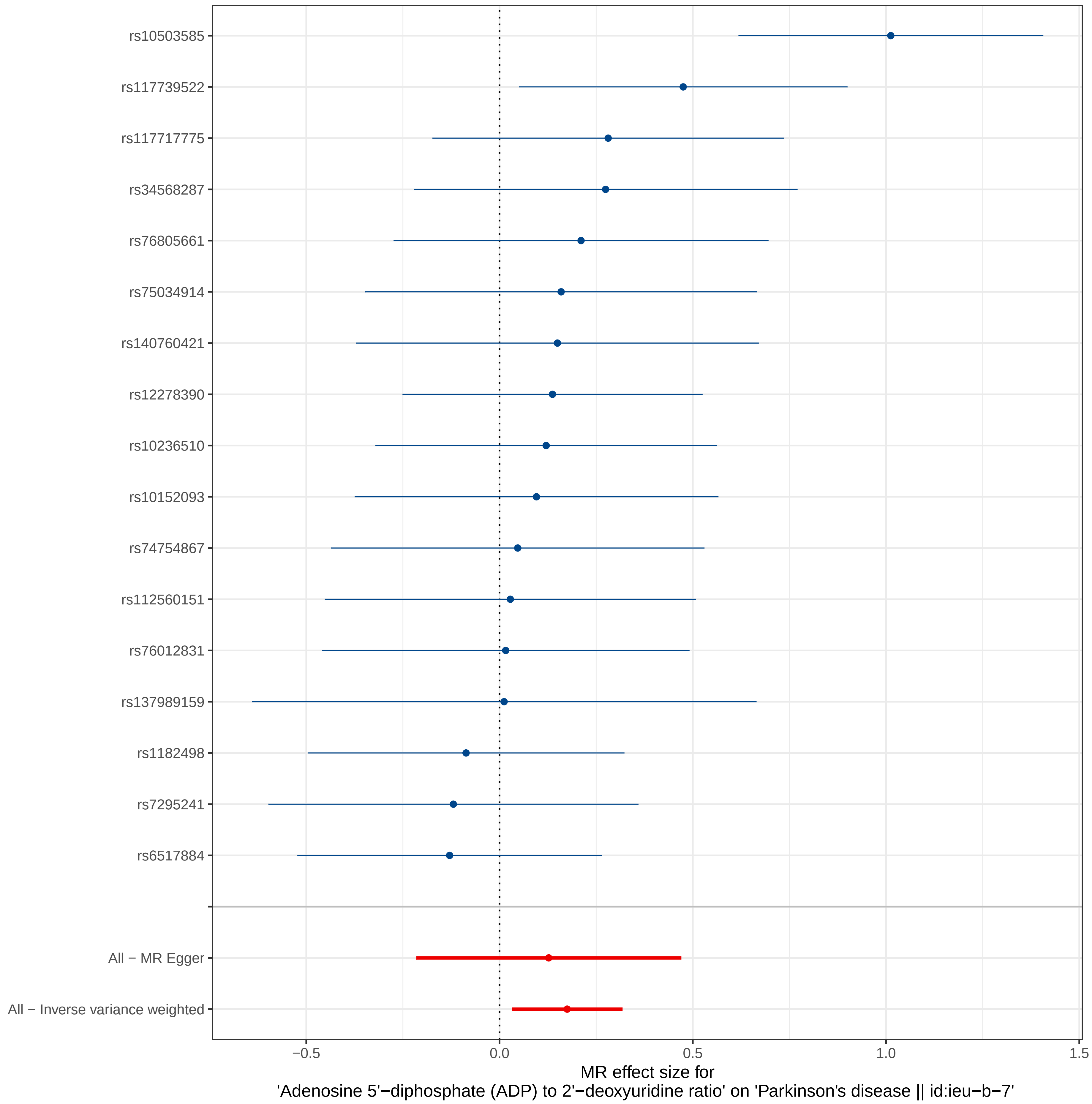

rs10503585
rs117739522
rs117717775
rs34568287
rs76805661
rs75034914
rs140760421
rs12278390
rs10236510
rs10152093
rs74754867
rs112560151
rs76012831
rs137989159
rs1182498
rs7295241
rs6517884
All − MR Egger
All − Inverse variance weighted
−0.5
0.0
0.5
1.0
1.5
MR effect size for
'Adenosine 5'−diphosphate (ADP) to 2'−deoxyuridine ratio' on 'Parkinson's disease || id:ieu−b−7'

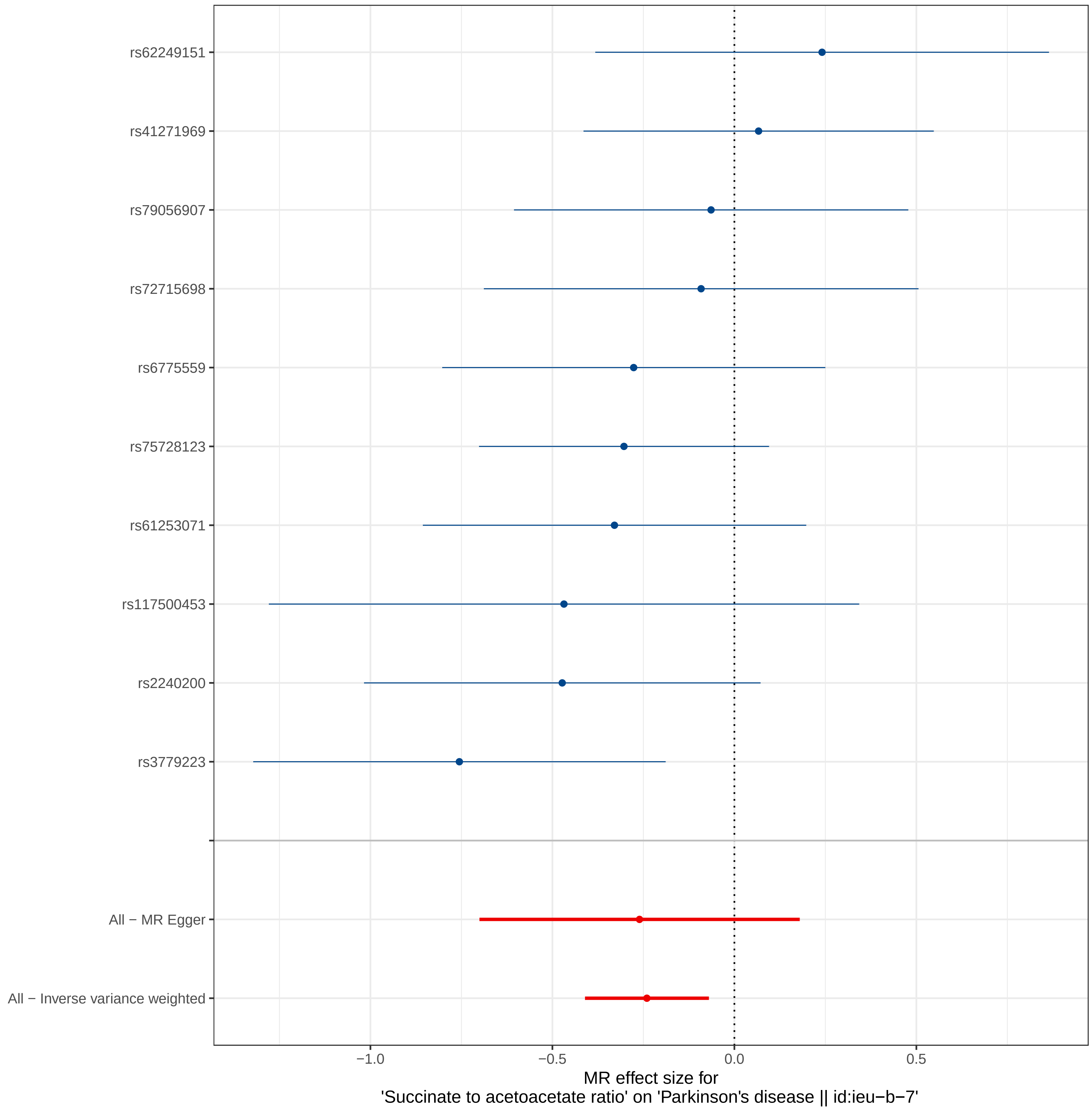

rs62249151
rs41271969
rs79056907
rs72715698
rs6775559
rs75728123
rs61253071
rs117500453
rs2240200
rs3779223
All − MR Egger
All − Inverse variance weighted
−1.0
−0.5
0.0
0.5
MR effect size for
'Succinate to acetoacetate ratio' on 'Parkinson's disease || id:ieu−b−7'

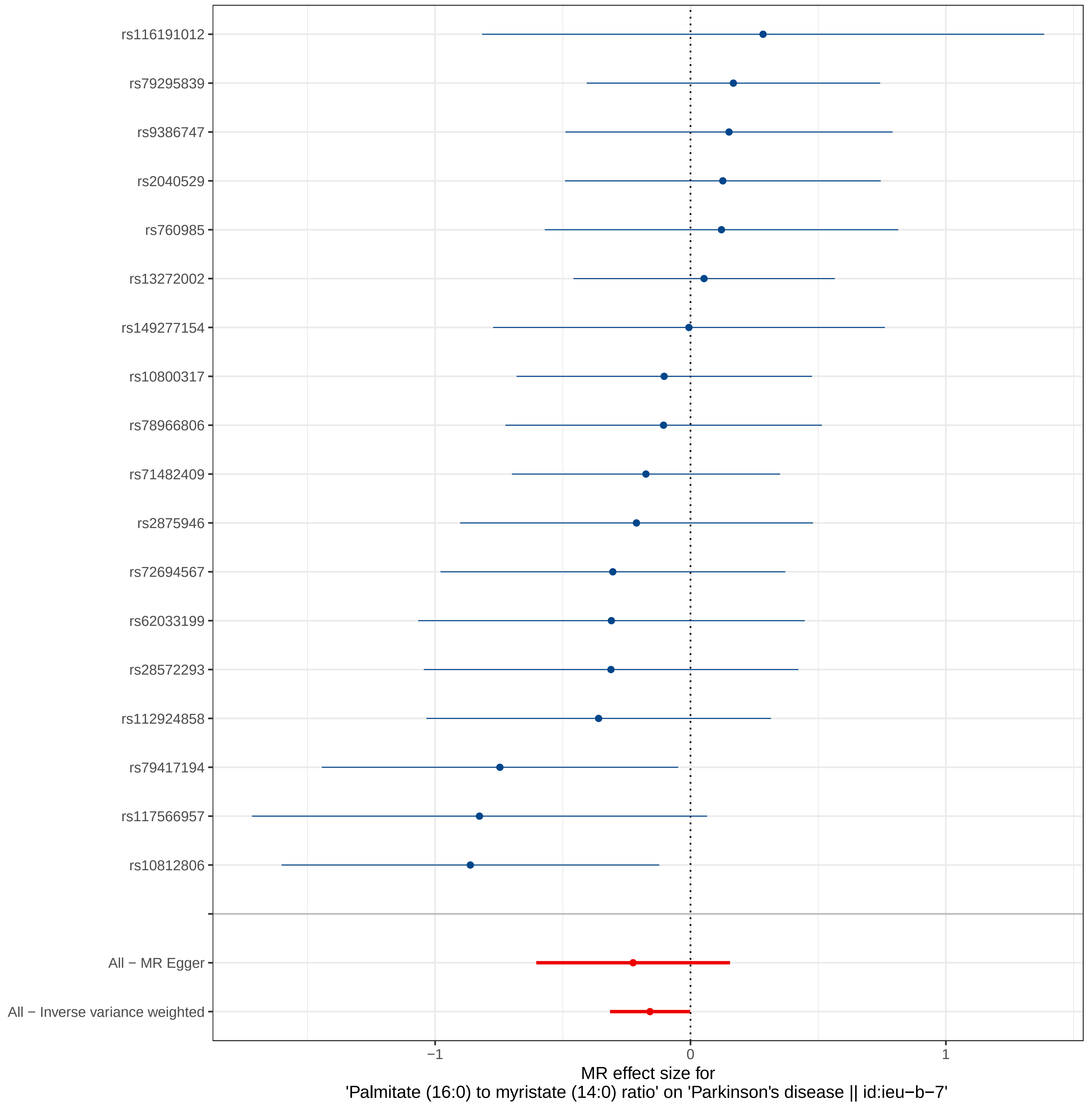

rs116191012
rs79295839
rs9386747
rs2040529
rs760985
rs13272002
rs149277154
rs10800317
rs78966806
rs71482409
rs2875946
rs72694567
rs62033199
rs28572293
rs112924858
rs79417194
rs117566957
rs10812806
All − MR Egger
All − Inverse variance weighted
−1
0
1
MR effect size for
'Palmitate (16:0) to myristate (14:0) ratio' on 'Parkinson's disease || id:ieu−b−7'

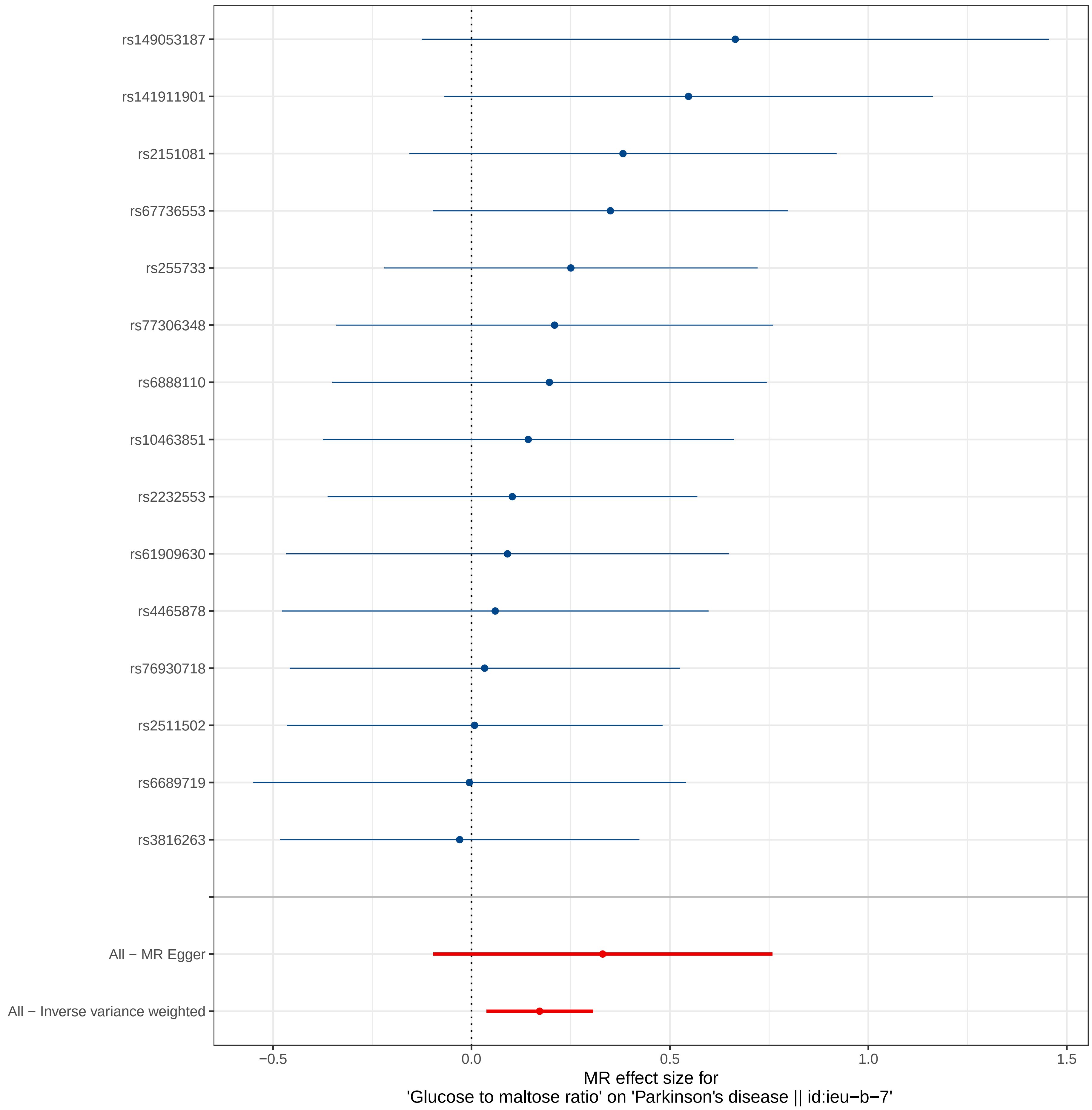

rs149053187
rs141911901
rs2151081
rs67736553
rs255733
rs77306348
rs6888110
rs10463851
rs2232553
rs61909630
rs4465878
rs76930718
rs2511502
rs6689719
rs3816263
All − MR Egger
All − Inverse variance weighted
−0.5
0.0
0.5
1.0
1.5
MR effect size for
'Glucose to maltose ratio' on 'Parkinson's disease || id:ieu−b−7'

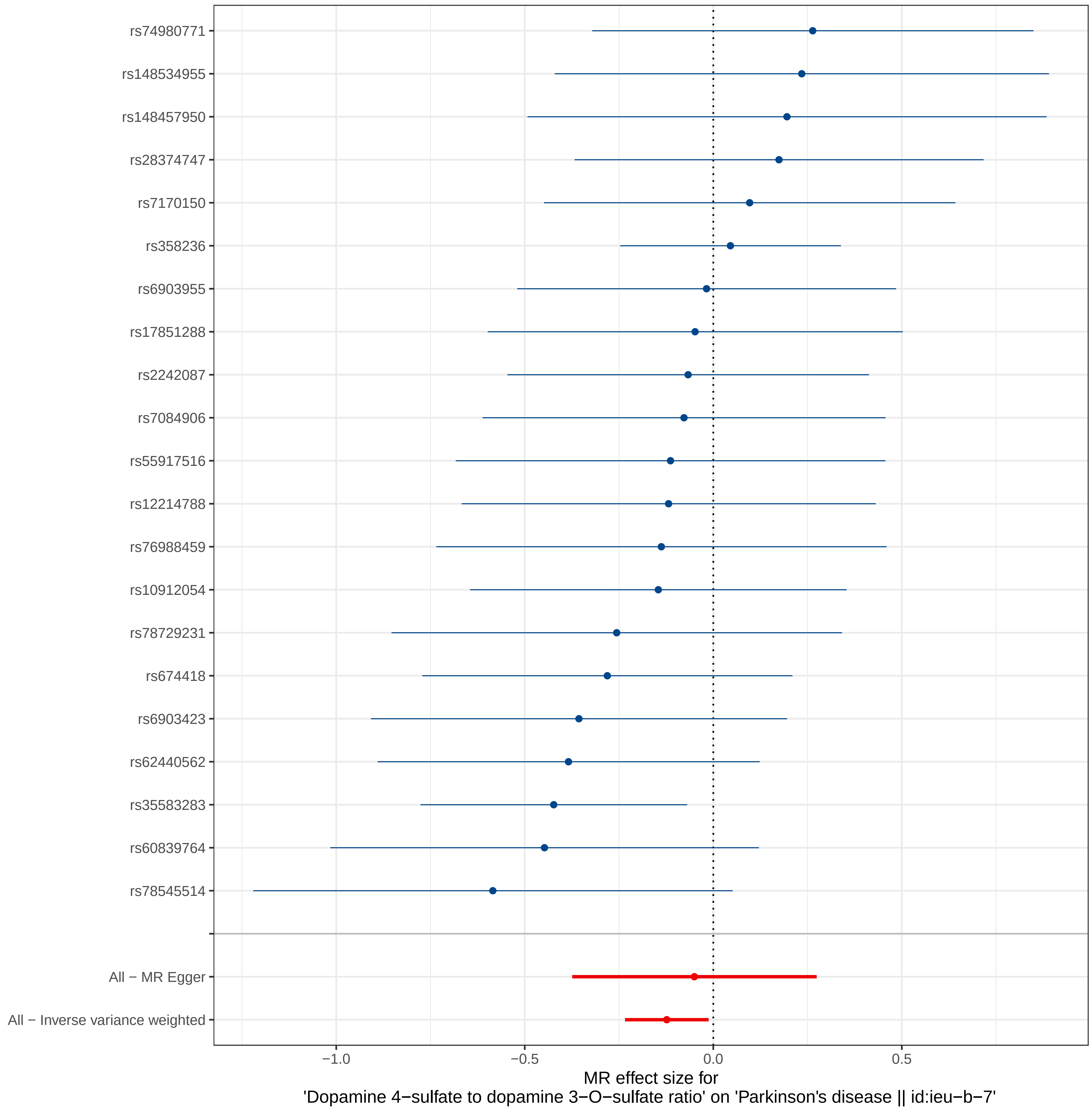
rs74980771
rs148534955
rs148457950
rs28374747
rs7170150
rs358236
rs6903955
rs17851288
rs2242087
rs7084906
rs55917516
rs12214788
rs76988459
rs10912054
rs78729231
rs674418
rs6903423
rs62440562
rs35583283
rs60839764
rs78545514
All − MR Egger
All − Inverse variance weighted
−1.0
−0.5
0.0
0.5
MR effect size for
'Dopamine 4−sulfate to dopamine 3−O−sulfate ratio' on 'Parkinson's disease || id:ieu−b−7'

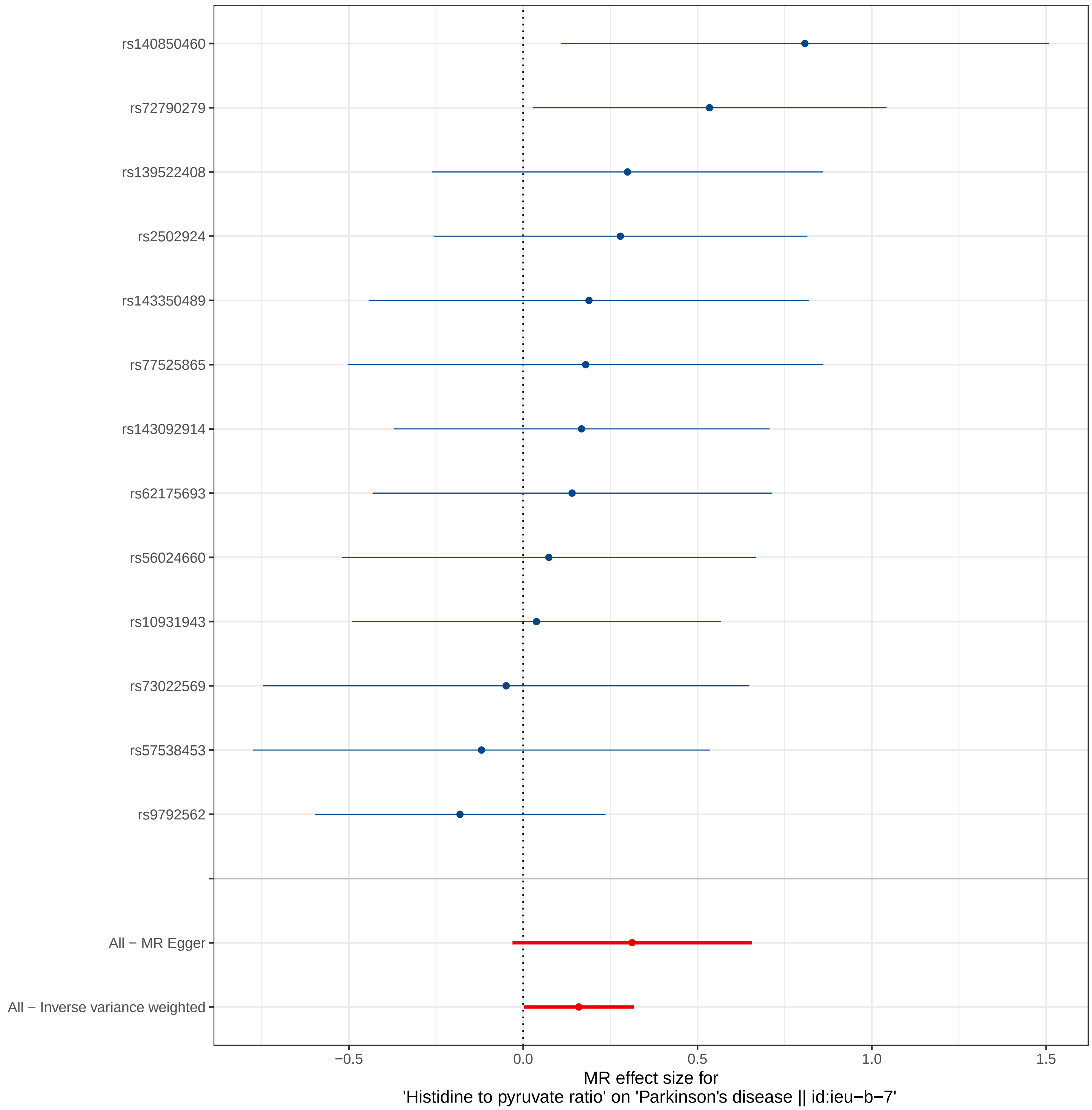

rs140850460
rs72790279
rs139522408
rs2502924
rs143350489
rs77525865
rs143092914
rs62175693
rs56024660
rs10931943
rs73022569
rs57538453
rs9792562
All − MR Egger
All − Inverse variance weighted
−0.5
0.0
0.5
1.0
1.5
MR effect size for
'Histidine to pyruvate ratio' on 'Parkinson's disease || id:ieu−b−7'

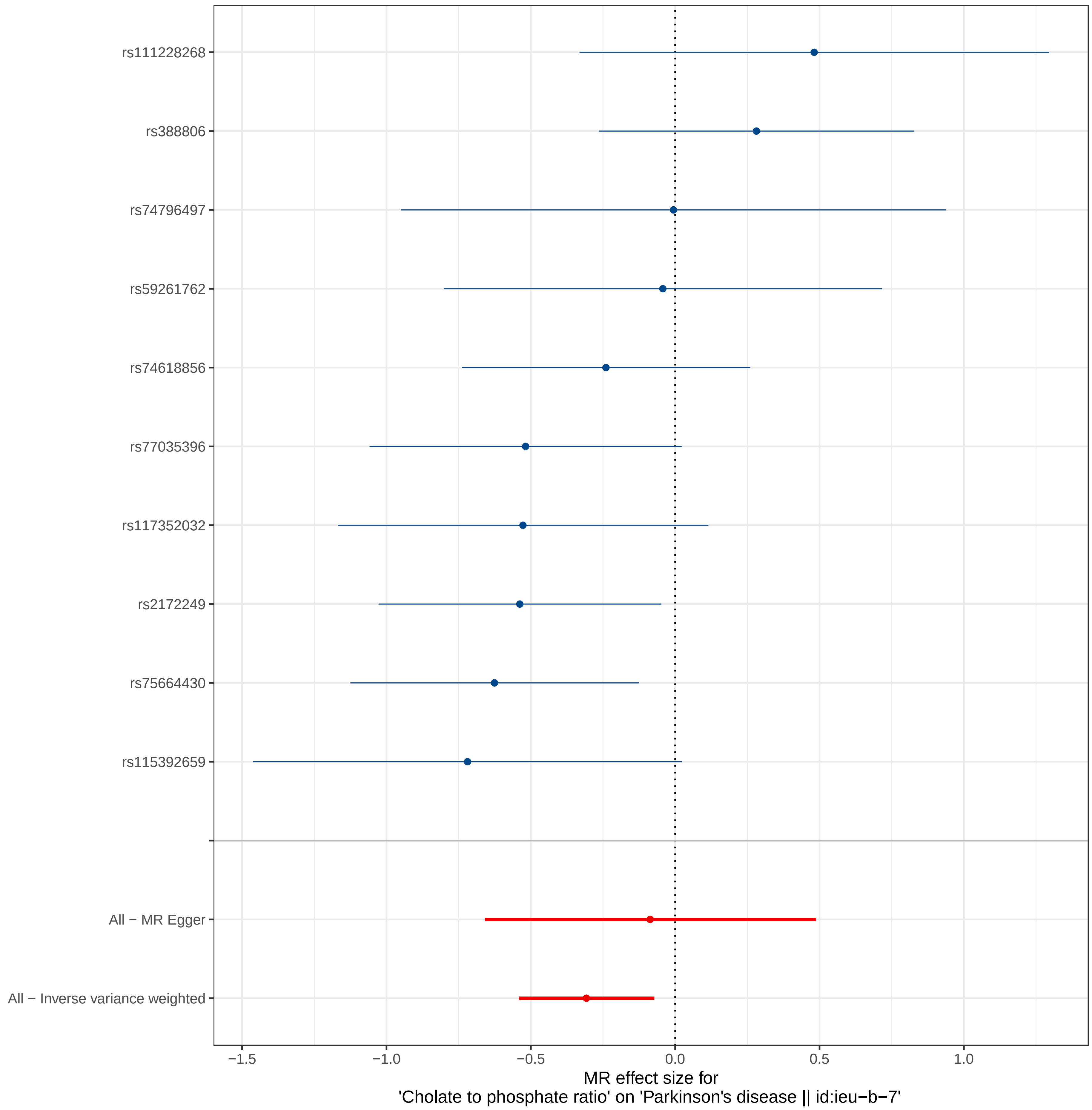
rs111228268
rs388806
rs74796497
rs59261762
rs74618856
rs77035396
rs117352032
rs2172249
rs75664430
rs115392659
All − MR Egger
All − Inverse variance weighted
−1.5
−1.0
−0.5
0.0
0.5
1.0
MR effect size for
'Cholate to phosphate ratio' on 'Parkinson's disease || id:ieu−b−7'

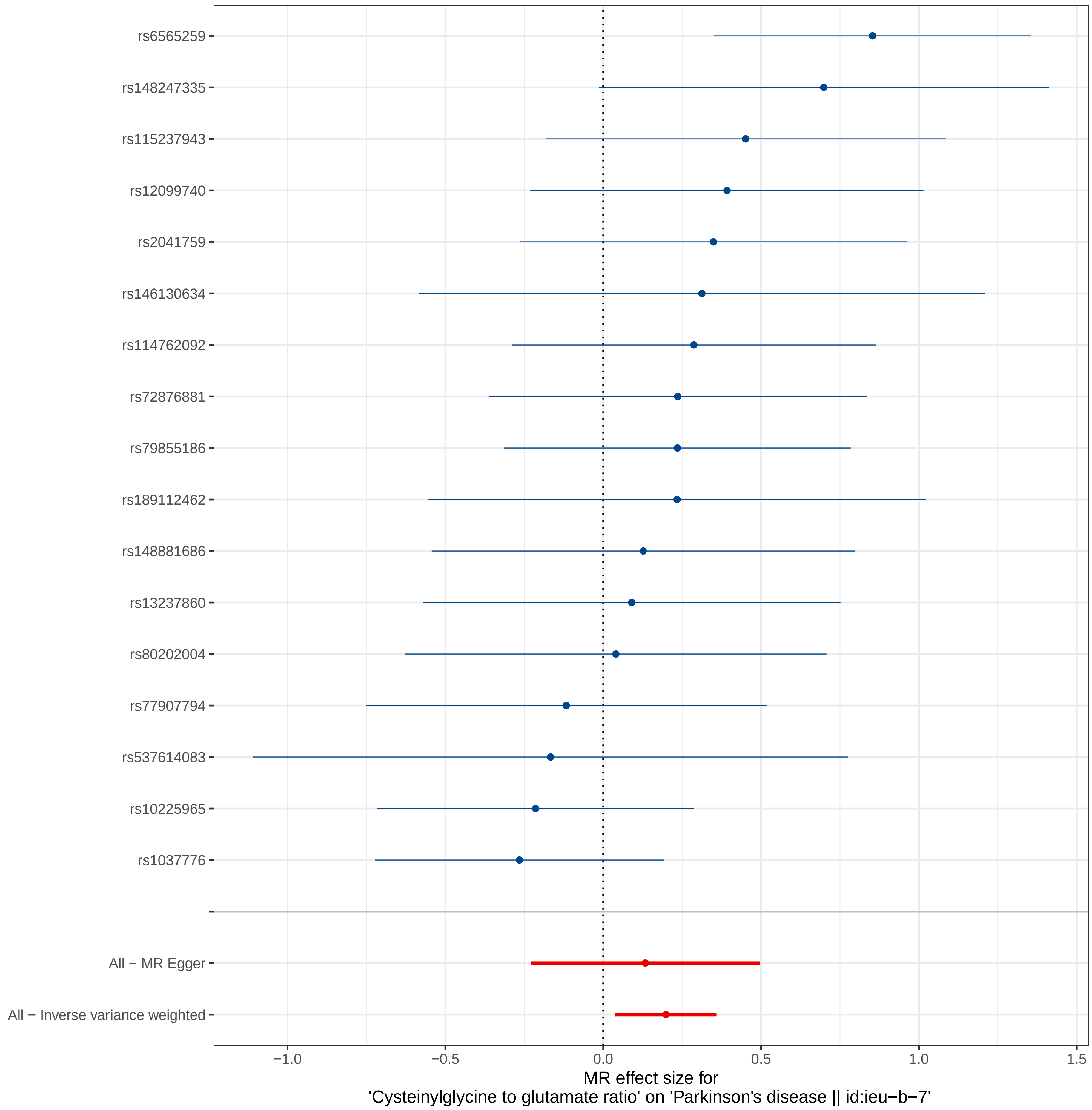
rs6565259
rs148247335
rs115237943
rs12099740
rs2041759
rs146130634
rs114762092
rs72876881
rs79855186
rs189112462
rs148881686
rs13237860
rs80202004
rs77907794
rs537614083
rs10225965
rs1037776
All − MR Egger
All − Inverse variance weighted
−1.0
−0.5
0.0
0.5
1.0
1.5
MR effect size for
'Cysteinylglycine to glutamate ratio' on 'Parkinson's disease || id:ieu−b−7'

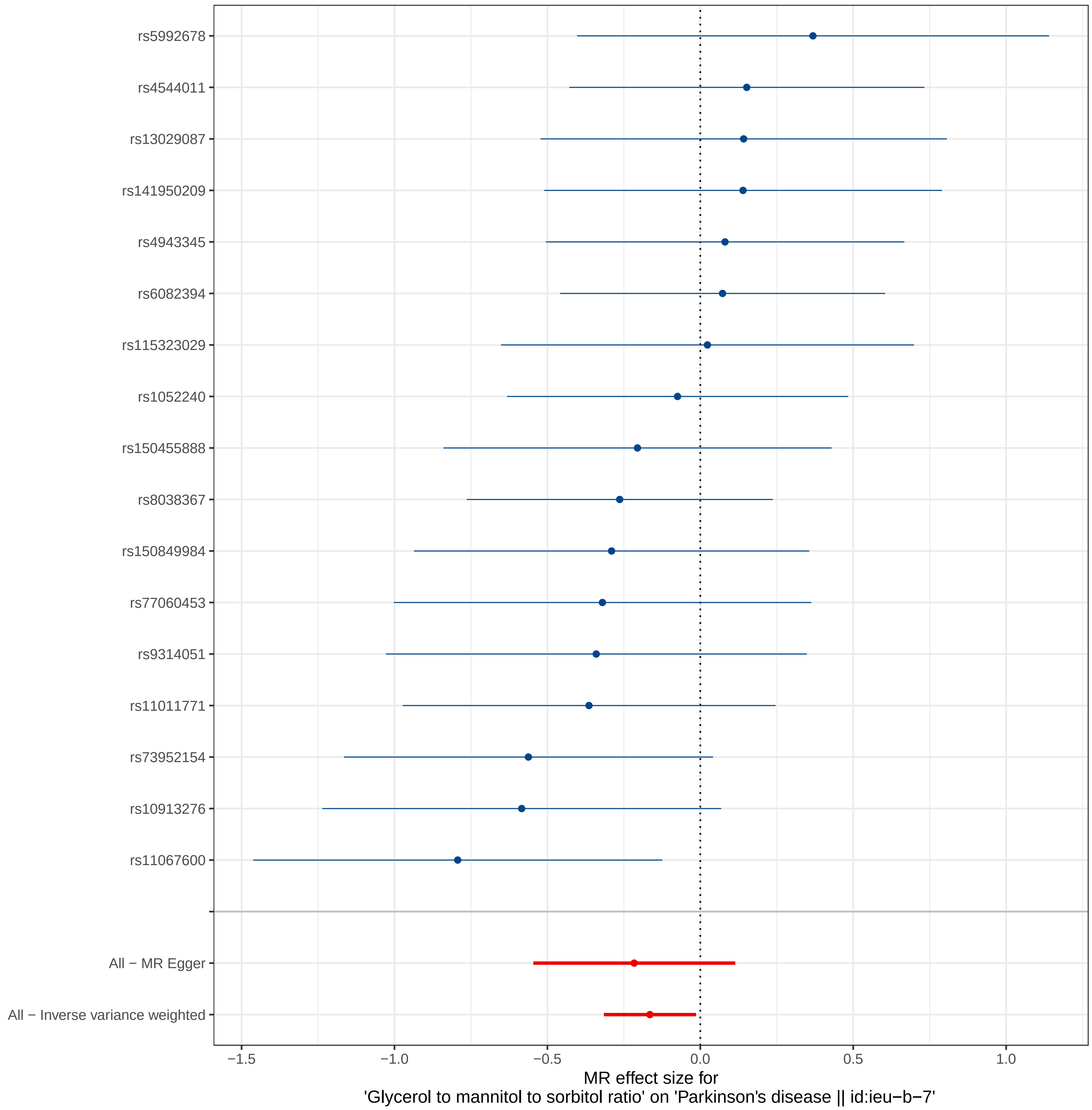

rs5992678
rs4544011
rs13029087
rs141950209
rs4943345
rs6082394
rs115323029
rs1052240
rs150455888
rs8038367
rs150849984
rs77060453
rs9314051
rs11011771
rs73952154
rs10913276
rs11067600
All − MR Egger
All − Inverse variance weighted
−1.5
−1.0
−0.5
0.0
0.5
1.0
MR effect size for
'Glycerol to mannitol to sorbitol ratio' on 'Parkinson's disease || id:ieu−b−7'

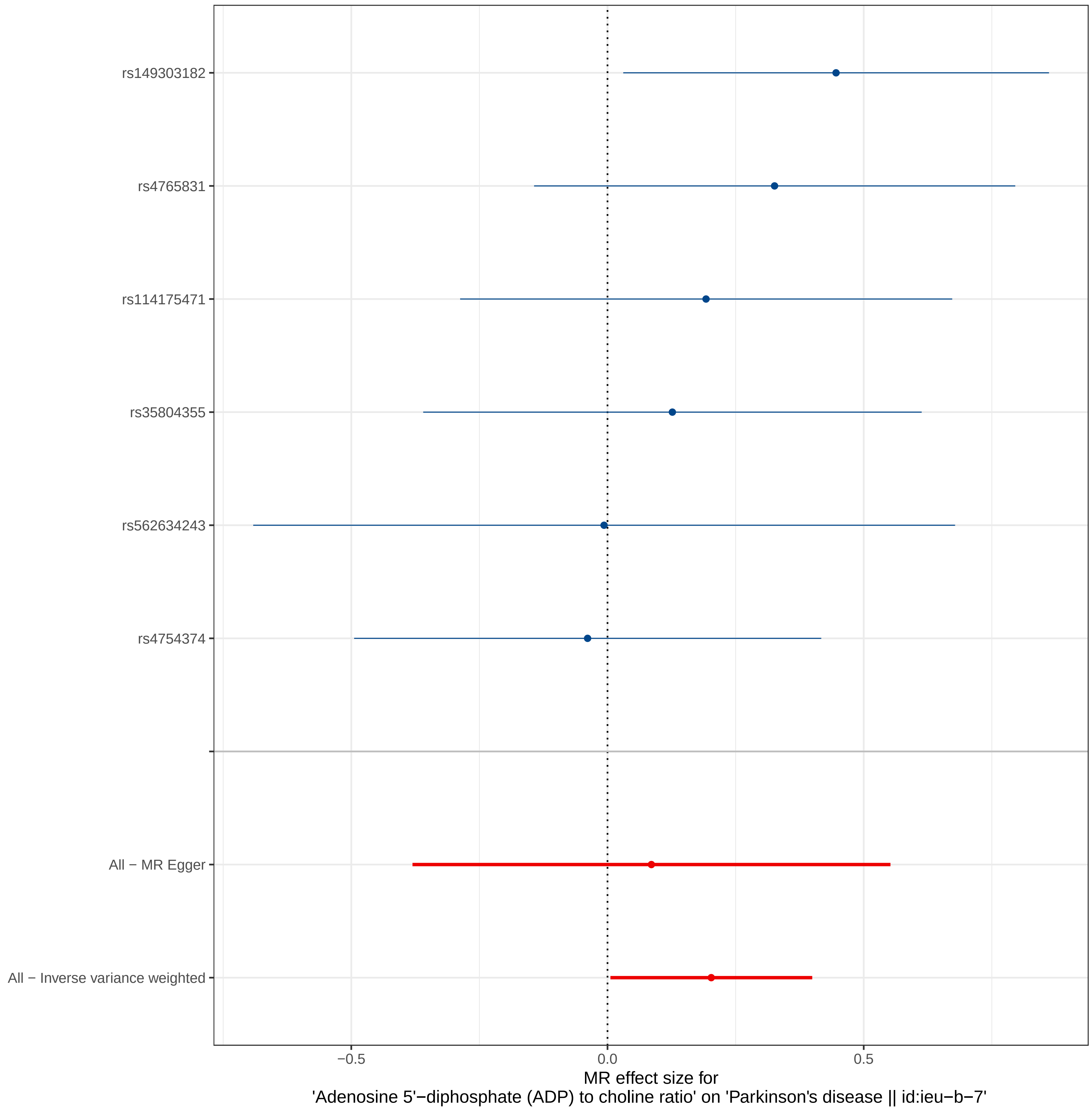

rs149303182
rs4765831
rs114175471
rs35804355
rs562634243
rs4754374
All − MR Egger
All − Inverse variance weighted
−0.5
0.0
0.5
MR effect size for
'Adenosine 5'−diphosphate (ADP) to choline ratio' on 'Parkinson's disease || id:ieu−b−7'

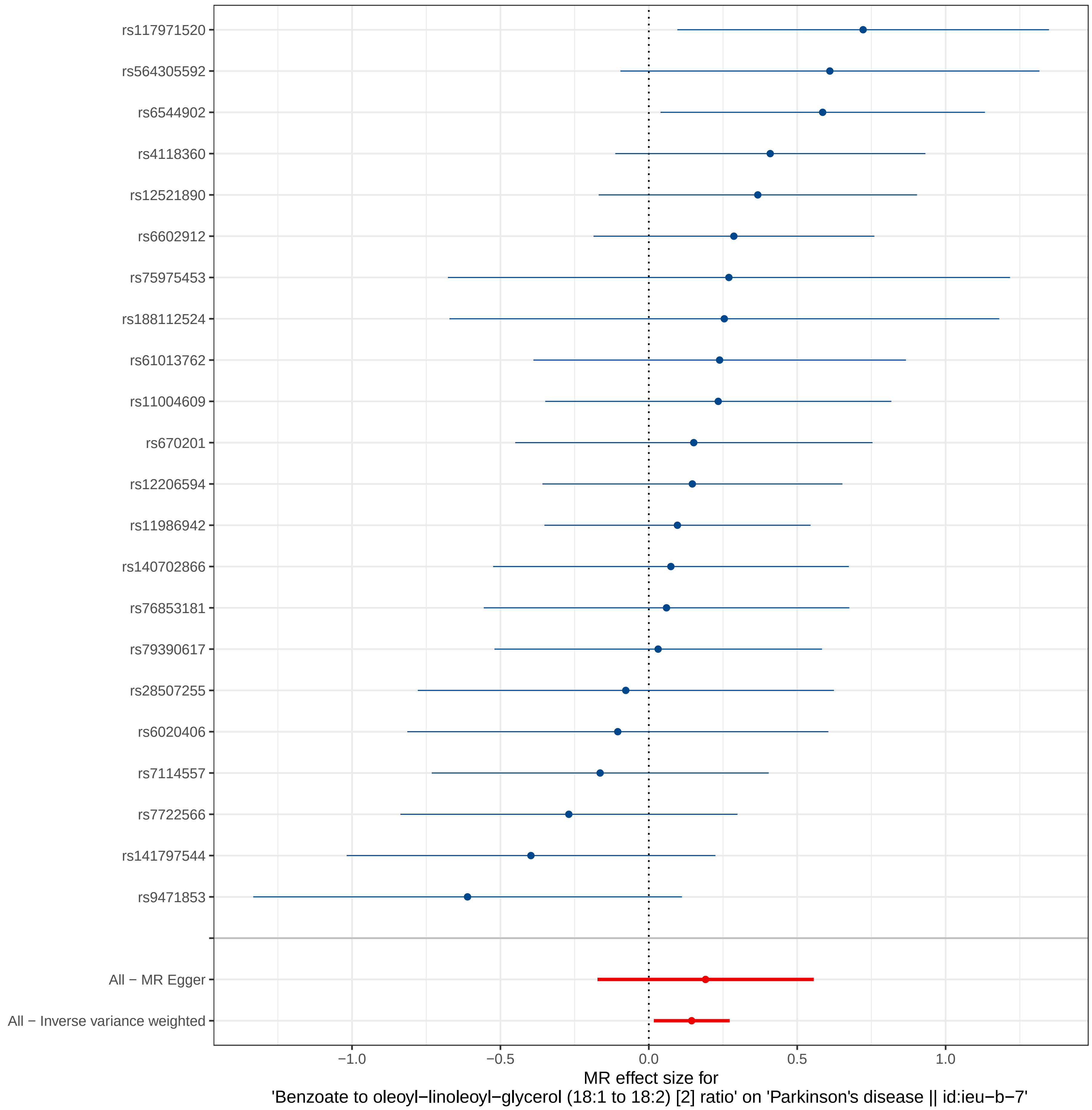
rs117971520
rs564305592
rs6544902
rs4118360
rs12521890
rs6602912
rs75975453
rs188112524
rs61013762
rs11004609
rs670201
rs12206594
rs11986942
rs140702866
rs76853181
rs79390617
rs28507255
rs6020406
rs7114557
rs7722566
rs141797544
rs9471853
All − MR Egger
All − Inverse variance weighted
−1.0
−0.5
0.0
0.5
1.0
MR effect size for
'Benzoate to oleoyl−linoleoyl−glycerol (18:1 to 18:2) [2] ratio' on 'Parkinson's disease || id:ieu−b−7'

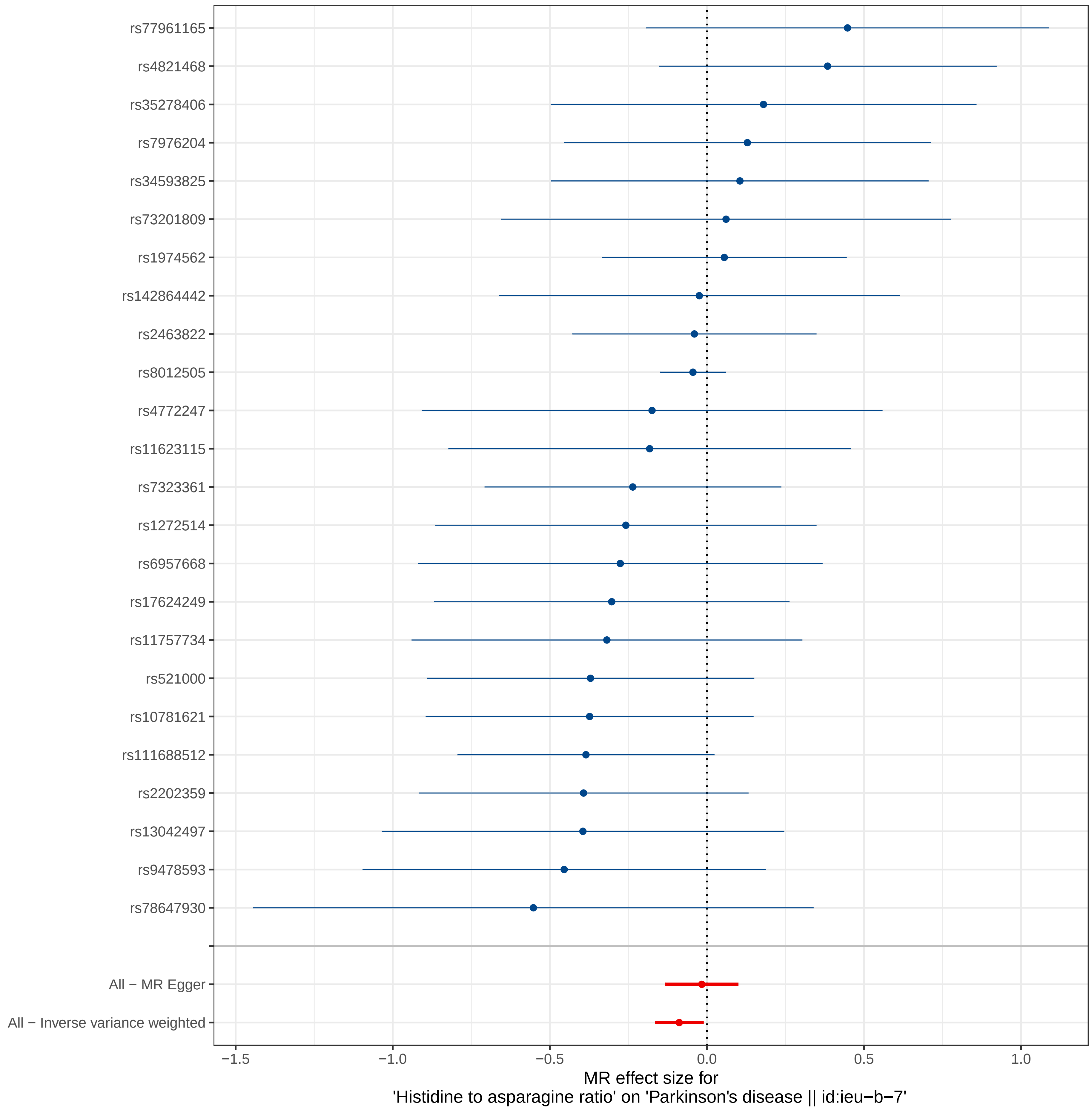
rs77961165
rs4821468
rs35278406
rs7976204
rs34593825
rs73201809
rs1974562
rs142864442
rs2463822
rs8012505
rs4772247
rs11623115
rs7323361
rs1272514
rs6957668
rs17624249
rs11757734
rs521000
rs10781621
rs111688512
rs2202359
rs13042497
rs9478593
rs78647930
All − MR Egger
All − Inverse variance weighted
−1.5
−1.0
−0.5
0.0
0.5
1.0
MR effect size for
'Histidine to asparagine ratio' on 'Parkinson's disease || id:ieu−b−7'

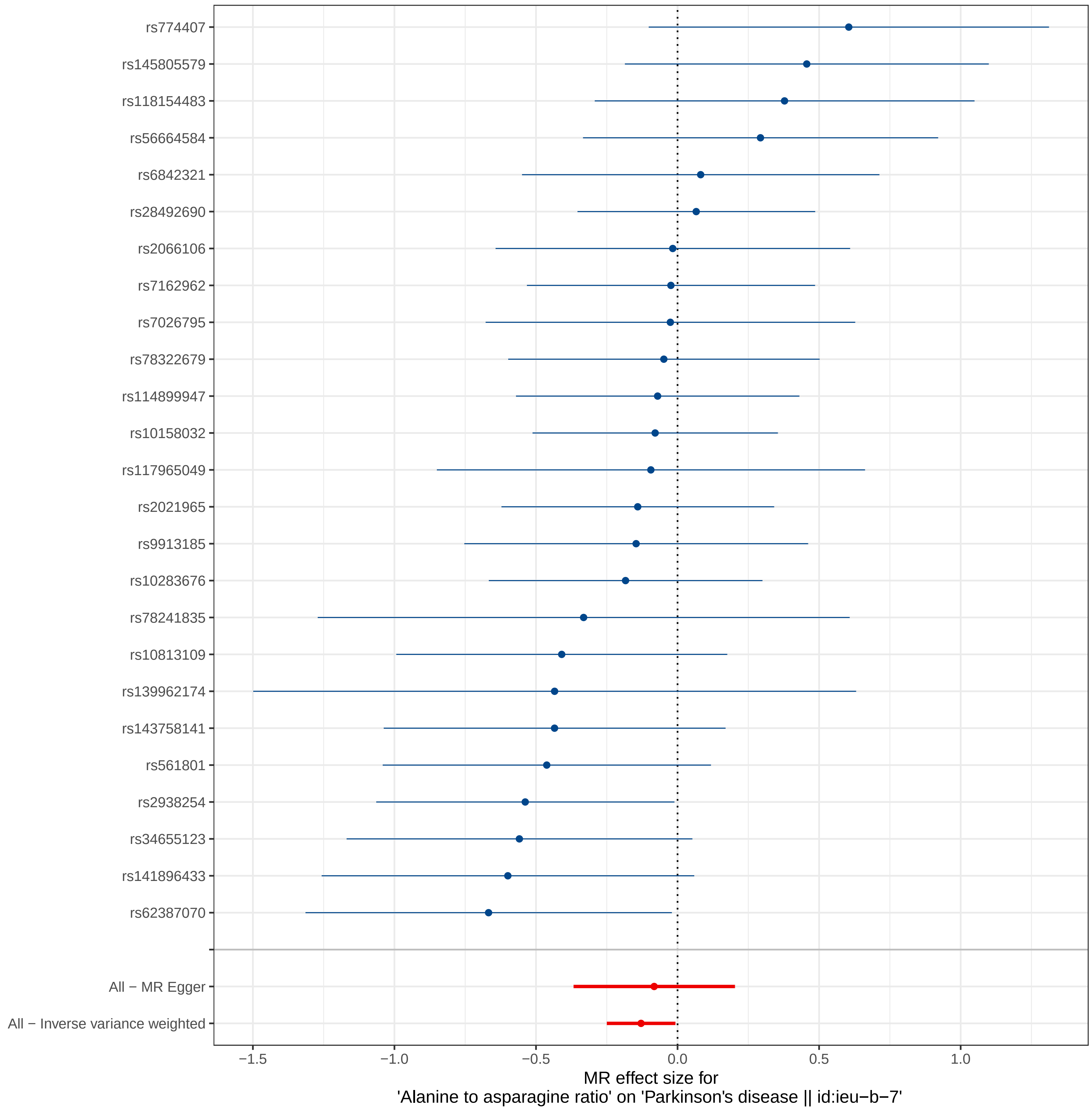
rs774407
rs145805579
rs118154483
rs56664584
rs6842321
rs28492690
rs2066106
rs7162962
rs7026795
rs78322679
rs114899947
rs10158032
rs117965049
rs2021965
rs9913185
rs10283676
rs78241835
rs10813109
rs139962174
rs143758141
rs561801
rs2938254
rs34655123
rs141896433
rs62387070
All − MR Egger
All − Inverse variance weighted
−1.5
−1.0
−0.5
0.0
0.5
1.0
MR effect size for
'Alanine to asparagine ratio' on 'Parkinson's disease || id:ieu−b−7'

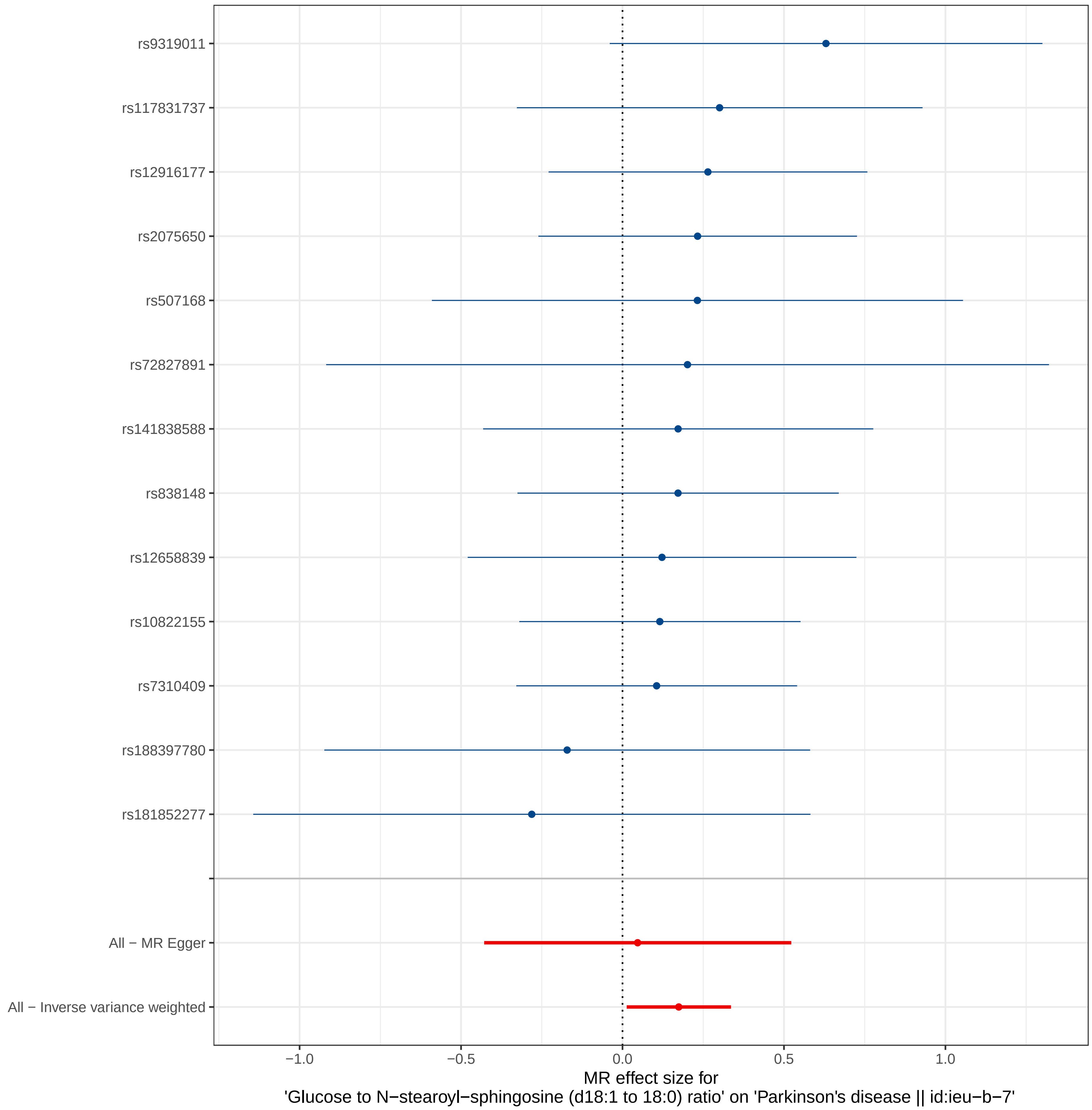

rs9319011
rs117831737
rs12916177
rs2075650
rs507168
rs72827891
rs141838588
rs838148
rs12658839
rs10822155
rs7310409
rs188397780
rs181852277
All − MR Egger
All − Inverse variance weighted
−1.0
−0.5
0.0
0.5
1.0
MR effect size for
'Glucose to N−stearoyl−sphingosine (d18:1 to 18:0) ratio' on 'Parkinson's disease || id:ieu−b−7'
