## Supplemental Figure 6 for "Cerebrospinal Fluid and Plasma Metabolites with Parkinson’s Disease: A Mendelian Randomization Study"

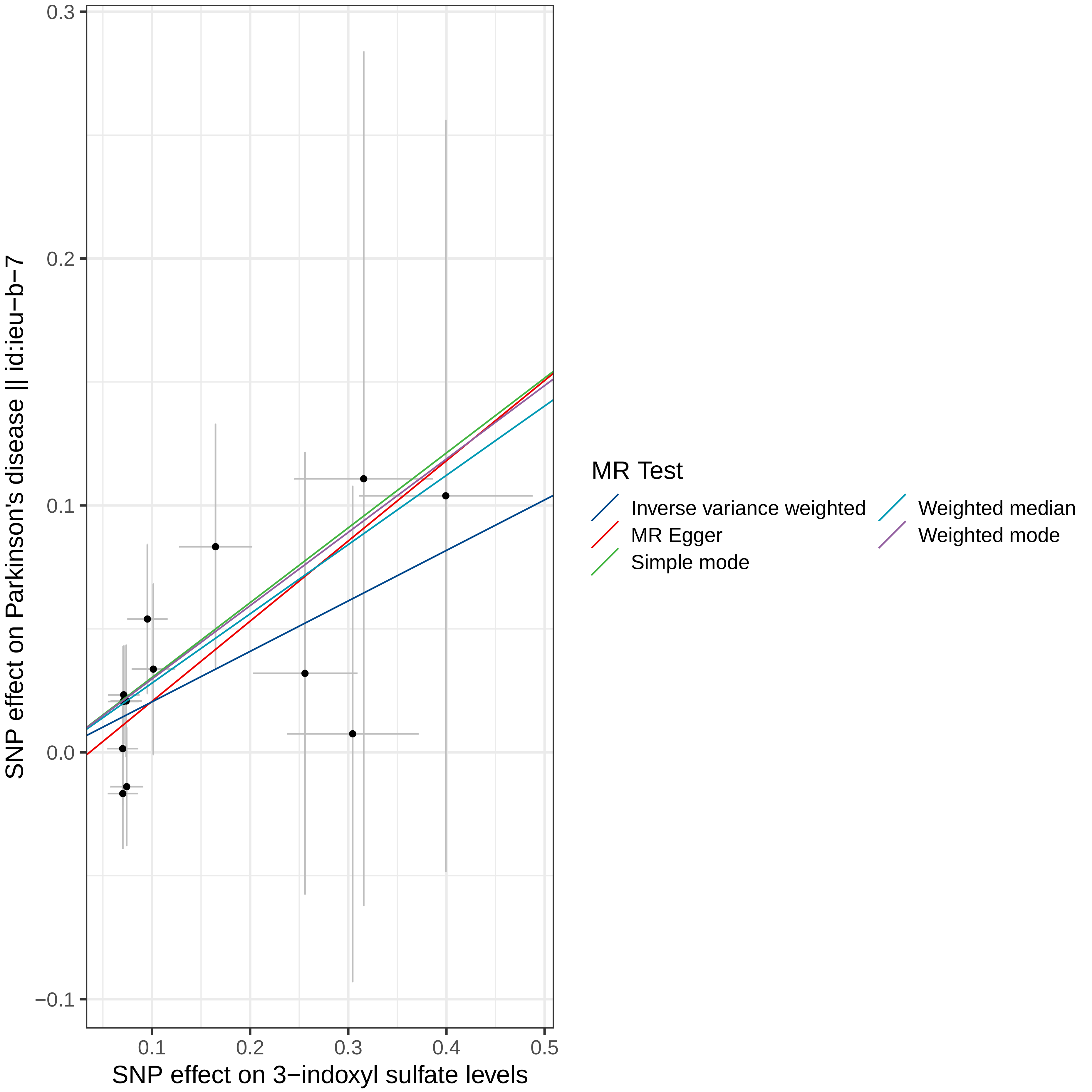
0.3
0.2
0.1
0.0
−0.1
0.1
0.2
0.3
0.4
0.5
SNP effect on Parkinson's disease || id:ieu−b−7
SNP effect on 3−indoxyl sulfate levels
MR Test
Inverse variance weighted
MR Egger
Simple mode
Weighted median
Weighted mode

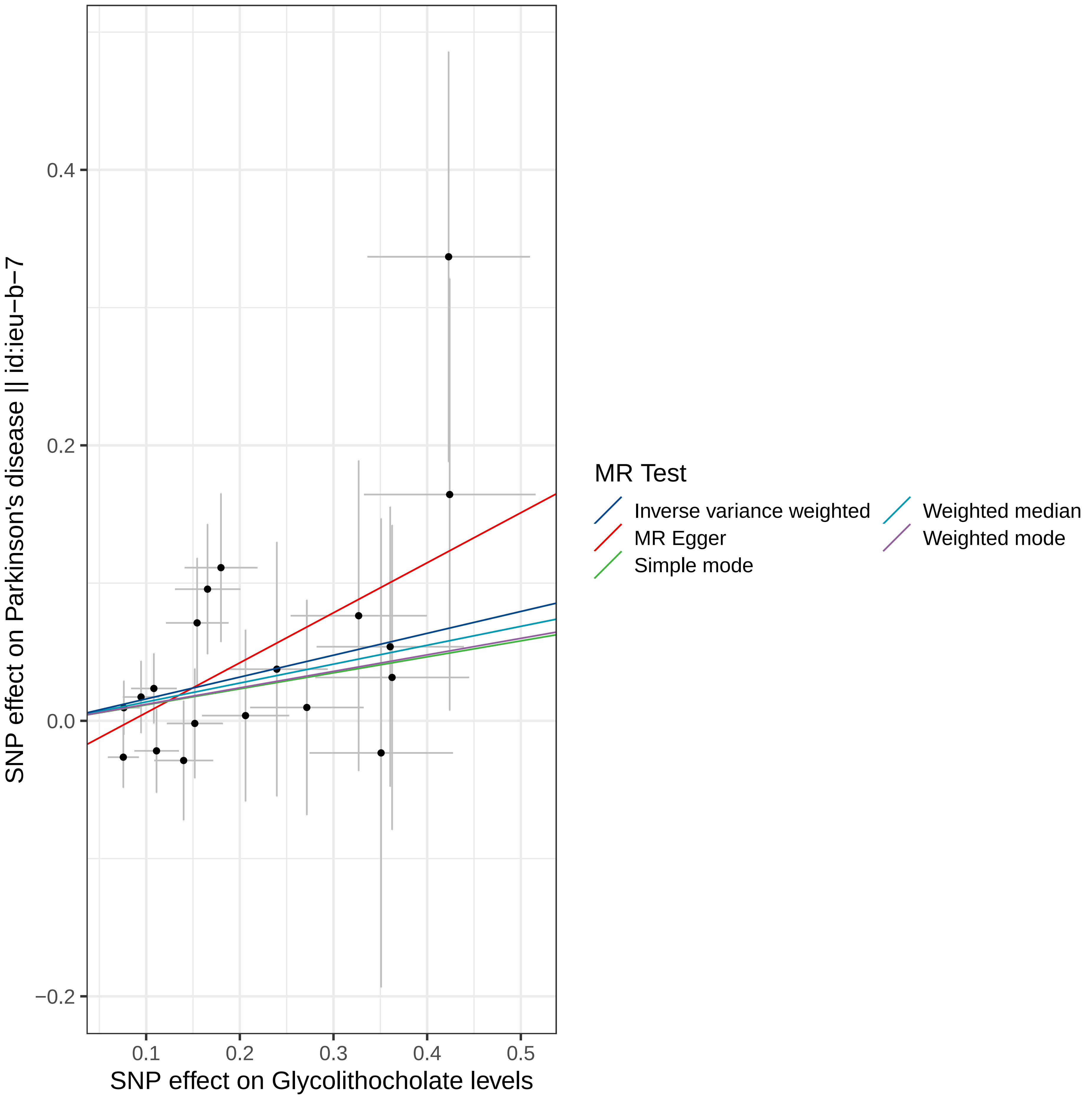

SNP effect on Parkinson's disease || id:ieu−b−7
0.4
0.2
0.0
−0.2
0.1
0.2
0.3
0.4
0.5
SNP effect on Glycolithocholate levels
MR Test
Inverse variance weighted
MR Egger
Simple mode
Weighted median
Weighted mode

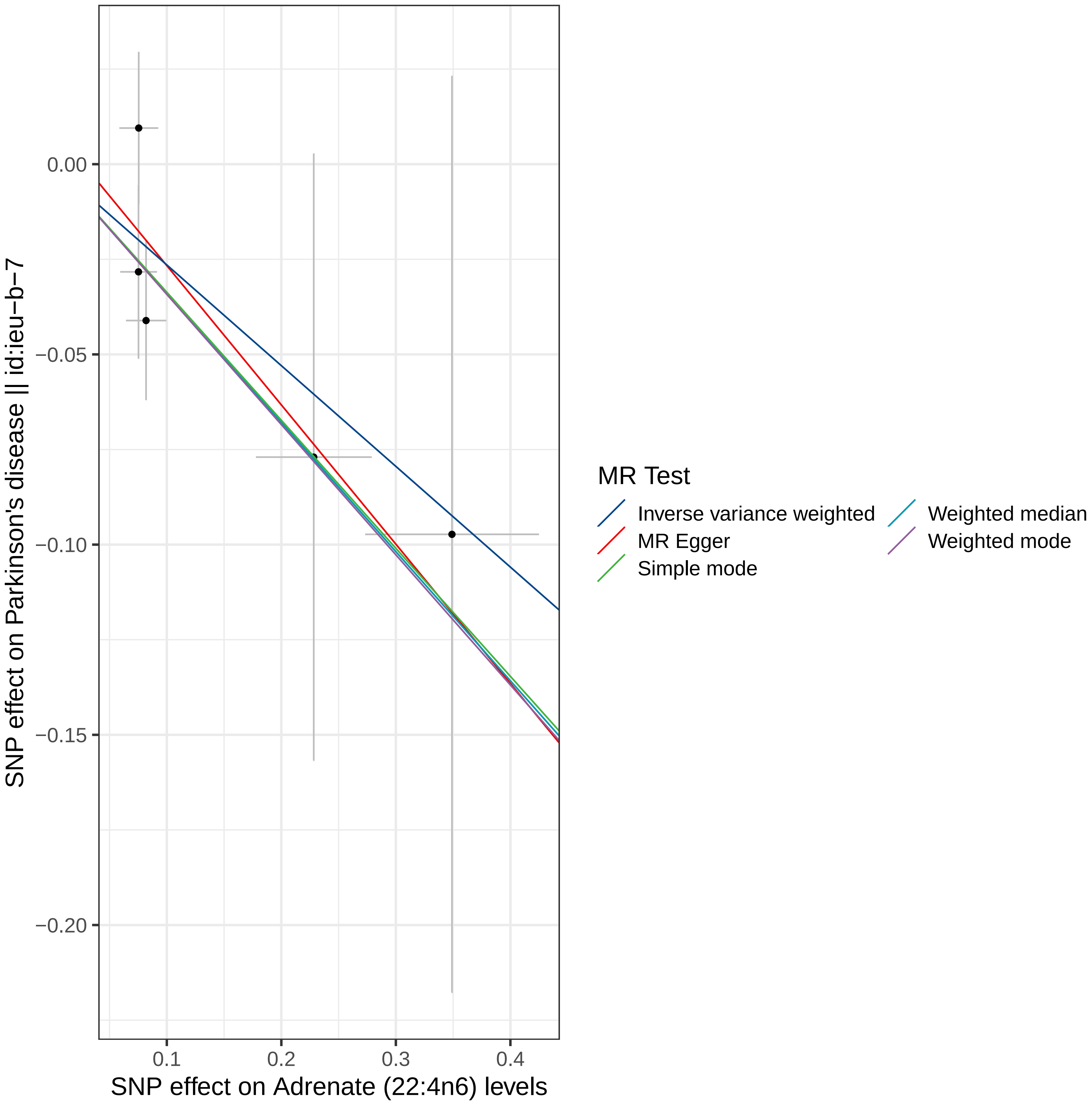

0.00
−0.05
−0.10
−0.15
−0.20
0.1
0.2
0.3
0.4
SNP effect on Parkinson's disease || id:ieu−b−7
SNP effect on Adrenate (22:4n6) levels
MR Test
Inverse variance weighted
MR Egger
Simple mode
Weighted median
Weighted mode

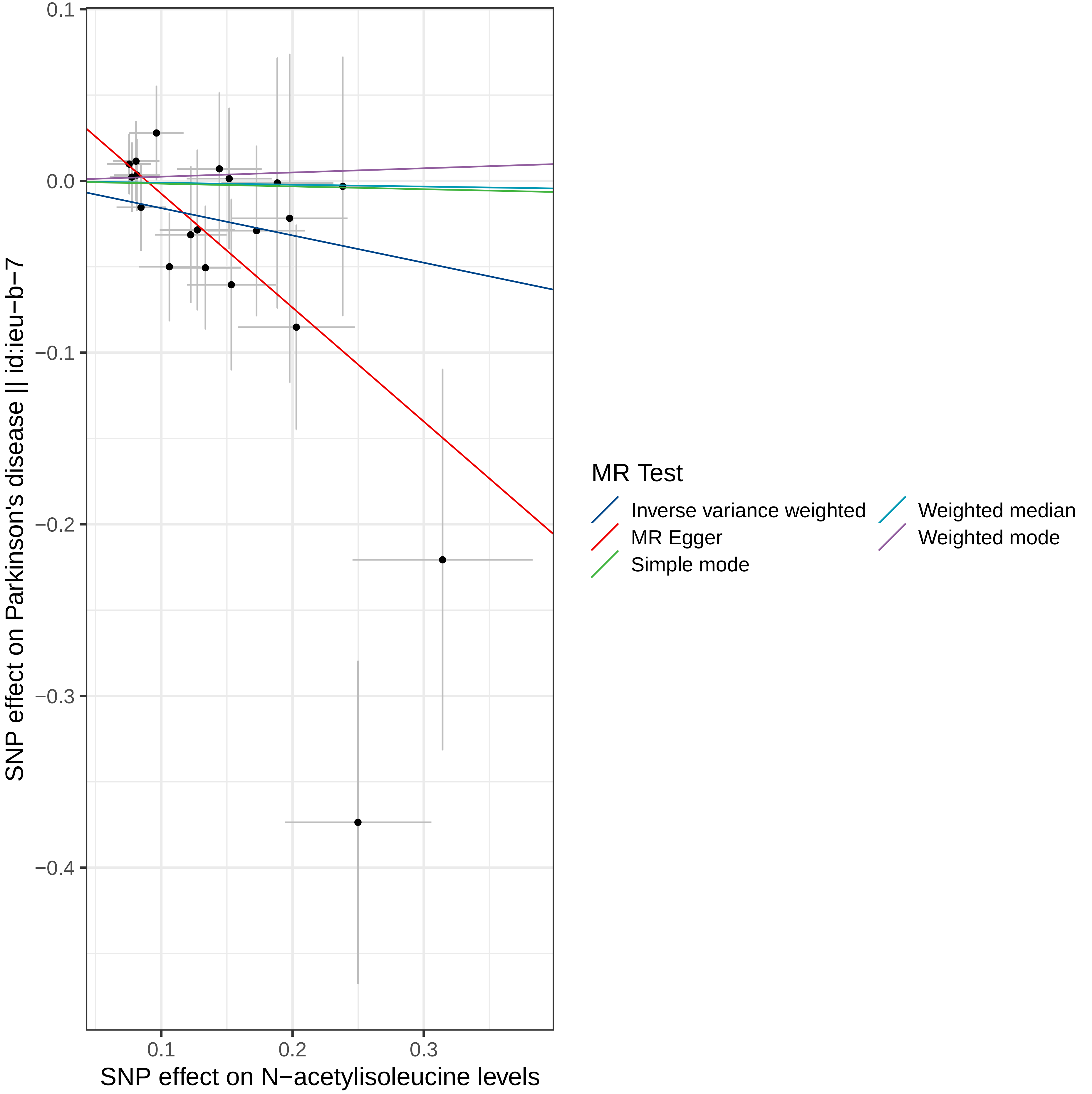

0.1
0.0
−0.1
−0.2
−0.3
−0.4
0.1
0.2
0.3
SNP effect on Parkinson's disease || id:ieu-b-7
SNP effect on N−acetylisoleucine levels
MR Test
Inverse variance weighted
MR Egger
Simple mode
Weighted median
Weighted mode

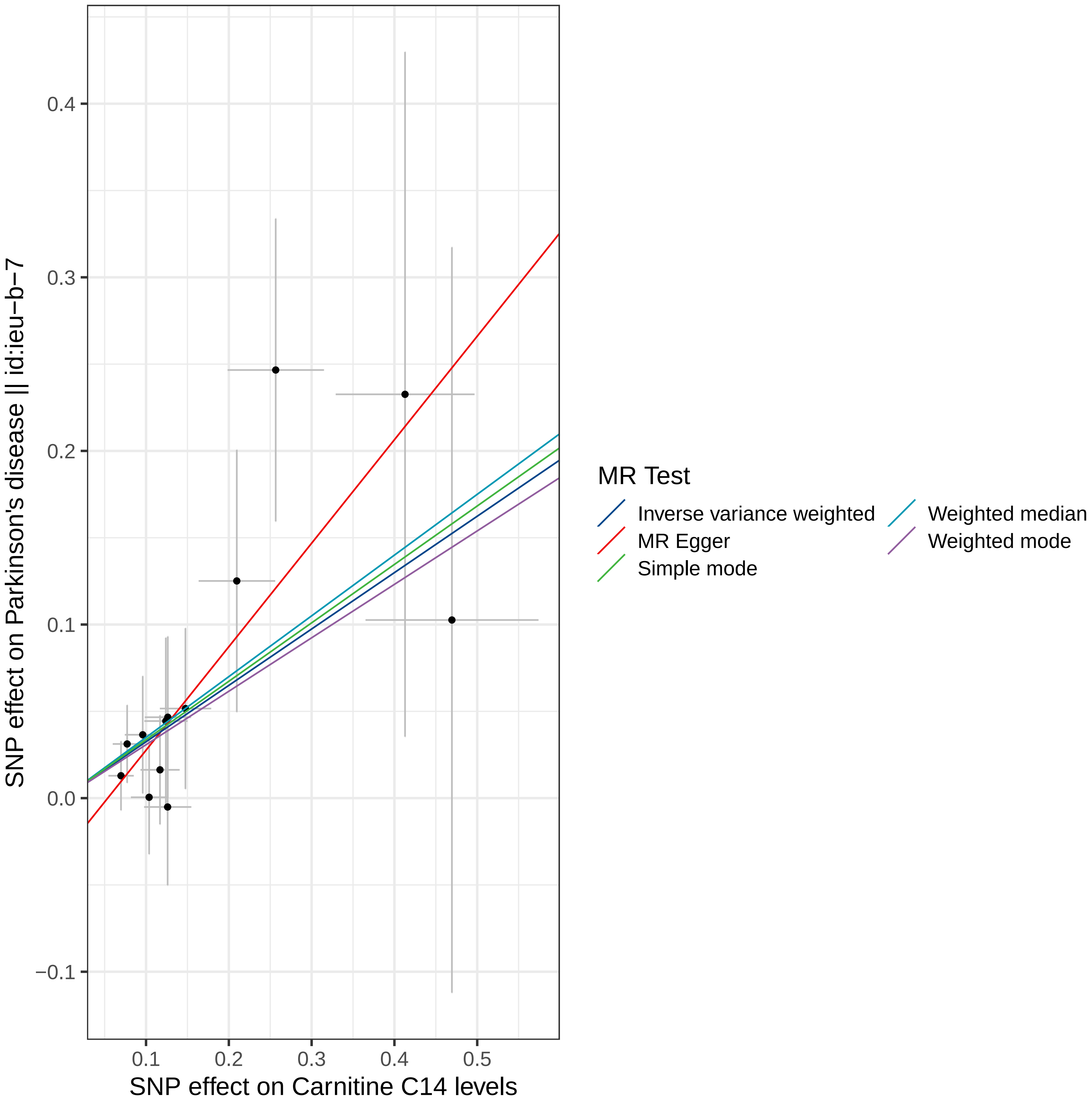

0.4
0.3
0.2
0.1
0.0
−0.1
0.1
0.2
0.3
0.4
0.5
SNP effect on Parkinson's disease || id:ieu−b−7
SNP effect on Carnitine C14 levels
MR Test
Inverse variance weighted
MR Egger
Simple mode
Weighted median
Weighted mode

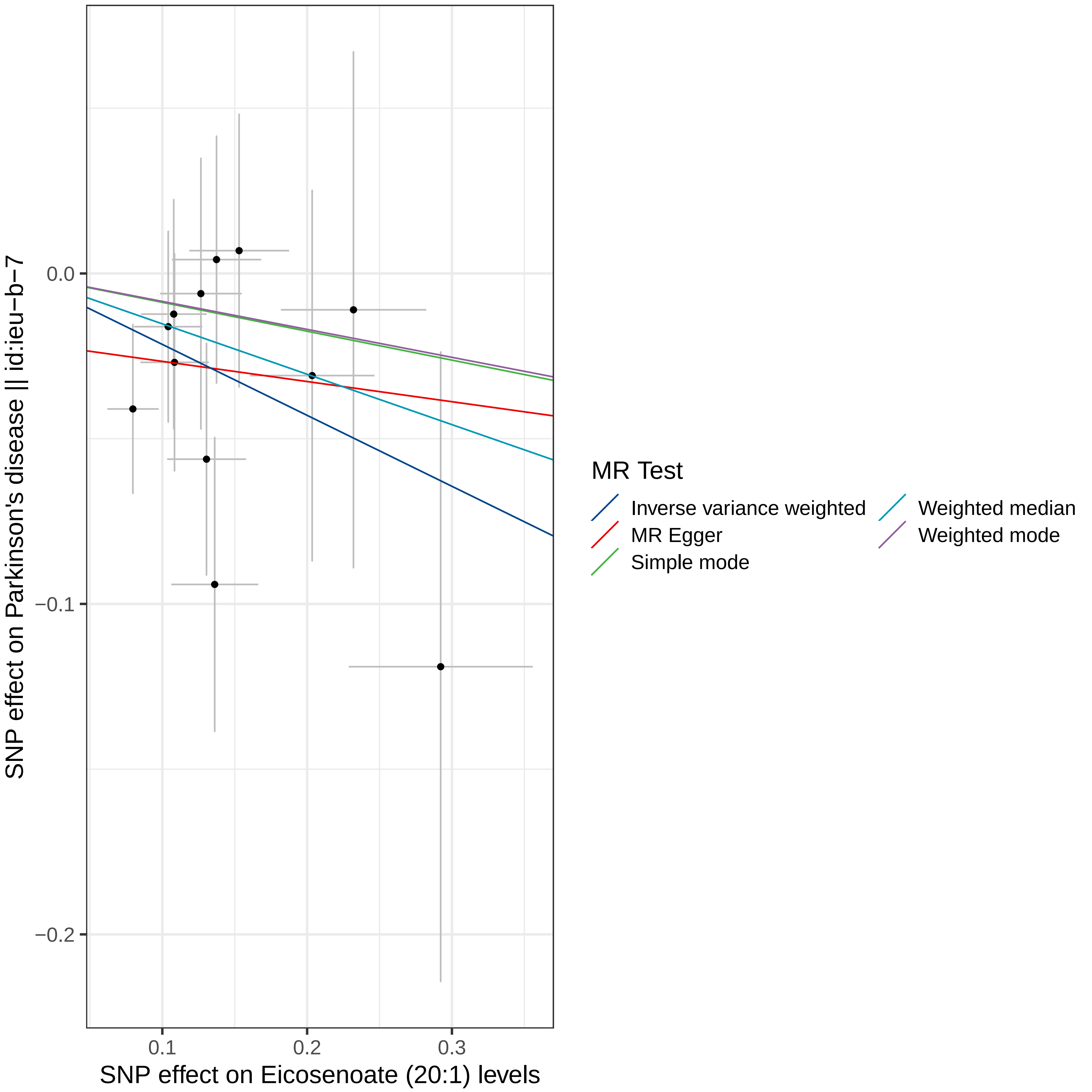

SNP effect on Parkinson's disease || id:ieu−b−7
0.0
−0.1
−0.2
0.1
0.2
0.3
SNP effect on Eicosenoate (20:1) levels
MR Test
Inverse variance weighted
MR Egger
Simple mode
Weighted median
Weighted mode

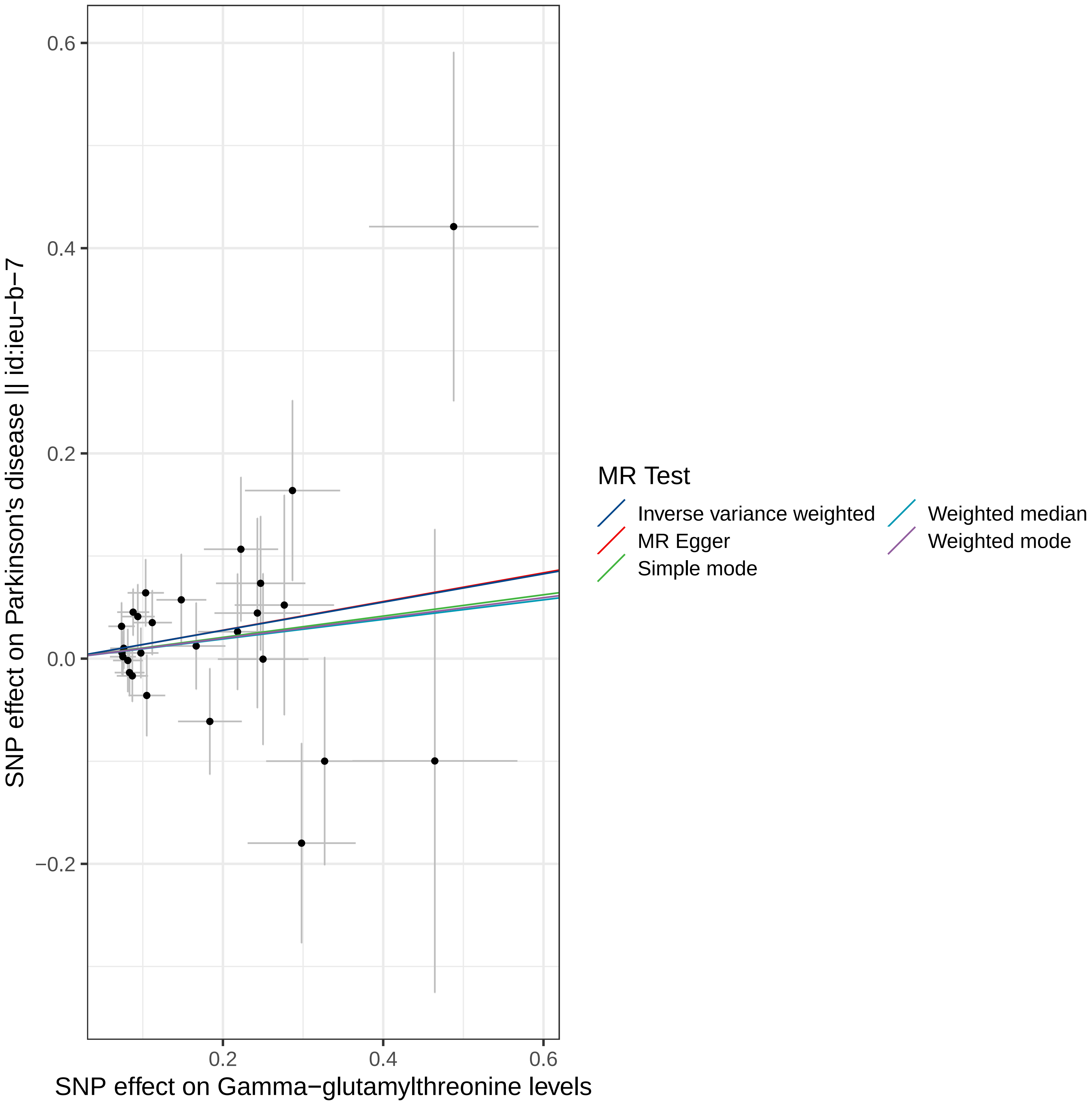

SNP effect on Parkinson's disease || id:ieu−b−7
SNP effect on Gamma−glutamylthreonine levels
0.6
0.4
0.2
0.0
−0.2
0.2
0.4
0.6
MR Test
Inverse variance weighted
MR Egger
Simple mode
Weighted median
Weighted mode

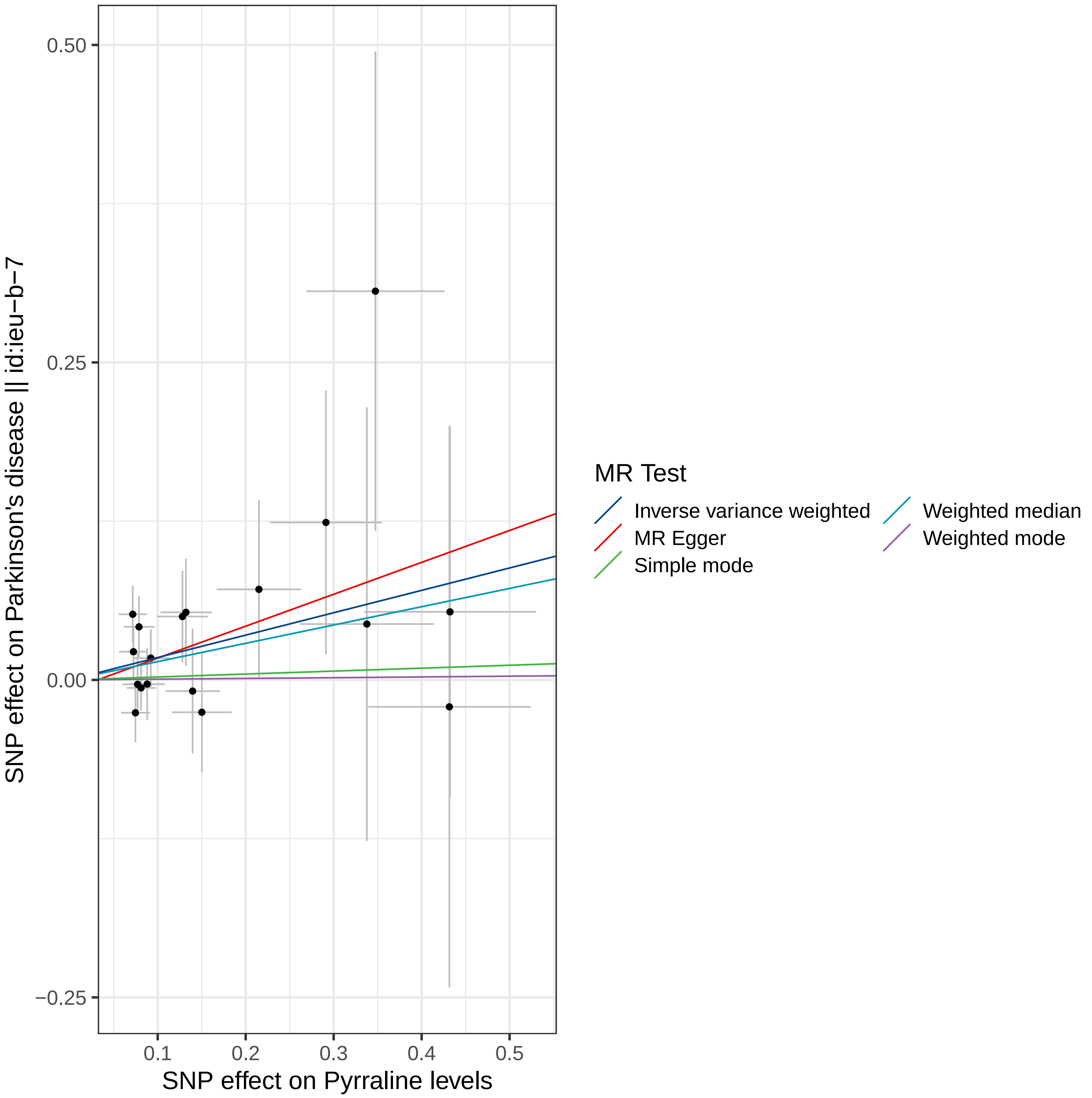

0.50
0.25
0.00
−0.25
0.1
0.2
0.3
0.4
0.5
SNP effect on Parkinson's disease || id:ieu−b−7
SNP effect on Pyrraline levels
MR Test
Inverse variance weighted
MR Egger
Simple mode
Weighted median
Weighted mode

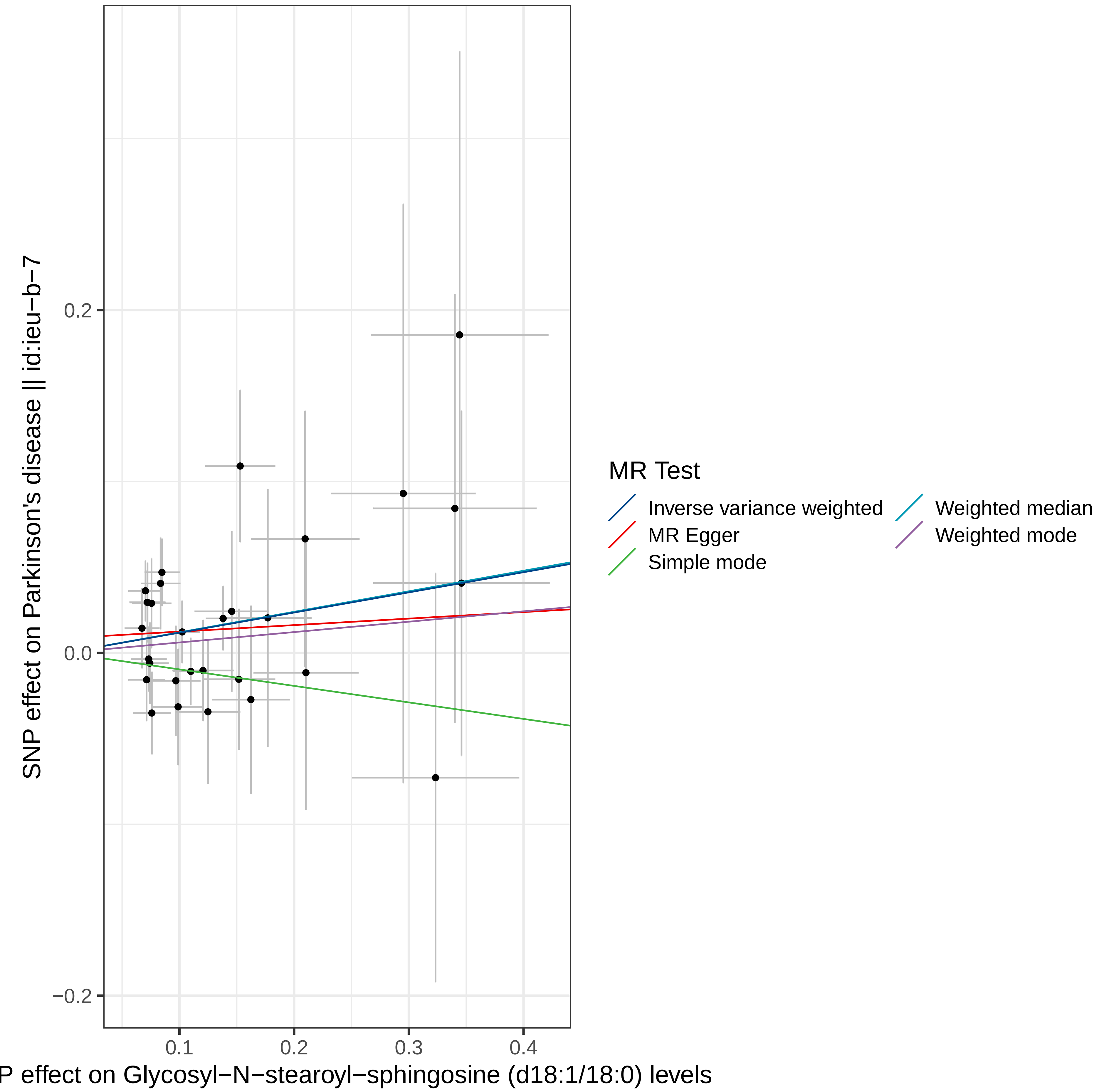

SNP effect on Parkinson's disease || id:ieu−b−7
0.2
0.0
−0.2
0.1
0.2
0.3
0.4
P effect on Glycosyl−N−stearoyl−sphingosine (d18:1/18:0) levels
MR Test
Inverse variance weighted
MR Egger
Simple mode
Weighted median
Weighted mode

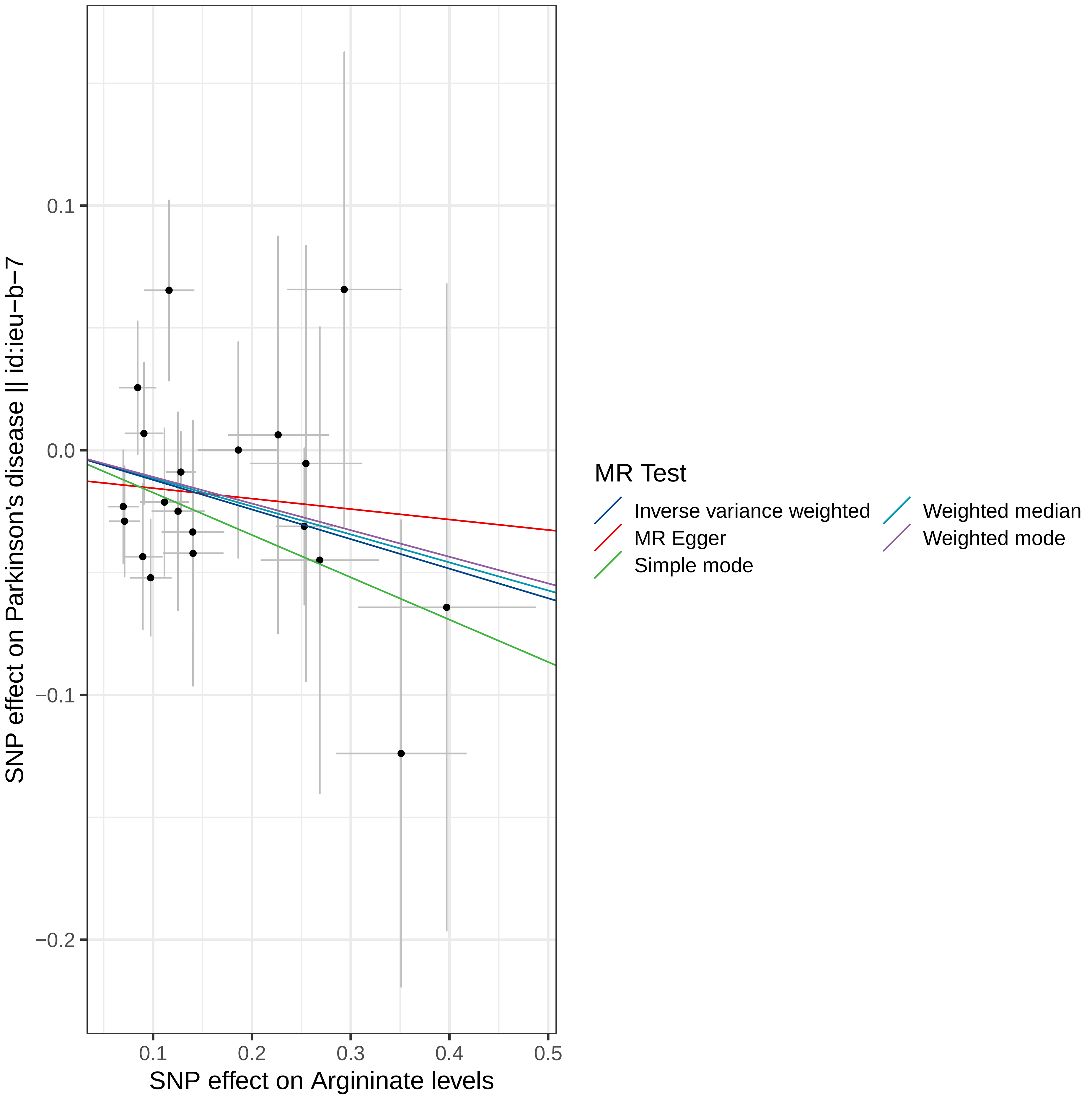

0.1
0.0
−0.1
−0.2
SNP effect on Parkinson's disease || id:ieu−b−7
0.1
0.2
0.3
0.4
0.5
SNP effect on Arginate levels
MR Test
Inverse variance weighted
MR Egger
Simple mode
Weighted median
Weighted mode

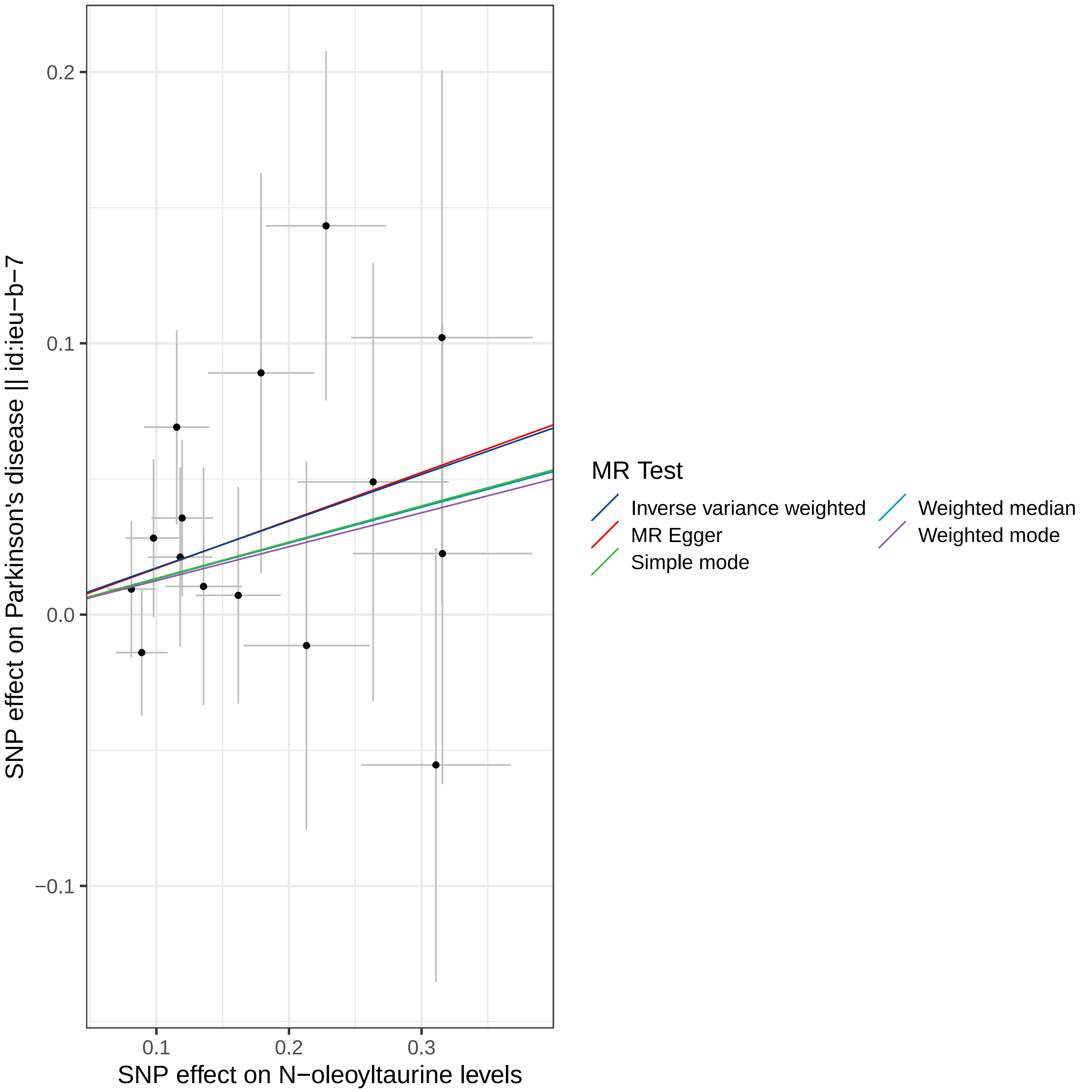

0.2
0.1
0.0
−0.1
0.1
0.2
0.3
SNP effect on Parkinson's disease || id:ieu−b−7
SNP effect on N−oleoyltaurine levels
MR Test
Inverse variance weighted
MR Egger
Simple mode
Weighted median
Weighted mode

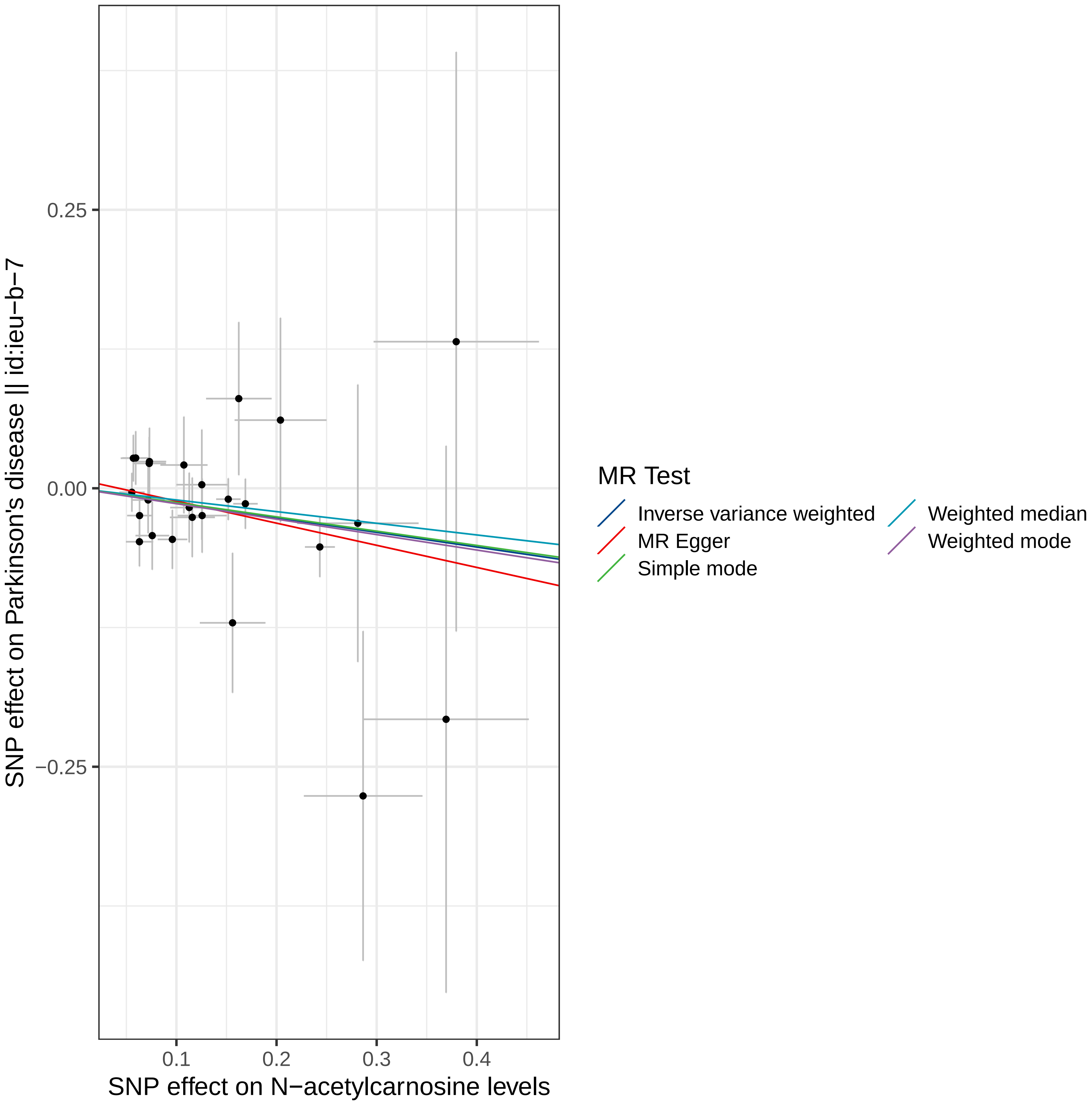

SNP effect on Parkinson's disease || id:ieu−b−7
0.25
0.00
−0.25
0.1
0.2
0.3
0.4
SNP effect on N−acetylcarnosine levels
MR Test
Inverse variance weighted
MR Egger
Simple mode
Weighted median
Weighted mode

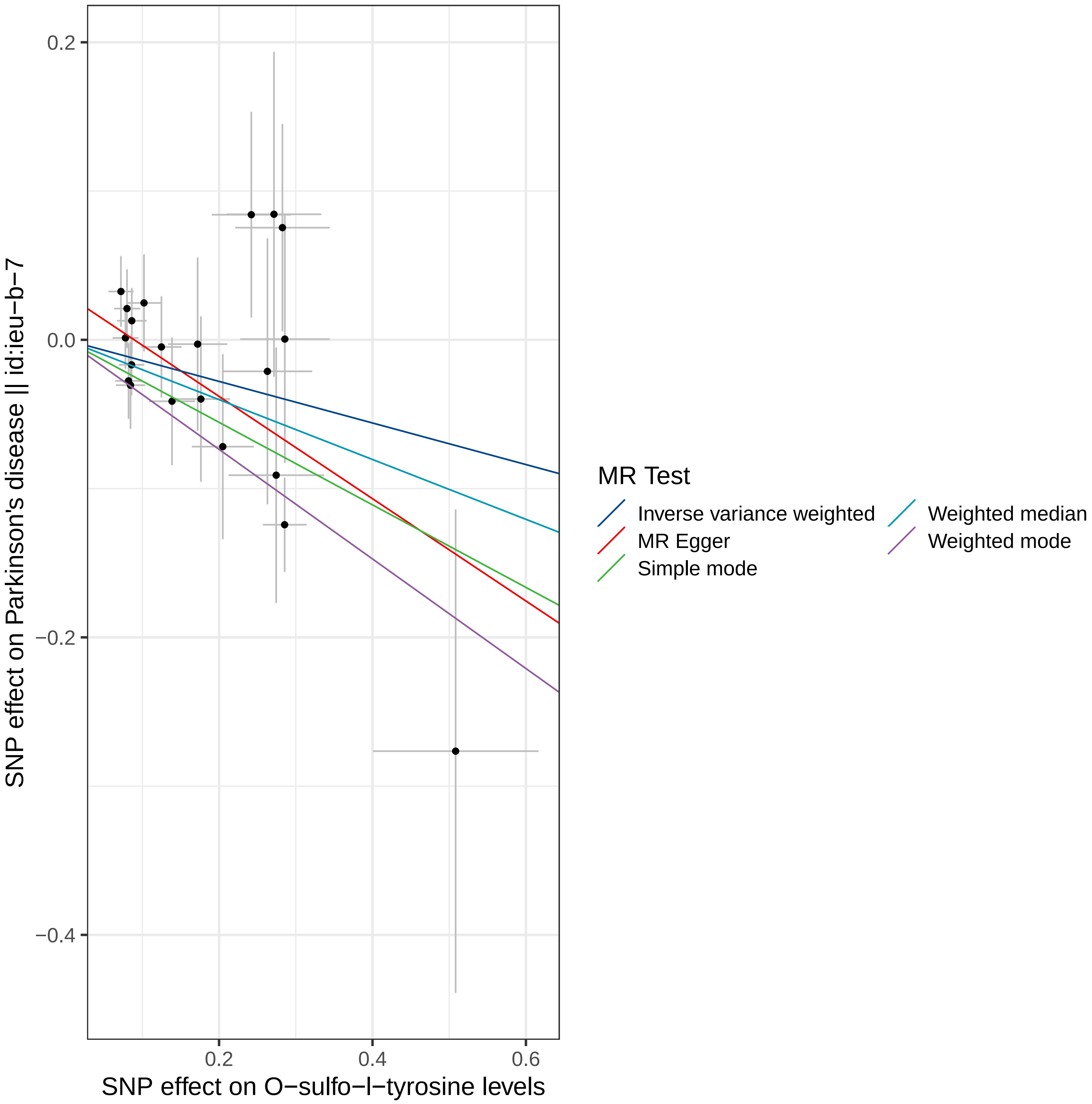

0.2
0.0
−0.2
−0.4
0.2
0.4
0.6
SNP effect on Parkinson's disease || id:ieu−b−7
SNP effect on O−sulfo−l−tyrosine levels
MR Test
Inverse variance weighted
MR Egger
Simple mode
Weighted median
Weighted mode

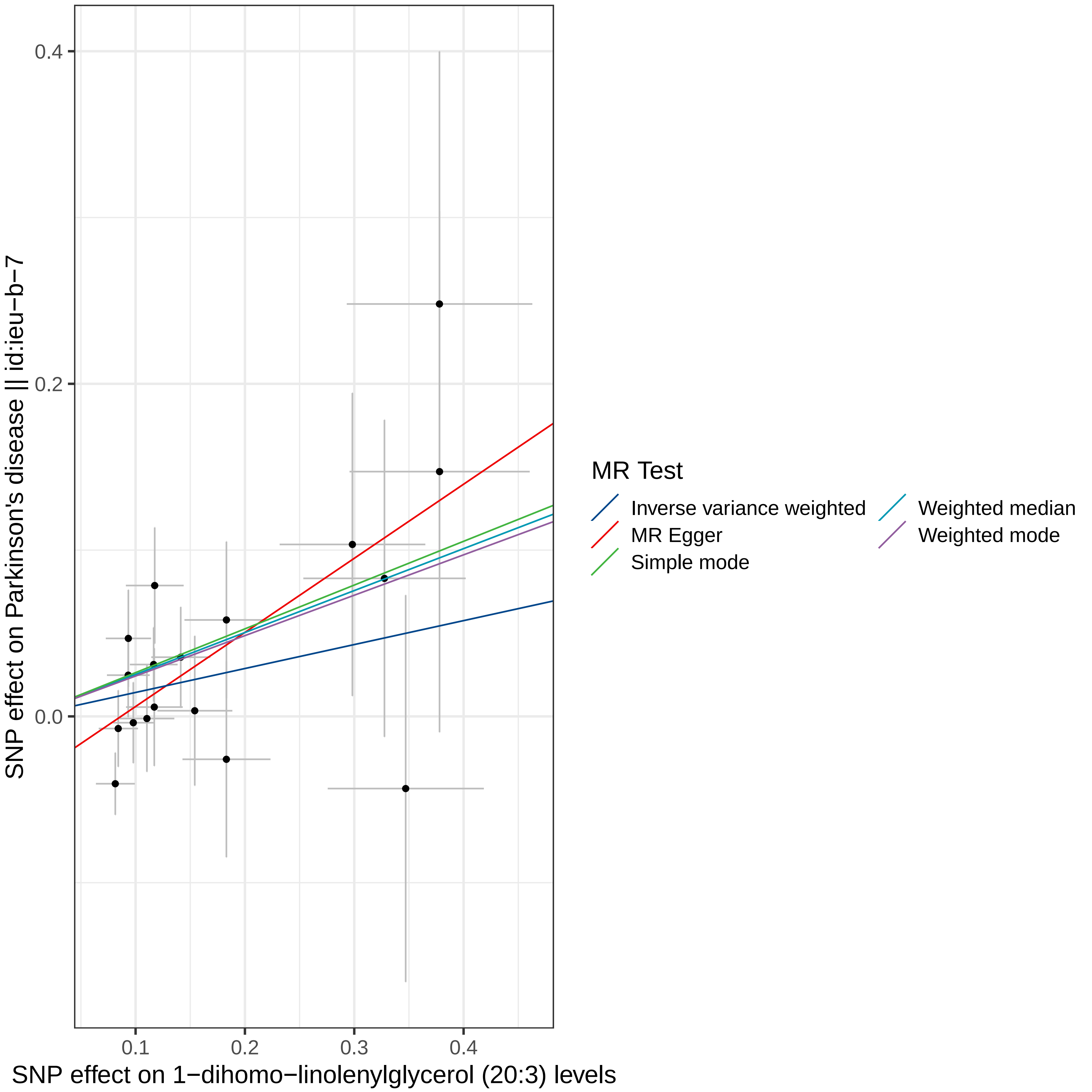

0.4
0.2
0.0
0.1
0.2
0.3
0.4
SNP effect on Parkinson's disease || id:ieu−b−7
SNP effect on 1−dihomo−linolenylglycerol (20:3) levels
MR Test
Inverse variance weighted
MR Egger
Simple mode
Weighted median
Weighted mode

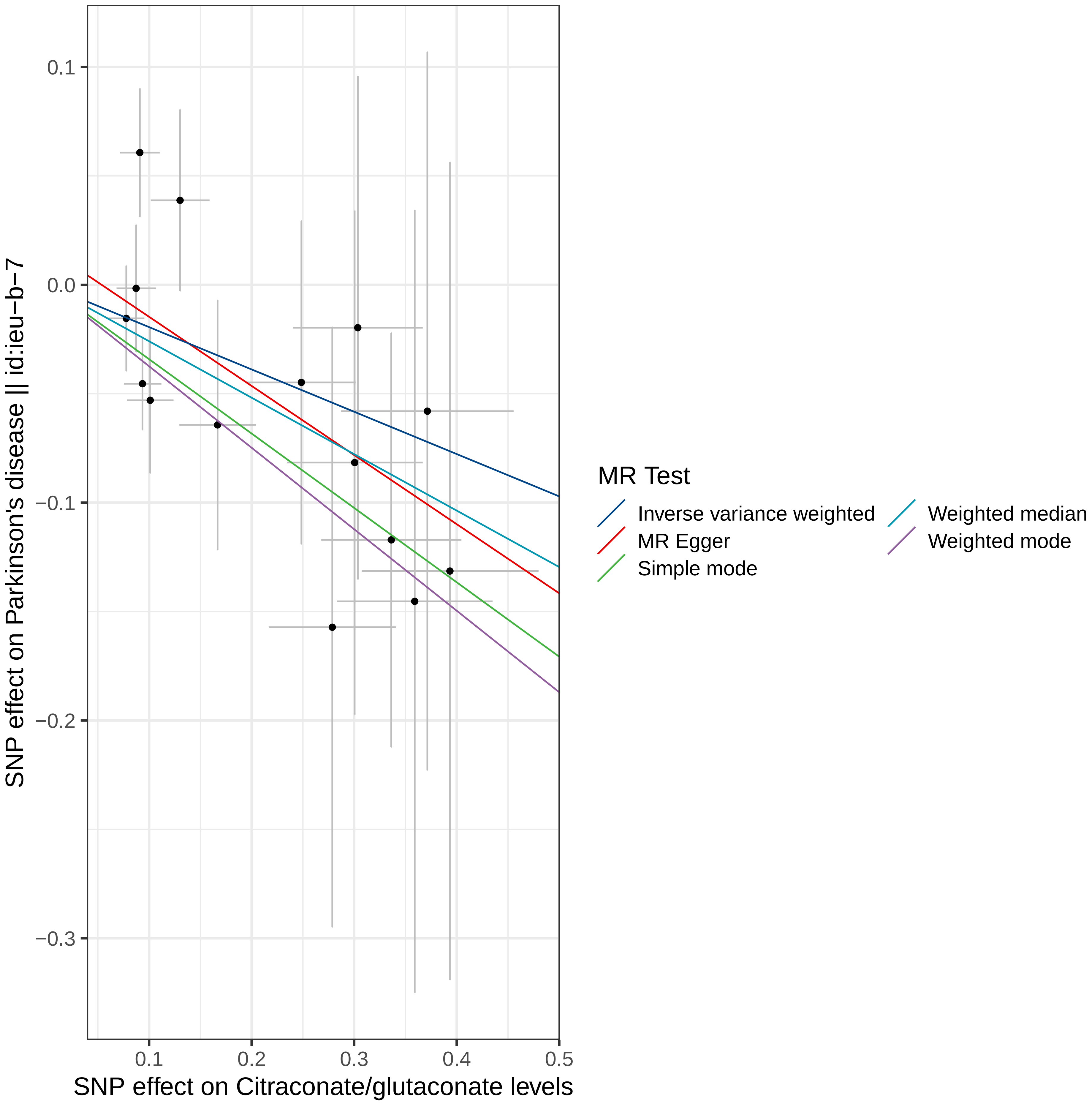

0.1
0.0
−0.1
−0.2
−0.3
0.1
0.2
0.3
0.4
0.5
SNP effect on Citraconate/glutaconate levels
SNP effect on Parkinson's disease || id:ieu−b−7
MR Test
Inverse variance weighted
MR Egger
Simple mode
Weighted median
Weighted mode

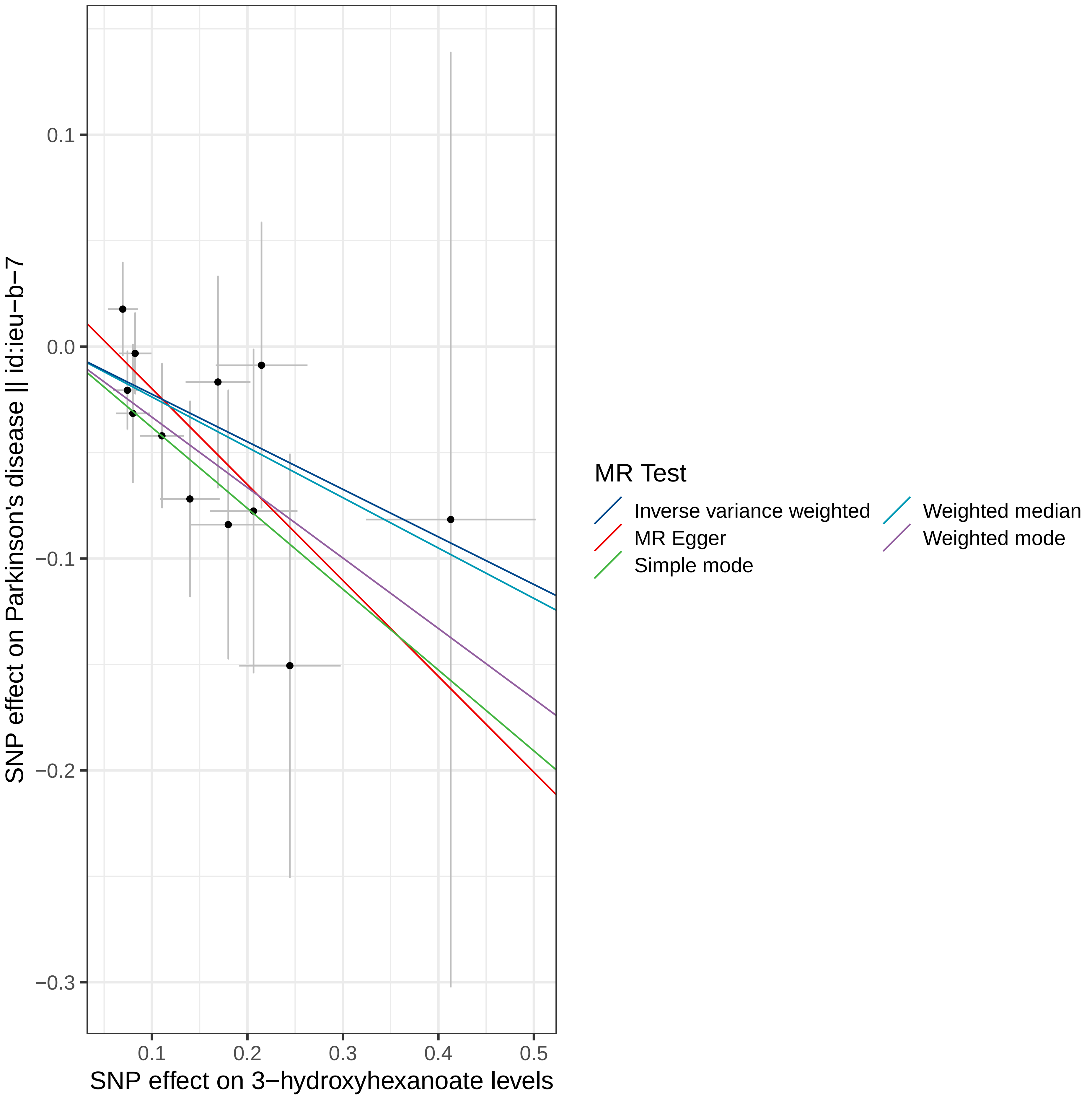

MR Test
Inverse variance weighted
MR Egger
Simple mode
Weighted median
Weighted mode
SNP effect on Parkinson's disease || id:ieu−b−7
SNP effect on 3−hydroxyhexanoate levels
0.1
0.0
−0.1
−0.2
−0.3
0.1
0.2
0.3
0.4
0.5

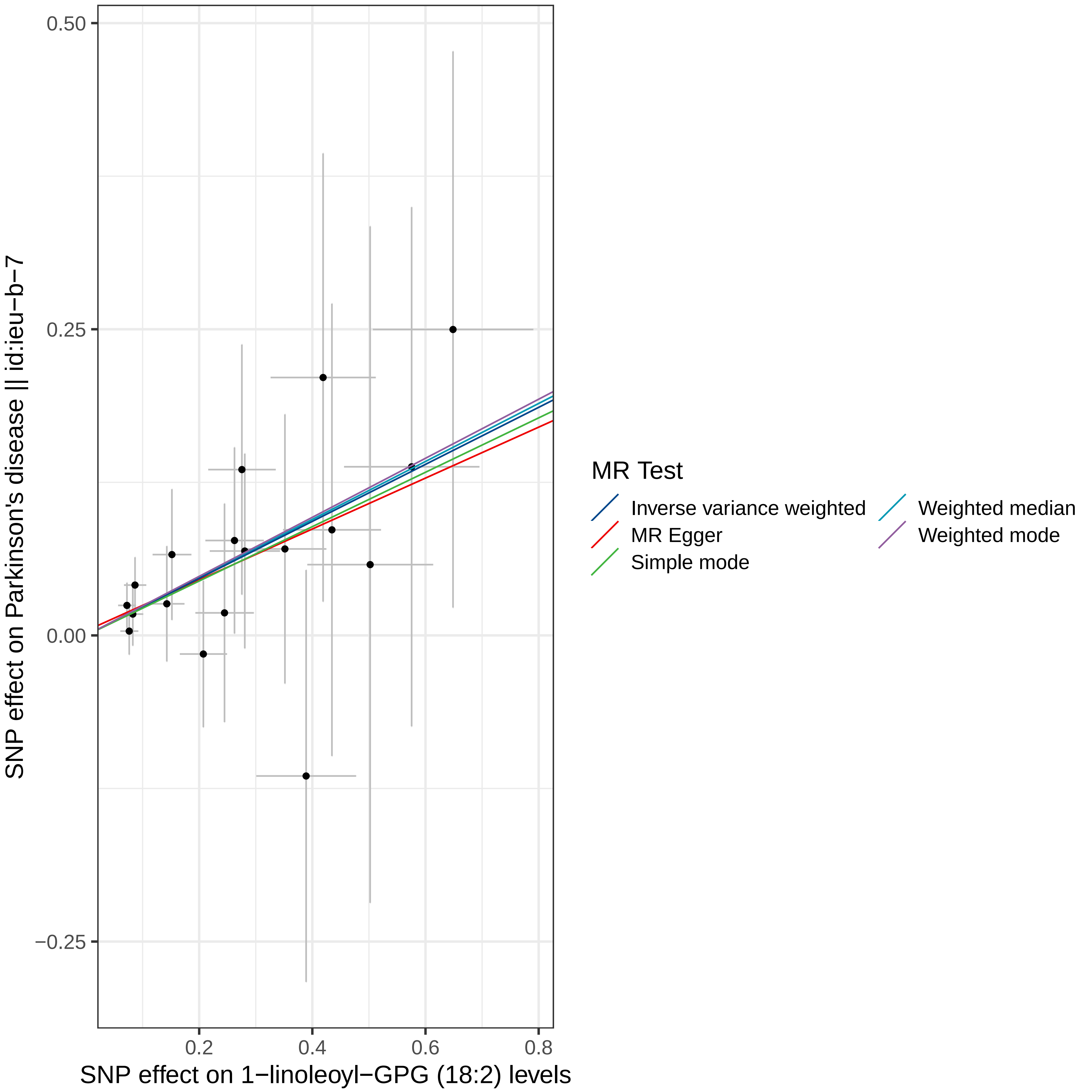

0.50
0.25
0.00
−0.25
0.2
0.4
0.6
0.8
SNP effect on Parkinson's disease || id:ieu−b−7
SNP effect on 1−linoleoyl−GPG (18:2) levels
MR Test
Inverse variance weighted
MR Egger
Simple mode
Weighted median
Weighted mode

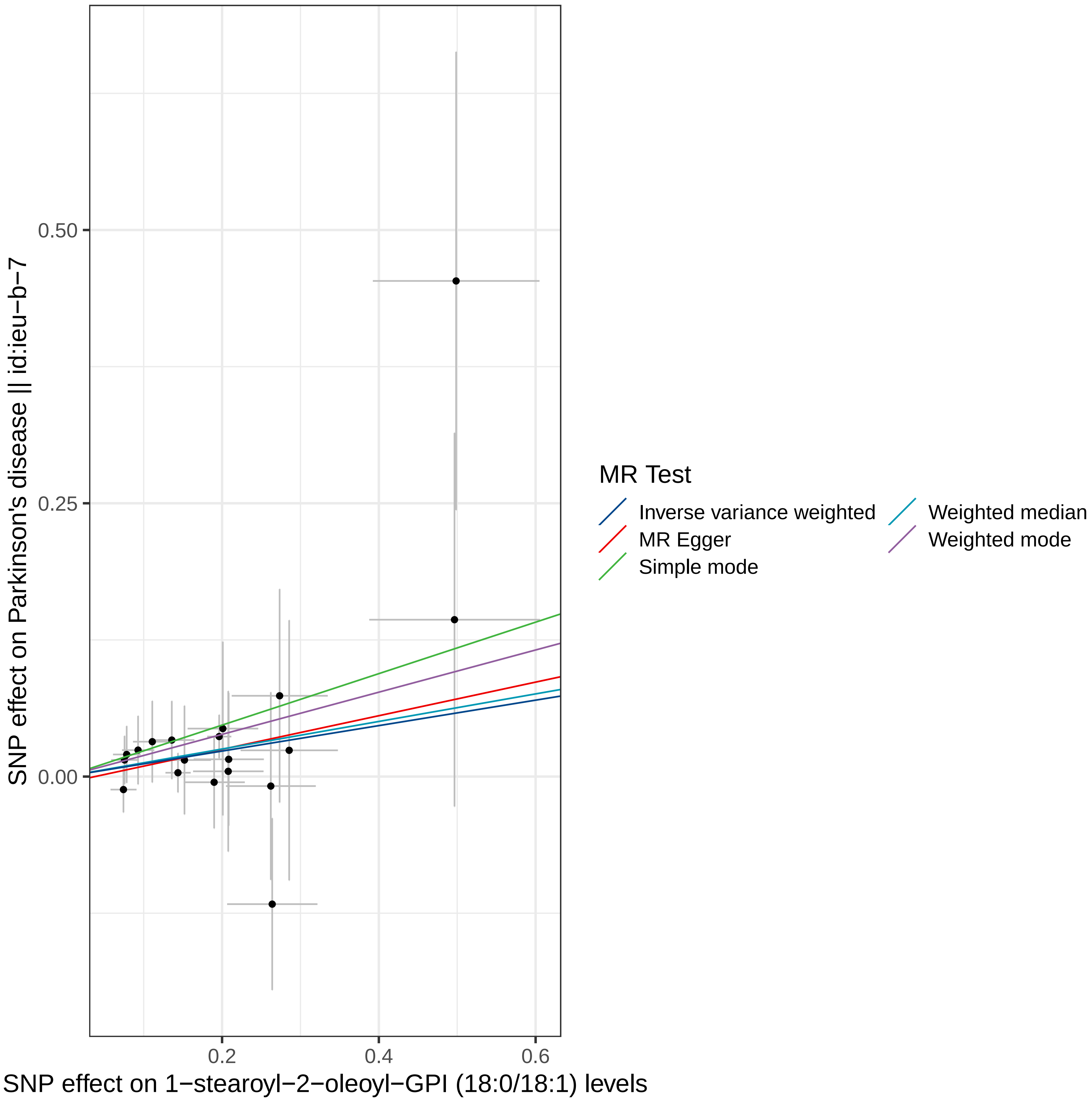

0.50
0.25
0.00
0.2
0.4
0.6
SNP effect on Parkinson's disease || id:ieu−b−7
SNP effect on 1−stearoyl−2−oleoyl−GPI (18:0/18:1) levels
MR Test
Inverse variance weighted
MR Egger
Simple mode
Weighted median
Weighted mode

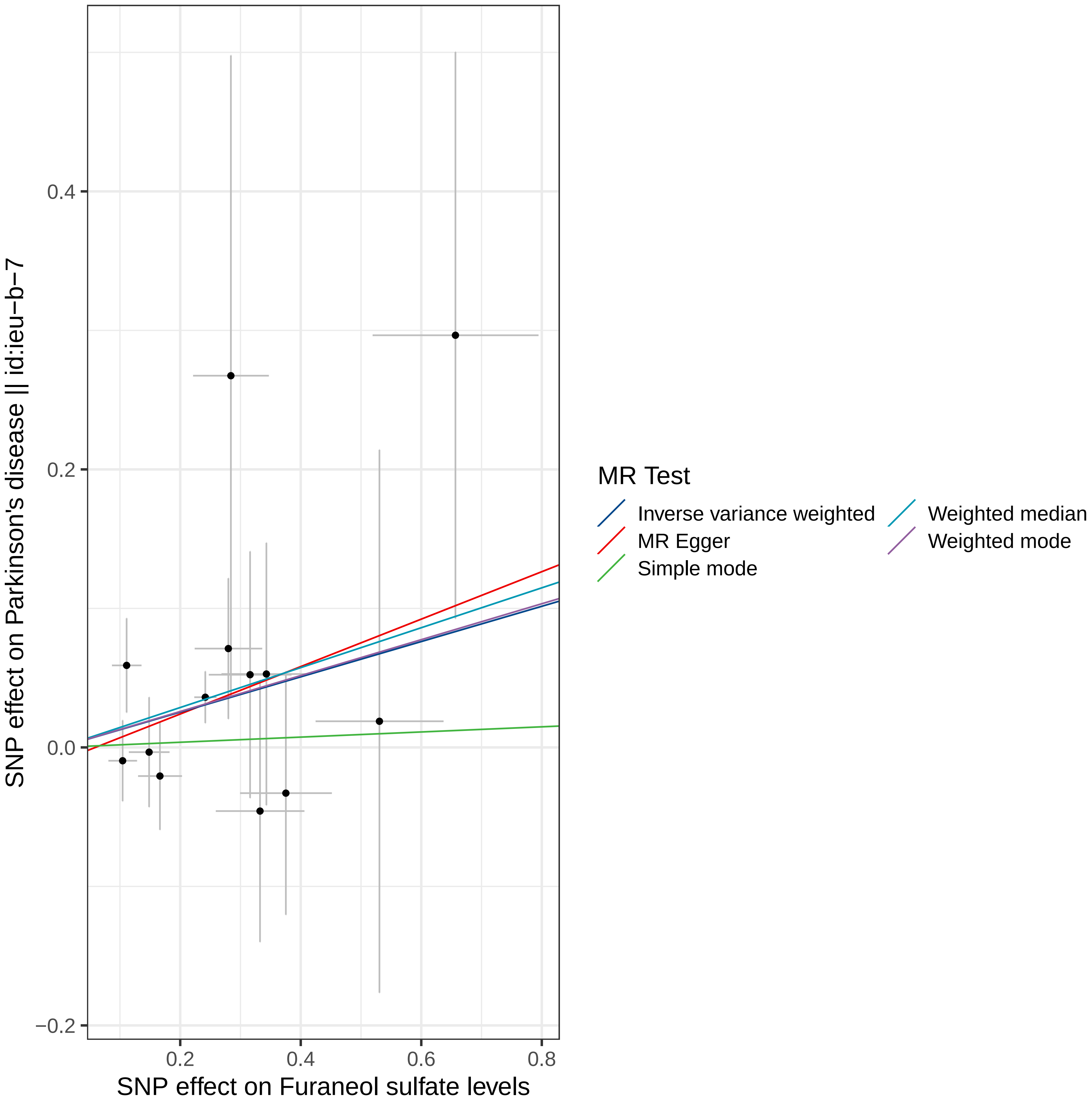
0.4
0.2
0.0
−0.2
0.2
0.4
0.6
0.8
SNP effect on Parkinson's disease || id:ieu−b−7
SNP effect on Furaneol sulfate levels
MR Test
Inverse variance weighted
MR Egger
Simple mode
Weighted median
Weighted mode

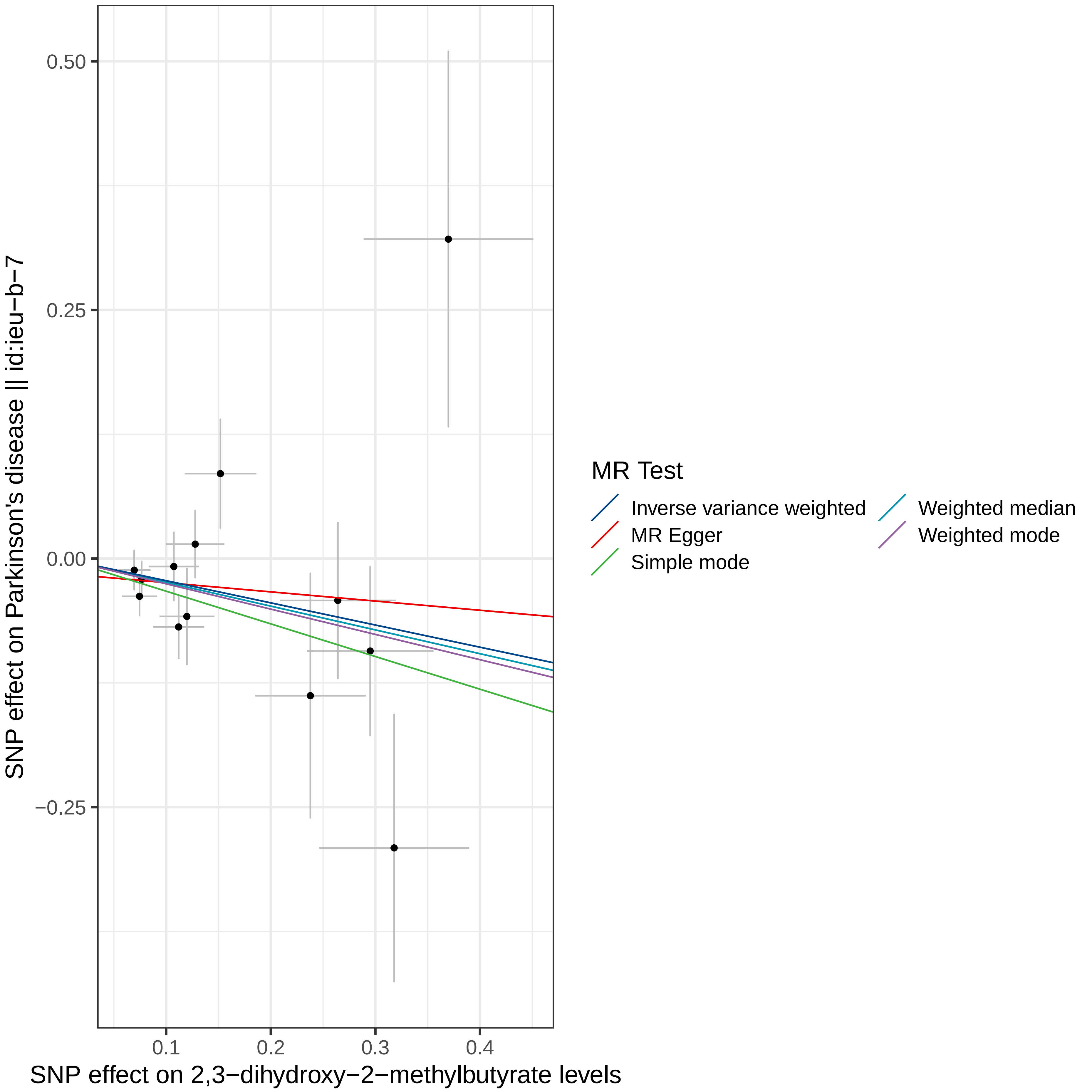

0.50
0.25
0.00
−0.25
0.1
0.2
0.3
0.4
SNP effect on Parkinson's disease || id:ieu−b−7
SNP effect on 2,3−dihydroxy−2−methylbutyrate levels
MR Test
Inverse variance weighted
MR Egger
Simple mode
Weighted median
Weighted mode

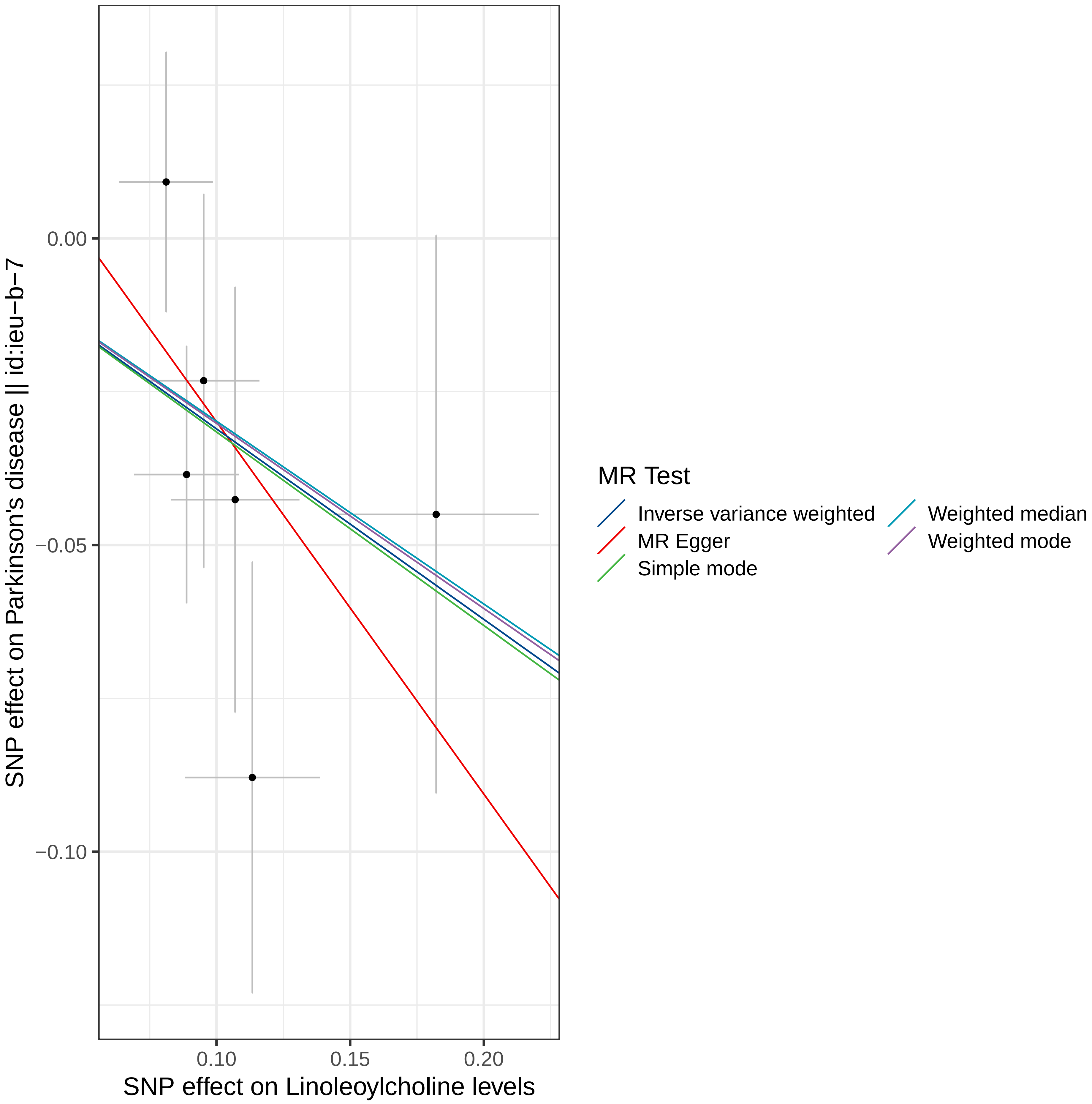

0.00
−0.05
−0.10
0.10
0.15
0.20
SNP effect on Linoleoylcholine levels
SNP effect on Parkinson's disease || id:ieu−b−7
MR Test
Inverse variance weighted
MR Egger
Simple mode
Weighted median
Weighted mode

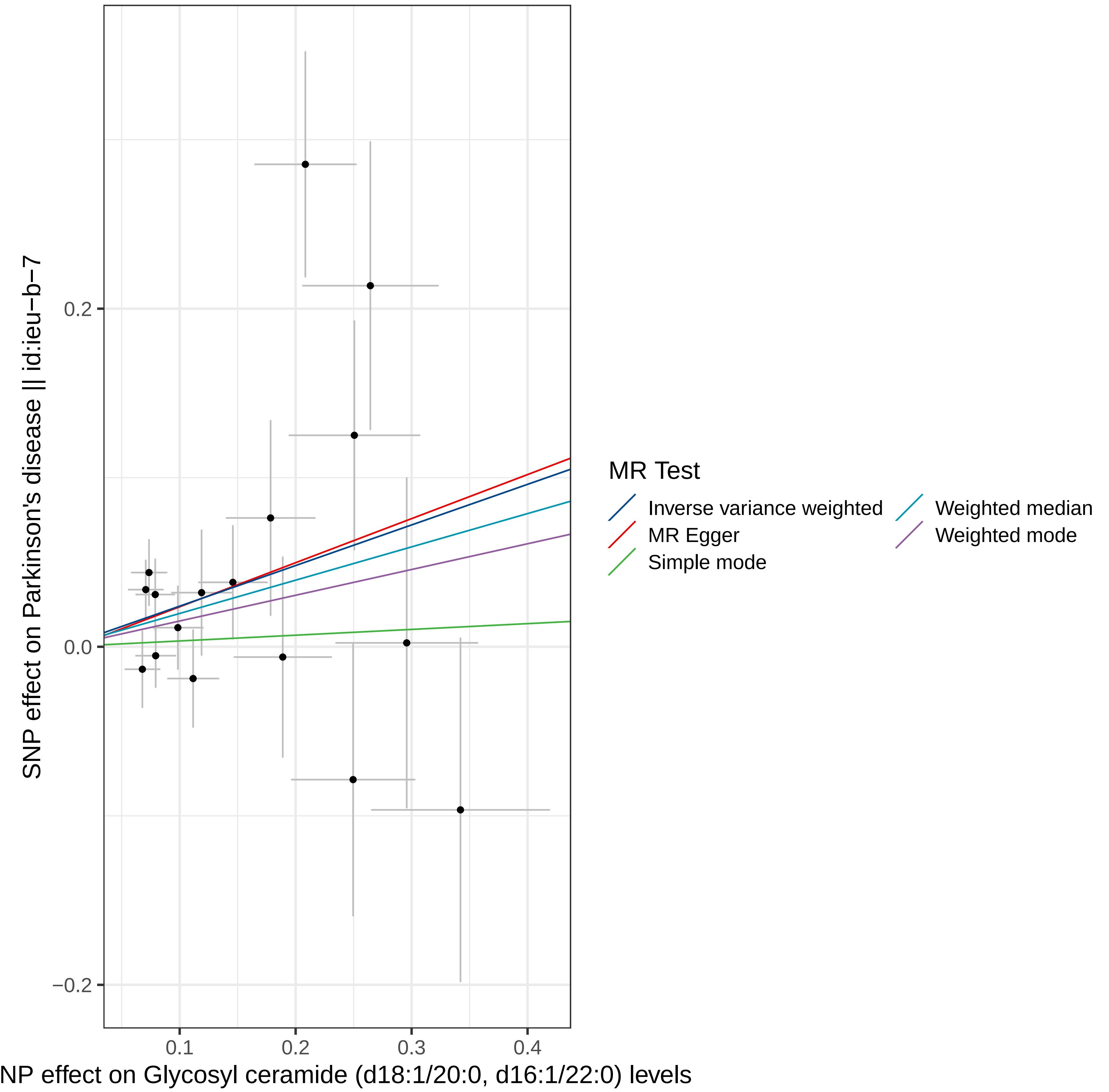

SNP effect on Parkinson's disease || id:ieu-b-7
0.2
0.0
−0.2
0.1
0.2
0.3
0.4
NP effect on Glycosyl ceramide (d18:1/20:0, d16:1/22:0) levels
MR Test
Inverse variance weighted
MR Egger
Simple mode
Weighted median
Weighted mode

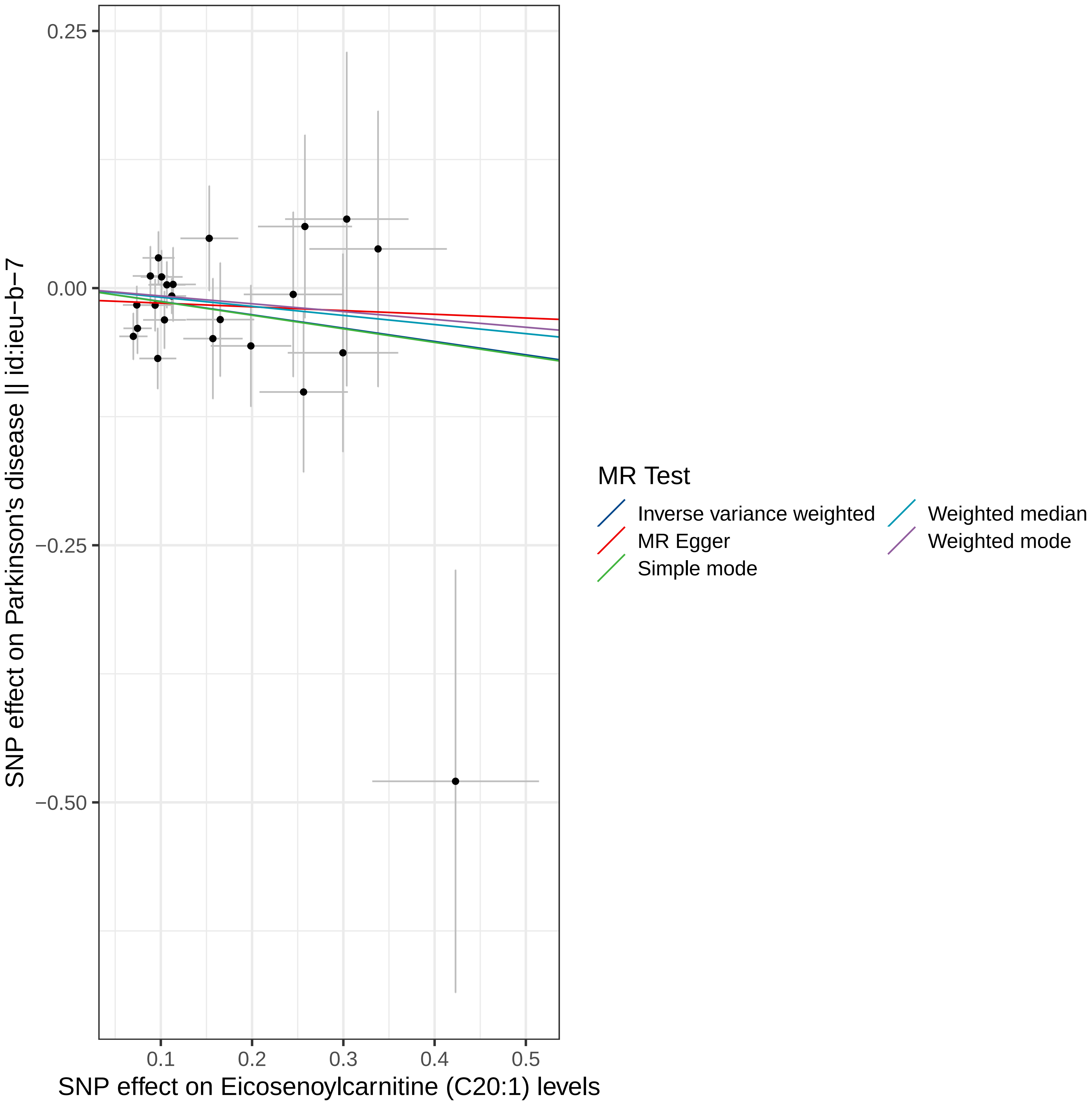

0.25
0.00
−0.25
−0.50
0.1
0.2
0.3
0.4
0.5
SNP effect on Eicosenoylcarnitine (C20:1) levels
SNP effect on Parkinson's disease || id:ieu−b−7
MR Test
Inverse variance weighted
MR Egger
Simple mode
Weighted median
Weighted mode

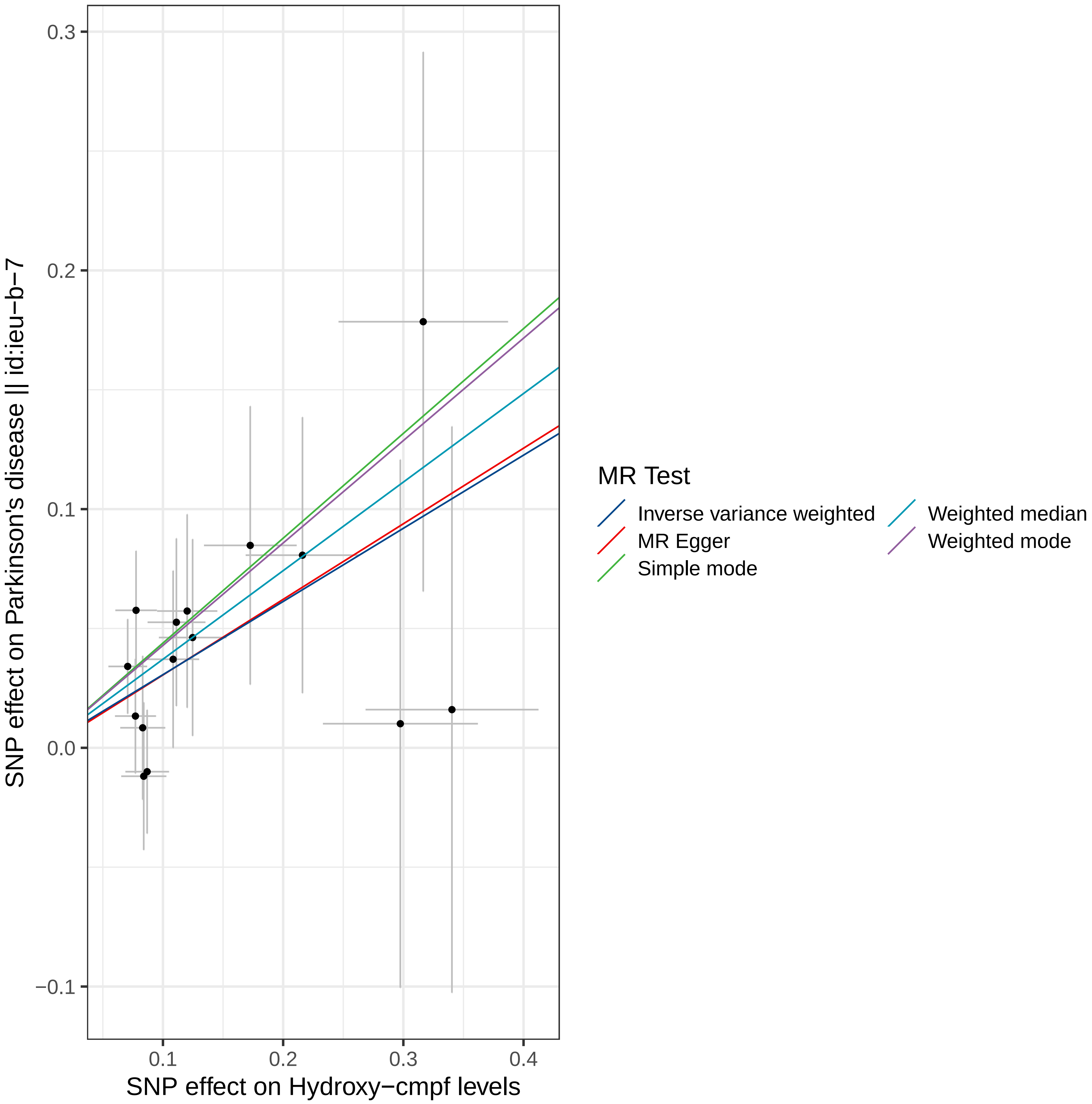

0.3
0.2
0.1
0.0
−0.1
0.1
0.2
0.3
0.4
SNP effect on Hydroxy−cmpf levels
SNP effect on Parkinson's disease || id:ieu−b−7
MR Test
Inverse variance weighted
MR Egger
Simple mode
Weighted median
Weighted mode

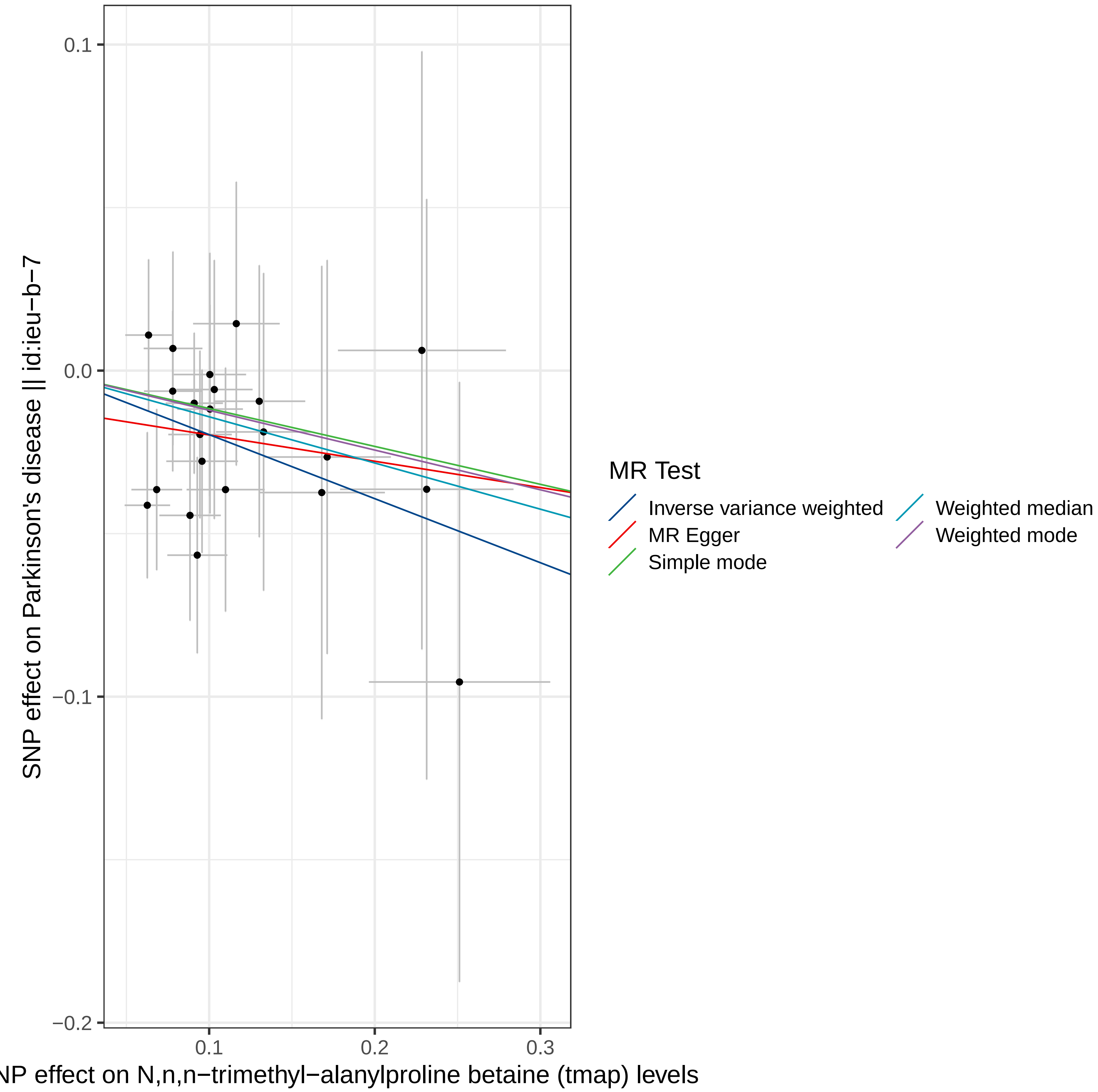

SNP effect on Parkinson's disease || id:ieu−b−7
0.1
0.0
−0.1
−0.2
0.1
0.2
0.3
NP effect on N,n,n−trimethyl−alanylproline betaine (tmap) levels
MR Test
Inverse variance weighted
MR Egger
Simple mode
Weighted median
Weighted mode

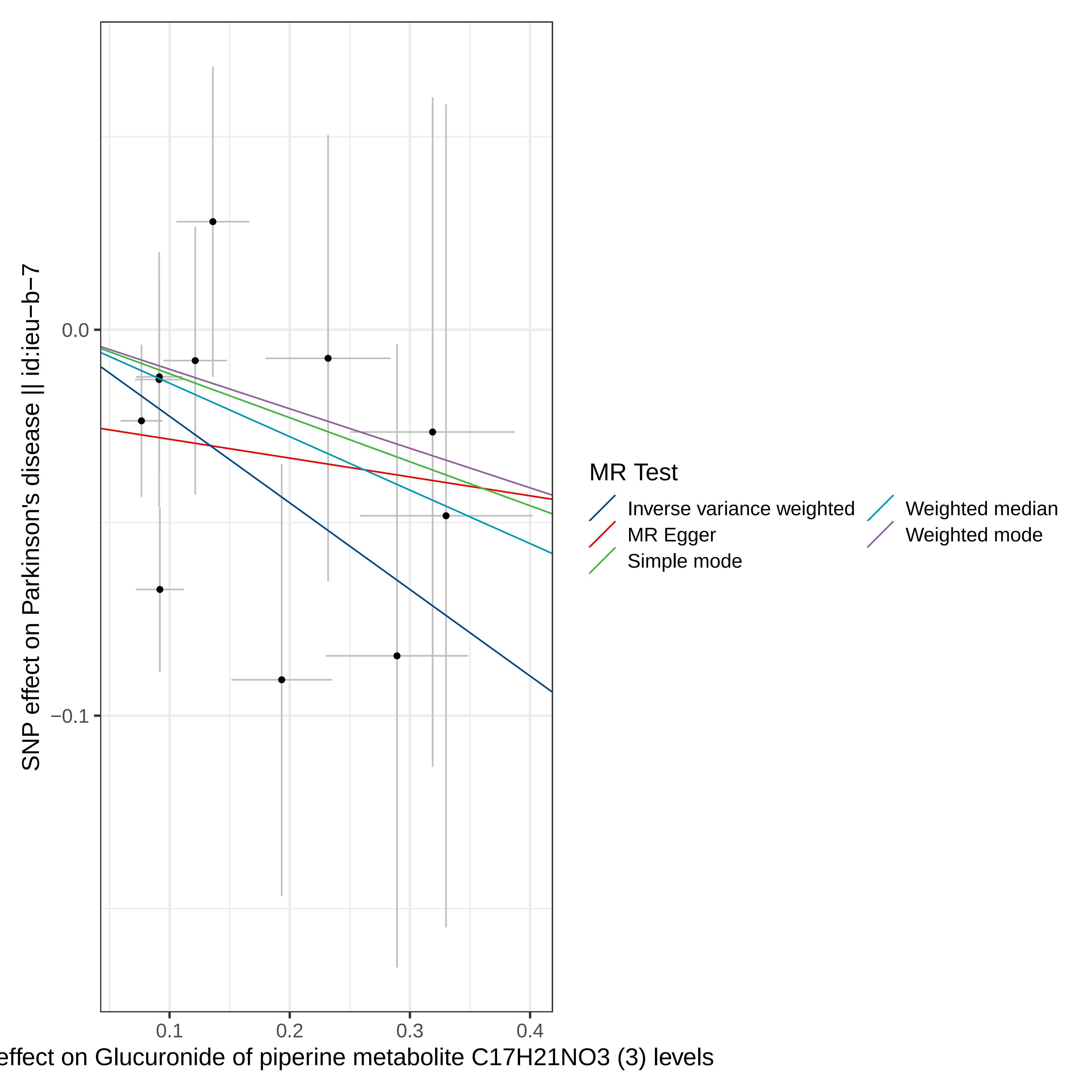

SNP effect on Parkinson's disease || id:ieu−b−7
0.0
−0.1
0.1
0.2
0.3
0.4
effect on Glucuronide of piperine metabolite C17H21NO3 (3) levels
MR Test
Inverse variance weighted
MR Egger
Simple mode
Weighted median
Weighted mode

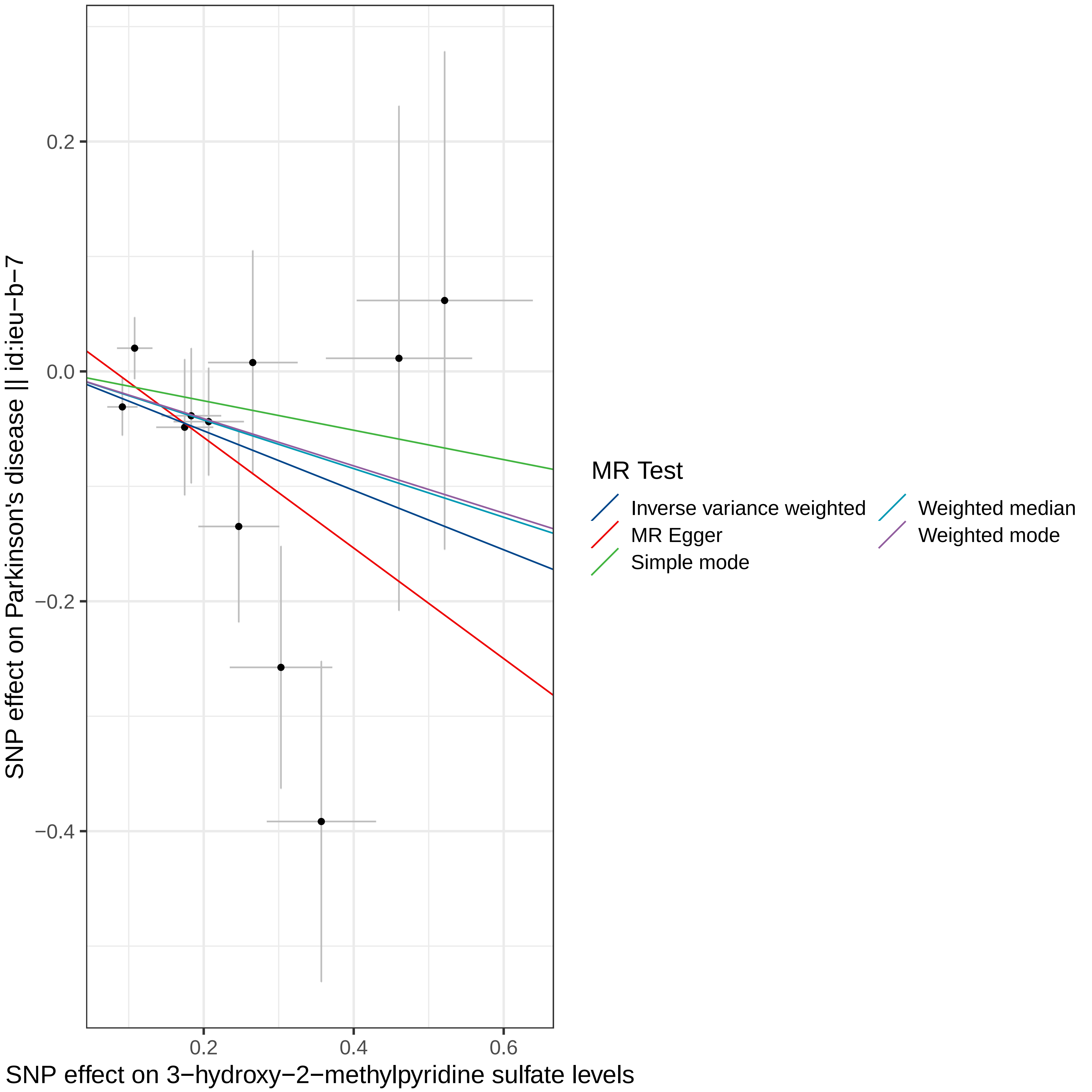

SNP effect on Parkinson's disease || id:ieu−b−7
0.2
0.0
−0.2
−0.4
0.2
0.4
0.6
SNP effect on 3−hydroxy−2−methylpyridine sulfate levels
MR Test
Inverse variance weighted
MR Egger
Simple mode
Weighted median
Weighted mode

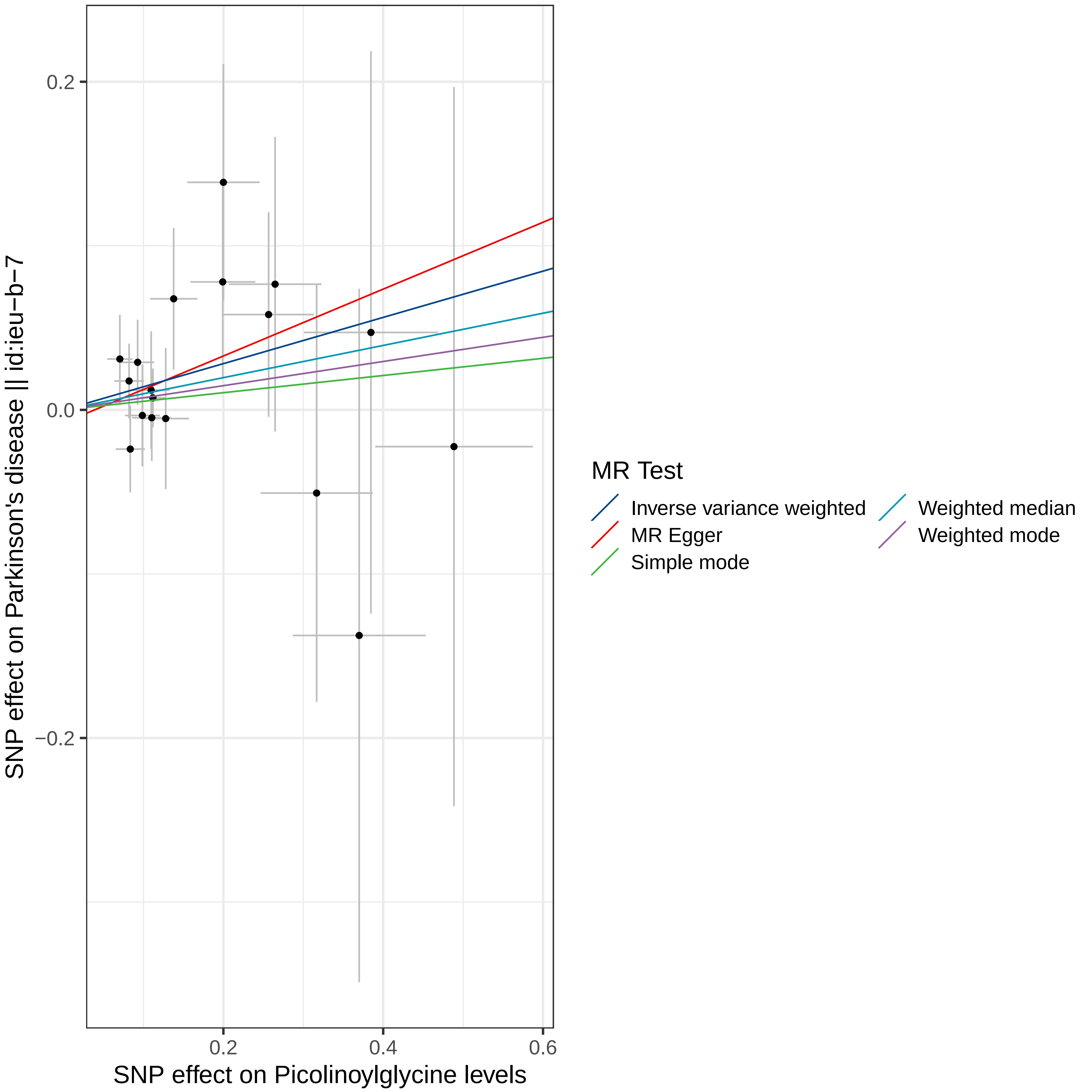

0.2
0.0
−0.2
0.2
0.4
0.6
SNP effect on Parkinson's disease || id:ieu−b−7
SNP effect on Picolinoylglycine levels
MR Test
Inverse variance weighted
MR Egger
Simple mode
Weighted median
Weighted mode

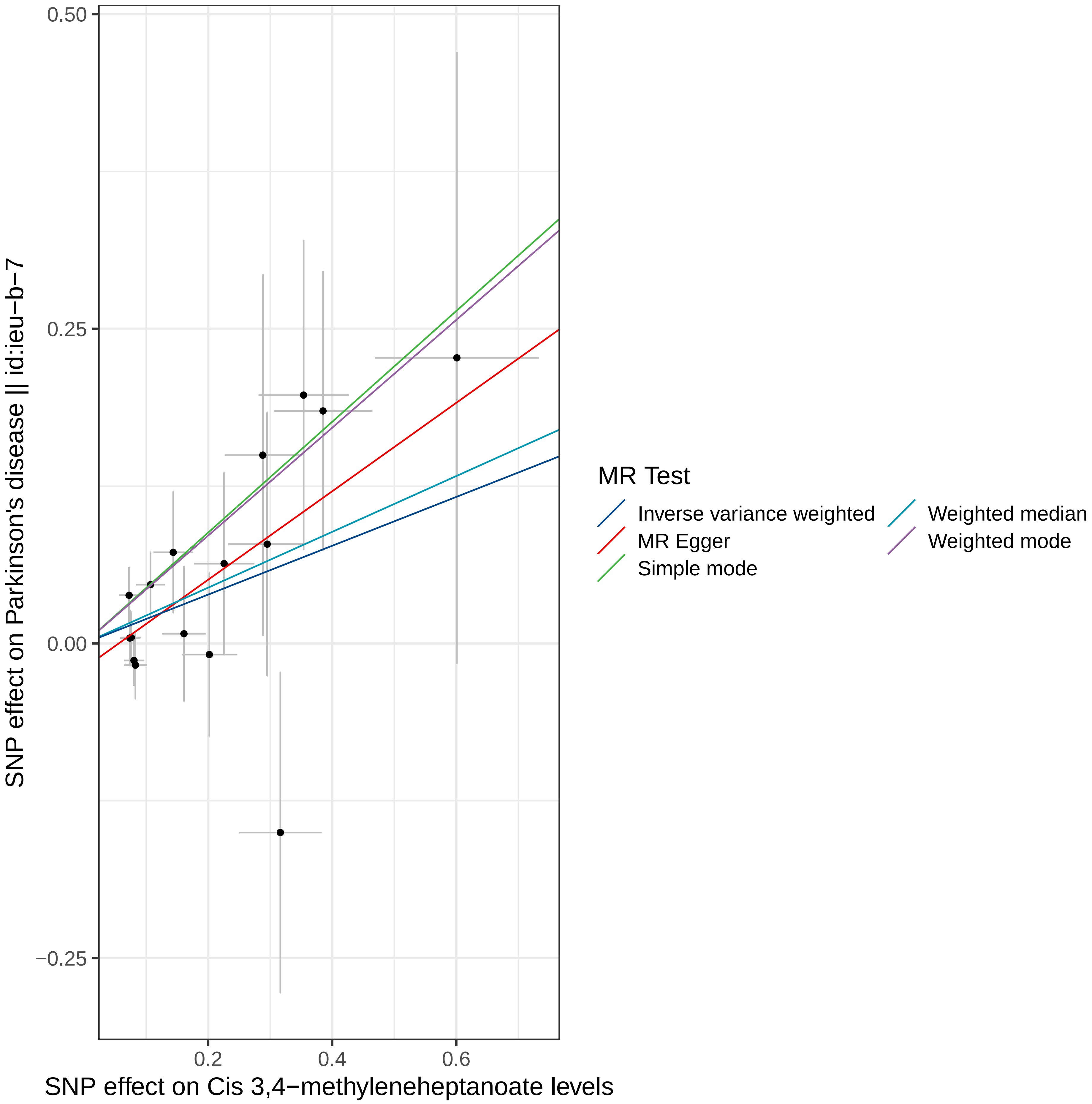

0.50
0.25
0.00
−0.25
0.2
0.4
0.6
SNP effect on Parkinson's disease || id:ieu−b−7
SNP effect on Cis 3,4−methyleneheptanoate levels
MR Test
Inverse variance weighted
MR Egger
Simple mode
Weighted median
Weighted mode

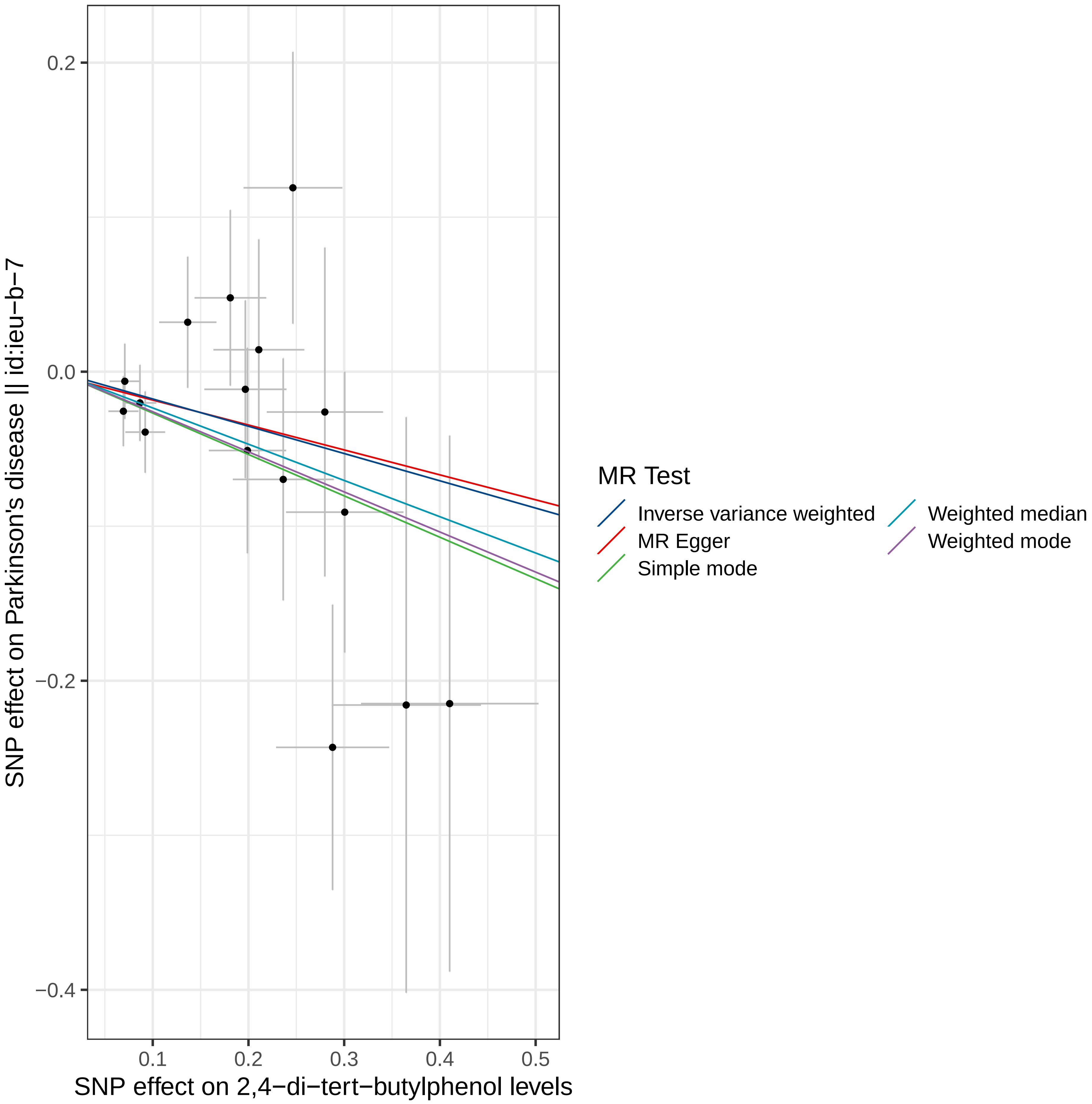

0.2
0.0
−0.2
−0.4
0.1
0.2
0.3
0.4
0.5
SNP effect on Parkinson's disease || id:ieu−b−7
SNP effect on 2,4−di−tert−butylphenol levels
MR Test
Inverse variance weighted
MR Egger
Simple mode
Weighted median
Weighted mode

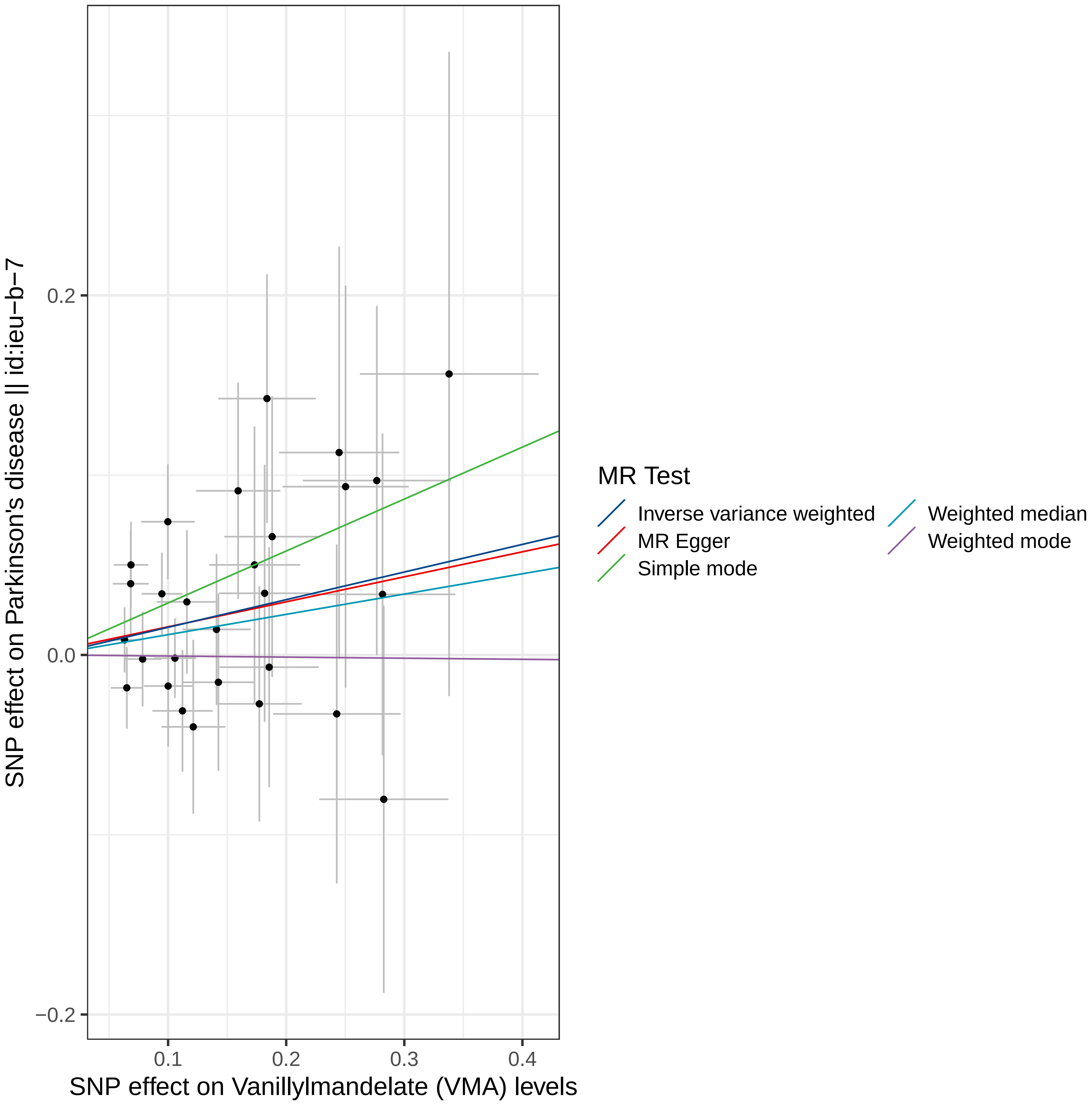

SNP effect on Parkinson's disease || id:ieu−b−7
0.2
0.0
−0.2
0.1
0.2
0.3
0.4
SNP effect on Vanillylmandelate (VMA) levels
MR Test
Inverse variance weighted
MR Egger
Simple mode
Weighted median
Weighted mode

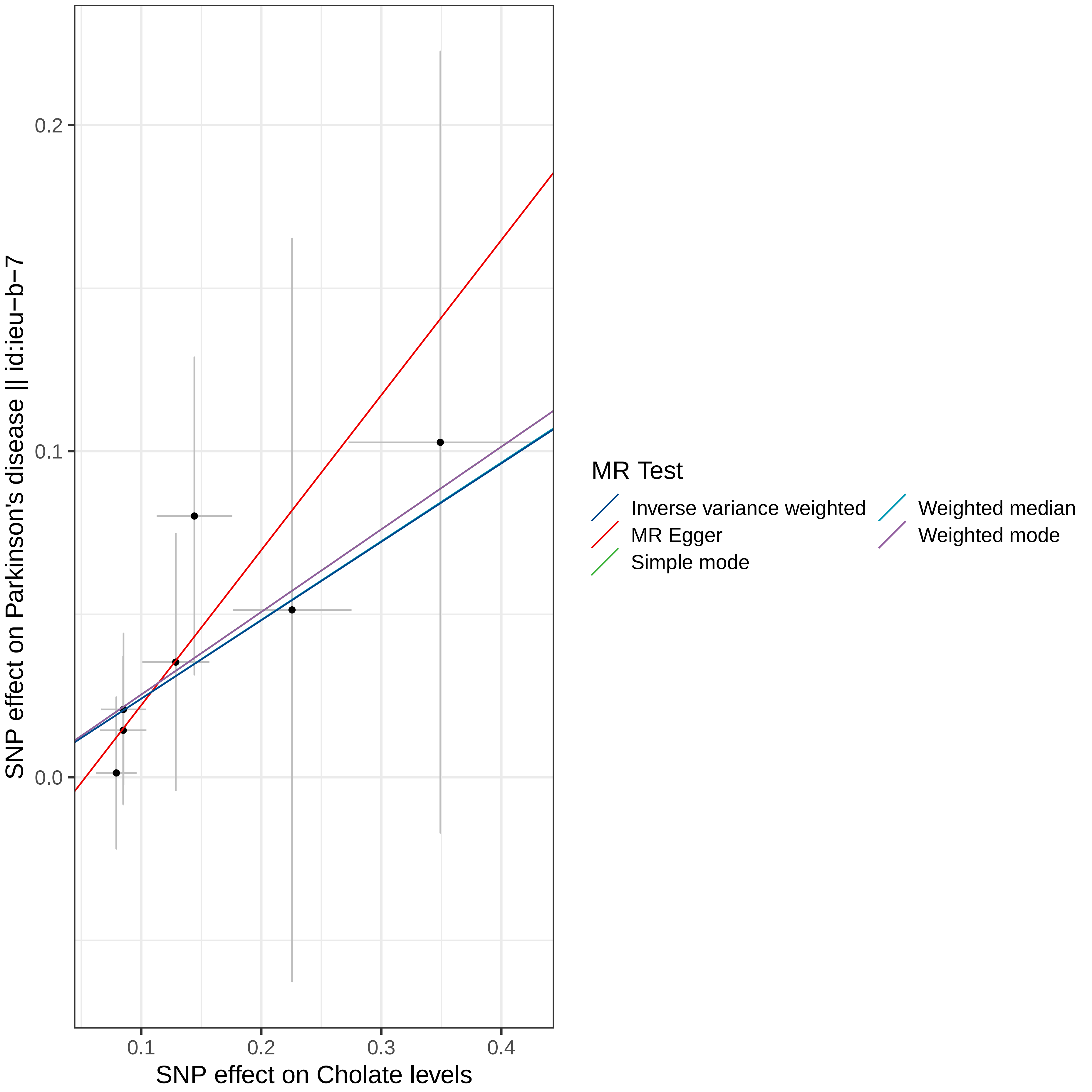

SNP effect on Parkinson's disease || id:ieu−b−7
SNP effect on Cholate levels
0.0
0.1
0.2
0.1
0.2
0.3
0.4
MR Test
Inverse variance weighted
MR Egger
Simple mode
Weighted median
Weighted mode

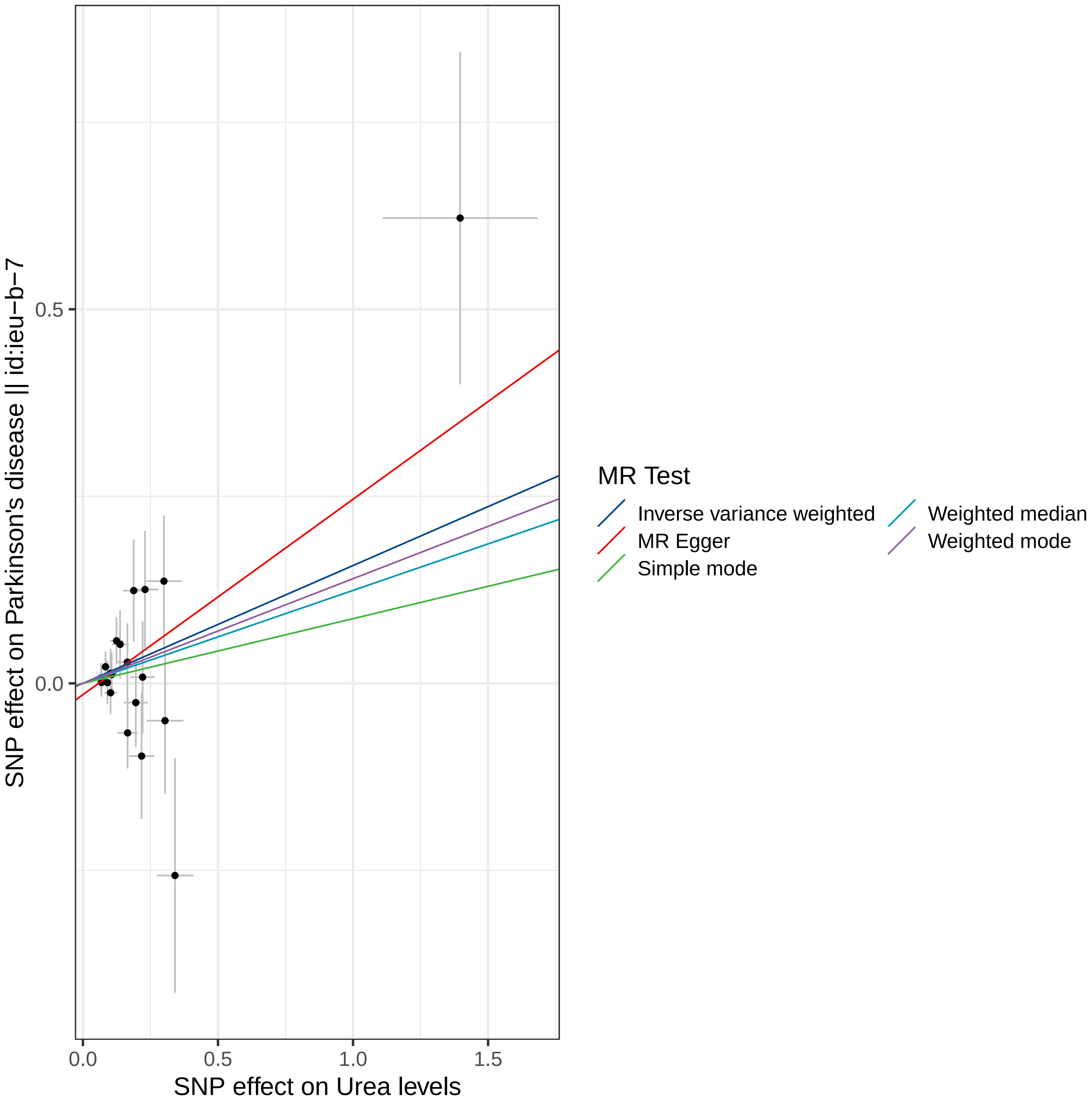

SNP effect on Parkinson's disease || id:ieu−b−7
0.5
0.0
0.0
0.5
1.0
1.5
SNP effect on Urea levels
MR Test
Inverse variance weighted
MR Egger
Simple mode
Weighted median
Weighted mode

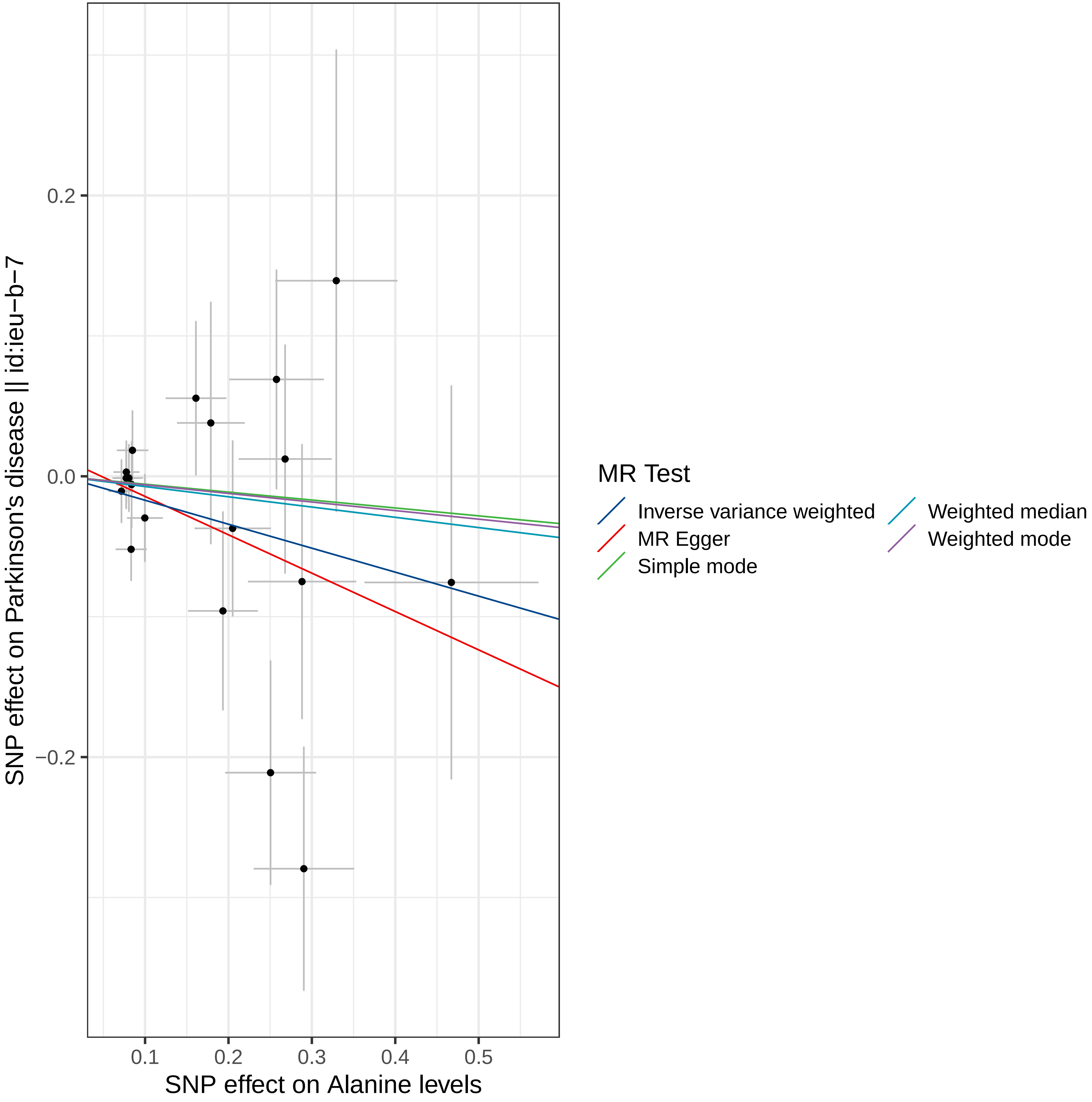
SNP effect on Parkinson's disease || id:ieu−b−7
0.2
0.0
−0.2
0.1
0.2
0.3
0.4
0.5
SNP effect on Alanine levels
MR Test
Inverse variance weighted
MR Egger
Simple mode
Weighted median
Weighted mode

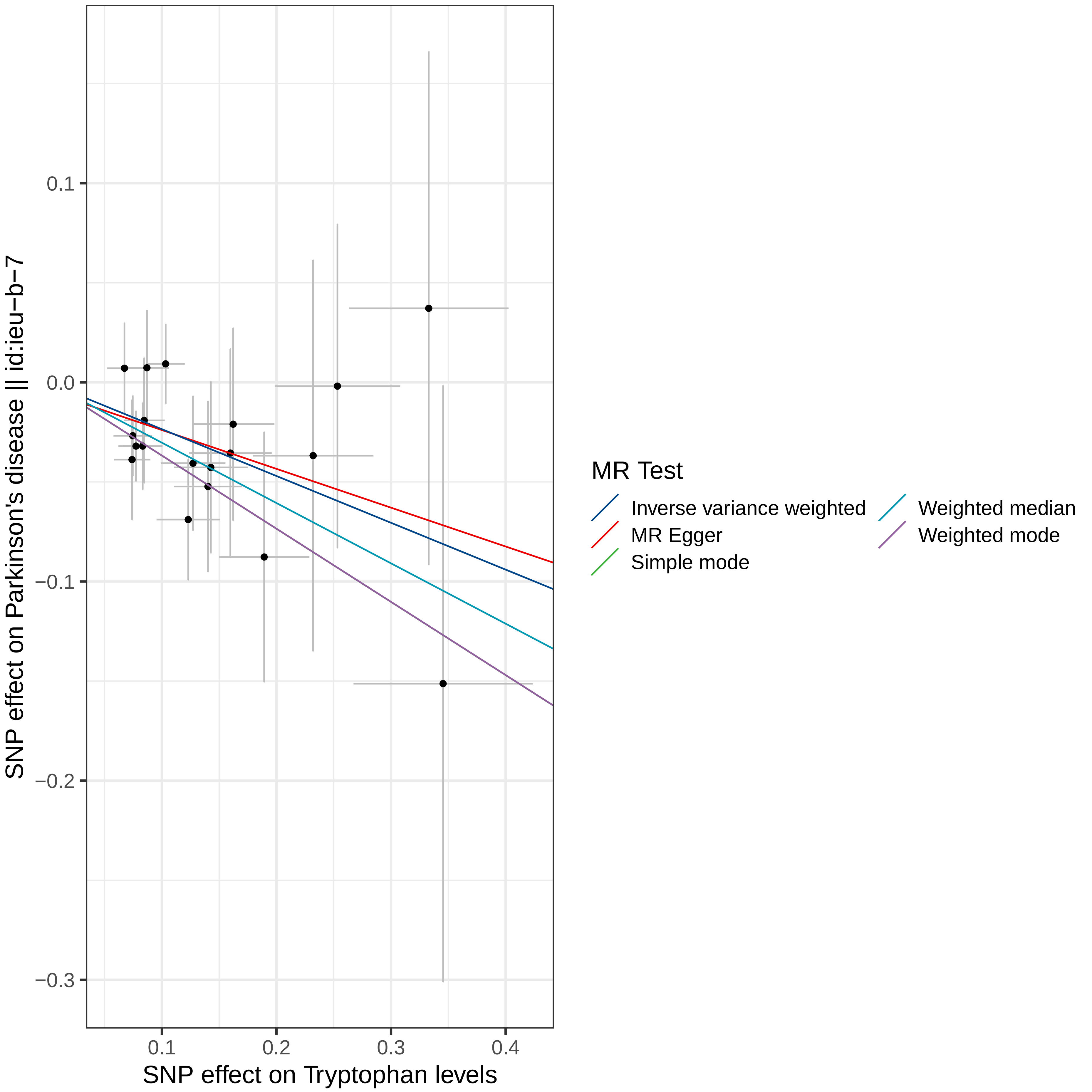

SNP effect on Parkinson's disease || id:ieu−b−7
SNP effect on Tryptophan levels
0.1
0.0
−0.1
−0.2
−0.3
0.1
0.2
0.3
0.4
MR Test
Inverse variance weighted
MR Egger
Simple mode
Weighted median
Weighted mode

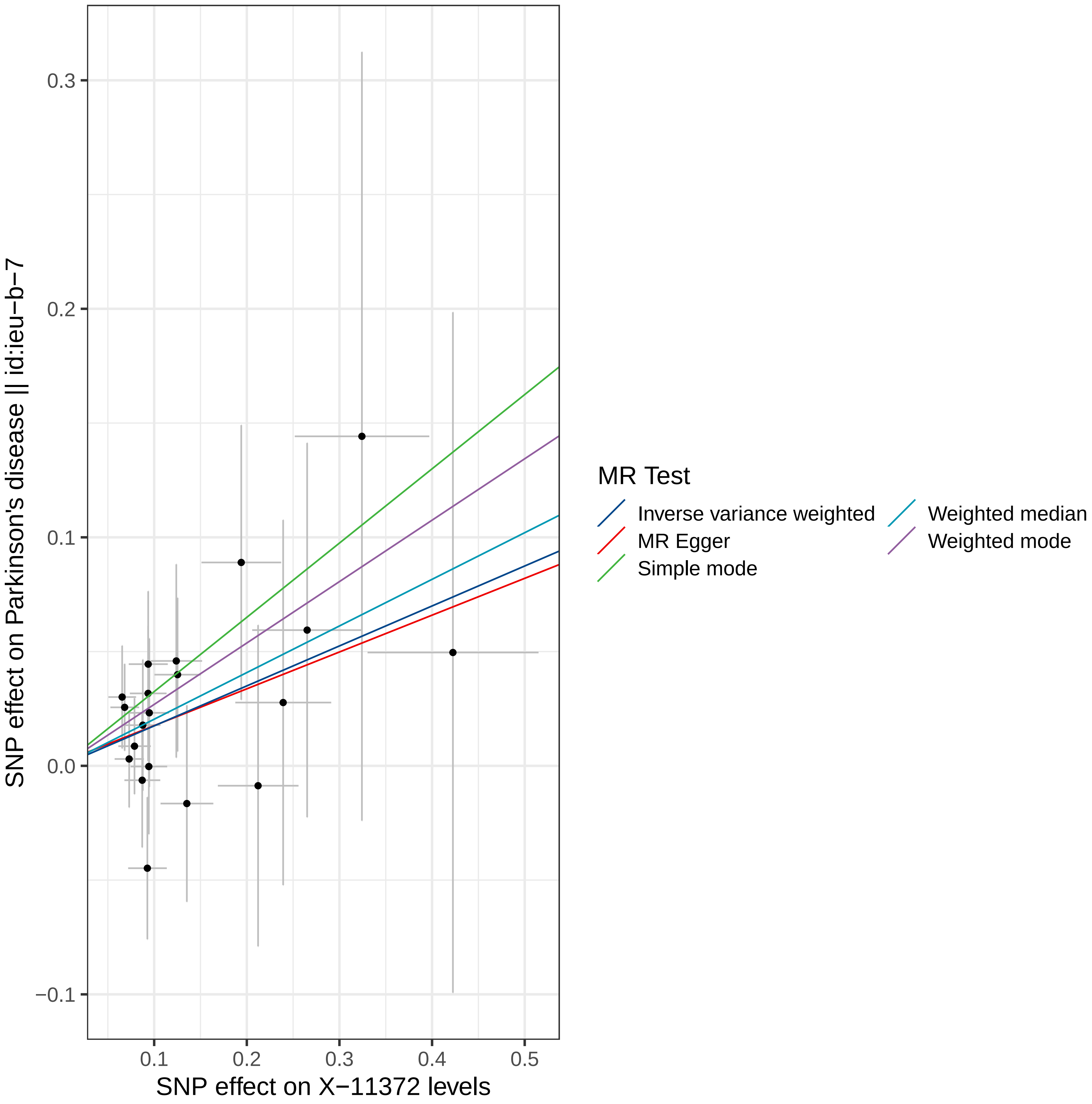

SNP effect on Parkinson's disease || id:ieu−b−7
SNP effect on X−11372 levels
0.3
0.2
0.1
0.0
−0.1
0.1
0.2
0.3
0.4
0.5
MR Test
Inverse variance weighted
MR Egger
Simple mode
Weighted median
Weighted mode

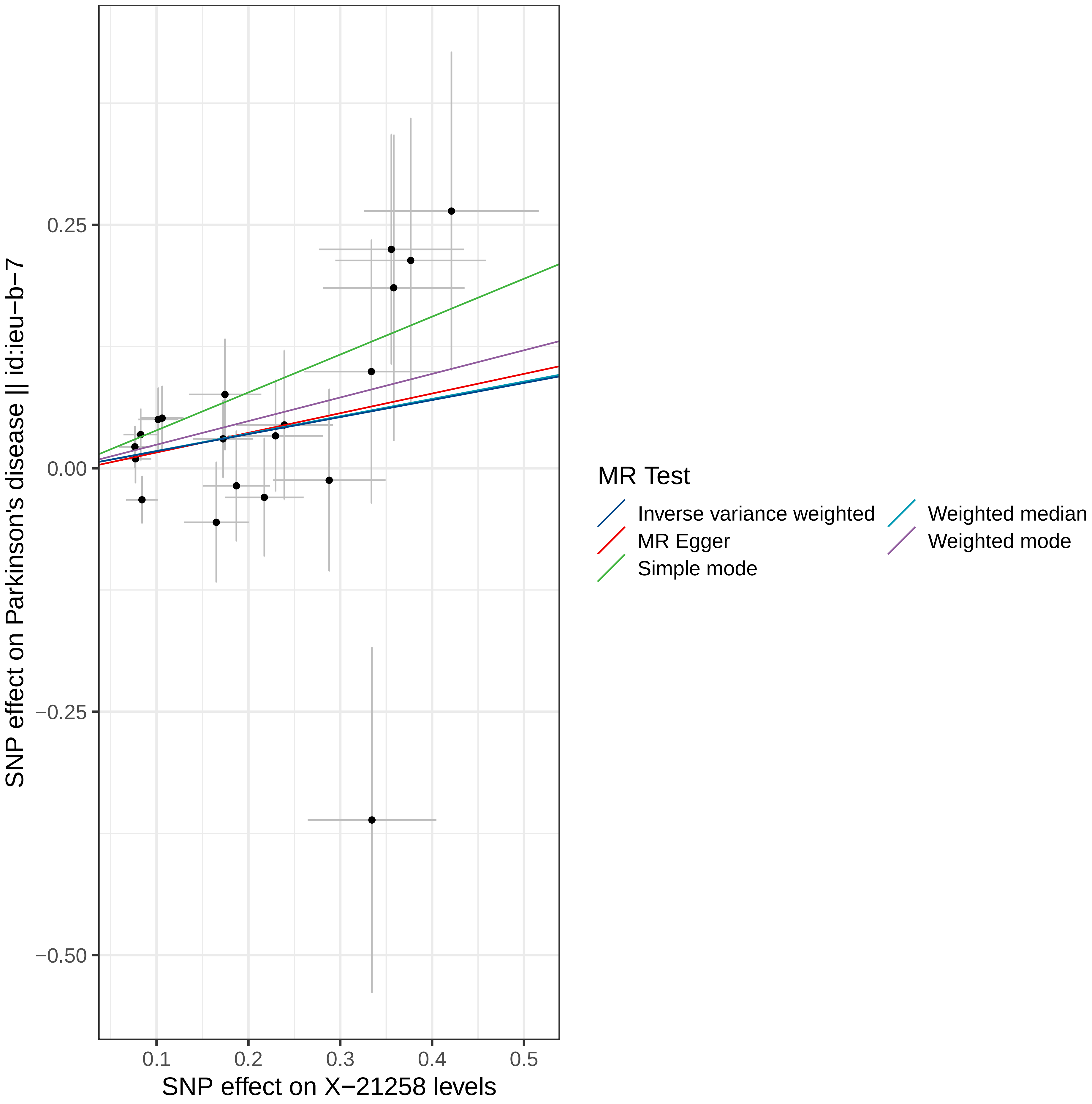

SNP effect on Parkinson's disease || id:ieu−b−7
SNP effect on X−21258 levels
0.25
0.00
−0.25
−0.50
0.1
0.2
0.3
0.4
0.5
MR Test
Inverse variance weighted
MR Egger
Simple mode
Weighted median
Weighted mode

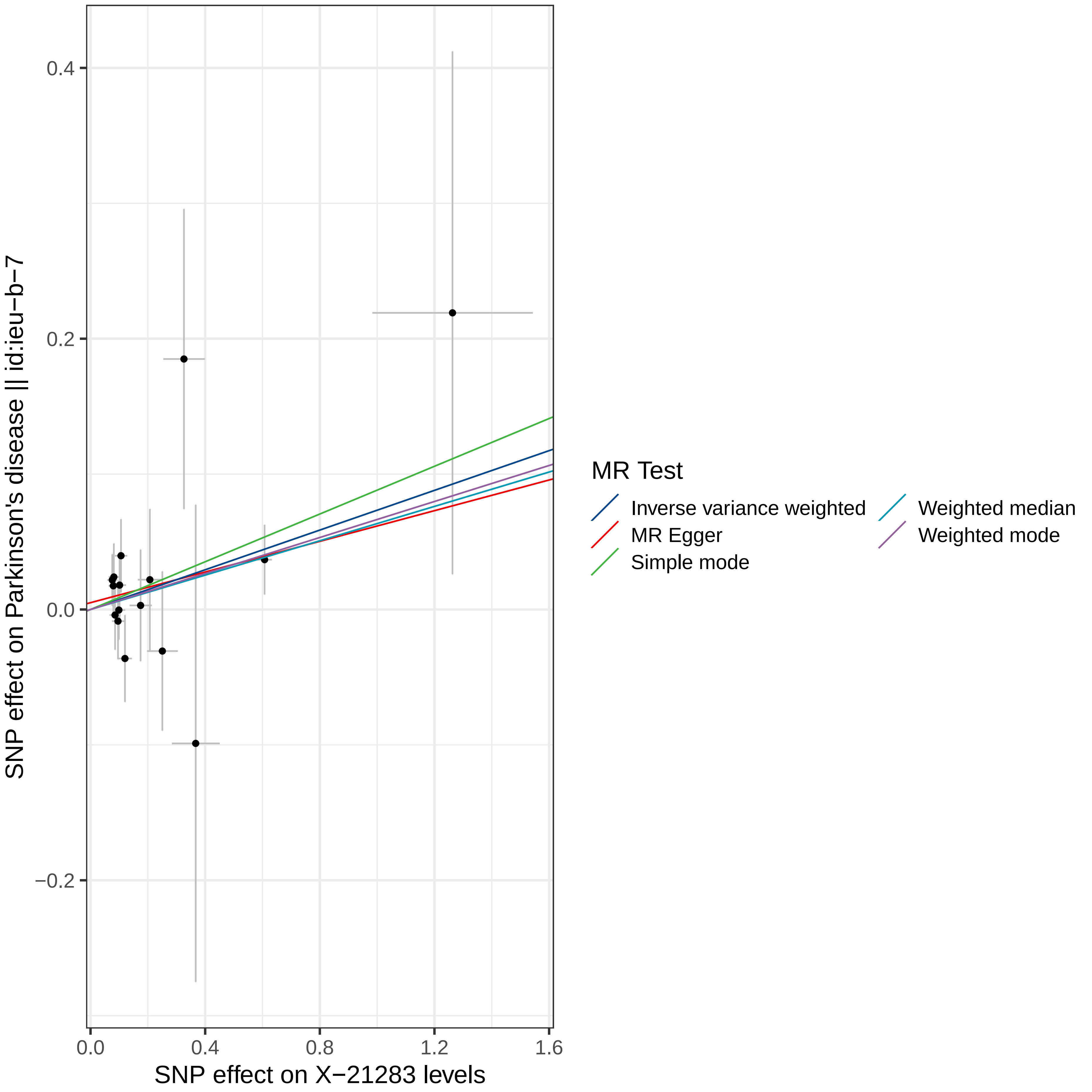

SNP effect on Parkinson's disease || id:ieu−b−7
SNP effect on X−21283 levels
0.4
0.2
0.0
−0.2
0.0
0.4
0.8
1.2
1.6
MR Test
Inverse variance weighted
MR Egger
Simple mode
Weighted median
Weighted mode

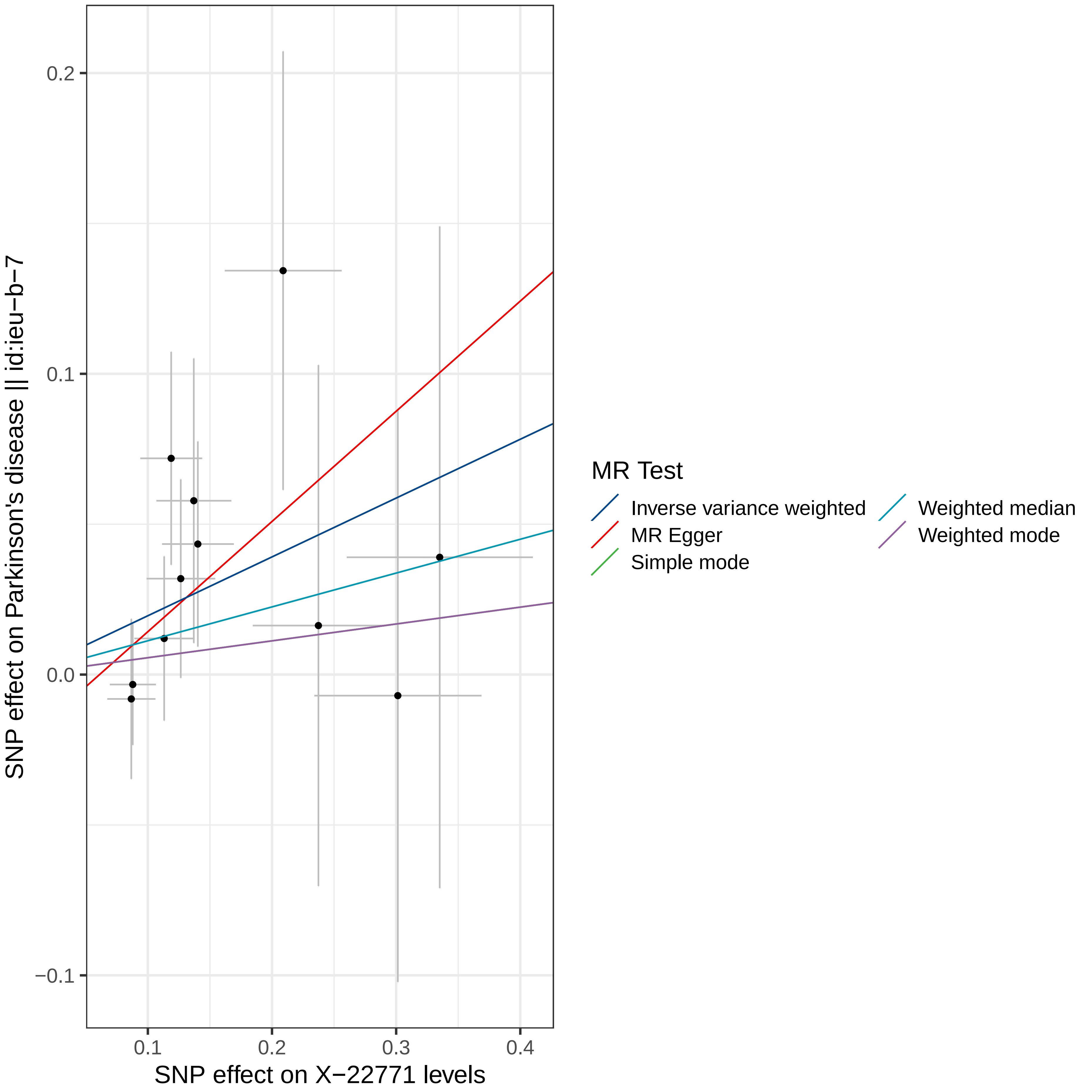

0.2
0.1
0.0
−0.1
0.1
0.2
0.3
0.4
SNP effect on X−22771 levels
SNP effect on Parkinson's disease || id:ieu−b−7
MR Test
Inverse variance weighted
MR Egger
Simple mode
Weighted median
Weighted mode

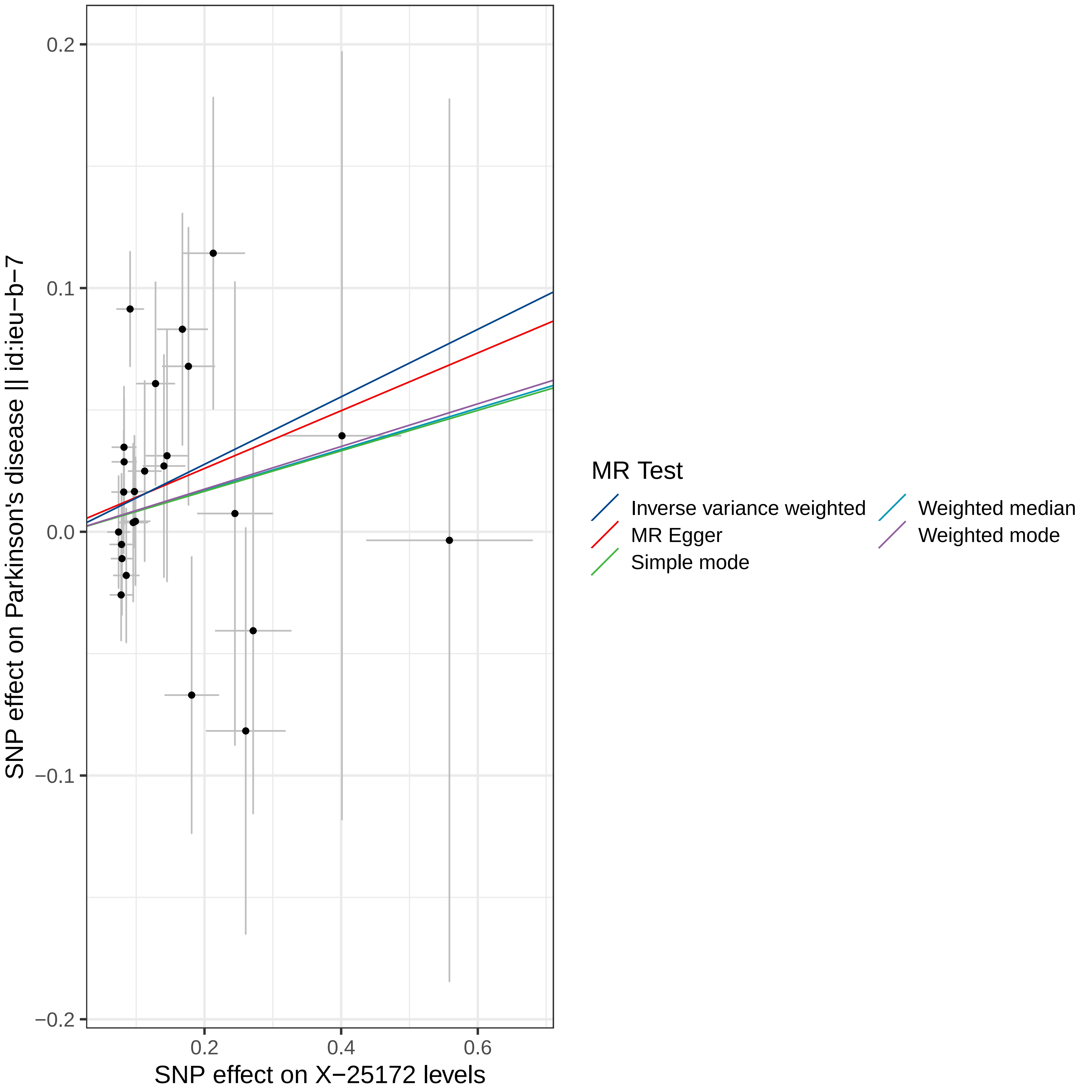

SNP effect on Parkinson's disease || id:ieu−b−7
0.2
0.1
0.0
−0.1
−0.2
0.2
0.4
0.6
SNP effect on X−25172 levels
MR Test
Inverse variance weighted
MR Egger
Simple mode
Weighted median
Weighted mode

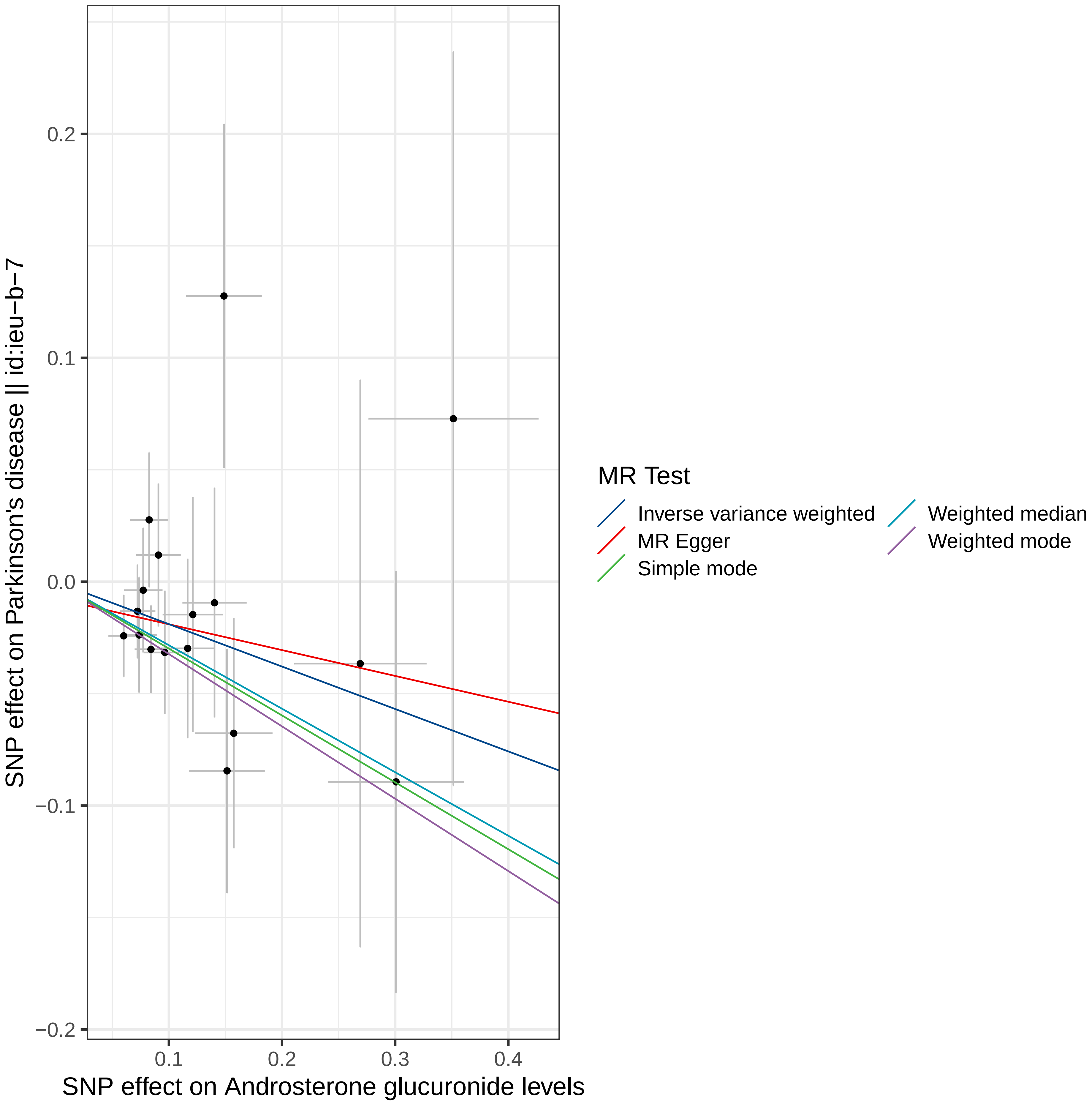

SNP effect on Parkinson's disease || id:ieu−b−7
0.2
0.1
0.0
−0.1
−0.2
0.1
0.2
0.3
0.4
SNP effect on Androsterone glucuronide levels
MR Test
Inverse variance weighted
MR Egger
Simple mode
Weighted median
Weighted mode

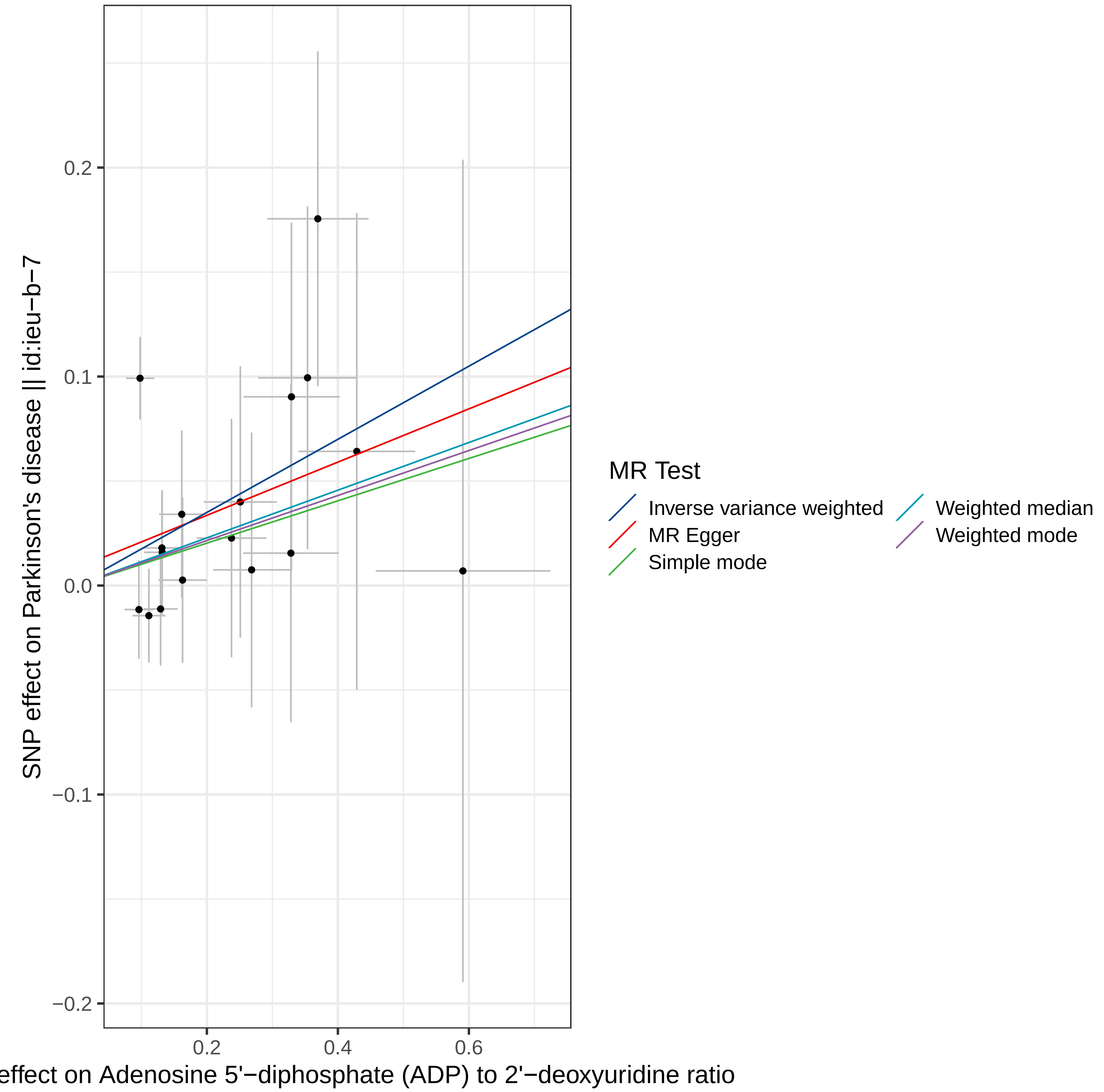

SNP effect on Parkinson's disease || id:ieu−b−7
0.2
0.1
0.0
−0.1
−0.2
0.2
0.4
0.6
effect on Adenosine 5'−diphosphate (ADP) to 2'−deoxyuridine ratio
MR Test
Inverse variance weighted
MR Egger
Simple mode
Weighted median
Weighted mode

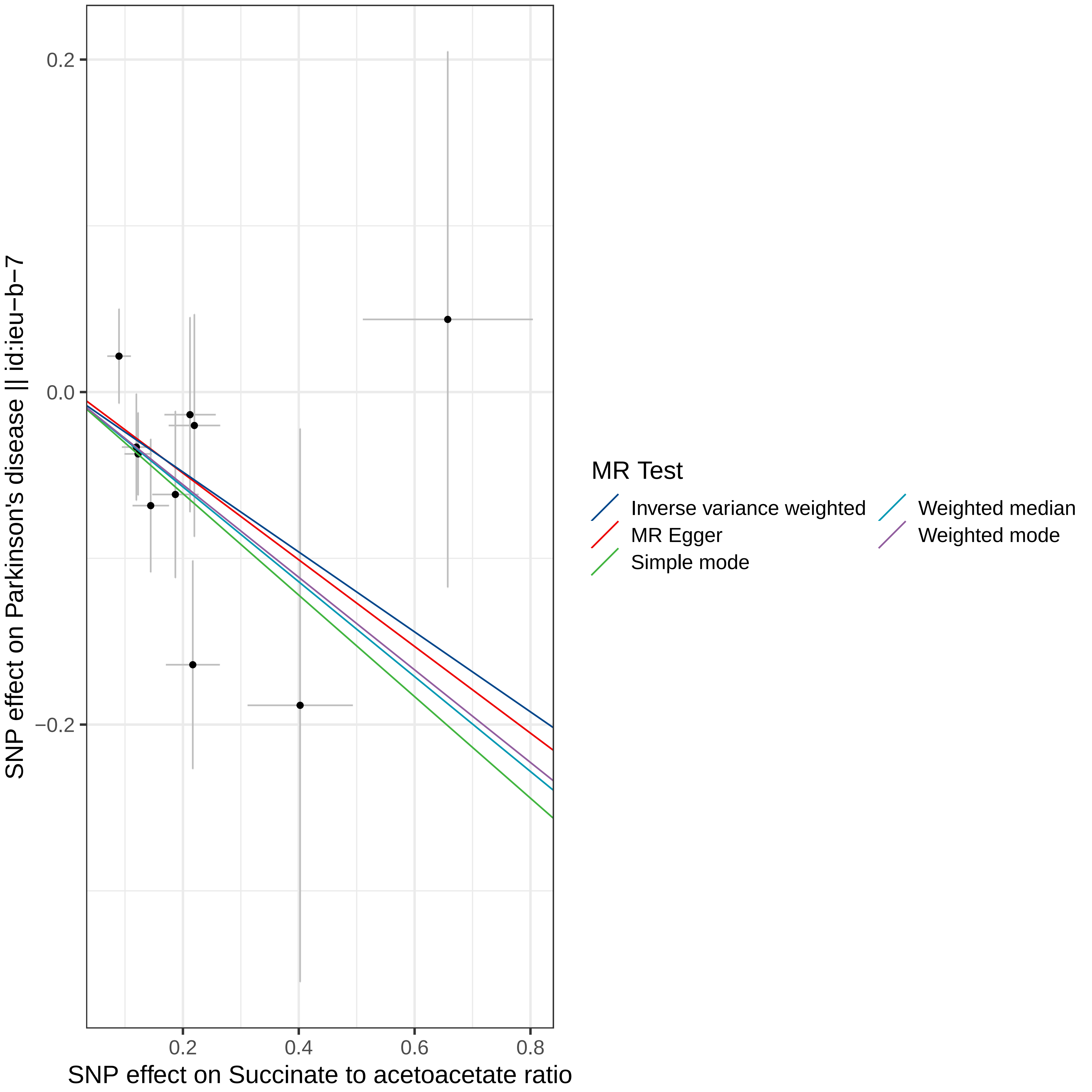

0.2
0.0
−0.2
0.2
0.4
0.6
0.8
SNP effect on Parkinson's disease || id:ieu−b−7
SNP effect on Succinate to acetoacetate ratio
MR Test
Inverse variance weighted
MR Egger
Simple mode
Weighted median
Weighted mode

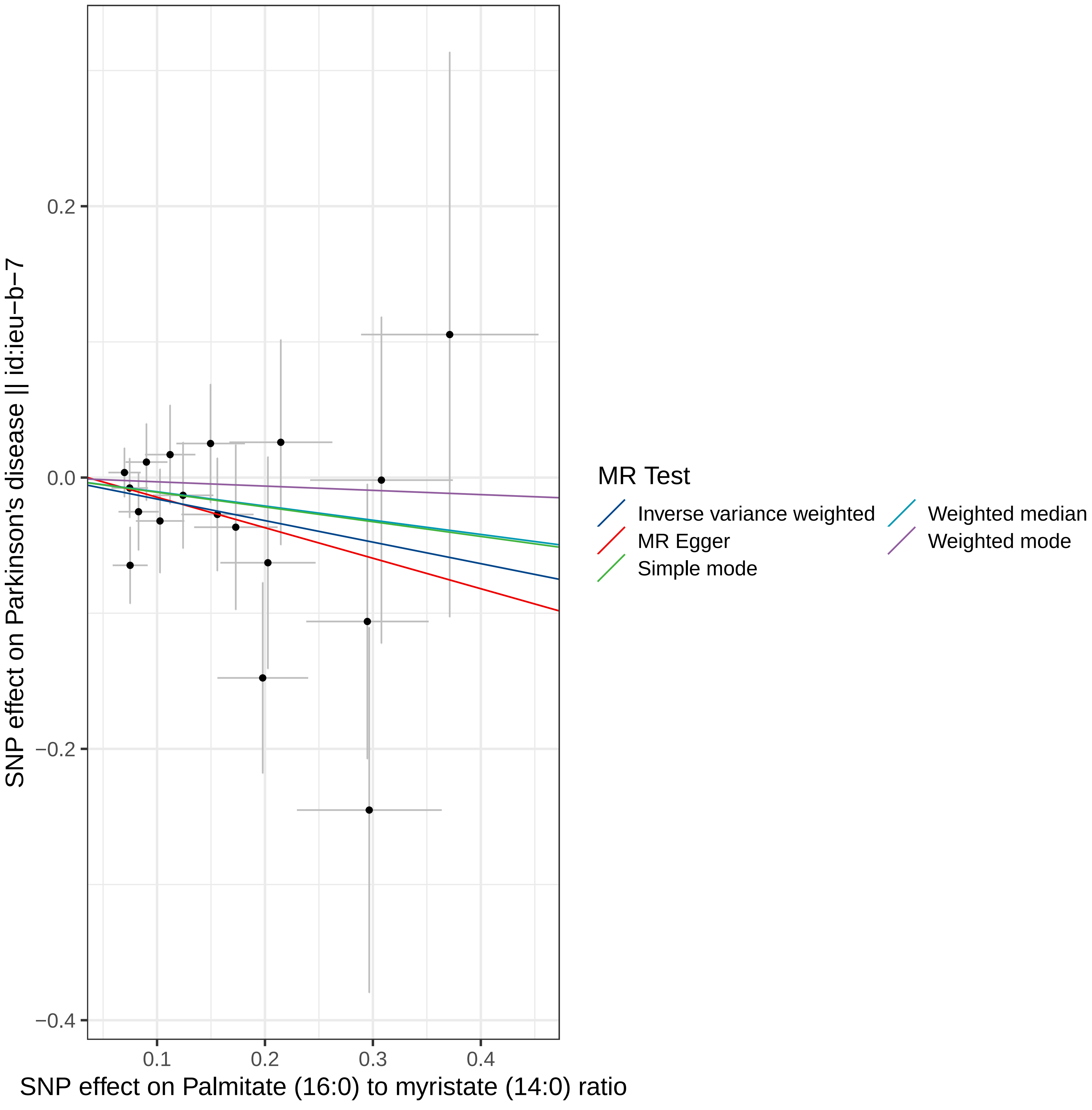

SNP effect on Parkinson's disease || id:ieu−b−7
0.2
0.0
−0.2
−0.4
0.1
0.2
0.3
0.4
SNP effect on Palmitate (16:0) to myristate (14:0) ratio
MR Test
Inverse variance weighted
MR Egger
Simple mode
Weighted median
Weighted mode

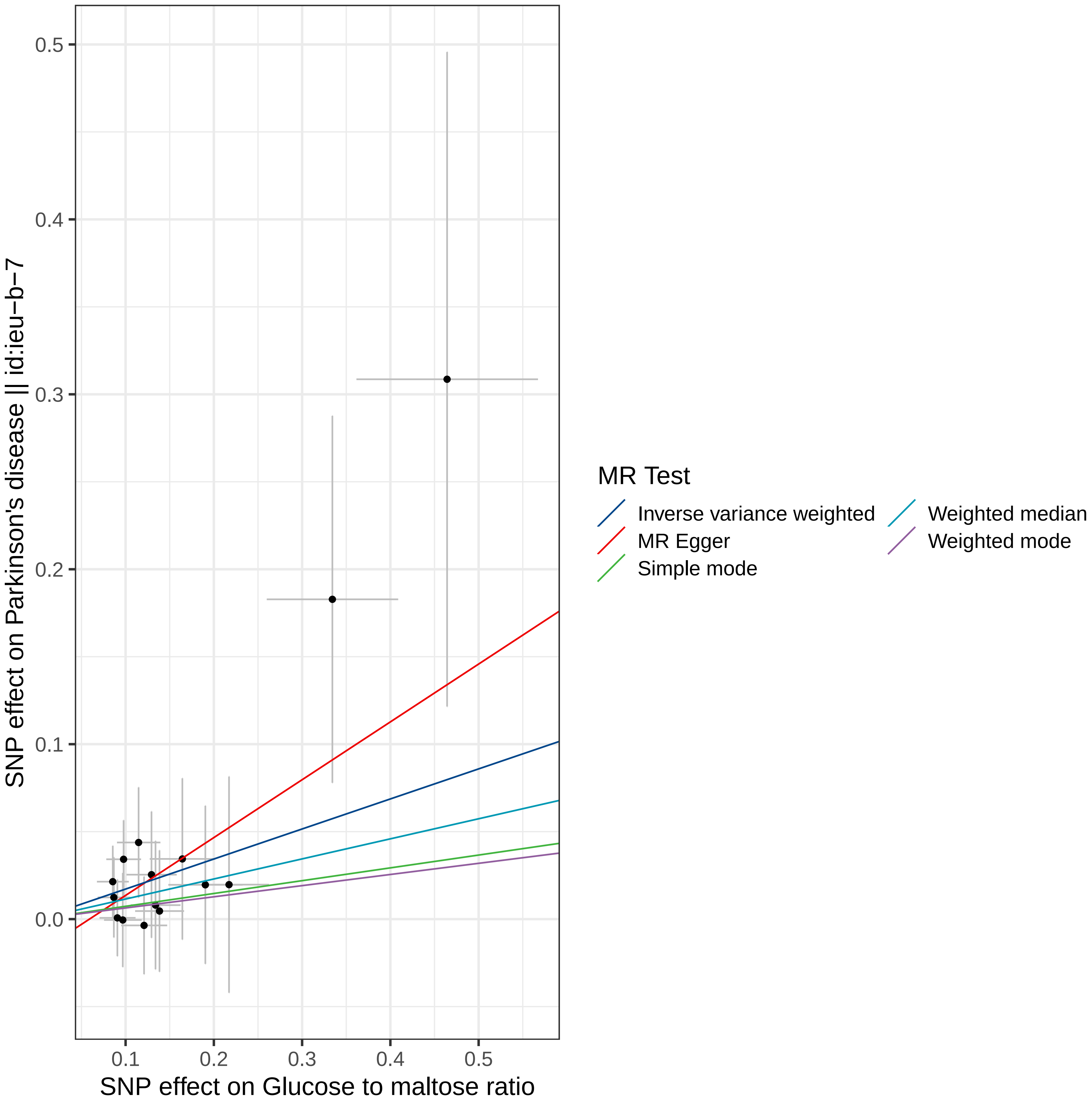

0.5
0.4
0.3
0.2
0.1
0.0
0.1
0.2
0.3
0.4
0.5
SNP effect on Parkinson's disease || id:ieu−b−7
SNP effect on Glucose to maltose ratio
MR Test
Inverse variance weighted
MR Egger
Simple mode
Weighted median
Weighted mode

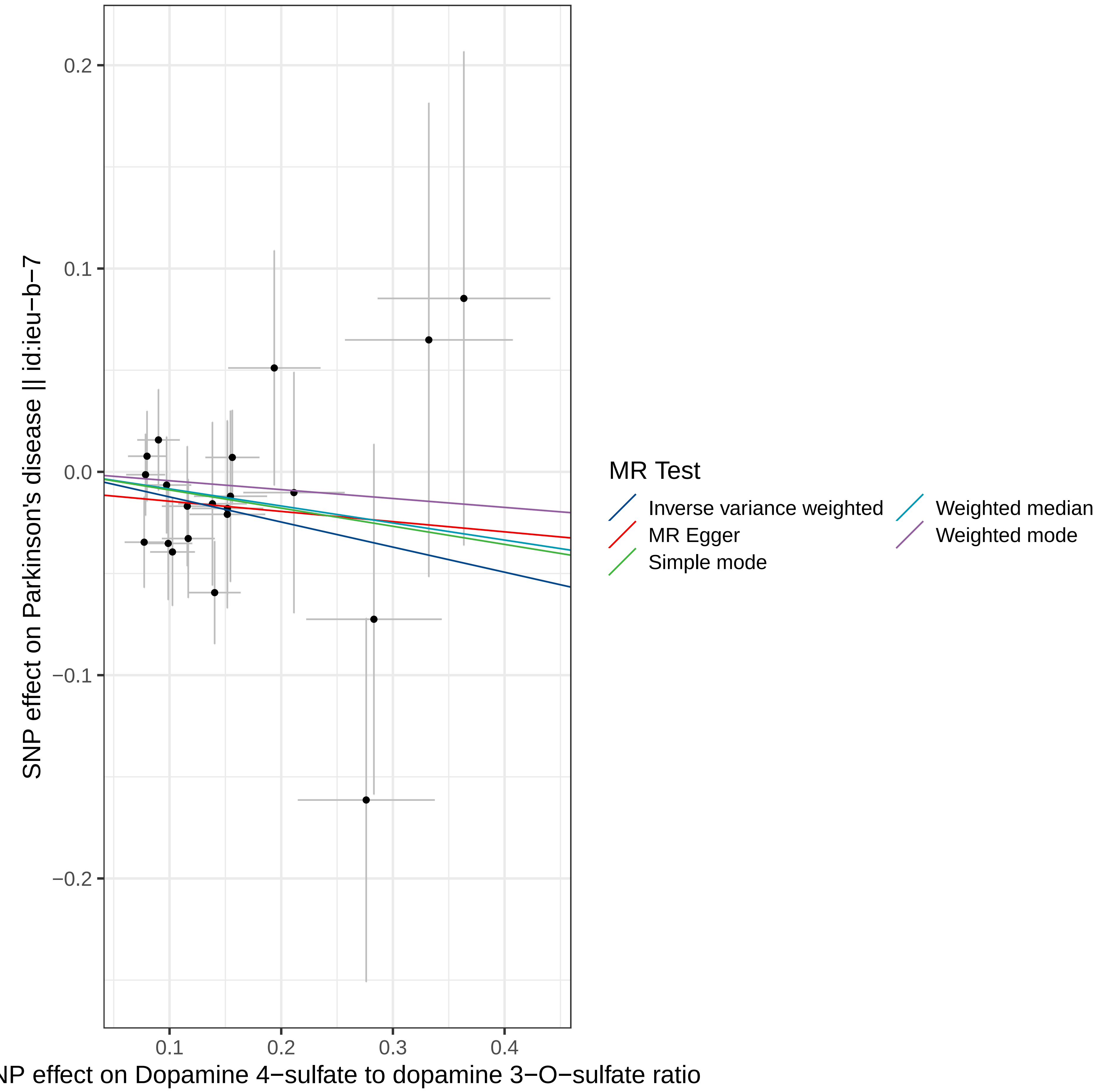

0.2
0.1
0.0
−0.1
−0.2
0.1
0.2
0.3
0.4
SNP effect on Parkinson's disease || id:ieu−b−7
NP effect on Dopamine 4−sulfate to dopamine 3−O−sulfate ratio
MR Test
Inverse variance weighted
MR Egger
Simple mode
Weighted median
Weighted mode

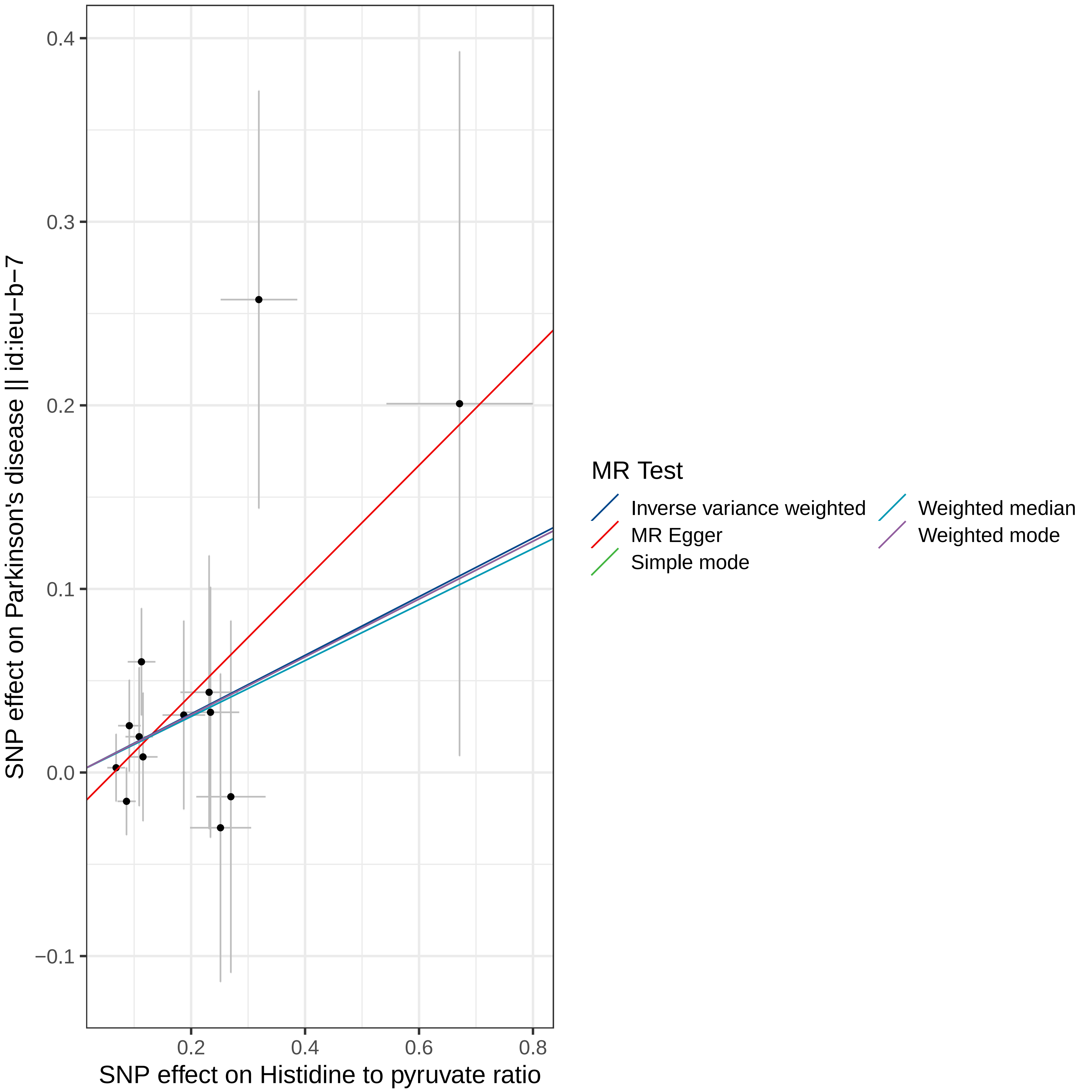

SNP effect on Parkinson's disease || id:ieu−b−7
SNP effect on Histidine to pyruvate ratio
0.4
0.3
0.2
0.1
0.0
−0.1
0.2
0.4
0.6
0.8
MR Test
Inverse variance weighted
MR Egger
Simple mode
Weighted median
Weighted mode

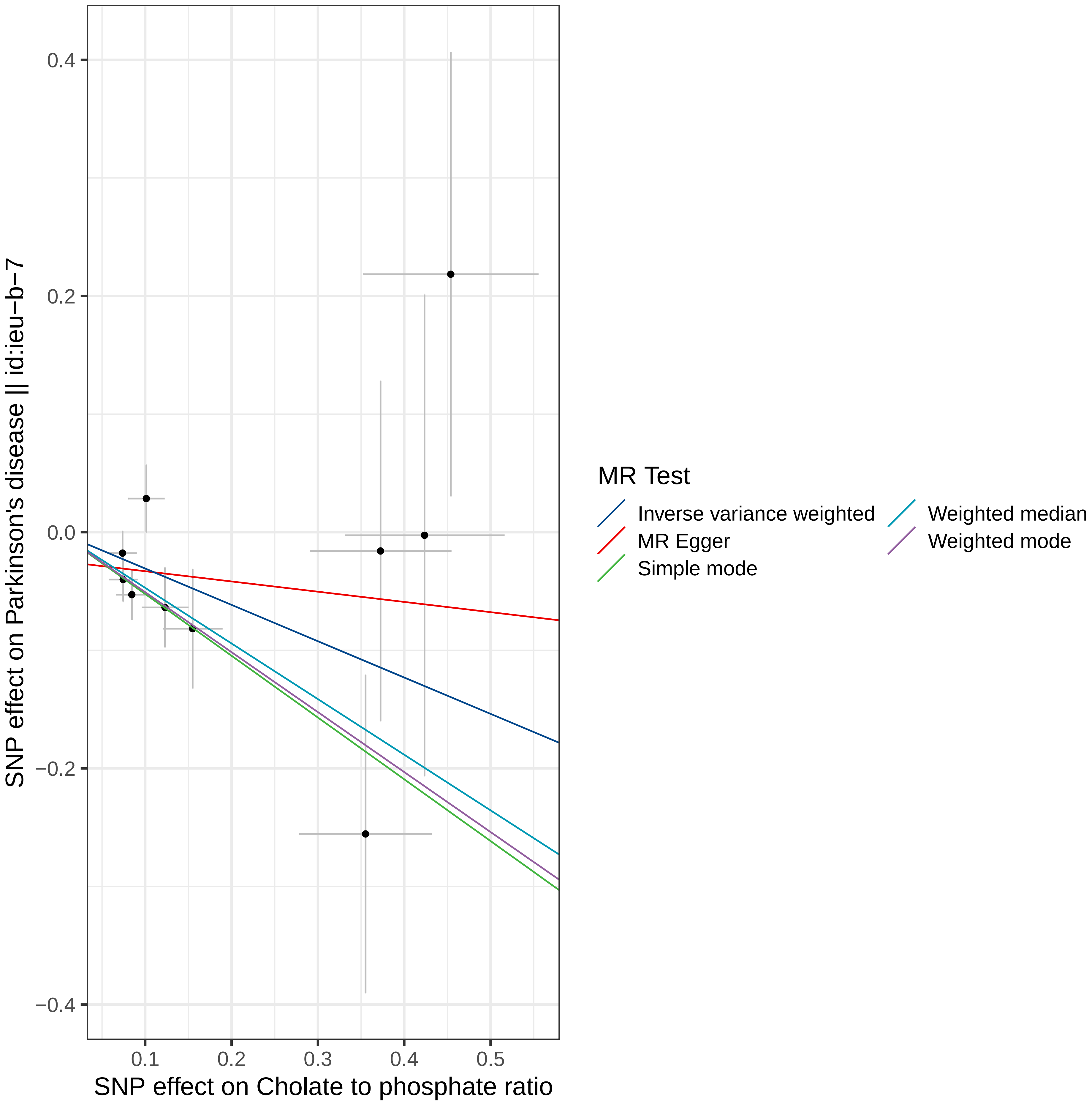

0.4
0.2
0.0
−0.2
−0.4
0.1
0.2
0.3
0.4
0.5
SNP effect on Parkinson's disease || id:ieu−b−7
SNP effect on Cholate to phosphate ratio
MR Test
Inverse variance weighted
MR Egger
Simple mode
Weighted median
Weighted mode

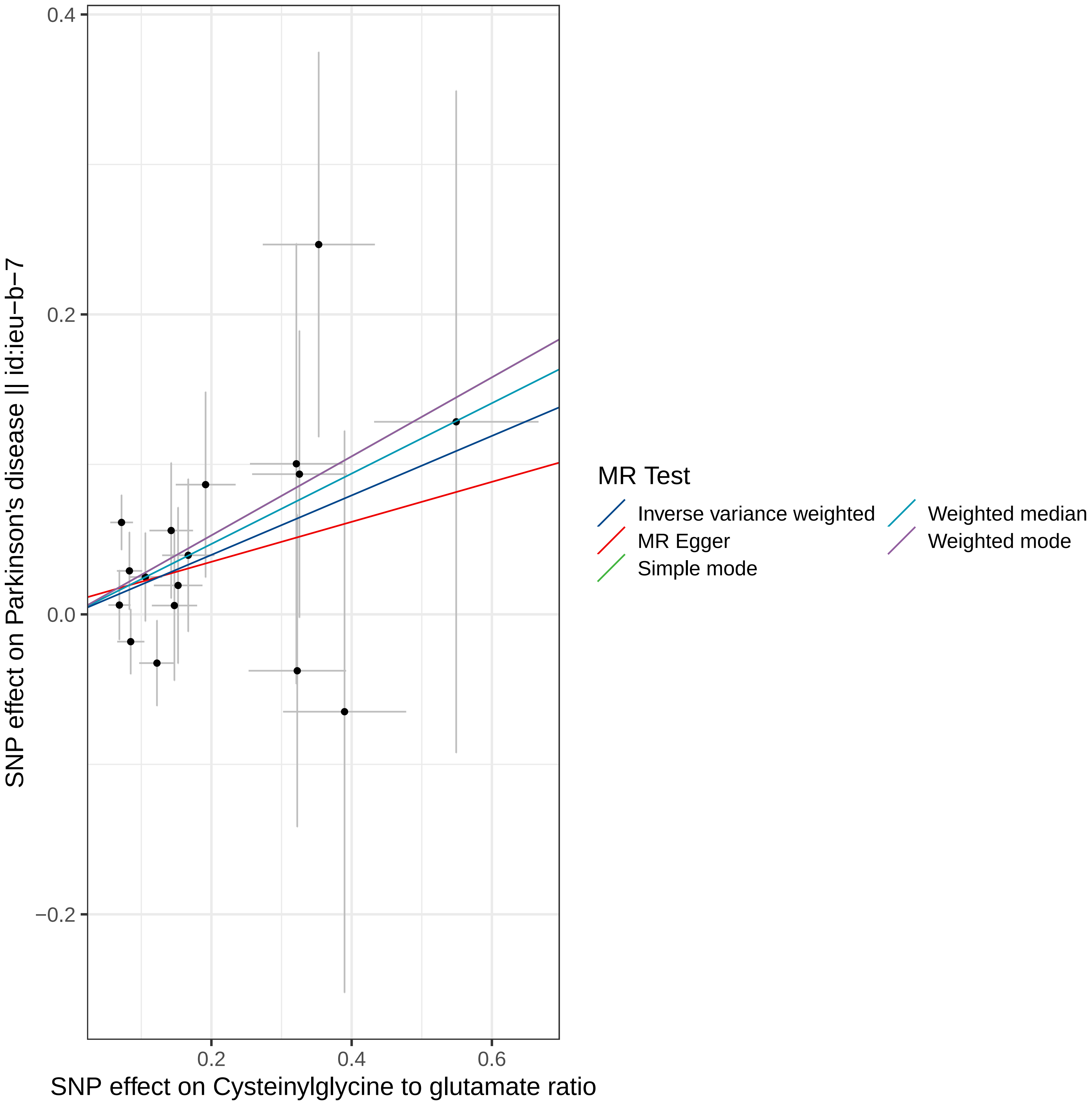

0.4
0.2
0.0
−0.2
0.2
0.4
0.6
SNP effect on Parkinson's disease || id:ieu−b−7
SNP effect on Cysteinylglycine to glutamate ratio
MR Test
Inverse variance weighted
MR Egger
Simple mode
Weighted median
Weighted mode

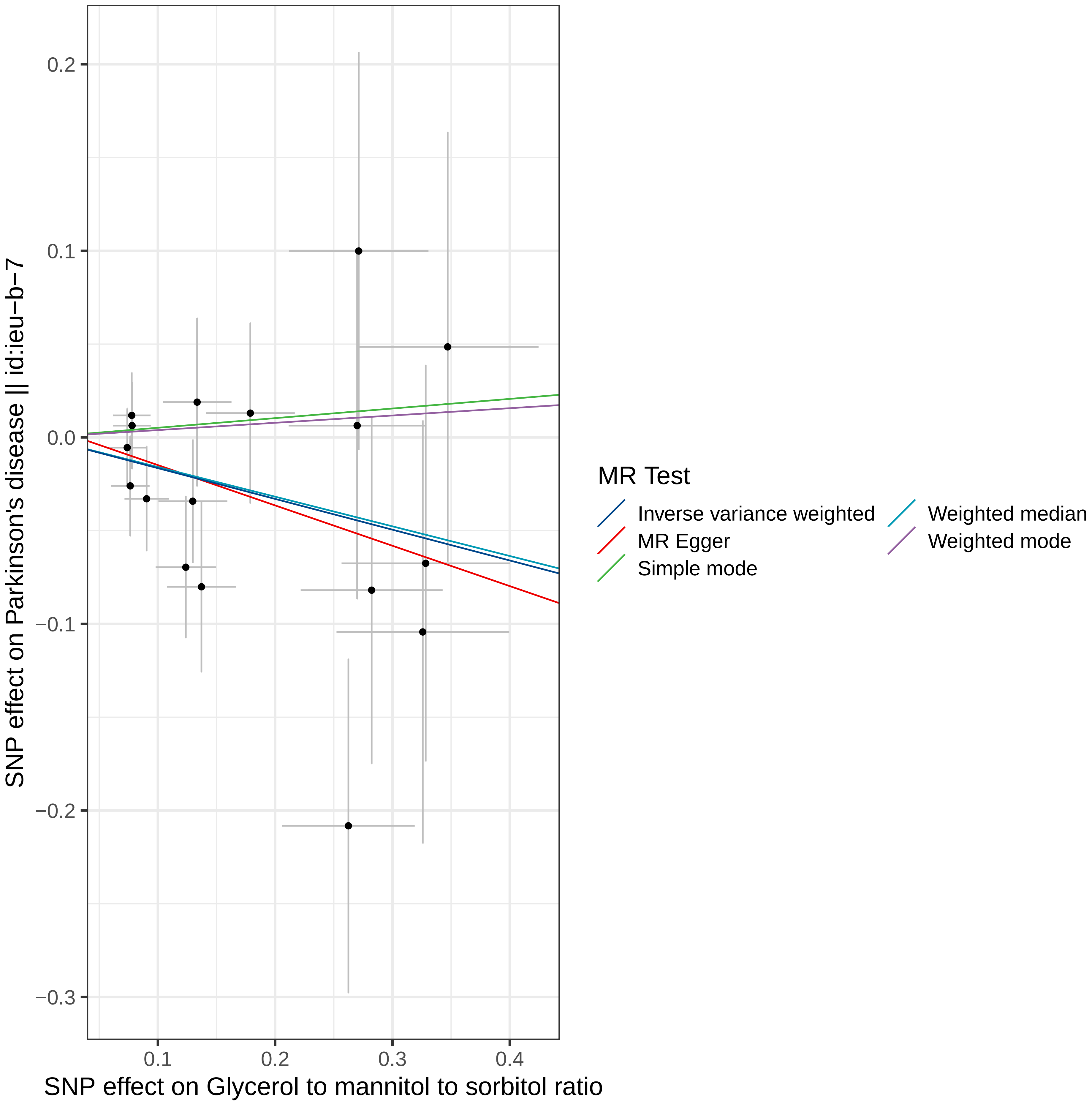
SNP effect on Parkinson's disease || id:ieu−b−7
0.2
0.1
0.0
−0.1
−0.2
−0.3
0.1
0.2
0.3
0.4
SNP effect on Glycerol to mannitol to sorbitol ratio
MR Test
Inverse variance weighted
MR Egger
Simple mode
Weighted median
Weighted mode

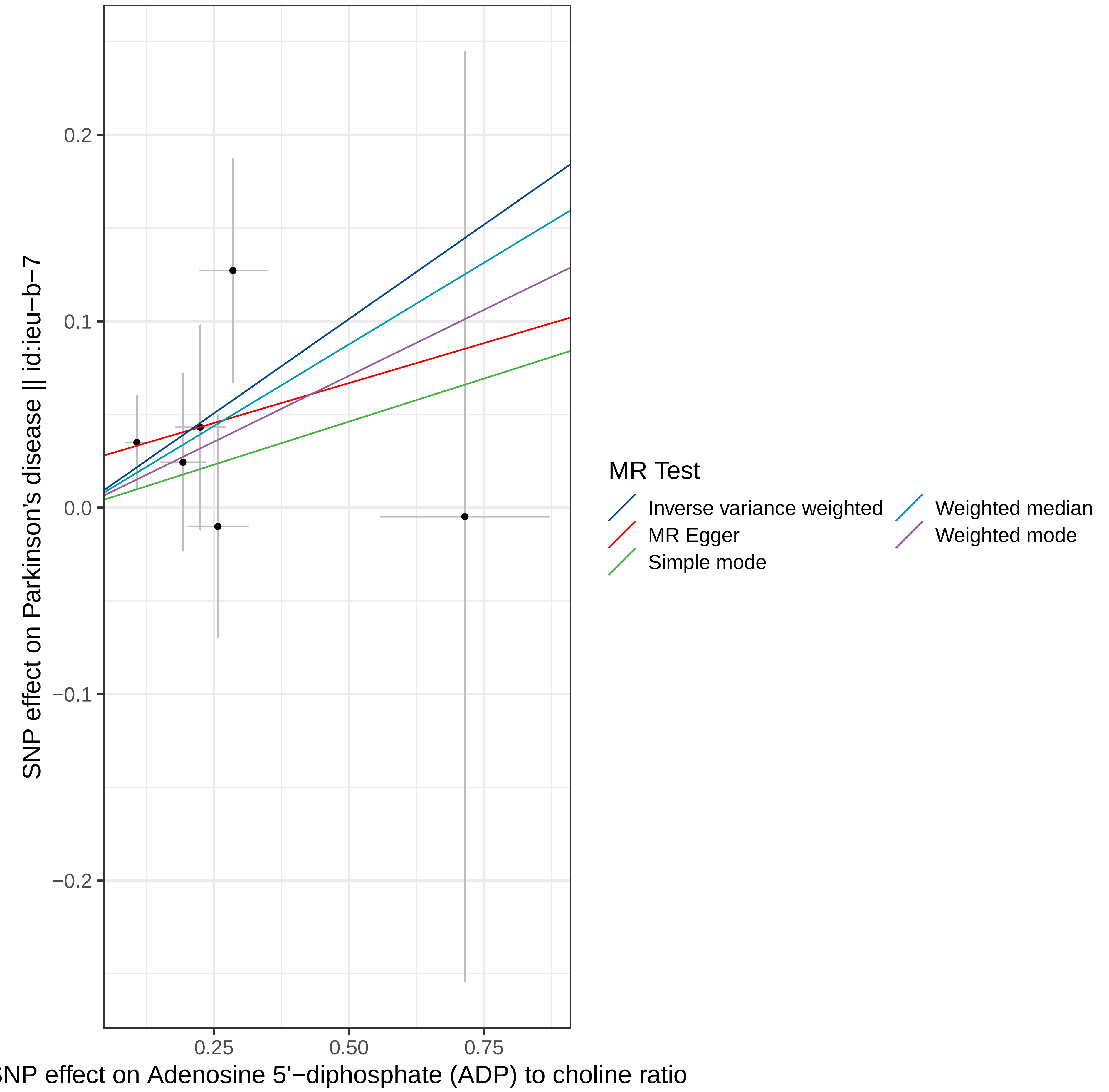

0.2
0.1
0.0
−0.1
−0.2
SNP effect on Parkinson's disease || id:ieu−b−7
0.25
0.50
0.75
SNP effect on Adenosine 5'−diphosphate (ADP) to choline ratio
MR Test
Inverse variance weighted
MR Egger
Simple mode
Weighted median
Weighted mode

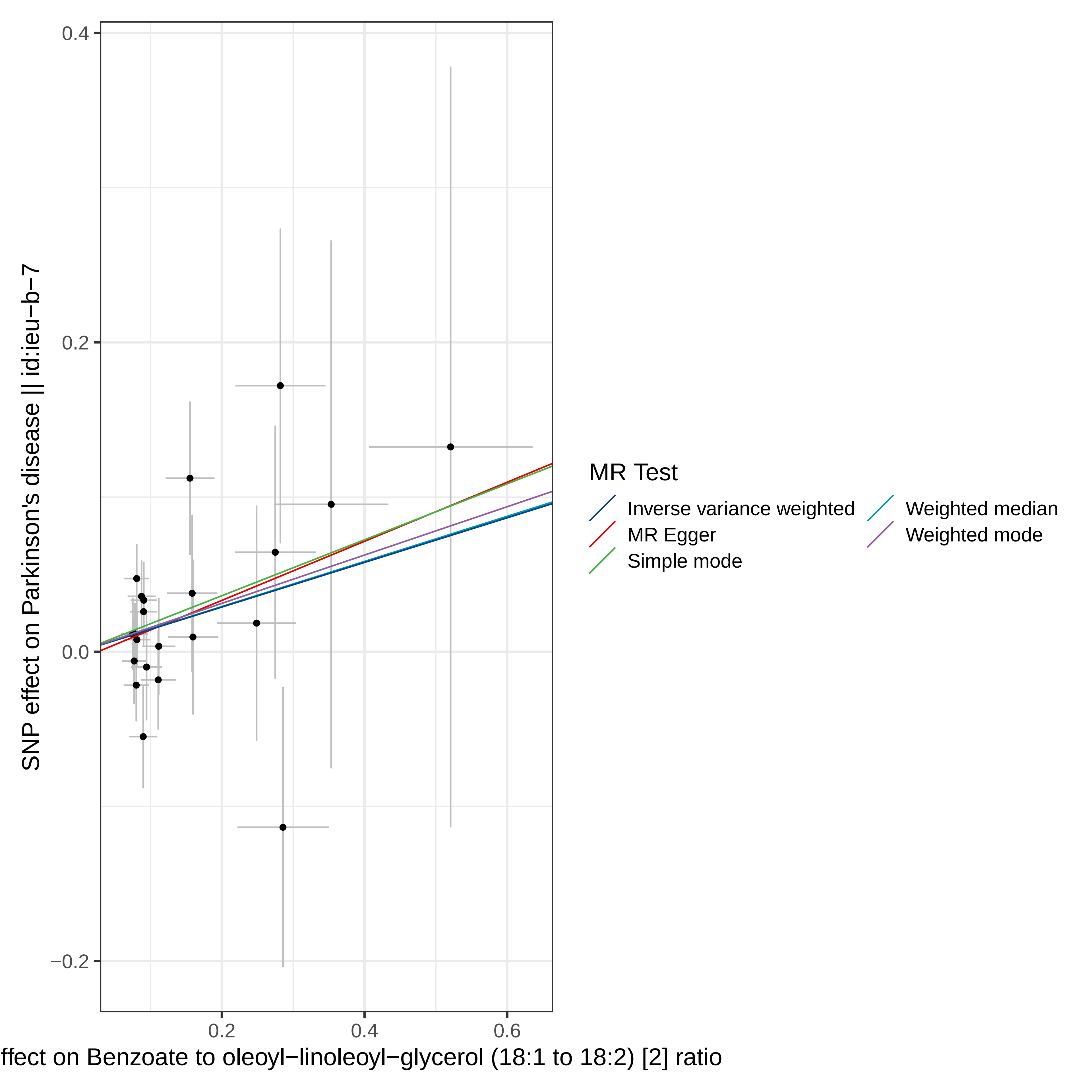
0.4
0.2
0.0
−0.2
0.2
0.4
0.6
SNP effect on Parkinson's disease || id:ieu−b−7
ffect on Benzoate to oleoyl−linoleoyl−glycerol (18:1 to 18:2) [2] ratio
MR Test
Inverse variance weighted
MR Egger
Simple mode
Weighted median
Weighted mode

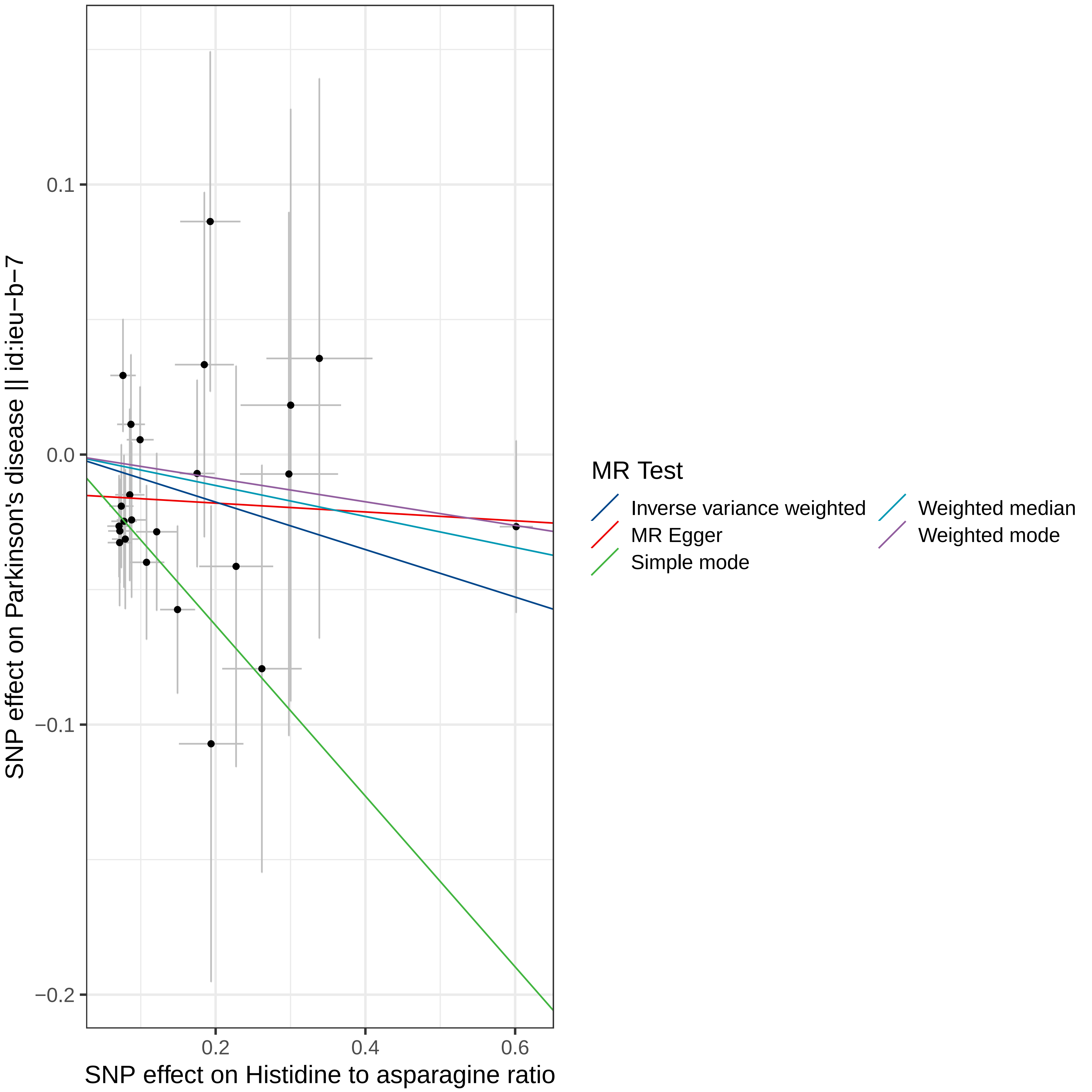

SNP effect on Parkinson's disease || id:ieu−b−7
0.1
0.0
−0.1
−0.2
0.2
0.4
0.6
SNP effect on Histidine to asparagine ratio
MR Test
Inverse variance weighted
MR Egger
Simple mode
Weighted median
Weighted mode

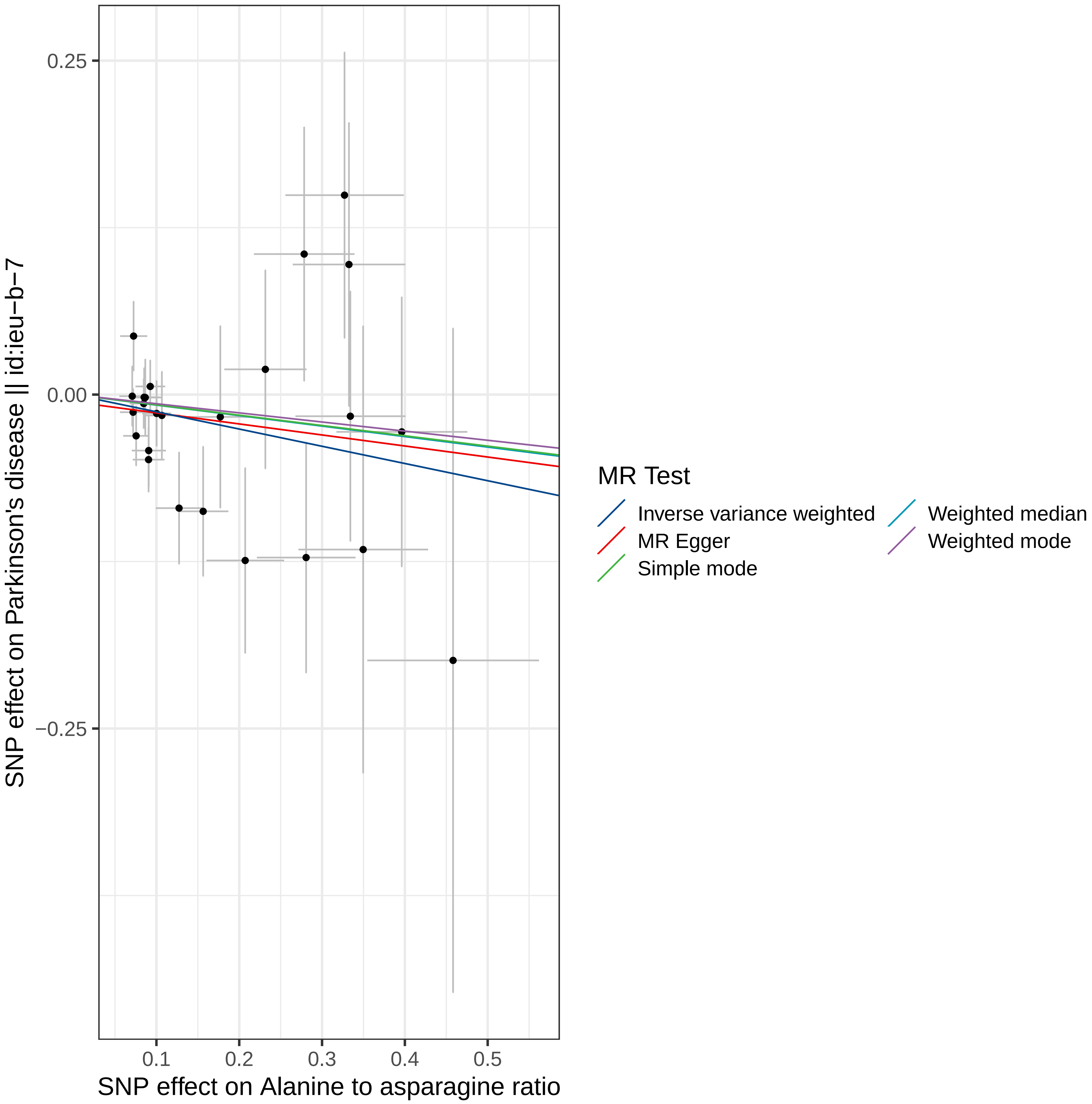

0.25
0.00
−0.25
0.1
0.2
0.3
0.4
0.5
SNP effect on Parkinson's disease || id:ieu−b−7
SNP effect on Alanine to asparagine ratio
MR Test
Inverse variance weighted
MR Egger
Simple mode
Weighted median
Weighted mode

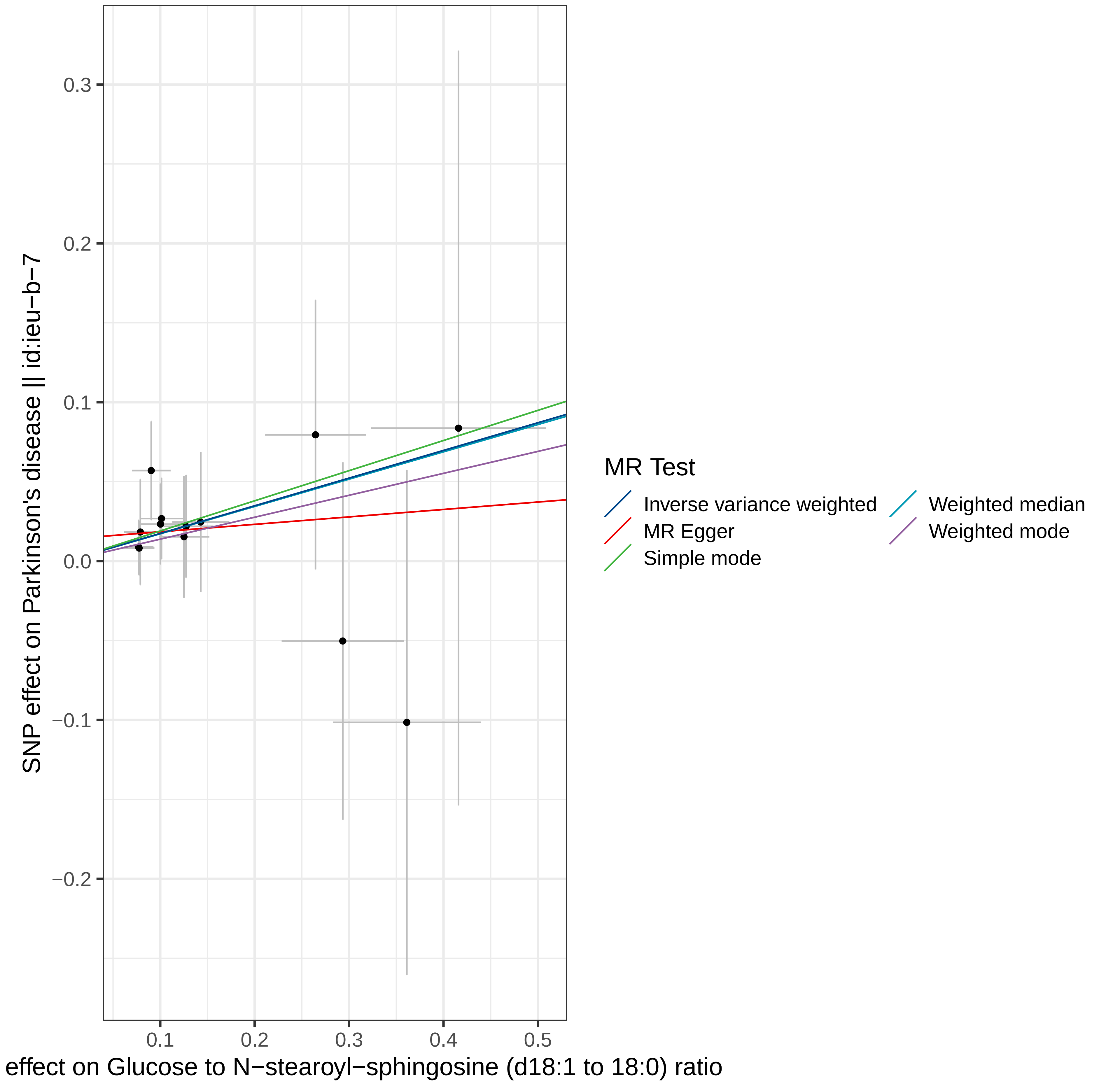

0.3
0.2
0.1
0.0
−0.1
−0.2
0.1
0.2
0.3
0.4
0.5
SNP effect on Parkinson's disease || id:ieu−b−7
effect on Glucose to N−stearoyl−sphingosine (d18:1 to 18:0) ratio
MR Test
Inverse variance weighted
MR Egger
Simple mode
Weighted median
Weighted mode
