## Supplemental Figure 8 for "Cerebrospinal Fluid and Plasma Metabolites with Parkinson’s Disease: A Mendelian Randomization Study"

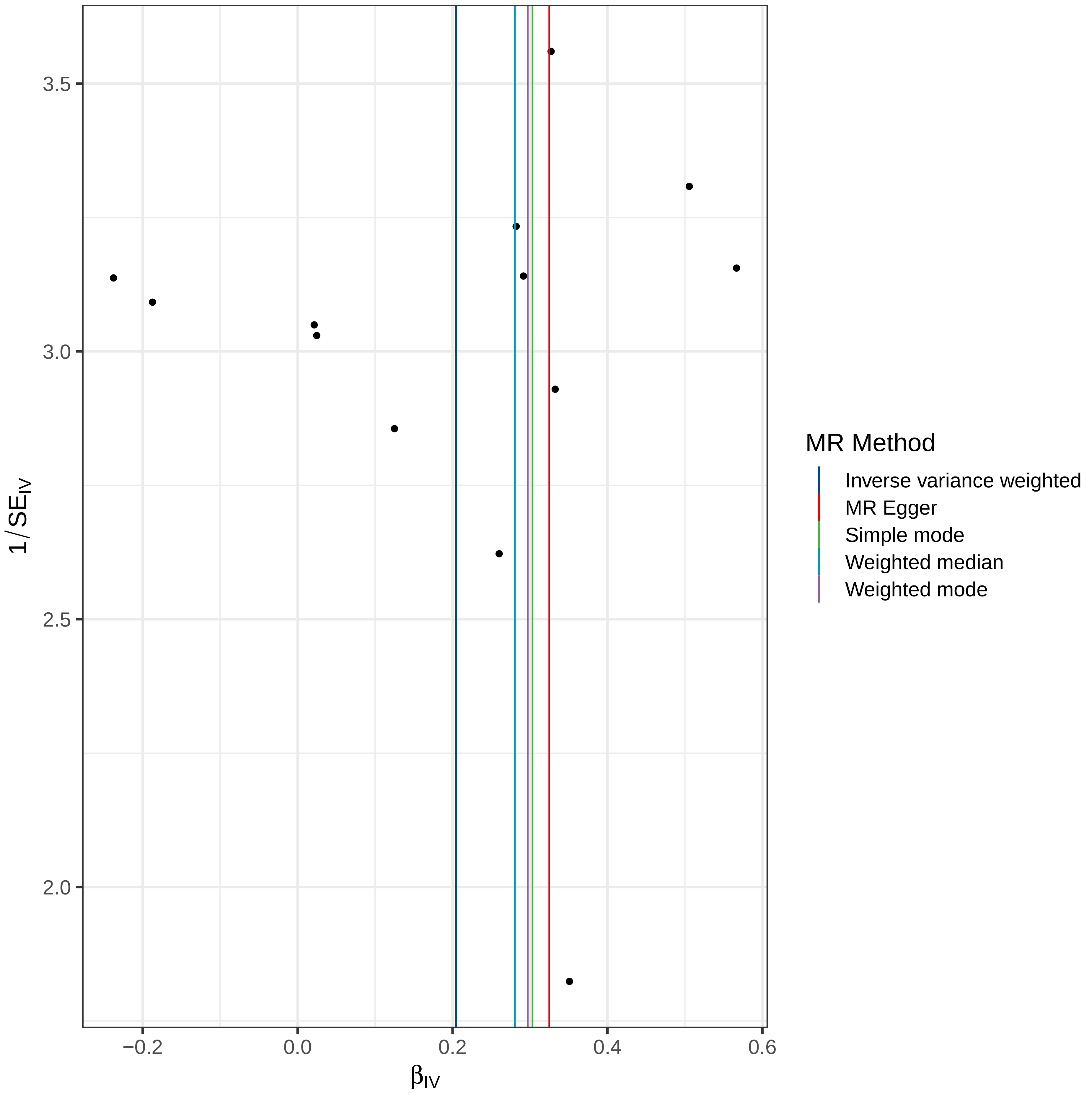

3.5
3.0
2.5
2.0
1/SE_IV
−0.2
0.0
0.2
0.4
0.6
β_IV
MR Method
Inverse variance weighted
MR Egger
Simple mode
Weighted median
Weighted mode

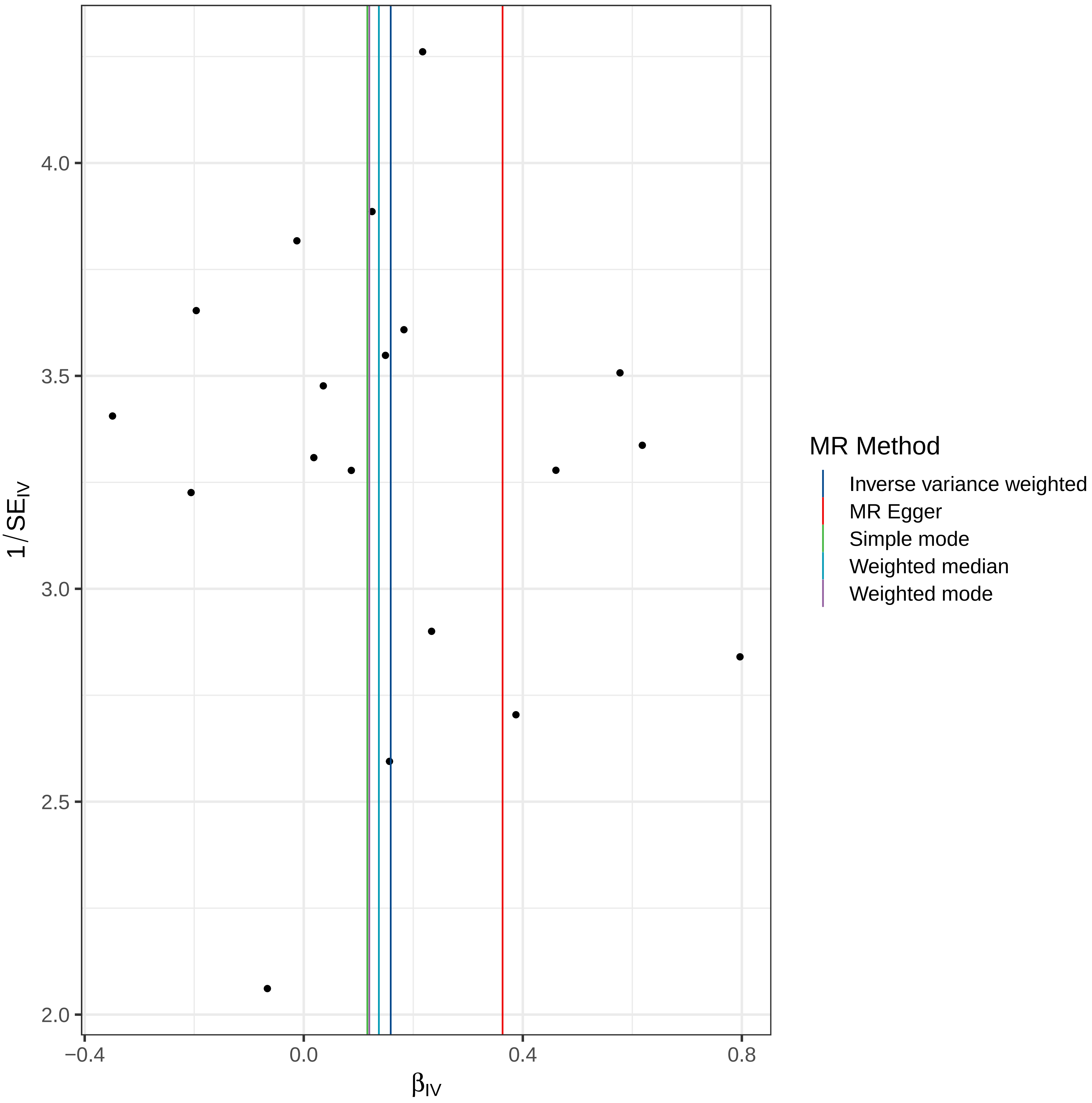

MR Method
Inverse variance weighted
MR Egger
Simple mode
Weighted median
Weighted mode
1/SE_IV
β_IV
4.0
3.5
3.0
2.5
2.0
−0.4
0.0
0.4
0.8

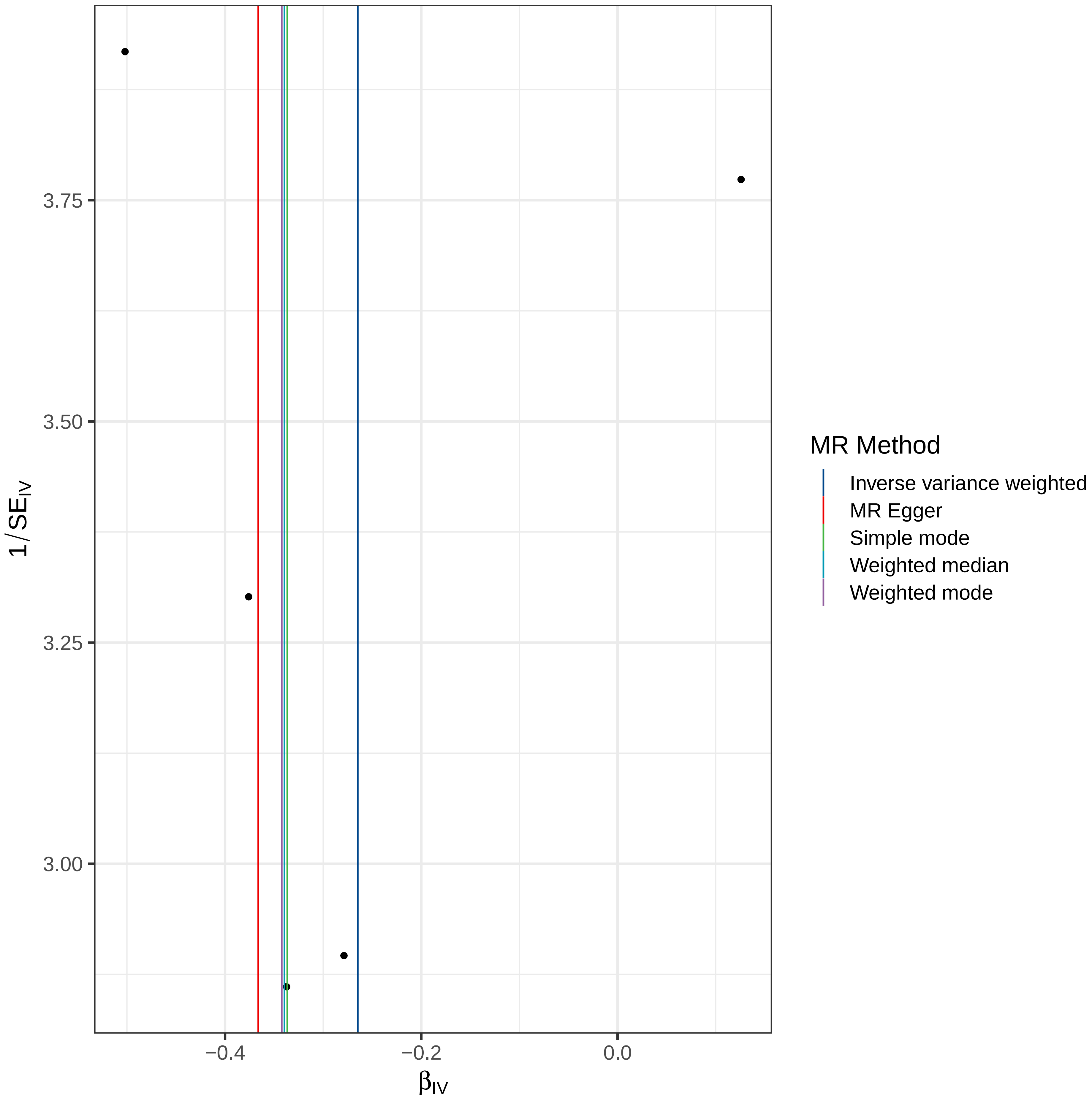

3.75
3.50
3.25
3.00
1/SE_IV
−0.4
−0.2
0.0
β_IV
MR Method
Inverse variance weighted
MR Egger
Simple mode
Weighted median
Weighted mode

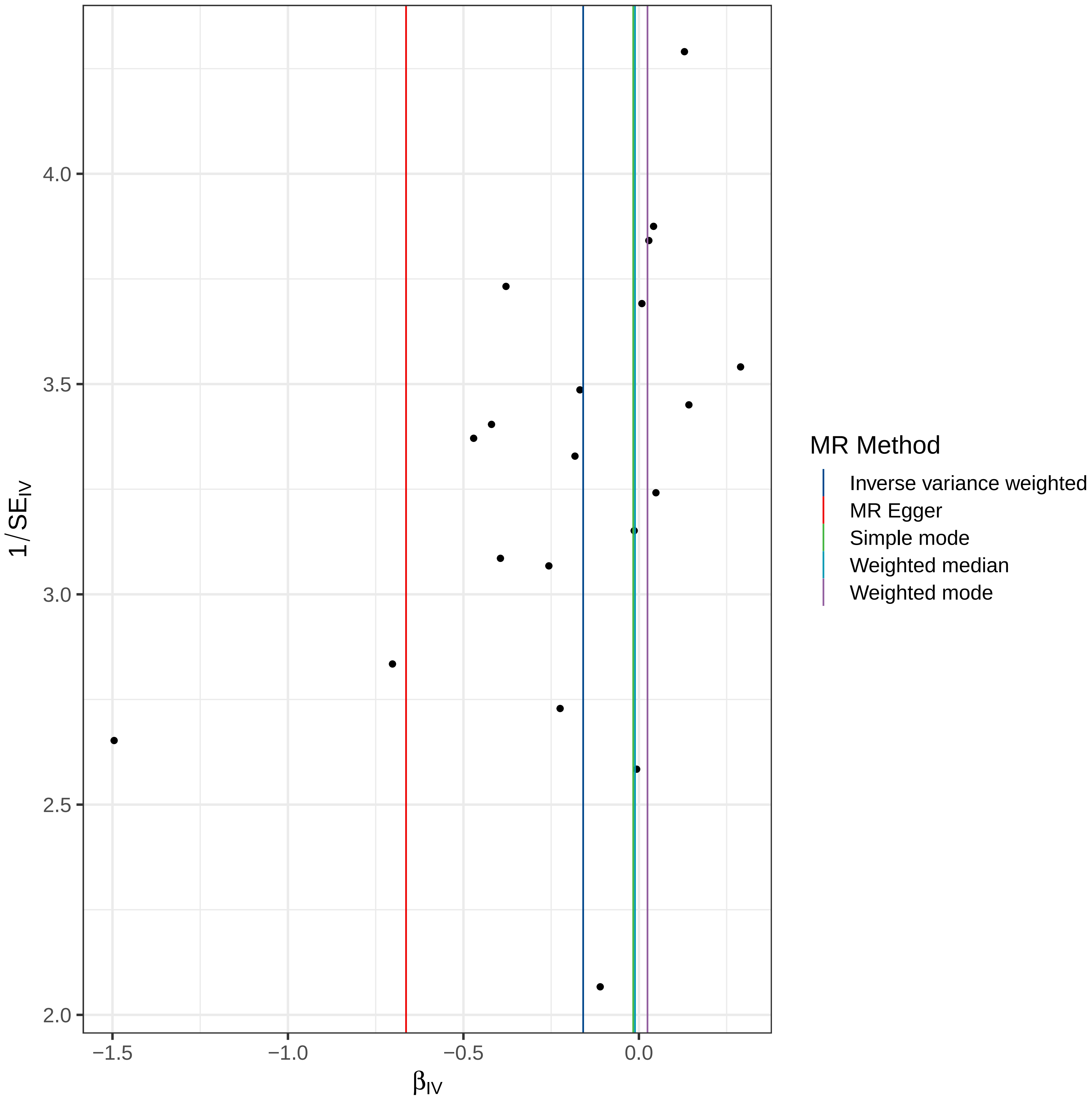

4.0
3.5
3.0
2.5
2.0
1/SE_IV
−1.5
−1.0
−0.5
0.0
β_IV
MR Method
Inverse variance weighted
MR Egger
Simple mode
Weighted median
Weighted mode

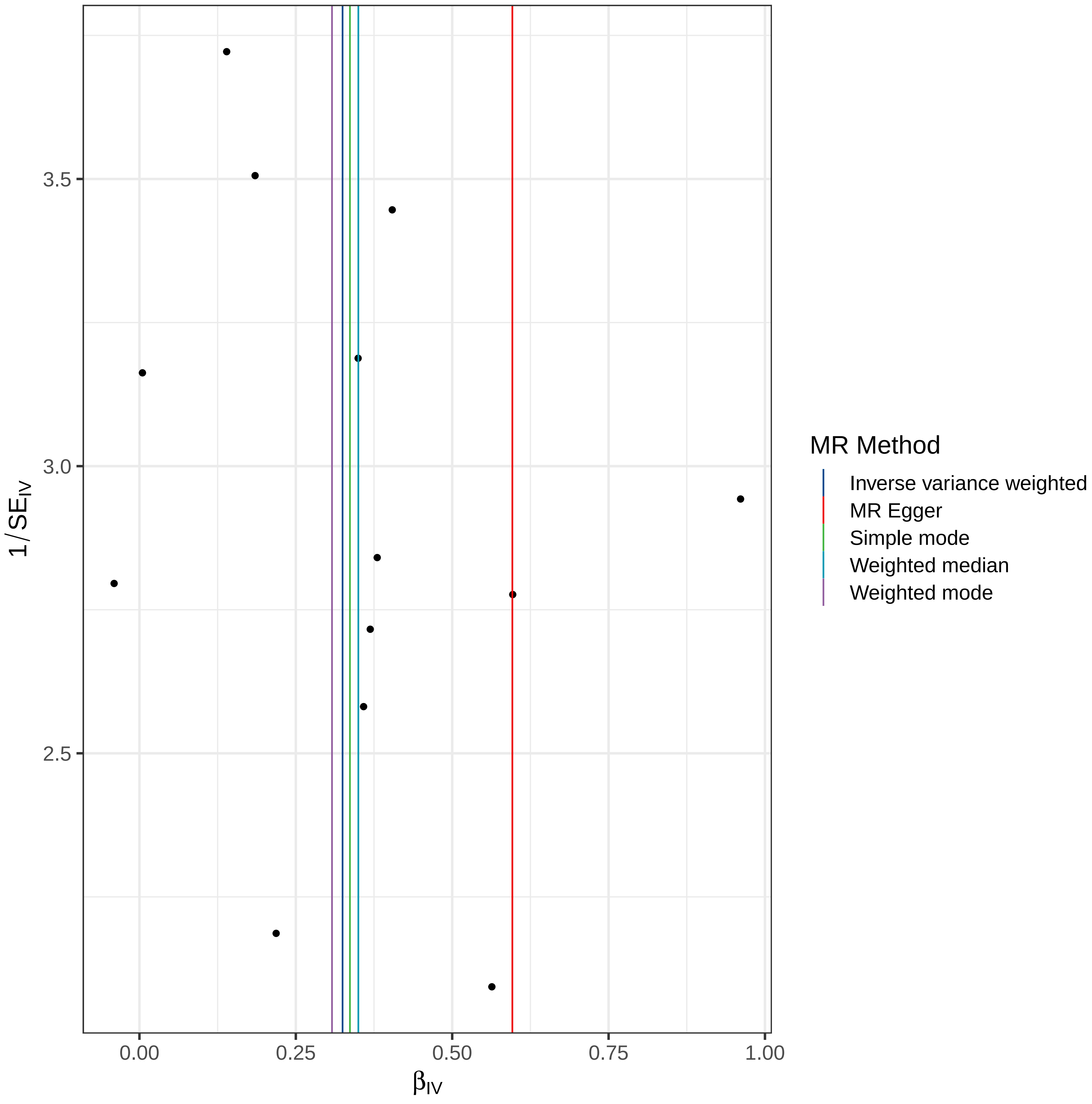

3.5
3.0
2.5
0.00
0.25
0.50
0.75
1.00
$\beta_{IV}$
$1/SE_{IV}$
MR Method
Inverse variance weighted
MR Egger
Simple mode
Weighted median
Weighted mode

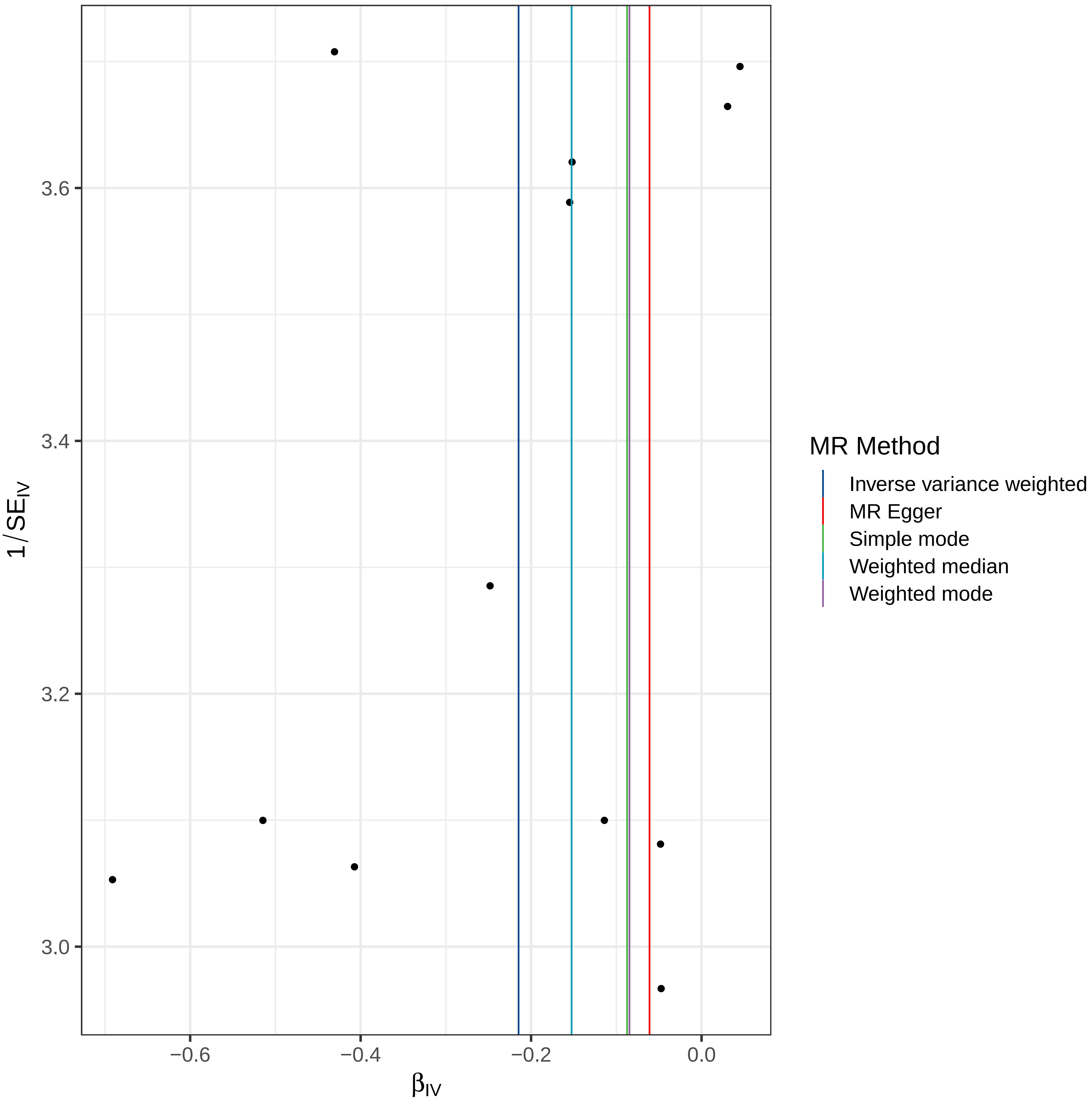

3.6
3.4
3.2
3.0
1/SE$_{IV}$
−0.6
−0.4
−0.2
0.0
β$_{IV}$
MR Method
Inverse variance weighted
MR Egger
Simple mode
Weighted median
Weighted mode

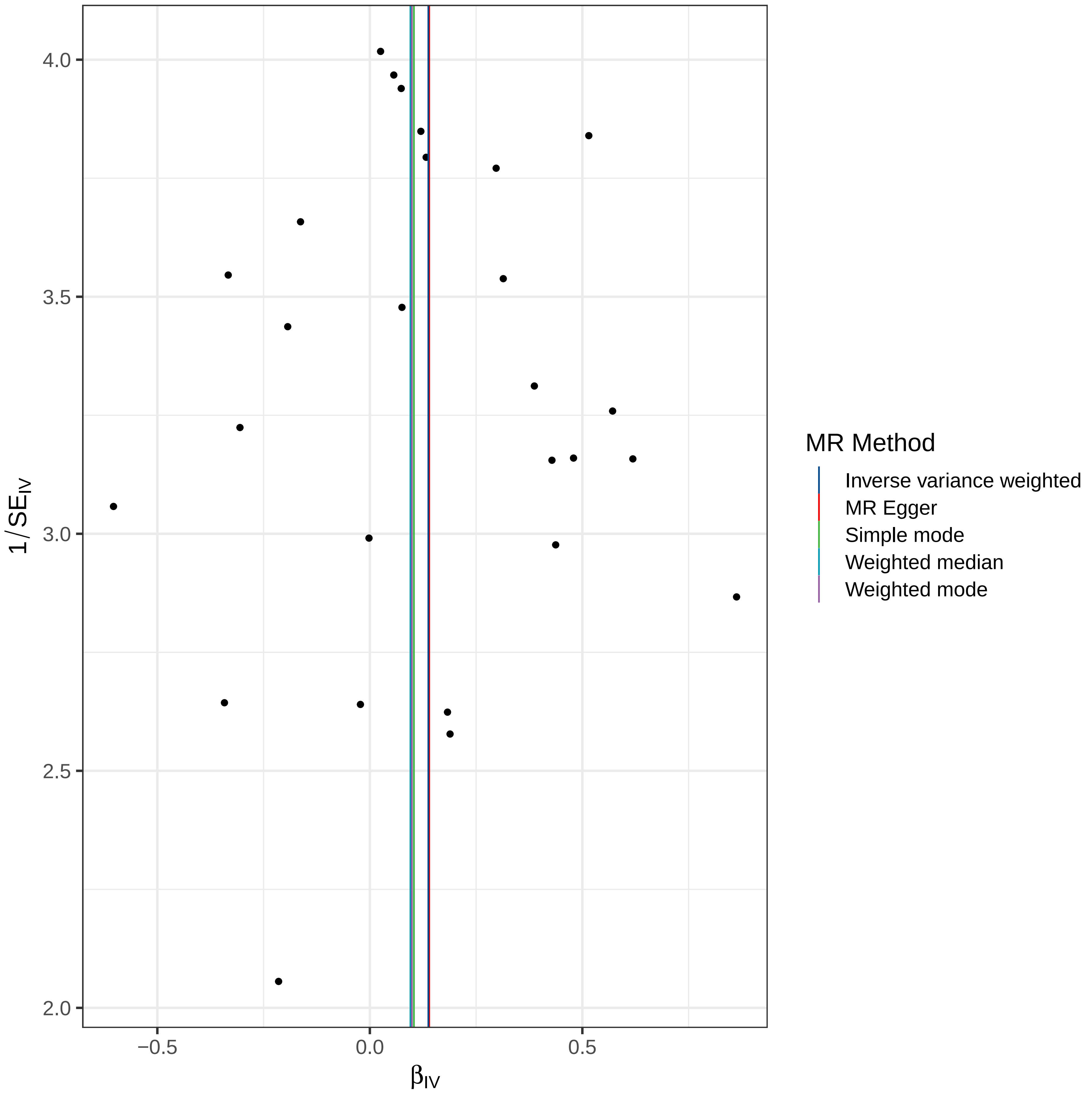
4.0
3.5
3.0
2.5
2.0
−0.5
0.0
0.5
1/SE_IV
β_IV
MR Method
Inverse variance weighted
MR Egger
Simple mode
Weighted median
Weighted mode

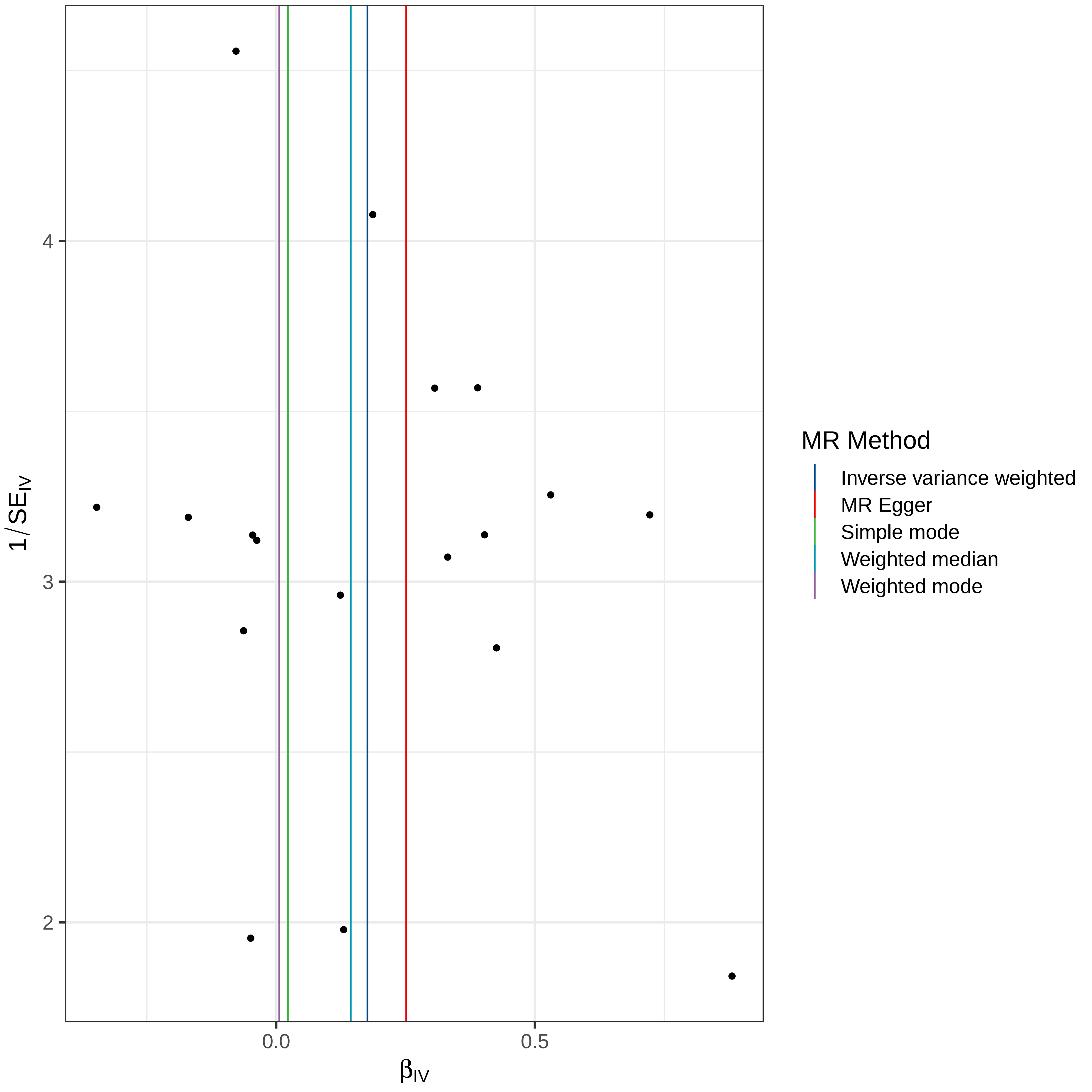

4
3
2
0.0
0.5
1/SE_IV
β_IV
MR Method
Inverse variance weighted
MR Egger
Simple mode
Weighted median
Weighted mode

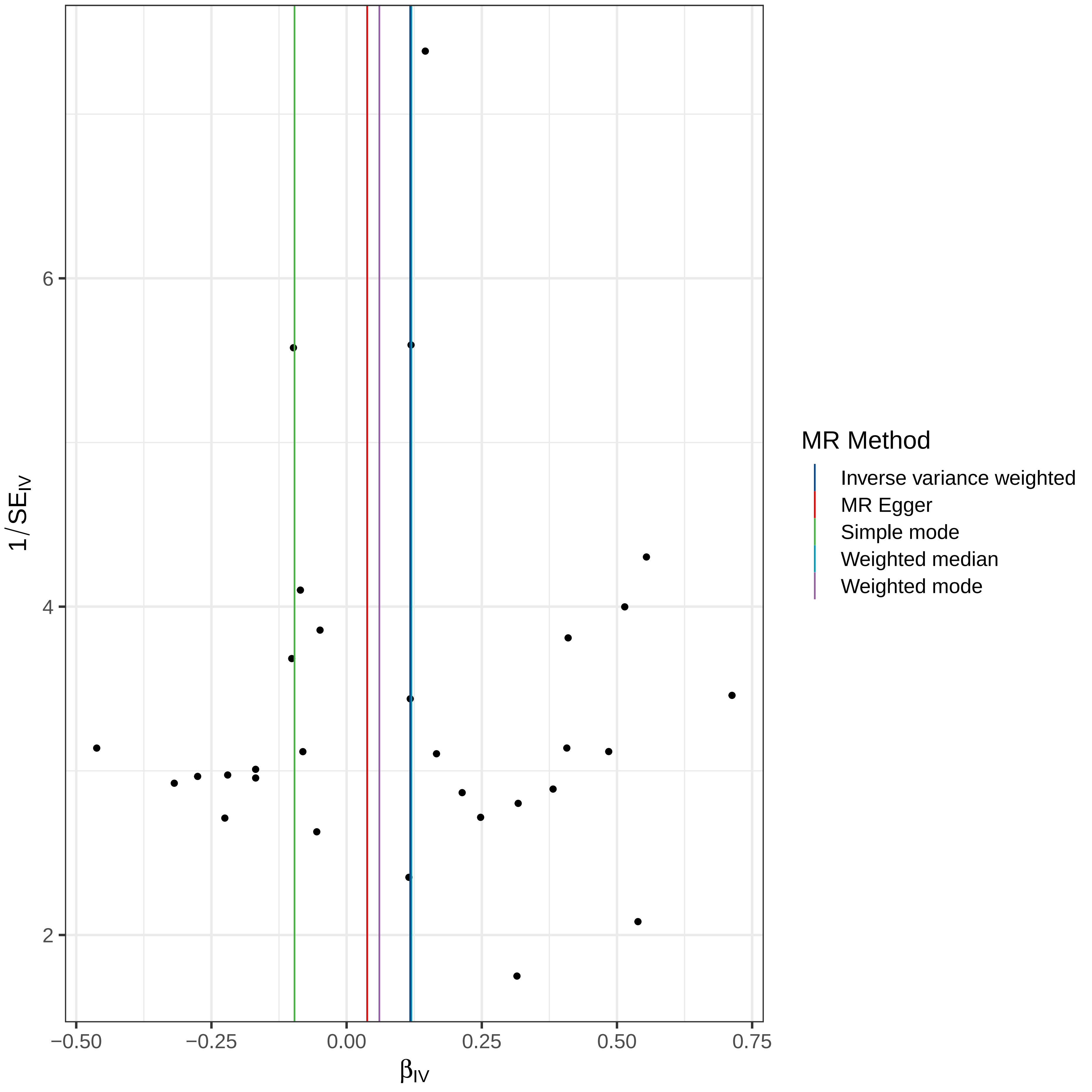

MR Method
Inverse variance weighted
MR Egger
Simple mode
Weighted median
Weighted mode
$1/SE_{IV}$
$\beta_{IV}$
6
4
2
−0.50
−0.25
0.00
0.25
0.50
0.75

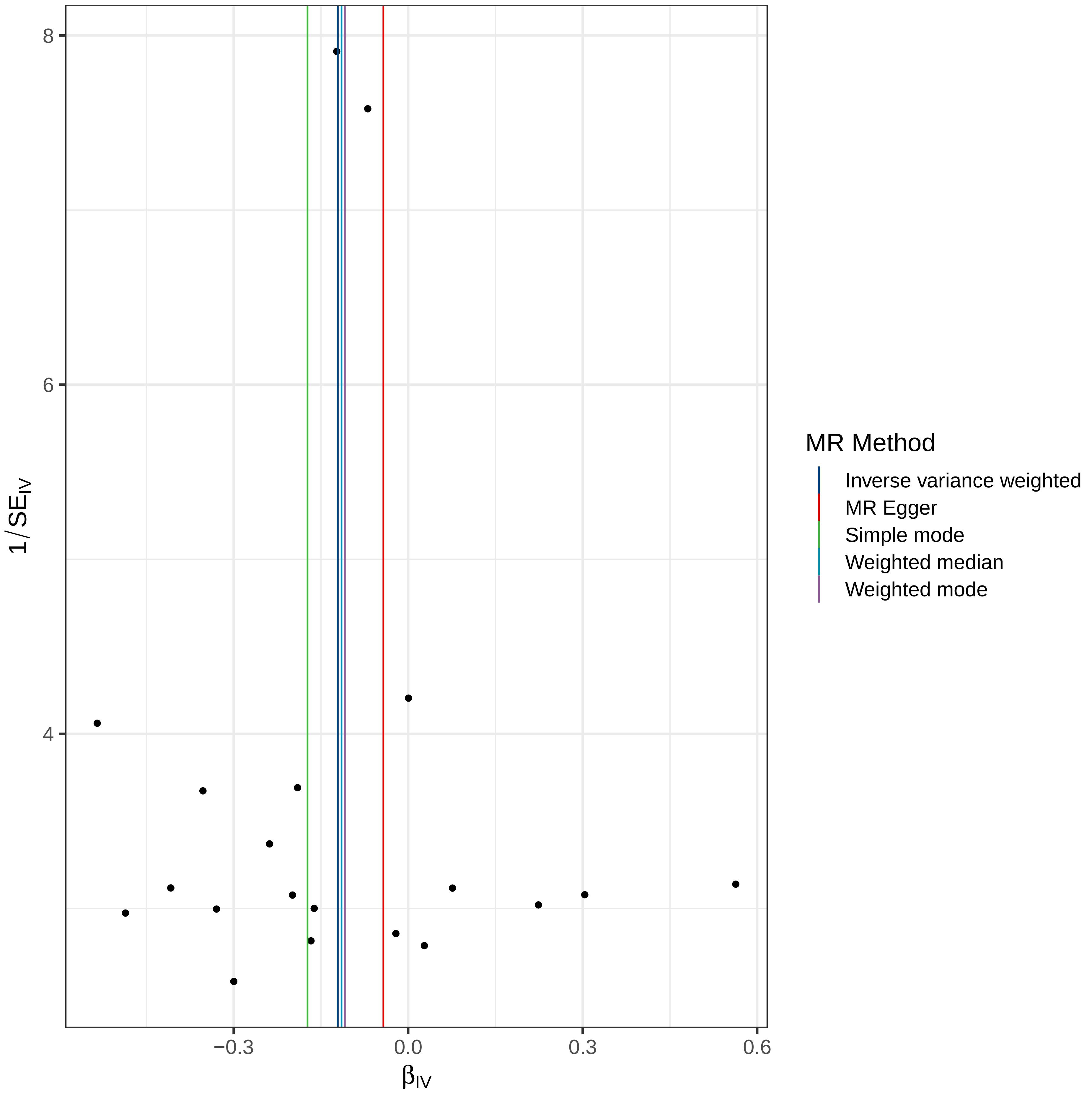
8
6
4
−0.3
0.0
0.3
0.6
$\beta_{IV}$
$1/SE_{IV}$
MR Method
Inverse variance weighted
MR Egger
Simple mode
Weighted median
Weighted mode

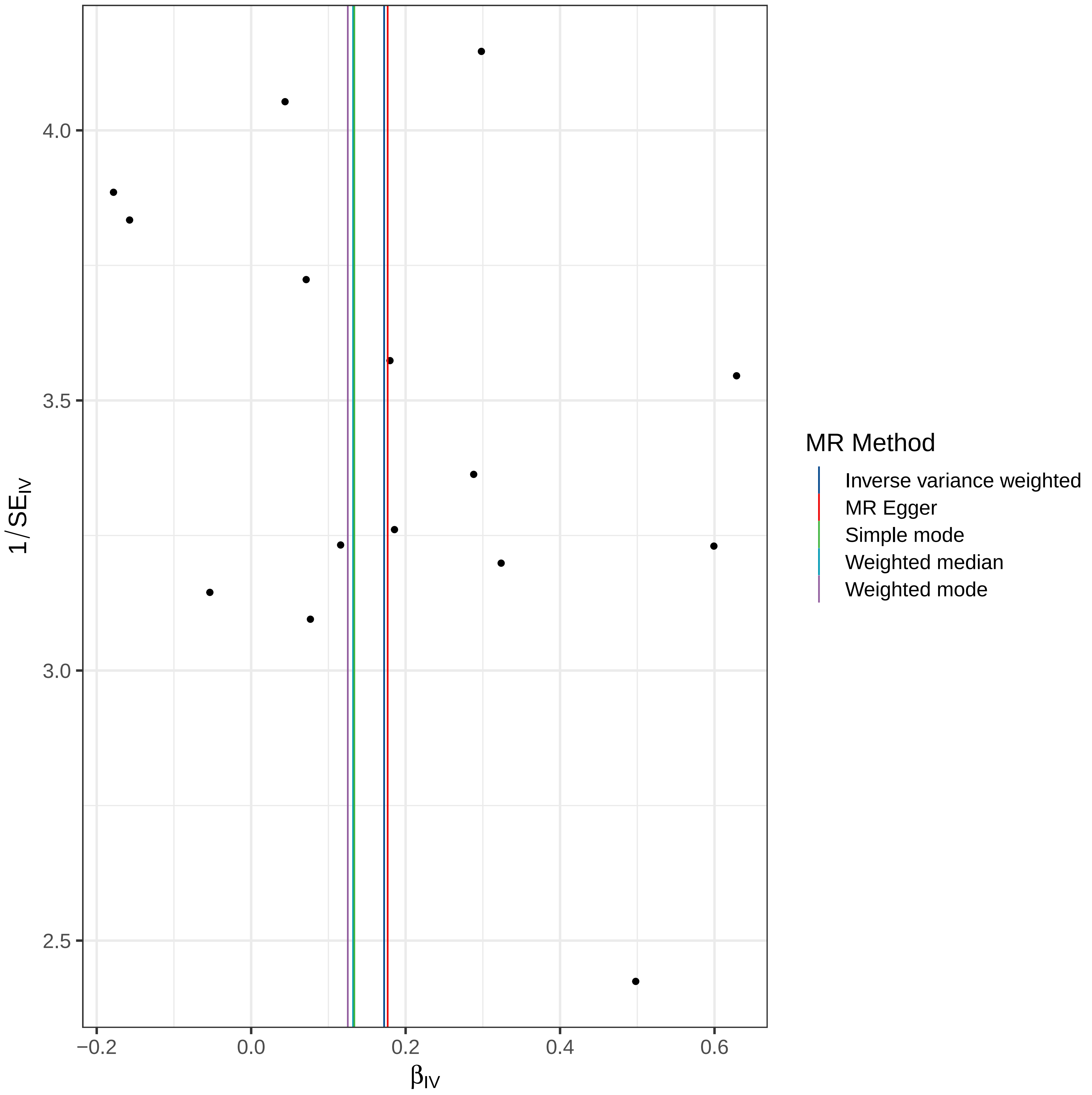

4.0
3.5
3.0
2.5
1/SE_IV
−0.2
0.0
0.2
0.4
0.6
β_IV
MR Method
Inverse variance weighted
MR Egger
Simple mode
Weighted median
Weighted mode

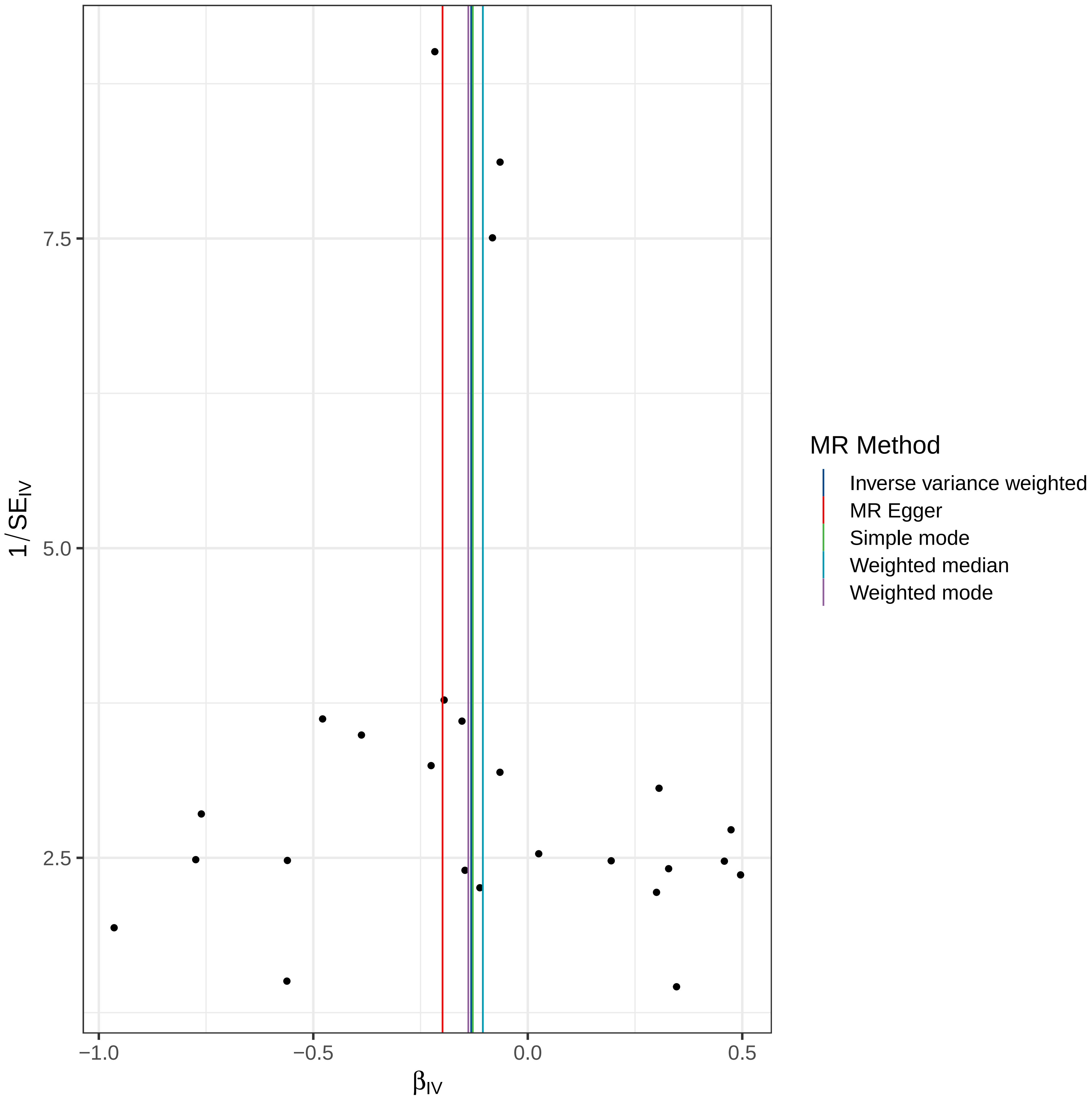

MR Method
Inverse variance weighted
MR Egger
Simple mode
Weighted median
Weighted mode
$1/SE_{IV}$
$\beta_{IV}$
7.5
5.0
2.5
−1.0
−0.5
0.0
0.5

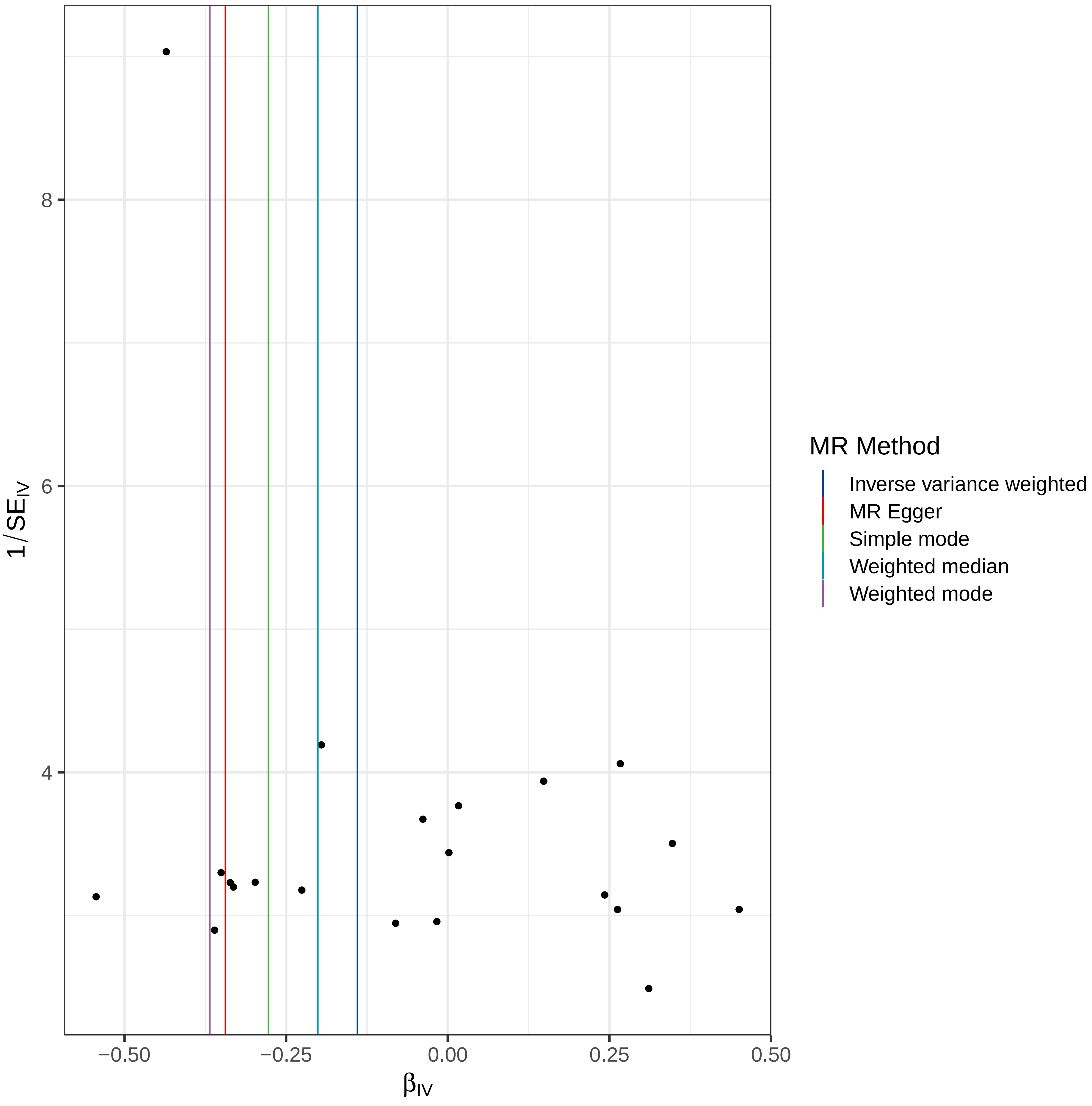

MR Method
Inverse variance weighted
MR Egger
Simple mode
Weighted median
Weighted mode
1/SE_IV
8
6
4
−0.50
−0.25
0.00
0.25
0.50
β_IV

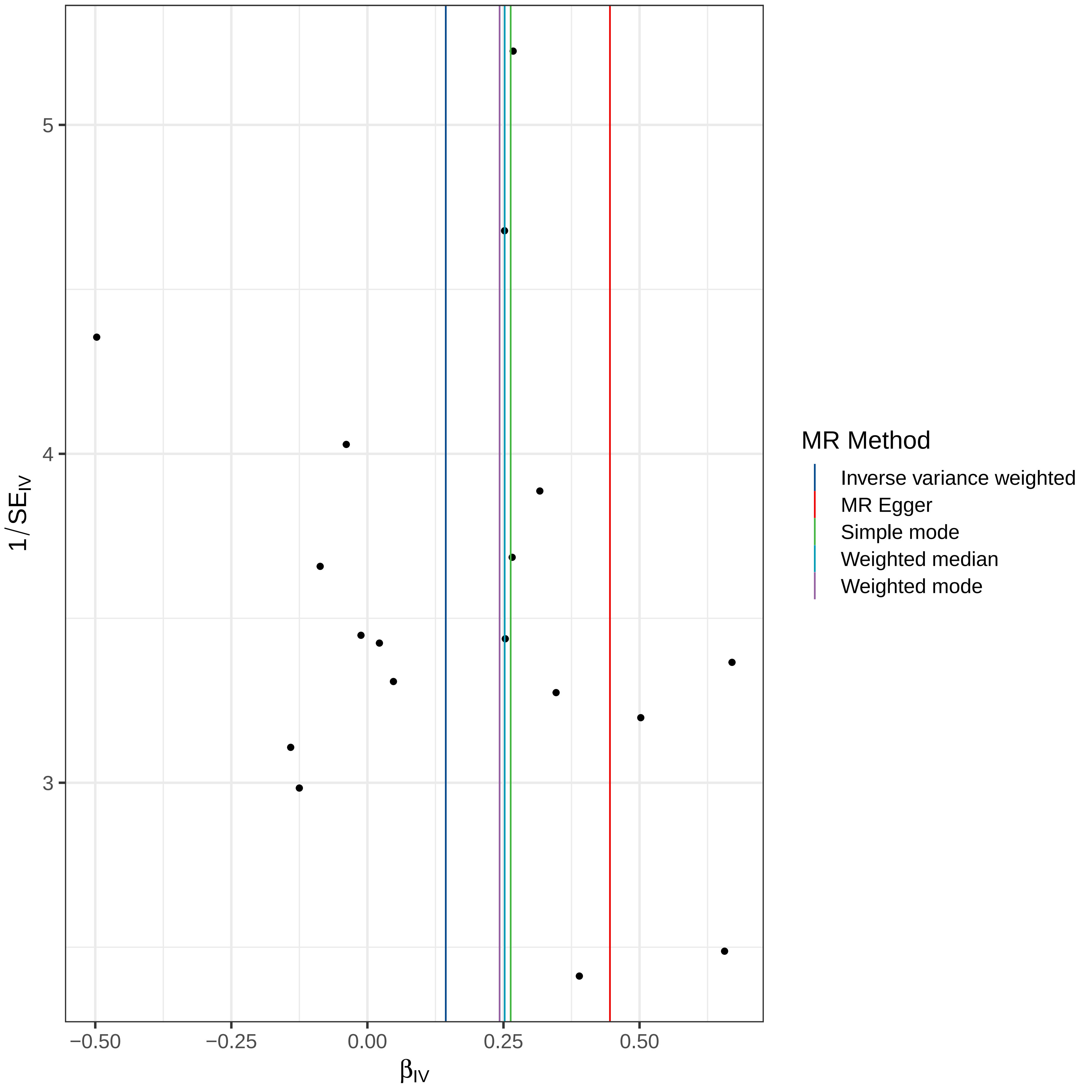

MR Method
Inverse variance weighted
MR Egger
Simple mode
Weighted median
Weighted mode
1/SE$_{IV}$
$\beta_{IV}$
5
4
3
−0.50
−0.25
0.00
0.25
0.50

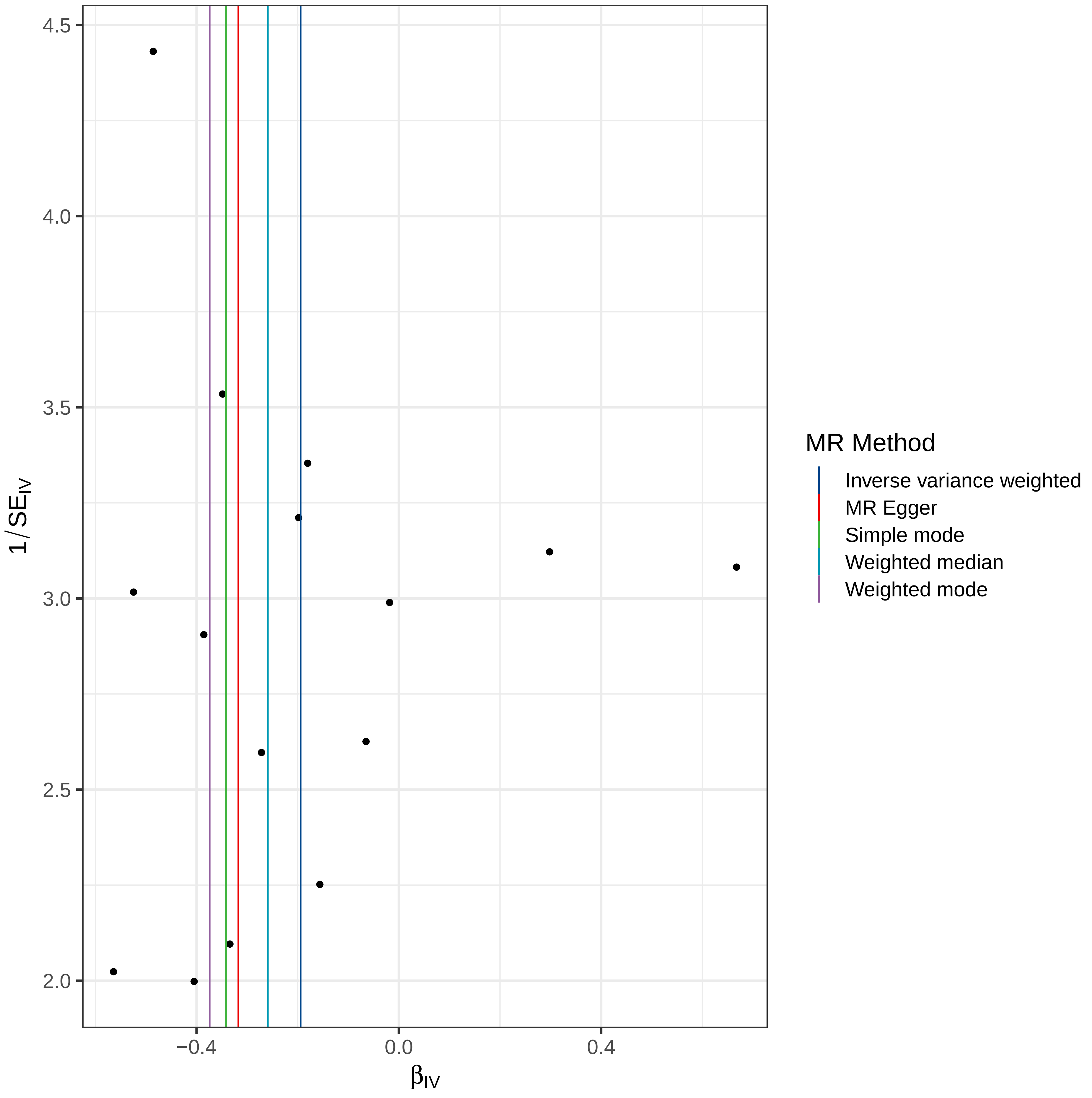

4.5
4.0
3.5
3.0
2.5
2.0
1/SE_IV
−0.4
0.0
0.4
β_IV
MR Method
Inverse variance weighted
MR Egger
Simple mode
Weighted median
Weighted mode

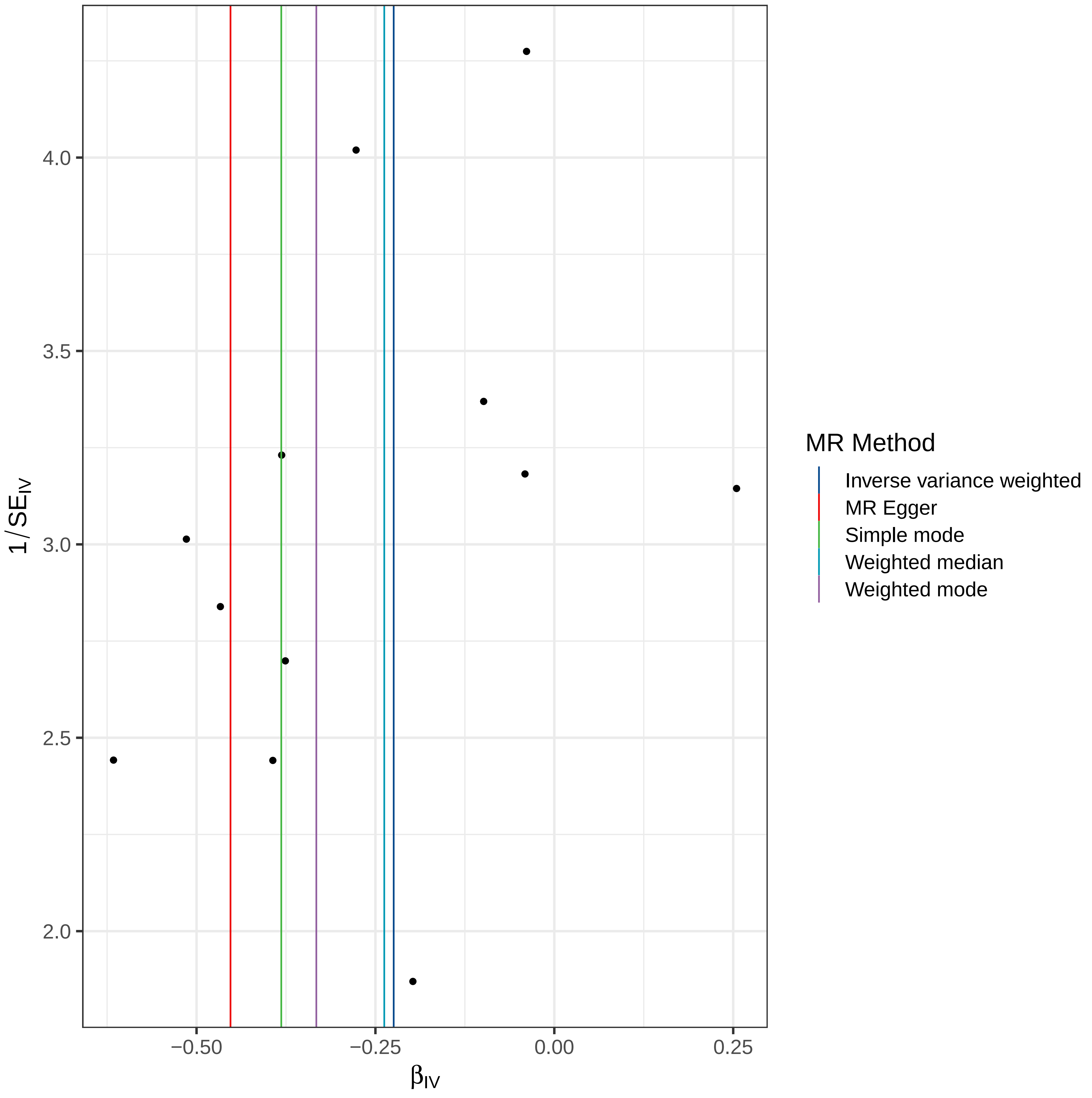

4.0
3.5
3.0
2.5
2.0
1/SE_IV
−0.50
−0.25
0.00
0.25
β_IV
MR Method
Inverse variance weighted
MR Egger
Simple mode
Weighted median
Weighted mode

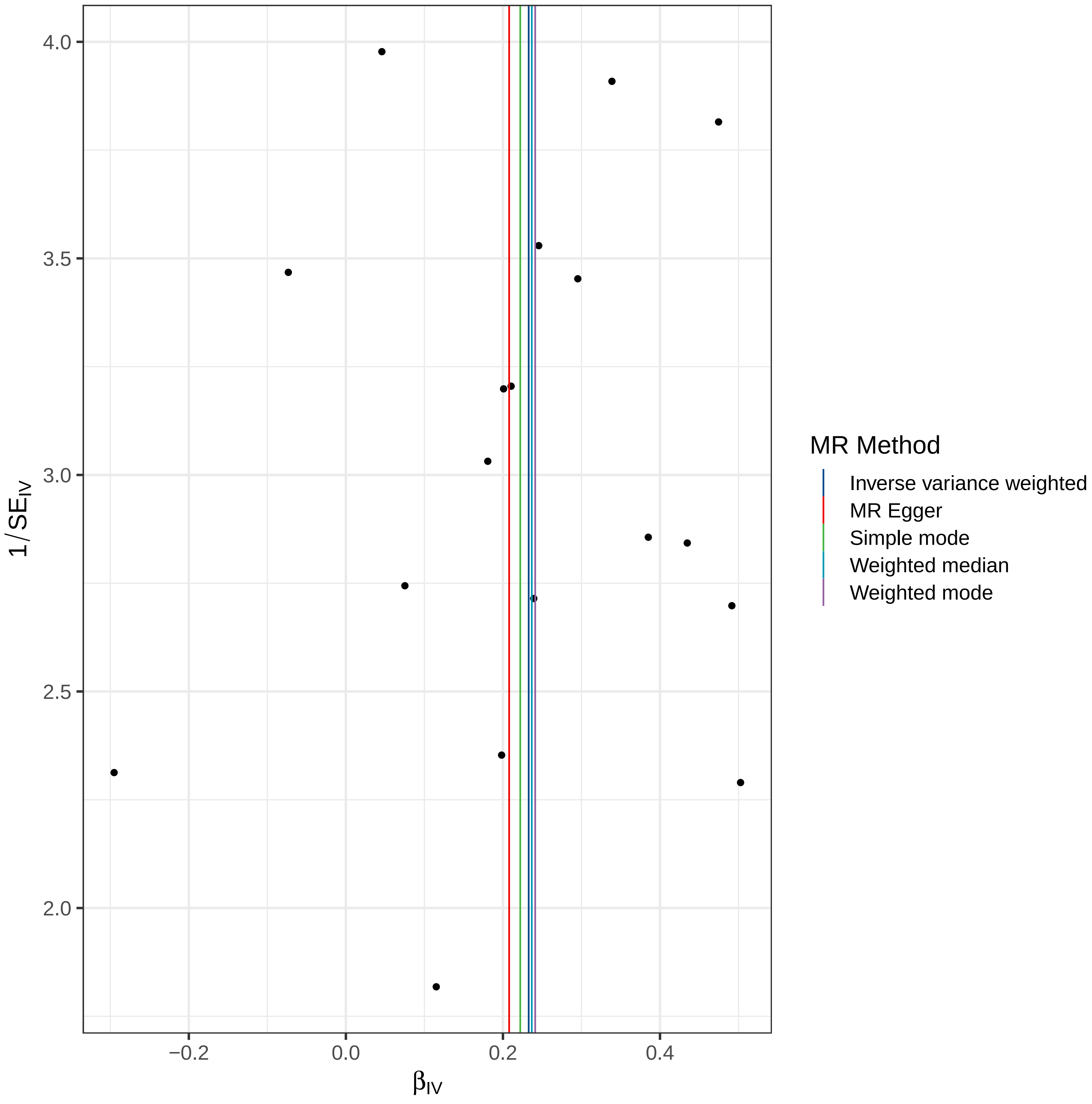

MR Method
Inverse variance weighted
MR Egger
Simple mode
Weighted median
Weighted mode
1/SE_IV
β_IV
4.0
3.5
3.0
2.5
2.0
−0.2
0.0
0.2
0.4

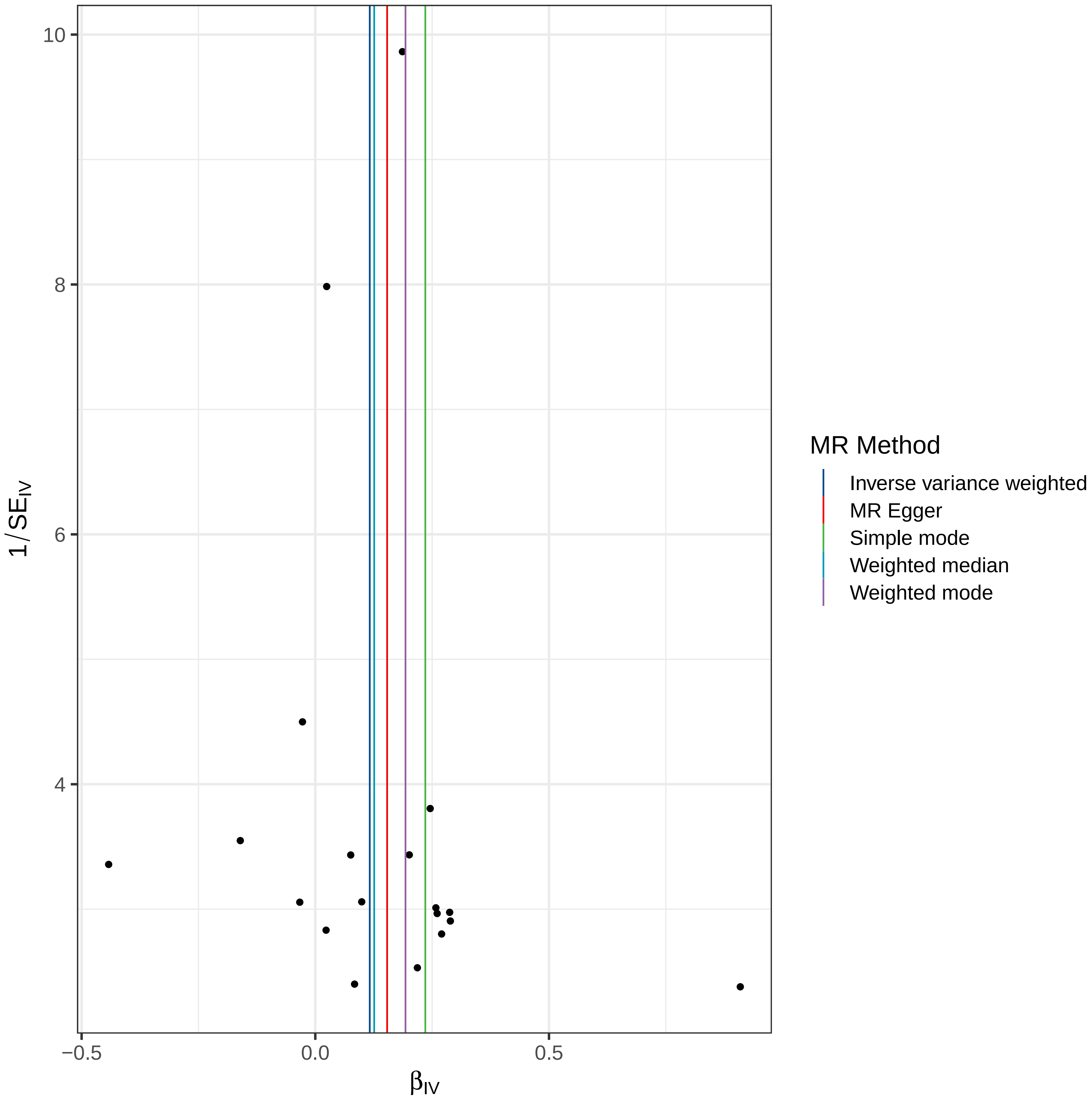

10
8
6
4
1/SE_IV
−0.5
0.0
0.5
β_IV
MR Method
Inverse variance weighted
MR Egger
Simple mode
Weighted median
Weighted mode

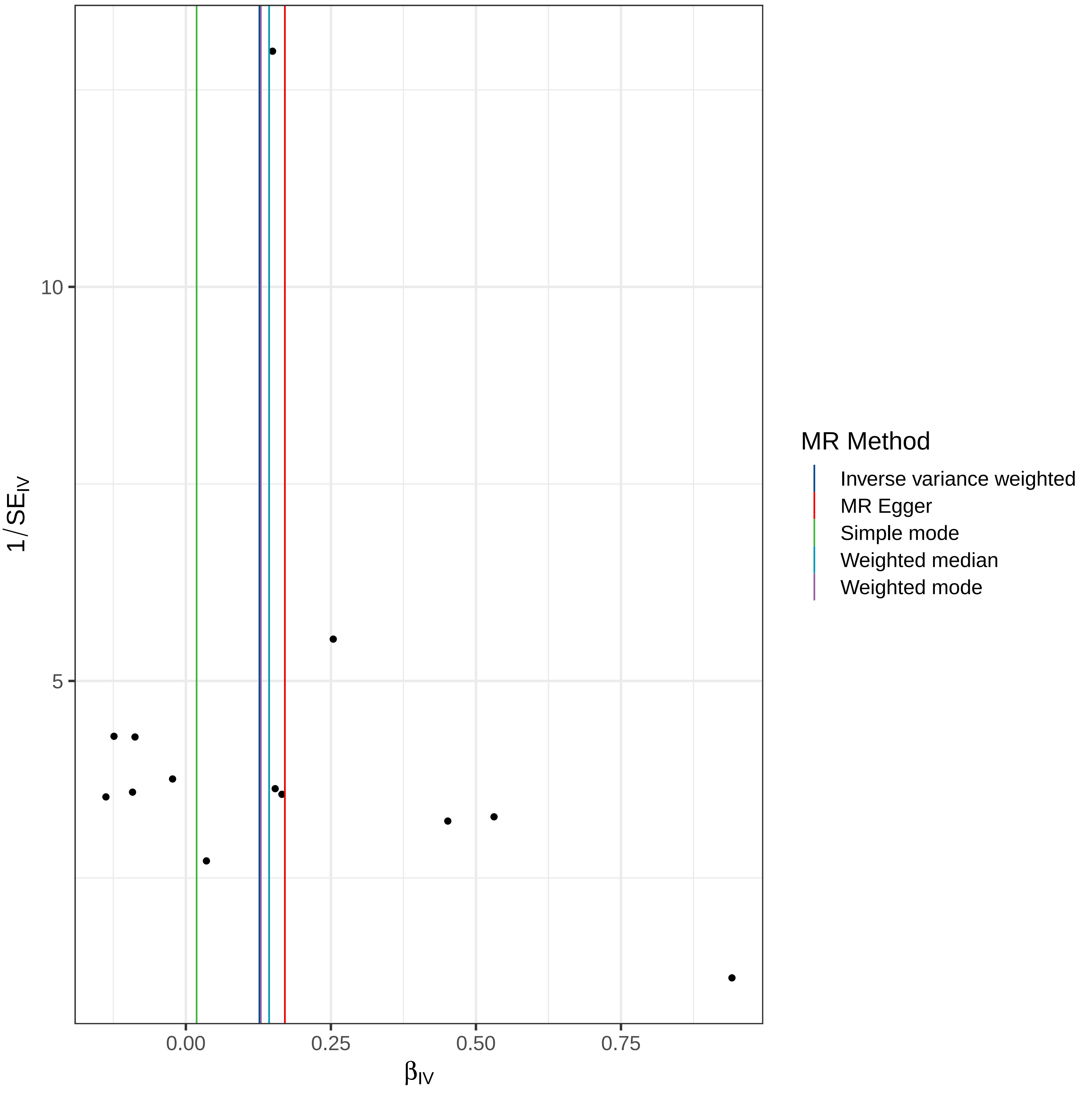

10
5
0.00
0.25
0.50
0.75
β_IV
1/SE_IV
MR Method
Inverse variance weighted
MR Egger
Simple mode
Weighted median
Weighted mode

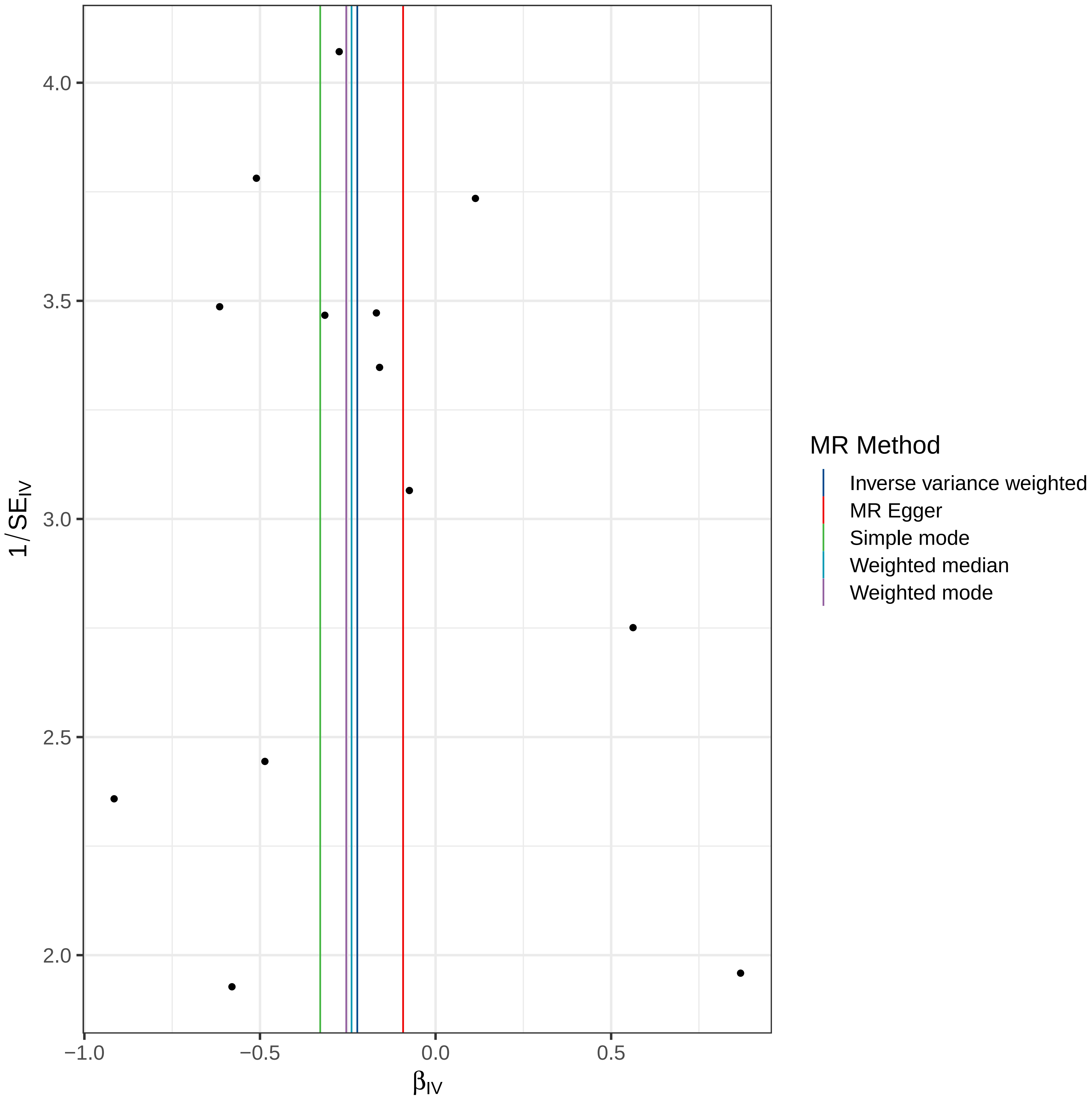

4.0
3.5
3.0
2.5
2.0
$1/SE_{IV}$
−1.0
−0.5
0.0
0.5
$\beta_{IV}$
MR Method
Inverse variance weighted
MR Egger
Simple mode
Weighted median
Weighted mode

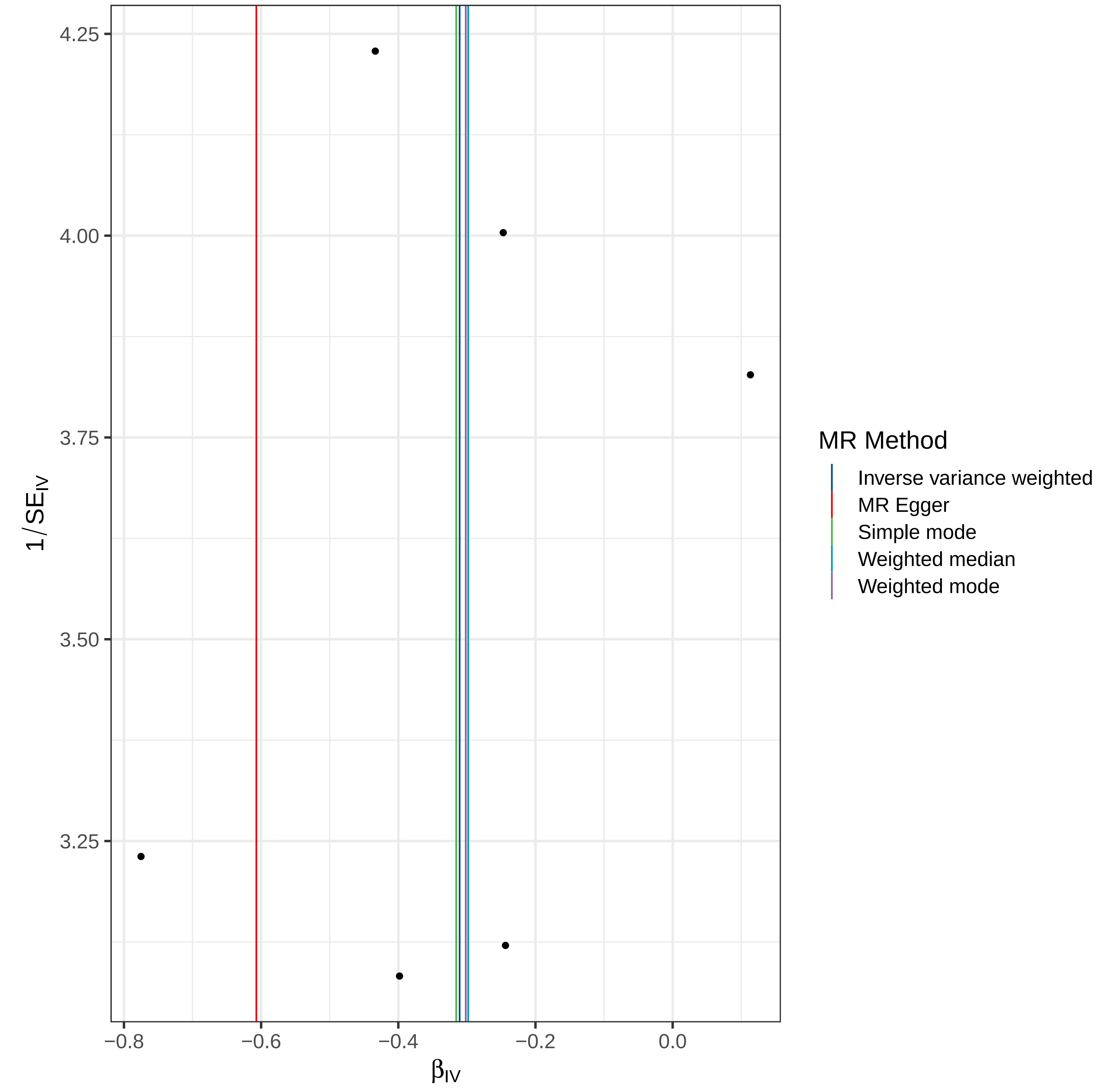

4.25
4.00
3.75
3.50
3.25
$1/SE_{IV}$
−0.8
−0.6
−0.4
−0.2
0.0
$\beta_{IV}$
MR Method
Inverse variance weighted
MR Egger
Simple mode
Weighted median
Weighted mode

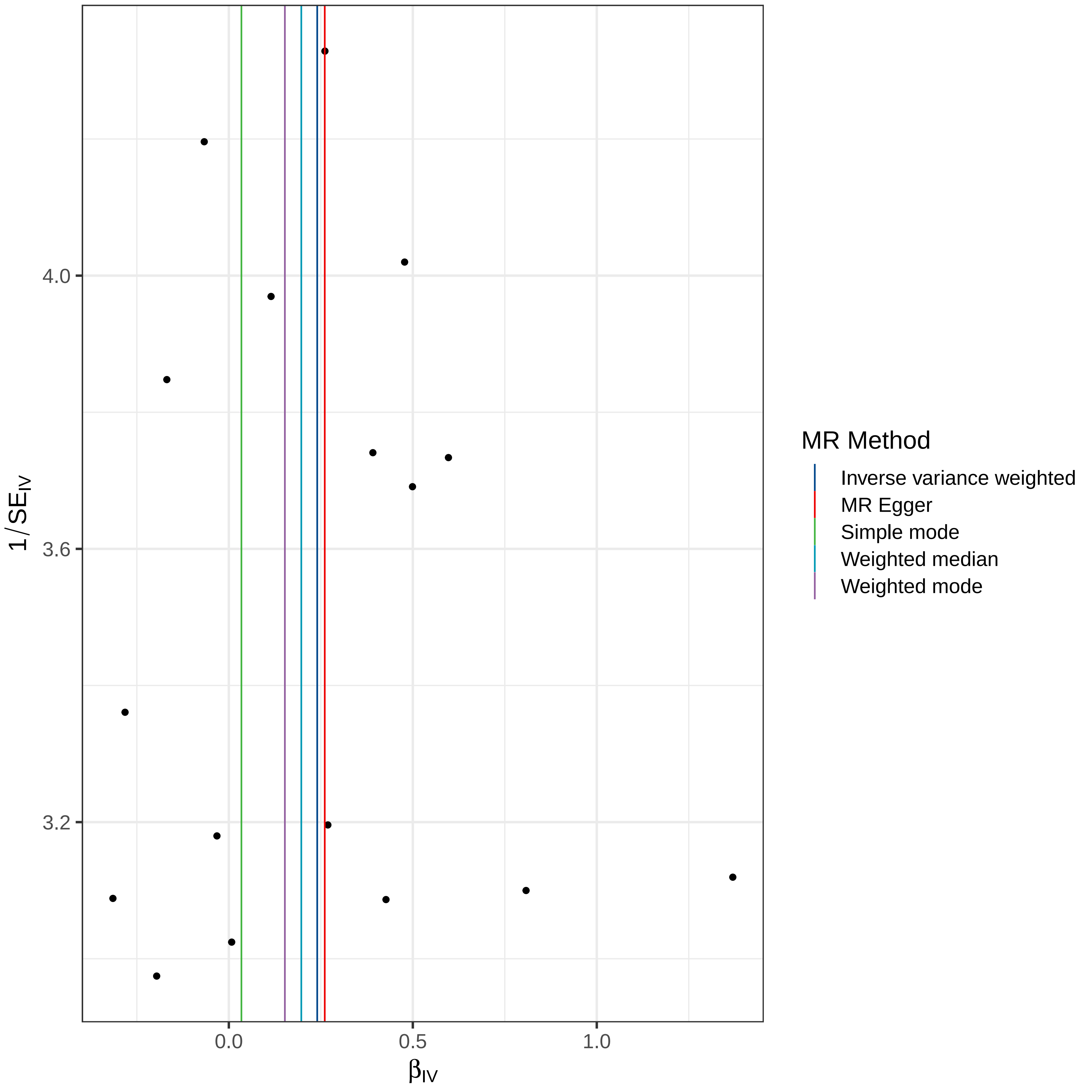

4.0
3.6
3.2
$1/SE_{IV}$
0.0
0.5
1.0
$\beta_{IV}$
MR Method
Inverse variance weighted
MR Egger
Simple mode
Weighted median
Weighted mode

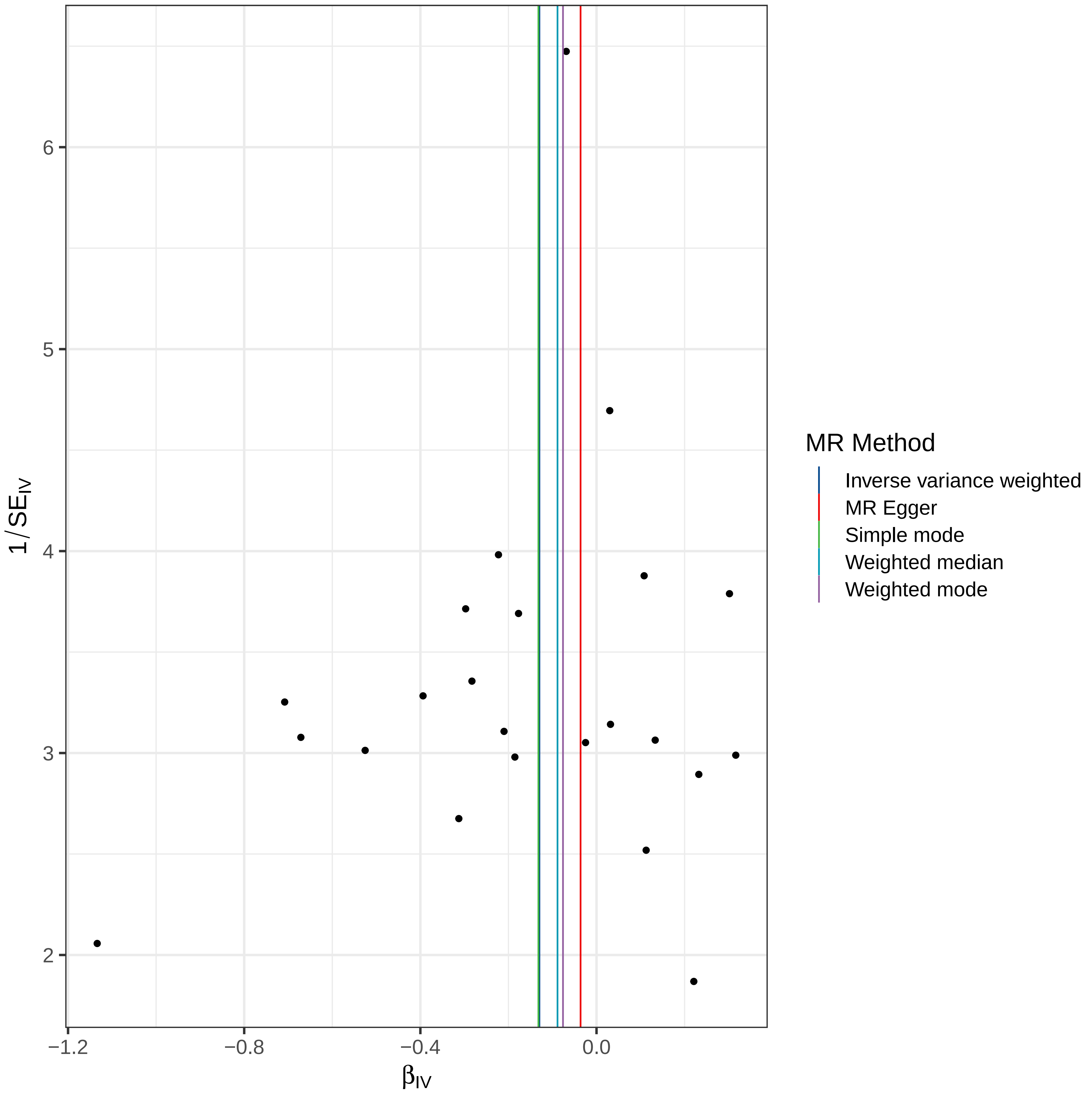

6
5
4
3
2
1/SE_IV
−1.2
−0.8
−0.4
0.0
β_IV
MR Method
Inverse variance weighted
MR Egger
Simple mode
Weighted median
Weighted mode

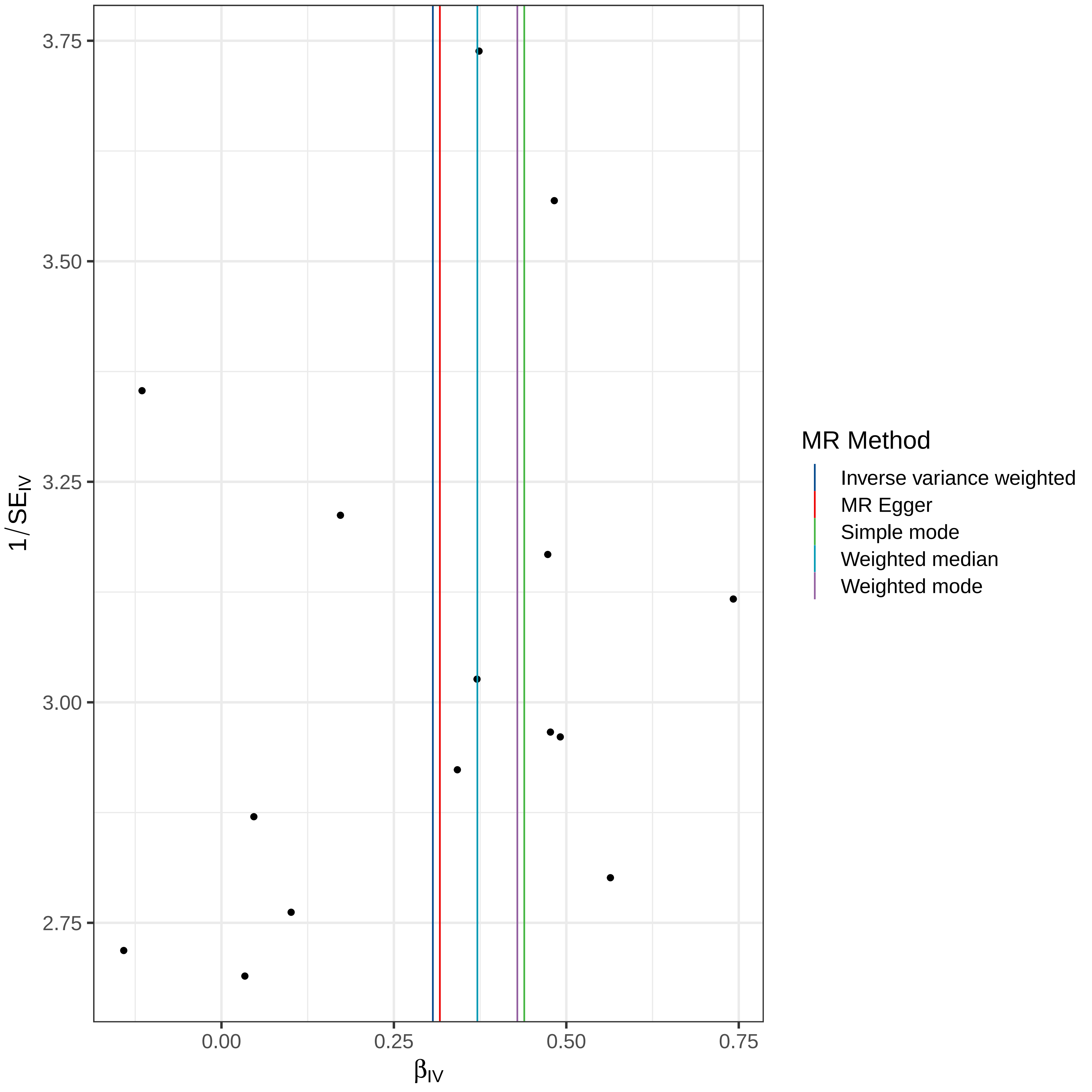

3.75
3.50
3.25
3.00
2.75
1/SE_IV
0.00
0.25
0.50
0.75
β_IV
MR Method
Inverse variance weighted
MR Egger
Simple mode
Weighted median
Weighted mode

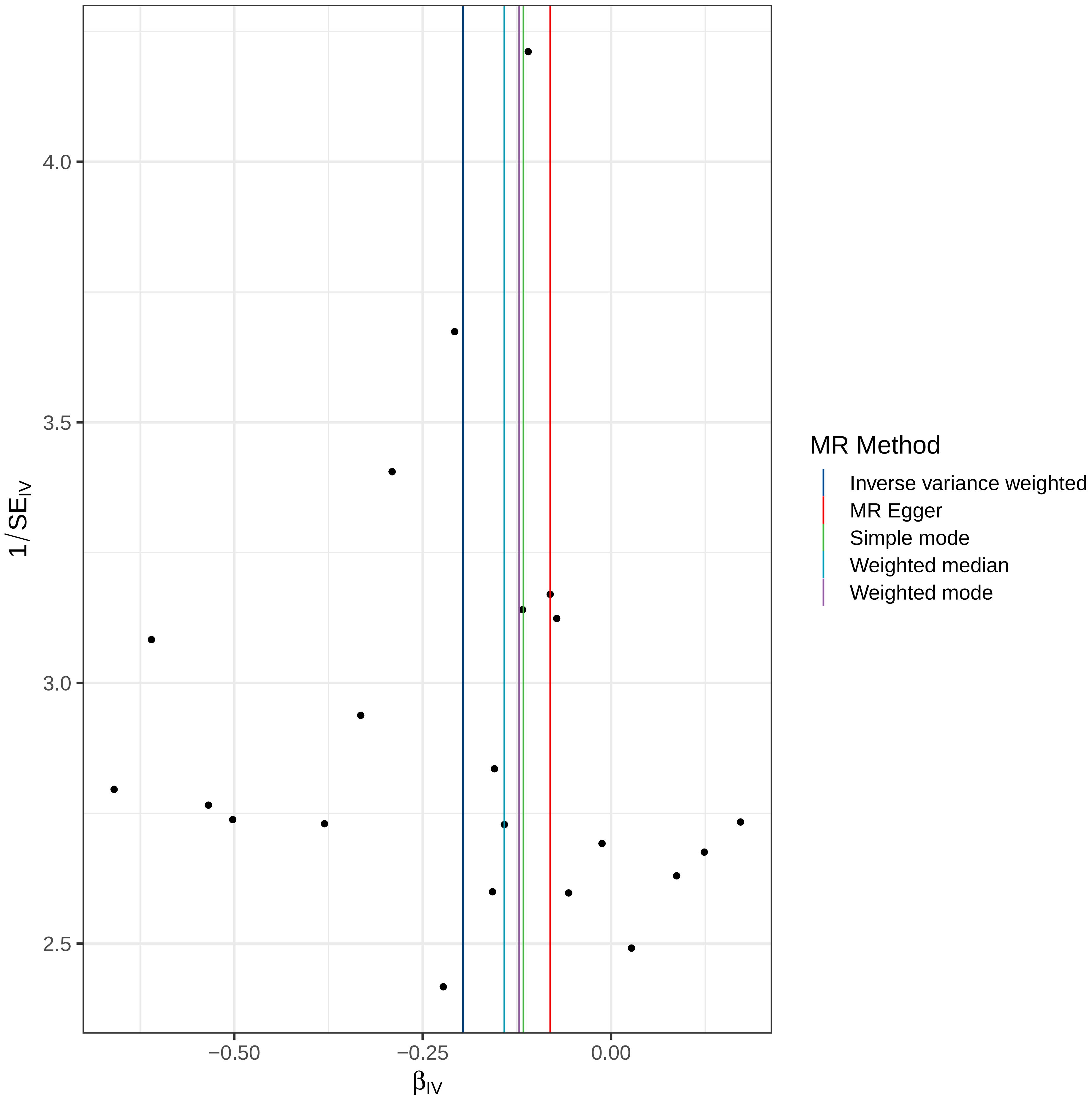

4.0
3.5
3.0
2.5
1/SE_IV
−0.50
−0.25
0.00
β_IV
MR Method
Inverse variance weighted
MR Egger
Simple mode
Weighted median
Weighted mode

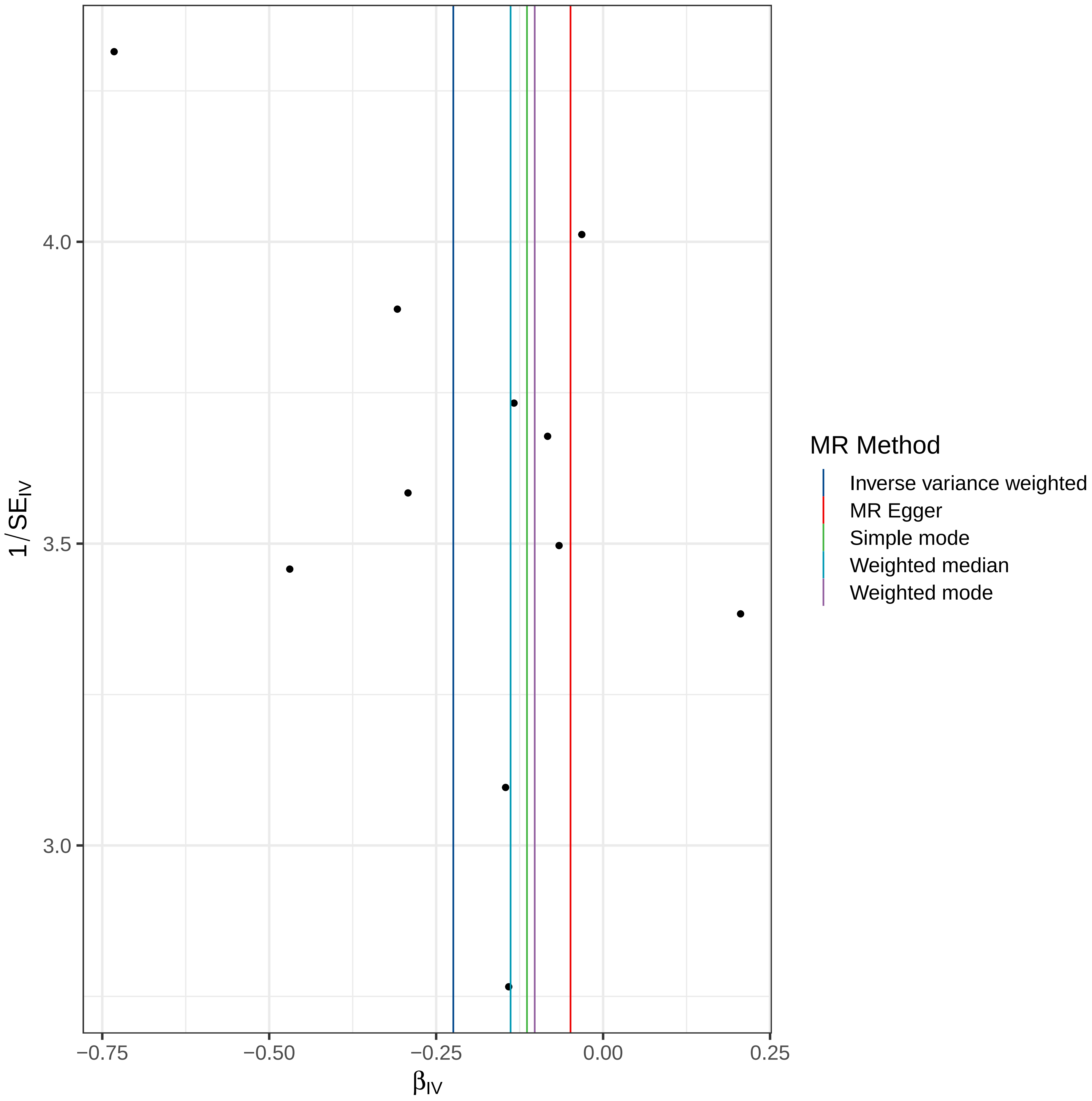

MR Method
Inverse variance weighted
MR Egger
Simple mode
Weighted median
Weighted mode
4.0
3.5
3.0
$1/SE_{IV}$
−0.75
−0.50
−0.25
0.00
0.25
$\beta_{IV}$

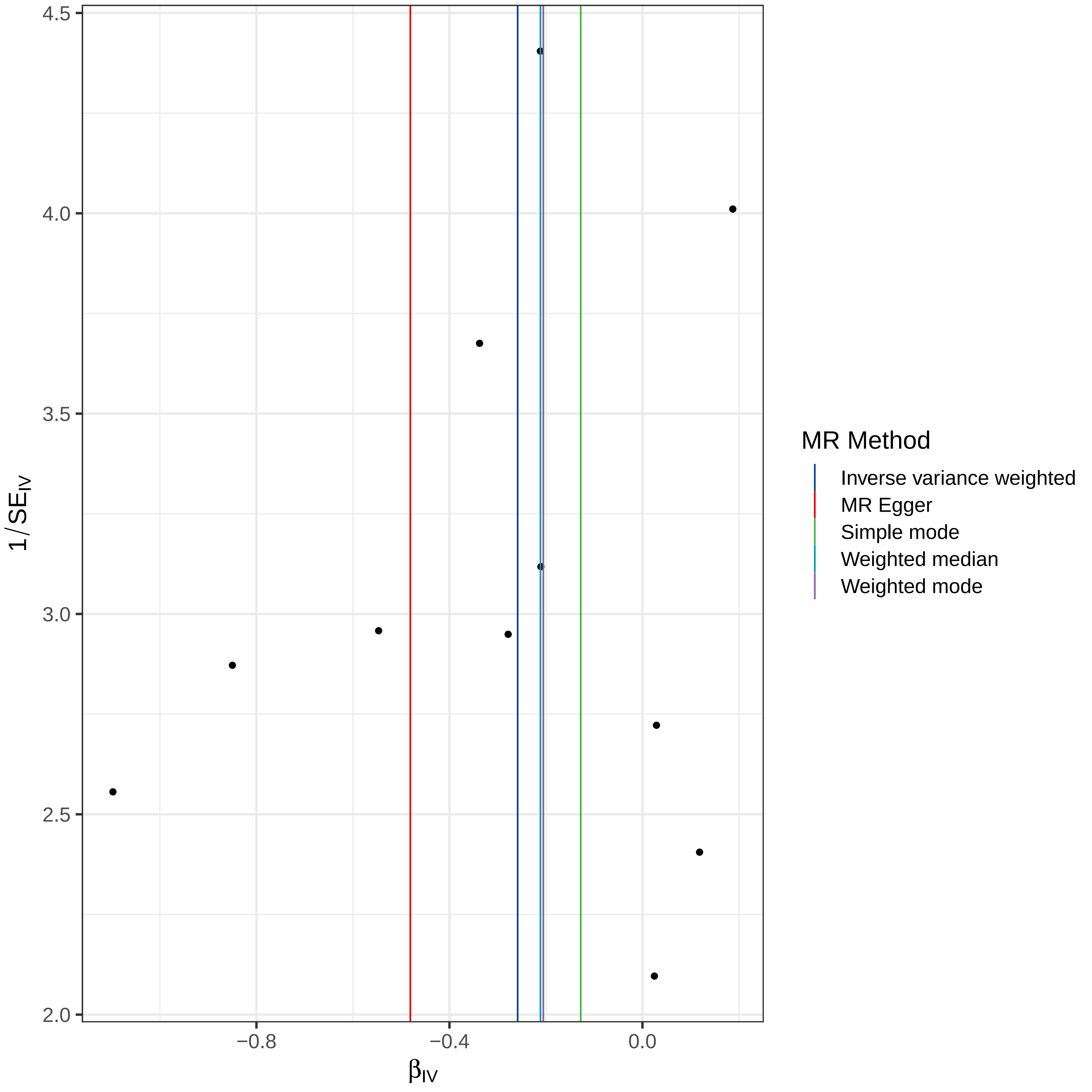
4.5
4.0
3.5
3.0
2.5
2.0
−0.8
−0.4
0.0
$1/SE_{IV}$
$\beta_{IV}$
MR Method
Inverse variance weighted
MR Egger
Simple mode
Weighted median
Weighted mode

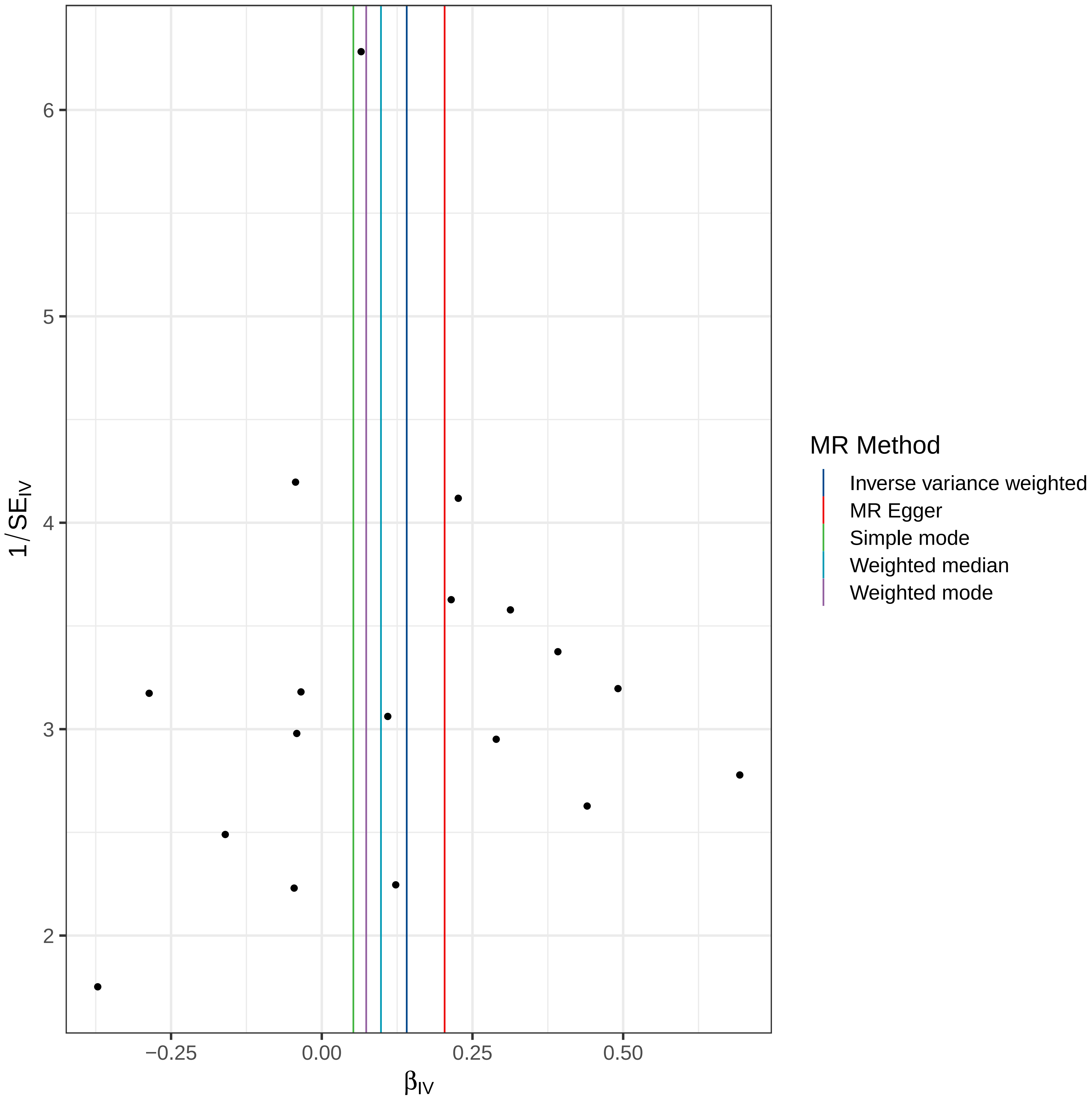

6
5
4
3
2
1/SE$_{IV}$
−0.25
0.00
0.25
0.50
β$_{IV}$
MR Method
Inverse variance weighted
MR Egger
Simple mode
Weighted median
Weighted mode

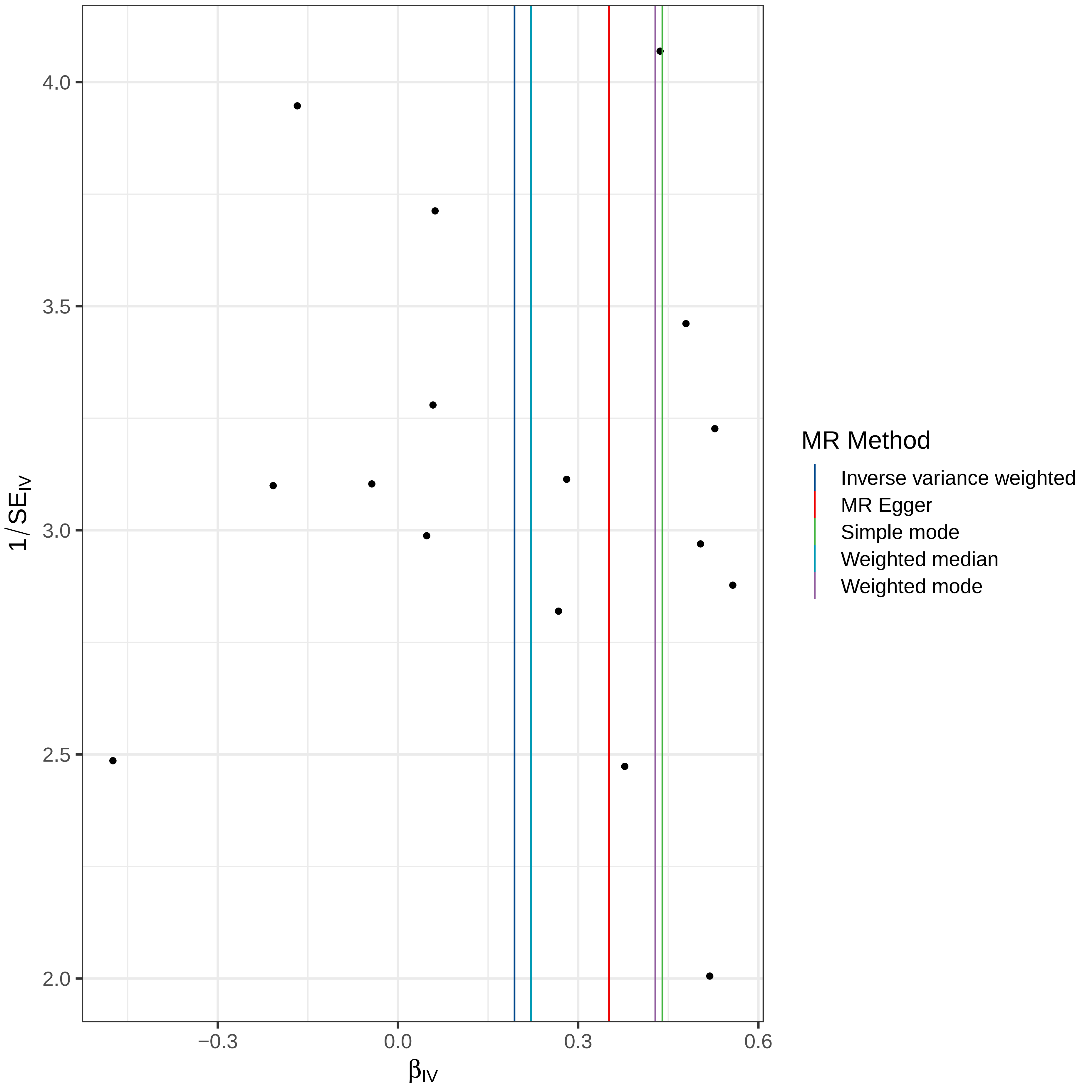

4.0
3.5
3.0
2.5
2.0
1/SE_IV
−0.3
0.0
0.3
0.6
β_IV
MR Method
Inverse variance weighted
MR Egger
Simple mode
Weighted median
Weighted mode

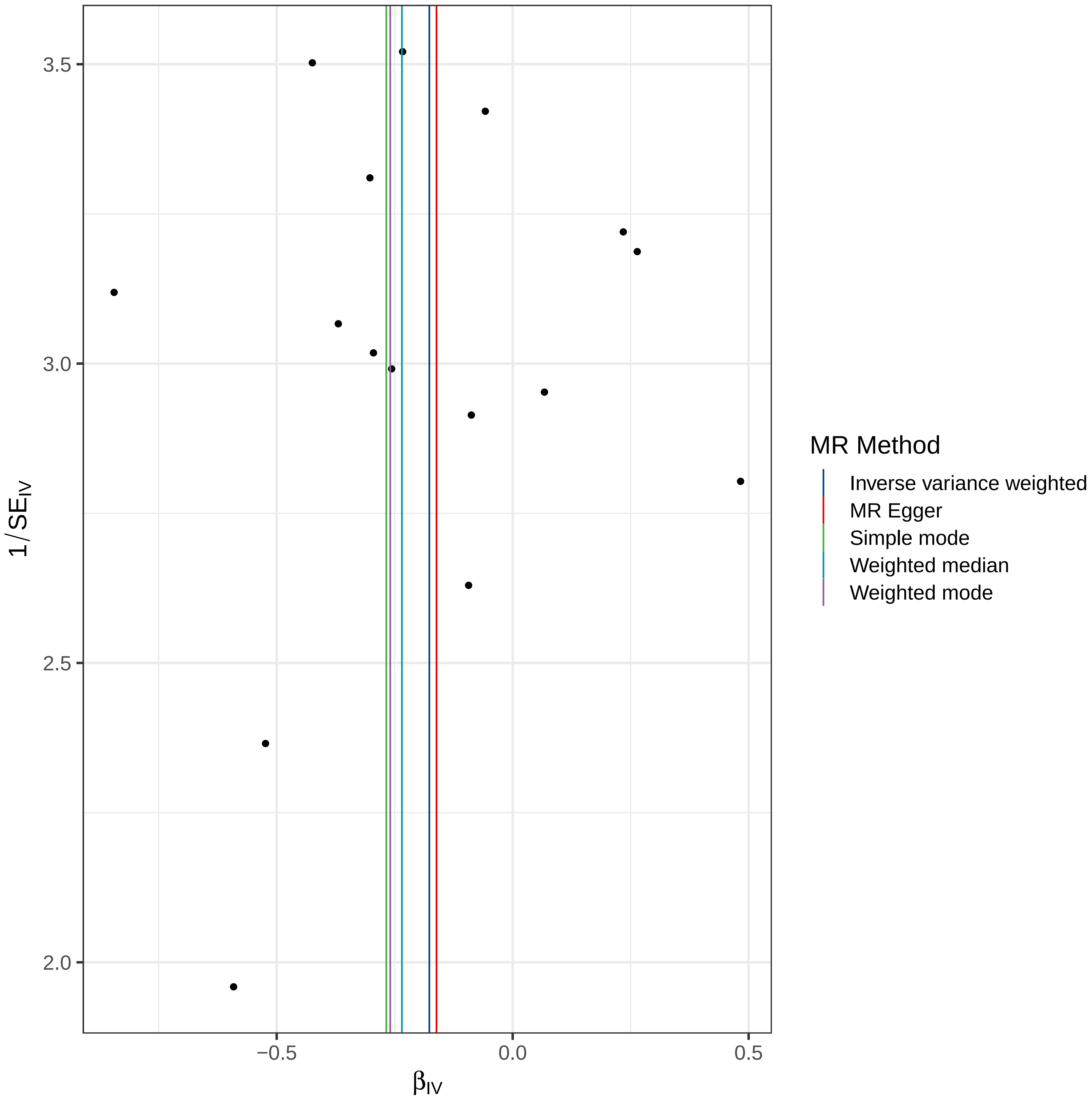

3.5
3.0
2.5
2.0
1/SE_IV
−0.5
0.0
0.5
β_IV
MR Method
Inverse variance weighted
MR Egger
Simple mode
Weighted median
Weighted mode

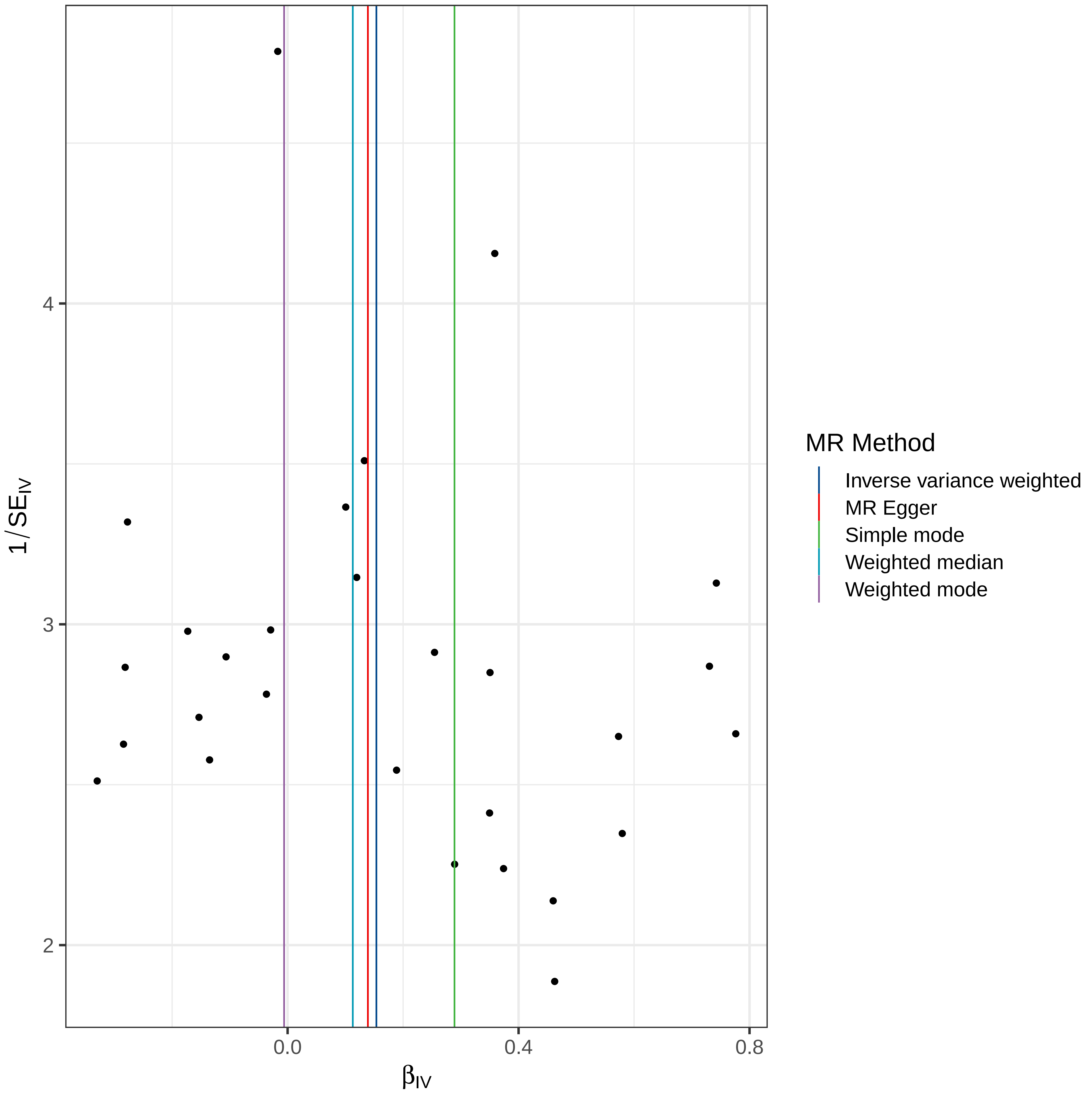

MR Method
Inverse variance weighted
MR Egger
Simple mode
Weighted median
Weighted mode
$1/SE_{IV}$
$\beta_{IV}$
2
3
4
0.0
0.4
0.8

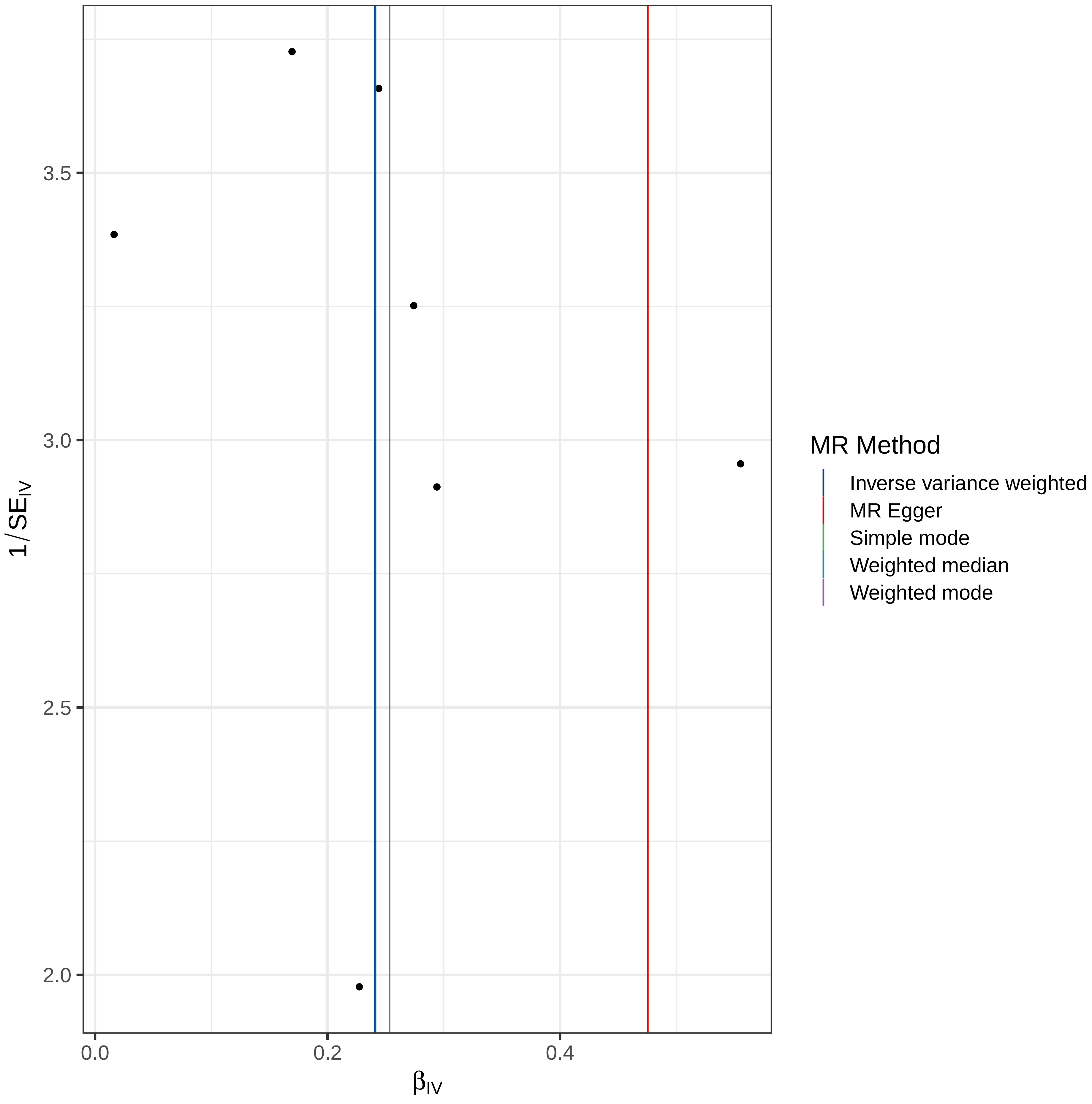

1/SE_IV
3.5
3.0
2.5
2.0
0.0
0.2
0.4
β_IV
MR Method
Inverse variance weighted
MR Egger
Simple mode
Weighted median
Weighted mode

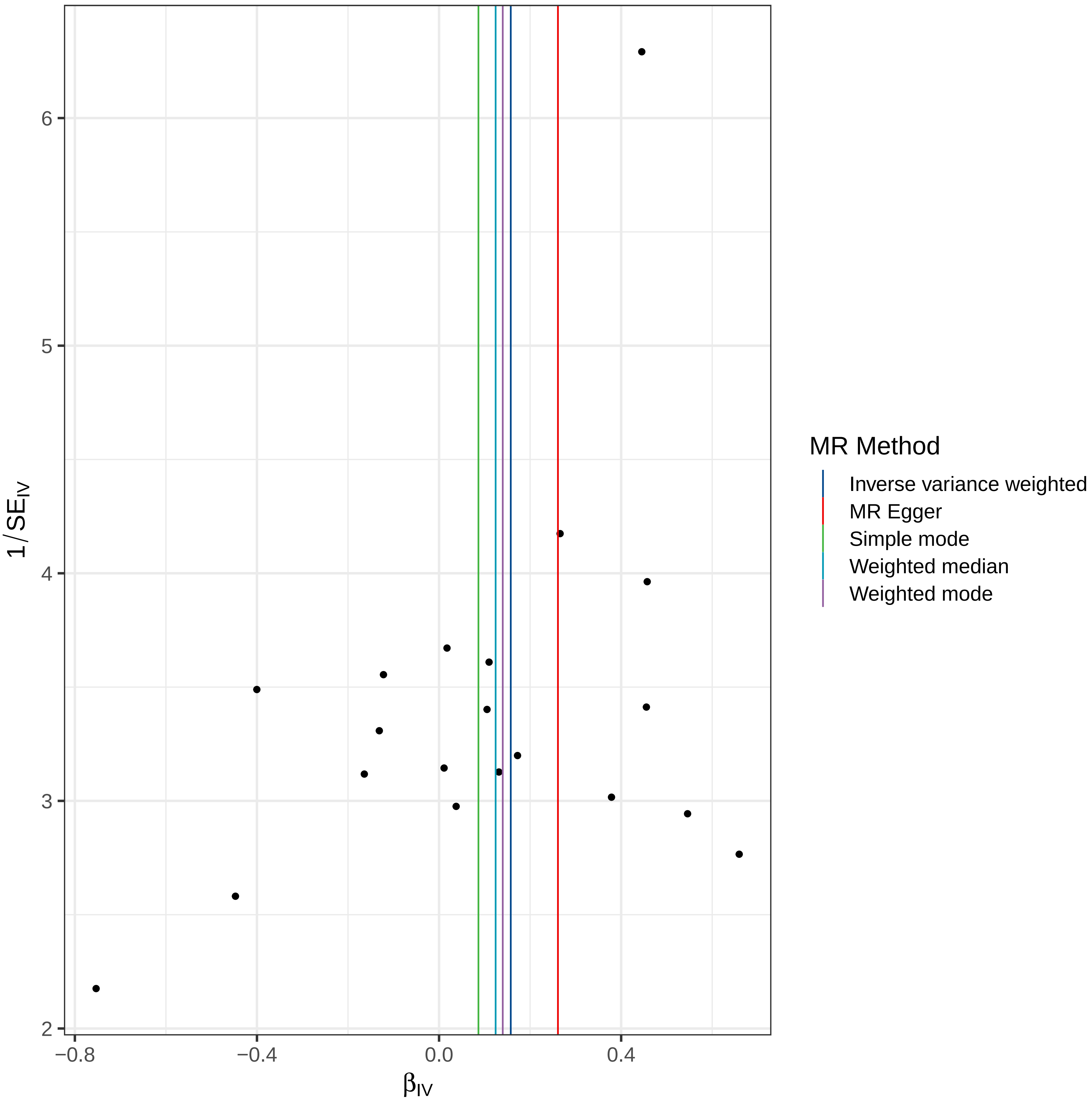

6
5
4
3
2
1/SE_IV
−0.8
−0.4
0.0
0.4
β_IV
MR Method
Inverse variance weighted
MR Egger
Simple mode
Weighted median
Weighted mode

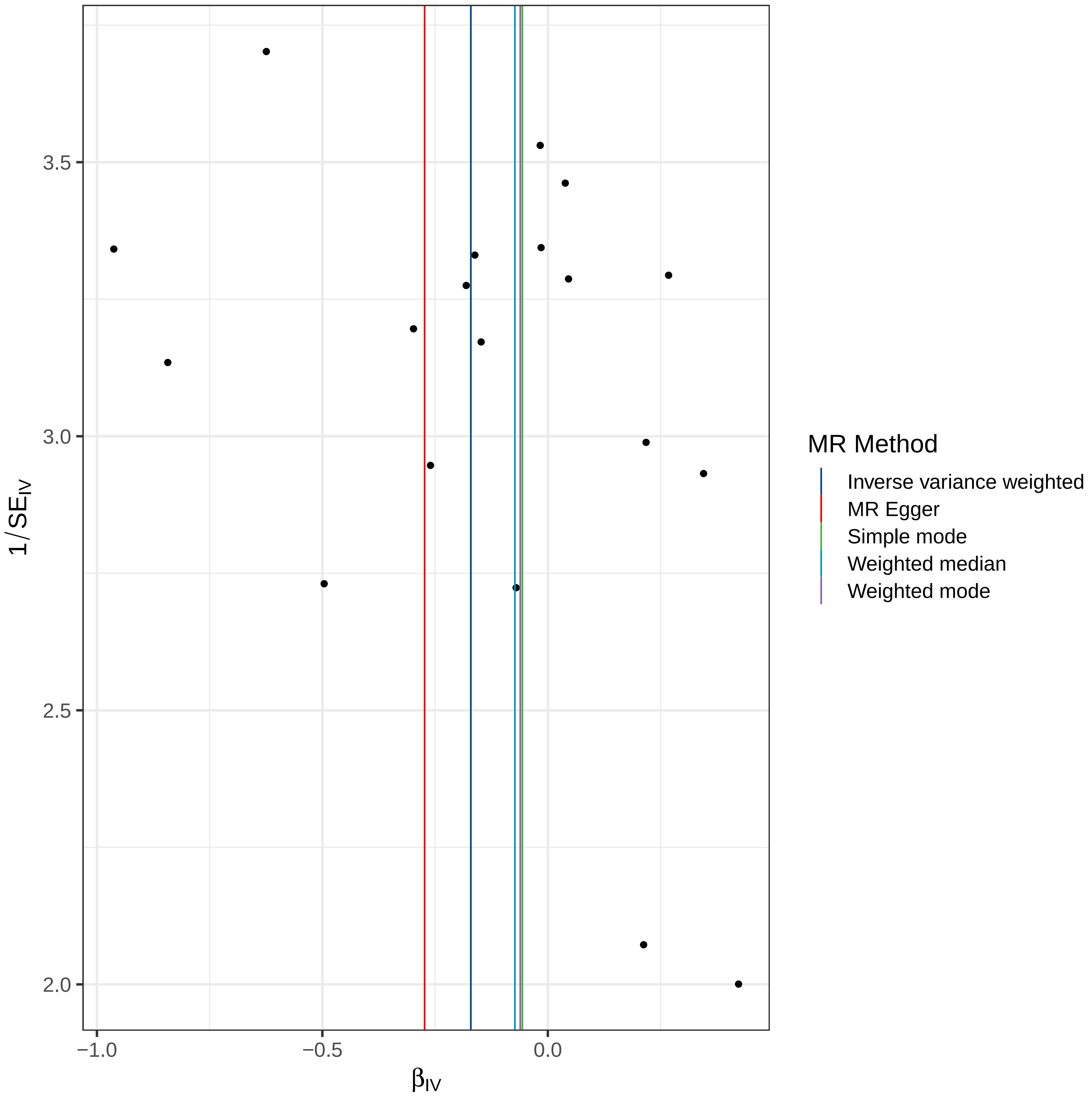

3.5
3.0
2.5
2.0
−1.0
−0.5
0.0
$1/SE_{IV}$
$\beta_{IV}$
MR Method
Inverse variance weighted
MR Egger
Simple mode
Weighted median
Weighted mode

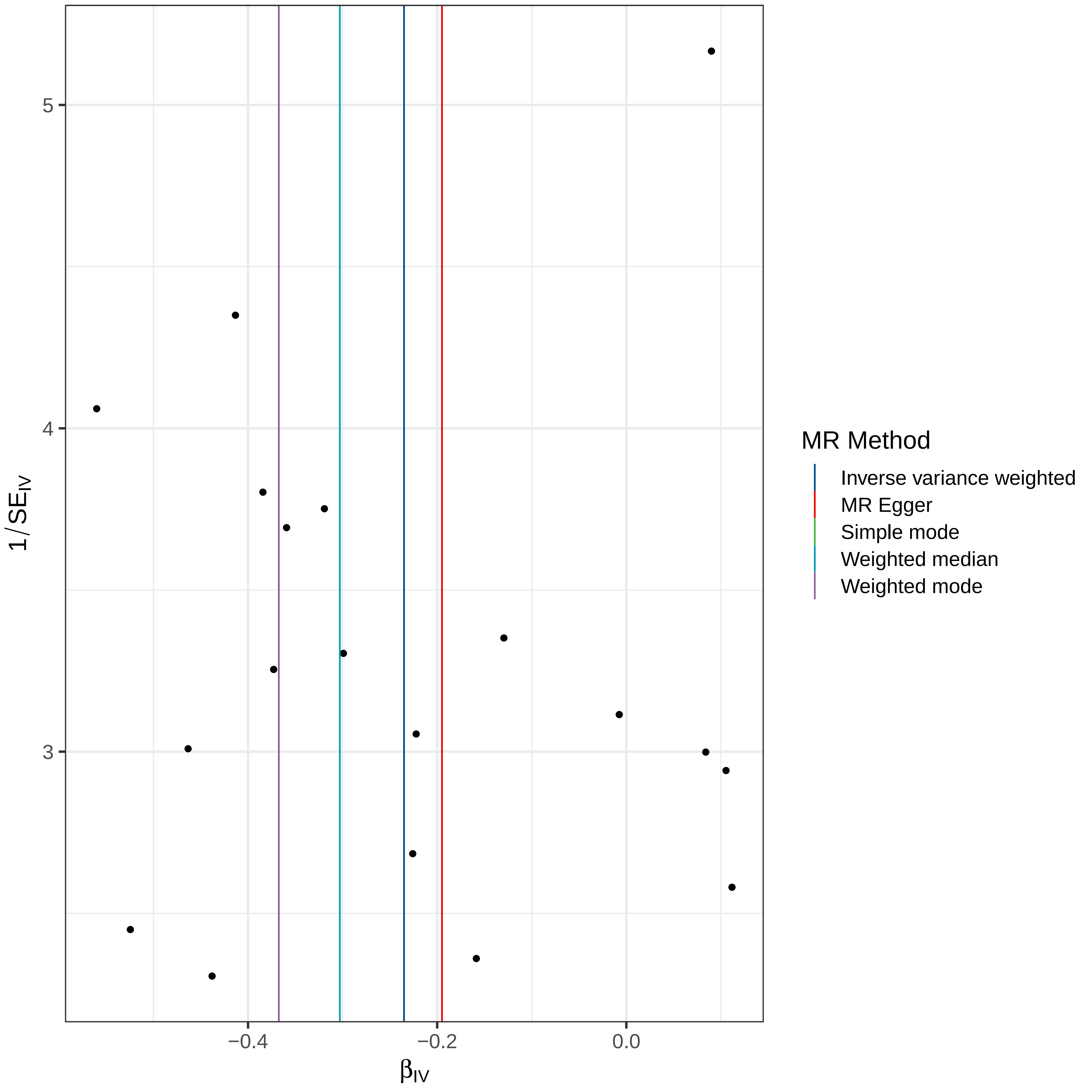

5
4
3
1/SE$_{IV}$
−0.4
−0.2
0.0
β$_{IV}$
MR Method
Inverse variance weighted
MR Egger
Simple mode
Weighted median
Weighted mode

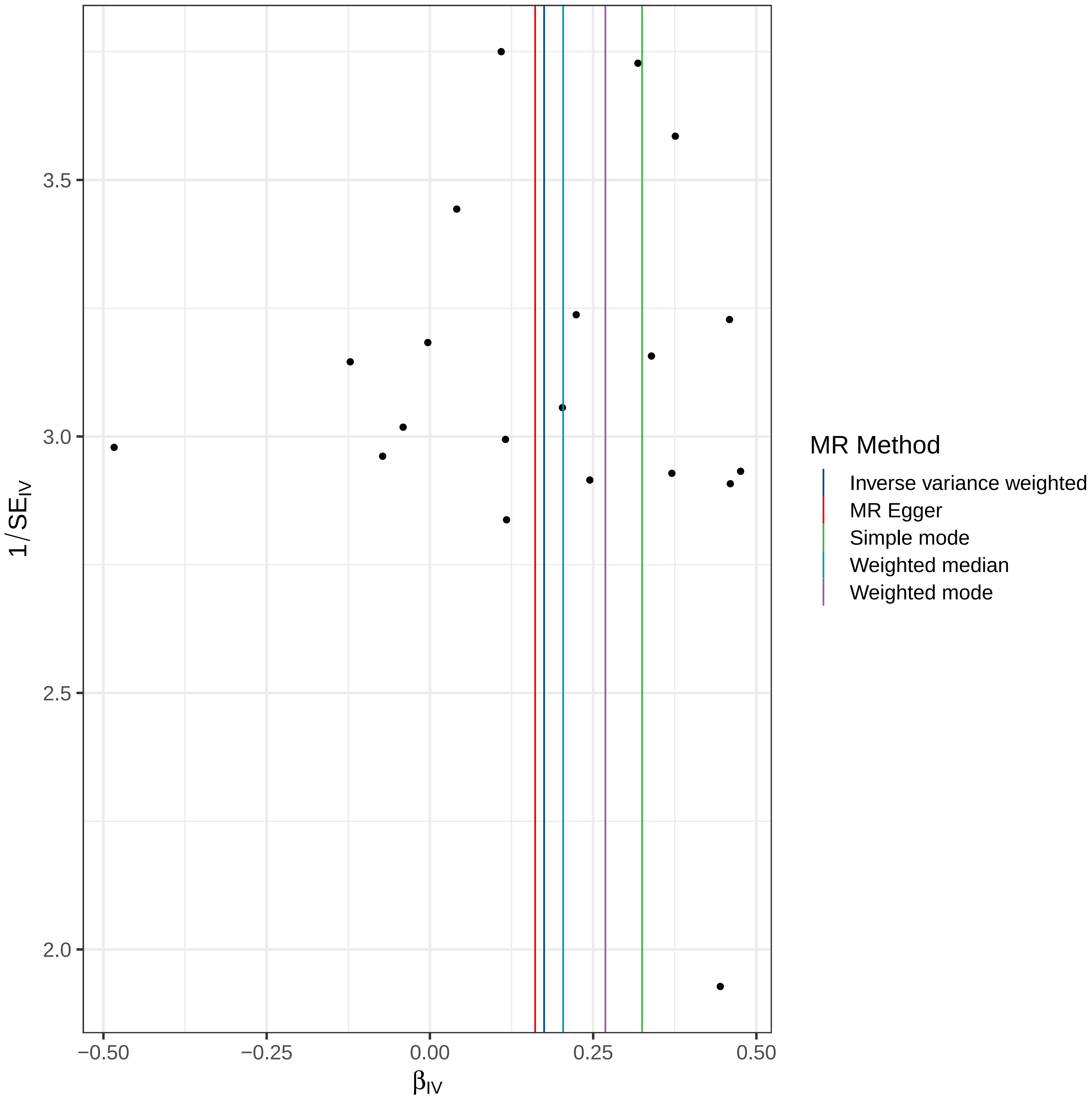

3.5
3.0
2.5
2.0
1/SE_IV
−0.50
−0.25
0.00
0.25
0.50
β_IV
MR Method
Inverse variance weighted
MR Egger
Simple mode
Weighted median
Weighted mode

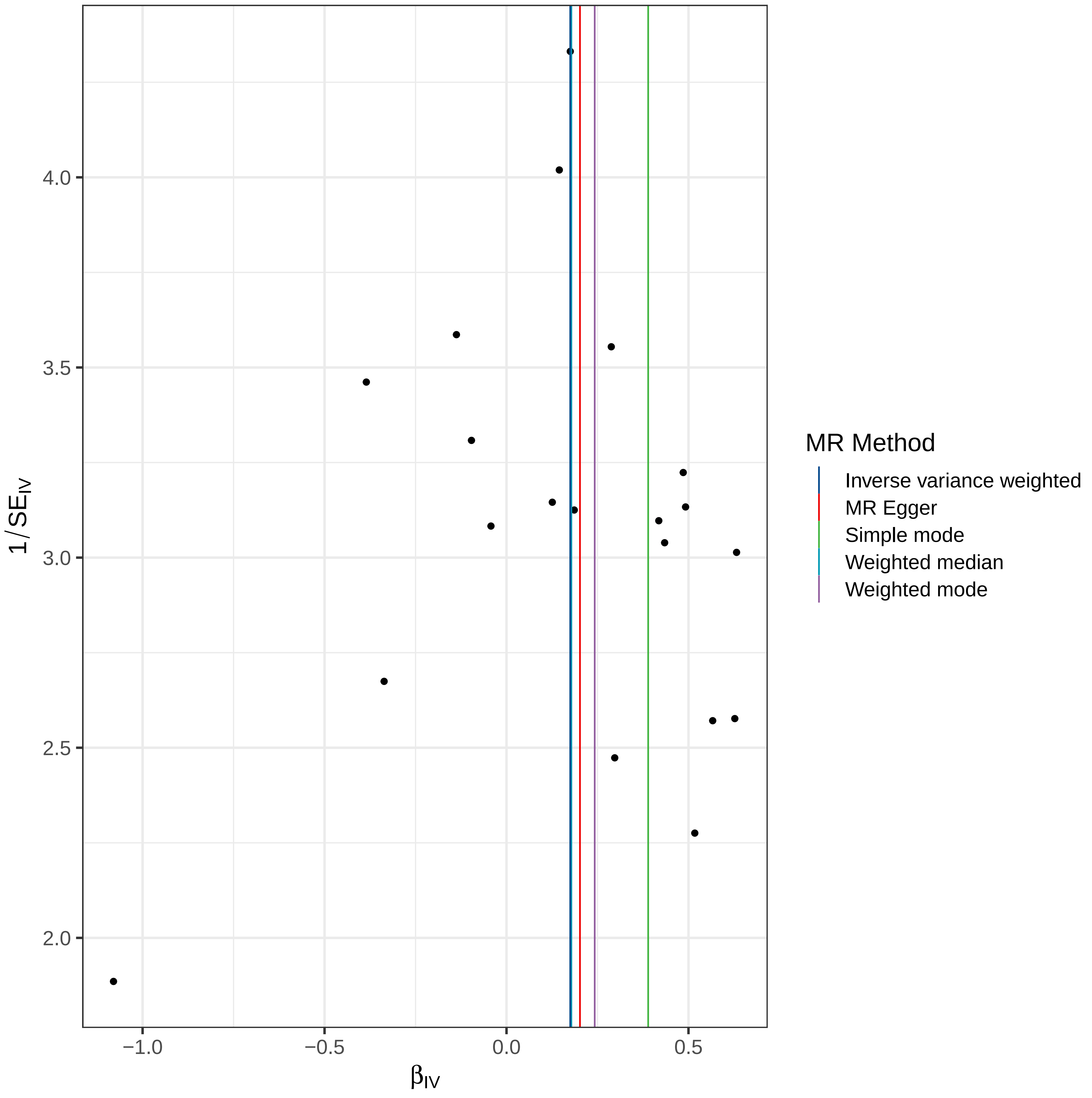

4.0
3.5
3.0
2.5
2.0
1/SE_IV
−1.0
−0.5
0.0
0.5
β_IV
MR Method
Inverse variance weighted
MR Egger
Simple mode
Weighted median
Weighted mode

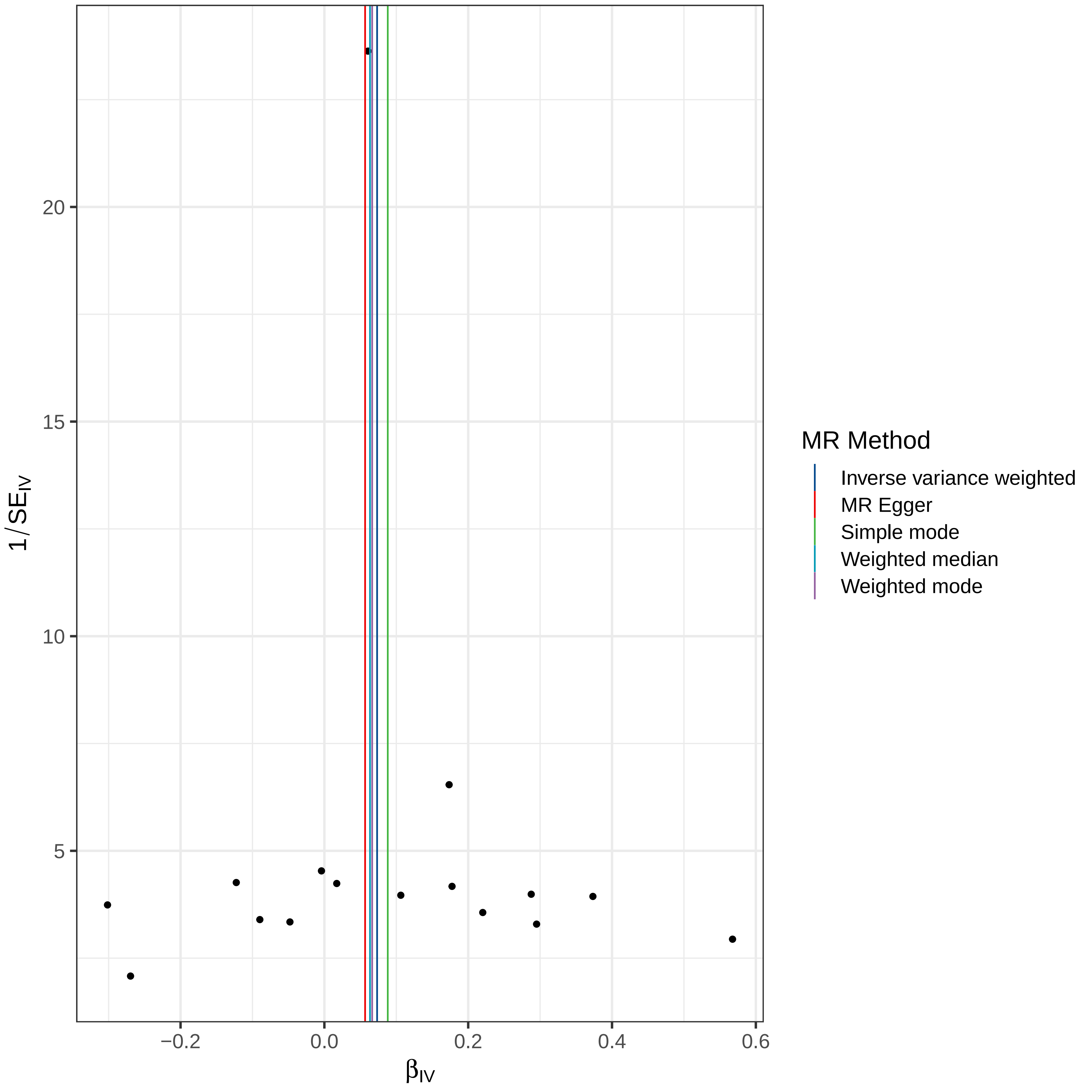

MR Method
Inverse variance weighted
MR Egger
Simple mode
Weighted median
Weighted mode
1/SE_IV
β_IV
20
15
10
5
−0.2
0.0
0.2
0.4
0.6

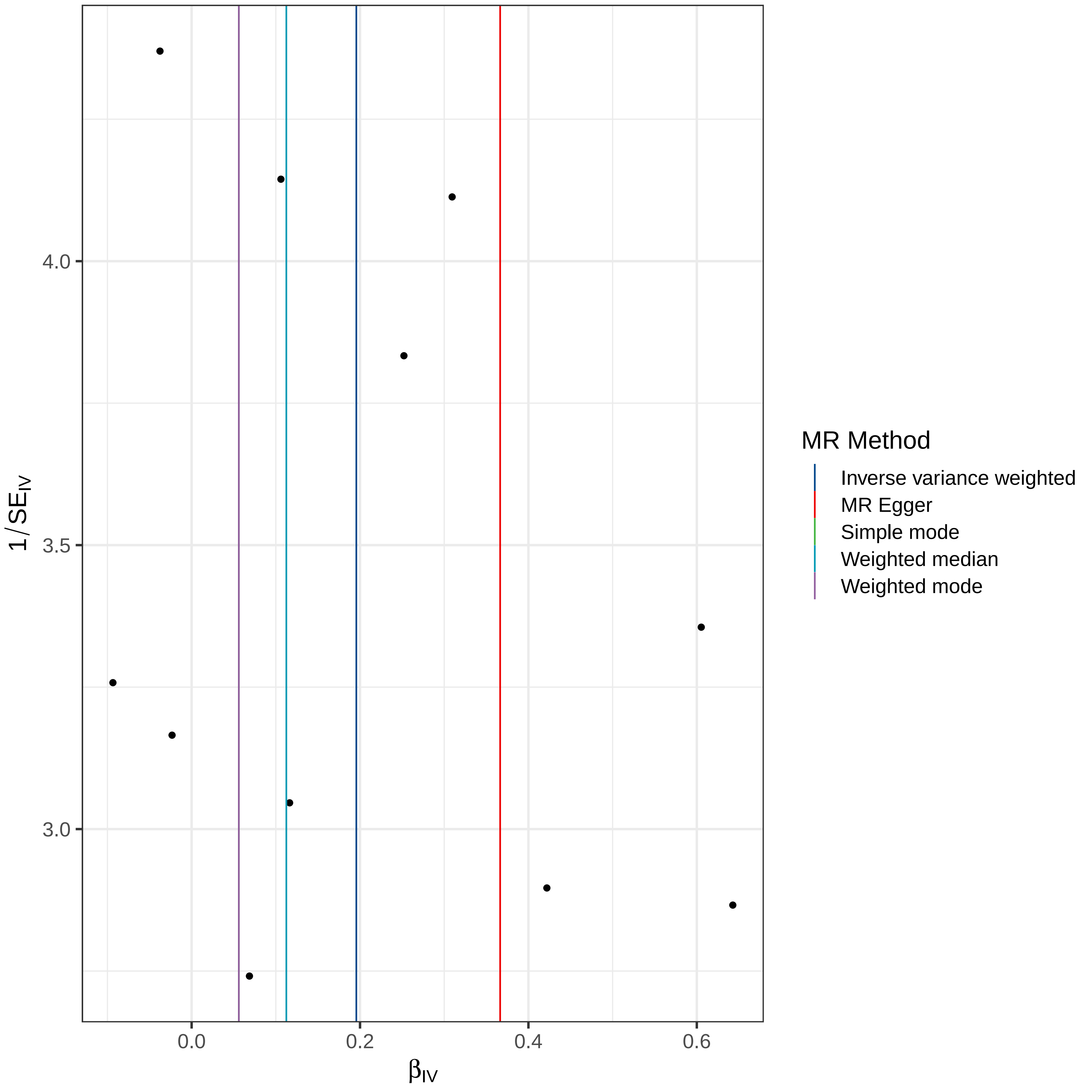

4.0
3.5
3.0
$1/SE_{IV}$
0.0
0.2
0.4
0.6
$\beta_{IV}$
MR Method
Inverse variance weighted
MR Egger
Simple mode
Weighted median
Weighted mode

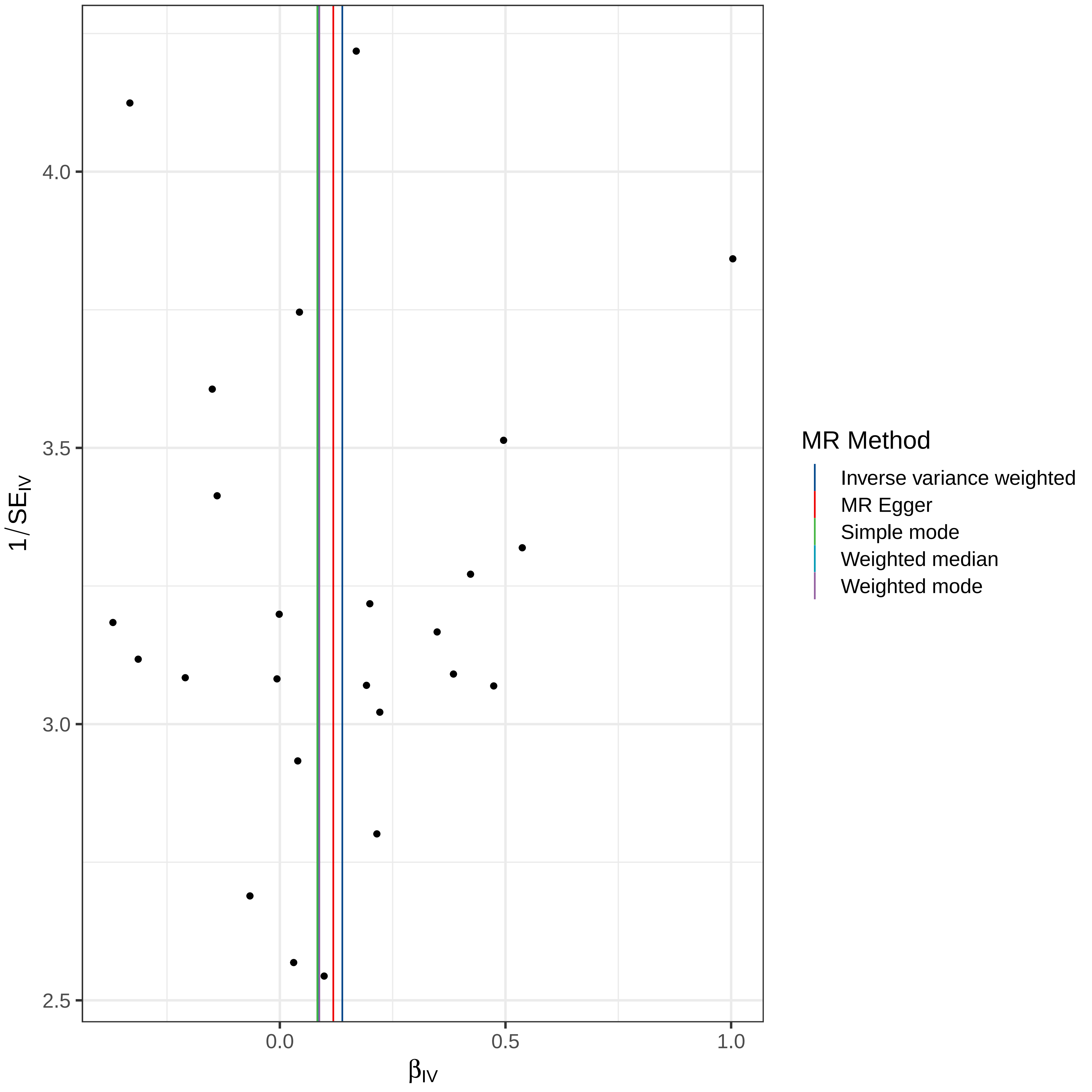

4.0
3.5
3.0
2.5
1/SE_IV
0.0
0.5
1.0
β_IV
MR Method
Inverse variance weighted
MR Egger
Simple mode
Weighted median
Weighted mode

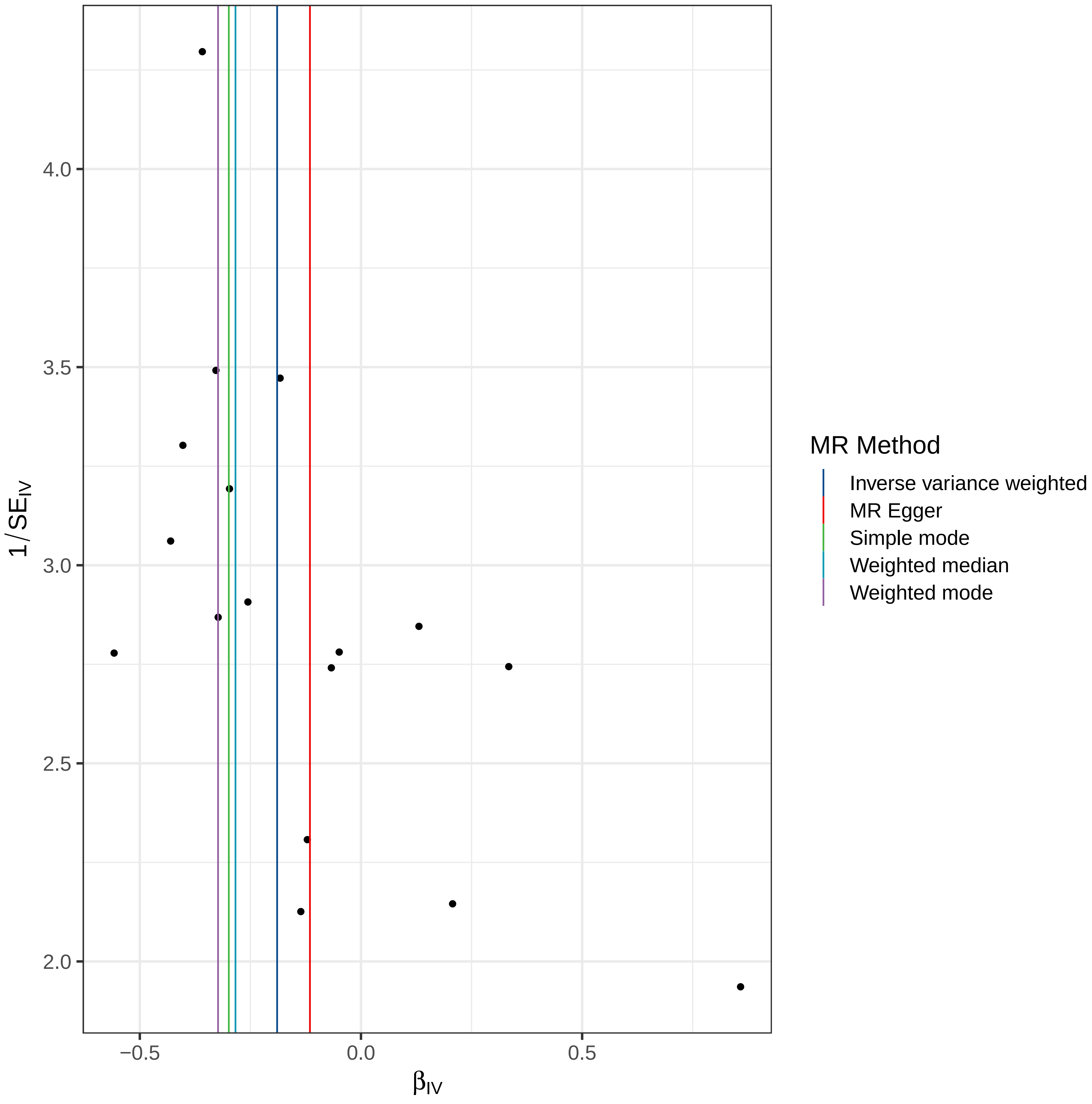

MR Method
Inverse variance weighted
MR Egger
Simple mode
Weighted median
Weighted mode
1/SE_IV
β_IV
4.0
3.5
3.0
2.5
2.0
−0.5
0.0
0.5

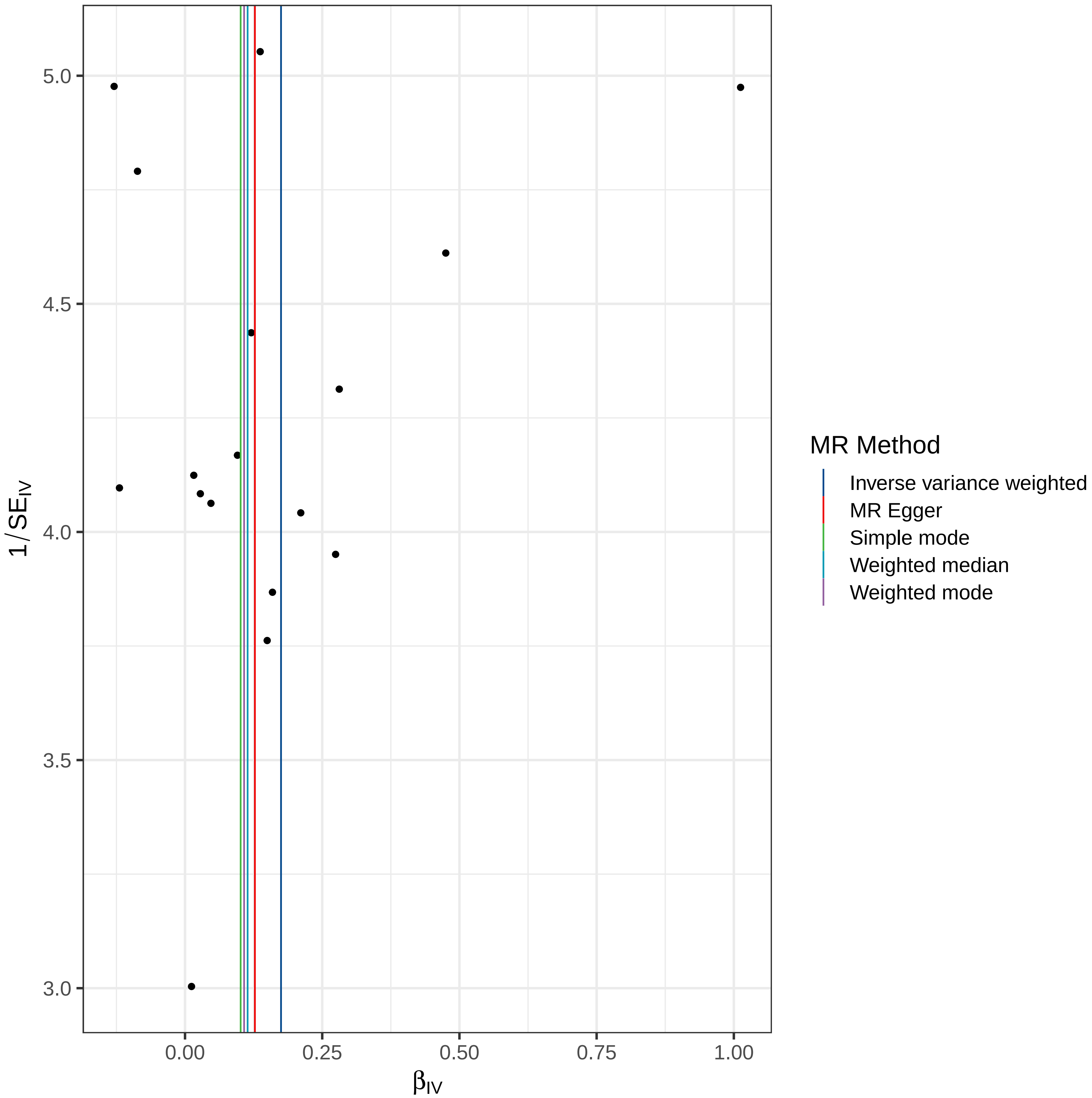

MR Method
Inverse variance weighted
MR Egger
Simple mode
Weighted median
Weighted mode
$1/SE_{IV}$
$\beta_{IV}$
5.0
4.5
4.0
3.5
3.0
0.00
0.25
0.50
0.75
1.00

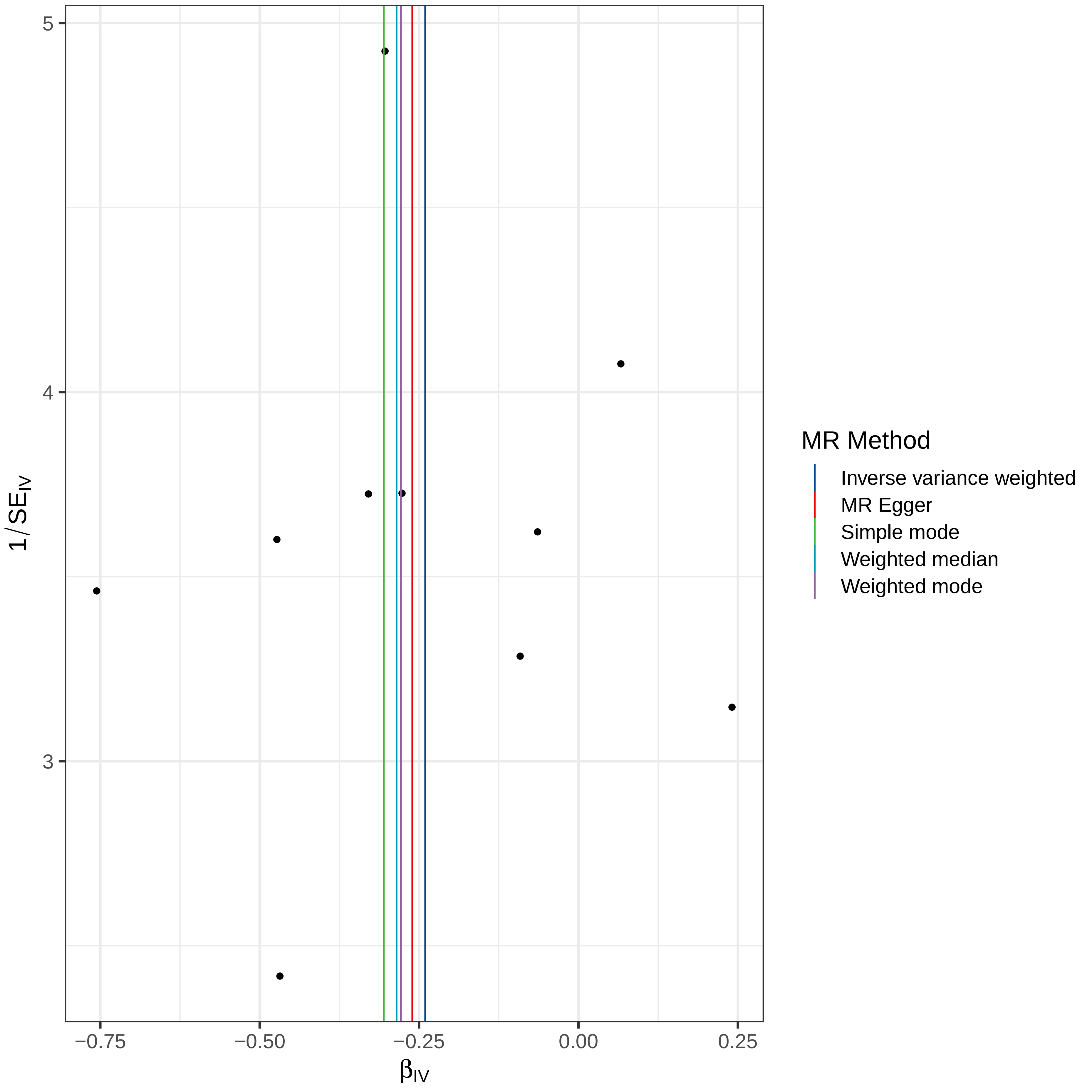

5
4
3
$1/SE_{IV}$
−0.75
−0.50
−0.25
0.00
0.25
$\beta_{IV}$
MR Method
Inverse variance weighted
MR Egger
Simple mode
Weighted median
Weighted mode

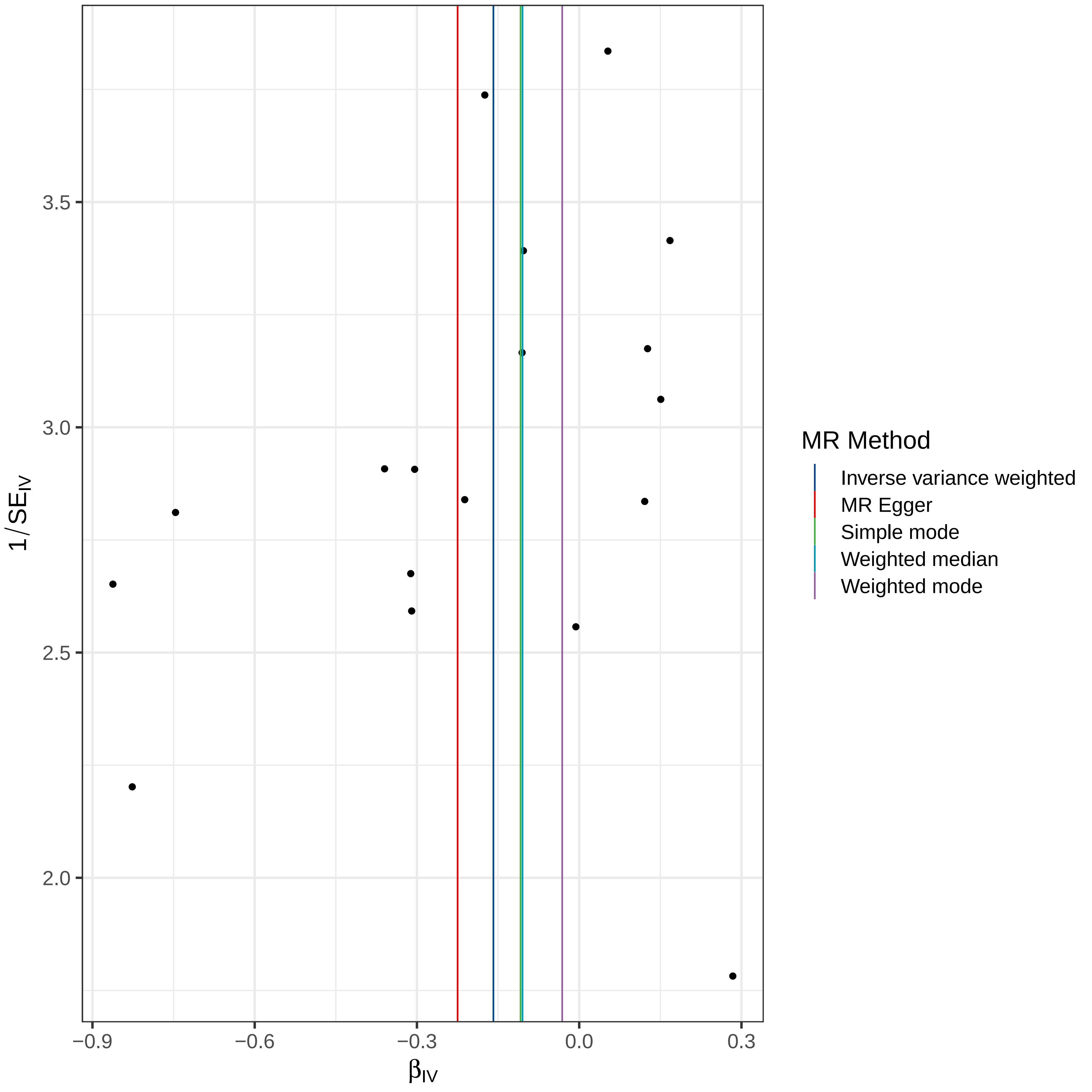

3.5
3.0
2.5
2.0
1/SE_IV
−0.9
−0.6
−0.3
0.0
0.3
β_IV
MR Method
Inverse variance weighted
MR Egger
Simple mode
Weighted median
Weighted mode

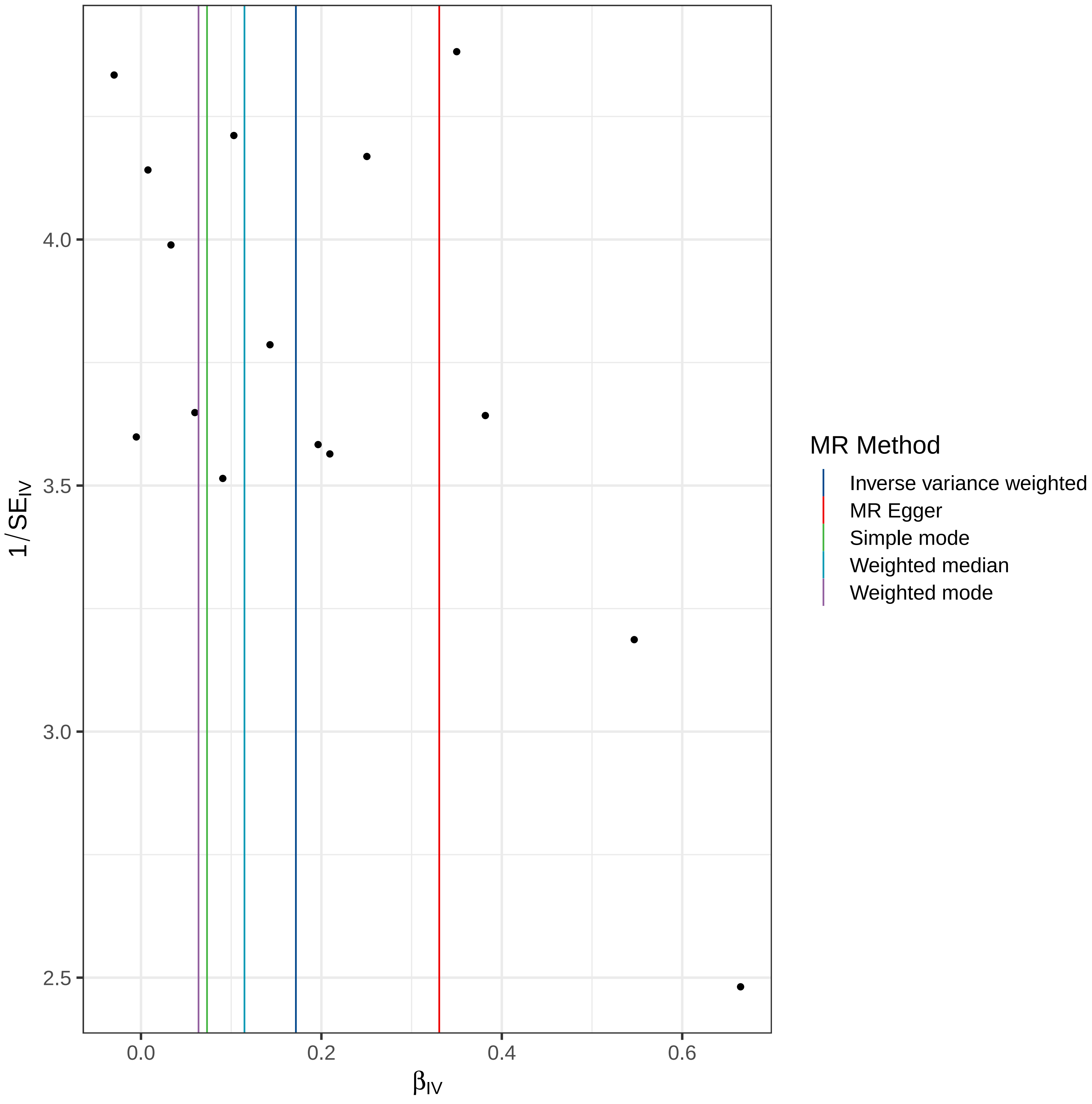

4.0
3.5
3.0
2.5
$1/SE_{IV}$
0.0
0.2
0.4
0.6
$\beta_{IV}$
MR Method
Inverse variance weighted
MR Egger
Simple mode
Weighted median
Weighted mode

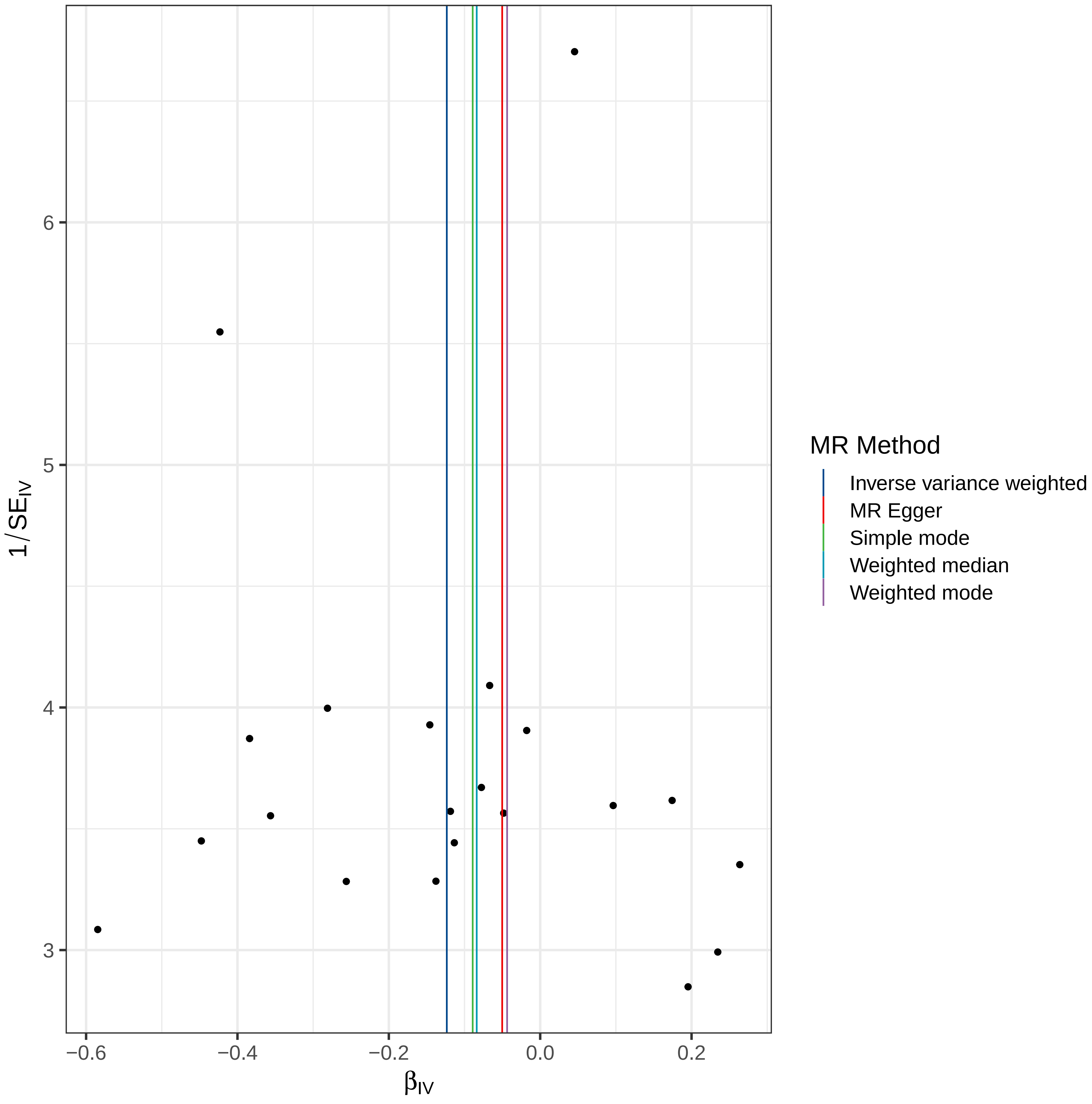

6
5
4
3
$1/SE_{IV}$
−0.6
−0.4
−0.2
0.0
0.2
$\beta_{IV}$
MR Method
Inverse variance weighted
MR Egger
Simple mode
Weighted median
Weighted mode

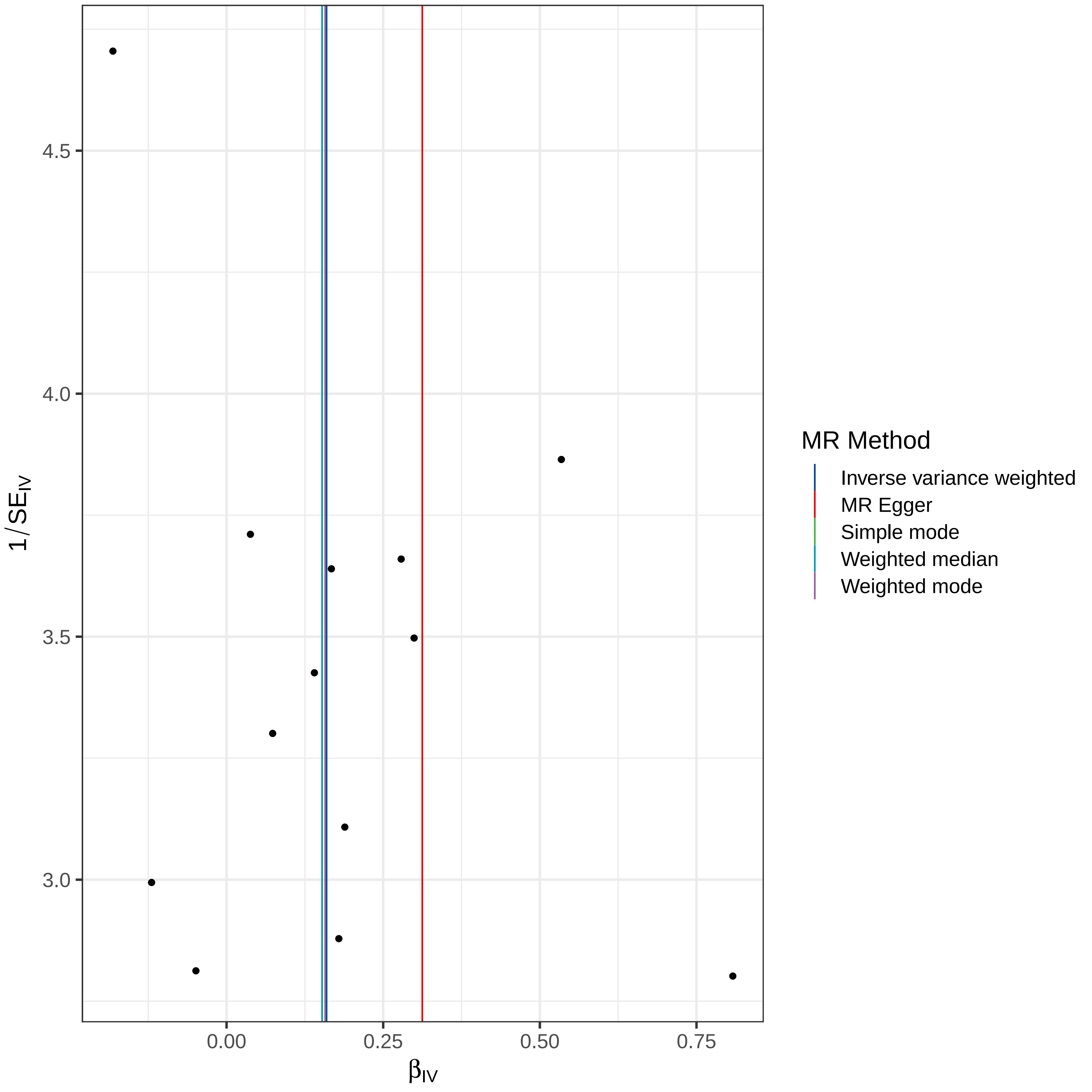

4.5
4.0
3.5
3.0
$1/SE_{IV}$
0.00
0.25
0.50
0.75
$\beta_{IV}$
MR Method
Inverse variance weighted
MR Egger
Simple mode
Weighted median
Weighted mode

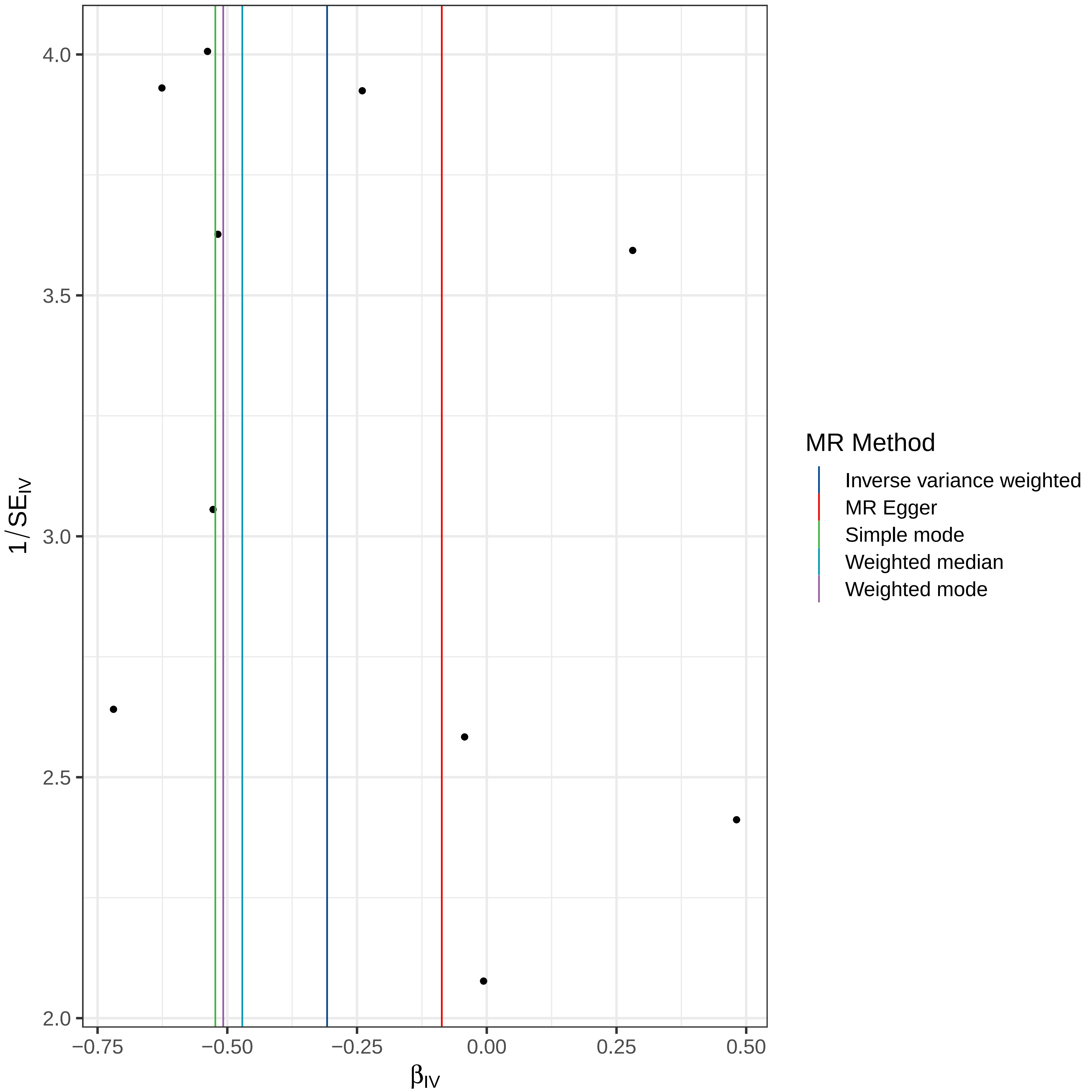

1/SE$_{IV}$
4.0
3.5
3.0
2.5
2.0
−0.75
−0.50
−0.25
0.00
0.25
0.50
β$_{IV}$
MR Method
Inverse variance weighted
MR Egger
Simple mode
Weighted median
Weighted mode

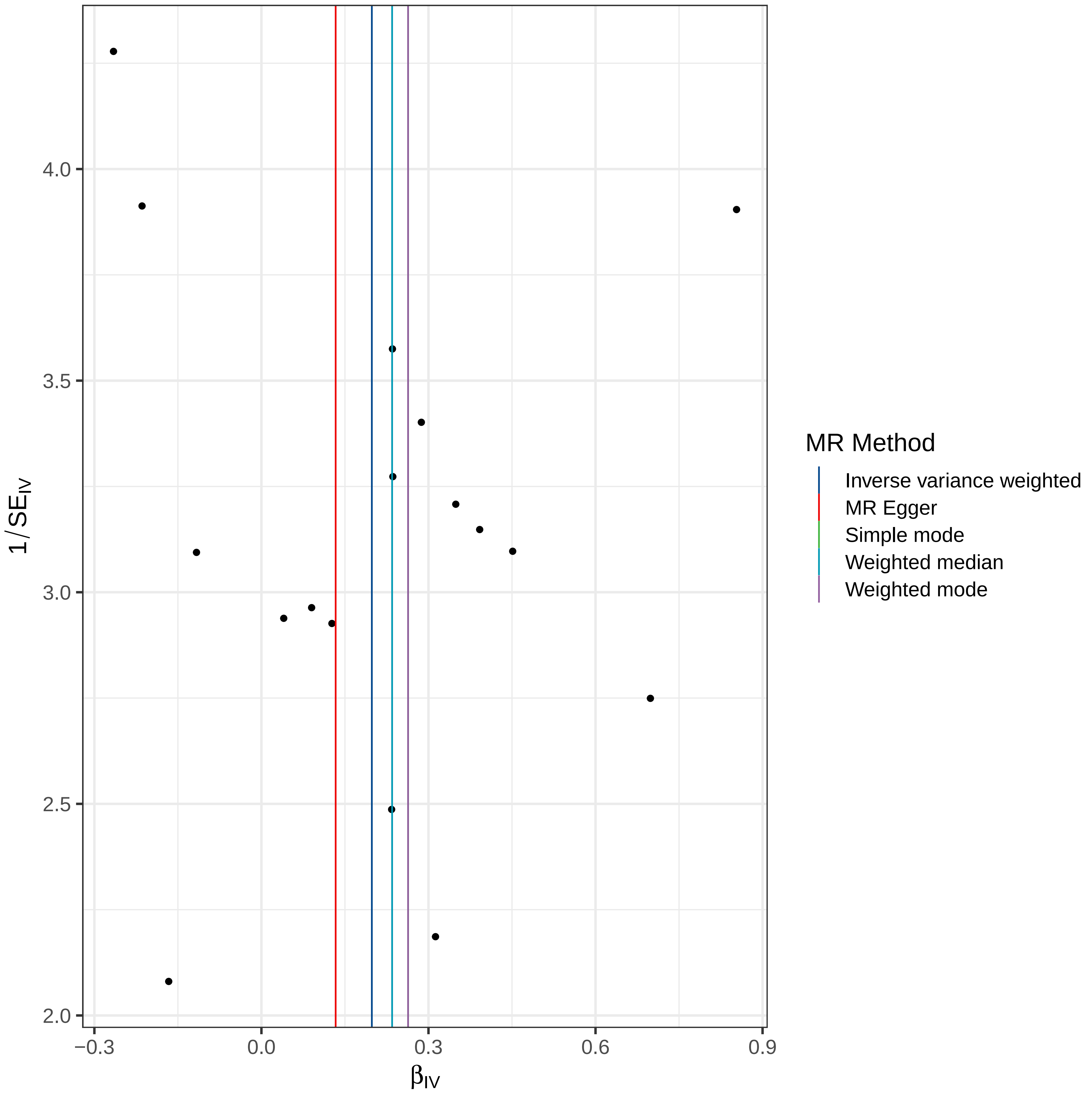

4.0
3.5
3.0
2.5
2.0
1/SE$_{IV}$
−0.3
0.0
0.3
0.6
0.9
β$_{IV}$
MR Method
Inverse variance weighted
MR Egger
Simple mode
Weighted median
Weighted mode

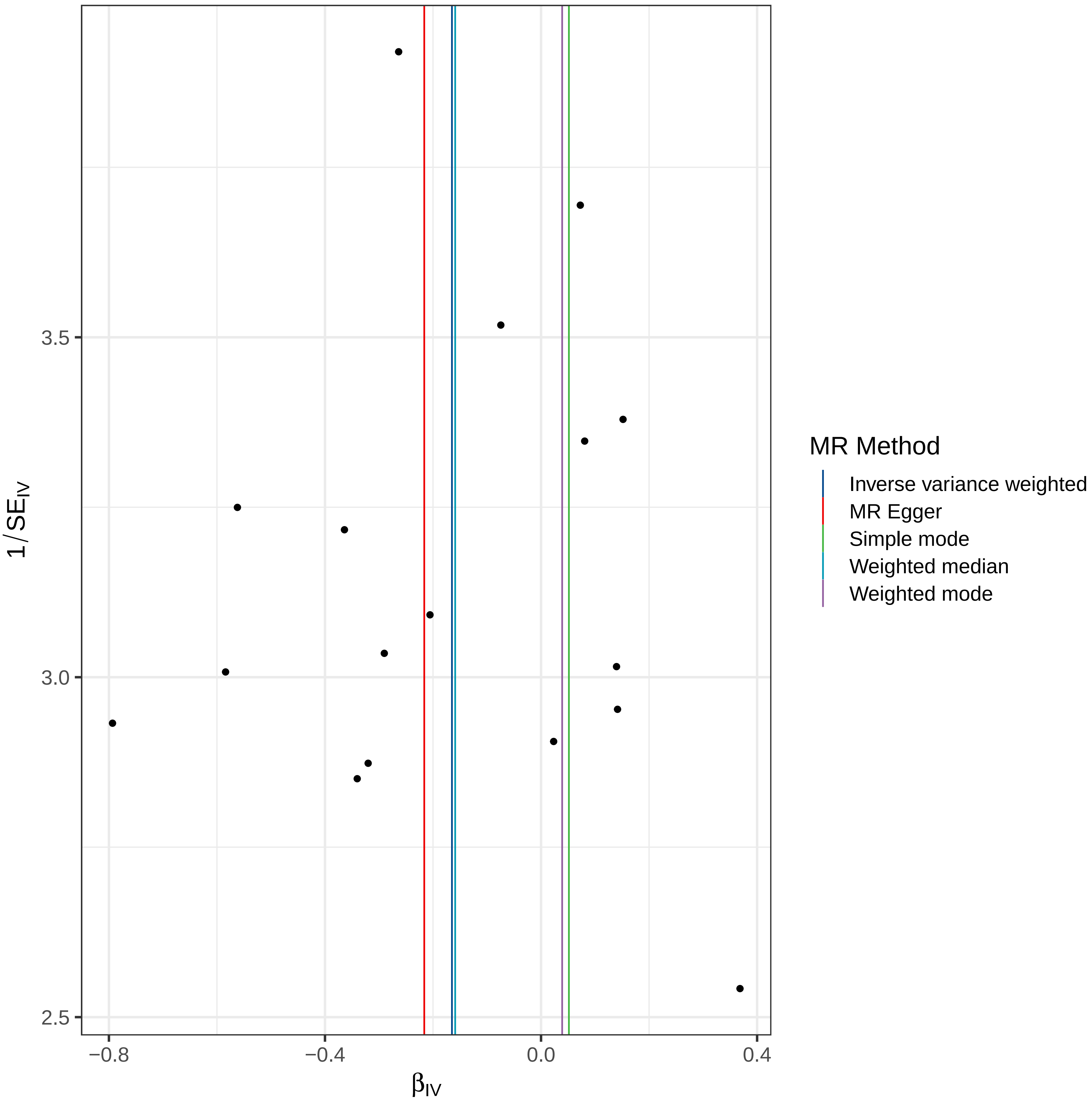

MR Method
Inverse variance weighted
MR Egger
Simple mode
Weighted median
Weighted mode
$1/SE_{IV}$
3.5
3.0
2.5
−0.8
−0.4
0.0
0.4
$\beta_{IV}$

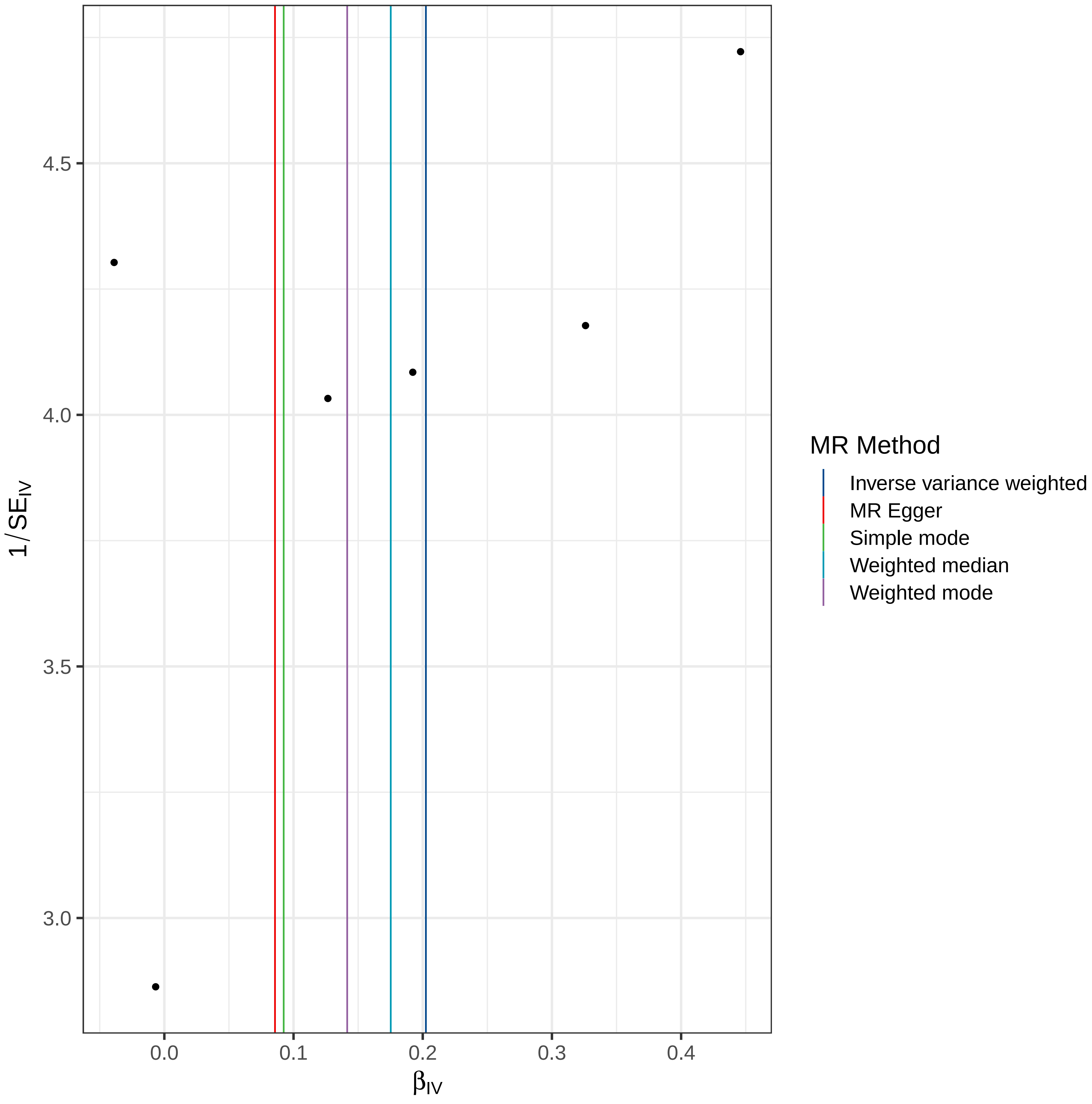

4.5
4.0
3.5
3.0
1/SE_IV
0.0
0.1
0.2
0.3
0.4
β_IV
MR Method
Inverse variance weighted
MR Egger
Simple mode
Weighted median
Weighted mode

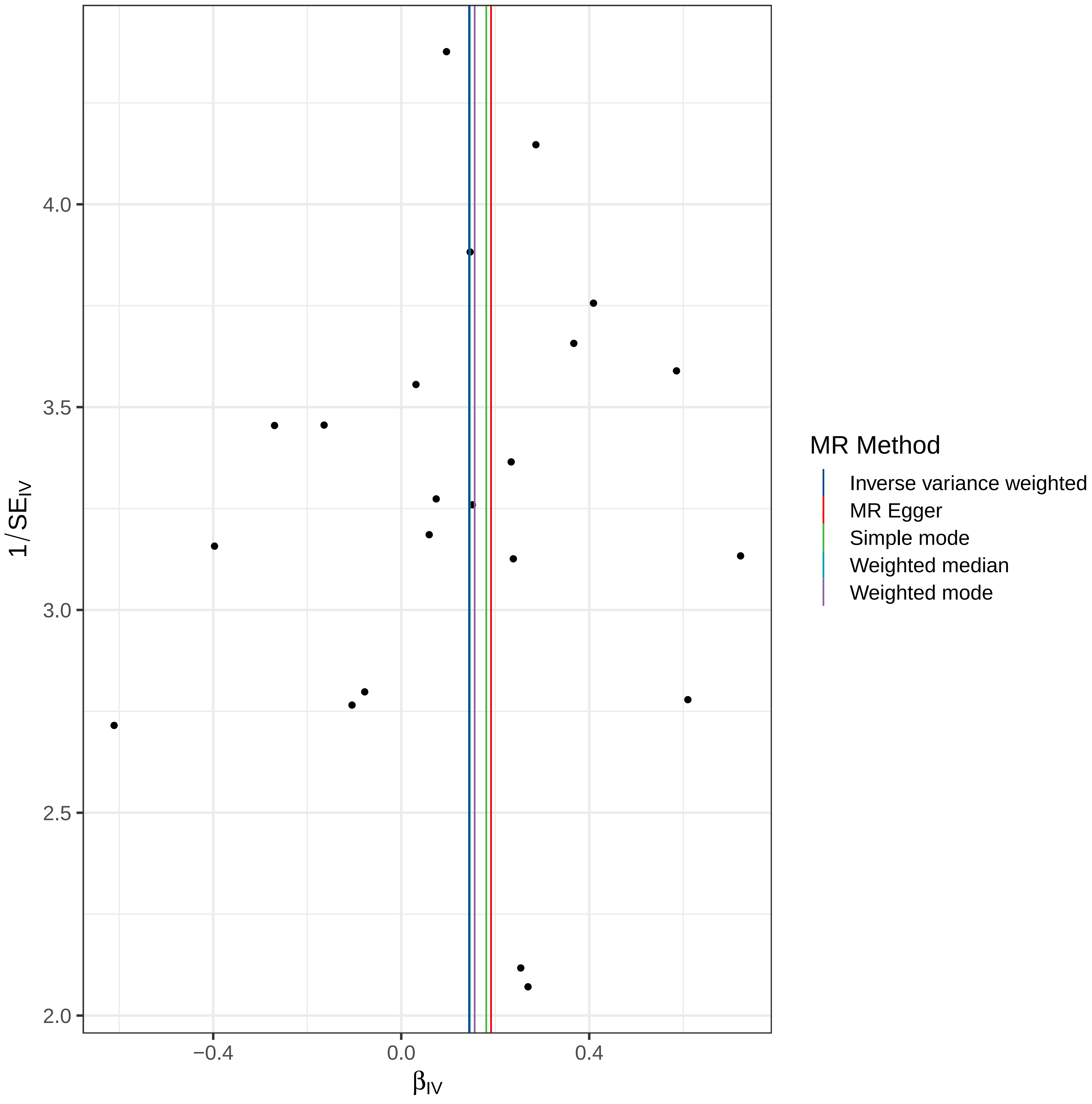

4.0
3.5
3.0
2.5
2.0
1/SE$_{IV}$
−0.4
0.0
0.4
β$_{IV}$
MR Method
Inverse variance weighted
MR Egger
Simple mode
Weighted median
Weighted mode

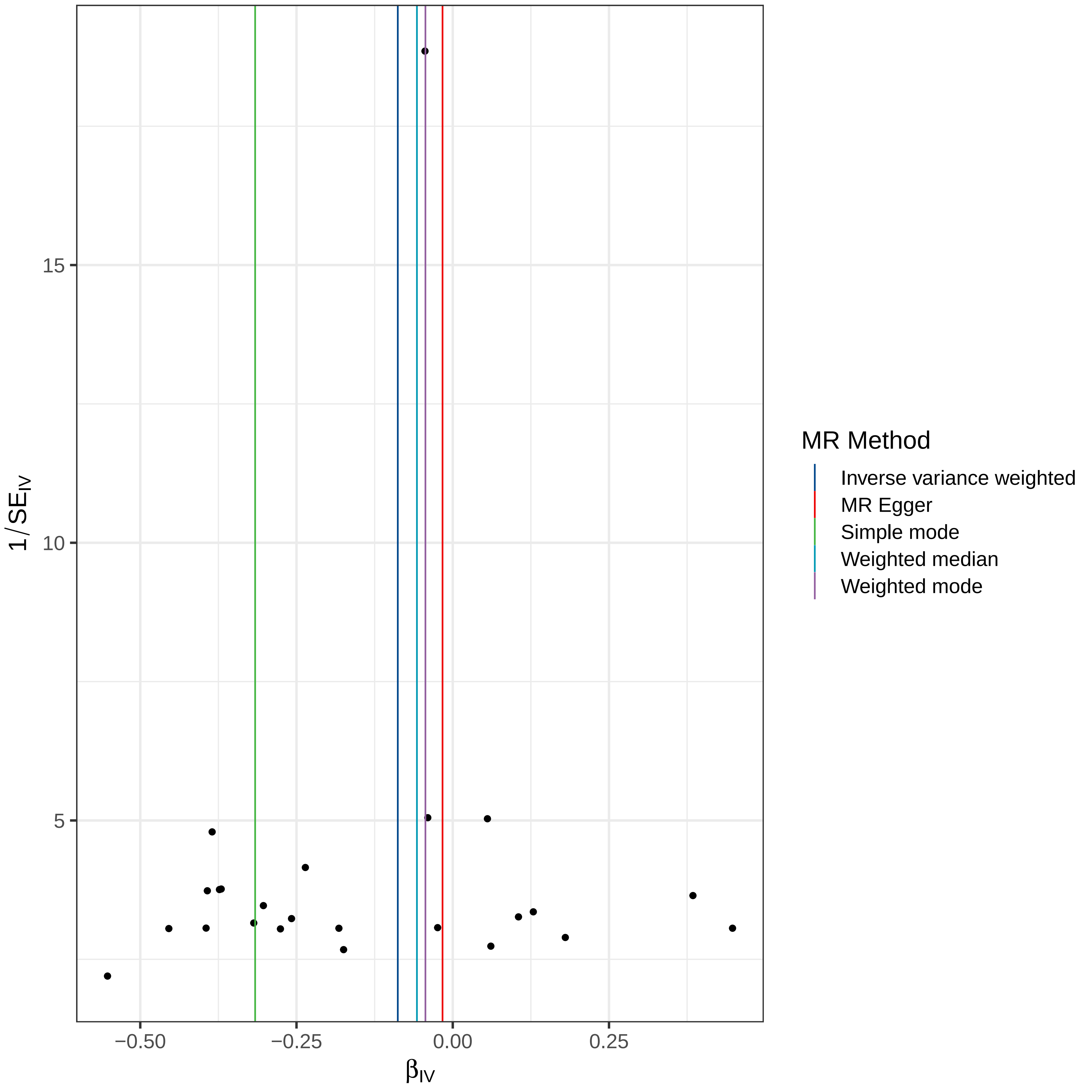

15
10
5
1/SE_IV
−0.50
−0.25
0.00
0.25
β_IV
MR Method
Inverse variance weighted
MR Egger
Simple mode
Weighted median
Weighted mode

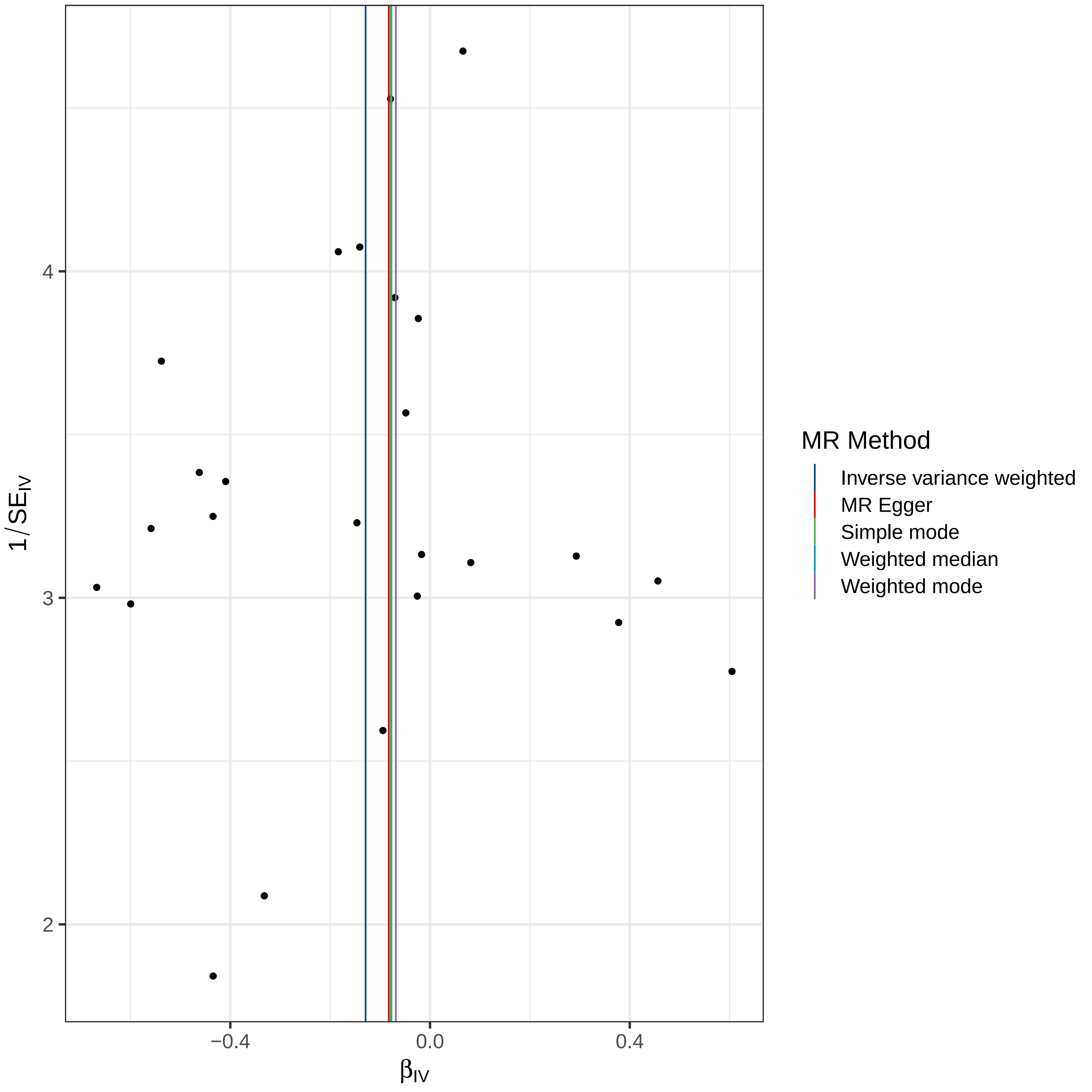

4
3
2
1/SE_IV
−0.4
0.0
0.4
β_IV
MR Method
Inverse variance weighted
MR Egger
Simple mode
Weighted median
Weighted mode

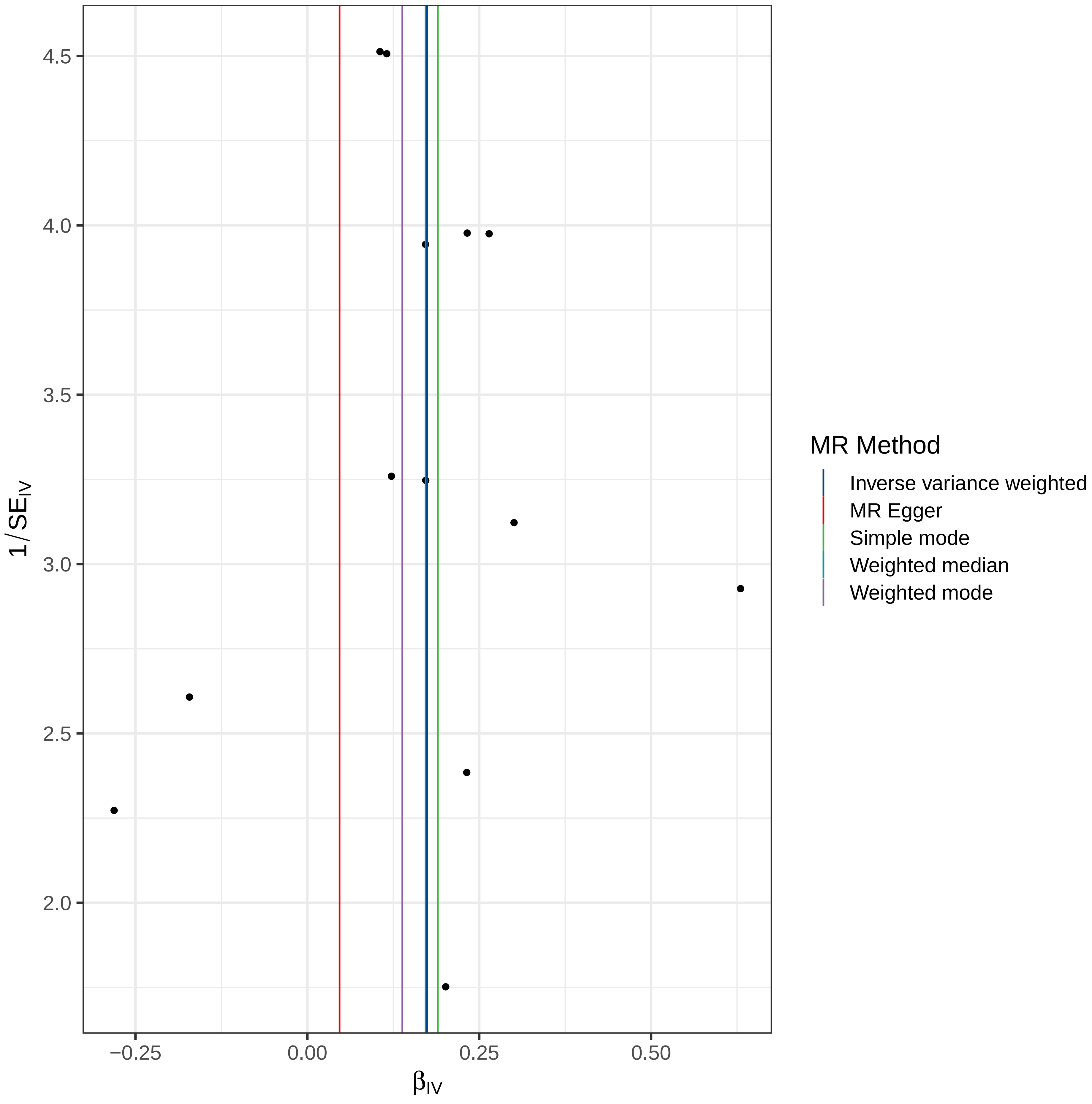

MR Method
Inverse variance weighted
MR Egger
Simple mode
Weighted median
Weighted mode
1/SE_IV
β_IV
4.5
4.0
3.5
3.0
2.5
2.0
−0.25
0.00
0.25
0.50
